# Genomic surveillance of SARS-CoV-2 in the state of Paraná, Southern Brazil, reveals the cocirculation of the VOC P.1, P.1-like-II lineage and a P.1 cluster harboring the S:E661D mutation

**DOI:** 10.1101/2021.07.14.21260508

**Authors:** Mauro de Medeiros Oliveira, Michelle Orane Schemberger, Andreia Akemi Suzukawa, Irina Nastassja Riediger, Maria do Carmo Debur, Guilherme Becker, Paola Cristina Resende, Tiago Gräf, Eduardo Balsanelli, Valter Antônio de Baura, Emanuel Maltempi de Souza, Fábio de Oliveira Pedrosa, Lysangela Ronalte Alves, Lucas Blanes, Sheila Cristina Nardeli, Alessandra De Melo Aguiar, Letusa Albrecht, Dalila Zanette, Andréa Rodrigues Ávila, Luis Gustavo Morello, Fabricio Klerynton Marchini, Hellen Geremias dos Santos, Fabio Passetti, Bruno Dallagiovanna, Helisson Faoro

## Abstract

We report a genomic surveillance of SARS-CoV-2 lineages circulating in Paraná, Southern Brazil, from March 2020 to April 2021. Our analysis, based on 333 genomes, revealed that the first variants detected in the state of Paraná in March 2020 were the B.1.1.33 and B.1.1.28 variants. The variants B.1.1.28 and B.1.1.33 were predominant throughout 2020 until the introduction of the variant P.2 in August 2020 and a variant of concern (VOC), P.1, in January 2021. Phylogenetic analyses of the SARS-CoV-2 genomes that were previously classified as the VOC P.1 lineage by PANGO showed that some genomes from February to April 2021 branched in a monophyletic clade and that these samples grouped together with genomes recently described with the lineage P.1-like-II. An extended phylogenetic analysis, including SARS-CoV-2 genomes from all over Brazil, showed that the P.1-like-II lineage appears at a high frequency in the southern region of the country. The P.1-like-II lineage genomes share some, but not all, defining mutations of the VOC P.1. For instance, it has the previously described ORF1a:D2980H and N:P383 L unique mutations and the newly detected ORF1a:P1213 L and ORF1b:K2340N mutations. Additionally, a new mutation (E661D) in the spike (S) protein has been identified in nearly 10% of the genomes classified as the VOC P.1 from Paraná in March and April 2021. We also report the identification of the S:W152C mutation in one genome from Paraná, classified as the N.10 variant. Finally, we analyzed the correlation between the lineage and the P.1 variant frequency, age group (patients younger or older than 60 years old) and the clinical data of 86 cases from the state of Paraná. This analysis does not support an association between the P.1 variant prevalence and COVID-19 severity or age strata. Our results provided a reliable picture of the evolution of the SARS-CoV-2 pandemic in the state of Paraná characterized by the dominance of the P.1 strain, as well as a high frequencies of the P.1-like-II lineage and the S:E661D mutations. Epidemiological and genomic surveillance efforts should be continued to unveil the biological relevance of the novel mutations detected in the VOC P.1 in Paraná.

## Introduction

Coronavirus disease 2019 (COVID-19) is caused by severe acute respiratory syndrome coronavirus 2 (SARS-CoV-2). Brazil is the third most affected country in the world, with over 16.5 million cases and over 462,000 deaths (https://covid19.who.int/). To date, Paraná has almost 1 million laboratory-confirmed COVID-19 cases and more than 195.9 deaths per 100,000 habitants (13). On the other hand, immunization efforts have progressed slowly: only 10% of Paraná citizens have been fully vaccinated thus far (17). The state of Paraná is strategically located in southern Brazil due to its borders with other Brazilian states (São Paulo, Santa Catarina and Mato Grosso do Sul) and with Argentina and Paraguay. This scenario of uncontrolled coronavirus spread may favor the emergence of potentially more contagious variants. Genomic surveillance efforts have been allowed to monitor the emergence and spread of new SARS-CoV-2 variants worldwide. Two lineages, the B.1.1.28 and B.1.1.33 variants, were dominant during the first wave of COVID-19 in Brazil (6, 35). However, the number of people infected with the VOC P.1, a ramification of the B.1.1.28 lineage first detected in Manaus (northern Brazil) that harbors the E484K and N501Y mutations in the spike (S) protein (20), has grown rapidly since December 2020 and was thought to be responsible for the deadly second wave of COVID-19 throughout Brazil (29, 30, 37). Compared to the first wave, more cases and deaths were registered in a shorter period of time during the second wave. According to official Brazilian Ministry of Health data, from the beginning of the pandemic until the end of November 2020, 282,645 cases of infection were registered in Paraná, which resulted in 6,160 deaths (11). Between December 2020 and April 2021, an additional 663,528 cases and 16,160 deaths were recorded (12). Although many factors may have contributed to this scenario, the introduction and spreading of new SARS-CoV-2 variants in the state of Paraná and the relationship between case severity and the prevalence of the VOC P.1 have not yet been investigated.

In this study, we carried out an analysis of 333 SARS-CoV-2 genomes of strains circulating in Paraná from February 2020 to April 2021. Our results demonstrate a cocirculation of the dominant VOC P.1 and the recently described P.1-like-II lineage (21). Notably, a new S:E661D mutation present in approximately 10% of the VOC P.1 genomes from March and April 2021 was identified. Mutations in the S protein are of particular interest because they may favor viral immune evasion and/or alter the efficacy of viral-cell interactions. Finally, we address the relationship between the prevalence of the P.1 variant, the severity of the cases and the age group strata.

## Materials and Methods

### Selection and recovery of samples

The 333 genomic sequences of SARS-CoV-2 from March 2020 to April 2021 were generated as part of the genomic surveillance carried out by Fiocruz in Brazil Genomahcov Fiocruz. The genomic sequences specifically identified in April 2021 were used to investigate the relationship between the frequency of the P.1 variant, the age group (patients younger or older than 60 years old) and the clinical severity (mild or severe cases), in addition to surveillance. For this purpose, SARS-CoV-2 samples were obtained from the two main laboratories responsible for the diagnosis of COVID-19 in the state of Paraná: the LACEN-PR (the Central Laboratory of the State of Paraná) and the LAC (COVID-19 Diagnostic Support, the Paraná State Unit - IBMP/Fiocruz). From the LACEN laboratory, samples of SARS-CoV-2 were taken from severe cases, while samples representing mild cases were obtained from the LAC. We aimed to obtain representative samples from all geographic areas of Paraná, and we considered the official division of the state into macroregions, each one with host cities: 1) EAST macroregion - host cities Curitiba and Paranaguá; 2) WEST macroregion - host cities Cascavel and Toledo; 3) NORTHWEST macroregion - host city Maringá; and 4) NORTH macroregion - host cities Londrina and Apucarana (Figure 1).

**Fig. 1.**
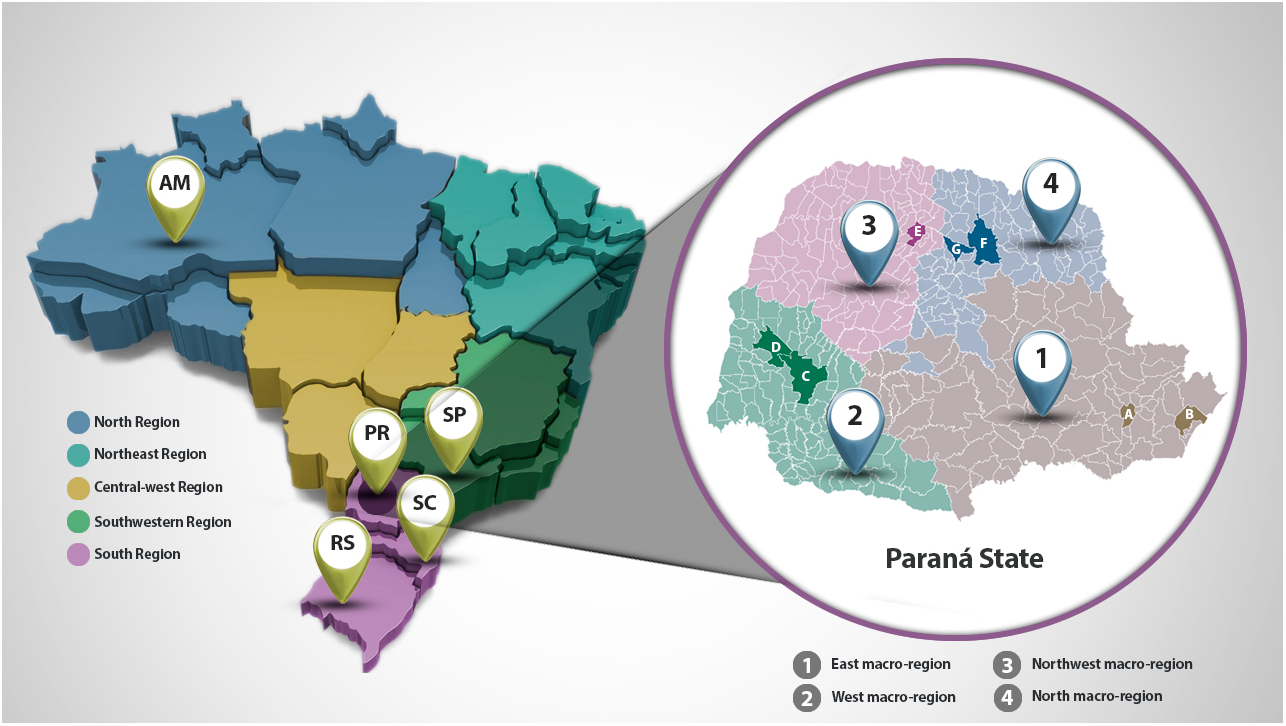
Sampling of SARS-CoV-2-positive patients was performed according to Paraná’s Secretary of Health official macroregional division. The map of Brazil is shown on the left. The color code represents the 5 regions of the country. The states of Amazonas (AM), São Paulo (SP), Paraná (PR), Santa Catarina (SC) and Rio Grande do Sul (RS) are indicated. The map of Paraná is shown on the right, and each macroregion was represented by at least one host city, as follows: (1) the east macroregion was represented by (A) Curitiba and (B) Paranaguá; (2) the west macroregion was represented by (C) Cascavel and (D) Toledo; (3) the northwest macroregion was represented by (E) Maringá; and (4) the north macroregion was represented by (F) Londrina and (G) Apucarana cities.

An equal allocation of samples were obtained in each clinical severity, host city/macroregion and age group strata. Thus, mild and severe cases were represented by 48 samples each, and a same number of samples were taken from each host city/microregion. Also, for each geographic region, an equal allocation of samples was used for each age strata (5 samples each). In the eastern macroregion, the municipality of Curitiba was considered separately from that of Paranaguá: the capital (Curitiba) contributed 20 samples (10 per laboratory and 5 per age group), and Paranaguá contributed 16 samples (8 per laboratory and 4 per age group). To assess whether there is an association between the frequency of the P.1 variant and the age group or the severity of COVID-19, we used a Fisher’s exact test at the 5% significance level. Additionally, we estimated the odds ratio (OR) and the corresponding 95% confidence interval (95% CI) considering age less than 60 years and mild case severity as the reference categories (OR=1.00).

### SARS-CoV-2 RNA purification and sequencing

Nasopharyngeal and pharyngeal swabs from infected patients (Ct < 25) were suspended in viral transport medium and maintained at -80 °C. Viral RNA was purified from these samples using the QIAamp Viral RNA Purification Kit (Qiagen). The purified RNA was converted to cDNA, and the viral genome was amplified by PCR using the Artic protocol (36). The genome was sequenced on the Illumina MiSeq platform in the 2×300 bp paired-end configuration. Variants were identified after read mapping in the reference genome for SARS-CoV-2 (RefSeq accession number NC_045512), and the analysis was performed on the Pangolin and Nextclade platforms. This study was approved by the Hospital do Trabalhador/SES/PR Ethics Committee: CAAE 31650020.5.0000.5225.

### SARS-CoV-2 genome assembly and variant calling

The bioinformatics analyzes were carried out at the Fiocruz Bioinformatic Platform Highz server (Fiocruz Rio - RPT04). The reads were trimmed with Trimmomatic v0.39 (5) and assembled using the tools SPAdes v3.14.1 (4) and MEGAHIT v1.2.9 (26). The scaffolds were built with RagTag v1.1.1 (1), ABACAS v1.3.1 (3) using the SARS-CoV-2 isolate Wuhan-Hu-1 as a template (NC_045512 RefSeq). Using this approach, we generated consensus sequences with a mean of 95% coverage (QUAST v5.0.2) (22). To assign the viral lineages, we used the nomenclature proposed by Rambaut et al (34), using the Pangolin standalone (pangolin v2.4.2 and pangoLEARN 2021-04-28). In parallel, we also assigned the viral lineages using the Nextclade web application (https://clades.nextstrain.org).

### Phylogenetic analysis

Phylogenetic analyses were performed using the 333 genomes generated as part of the Fiocruz COVID-19 Genomic Surveillance Network but also included other SARS-CoV-2 genomes from the state of Paraná and other states in Brazil that were deposited in the GISAID (Appendix S1). We retrieved all high-quality (< 5% of N) complete (> 29 kb) SARS-CoV-2 genomes of lineages B.1.1.28, B.1.1.33, P.1 and P.2 sampled in the state of Paraná, the B.1.1.28 lineage sampled in the state of Amazonas, and the P.1 variant sampled in the states of Amazonas, Santa Catarina, Rio Grande do Sul and from other regions of Brazil that were available on the GISAID (39) as of May 14, 2021. The set of sequences was divided into two datasets: 1) Paraná sequences (n=396) and 2) Paraná + Out Paraná sequences (n=354). The two groups of sequences were aligned using MAFFT v7.475 (24). The datasets were subjected to a maximum likelihood (ML) and a phylogenetic analysis using IQ-TREE v2.1.2 (31) under the GTR+G4+F nucleotide substitution model, and branch support was assessed by the approximate likelihood-ratio test based on the Shimodaira–Hasegawa-like procedure (SH-aLRT) with 1,000 replicates. Trees were visualized on the web application Interactive Tree Of Life (iTOL v5) (25). To highlight the mutations and categorize them by the frequency, genomic location and effect on protein sequences, we performed single nucleotide polymorphisms (SNPs) analysis with Snippy tools and Coronapp. The SNPs were assessed in each sample using the Wuhan-Hu-1 genome sequence as the reference. The results of the two tools were considered positive when the SNPs observed were predicted in the two tools.

### Time-scaled phylogenetic analysis

The time-scale phylogenetic tree was built using the Bayesian Markov Chain Monte Carlo (MCMC) approach implemented in BEAST 1.10.4 with the BEAGLE v3.1.0 library to optimize the computational time. The Bayesian tree was reconstructed using the HKY substitution model and the gamma distribution to account for rate heterogeneity across sites. The tree prior with parameter was set up using the coalescent Bayesian skyline model to create a molecular clock using the lognormal relaxed-clock model with an exponential substitution rate (8 × 10^*-*4^ substitutions/site/year). Two MCMC chains were run for 200 million generations and then were combined to ensure stationarity and good mixing. Convergence (effective sample size> 200) in parameter estimates was assessed using TRACER v1.7.2. The maximum clade credibility (MCC) tree was summarized with TreeAnnotator v1.10.4 ML, and MCC trees were visualized using FigTree v1.4.4 (18, 19). In parallel, we built phylogenetic trees on a time scale with the same SARS-CoV-2 samples using the TreeTime tool with default parameters (38). The time-scaled phylogenetic tree was constructed with up to 3 random sequences from each state in Brazil in each of the months of 2021 (Period 2021-01-01 to 2021-05-14; GISAID). Furthermore, the sequences were identified as the B.1.1.28 lineage from the Amazonas state (n=15) and the P.1-like-II lineage genome from different Brazilian states: Paraná (n = 20), Santa Catarina (n=30), São Paulo (n=14), Rio de Janeiro (n=5), Rio Grande do Sul (n=5), Minas Gerais (n=3), Espírito Santo (n=1) and Alagoas (n=1).

## Results

The analysis of the accumulated frequencies from March 2020 to April 2021 (Figure 2) showed the B.1.1.33 and B.1.1.28 variants as the main variants that predominated in the state in 2020.

**Fig. 2.**
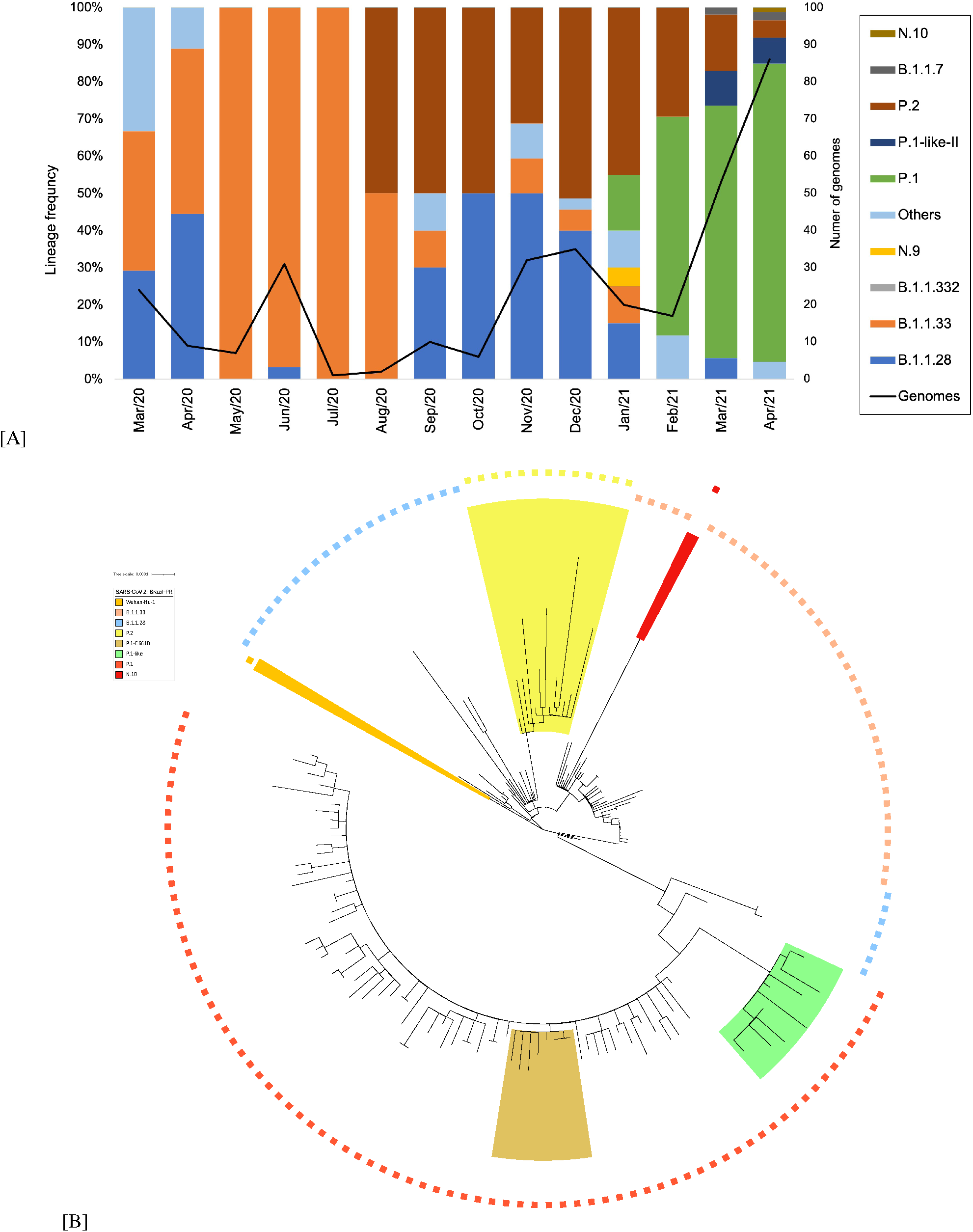
SARS-CoV-2 variants in Paraná state identified by genome sequencing. (**A**) Frequency and temporal distribution of the SARS-CoV-2 variant from February 2020 to April 2021. Information on the number of sequenced genomes and identification of variants from February 2020 to February 2021 were retrieved from the website genomahcov.fiocruz.br. The “Genomes” line indicates the number of representative sequences that were used to estimate the variant frequency at each time point. (**B**) Phylogenetic analysis of SARS-CoV-2 genomes identified in the state of Paraná. High-quality genomic sequences (n <95%) were aligned using Mafft software in the default configuration. The phylogenetic tree was built using the maximum likelihood method in IQTree software and visualized in IToL. The scale of the phylogenetic branches is given as substitutions per nucleotide site. Each symbol represents a SARS-CoV-2 lineage, as shown in the legend. The lineage N.10 is highlighted in red; the lineage P.2 is highlighted in light yellow; the P.1-like-II genomes are highlighted in light green; P.1 sequences harboring S:E661D are highlighted in shadow golden and Wuhan-Hu-1 is highlighted in orange.

Lineage P.2, derived from the B.1.1.28 variant, was identified in the state in August 2020 and had surpassed the previous two, reaching a frequency of 50% of the total genomes sequenced in September 2020 (Figure 2). This scenario started to change in December 2020 with the emergence of the VOC P.1 in the Brazilian state of Amazonas and the subsequent dissemination in the state of Paraná. The P.1 variant was identified in Paraná in January 2021 (15% of the genomes), and in February 2021, it already corresponded to 58% of the sequenced genomes, replacing the P.2 variant as the dominant variant (Figure 2). In April, the P.1 variant was identified in 87.2% of all the sequenced genomes.

To refine our knowledge about the prevalence of the VOC P.1 in the state of Paraná and its relationship with age strata (patients <60 or 60 years old) and clinical severity (mild or severe COVID-19 cases), we selected a cohort of 96 patients who tested positive for COVID-19 (Ct <25) in April 2021 to sequence the genome of the virus that infected them. Of the 86 genomes successfully sequenced, the variant P.1 was the most frequent (n = 75; 87.2%), followed by the P.2 variant (n = 4; 4.7%) (Figure S1). The VOC P.1 was predominant in all macroregions, ranging from 58.8% in the western region to 100% in Curitiba (eastern region), and in both age groups (Figure S1), although a higher diversity of variants was observed in the <60-year-old stratum (Figure S3). Regarding the sampling strata established, we did not observe any association between the frequency of the P.1 variant and age group (p-value 0.33) or the case severity (p-value 0.75) (Figure S3 and Figure S4) in this cohort.

A phylogenetic analysis using the 333 SARS-CoV-2 genomes of the samples from the state of Paraná confirmed the previous analyses (6, 35), with variants B.1.1.28 and B.1.1.33 being the most dominant in 2020 (Figure 2). We also identified a group of 11 genomes, 5 from March and 6 from April 2021, which were previously classified as the P.1 lineage by PANGO but truly constituted a separate clade related to the P.1 variant (Figure 2). The divergent group had lineage defining mutations compatible with the recently described lineage of the P.1-like-II lineage (21). These samples were mainly collected in the host cities Toledo and Cascavel from the West macroregion during the months of March and April 2020. Notably, in this macroregion, we found a higher P.1-like-II lineage prevalence (29.4%) and a lower P.1 variant (58.8%) prevalence in the cohort analysis conducted in April 2021 (Figure S2). No P.1-like-II lineage genomes were retrieved from the northern or northwestern regions of Paraná. To deepen the understanding of the presence of the P.1-like-II lineage in the state of Paraná, we recovered the genomic sequences identified as the P.1-like-II lineage in the other two states in the southern region of Brazil (21), Rio Grande do Sul (n = 4) and Santa Catarina (n = 30), as well as genomic sequences of the true VOC P.1 identified in these two states. Santa Catarina was of particular interest because of its geographical proximity to Paraná and because it was the Brazilian state where most of the P.1-like-II genomes were recovered in previous studies (21). We also included the P.1 variant genome sequences from the state of Amazonas, where it first emerged, and other P.1 variant genomes from Paraná available in the GISAID. The phylogenetic analysis involving this set of genomes supported the identification of representatives of the P.1-like-II lineage in the state of Paraná (Figure 3), since they grouped together with genomes previously identified as the P.1-like-II lineage from the states of Santa Catarina and Rio Grande do Sul but were separate from the branch composed of the VOC P.1 genomes (SH-aLRT =99.5%). Additionally, the number of the P.1 like-II lineage genomes from Paraná increased from 11 to 20 (Figure 3), with the additional genomes corresponding to samples from Mach and April. An independent cluster was comprised solely by the P.1-like-II lineage genomes from Paraná, which indicated a possible local transmission of this lineage.

**Fig. 3.**
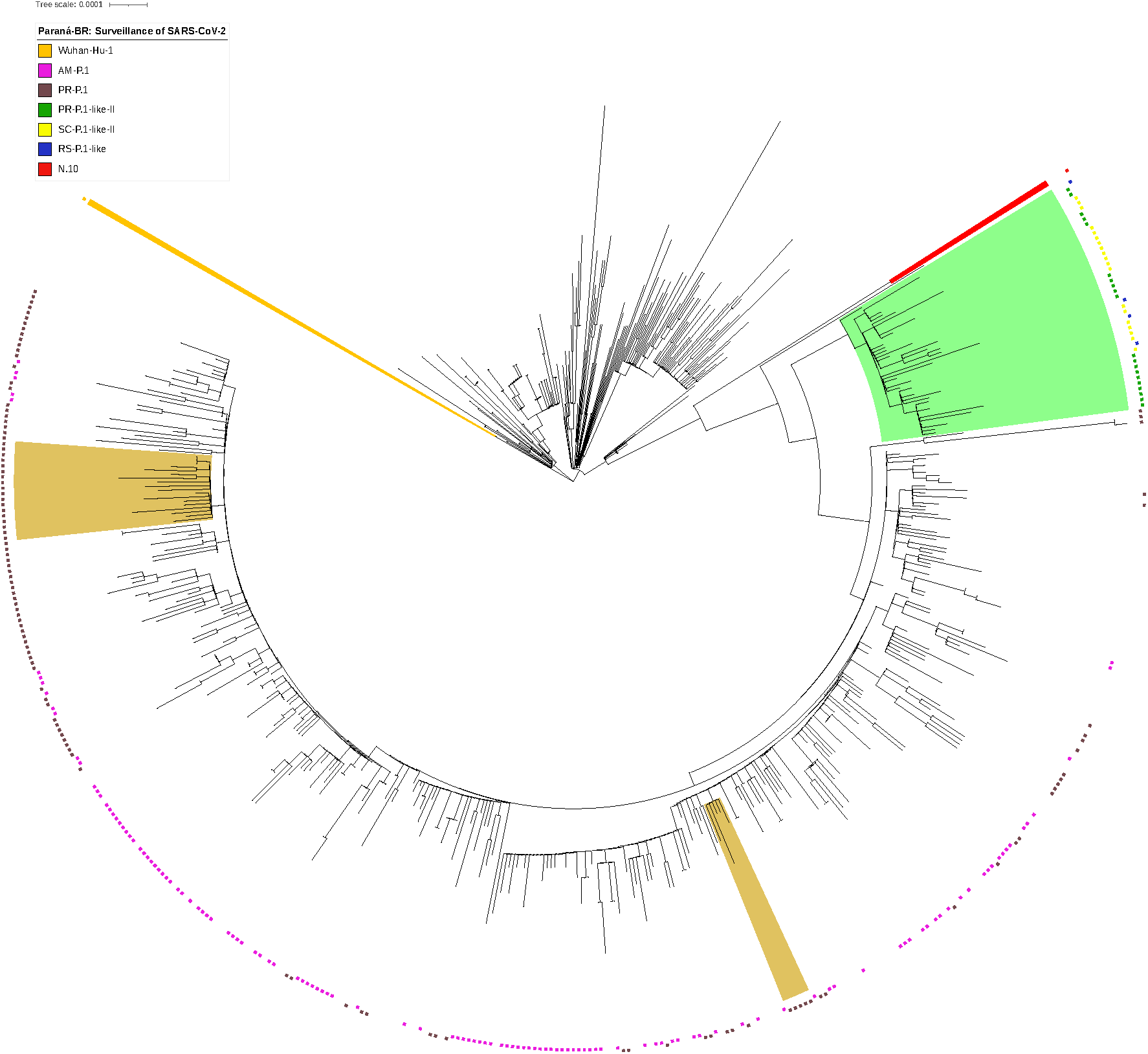
Phylogenetic tree of the VOC P.1 strain isolated in the State of Paraná from 2020-03-05 to 2021-04-28. High-quality genomic sequences (n <5%) were aligned using Mafft software in the default configuration. The phylogenetic tree was built using the maximum likelihood method in IQTree software and visualized in IToL. Phylogenetic tree highlighting the P.1 variant containing the sequences of Amazonas, Santa Catarina and Rio Grande do Sul and closely related sequences, including those of the subclad P.1-like-II. The aLRT support values are indicated in key branches, and the scale of the phylogenetic branches is given as substitutions per nucleotide site. Each symbol represents a SARS-CoV-2 lineage, as shown in the legend. The lineage N.10 is highlighted in red; the P.1-like-II genomes are highlighted in light green; P.1 sequences harboring S:E661D are highlighted in shadow golden and Wuhan-Hu-1 is highlighted in orange.

The current hypothesis is that the P.1 and P.1-like-II lineages diverged from a common ancestor (21) at a time when the mutations were arising and accumulating. To understand this question, we selected a representative group of genomes to perform a time-scaled phylogenetic tree analysis. The time-scaled phylogenetic tree (Figure 4) supports the hypothesis that the P.1-like-II and P.1 lineages have closer phylogenetic relationships than the other variants but belong to distinct clades and accumulate more mutations than the B.1.1.28 lineage from Amazonas. Our results also reinforce that the P.1-like-II lineage and the VOC P.1 probably diverged in late August/early September (Figure 4). Previous analysis based on SARS-CoV-2 genomes from Brazil obtained up until March 2021 indicated that the P.1-like-II lineage was mainly distributed in the south and southeast regions of Brazil, with a high prevalence in the state of Santa Catarina but with a unique representative from the state of Paraná (21). Here, including the finding of novel genomic sequences, we verified that the P.1-like-II lineage is also present in the western region of the state of Paraná at a relatively high frequency. Of note, the first representative of the P.1-like-II genome in Paraná was identified in February 2021, while the more recent genome was found in April 2021. From this analysis, we demonstrate that the genomes classified as the P.1-like-II lineage in the states of Paraná, Rio Grande do Sul and Santa Catarina emerged after the emergence of the P.1 variant. It would be expected that genomes referring to the P.1-like-II lineage were found among the sequenced genomes in the state of Amazonas, which did not occur.

**Fig. 4.**
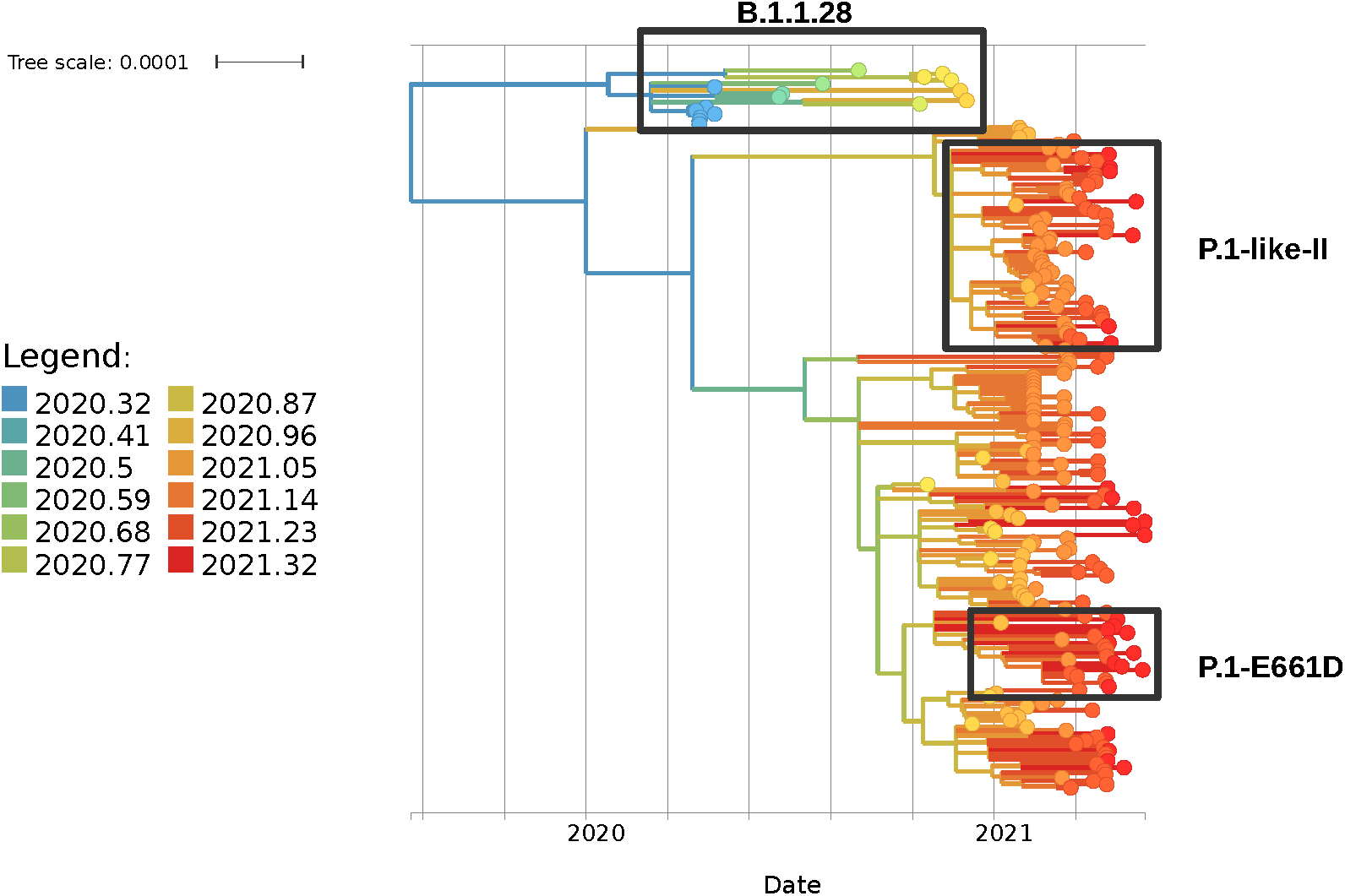
Time-scaled phylogenetic trees of SARS-CoV-2. The picture shows a time-stamped phylogeny of the P.1 lineage in the State of Paraná in comparison with sets of P.1 sequences from the States of Amazonas, Santa Catarina and Rio Grande do Sul. In the figure, the length of the branches corresponds to years and the number of mutations, and these are colored according to the legend of colors. In addition, the SARS CoV-2 reference sequence, the P.1-like-II sequence and the P.1 sequence group with the spike protein mutation S:E661D are highlighted.

The analyses of the phylogenetic tree constructed with the P.1 sequences from Paraná state (Figure 2) revealed the existence of a small cluster formed by only 8 P.1 genomes. The analysis of mutations in these 8 genomes revealed an additional substitution in the 661 amino acid residue of the S protein in this group of P.1 variants (Table 1). A glutamic acid residue is changed to an aspartic acid residue (S:E661D). When we exclusively considered the cohort of patients analyzed in April 2021, this mutation occurred in 8 of the 75 (10.8%) P.1 genomes from Paraná, with particular prevalence in the eastern and northern macroregions: approximately 25% of the VOC P.1 samples from Curitiba (5/20) and 3/12 samples from Londrina and Apucarana harbored the S:E661D muta tion. In March 2021, the number of P.1 genomes with this mutation was 3 of the 41 analyzed genomes (7.3%) (data not shown). This result suggests that the S:E661D mutation increases in frequency over time. Among the 8 cases identified in April 2021 carrying the P.1 variant with the E661D mutation, one death was reported, 6 patients were hospitalized for more than a month, and only one patient was cured. To determine the frequencies of the P.1-like-II lineage and the S:E661D mutation in other Brazilian states, we constructed a phylogenetic tree with all P.1 genomes sequenced in Brazil (n=3,648) (Figure 5).

**Table 1.**
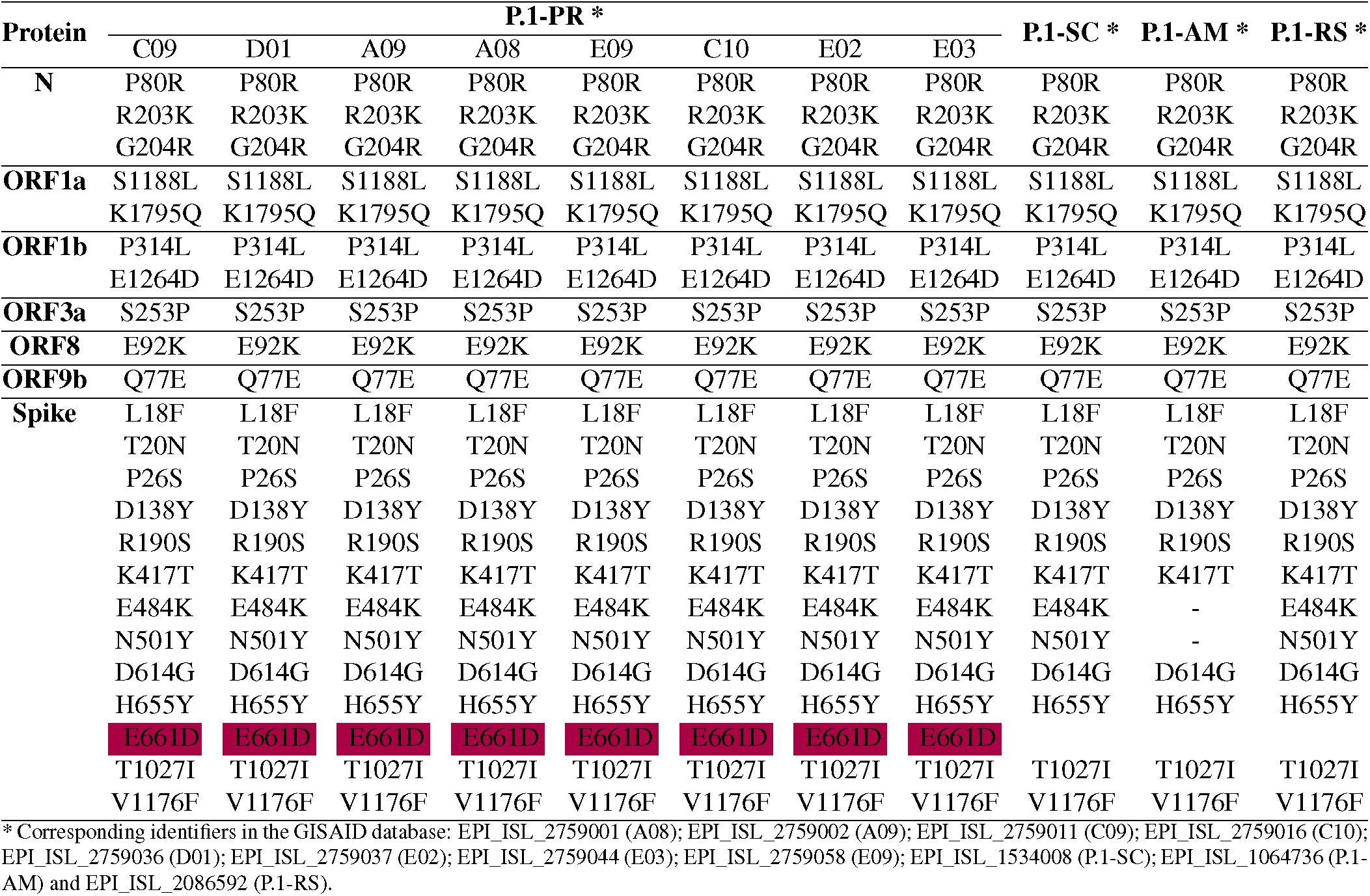
Comparison between mutations identified in variant P.1 described in Amazonas, Santa Catarina and Rio Grande do Sul (P.1-AM, P.1-SC and P.1-RS) and in eight P.1 samples from Paraná (P.1-PR).

**Fig. 5.**
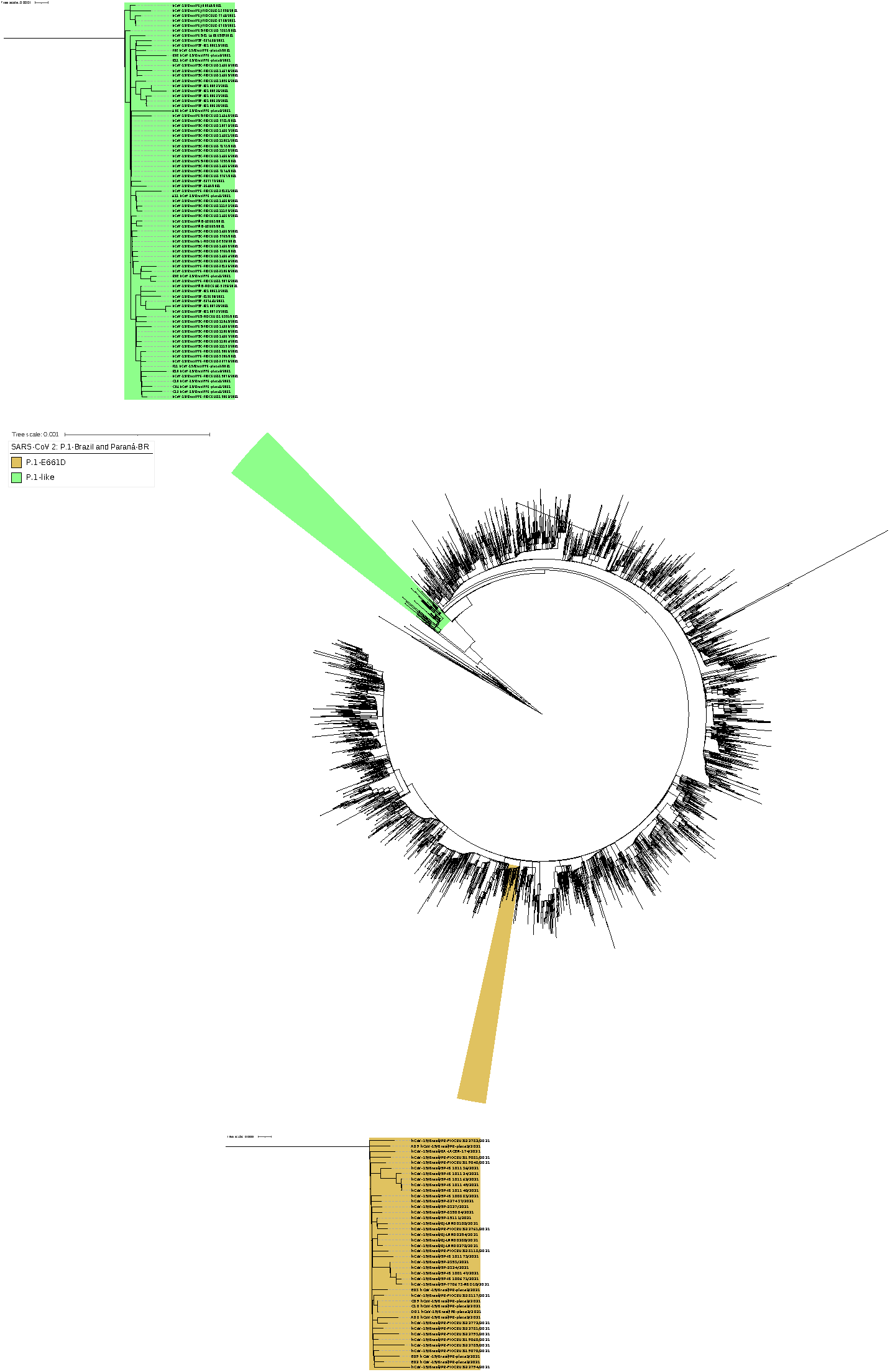
Phylogenetic tree of the Brazilian VOC P.1. VOC P.1 genomic sequences from all Brazilian states, produced by the Fiocruz genomic network or retrieved from GISAID, were aligned using MAFFT and used to construct a phylogenetic tree. Highlighted in the boxes are the genomes of the P.1-like-II lineage (green) and the P.1-E661D mutation (golden). The states from which the sequences originated were represented as PR-Paraná; SC-Santa Catarina; RS-Rio Grande do Sul; SP-São Paulo; RJ-Rio de Janeiro; MG-Minas Gerais; ES-Espírito Santo; AL-Alagoas; BA-Bahia.

As previously described (21), the P.1-like-II lineage is distributed mainly in the south and southeast regions of Brazil, with a higher prevalence in the states of Paraná and Santa Catarina. Related to the S:E661D mutation, we found, in addition to those genomes from Paraná, 25 genomes whose S protein contained the mutation, S:E661D, all from 2021.

These were from Minas Gerais (n=1), São Paulo (n=19), Bahia (n=1) and Rio de Janeiro (n=4) (Figure 5). When we considered the frequency in relation to the total number of P.1 variants identified, the E661D mutation appeared in 10.24% of the genomes from Paraná, followed by Minas Gerais (2.94%), Bahia (1.92%), São Paulo (0.90%) and Rio de Janeiro (0.86%). We also highlight the identification of 2 high-quality genomes of the variant B.1.1.7, first identified in the United Kingdom, and one high-quality genome of the N.10 variant, recently identified in the Brazilian state of Maranhão (32). The N.10-MA variant (a term used in this article to differentiate it from the variant found in Paraná, N.10-PR) was described as a variant of interest (VOI), was derived from the B.1.1.33 variant, and carried important mutations in the S protein, including the S:V445A and S:E484K mutations. A more detailed analysis of the mutations that occurred in the N.10-PR variant revealed one additional mutation in the S protein, S:W152C, which is absent in the N.10-MA variant. The S:W152C mutation occurs in the N-terminal domain (NTD) of the S protein and was previously described in another VOI, named B.1.429 (CAL.20C) (Table S1). Of note, there was no evidence for the circulation of the B.1.617 variant (first observed in India) or the B.1.351 variant (first observed in South Africa) in Paraná. However, in June 2021, the State Secretary of Health of Paraná reported the identification of 3 cases of COVID-19 from Apucarana, which were caused by the variant B.1.617 (14). These infections occurred in April (the same period of sample collection of this study), which suggests that the VOC B.1.617 was already present but had not broadly disseminated in the state by that time.

## Discussion

In this work, we performed an analysis of the evolution of the SARS-CoV-2 pandemic in the state of Paraná, Southern Brazil. Our study was based on 333 genomes identified between March 2020 and April 2021 by the Genomic Surveillance Network from Fiocruz and other sequences available in the GISAID database (Appendix S1). As in other Brazilian states, the year 2020 was marked by the predominance of the variants B.1.1.28 and B.1.1.33 and, from August 2020 onwards, by the VOI P.2 (6, 35). From January 2021 and in subsequent months, the VOC P.1 emerged in the state and spread fast, reaching 87.2% of all genomes in April 2021. No other variant introduced in Brazil has thus far reached such high prevalence values in the state of Paraná. This may be related to the low social isolation rates observed in Paraná, as well in Brazil, during the COVID-19 pandemic (23). Our analysis reveals that the P.1 variant, first identified in January 2021 in Japanese travelers returning from a trip to the Amazon (20), was already circulating in the state of Paraná by that time. Additionally, in January 2021, genomes of the VOC B.1.1.7, which are dominant in the United Kingdom (2), were identified in the state of Paraná. Despite the presence of two VOCs in the state of Paraná, it is noticeable that variant B.1.1.7 did not replace the most frequent variant or even spread in the state and remained at low frequencies. The dominance of the P.1 variant, even in the presence of another VOC, may be related to the fact that P.1 variant infections are associated with higher viral loads in the upper respiratory tract when compared to non-P.1 variant patients (29).

A study conducted in the United Kingdom that evaluated the relationship between the B.1.1.7 variant and case severity estimated that the risk of death associated with B.1.1.7 is 61% higher than that associated with pre-existing non-VOC lineages (9). In this study, the presence of the B.1.1.7 variant was confirmed by PCR based on S gene amplification failure instead of the complete genome. In our study, we analyzed whether the incidence of the P.1 variant would be related to different age groups or clinical condition strata based on a specific cohort composed of 86 samples from April 2021 that were completely sequenced. Our results did not reveal an association between the P.1 variant prevalence, age range and severity. The P.1 variant was dispersed in all groups with similar severity indices; however, we emphasize that in our April cohort, the P.1-like-II lineage was identified only in patients under the age of 60 years. A limitation of our study includes the impossibility of follow-up in the cases to make a more precise correlation between the variants and the clinical outcomes, and, for the association analysis, clinical severity was determined from the data from the laboratory that provided the samples. Therefore, we infer, based on the available data, that the P.1 variant may not be related to a higher severity of COVID-19, although further studies should be carried out to carefully explore this issue.

Our phylogenetic analysis performed with the P.1 variant genomes identified in the state of Paraná and in the other states of Brazil showed that the recently described lineage P.1-like-II (21) was also present at moderate frequencies in the state of Paraná (Figure 3 and 5). The P.1-like-II lineage presents 15 of the 22 mutations of the P.1 variant, including the three main mutations in the RBD domain of the S protein: K417T, E484K and N501Y. On the other hand, they present some unique substitutions: ORF1ab: C8905T, C16954T and A20931G; NSP4:D2980H, intergenic region E/M A26492T and N:P383 L. The analysis of the mutations in the first 11 P.1-like-II genomes from Paraná identified 3 of the 4 unique substitutions of the P.1-like-II lineage (C8905T, C16954T, A26492T) in addition to the N:P383 L and ORF1a:D2980H mutations. However, we also identified some unique substitutions, which were not yet described, for the P.1 variant and P.1-like-II lineage, and these substitutions included ORF1a:P1213 L and ORF1b:K23240N, which appeared in 45% of the genomes (Table S2). Our results, in agreement with the initial study (21, 29), show that the VOC P.1 and the P.1-like-II lineages diverged from a common ancestor that contained some, but not all, of the defining mutations found in the P.1 variant. Among the main mutations common between the two groups, we highlight those that occur in the S protein: S:E484K, S:N501Y and S:D614G. This hypothesis also justifies the absence of other mutations in the S protein, such as S:T20N, in the P.1-like-II lineage. Our phylogenetic analysis over time showed that the P.1-like-II lineage emerged between 2 and 4 months later than the P.1 variant. Interestingly, unlike the VOC P.1, the P.1-like-II lineage genomes were not dispersed throughout the state of Paraná but were clustered in western Paraná (represented by the host cities, Toledo and Cascavel). Particularly in this region, we found the lowest VOC P.1 frequency of the state, while the P.1-like-II lineage genomes accounted for almost 30% of the samples sequenced in the cohort study in April 2021. Conversely, the phylogenetic analysis supports a local transmission of the P.1-like-II lineage in Paraná. To date, Santa Catarina is the Brazilian state where the largest number of the P.1-like-II lineage genomes has been identified: 30 in total, which corresponds to 71% of the genomes of that state that were previously classified as the P.1 variant by PANGO (Figure 5). Of this, 20 P.1-like-II representatives were identified in the city of Chapecó, which is also located in the western region, approximately 360 km from Toledo and Cascavel. For instance, in February 2021, Chapecó was responsible for the largest outbreak of COVID-19 in the state of Santa Catarina (16). According to our tree-scaled analysis (Figure 4), the emergence of the P.1-like-II lineages in the state of Paraná is estimated to occur at the same time as the appearance of the P.1-like-II lineages in Santa Catarina. On the other hand, the P.1-like-II lineages from the other states (São Paulo, Minas Gerais, Alagoas, Espírito Santo) appeared after the variants observed in Paraná and Santa Catarina. These data, together with genomic surveillance information, reinforce the hypothesis that the P.1-like-II lineage may have arisen in the western area of the state of Santa Catarina. It is possible that the P.1-like-II lineage may have emerged during the outbreak in the city of Chapecó and initially spread to the western region of Paraná, which could explain the localized expansion of the P.1-like-II lineage. This result suggests that, unlike the B.1.1.7 variant that failed to establish a prevalence in the state and despite having emerged after the VOC P.1, the constellation of mutations of the P.1-like-II lineage allowed it to increase in frequency and then cocirculate with the P.1 variant in some specific geographic regions.

During the phylogenetic analysis, we noticed that a group formed by 8 genomes from the state of Paraná stood out. Analysis of this group of genomes, classified as belonging to the P.1 variant, revealed the presence of an additional mutation in the S protein (S:E661D). A genomic surveillance study carried out in the United Kingdom identified the S:E661D mutation in viral genomes; however, they were at an extremely low frequency, because 6 genomes out of 142,859 were identified (10). In our study, the E661D mutation appeared at a much higher frequency and occurred in the P.1 variant, which already has a high number of mutations. We also looked for the presence of this mutation in other P.1 variant genomes from Brazil deposited in the GISAID. We verified the existence of 46 Brazilian P.1 variant genomes with the S:E661D mutation, 20 of which were identified in April 2021. However, our attention was drawn to the high frequency of this mutation in genomes in the state of Paraná, which occurred in 11.35% of the P.1 variant genomes. In comparison, the second state with the highest frequency of the S:E661D mutation was Minas Gerais, with 2.94%.

Notably, the S:E661D mutation was not detected throughout the state of Paraná but was observed specifically in northern Paraná, as well as in the capital Curitiba, which accounted for nearly 25% of the P.1 variant genomes. Conversely, the phylogenetic analysis revealed clusterization of the P.1 genomes harboring S:E661D mutation as a function of the state from which the samples were collected, indicating that this mutation had not occurred randomly, but this mutation happened systematically in some geographical regions (Figure 5).

Cheng and coworkers (8) used structure-based computational models to demonstrate that the SARS-CoV-2 S protein exhibits a high affinity motif for binding to T cell receptors (8). This predicted motif extends from the amino acid residues E661 to R685 of the S protein. Despite the amino acid change, the E661D mutation maintained the acidic character of the motif, suggesting that this feature may be important for the function of the protein. Additionally, the comparison of the predicted superantigenic region of the SARS-CoV-2 S protein with the same region within bacterial superantigenic proteins showed that there is conservation of aspartic acid residues in bacterial proteins. Although there is no evidence regarding this additional mutation on any aspect of the biology of the virus, it is noteworthy that mutations in the S protein are frequently associated with altered viral fitness and/or immune evasion. For example, the D614G substitution enhanced viral stability and infectivity (33), and the E484 mutation in the receptor-binding domain (RBD) conferred a partial resistance of SARS-CoV-2 to neutralizing antibodies (40). Our results reinforce the importance of genomic vigilance to track possible novel VOIs that may arise from the current scenario of low vaccination rates and uncontrolled dissemination of SARS-CoV-2 in Brazil, as previously documented (27).

The classification of genomes into variants performed by Nexclade and Pangolin revealed the presence of an N.10 variant genome in the state of Paraná (N.10-PR). Variant N.10, identified in samples from the Brazilian state of Maranhão, is considered a VOI due to the set of mutations it carries, mainly in the S protein (26). However, unlike the N.10 variant originally described (N.10-MA), the N.10-PR variant has the S:W152C mutation in addition to the other mutations previously described. The S:W152C mutation was initially described in the VOI B.1.429 (CAL.20C), which was identified in the state of California, United States (Table S1). The importance of the S:W152C mutation is the fact that the NTD is a target for neutralizing antibodies (7). The VOI B.1.429, which also has a very important S:L452R mutation (40), was identified in California, and to date, there are no reports that it has arrived in Brazil (28, 41). The other mutations identified in the B.1.429 variant, with the exception of S:D614G, do not match the N.10-PR variant. These results suggest that the N.10-PR S:W152C mutation occurred independently in this variant and has no relation with the VOI B.1.429. The B.1.429 variant has been extensively studied for its infective capacity and binding to neutralizing antibodies. The infective capacities of mutations occurring in the B.1.429 variant were investigated using pseudoviruses carrying the D614G mutation together with the L452R or W152C mutations infected in 293T cells expressing the ACE2 receptor as well as the cofactor TMPRSS2 (15). Although the results show an increase in infectivity close to 20-fold for the D614G/L452R protein, the increase in infectivity for D614G/W152C was 4-fold when compared to the S protein containing only the D614G mutation (15). A neutralizing antibody activity study using convalescent serum from patients or serum from vaccinated individuals showed a 3-6-fold reduction against the B.1.429 variant, and furthermore, the neutralizing activity of mAbs targeting NTD was completely blocked due to the combined mutations S:S13I and S:W152C in that domain (28). Additional information from the GISAID (GISAID - 2021-06-01) showed that the S:W152C mutation emerged 41,272 times (2.38% of all samples with spike sequences) in 39 countries. The first strain with this change, collected in July 2020, was hCoV-19/Mexico/ROO-InDRE_243/2020. The most recent occurred in strain hCoV-19/USA/KS-KHEL-1975/2021, collected in May 2021, but none of them were identified in Brazil according to the GISAID data. The concern with this N.10-PR variant is the emergence of the S:W152C mutation, and several studies have already showed its relevance to the biology of SARS-CoV-2 due to its S protein that has already accumulated important mutations such as S:V445A, S:E484k and S:D614G. At this point, the N.10-PR variant has as many mutations in the S protein as the VOCs B.1.1.7 and B.1.645, which are seven in total.

## Conclusions

A genomic survey from March 2020 onwards in the state of Paraná state, which is based on 333 SARS-CoV-2 genomes, allowed us to map the entire dynamics of detection, distribution and replacement of variants in the state of Paraná since the beginning of the pandemic. This work represents the most complete SARS-CoV-2 genomic surveillance study carried out in the state of Paraná. A phylogenetic analysis revealed that a group of genomes branched into a monophyletic clade, identified as the P.1-like-II lineage, which probably emerged between February and March 2021. The P.1-like-II lineage was proportionally more frequently detected in the state of Santa Catarina, which borders the state of Paraná. Interestingly, unlike other lineages, the prevalence of the P.1-like-II variant in Paraná was not negatively affected by the advancement of the P.1 variant during the period analyzed. We also detected the occurrence of an additional mutation S:(E661D) in the P.1 variant genomes. Finally, in our work, we did not detect a relationship between the P.1 variant, age group and case severity. However, cases associated with the P.1 variant with the E661D mutation were more likely to develop a severe form of the disease.

## Data Availability

Supplementary figures
Figure S1: SARS-CoV-2 variants identified at Paraná state in April 2021.
Figure S2: Distribution of SARS-CoV-2 variants identified in April 2021, according to macro-regions of Paraná state.
Figure S3: SARS-CoV-2 variants identified at Paraná state in April 2021, by age group.
Figure S4: SARS-CoV-2 variants identified at Paraná state in April 2021, by clinical severity at the moment of sample collection (LAC: mild cases; LACEN: severe cases).
Supplementary list
List S1: GISAID accession numbers of 245 news genomes from Paraná produced by the Fiocruz COVID-19 Genomic Surveillance Network.
Supplementary tables
Table S1: Non-synonymous mutations described for VOIs N.10-PR, N.10-MA and B.1.429.
Table S2: Non-synonymous mutations described for P.1-like-II.
Supplementary appendix
Appendix S1: GISAID acknowledgment.

https://github.com/mauromedeirosoliveira/Fiocruz-ICC

## Supplementary Material 1: here

### A. Supplementary figures

Figure S1: SARS-CoV-2 variants identified in Paraná state in April 2021. Figure S2: Distribution of SARS-CoV-2 variants identified in April 2021, according to macroregions of Paraná state. Figure S3: SARS-CoV-2 variants identified in Paraná state in April 2021 by age group. Figure S4: SARS-CoV-2 variants identified at Paraná state in April 2021 by clinical severity at the moment of sample collection (LAC: mild cases; LACEN: severe cases).

### B. Supplementary list

List S1: Number and source of SARS-CoV-2 genomes used in this study.

### C. Supplementary tables

Table S1: Nonsynonymous mutations described for VOIs N.10-PR, N.10-MA and B.1.429. Table S2: Nonsynonymous mutations described for P.1-like-II.

## Supplementary Material 2: here

### A. Supplementary appendix

Appendix S1: GISAID acknowledgment file.

## Availability of Data

The data are publicly available at NCBI with Gen-Bank accession numbers: MZ477744 - MZ477859. The data are available at GISAID with accession numbers: EPI_ISL_2775450 - EPI_ISL_2775468 and EPI_ISL_2759020-EPI_ISL_2758965.

## Acknowledgements

We would like to thank the Fiocruz COVID-19 Genomic Surveillance Network and Instituto Carlos Chagas for their support; the Laboratório Central do Estado (LACEN-PR), the Instituto de Biologia Molecular do Paraná (IBMP), Secretaria da Saúde of the Paraná state (Sesa-PR) and the Laboratório de Apoio ao Diagnóstico da Covid-19 (LAC) for providing the samples and metadata used in that study. We thank the Universidade Federal do Paraná (UFPR) for the use of the Illumina MiSEq platform. FP and BD LRA are research fellow awardees from Conselho Nacional de Pesquisa (CNPQ). MM and MO are postdoctoral fellows from CAPES (CAPES Finance Code 001). Finally, we would like to thank the digital designer Wagner Nagib from Instituto Carlos Chagas for helping us with the design of the figures. We would also like to thank all Brazilian researchers who have dedicated their time and efforts to carry out genomic surveillance of SARS-CoV-2 in Brazil. A GISAID acknowledgment table containing sequences used in this study is shown in Supplementary Appendix S1.

