## Supplementary Material 1 for "Genomic surveillance of SARS-CoV-2 in the state of Paraná, Southern Brazil, reveals the cocirculation of the VOC P.1, P.1-like-II lineage and a P.1 cluster harboring the S:E661D mutation"

**Figure S1:** SARS-CoV-2 variants identified in Paraná state in April 2021.

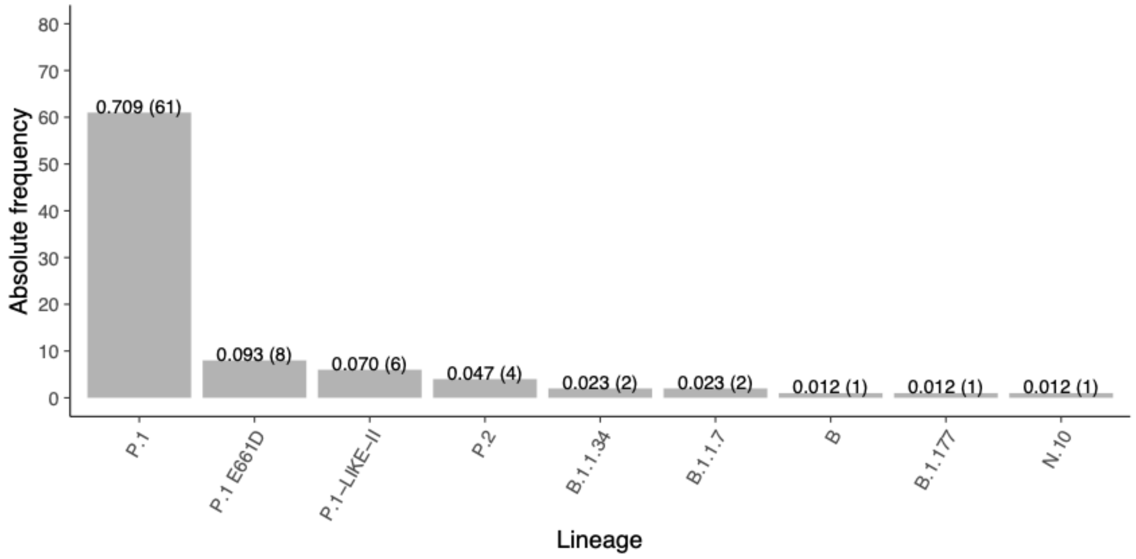


**Figure S2:** Distribution of SARS-CoV-2 variants identified in April 2021, according to macroregions of Paraná state.

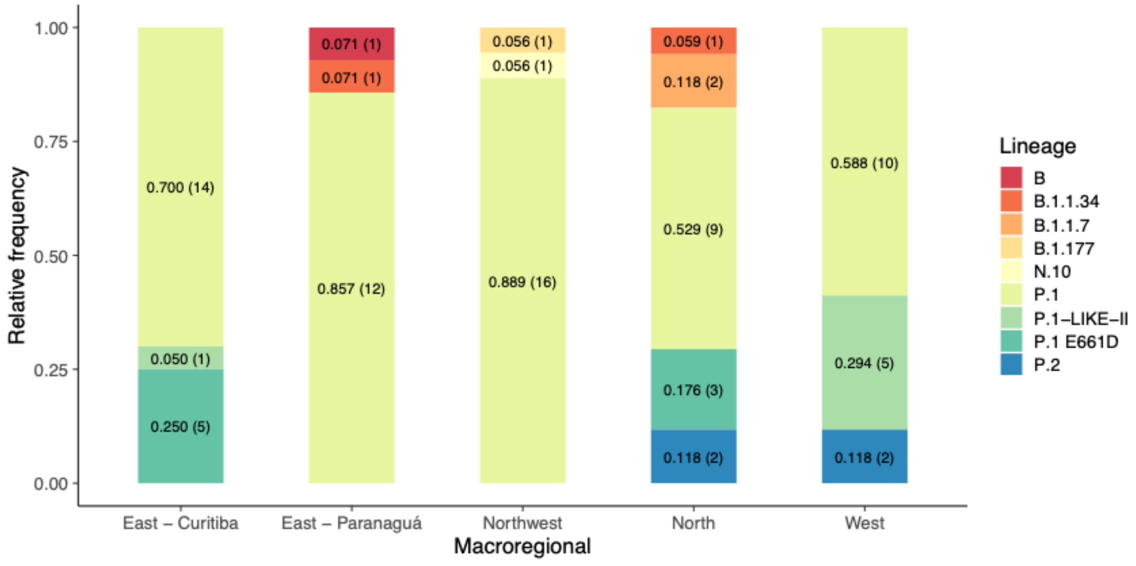

**Figure S3:** SARS-CoV-2 variants identified in Paraná state in April 2021 by age group.

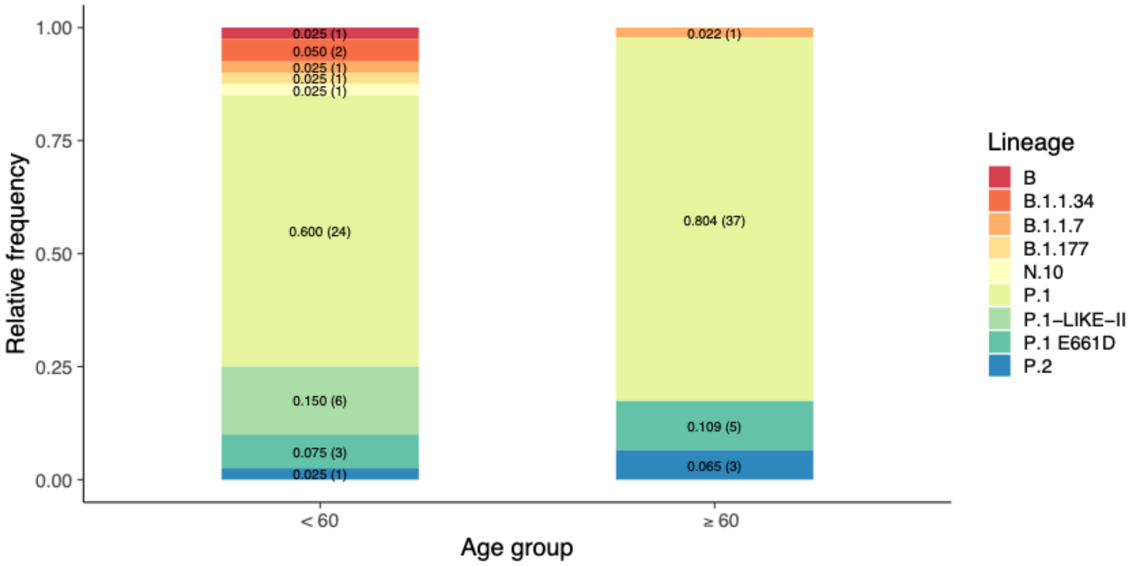


**Figure S4:** SARS-CoV-2 variants identified at Paraná state in April 2021 by clinical severity at the moment of sample collection (LAC: mild cases; LACEN: severe cases).

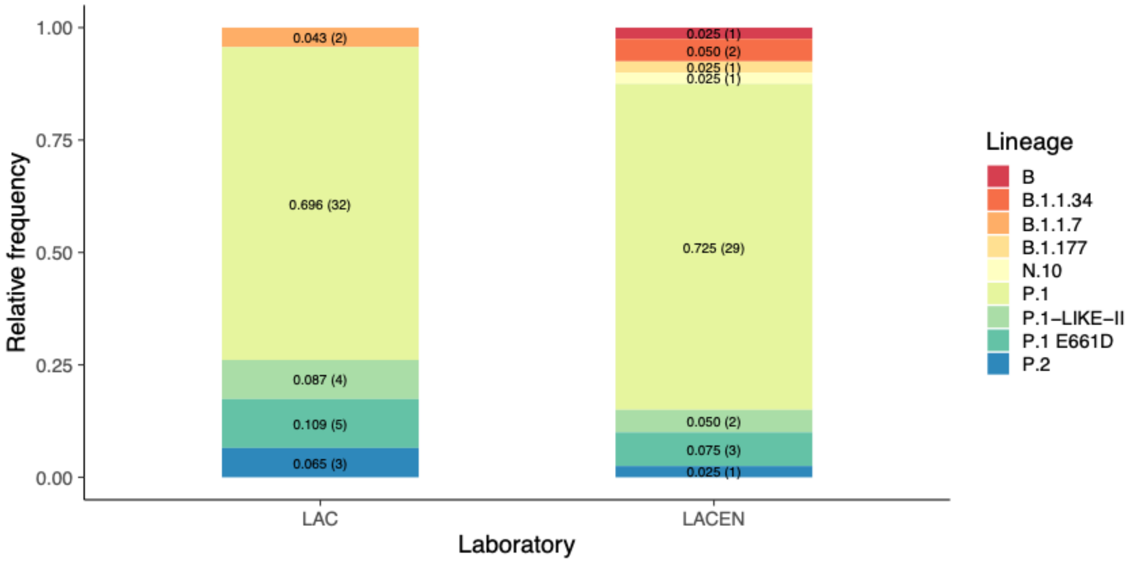


**List S1:** Number and source of SARS-CoV-2 genomes used in this study.

EPI_ISL_2775468, EPI_ISL_2775467, EPI_ISL_2775469, EPI_ISL_2775464, EPI_ISL_2775463, EPI_ISL_2775466, EPI_ISL_2775465, EPI_ISL_2775460, EPI_ISL_2775462, EPI_ISL_2775461, EPI_ISL_2775479, EPI_ISL_2775478, EPI_ISL_2775475, EPI_ISL_2775474, EPI_ISL_2775477, EPI_ISL_2775476, EPI_ISL_2775471, EPI_ISL_2775470, EPI_ISL_2775473, EPI_ISL_2775472, EPI_ISL_2775402, EPI_ISL_2775401, EPI_ISL_2775489, EPI_ISL_2775404, EPI_ISL_2775403, EPI_ISL_2775486, EPI_ISL_2775485, EPI_ISL_2775400, EPI_ISL_2775488, EPI_ISL_2775487, EPI_ISL_2775409, EPI_ISL_2775406, EPI_ISL_2775405, EPI_ISL_2775408, EPI_ISL_2775407, EPI_ISL_2775482, EPI_ISL_2775481, EPI_ISL_2775484, EPI_ISL_2775483, EPI_ISL_2775480, EPI_ISL_2775413, EPI_ISL_2775412, EPI_ISL_2775415, EPI_ISL_2775414, EPI_ISL_2775497, EPI_ISL_2775496, EPI_ISL_2775411, EPI_ISL_2775499, EPI_ISL_2775410, EPI_ISL_2775498, EPI_ISL_2775417, EPI_ISL_2775416, EPI_ISL_2775419, EPI_ISL_2775418, EPI_ISL_2775493, EPI_ISL_2775492, EPI_ISL_2775495, EPI_ISL_2775494, EPI_ISL_2775491, EPI_ISL_2775490, EPI_ISL_2775424, EPI_ISL_2775423, EPI_ISL_2775426, EPI_ISL_2775425, EPI_ISL_2775420, EPI_ISL_2775422, EPI_ISL_2775421, EPI_ISL_2775428, EPI_ISL_2775427, EPI_ISL_2775429, EPI_ISL_2775435, EPI_ISL_2775434, EPI_ISL_2775437, EPI_ISL_2775436, EPI_ISL_2775431, EPI_ISL_2775398, EPI_ISL_2775430, EPI_ISL_2775397, EPI_ISL_2775433, EPI_ISL_2775432, EPI_ISL_2775399, EPI_ISL_2775439, EPI_ISL_2775438, EPI_ISL_2775394, EPI_ISL_2775393, EPI_ISL_2775396, EPI_ISL_2775395, EPI_ISL_2775390, EPI_ISL_2775392, EPI_ISL_2775391, EPI_ISL_2775446, EPI_ISL_2775445, EPI_ISL_2775448, EPI_ISL_2775447, EPI_ISL_2775442, EPI_ISL_2775441, EPI_ISL_2775444, EPI_ISL_2775443, EPI_ISL_2775449, EPI_ISL_2775440, EPI_ISL_2775457, EPI_ISL_2775456, EPI_ISL_2775459, EPI_ISL_2775458, EPI_ISL_2775453, EPI_ISL_2775452, EPI_ISL_2775455, EPI_ISL_2775454, EPI_ISL_2775451, EPI_ISL_2775450, EPI_ISL_2758965, EPI_ISL_2758966, EPI_ISL_2758963, EPI_ISL_2759018, EPI_ISL_2758964, EPI_ISL_2759019, EPI_ISL_2758969, EPI_ISL_2758967, EPI_ISL_2758968, EPI_ISL_2759012, EPI_ISL_2759013, EPI_ISL_2759010, EPI_ISL_2759011, EPI_ISL_2758961, EPI_ISL_2759016, EPI_ISL_2758962, EPI_ISL_2759017, EPI_ISL_2759014, EPI_ISL_2758960, EPI_ISL_2759015, EPI_ISL_2758954, EPI_ISL_2759009,

EPI_ISL_2758955, EPI_ISL_2758952, EPI_ISL_2759007, EPI_ISL_2758953, EPI_ISL_2759008, EPI_ISL_2758958, EPI_ISL_2758959, EPI_ISL_2758956, EPI_ISL_2758957, EPI_ISL_2759001, EPI_ISL_2759002, EPI_ISL_2759000, EPI_ISL_2758950, EPI_ISL_2759005, EPI_ISL_2758951, EPI_ISL_2759006, EPI_ISL_2759003, EPI_ISL_2759004, EPI_ISL_2758949, EPI_ISL_2758943, EPI_ISL_2758944, EPI_ISL_2758941, EPI_ISL_2758942, EPI_ISL_2758947, EPI_ISL_2758948, EPI_ISL_2758945, EPI_ISL_2758946, EPI_ISL_2758940, EPI_ISL_2759070, EPI_ISL_2759071, EPI_ISL_2758938, EPI_ISL_2758939, EPI_ISL_2759067, EPI_ISL_2759068, EPI_ISL_2759065, EPI_ISL_2759066, EPI_ISL_2759069, EPI_ISL_2759060, EPI_ISL_2759063, EPI_ISL_2759064, EPI_ISL_2759061, EPI_ISL_2759062, EPI_ISL_2759056, EPI_ISL_2759057, EPI_ISL_2759054, EPI_ISL_2759055, EPI_ISL_2759058, EPI_ISL_2759059, EPI_ISL_2759052, EPI_ISL_2759053, EPI_ISL_2759050, EPI_ISL_2759051, EPI_ISL_2758998, EPI_ISL_2758999, EPI_ISL_2758996, EPI_ISL_2758997, EPI_ISL_2758990, EPI_ISL_2759045, EPI_ISL_2758991, EPI_ISL_2759046, EPI_ISL_2759043, EPI_ISL_2759044, EPI_ISL_2758994, EPI_ISL_2759049, EPI_ISL_2758995, EPI_ISL_2758992, EPI_ISL_2759047, EPI_ISL_2758993, EPI_ISL_2759048, EPI_ISL_2759041, EPI_ISL_2759042, EPI_ISL_2759040, EPI_ISL_2758987, EPI_ISL_2758988, EPI_ISL_2758985, EPI_ISL_2758986, EPI_ISL_2758989, EPI_ISL_2759034, EPI_ISL_2758980, EPI_ISL_2759035, EPI_ISL_2759032, EPI_ISL_2759033, EPI_ISL_2758983, EPI_ISL_2759038, EPI_ISL_2758984, EPI_ISL_2759039, EPI_ISL_2758981, EPI_ISL_2759036, EPI_ISL_2758982, EPI_ISL_2759037, EPI_ISL_2759030, EPI_ISL_2759031, EPI_ISL_2758976, EPI_ISL_2758977, EPI_ISL_2758974, EPI_ISL_2759029, EPI_ISL_2758975, EPI_ISL_2758978, EPI_ISL_2758979, EPI_ISL_2759023, EPI_ISL_2759024, EPI_ISL_2759021, EPI_ISL_2759022, EPI_ISL_2758972, EPI_ISL_2759027, EPI_ISL_2758973, EPI_ISL_2759028, EPI_ISL_2758970, EPI_ISL_2759025, EPI_ISL_2758971, EPI_ISL_2759026, EPI_ISL_2759020.

**Table S1:** Nonsynonymous mutations described for VOIs N.10-PR, N.10-MA and B.1.429.

| | VOI | | |
|---|---|---|---|
| Protein | N.10-PR | N.10-MA | B.1.429 (CAL.20C) |
| S | S:P9L | S:P9L | - |
| | - | - | S:S13I |
| | S:W152C | - | S:W152C |
| | S:I210V | S:I210V | - |
| | S:N211I | S:N211I | - |
| | S:V445A | S:V445A | - |
| | - | - | S:L452R |
| | S:E484K | S:E484K | - |
| | S:D614G | S:D614G | S:D614G |
| ORF1a | ORF1a:H712Y | - | - |
| | ORF1a:P1640L | ORF1a:P1640L | - |
| | ORF1a:P3371S | ORF1a:P3371S | - |
| | ORF1a:P3395S | ORF1a:P3395S | - |
| | ORF1a:V3718A | ORF1a:V3718A | - |
| - | - | - | ORF1a:I4205V |
| N | N:Q70R | N:Q70R | - |
| | N:R203K | N:R203K | - |
| | N:G204R | N:G204R | - |
| | N:I292T | N:I292T | - |
| ORF1b | ORF1b:P314L | ORF1b:P314L | - |
| ORF6 | ORF6:I33T | ORF6:I33T | - |
| ORF7b | ORF7b:F13X | ORF7b:F13X | - |
| ORF9b | ORF9b:K67E | ORF9b:K67E | - |

**Table S2:** Nonsynonymous mutations described for P.1-like-II.

| | P.1-like-II | | | | | | | | | | | | |
|---|---|---|---|---|---|---|---|---|---|---|---|---|---|
| Protein | RS | SC | PR-A11-PL1 | PR-F08-PL2 | PR-C04-PL1 | PR-C10-PL1 | PR-C12-PL1 | PR-A05-PL2 | BR-B08-PL2 | PR-B10-PL2 | PR-B11-PL2 | PR-F08-PL2 | PR-F11_PL2 |
| N | N:P80R | N:P80R | N:P80R | N:P80R | N:P80R | N:P80R | N:P80R | N:P80R | N:P80R | N:P80R | N:P80R | N:P80R | N:P80R |
| | N:R203K | N:R203K | N:R203K | N:R203K | N:R203K | N:R203K | N:R203K | N:R203K | N:R203K | N:R203K | N:R203K | N:R203K | N:R203K |
| | N:G204R | N:G204R | N:G204R | N:G204R | N:G204R | N:G204R | N:G204R | N:G204R | N:G204R | N:G204R | N:G204R | N:G204R | N:G204R |
| | | N:P383L | N:P383L | N:P383L | N:P383L | N:P383L | N:P383L | N:P383L | N:P383L | N:P383L | N:P383L | N:P383L | N:P383L |
| | | | | | | | N:T271I | | | | | | |
| ORF1a | | | | | | | | ORF1a:S40L | | | | | |
| | | | | | | | | ORF1a:F548S | | | | | |
| | | | | | | | | | ORF1a:S17I | | | | |
| | ORF1a:S1188L | ORF1a:S1188L | ORF1a:S1188L | ORF1a:S1188L | ORF1a:S1188L | ORF1a:S1188L | ORF1a:S1188L | ORF1a:S1188L | ORF1a:S1188L | ORF1a:S1188L | ORF1a:S1188L | ORF1a:S1188L | ORF1a:S1188L |
| | | | | | ORF1a:P1213L | ORF1a:P1213L | ORF1a:P1213L | | | ORF1a:P1213L | | | ORF1a:P1213L |
| | ORF1a:K1795Q | ORF1a:K1795Q | ORF1a:K1795Q | ORF1a:K1795Q | ORF1a:K1795Q | ORF1a:K1795Q | ORF1a:K1795Q | ORF1a:K1795Q | ORF1a:K1795Q | ORF1a:K1795Q | ORF1a:K1795Q | ORF1a:K1795Q | ORF1a:K1795Q |
| | | | | | | | | ORF1a:M2347I | | | | | |
| | ORF1a:D2980H | ORF1a:D2980H | ORF1a:D2980H | ORF1a:D2980H | ORF1a:D2980H | ORF1a:D2980H | ORF1a:D2980H | ORF1a:D2980H | ORF1a:D2980H | ORF1a:D2980H | ORF1a:D2980H | ORF1a:D2980H | ORF1a:D2980H |
| | | | | | ORF1a:I2729V | | | | | | | | |
| | | | | | | | ORF1a:N2405S | | | | | | |
| | | | | | | | | ORF1a:K3353R | | | | | |
| ORF1b | ORF1b:P314L | ORF1b:P314L | ORF1b:P314L | ORF1b:P314L | ORF1b:P314L | ORF1b:P314L | ORF1b:P314L | ORF1b:P314L | ORF1b:P314L | ORF1b:P314L | ORF1b:P314L | ORF1b:P314L | ORF1b:P314L |
| | ORF1b:S1187N | | | | | | | | | | | | |
| | | | | | | | | ORF1b:K2557R | | | | | |
| | | | | | | | | | ORF1b:T730I | | | | |
| | | | | | | | ORF1b:K1873N | | | | | | |
| | | | | | | | | | | ORF1b:I2568V | | | |
| | | | | | ORF1b:K2340N | ORF1b:K2340N | ORF1b:K2340N | | | ORF1b:K2340N | | | ORF1b:K2340N |
| ORF3a | ORF3a:S253P | ORF3a:S253P | ORF3a:S253P | ORF3a:S253P | ORF3a:S253P | ORF3a:S253P | ORF3a:S253P | ORF3a:S253P | ORF3a:S253P | ORF3a:S253P | ORF3a:S253P | ORF3a:S253P | |
| ORF6 | | | | | | | | | | | ORF6:L16X | | |
| | | | | | ORF6:W27L | | | | | | | | |
| ORF9b | ORF9b:Q77E | ORF9b:Q77E | ORF9b:Q77E | ORF9b:Q77E | ORF9b:Q77E | ORF9b:Q77E | ORF9b:Q77E | ORF9b:Q77E | ORF9b:Q77E | ORF9b:Q77E | ORF9b:Q77E | ORF9b:Q77E | ORF9b:Q77E |
| S | S:L18F | S:L18F | S:L18F | S:L18F | S:L18F | S:L18F | S:L18F | S:L18F | S:L18F | S:L18F | S:L18F | S:L18F | S:L18F |
| | S:P26S | S:P26S | S:P26S | S:P26S | S:P26S | S:P26S | S:P26S | S:P26S | S:P26S | S:P26S | S:P26S | S:P26S | S:P26S |
| | S:D138Y | S:D138Y | S:D138Y | S:D138Y | S:D138Y | S:D138Y | S:D138Y | S:D138Y | S:D138Y | S:D138Y | S:D138Y | S:D138Y | S:D138Y |
| | S:R190S | S:R190S | S:R190S | S:R190S | S:R190S | S:R190S | S:R190S | S:R190S | S:R190S | S:R190S | S:R190S | S:R190S | S:R190S |
| | S:K417T | S:K417T | S:K417T | S:K417T | S:K417T | S:K417T | S:K417T | S:K417T | S:K417T | S:K417T | S:K417T | S:K417T | S:K417T |
| | S:E484K | S:E484K | S:E484K | S:E484K | S:E484K | | S:E484K | S:E484K | S:E484K | S:E484K | S:E484K | S:E484K | S:E484K |
| | S:N501Y | S:N501Y | S:N501Y | S:N501Y | S:N501Y | | S:N501Y | S:N501Y | S:N501Y | S:N501Y | S:N501Y | S:N501Y | S:N501Y |
| | | | | | | | | | | S:T547I | | | |
| | S:D614G | S:D614G | S:D614G | S:D614G | S:D614G | S:D614G | S:D614G | S:D614G | S:D614G | S:D614G | S:D614G | S:D614G | S:D614G |
| | S:H655Y | S:H655Y | S:H655Y | S:H655Y | S:H655Y | | S:H655Y | S:H655Y | S:H655Y | S:H655Y | S:H655Y | S:H655Y | S:H655Y |
| | | | S:Q675H | | | | | | | | | | |
| | S:T1027I | | S:T1027I | S:T1027I | S:T1027I | | S:T1027I | S:T1027I | S:T1027I | S:T1027I | S:T1027I | S:T1027I | S:T1027I |
| | S:V1176F | S:V1176F | S:V1176F | S:V1176F | S:V1176F | S:V1176F | S:V1176F | S:V1176F | S:V1176F | | S:V1176F | S:V1176F | S:V1176F |
