## Supplementary Material 2 for "Genomic surveillance of SARS-CoV-2 in the state of Paraná, Southern Brazil, reveals the cocirculation of the VOC P.1, P.1-like-II lineage and a P.1 cluster harboring the S:E661D mutation"

We gratefully acknowledge the following Authors from the Originating laboratories responsible for obtaining the specimens, as well as the Submitting laboratories where the genome data were generated and shared via GISAID, on which this research is based.

All Submitters of data may be contacted directly via www.gisaid.org

Authors are sorted alphabetically.

| Accession ID | Originating Laboratory | Submitting Laboratory | Authors |
| --- | --- | --- | --- |
| EPI_ISL_1000671, EPI_ISL_1000673 | Instituto de Biotecnologia - UNESP-Botucatu-SP | Instituto de Biotecnologia - UNESP-Botucatu-SP | Leila Sabrina Ullmann; Fábio Sossai Possebon, Camila Dantas Malossi, Paula Rahal, Paulo Inacio da Costa, João Pessoa Araújo Jr. |
| EPI_ISL_1004233, EPI_ISL_1004234, EPI_ISL_1004235, EPI_ISL_1004238, EPI_ISL_1004239, EPI_ISL_1004240, EPI_ISL_1004241, EPI_ISL_1004242 | Laboratório de Virologia - Instituto de Medicina Tropical - Universidade de São Paulo | Laboratório de Parasitologia Médica - Instituto de Medicina Tropical - Universidade de São Paulo | Camila Malta Romano, Jaqueline Goes de Jesus, Giulia Magalhães Ferreira, Pamela dos Santos Andrade, Esmenia Coelho, Alvina Clara Felix, Anderson de Paula, Darlan Candido, Ingra Morales Claro, Franciane Mendes, Midiã Ferreira, Lucas A. Moyses Franco, Flavia Cristina Sales, Nuno Faria, Ester C. Sabino; Brazil-UK Centre for Arbovirus Discovery Diagnosis Genomics and Epidemiology (CADDE) Genomic Network - Instituto de Medicina Tropical |
| EPI_ISL_1039691, EPI_ISL_1039692, EPI_ISL_1039693, EPI_ISL_1039694, EPI_ISL_1039695, EPI_ISL_1041509 | LACEN do Estado de Goias | Instituto Adolfo Lutz, Interdiciplinary Procedures Center, Strategic Laboratory | Claudio Tavares Sacchi, Claudia Regina Gonçalves, Erica Valessa Ramos Gomes, Karoline Rodrigues Campos |
| EPI_ISL_1060877, EPI_ISL_1060879, EPI_ISL_1060880, EPI_ISL_1060881, EPI_ISL_1060888, EPI_ISL_1060889, EPI_ISL_1060890, EPI_ISL_1060892, EPI_ISL_1060894, EPI_ISL_1060895, EPI_ISL_1060896, EPI_ISL_1060897, EPI_ISL_1060898, EPI_ISL_1060899, EPI_ISL_1060906, EPI_ISL_1060911, EPI_ISL_1060912, EPI_ISL_1060913, EPI_ISL_1060915, EPI_ISL_1064736, EPI_ISL_1064737 | | | |
| see above | CDL Laboratorio Santos e Vidal LTDA. | Instituto de Medicina Tropical de Sao Paulo | Brazil-UK Centre for Arbovirus Discovery Diagnosis Genomics and Epidemiology (CADDE) Genomic Network - Instituto de Medicina Tropical |
| EPI_ISL_1067728 | Center for Biotechnology and Cell Therapy, São Rafael Hospital, Salvador, Brazil | Central Public Health Laboratory - LACEN -Bahia, Salvador, Brazil | Stephane Tosta, Luciana Oliveira, Vanessa Nardy,Patrícia Cajado,Marcela Gómez, Breno Dominguez, Jaqueline Gomes, Vagner Fonseca,Marta Giovanetti,Luiz Alcantara, Felicidade Pereira, Arabela Leal |
| EPI_ISL_1067729, EPI_ISL_1067730, EPI_ISL_1067731 | Central Public Health Laboratory - LACEN -Bahia, Salvador, Brazil | Central Public Health Laboratory - LACEN -Bahia, Salvador, Brazil | Stephane Tosta, Luciana Oliveira, Vanessa Nardy,Patrícia Cajado,Marcela Gómez, Breno Dominguez, Jaqueline Gomes, Vagner Fonseca,Marta Giovanetti,Luiz Alcantara, Felicidade Pereira, Arabela Leal |
| EPI_ISL_1067732 | Center for Biotechnology and Cell Therapy, São Rafael Hospital, Salvador, Brazil | Central Public Health Laboratory - LACEN -Bahia, Salvador, Brazil | Stephane Tosta, Luciana Oliveira, Vanessa Nardy,Patrícia Cajado,Marcela Gómez, Breno Dominguez, Jaqueline Gomes, Vagner Fonseca,Marta Giovanetti,Luiz Alcantara, Felicidade Pereira, Arabela Leal |
| EPI_ISL_1067733, EPI_ISL_1067734, EPI_ISL_1067735 | Central Public Health Laboratory - LACEN -Bahia, Salvador, Brazil | Central Public Health Laboratory - LACEN -Bahia, Salvador, Brazil | Stephane Tosta, Luciana Oliveira, Vanessa Nardy,Patrícia Cajado,Marcela Gómez, Breno Dominguez, Jaqueline Gomes, Vagner Fonseca,Marta Giovanetti,Luiz Alcantara, Felicidade Pereira, Arabela Leal |
| EPI_ISL_1067736 | Center for Biotechnology and Cell Therapy, São Rafael Hospital, Salvador, Brazil | Central Public Health Laboratory - LACEN -Bahia, Salvador, Brazil | Stephane Tosta, Luciana Oliveira, Vanessa Nardy,Patrícia Cajado,Marcela Gómez, Breno Dominguez, Jaqueline Gomes, Vagner Fonseca,Marta Giovanetti,Luiz Alcantara, Felicidade Pereira, Arabela Leal |
| EPI_ISL_1067737, EPI_ISL_1067738 | Central Public Health Laboratory - LACEN -Bahia, Salvador, Brazil | Central Public Health Laboratory - LACEN -Bahia, Salvador, Brazil | Stephane Tosta, Luciana Oliveira, Vanessa Nardy,Patrícia Cajado,Marcela Gómez, Breno Dominguez, Jaqueline Gomes, Vagner Fonseca,Marta Giovanetti,Luiz Alcantara, Felicidade Pereira, Arabela Leal |
| EPI_ISL_1078992, EPI_ISL_1079002, EPI_ISL_1079008, EPI_ISL_1079162 | IAL Regional de Bauru | Instituto Adolfo Lutz, Interdiciplinary Procedures Center, Strategic Laboratory | Claudio Tavares Sacchi, Claudia Regina Gonçalves, Erica Valessa Ramos Gomes, Karoline Rodrigues Campos |
| EPI_ISL_1086035, EPI_ISL_1086036 | IAL Regional de Bauru | Instituto Adolfo Lutz, Interdiciplinary Procedures Center, Strategic Laboratory | Claudio Tavares Sacchi, Claudia Regina Gonçalves, Erica Valessa Ramos Gomes, Karoline Rodrigues Campos, Caio Vinicius Dias Lopes |
| EPI_ISL_1086037, EPI_ISL_1086038, EPI_ISL_1086039, EPI_ISL_1086040, EPI_ISL_1086041, EPI_ISL_1086043 | Diagnosticos da America - DASA | Instituto Adolfo Lutz, Interdiciplinary Procedures Center, Strategic Laboratory | Claudio Tavares Sacchi, Claudia Regina Gonçalves, Erica Valessa Ramos Gomes, Karoline Rodrigues Campos, Caio Vinicius Dias Lopes |
| EPI_ISL_1086044, EPI_ISL_1086045, EPI_ISL_1086046, EPI_ISL_1086047, EPI_ISL_1096120 | IAL Regional de Bauru | Instituto Adolfo Lutz, Interdiciplinary Procedures Center, Strategic Laboratory | Claudio Tavares Sacchi, Claudia Regina Gonçalves, Erica Valessa Ramos Gomes, Karoline Rodrigues Campos, Caio Vinicius Dias Lopes |
| EPI_ISL_1096121 | Diagnosticos da America - DASA | Instituto Adolfo Lutz, Interdiciplinary Procedures Center, Strategic Laboratory | Claudio Tavares Sacchi, Claudia Regina Gonçalves, Erica Valessa Ramos Gomes, Karoline Rodrigues Campos, Caio Vinicius Dias Lopes |
| EPI_ISL_1096122, EPI_ISL_1096124, EPI_ISL_1096134 | IAL Regional de Bauru | Instituto Adolfo Lutz, Interdiciplinary Procedures Center, Strategic Laboratory | Claudio Tavares Sacchi, Claudia Regina Gonçalves, Erica Valessa Ramos Gomes, Karoline Rodrigues Campos, Caio Vinicius Dias Lopes |
| EPI_ISL_1096135 | Diagnosticos da America - DASA | Instituto Adolfo Lutz, Interdiciplinary Procedures Center, Strategic Laboratory | Claudio Tavares Sacchi, Claudia Regina Gonçalves, Erica Valessa Ramos Gomes, Karoline Rodrigues Campos, Caio Vinicius Dias Lopes |
| EPI_ISL_1121307 | Hospital E Antonio Policarpo de Oliveira | Instituto Adolfo Lutz, Interdiciplinary Procedures Center, Strategic Laboratory | Claudio Tavares Sacchi, Claudia Regina Gonçalves, Erica Valessa Ramos Gomes, Karoline Rodrigues Campos, Caio Vinicius Dias Lopes |
| EPI_ISL_1121308 | Hospital e Pronto Socorro Portinari | Instituto Adolfo Lutz, Interdiciplinary Procedures Center, Strategic Laboratory | Claudio Tavares Sacchi, Claudia Regina Gonçalves, Erica Valessa Ramos Gomes, Karoline Rodrigues Campos, Caio Vinicius Dias Lopes |
| EPI_ISL_1121309 | UBS Jose Francisco Rezende | Instituto Adolfo Lutz, Interdiciplinary Procedures Center, Strategic Laboratory | Claudio Tavares Sacchi, Claudia Regina Gonçalves, Erica Valessa Ramos Gomes, Karoline Rodrigues Campos, Caio Vinicius Dias Lopes |
| EPI_ISL_1121310 | IAL Regional de Bauru | Instituto Adolfo Lutz, Interdiciplinary Procedures Center, Strategic Laboratory | Claudio Tavares Sacchi, Claudia Regina Gonçalves, Erica Valessa Ramos Gomes, Karoline Rodrigues Campos, Caio Vinicius Dias Lopes |
| EPI_ISL_1121311 | Centro de Saude II Dr. Jose Paione Mococa | Instituto Adolfo Lutz, Interdiciplinary Procedures Center, Strategic Laboratory | Claudio Tavares Sacchi, Claudia Regina Gonçalves, Erica Valessa Ramos Gomes, Karoline Rodrigues Campos, Caio Vinicius Dias Lopes |
| EPI_ISL_1121312, EPI_ISL_1121313, EPI_ISL_1121314, EPI_ISL_1121315 | IAL Regional de Bauru | Instituto Adolfo Lutz, Interdiciplinary Procedures Center, Strategic Laboratory | Claudio Tavares Sacchi, Claudia Regina Gonçalves, Erica Valessa Ramos Gomes, Karoline Rodrigues Campos, Caio Vinicius Dias Lopes |
| EPI_ISL_1121316 | LACEN do Rio Grande do Sul | Instituto Adolfo Lutz, Interdiciplinary Procedures Center, Strategic Laboratory | Claudio Tavares Sacchi, Claudia Regina Gonçalves, Erica Valessa Ramos Gomes, Karoline Rodrigues Campos, Caio Vinicius Dias Lopes |
| EPI_ISL_1123373 | Grupo Tecnico de Vigilancia Sanitaria e Epidemiologica | Instituto Adolfo Lutz, Interdiciplinary Procedures Center, Strategic Laboratory | Claudio Tavares Sacchi, Claudia Regina Gonçalves, Erica Valessa Ramos Gomes, Karoline Rodrigues Campos, Caio Vinicius Dias Lopes |
| EPI_ISL_1133120, EPI_ISL_1133121, EPI_ISL_1133122, EPI_ISL_1133123, EPI_ISL_1133124, EPI_ISL_1133125, EPI_ISL_1133126, EPI_ISL_1133127, EPI_ISL_1133128, EPI_ISL_1133129, EPI_ISL_1133130, EPI_ISL_1133131, EPI_ISL_1133132, EPI_ISL_1133134, EPI_ISL_1133135, EPI_ISL_1133136, EPI_ISL_1133137, EPI_ISL_1133138, EPI_ISL_1133139, EPI_ISL_1133140, EPI_ISL_1133141, EPI_ISL_1133142, EPI_ISL_1133143, EPI_ISL_1133144 | | | |
| see above | LABCOVID_HCPA | LABRESIS_HCPA | Martins AF, Wink PL, Volpato F, Rosset C, de Paris F, Monteiro F, Barth AL |
| EPI_ISL_1139070 | IAL Regional de Ribeirao Preto | Instituto Adolfo Lutz, Interdiciplinary Procedures Center, Strategic Laboratory | Claudio Tavares Sacchi, Claudia Regina Gonçalves, Erica Valessa Ramos Gomes, Karoline Rodrigues Campos, Caio Vinicius Dias Lopes |
| EPI_ISL_1164970, EPI_ISL_1164971 | LACEN - Laboratório Central de Saúde Pública do Ceará | Evandro Chagas Institute | Santos, M.C.; Silva, A.M.; Junior, W.D.C.; Barbagelata, L.S.; Ferreira, J.A.; Sousa, E.M.A.; da Silva, P.S.; Pinheiro, K.C.; L.C.; Sousa Junior, E.C. |

| | | | |
|---|---|---|---|
| EPI_ISL_1164972 | LACEN - Laboratório Central de Saúde Pública do Pará | Evandro Chagas Institute | Santos, M.C.; Silva, A.M.; Junior, W.D.C.; Barbagelata, L.S.; Ferreira, J.A.; Sousa, E.M.A.; da Silva, P.S.; Pinheiro, K.C.; L.C.; Sousa Junior, E.C. |
| EPI_ISL_1164973 | LACEN - Laboratório Central de Saúde Pública do Ceará | Evandro Chagas Institute | Santos, M.C.; Silva, A.M.; Junior, W.D.C.; Barbagelata, L.S.; Ferreira, J.A.; Sousa, E.M.A.; da Silva, P.S.; Pinheiro, K.C.; L.C.; Sousa Junior, E.C. |
| EPI_ISL_1164978 | LACEN - Laboratório Central de Saúde Pública do Pará | Evandro Chagas Institute | Santos, M.C.; Silva, A.M.; Junior, W.D.C.; Barbagelata, L.S.; Ferreira, J.A.; Sousa, E.M.A.; da Silva, P.S.; Pinheiro, K.C.; L.C.; Sousa Junior, E.C. |
| EPI_ISL_1164980 | LACEN - Laboratório Central de Saúde Pública do Ceará | Evandro Chagas Institute | Santos, M.C.; Silva, A.M.; Junior, W.D.C.; Barbagelata, L.S.; Ferreira, J.A.; Sousa, E.M.A.; da Silva, P.S.; Pinheiro, K.C.; L.C.; Sousa Junior, E.C. |
| EPI_ISL_1164984 | LACEN - Laboratório Central de Saúde Pública do Amapá | Evandro Chagas Institute | Santos, M.C.; Silva, A.M.; Junior, W.D.C.; Barbagelata, L.S.; Ferreira, J.A.; Sousa, E.M.A.; da Silva, P.S.; Pinheiro, K.C.; L.C.; Sousa Junior, E.C. |
| EPI_ISL_1164991 | LACEN - Laboratório Central de Saúde Pública do Paraiba | Evandro Chagas Institute | Santos, M.C.; Silva, A.M.; Junior, W.D.C.; Barbagelata, L.S.; Ferreira, J.A.; Sousa, E.M.A.; da Silva, P.S.; Pinheiro, K.C.; L.C.; Sousa Junior, E.C. |
| EPI_ISL_1166615 | LACEN - Laboratório Central de Saúde Pública do Rio Grande do Norte | Evandro Chagas Institute Virology | Santos, M.C.; Silva, A.M.; Junior, W.D.C.; Barbagelata, L.S.; Ferreira, J.A.; Sousa, E.M.A.; da Silva, P.S.; Pinheiro, K.C.; L.C.; Sousa Junior, E.C. |
| EPI_ISL_1171619 | IAL Regional de Presidente Prudente | Instituto Adolfo Lutz, Interdiciplinary Procedures Center, Strategic Laboratory | Claudio Tavares Sacchi, Claudia Regina Gonçalves, Erica Valessa Ramos Gomes, Karoline Rodrigues Campos |
| EPI_ISL_1171648, EPI_ISL_1171649, EPI_ISL_1171650 | Centro de Saude Dr. Jose Paione em Mococa | Instituto Adolfo Lutz, Interdiciplinary Procedures Center, Strategic Laboratory | Claudio Tavares Sacchi, Claudia Regina Gonçalves, Erica Valessa Ramos Gomes, Karoline Rodrigues Campos, Caio Vinicius Dias Lopes |
| EPI_ISL_1171653, EPI_ISL_1171656, EPI_ISL_1171657, EPI_ISL_1171658, EPI_ISL_1171666, EPI_ISL_1171672, EPI_ISL_1171674 | IAL Regional de Presidente Prudente | Instituto Adolfo Lutz, Interdiciplinary Procedures Center, Strategic Laboratory | Claudio Tavares Sacchi, Claudia Regina Gonçalves, Erica Valessa Ramos Gomes, Karoline Rodrigues Campos, Caio Vinicius Dias Lopes |
| EPI_ISL_1182542 | Laboratório Central do Estado do Paraná | Coordenação Geral de Laboratórios de Saúde Pública (CGLAB/DAEVS/SVS/MS) | Vagner Fonseca, et al. |
| EPI_ISL_1182543 | Fundação Ezequiel Dias (FUNED) | Coordenação Geral de Laboratórios de Saúde Pública (CGLAB/DAEVS/SVS/MS) | Vagner Fonseca, et al. |
| EPI_ISL_1182546, EPI_ISL_1182562, EPI_ISL_1182563, EPI_ISL_1182565, EPI_ISL_1182567, EPI_ISL_1182568, EPI_ISL_1182571, EPI_ISL_1182572, EPI_ISL_1182575, EPI_ISL_1182576, EPI_ISL_1182580, EPI_ISL_1182581, EPI_ISL_1182582, EPI_ISL_1182583, EPI_ISL_1182584, EPI_ISL_1182588, EPI_ISL_1182589, EPI_ISL_1182592, EPI_ISL_1182594, EPI_ISL_1182595, EPI_ISL_1182596, EPI_ISL_1182603, EPI_ISL_1182604, EPI_ISL_1182613, EPI_ISL_1182614, EPI_ISL_1182617, EPI_ISL_1182622 | | | |
| see above | Laboratório Central do Estado do Paraná | Coordenação Geral de Laboratórios de Saúde Pública (CGLAB/DAEVS/SVS/MS) | Vagner Fonseca, et al. |
| EPI_ISL_1196296 | Centro de Saude II Dr. Jose Paione Mococa | Instituto Adolfo Lutz, Interdiciplinary Procedures Center, Strategic Laboratory | Claudio Tavares Sacchi, Claudia Regina Gonçalves, Erica Valessa Ramos Gomes, Karoline Rodrigues Campos, Caio Vinicius Dias Lopes |
| EPI_ISL_1196299 | IAL Regional de Marilia | Instituto Adolfo Lutz, Interdiciplinary Procedures Center, Strategic Laboratory | Claudio Tavares Sacchi, Claudia Regina Gonçalves, Erica Valessa Ramos Gomes, Karoline Rodrigues Campos, Caio Vinicius Dias Lopes |
| EPI_ISL_1201884 | Aeroporto Internacional de Guarulhos | Instituto Adolfo Lutz, Interdiciplinary Procedures Center, Strategic Laboratory | Claudio Tavares Sacchi, Claudia Regina Gonçalves, Erica Valessa Ramos Gomes, Karoline Rodrigues Campos, Caio Vinicius Dias Lopes |
| EPI_ISL_1201893 | IAL Regional de Sorocaba | Instituto Adolfo Lutz, Interdiciplinary Procedures Center, Strategic Laboratory | Claudio Tavares Sacchi, Claudia Regina Gonçalves, Erica Valessa Ramos Gomes, Karoline Rodrigues Campos, Caio Vinicius Dias Lopes |
| EPI_ISL_1213149, EPI_ISL_1213151, EPI_ISL_1213158 | Laboratório HLA/UERJ | Bioinformatics Laboratory / LNCC | Alessandra P Lamarca, Luiz G P de Almeida, Ronaldo da Silva Francisco Jr, Lucymara Fassarella Agnez Lima, Kátia Castanho Scortecci, Vinícius Pietta Perez, Otavio J. Brustolini, Eduardo Sérgio Soares Sousa, Danielle Angst Secco, Angela Maria Guimarães Santos, George Rego Albuquerque, Ana Paula Melo Mariano, Bianca Mendes Maciel, Alexandra L Gerber, Ana Paula de C Guimarães, Paulo Ricardo Nascimento, Francisco Paulo Freire Neto, Sandra Rocha Gadelha, Luís Cristóvão Porto, Eloiza Helena Campana, Selma Maria Bezerra Jeronimo, Ana Tereza R Vasconcelos |
| EPI_ISL_1213168, EPI_ISL_1213170 | LAFEM/UESC | Bioinformatics Laboratory / LNCC | Alessandra P Lamarca, Luiz G P de Almeida, Ronaldo da Silva Francisco Jr, Lucymara Fassarella Agnez Lima, Kátia Castanho Scortecci, Vinícius Pietta Perez, Otavio J. Brustolini, Eduardo Sérgio Soares Sousa, Danielle Angst Secco, Angela Maria Guimarães Santos, George Rego Albuquerque, Ana Paula Melo Mariano, Bianca Mendes Maciel, Alexandra L Gerber, Ana Paula de C Guimarães, Paulo Ricardo Nascimento, Francisco Paulo Freire Neto, Sandra Rocha Gadelha, Luís Cristóvão Porto, Eloiza Helena Campana, Selma Maria Bezerra Jeronimo, Ana Tereza R Vasconcelos |
| EPI_ISL_1213171 | Laboratório HLA/UERJ | Bioinformatics Laboratory / LNCC | Alessandra P Lamarca, Luiz G P de Almeida, Ronaldo da Silva Francisco Jr, Lucymara Fassarella Agnez Lima, Kátia Castanho Scortecci, Vinícius Pietta Perez, Otavio J. Brustolini, Eduardo Sérgio Soares Sousa, Danielle Angst Secco, Angela Maria Guimarães Santos, George Rego Albuquerque, Ana Paula Melo Mariano, Bianca Mendes Maciel, Alexandra L Gerber, Ana Paula de C Guimarães, Paulo Ricardo Nascimento, Francisco Paulo Freire Neto, Sandra Rocha Gadelha, Luís Cristóvão Porto, Eloiza Helena Campana, Selma Maria Bezerra Jeronimo, Ana Tereza R Vasconcelos |
| EPI_ISL_1213173, EPI_ISL_1213175, EPI_ISL_1213177, EPI_ISL_1213178, EPI_ISL_1213180, EPI_ISL_1213182, EPI_ISL_1213183, EPI_ISL_1213185, EPI_ISL_1213187 | IMT-UFRN/RN | Bioinformatics Laboratory / LNCC | Alessandra P Lamarca, Luiz G P de Almeida, Ronaldo da Silva Francisco Jr, Lucymara Fassarella Agnez Lima, Kátia Castanho Scortecci, Vinícius Pietta Perez, Otavio J. Brustolini, Eduardo Sérgio Soares Sousa, Danielle Angst Secco, Angela Maria Guimarães Santos, George Rego Albuquerque, Ana Paula Melo Mariano, Bianca Mendes Maciel, Alexandra L Gerber, Ana Paula de C Guimarães, Paulo Ricardo Nascimento, Francisco Paulo Freire Neto, Sandra Rocha Gadelha, Luís Cristóvão Porto, Eloiza Helena Campana, Selma Maria Bezerra Jeronimo, Ana Tereza R Vasconcelos |
| EPI_ISL_1213189, EPI_ISL_1213190 | LBM/UFPB | Bioinformatics Laboratory / LNCC | Alessandra P Lamarca, Luiz G P de Almeida, Ronaldo da Silva Francisco Jr, Lucymara Fassarella Agnez Lima, Kátia Castanho Scortecci, Vinícius Pietta Perez, Otavio J. Brustolini, Eduardo Sérgio Soares Sousa, Danielle Angst Secco, Angela Maria Guimarães Santos, George Rego Albuquerque, Ana Paula Melo Mariano, Bianca Mendes Maciel, Alexandra L Gerber, Ana Paula de C Guimarães, Paulo Ricardo Nascimento, Francisco Paulo Freire Neto, Sandra Rocha Gadelha, Luís Cristóvão Porto, Eloiza Helena Campana, Selma Maria Bezerra Jeronimo, Ana Tereza R Vasconcelos |
| EPI_ISL_1213192, EPI_ISL_1213194, EPI_ISL_1213196, EPI_ISL_1213197, EPI_ISL_1213199 | IMT-UFRN/RN | Bioinformatics Laboratory / LNCC | Alessandra P Lamarca, Luiz G P de Almeida, Ronaldo da Silva Francisco Jr, Lucymara Fassarella Agnez Lima, Kátia Castanho Scortecci, Vinícius Pietta Perez, Otavio J. Brustolini, Eduardo Sérgio Soares Sousa, Danielle Angst Secco, Angela Maria Guimarães Santos, George Rego Albuquerque, Ana Paula Melo Mariano, Bianca Mendes Maciel, Alexandra L Gerber, Ana Paula de C Guimarães, Paulo Ricardo Nascimento, Francisco Paulo Freire Neto, Sandra Rocha Gadelha, Luís Cristóvão Porto, Eloiza Helena Campana, Selma Maria Bezerra Jeronimo, Ana Tereza R Vasconcelos |
| EPI_ISL_1213201, EPI_ISL_1213202, EPI_ISL_1213204, EPI_ISL_1213206, EPI_ISL_1213207 | LBM/UFPB | Bioinformatics Laboratory / LNCC | Alessandra P Lamarca, Luiz G P de Almeida, Ronaldo da Silva Francisco Jr, Lucymara Fassarella Agnez Lima, Kátia Castanho Scortecci, Vinícius Pietta Perez, Otavio J. Brustolini, Eduardo Sérgio Soares Sousa, Danielle Angst Secco, Angela Maria Guimarães Santos, George Rego Albuquerque, Ana Paula Melo Mariano, Bianca Mendes Maciel, Alexandra L Gerber, Ana Paula de C Guimarães, Paulo Ricardo Nascimento, Francisco Paulo Freire Neto, Sandra Rocha Gadelha, Luís Cristóvão Porto, Eloiza Helena Campana, Selma Maria Bezerra Jeronimo, Ana Tereza R Vasconcelos |
| EPI_ISL_1213209 | IMT-UFRN/RN | Bioinformatics Laboratory / LNCC | Alessandra P Lamarca, Luiz G P de Almeida, Ronaldo da Silva Francisco Jr, Lucymara Fassarella Agnez Lima, Kátia Castanho Scortecci, Vinícius Pietta Perez, Otavio J. Brustolini, Eduardo Sérgio Soares Sousa, Danielle Angst Secco, Angela Maria Guimarães Santos, George Rego Albuquerque, Ana Paula Melo Mariano, Bianca Mendes Maciel, Alexandra L Gerber, Ana Paula de C Guimarães, Paulo Ricardo Nascimento, Francisco Paulo Freire Neto, Sandra Rocha Gadelha, Luís Cristóvão Porto, Eloiza Helena Campana, Selma Maria Bezerra Jeronimo, Ana Tereza R Vasconcelos |
| EPI_ISL_1213215 | LBM/UFPB | Bioinformatics Laboratory / LNCC | Alessandra P Lamarca, Luiz G P de Almeida, Ronaldo da Silva Francisco Jr, Lucymara Fassarella Agnez Lima, Kátia Castanho Scortecci, Vinícius Pietta Perez, Otavio J. Brustolini, Eduardo Sérgio Soares Sousa, Danielle Angst Secco, Angela Maria Guimarães Santos, George Rego Albuquerque, Ana Paula Melo Mariano, Bianca Mendes Maciel, Alexandra L Gerber, Ana Paula de C Guimarães, Paulo Ricardo Nascimento, Francisco Paulo Freire Neto, Sandra Rocha Gadelha, Luís Cristóvão Porto, Eloiza Helena Campana, Selma Maria Bezerra Jeronimo, Ana Tereza R Vasconcelos |
| EPI_ISL_1219021 | Hospital Aviccena | Instituto Adolfo Lutz, Interdiciplinary Procedures Center, Strategic Laboratory | Claudio Tavares Sacchi, Claudia Regina Gonçalves, Erica Valessa Ramos Gomes, Karoline Rodrigues Campos, Caio Vinicius Dias Lopes |
| EPI_ISL_1219025 | IAL Regional de Sorocaba | Instituto Adolfo Lutz, Interdiciplinary Procedures Center, Strategic Laboratory | Claudio Tavares Sacchi, Claudia Regina Gonçalves, Erica Valessa Ramos Gomes, Karoline Rodrigues Campos, Caio Vinicius Dias Lopes |

| | | | |
|---|---|---|---|
| EPI_ISL_1219029, EPI_ISL_1219030, EPI_ISL_1219033, EPI_ISL_1219036 | Aeroporto Internacional de Guarulhos | Instituto Adolfo Lutz, Interdiciplinary Procedures Center, Strategic Laboratory | Claudio Tavares Sacchi, Claudia Regina Gonçalves, Erica Valessa Ramos Gomes, Karoline Rodrigues Campos, Caio Vinicius Dias Lopes |
| EPI_ISL_1220095 | Hospital Municipal Gov. Mario Covas Jr. | Instituto Butantan (genome assembly and bioinformatics) and Mendelics (sequencing) | Maria Carolina Quartim Barbosa Elias Sabbaga, Jose Patane, Simone Haddad, Rafael dos Santos Bezerra, Sandra Coccuzzo Sampaio Vessoni, Antonio Jorge Martins, Dimas Tadeu Covas |
| EPI_ISL_1239111 | Laboratório Central de Saúde Pública Noel Nutels | Coordenação Geral de Laboratórios de Saúde Pública (CGLAB) | Vagner Fonseca et al, |
| EPI_ISL_1239119 | Laboratório Central de Saúde Pública do Espírito Santo | Coordenação Geral de Laboratórios de Saúde Pública (CGLAB) | Vagner Fonseca et al, |
| EPI_ISL_1239137, EPI_ISL_1239138 | Fundação Ezequiel Dias | Coordenação Geral de Laboratórios de Saúde Pública (CGLAB) | Vagner Fonseca et al, |
| EPI_ISL_1261683 | LACEN - Laboratório Central de Saúde Pública do Amazonas | Evandro Chagas Institute | Santos, M.C.; Silva, A.M.; Junior, W.D.C.; Barbagelata, L.S.; Ferreira, J.A.; Sousa, E.M.A.; da Silva, P.S.; Pinheiro, K.C.; L.C.; Sousa Junior, E.C. |
| EPI_ISL_1261684 | LACEN - Laboratório Central de Saúde Pública do Ceará | Evandro Chagas Institute | Santos, M.C.; Silva, A.M.; Junior, W.D.C.; Barbagelata, L.S.; Ferreira, J.A.; Sousa, E.M.A.; da Silva, P.S.; Pinheiro, K.C.; L.C.; Sousa Junior, E.C. |
| EPI_ISL_1261685 | LACEN - Laboratório Central de Saúde Pública do Amazonas | Evandro Chagas Institute | Santos, M.C.; Silva, A.M.; Junior, W.D.C.; Barbagelata, L.S.; Ferreira, J.A.; Sousa, E.M.A.; da Silva, P.S.; Pinheiro, K.C.; L.C.; Sousa Junior, E.C. |
| EPI_ISL_1261686 | LACEN - Laboratório Central de Saúde Pública do Amapá | Evandro Chagas Institute | Santos, M.C.; Silva, A.M.; Junior, W.D.C.; Barbagelata, L.S.; Ferreira, J.A.; Sousa, E.M.A.; da Silva, P.S.; Pinheiro, K.C.; L.C.; Sousa Junior, E.C. |
| EPI_ISL_1261687 | LACEN - Laboratório Central de Saúde Pública de Roraima | Evandro Chagas Institute | Santos, M.C.; Silva, A.M.; Junior, W.D.C.; Barbagelata, L.S.; Ferreira, J.A.; Sousa, E.M.A.; da Silva, P.S.; Pinheiro, K.C.; L.C.; Sousa Junior, E.C. |
| EPI_ISL_1261688, EPI_ISL_1261689 | LACEN - Laboratório Central de Saúde Pública do Amapá | Evandro Chagas Institute | Santos, M.C.; Silva, A.M.; Junior, W.D.C.; Barbagelata, L.S.; Ferreira, J.A.; Sousa, E.M.A.; da Silva, P.S.; Pinheiro, K.C.; L.C.; Sousa Junior, E.C. |
| EPI_ISL_1261690 | LACEN - Laboratório Central de Saúde Pública do Amazonas | Evandro Chagas Institute | Santos, M.C.; Silva, A.M.; Junior, W.D.C.; Barbagelata, L.S.; Ferreira, J.A.; Sousa, E.M.A.; da Silva, P.S.; Pinheiro, K.C.; L.C.; Sousa Junior, E.C. |
| EPI_ISL_1261691 | Laboratório Paulo C. Azevedo | Evandro Chagas Institute | Santos, M.C.; Silva, A.M.; Junior, W.D.C.; Barbagelata, L.S.; Ferreira, J.A.; Sousa, E.M.A.; da Silva, P.S.; Pinheiro, K.C.; L.C.; Sousa Junior, E.C. |
| EPI_ISL_1261692 | LACEN - Laboratório Central de Saúde Pública do Amapá | Evandro Chagas Institute | Santos, M.C.; Silva, A.M.; Junior, W.D.C.; Barbagelata, L.S.; Ferreira, J.A.; Sousa, E.M.A.; da Silva, P.S.; Pinheiro, K.C.; L.C.; Sousa Junior, E.C. |
| EPI_ISL_1261693 | LACEN - Laboratório Central de Saúde Pública do Ceará | Evandro Chagas Institute | Santos, M.C.; Silva, A.M.; Junior, W.D.C.; Barbagelata, L.S.; Ferreira, J.A.; Sousa, E.M.A.; da Silva, P.S.; Pinheiro, K.C.; L.C.; Sousa Junior, E.C. |
| EPI_ISL_1261694 | LACEN - Laboratório Central de Saúde Pública do Amazonas | Evandro Chagas Institute | Santos, M.C.; Silva, A.M.; Junior, W.D.C.; Barbagelata, L.S.; Ferreira, J.A.; Sousa, E.M.A.; da Silva, P.S.; Pinheiro, K.C.; L.C.; Sousa Junior, E.C. |
| EPI_ISL_1272188 | Universidade Federal do Norte do Tocantins (UFNT) | Laboratório de Bioinformática e Biotecnologia (Labinftec/UFT) | Ueric José Borges de Souza, Fabrício Souza Campos, Raíssa Nunes dos Santos, José Carlos Ribeiro Júnior, Rogério Fernandes Carvalho, Monike da Silva Oliveira, Bergmann Morais Ribeiro, Fernando Lucas Melo |
| EPI_ISL_1278274, EPI_ISL_1289956, EPI_ISL_1289957, EPI_ISL_1289958, EPI_ISL_1289959, EPI_ISL_1289960 | Laboratory of Virology, Ribeirão Preto General Hospital, Ribeirão Preto Medical School, University of São Paulo | Laboratory of Oncology, Blood Center of Ribeirão Preto, Ribeirão Preto School of Medicine, University of São Paulo | CAMPOS, MR; SANTOS, A.L.P.; YAMAMOTO, A.Y.; COLLI, L.M.; FONSECA, B.A.L.; BELLISSIMO-RODRIGUES, F. |
| EPI_ISL_1290802 | Genomic and molecular Biology Group, A.C.Camargo Cancer Center | Laboratory of Bioinformatics and Computational Biology, A.C.Camargo Cancer Center | Giovana Torrezan, Dirce Carraro, Israel Tojal, Alexandre Defelicibus |
| EPI_ISL_1293053, EPI_ISL_1293054, EPI_ISL_1293055 | LACEN de Rondonia | Instituto Adolfo Lutz, Interdiciplinary Procedures Center, Strategic Laboratory | Claudio Tavares Sacchi, Claudia Regina Gonçalves, Erica Valessa Ramos Gomes, Karoline Rodrigues Campos, Caio Vinicius Dias Lopes |
| EPI_ISL_1293059, EPI_ISL_1293063, EPI_ISL_1293067, EPI_ISL_1293075, EPI_ISL_1293077, EPI_ISL_1293080 | IAL Regional de Sorocaba | Instituto Adolfo Lutz, Interdiciplinary Procedures Center, Strategic Laboratory | Claudio Tavares Sacchi, Claudia Regina Gonçalves, Erica Valessa Ramos Gomes, Karoline Rodrigues Campos, Caio Vinicius Dias Lopes |
| EPI_ISL_1303503 | LACEN de Rondonia | Instituto Adolfo Lutz, Interdiciplinary Procedures Center, Strategic Laboratory | Claudio Tavares Sacchi, Claudia Regina Gonçalves, Erica Valessa Ramos Gomes, Karoline Rodrigues Campos, Caio Vinicius Dias Lopes |
| EPI_ISL_1303515 | LACEN do Estado de Goias | Instituto Adolfo Lutz, Interdiciplinary Procedures Center, Strategic Laboratory | Claudio Tavares Sacchi, Claudia Regina Gonçalves, Erica Valessa Ramos Gomes, Karoline Rodrigues Campos, Caio Vinicius Dias Lopes |
| EPI_ISL_1303522, EPI_ISL_1303525, EPI_ISL_1303529 | IAL Regional de São Jose do Rio Preto | Instituto Adolfo Lutz, Interdiciplinary Procedures Center, Strategic Laboratory | Claudio Tavares Sacchi, Claudia Regina Gonçalves, Erica Valessa Ramos Gomes, Karoline Rodrigues Campos, Caio Vinicius Dias Lopes |
| EPI_ISL_1303536 | Hospital Heliopolis | Instituto Adolfo Lutz, Interdiciplinary Procedures Center, Strategic Laboratory | Claudio Tavares Sacchi, Claudia Regina Gonçalves, Erica Valessa Ramos Gomes, Karoline Rodrigues Campos, Caio Vinicius Dias Lopes |
| EPI_ISL_1303537 | Hospital Estadual de Vila Alpina | Instituto Adolfo Lutz, Interdiciplinary Procedures Center, Strategic Laboratory | Claudio Tavares Sacchi, Claudia Regina Gonçalves, Erica Valessa Ramos Gomes, Karoline Rodrigues Campos, Caio Vinicius Dias Lopes |
| EPI_ISL_1303539 | Hospital Presidente | Instituto Adolfo Lutz, Interdiciplinary Procedures Center, Strategic Laboratory | Claudio Tavares Sacchi, Claudia Regina Gonçalves, Erica Valessa Ramos Gomes, Karoline Rodrigues Campos, Caio Vinicius Dias Lopes |
| EPI_ISL_1303541 | Hospital Municipal Cidade Tiradentes Carmen Prudente | Instituto Adolfo Lutz, Interdiciplinary Procedures Center, Strategic Laboratory | Claudio Tavares Sacchi, Claudia Regina Gonçalves, Erica Valessa Ramos Gomes, Karoline Rodrigues Campos, Caio Vinicius Dias Lopes |
| EPI_ISL_1303542, EPI_ISL_1303543 | Hospital Estadual de Campanha Barradas | Instituto Adolfo Lutz, Interdiciplinary Procedures Center, Strategic Laboratory | Claudio Tavares Sacchi, Claudia Regina Gonçalves, Erica Valessa Ramos Gomes, Karoline Rodrigues Campos, Caio Vinicius Dias Lopes |
| EPI_ISL_1303545 | Hospital Municipal Cidade Tiradentes Carmen Prudente | Instituto Adolfo Lutz, Interdiciplinary Procedures Center, Strategic Laboratory | Claudio Tavares Sacchi, Claudia Regina Gonçalves, Erica Valessa Ramos Gomes, Karoline Rodrigues Campos, Caio Vinicius Dias Lopes |
| EPI_ISL_1303546, EPI_ISL_1303547, EPI_ISL_1303548, EPI_ISL_1303549 | UPA Vila Santa Catarina | Instituto Adolfo Lutz, Interdiciplinary Procedures Center, Strategic Laboratory | Claudio Tavares Sacchi, Claudia Regina Gonçalves, Erica Valessa Ramos Gomes, Karoline Rodrigues Campos, Caio Vinicius Dias Lopes |
| EPI_ISL_1358285 | Centro de Treinamento e Referencia DST AIDS | Instituto Adolfo Lutz, Interdiciplinary Procedures Center, Strategic Laboratory | Claudio Tavares Sacchi, Claudia Regina Gonçalves, Erica Valessa Ramos Gomes, Karoline Rodrigues Campos, Caio Vinicius Dias Lopes |
| EPI_ISL_1358319, EPI_ISL_1358320, EPI_ISL_1358321 | UPA Vila Santa Catarina | Instituto Adolfo Lutz, Interdiciplinary Procedures Center, Strategic Laboratory | Claudio Tavares Sacchi, Claudia Regina Gonçalves, Erica Valessa Ramos Gomes, Karoline Rodrigues Campos, Caio Vinicius Dias Lopes |
| EPI_ISL_1365747 | Associação Fundo de Incentivo a Pesquisa | Associação Fundo de Incentivo a Pesquisa | Priscila Farias Tempaku, Juliana Nogueira Martins Rodrigues, Erika Rodrigues de Oliveira, Soraya Sgambatti de Andrade, Debora Ribeiro Ramadan, Sergio Tufik |
| EPI_ISL_1381045, EPI_ISL_1381047, EPI_ISL_1381048, EPI_ISL_1381050, EPI_ISL_1381052, EPI_ISL_1381059, EPI_ISL_1381060, EPI_ISL_1381061, EPI_ISL_1381062, EPI_ISL_1381065 | IAL Regional de Santo Andre | Instituto Adolfo Lutz, Interdiciplinary Procedures Center, Strategic Laboratory | Claudio Tavares Sacchi, Claudia Regina Gonçalves, Erica Valessa Ramos Gomes, Karoline Rodrigues Campos, Caio Vinicius Dias Lopes |
| EPI_ISL_1381068 | Conjunto Hospitalar do Mandaqui de Sao Paulo | Instituto Adolfo Lutz, Interdiciplinary Procedures Center, Strategic Laboratory | Claudio Tavares Sacchi, Claudia Regina Gonçalves, Erica Valessa Ramos Gomes, Karoline Rodrigues Campos, Caio Vinicius Dias Lopes |
| EPI_ISL_1381069 | Hospital Heliopolis | Instituto Adolfo Lutz, Interdiciplinary Procedures Center, Strategic Laboratory | Claudio Tavares Sacchi, Claudia Regina Gonçalves, Erica Valessa Ramos Gomes, Karoline Rodrigues Campos, Caio Vinicius Dias Lopes |
| EPI_ISL_1381070, EPI_ISL_1381071 | Hospital Municipal Cidade Tiradentes Carmem Prudente | Instituto Adolfo Lutz, Interdiciplinary Procedures Center, | Claudio Tavares Sacchi, Claudia Regina Gonçalves, Erica Valessa Ramos Gomes, Karoline Rodrigues Campos, Caio Vinicius Dias Lopes |

| | | | |
|---|---|---|---|
| | | Strategic Laboratory | |
| EPI_ISL_1443196, EPI_ISL_1443197, EPI_ISL_1443198 | Hospital Aliança | Hospital São Rafael - IDOR | Isadora Cristina de Siqueira, Aquiles Assunção Camelier, Elves A.P. Maciel, Margarida Celia L. C. Neves, Carolina Kymie Vasques Nonaka, Karoline Almeida Félix de Sousa, Victor Costa Araujo, Yasmin Santos Freitas Macêdo, Aurea Angelica Paste, Bruno Solano de Freitas Souza, Tiago Gräf |
| EPI_ISL_1445071 | USF TRES PONTES | Instituto Butantan / Mendelics | Dimas Tadeu Covas, Sandra Coccuzzo Sampaio, Maria Carolina Elias, José Salvatore Leister Patané, Vincent Louis Viala, Antonio Jorge Martins, Ricardo Haddad, Claudia Renata dos Santos Barros, Elaine Cristina Marqueze, Raul Machado Neto, Debora Botequio Moretti, Bibiana Santos, João Paulo Kitajima, Erika Freitas, David Schlesinger, Simone Kashima, Evandra Strazza Rodrigues, Svetoslav Nanev Slavov, Elaine Vieira dos Santos, Rafael dos Santos Bezerra, Luiz Carlos Junior de Alcantara, Marta Giovanetti, Vagner Fonseca, Flavia Aburjaile, Rodrigo Tocantins Calado. |
| EPI_ISL_1445072 | USF JARDIM SAO DIMAS | Instituto Butantan / Mendelics | Dimas Tadeu Covas, Sandra Coccuzzo Sampaio, Maria Carolina Elias, José Salvatore Leister Patané, Vincent Louis Viala, Antonio Jorge Martins, Ricardo Haddad, Claudia Renata dos Santos Barros, Elaine Cristina Marqueze, Raul Machado Neto, Debora Botequio Moretti, Bibiana Santos, João Paulo Kitajima, Erika Freitas, David Schlesinger, Simone Kashima, Evandra Strazza Rodrigues, Svetoslav Nanev Slavov, Elaine Vieira dos Santos, Rafael dos Santos Bezerra, Luiz Carlos Junior de Alcantara, Marta Giovanetti, Vagner Fonseca, Flavia Aburjaile, Rodrigo Tocantins Calado. |
| EPI_ISL_1445078 | CENTRO DE SAUDE II DR GABRIEL MESQUITA VARGEM GDE DO SUL | Instituto Butantan / Mendelics | Dimas Tadeu Covas, Sandra Coccuzzo Sampaio, Maria Carolina Elias, José Salvatore Leister Patané, Vincent Louis Viala, Antonio Jorge Martins, Ricardo Haddad, Claudia Renata dos Santos Barros, Elaine Cristina Marqueze, Raul Machado Neto, Debora Botequio Moretti, Bibiana Santos, João Paulo Kitajima, Erika Freitas, David Schlesinger, Simone Kashima, Evandra Strazza Rodrigues, Svetoslav Nanev Slavov, Elaine Vieira dos Santos, Rafael dos Santos Bezerra, Luiz Carlos Junior de Alcantara, Marta Giovanetti, Vagner Fonseca, Flavia Aburjaile, Rodrigo Tocantins Calado. |
| EPI_ISL_1445082 | SECRETARIA MUNICIPAL DE SAUDE SOROCABA | Instituto Butantan / Mendelics | Dimas Tadeu Covas, Sandra Coccuzzo Sampaio, Maria Carolina Elias, José Salvatore Leister Patané, Vincent Louis Viala, Antonio Jorge Martins, Ricardo Haddad, Claudia Renata dos Santos Barros, Elaine Cristina Marqueze, Raul Machado Neto, Debora Botequio Moretti, Bibiana Santos, João Paulo Kitajima, Erika Freitas, David Schlesinger, Simone Kashima, Evandra Strazza Rodrigues, Svetoslav Nanev Slavov, Elaine Vieira dos Santos, Rafael dos Santos Bezerra, Luiz Carlos Junior de Alcantara, Marta Giovanetti, Vagner Fonseca, Flavia Aburjaile, Rodrigo Tocantins Calado. |
| EPI_ISL_1445084 | SMS SECRETARIA MUNICIPAL DE SAUDE DE BOITUVA | Instituto Butantan / Mendelics | Dimas Tadeu Covas, Sandra Coccuzzo Sampaio, Maria Carolina Elias, José Salvatore Leister Patané, Vincent Louis Viala, Antonio Jorge Martins, Ricardo Haddad, Claudia Renata dos Santos Barros, Elaine Cristina Marqueze, Raul Machado Neto, Debora Botequio Moretti, Bibiana Santos, João Paulo Kitajima, Erika Freitas, David Schlesinger, Simone Kashima, Evandra Strazza Rodrigues, Svetoslav Nanev Slavov, Elaine Vieira dos Santos, Rafael dos Santos Bezerra, Luiz Carlos Junior de Alcantara, Marta Giovanetti, Vagner Fonseca, Flavia Aburjaile, Rodrigo Tocantins Calado. |
| EPI_ISL_1445088 | SECRETARIA MUNICIPAL DE SAUDE SOROCABA | Instituto Butantan / Mendelics | Dimas Tadeu Covas, Sandra Coccuzzo Sampaio, Maria Carolina Elias, José Salvatore Leister Patané, Vincent Louis Viala, Antonio Jorge Martins, Ricardo Haddad, Claudia Renata dos Santos Barros, Elaine Cristina Marqueze, Raul Machado Neto, Debora Botequio Moretti, Bibiana Santos, João Paulo Kitajima, Erika Freitas, David Schlesinger, Simone Kashima, Evandra Strazza Rodrigues, Svetoslav Nanev Slavov, Elaine Vieira dos Santos, Rafael dos Santos Bezerra, Luiz Carlos Junior de Alcantara, Marta Giovanetti, Vagner Fonseca, Flavia Aburjaile, Rodrigo Tocantins Calado. |
| EPI_ISL_1445091, EPI_ISL_1445092, EPI_ISL_1445093, EPI_ISL_1445094, EPI_ISL_1445095 | SECRETARIA MUNICIPAL DE SAUDE DE PIRACAIA | Instituto Butantan / Mendelics | Dimas Tadeu Covas, Sandra Coccuzzo Sampaio, Maria Carolina Elias, José Salvatore Leister Patané, Vincent Louis Viala, Antonio Jorge Martins, Ricardo Haddad, Claudia Renata dos Santos Barros, Elaine Cristina Marqueze, Raul Machado Neto, Debora Botequio Moretti, Bibiana Santos, João Paulo Kitajima, Erika Freitas, David Schlesinger, Simone Kashima, Evandra Strazza Rodrigues, Svetoslav Nanev Slavov, Elaine Vieira dos Santos, Rafael dos Santos Bezerra, Luiz Carlos Junior de Alcantara, Marta Giovanetti, Vagner Fonseca, Flavia Aburjaile, Rodrigo Tocantins Calado. |
| EPI_ISL_1445096, EPI_ISL_1445097, EPI_ISL_1445098, EPI_ISL_1445100 | PRONTO ATENDIMENTO VILA PADRE ANCHIETA | Instituto Butantan / Mendelics | Dimas Tadeu Covas, Sandra Coccuzzo Sampaio, Maria Carolina Elias, José Salvatore Leister Patané, Vincent Louis Viala, Antonio Jorge Martins, Ricardo Haddad, Claudia Renata dos Santos Barros, Elaine Cristina Marqueze, Raul Machado Neto, Debora Botequio Moretti, Bibiana Santos, João Paulo Kitajima, Erika Freitas, David Schlesinger, Simone Kashima, Evandra Strazza Rodrigues, Svetoslav Nanev Slavov, Elaine Vieira dos Santos, Rafael dos Santos Bezerra, Luiz Carlos Junior de Alcantara, Marta Giovanetti, Vagner Fonseca, Flavia Aburjaile, Rodrigo Tocantins Calado. |
| EPI_ISL_1445101 | SECRETARIA MUNICIPAL DE SAUDE DE PIRACAIA | Instituto Butantan / Mendelics | Dimas Tadeu Covas, Sandra Coccuzzo Sampaio, Maria Carolina Elias, José Salvatore Leister Patané, Vincent Louis Viala, Antonio Jorge Martins, Ricardo Haddad, Claudia Renata dos Santos Barros, Elaine Cristina Marqueze, Raul Machado Neto, Debora Botequio Moretti, Bibiana Santos, João Paulo Kitajima, Erika Freitas, David Schlesinger, Simone Kashima, Evandra Strazza Rodrigues, Svetoslav Nanev Slavov, Elaine Vieira dos Santos, Rafael dos Santos Bezerra, Luiz Carlos Junior de Alcantara, Marta Giovanetti, Vagner Fonseca, Flavia Aburjaile, Rodrigo Tocantins Calado. |
| EPI_ISL_1445102, EPI_ISL_1445103, EPI_ISL_1445104, EPI_ISL_1445106 | PRONTO ATENDIMENTO VILA PADRE ANCHIETA | Instituto Butantan / Mendelics | Dimas Tadeu Covas, Sandra Coccuzzo Sampaio, Maria Carolina Elias, José Salvatore Leister Patané, Vincent Louis Viala, Antonio Jorge Martins, Ricardo Haddad, Claudia Renata dos Santos Barros, Elaine Cristina Marqueze, Raul Machado Neto, Debora Botequio Moretti, Bibiana Santos, João Paulo Kitajima, Erika Freitas, David Schlesinger, Simone Kashima, Evandra Strazza Rodrigues, Svetoslav Nanev Slavov, Elaine Vieira dos Santos, Rafael dos Santos Bezerra, Luiz Carlos Junior de Alcantara, Marta Giovanetti, Vagner Fonseca, Flavia Aburjaile, Rodrigo Tocantins Calado. |
| EPI_ISL_1445109, EPI_ISL_1445110 | COMPLEXO HOSPITALAR OURO VERDE DE CAMPINAS | Instituto Butantan / Mendelics | Dimas Tadeu Covas, Sandra Coccuzzo Sampaio, Maria Carolina Elias, José Salvatore Leister Patané, Vincent Louis Viala, Antonio Jorge Martins, Ricardo Haddad, Claudia Renata dos Santos Barros, Elaine Cristina Marqueze, Raul Machado Neto, Debora Botequio Moretti, Bibiana Santos, João Paulo Kitajima, Erika Freitas, David Schlesinger, Simone Kashima, Evandra Strazza Rodrigues, Svetoslav Nanev Slavov, Elaine Vieira dos Santos, Rafael dos Santos Bezerra, Luiz Carlos Junior de Alcantara, Marta Giovanetti, Vagner Fonseca, Flavia Aburjaile, Rodrigo Tocantins Calado. |
| EPI_ISL_1445111, EPI_ISL_1445112 | PRONTO ATENDIMENTO VILA PADRE ANCHIETA | Instituto Butantan / Mendelics | Dimas Tadeu Covas, Sandra Coccuzzo Sampaio, Maria Carolina Elias, José Salvatore Leister Patané, Vincent Louis Viala, Antonio Jorge Martins, Ricardo Haddad, Claudia Renata dos Santos Barros, Elaine Cristina Marqueze, Raul Machado Neto, Debora Botequio Moretti, Bibiana Santos, João Paulo Kitajima, Erika Freitas, David Schlesinger, Simone Kashima, Evandra Strazza Rodrigues, Svetoslav Nanev Slavov, Elaine Vieira dos Santos, Rafael dos Santos Bezerra, Luiz Carlos Junior de Alcantara, Marta Giovanetti, Vagner Fonseca, Flavia Aburjaile, Rodrigo Tocantins Calado. |
| EPI_ISL_1445115, EPI_ISL_1445116, EPI_ISL_1445117, EPI_ISL_1445118 | COMPLEXO HOSPITALAR OURO VERDE DE CAMPINAS | Instituto Butantan / Mendelics | Dimas Tadeu Covas, Sandra Coccuzzo Sampaio, Maria Carolina Elias, José Salvatore Leister Patané, Vincent Louis Viala, Antonio Jorge Martins, Ricardo Haddad, Claudia Renata dos Santos Barros, Elaine Cristina Marqueze, Raul Machado Neto, Debora Botequio Moretti, Bibiana Santos, João Paulo Kitajima, Erika Freitas, David Schlesinger, Simone Kashima, Evandra Strazza Rodrigues, Svetoslav Nanev Slavov, Elaine Vieira dos Santos, Rafael dos Santos Bezerra, Luiz Carlos Junior de Alcantara, Marta Giovanetti, Vagner Fonseca, Flavia Aburjaile, Rodrigo Tocantins Calado. |
| EPI_ISL_1445120, EPI_ISL_1445121, EPI_ISL_1445122, EPI_ISL_1445123, EPI_ISL_1445124, EPI_ISL_1445126 | UNIDADE DE PRONTO ATENDIMENTO UPA DRA ANA OLIVIA BENTIVOGLIO | Instituto Butantan / Mendelics | Dimas Tadeu Covas, Sandra Coccuzzo Sampaio, Maria Carolina Elias, José Salvatore Leister Patané, Vincent Louis Viala, Antonio Jorge Martins, Ricardo Haddad, Claudia Renata dos Santos Barros, Elaine Cristina Marqueze, Raul Machado Neto, Debora Botequio Moretti, Bibiana Santos, João Paulo Kitajima, Erika Freitas, David Schlesinger, Simone Kashima, Evandra Strazza Rodrigues, Svetoslav Nanev Slavov, Elaine Vieira dos Santos, Rafael dos Santos Bezerra, Luiz Carlos Junior de Alcantara, Marta Giovanetti, Vagner Fonseca, Flavia Aburjaile, Rodrigo Tocantins Calado. |
| EPI_ISL_1445127, EPI_ISL_1445128, EPI_ISL_1445129 | UNIDADE RESPIRATORIA NOVA HORTOLANDIA | Instituto Butantan / Mendelics | Dimas Tadeu Covas, Sandra Coccuzzo Sampaio, Maria Carolina Elias, José Salvatore Leister Patané, Vincent Louis Viala, Antonio Jorge Martins, Ricardo Haddad, Claudia Renata dos Santos Barros, Elaine Cristina Marqueze, Raul Machado Neto, Debora Botequio Moretti, Bibiana Santos, João Paulo Kitajima, Erika Freitas, David Schlesinger, Simone Kashima, Evandra Strazza Rodrigues, Svetoslav Nanev Slavov, Elaine Vieira dos Santos, Rafael dos Santos Bezerra, Luiz Carlos Junior de Alcantara, Marta Giovanetti, Vagner Fonseca, Flavia Aburjaile, Rodrigo Tocantins Calado. |
| EPI_ISL_1445132 | PRONTO ATENDIMENTO DA PONTE DE SAO JOAO | Instituto Butantan / Mendelics | Dimas Tadeu Covas, Sandra Coccuzzo Sampaio, Maria Carolina Elias, José Salvatore Leister Patané, Vincent Louis Viala, Antonio Jorge Martins, Ricardo Haddad, Claudia Renata dos Santos Barros, Elaine Cristina Marqueze, Raul Machado Neto, Debora Botequio Moretti, Bibiana Santos, João Paulo Kitajima, Erika Freitas, David Schlesinger, Simone Kashima, Evandra Strazza Rodrigues, Svetoslav Nanev Slavov, Elaine Vieira dos Santos, Rafael dos Santos Bezerra, Luiz Carlos Junior de Alcantara, Marta Giovanetti, Vagner Fonseca, Flavia Aburjaile, Rodrigo Tocantins Calado. |
| EPI_ISL_1445133, EPI_ISL_1445135 | SECRETARIA MUNICIPAL DE SAUDE SOROCABA | Instituto Butantan / Mendelics | Dimas Tadeu Covas, Sandra Coccuzzo Sampaio, Maria Carolina Elias, José Salvatore Leister Patané, Vincent Louis Viala, Antonio Jorge Martins, Ricardo Haddad, Claudia Renata dos Santos Barros, Elaine Cristina Marqueze, Raul Machado Neto, Debora Botequio Moretti, Bibiana Santos, João Paulo Kitajima, Erika Freitas, David Schlesinger, Simone Kashima, Evandra Strazza Rodrigues, Svetoslav Nanev Slavov, Elaine Vieira dos Santos, Rafael dos Santos Bezerra, Luiz Carlos Junior de Alcantara, Marta Giovanetti, Vagner Fonseca, Flavia Aburjaile, Rodrigo Tocantins Calado. |
| EPI_ISL_1445136 | SMS IPERO | Instituto Butantan / Mendelics | Dimas Tadeu Covas, Sandra Coccuzzo Sampaio, Maria Carolina Elias, José Salvatore Leister Patané, Vincent Louis Viala, Antonio Jorge Martins, Ricardo Haddad, Claudia Renata dos Santos Barros, Elaine Cristina Marqueze, Raul Machado Neto, Debora Botequio Moretti, Bibiana Santos, João Paulo Kitajima, Erika Freitas, David Schlesinger, Simone Kashima, Evandra Strazza Rodrigues, Svetoslav Nanev Slavov, Elaine Vieira dos Santos, Rafael dos Santos Bezerra, Luiz Carlos Junior de Alcantara, Marta Giovanetti, Vagner Fonseca, Flavia Aburjaile, Rodrigo Tocantins Calado. |
| EPI_ISL_1445138, EPI_ISL_1445139 | SECRETARIA MUNICIPAL DE SAUDE SOROCABA | Instituto Butantan / Mendelics | Dimas Tadeu Covas, Sandra Coccuzzo Sampaio, Maria Carolina Elias, José Salvatore Leister Patané, Vincent Louis Viala, Antonio Jorge Martins, Ricardo Haddad, Claudia Renata dos Santos Barros, Elaine Cristina Marqueze, Raul Machado Neto, Debora Botequio Moretti, Bibiana Santos, João Paulo |

| | | | |
|---|---|---|---|
| | | | Kitajima, Erika Freitas, David Schlesinger, Simone Kashima, Evandra Strazza Rodrigues, Svetoslav Nanev Slavov, Elaine Vieira dos Santos, Rafael dos Santos Bezerra, Luiz Carlos Junior de Alcantara, Marta Giovanetti, Vagner Fonseca, Flavia Aburjaile, Rodrigo Tocantins Calado. |
| EPI_ISL_1445141 | UBS SALTO DE SAO JOSE | Instituto Butantan / Mendelics | Dimas Tadeu Covas, Sandra Coccuzzo Sampaio, Maria Carolina Elias, José Salvatore Leister Patané, Vincent Louis Viala, Antonio Jorge Martins, Ricardo Haddad, Claudia Renata dos Santos Barros, Elaine Cristina Marqueze, Raul Machado Neto, Debora Botequio Moretti, Bibiana Santos, João Paulo Kitajima, Erika Freitas, David Schlesinger, Simone Kashima, Evandra Strazza Rodrigues, Svetoslav Nanev Slavov, Elaine Vieira dos Santos, Rafael dos Santos Bezerra, Luiz Carlos Junior de Alcantara, Marta Giovanetti, Vagner Fonseca, Flavia Aburjaile, Rodrigo Tocantins Calado. |
| EPI_ISL_1445142, EPI_ISL_1445143, EPI_ISL_1445145 | CS II EGIDIO BRUNHARA MORRO AGUDO | Instituto Butantan / Mendelics | Dimas Tadeu Covas, Sandra Coccuzzo Sampaio, Maria Carolina Elias, José Salvatore Leister Patané, Vincent Louis Viala, Antonio Jorge Martins, Ricardo Haddad, Claudia Renata dos Santos Barros, Elaine Cristina Marqueze, Raul Machado Neto, Debora Botequio Moretti, Bibiana Santos, João Paulo Kitajima, Erika Freitas, David Schlesinger, Simone Kashima, Evandra Strazza Rodrigues, Svetoslav Nanev Slavov, Elaine Vieira dos Santos, Rafael dos Santos Bezerra, Luiz Carlos Junior de Alcantara, Marta Giovanetti, Vagner Fonseca, Flavia Aburjaile, Rodrigo Tocantins Calado. |
| EPI_ISL_1445147, EPI_ISL_1445148, EPI_ISL_1445150, EPI_ISL_1445151, EPI_ISL_1445152 | CENTRO DE REFERENCIA DO IDOSO DR HUMBERTO MENDES DE CARVALHO | Instituto Butantan / Mendelics | Dimas Tadeu Covas, Sandra Coccuzzo Sampaio, Maria Carolina Elias, José Salvatore Leister Patané, Vincent Louis Viala, Antonio Jorge Martins, Ricardo Haddad, Claudia Renata dos Santos Barros, Elaine Cristina Marqueze, Raul Machado Neto, Debora Botequio Moretti, Bibiana Santos, João Paulo Kitajima, Erika Freitas, David Schlesinger, Simone Kashima, Evandra Strazza Rodrigues, Svetoslav Nanev Slavov, Elaine Vieira dos Santos, Rafael dos Santos Bezerra, Luiz Carlos Junior de Alcantara, Marta Giovanetti, Vagner Fonseca, Flavia Aburjaile, Rodrigo Tocantins Calado. |
| EPI_ISL_1445153, EPI_ISL_1445156 | SAE SERVICO DE ATENDIMENTO ESPECIALIZADO | Instituto Butantan / Mendelics | Dimas Tadeu Covas, Sandra Coccuzzo Sampaio, Maria Carolina Elias, José Salvatore Leister Patané, Vincent Louis Viala, Antonio Jorge Martins, Ricardo Haddad, Claudia Renata dos Santos Barros, Elaine Cristina Marqueze, Raul Machado Neto, Debora Botequio Moretti, Bibiana Santos, João Paulo Kitajima, Erika Freitas, David Schlesinger, Simone Kashima, Evandra Strazza Rodrigues, Svetoslav Nanev Slavov, Elaine Vieira dos Santos, Rafael dos Santos Bezerra, Luiz Carlos Junior de Alcantara, Marta Giovanetti, Vagner Fonseca, Flavia Aburjaile, Rodrigo Tocantins Calado. |
| EPI_ISL_1445157, EPI_ISL_1445158, EPI_ISL_1445159, EPI_ISL_1445160 | SECAO CENTRO DE DIAGNOSTICO SECEDI | Instituto Butantan / Mendelics | Dimas Tadeu Covas, Sandra Coccuzzo Sampaio, Maria Carolina Elias, José Salvatore Leister Patané, Vincent Louis Viala, Antonio Jorge Martins, Ricardo Haddad, Claudia Renata dos Santos Barros, Elaine Cristina Marqueze, Raul Machado Neto, Debora Botequio Moretti, Bibiana Santos, João Paulo Kitajima, Erika Freitas, David Schlesinger, Simone Kashima, Evandra Strazza Rodrigues, Svetoslav Nanev Slavov, Elaine Vieira dos Santos, Rafael dos Santos Bezerra, Luiz Carlos Junior de Alcantara, Marta Giovanetti, Vagner Fonseca, Flavia Aburjaile, Rodrigo Tocantins Calado. |
| EPI_ISL_1445162, EPI_ISL_1445163, EPI_ISL_1445164, EPI_ISL_1445165, EPI_ISL_1445166, EPI_ISL_1445167 | SERV DE VIG SANITARIA EPIDEMIO E CTRL DE ZOONOZES GUARUJA | Instituto Butantan / Mendelics | Dimas Tadeu Covas, Sandra Coccuzzo Sampaio, Maria Carolina Elias, José Salvatore Leister Patané, Vincent Louis Viala, Antonio Jorge Martins, Ricardo Haddad, Claudia Renata dos Santos Barros, Elaine Cristina Marqueze, Raul Machado Neto, Debora Botequio Moretti, Bibiana Santos, João Paulo Kitajima, Erika Freitas, David Schlesinger, Simone Kashima, Evandra Strazza Rodrigues, Svetoslav Nanev Slavov, Elaine Vieira dos Santos, Rafael dos Santos Bezerra, Luiz Carlos Junior de Alcantara, Marta Giovanetti, Vagner Fonseca, Flavia Aburjaile, Rodrigo Tocantins Calado. |
| EPI_ISL_1445169, EPI_ISL_1445170 | SECAO CENTRO DE DIAGNOSTICO SECEDI | Instituto Butantan / Mendelics | Dimas Tadeu Covas, Sandra Coccuzzo Sampaio, Maria Carolina Elias, José Salvatore Leister Patané, Vincent Louis Viala, Antonio Jorge Martins, Ricardo Haddad, Claudia Renata dos Santos Barros, Elaine Cristina Marqueze, Raul Machado Neto, Debora Botequio Moretti, Bibiana Santos, João Paulo Kitajima, Erika Freitas, David Schlesinger, Simone Kashima, Evandra Strazza Rodrigues, Svetoslav Nanev Slavov, Elaine Vieira dos Santos, Rafael dos Santos Bezerra, Luiz Carlos Junior de Alcantara, Marta Giovanetti, Vagner Fonseca, Flavia Aburjaile, Rodrigo Tocantins Calado. |
| EPI_ISL_1445172 | UNIDADE MISTA ARACOIABA DA SERRA | Instituto Butantan / Mendelics | Dimas Tadeu Covas, Sandra Coccuzzo Sampaio, Maria Carolina Elias, José Salvatore Leister Patané, Vincent Louis Viala, Antonio Jorge Martins, Ricardo Haddad, Claudia Renata dos Santos Barros, Elaine Cristina Marqueze, Raul Machado Neto, Debora Botequio Moretti, Bibiana Santos, João Paulo Kitajima, Erika Freitas, David Schlesinger, Simone Kashima, Evandra Strazza Rodrigues, Svetoslav Nanev Slavov, Elaine Vieira dos Santos, Rafael dos Santos Bezerra, Luiz Carlos Junior de Alcantara, Marta Giovanetti, Vagner Fonseca, Flavia Aburjaile, Rodrigo Tocantins Calado. |
| EPI_ISL_1445174 | PRONTO SOCORRO MUNICIPAL DE TATUI | Instituto Butantan / Mendelics | Dimas Tadeu Covas, Sandra Coccuzzo Sampaio, Maria Carolina Elias, José Salvatore Leister Patané, Vincent Louis Viala, Antonio Jorge Martins, Ricardo Haddad, Claudia Renata dos Santos Barros, Elaine Cristina Marqueze, Raul Machado Neto, Debora Botequio Moretti, Bibiana Santos, João Paulo Kitajima, Erika Freitas, David Schlesinger, Simone Kashima, Evandra Strazza Rodrigues, Svetoslav Nanev Slavov, Elaine Vieira dos Santos, Rafael dos Santos Bezerra, Luiz Carlos Junior de Alcantara, Marta Giovanetti, Vagner Fonseca, Flavia Aburjaile, Rodrigo Tocantins Calado. |
| EPI_ISL_1445178, EPI_ISL_1445180 | SECRETARIA MUNICIPAL DE SAUDE SOROCABA | Instituto Butantan / Mendelics | Dimas Tadeu Covas, Sandra Coccuzzo Sampaio, Maria Carolina Elias, José Salvatore Leister Patané, Vincent Louis Viala, Antonio Jorge Martins, Ricardo Haddad, Claudia Renata dos Santos Barros, Elaine Cristina Marqueze, Raul Machado Neto, Debora Botequio Moretti, Bibiana Santos, João Paulo Kitajima, Erika Freitas, David Schlesinger, Simone Kashima, Evandra Strazza Rodrigues, Svetoslav Nanev Slavov, Elaine Vieira dos Santos, Rafael dos Santos Bezerra, Luiz Carlos Junior de Alcantara, Marta Giovanetti, Vagner Fonseca, Flavia Aburjaile, Rodrigo Tocantins Calado. |
| EPI_ISL_1445181 | HOSPITAL REGIONAL DE ITAPETININGA | Instituto Butantan / Mendelics | Dimas Tadeu Covas, Sandra Coccuzzo Sampaio, Maria Carolina Elias, José Salvatore Leister Patané, Vincent Louis Viala, Antonio Jorge Martins, Ricardo Haddad, Claudia Renata dos Santos Barros, Elaine Cristina Marqueze, Raul Machado Neto, Debora Botequio Moretti, Bibiana Santos, João Paulo Kitajima, Erika Freitas, David Schlesinger, Simone Kashima, Evandra Strazza Rodrigues, Svetoslav Nanev Slavov, Elaine Vieira dos Santos, Rafael dos Santos Bezerra, Luiz Carlos Junior de Alcantara, Marta Giovanetti, Vagner Fonseca, Flavia Aburjaile, Rodrigo Tocantins Calado. |
| EPI_ISL_1445182 | SECRETARIA MUNICIPAL DE SAUDE SOROCABA | Instituto Butantan / Mendelics | Dimas Tadeu Covas, Sandra Coccuzzo Sampaio, Maria Carolina Elias, José Salvatore Leister Patané, Vincent Louis Viala, Antonio Jorge Martins, Ricardo Haddad, Claudia Renata dos Santos Barros, Elaine Cristina Marqueze, Raul Machado Neto, Debora Botequio Moretti, Bibiana Santos, João Paulo Kitajima, Erika Freitas, David Schlesinger, Simone Kashima, Evandra Strazza Rodrigues, Svetoslav Nanev Slavov, Elaine Vieira dos Santos, Rafael dos Santos Bezerra, Luiz Carlos Junior de Alcantara, Marta Giovanetti, Vagner Fonseca, Flavia Aburjaile, Rodrigo Tocantins Calado. |
| EPI_ISL_1445183 | UNIDADE BASICA DE SAUDE DA FAMILIA BILAC | Instituto Butantan / Mendelics | Dimas Tadeu Covas, Sandra Coccuzzo Sampaio, Maria Carolina Elias, José Salvatore Leister Patané, Vincent Louis Viala, Antonio Jorge Martins, Ricardo Haddad, Claudia Renata dos Santos Barros, Elaine Cristina Marqueze, Raul Machado Neto, Debora Botequio Moretti, Bibiana Santos, João Paulo Kitajima, Erika Freitas, David Schlesinger, Simone Kashima, Evandra Strazza Rodrigues, Svetoslav Nanev Slavov, Elaine Vieira dos Santos, Rafael dos Santos Bezerra, Luiz Carlos Junior de Alcantara, Marta Giovanetti, Vagner Fonseca, Flavia Aburjaile, Rodrigo Tocantins Calado. |
| EPI_ISL_1445187 | PRONTO ATENDIMENTO SAO JOSE | Instituto Butantan / Mendelics | Dimas Tadeu Covas, Sandra Coccuzzo Sampaio, Maria Carolina Elias, José Salvatore Leister Patané, Vincent Louis Viala, Antonio Jorge Martins, Ricardo Haddad, Claudia Renata dos Santos Barros, Elaine Cristina Marqueze, Raul Machado Neto, Debora Botequio Moretti, Bibiana Santos, João Paulo Kitajima, Erika Freitas, David Schlesinger, Simone Kashima, Evandra Strazza Rodrigues, Svetoslav Nanev Slavov, Elaine Vieira dos Santos, Rafael dos Santos Bezerra, Luiz Carlos Junior de Alcantara, Marta Giovanetti, Vagner Fonseca, Flavia Aburjaile, Rodrigo Tocantins Calado. |
| EPI_ISL_1445188, EPI_ISL_1445189, EPI_ISL_1445190, EPI_ISL_1445191, EPI_ISL_1445192 | PRONTO ATENDIMENTO VILA PADRE ANCHIETA | Instituto Butantan / Mendelics | Dimas Tadeu Covas, Sandra Coccuzzo Sampaio, Maria Carolina Elias, José Salvatore Leister Patané, Vincent Louis Viala, Antonio Jorge Martins, Ricardo Haddad, Claudia Renata dos Santos Barros, Elaine Cristina Marqueze, Raul Machado Neto, Debora Botequio Moretti, Bibiana Santos, João Paulo Kitajima, Erika Freitas, David Schlesinger, Simone Kashima, Evandra Strazza Rodrigues, Svetoslav Nanev Slavov, Elaine Vieira dos Santos, Rafael dos Santos Bezerra, Luiz Carlos Junior de Alcantara, Marta Giovanetti, Vagner Fonseca, Flavia Aburjaile, Rodrigo Tocantins Calado. |
| EPI_ISL_1445193, EPI_ISL_1445194 | PRONTO ATENDIMENTO SAO JOSE | Instituto Butantan / Mendelics | Dimas Tadeu Covas, Sandra Coccuzzo Sampaio, Maria Carolina Elias, José Salvatore Leister Patané, Vincent Louis Viala, Antonio Jorge Martins, Ricardo Haddad, Claudia Renata dos Santos Barros, Elaine Cristina Marqueze, Raul Machado Neto, Debora Botequio Moretti, Bibiana Santos, João Paulo Kitajima, Erika Freitas, David Schlesinger, Simone Kashima, Evandra Strazza Rodrigues, Svetoslav Nanev Slavov, Elaine Vieira dos Santos, Rafael dos Santos Bezerra, Luiz Carlos Junior de Alcantara, Marta Giovanetti, Vagner Fonseca, Flavia Aburjaile, Rodrigo Tocantins Calado. |
| EPI_ISL_1445196 | UBS SAO JUDAS | Instituto Butantan / Mendelics | Dimas Tadeu Covas, Sandra Coccuzzo Sampaio, Maria Carolina Elias, José Salvatore Leister Patané, Vincent Louis Viala, Antonio Jorge Martins, Ricardo Haddad, Claudia Renata dos Santos Barros, Elaine Cristina Marqueze, Raul Machado Neto, Debora Botequio Moretti, Bibiana Santos, João Paulo Kitajima, Erika Freitas, David Schlesinger, Simone Kashima, Evandra Strazza Rodrigues, Svetoslav Nanev Slavov, Elaine Vieira dos Santos, Rafael dos Santos Bezerra, Luiz Carlos Junior de Alcantara, Marta Giovanetti, Vagner Fonseca, Flavia Aburjaile, Rodrigo Tocantins Calado. |
| EPI_ISL_1445198 | UNIDADE MISTA DE TAIACU | Instituto Butantan / Mendelics | Dimas Tadeu Covas, Sandra Coccuzzo Sampaio, Maria Carolina Elias, José Salvatore Leister Patané, Vincent Louis Viala, Antonio Jorge Martins, Ricardo Haddad, Claudia Renata dos Santos Barros, Elaine Cristina Marqueze, Raul Machado Neto, Debora Botequio Moretti, Bibiana Santos, João Paulo Kitajima, Erika Freitas, David Schlesinger, Simone Kashima, Evandra Strazza Rodrigues, Svetoslav Nanev Slavov, Elaine Vieira dos Santos, Rafael dos Santos Bezerra, Luiz Carlos Junior de Alcantara, Marta Giovanetti, Vagner Fonseca, Flavia Aburjaile, Rodrigo Tocantins Calado. |
| EPI_ISL_1445200 | HOSPITAL E MATERNIDADE NOSSA SENHORA DA AJUDA | Instituto Butantan / Mendelics | Dimas Tadeu Covas, Sandra Coccuzzo Sampaio, Maria Carolina Elias, José Salvatore Leister Patané, Vincent Louis Viala, Antonio Jorge Martins, Ricardo Haddad, Claudia Renata dos Santos Barros, Elaine Cristina Marqueze, Raul Machado Neto, Debora Botequio Moretti, Bibiana Santos, João Paulo Kitajima, Erika Freitas, David Schlesinger, Simone Kashima, Evandra Strazza Rodrigues, Svetoslav Nanev Slavov, Elaine Vieira dos Santos, Rafael dos Santos Bezerra, Luiz Carlos Junior de Alcantara, Marta Giovanetti, Vagner Fonseca, Flavia Aburjaile, Rodrigo Tocantins Calado. |

| | | | |
|---|---|---|---|
| EPI_ISL_1445202 | USAFA MARACANA | Instituto Butantan / Mendelics | Dimas Tadeu Covas, Sandra Coccuzzo Sampaio, Maria Carolina Elias, José Salvatore Leister Patané, Vincent Louis Viala, Antonio Jorge Martins, Ricardo Haddad, Claudia Renata dos Santos Barros, Elaine Cristina Marqueze, Raul Machado Neto, Debora Botequio Moretti, Bibiana Santos, João Paulo Kitajima, Erika Freitas, David Schlesinger, Simone Kashima, Evandra Strazza Rodrigues, Svetoslav Nanev Slavov, Elaine Vieira dos Santos, Rafael dos Santos Bezerra, Luiz Carlos Junior de Alcantara, Marta Giovanetti, Vagner Fonseca, Flavia Aburjaile, Rodrigo Tocantins Calado. |
| EPI_ISL_1445203 | USAFA GUILHERMINA | Instituto Butantan / Mendelics | Dimas Tadeu Covas, Sandra Coccuzzo Sampaio, Maria Carolina Elias, José Salvatore Leister Patané, Vincent Louis Viala, Antonio Jorge Martins, Ricardo Haddad, Claudia Renata dos Santos Barros, Elaine Cristina Marqueze, Raul Machado Neto, Debora Botequio Moretti, Bibiana Santos, João Paulo Kitajima, Erika Freitas, David Schlesinger, Simone Kashima, Evandra Strazza Rodrigues, Svetoslav Nanev Slavov, Elaine Vieira dos Santos, Rafael dos Santos Bezerra, Luiz Carlos Junior de Alcantara, Marta Giovanetti, Vagner Fonseca, Flavia Aburjaile, Rodrigo Tocantins Calado. |
| EPI_ISL_1445204 | USAFA MARACANA | Instituto Butantan / Mendelics | Dimas Tadeu Covas, Sandra Coccuzzo Sampaio, Maria Carolina Elias, José Salvatore Leister Patané, Vincent Louis Viala, Antonio Jorge Martins, Ricardo Haddad, Claudia Renata dos Santos Barros, Elaine Cristina Marqueze, Raul Machado Neto, Debora Botequio Moretti, Bibiana Santos, João Paulo Kitajima, Erika Freitas, David Schlesinger, Simone Kashima, Evandra Strazza Rodrigues, Svetoslav Nanev Slavov, Elaine Vieira dos Santos, Rafael dos Santos Bezerra, Luiz Carlos Junior de Alcantara, Marta Giovanetti, Vagner Fonseca, Flavia Aburjaile, Rodrigo Tocantins Calado. |
| EPI_ISL_1445205 | UBS FRANCISCA LIMA DE LIRA | Instituto Butantan / Mendelics | Dimas Tadeu Covas, Sandra Coccuzzo Sampaio, Maria Carolina Elias, José Salvatore Leister Patané, Vincent Louis Viala, Antonio Jorge Martins, Ricardo Haddad, Claudia Renata dos Santos Barros, Elaine Cristina Marqueze, Raul Machado Neto, Debora Botequio Moretti, Bibiana Santos, João Paulo Kitajima, Erika Freitas, David Schlesinger, Simone Kashima, Evandra Strazza Rodrigues, Svetoslav Nanev Slavov, Elaine Vieira dos Santos, Rafael dos Santos Bezerra, Luiz Carlos Junior de Alcantara, Marta Giovanetti, Vagner Fonseca, Flavia Aburjaile, Rodrigo Tocantins Calado. |
| EPI_ISL_1445206 | UBS DARCY ALVES E ROBALINHO | Instituto Butantan / Mendelics | Dimas Tadeu Covas, Sandra Coccuzzo Sampaio, Maria Carolina Elias, José Salvatore Leister Patané, Vincent Louis Viala, Antonio Jorge Martins, Ricardo Haddad, Claudia Renata dos Santos Barros, Elaine Cristina Marqueze, Raul Machado Neto, Debora Botequio Moretti, Bibiana Santos, João Paulo Kitajima, Erika Freitas, David Schlesinger, Simone Kashima, Evandra Strazza Rodrigues, Svetoslav Nanev Slavov, Elaine Vieira dos Santos, Rafael dos Santos Bezerra, Luiz Carlos Junior de Alcantara, Marta Giovanetti, Vagner Fonseca, Flavia Aburjaile, Rodrigo Tocantins Calado. |
| EPI_ISL_1445207 | PPA MARCIO VALDEVINO BATISTA | Instituto Butantan / Mendelics | Dimas Tadeu Covas, Sandra Coccuzzo Sampaio, Maria Carolina Elias, José Salvatore Leister Patané, Vincent Louis Viala, Antonio Jorge Martins, Ricardo Haddad, Claudia Renata dos Santos Barros, Elaine Cristina Marqueze, Raul Machado Neto, Debora Botequio Moretti, Bibiana Santos, João Paulo Kitajima, Erika Freitas, David Schlesinger, Simone Kashima, Evandra Strazza Rodrigues, Svetoslav Nanev Slavov, Elaine Vieira dos Santos, Rafael dos Santos Bezerra, Luiz Carlos Junior de Alcantara, Marta Giovanetti, Vagner Fonseca, Flavia Aburjaile, Rodrigo Tocantins Calado. |
| EPI_ISL_1445208 | UBS DARCY ALVES E ROBALINHO | Instituto Butantan / Mendelics | Dimas Tadeu Covas, Sandra Coccuzzo Sampaio, Maria Carolina Elias, José Salvatore Leister Patané, Vincent Louis Viala, Antonio Jorge Martins, Ricardo Haddad, Claudia Renata dos Santos Barros, Elaine Cristina Marqueze, Raul Machado Neto, Debora Botequio Moretti, Bibiana Santos, João Paulo Kitajima, Erika Freitas, David Schlesinger, Simone Kashima, Evandra Strazza Rodrigues, Svetoslav Nanev Slavov, Elaine Vieira dos Santos, Rafael dos Santos Bezerra, Luiz Carlos Junior de Alcantara, Marta Giovanetti, Vagner Fonseca, Flavia Aburjaile, Rodrigo Tocantins Calado. |
| EPI_ISL_1445209 | UBS OTACILIO FIRMINO LOPES | Instituto Butantan / Mendelics | Dimas Tadeu Covas, Sandra Coccuzzo Sampaio, Maria Carolina Elias, José Salvatore Leister Patané, Vincent Louis Viala, Antonio Jorge Martins, Ricardo Haddad, Claudia Renata dos Santos Barros, Elaine Cristina Marqueze, Raul Machado Neto, Debora Botequio Moretti, Bibiana Santos, João Paulo Kitajima, Erika Freitas, David Schlesinger, Simone Kashima, Evandra Strazza Rodrigues, Svetoslav Nanev Slavov, Elaine Vieira dos Santos, Rafael dos Santos Bezerra, Luiz Carlos Junior de Alcantara, Marta Giovanetti, Vagner Fonseca, Flavia Aburjaile, Rodrigo Tocantins Calado. |
| EPI_ISL_1445210, EPI_ISL_1445211 | UBS JOSE SABINO FERREIRA | Instituto Butantan / Mendelics | Dimas Tadeu Covas, Sandra Coccuzzo Sampaio, Maria Carolina Elias, José Salvatore Leister Patané, Vincent Louis Viala, Antonio Jorge Martins, Ricardo Haddad, Claudia Renata dos Santos Barros, Elaine Cristina Marqueze, Raul Machado Neto, Debora Botequio Moretti, Bibiana Santos, João Paulo Kitajima, Erika Freitas, David Schlesinger, Simone Kashima, Evandra Strazza Rodrigues, Svetoslav Nanev Slavov, Elaine Vieira dos Santos, Rafael dos Santos Bezerra, Luiz Carlos Junior de Alcantara, Marta Giovanetti, Vagner Fonseca, Flavia Aburjaile, Rodrigo Tocantins Calado. |
| EPI_ISL_1445212 | UBS IRMA AGUEDA MARIA JAIME | Instituto Butantan / Mendelics | Dimas Tadeu Covas, Sandra Coccuzzo Sampaio, Maria Carolina Elias, José Salvatore Leister Patané, Vincent Louis Viala, Antonio Jorge Martins, Ricardo Haddad, Claudia Renata dos Santos Barros, Elaine Cristina Marqueze, Raul Machado Neto, Debora Botequio Moretti, Bibiana Santos, João Paulo Kitajima, Erika Freitas, David Schlesinger, Simone Kashima, Evandra Strazza Rodrigues, Svetoslav Nanev Slavov, Elaine Vieira dos Santos, Rafael dos Santos Bezerra, Luiz Carlos Junior de Alcantara, Marta Giovanetti, Vagner Fonseca, Flavia Aburjaile, Rodrigo Tocantins Calado. |
| EPI_ISL_1445213 | UBS OTACILIO FIRMINO LOPES | Instituto Butantan / Mendelics | Dimas Tadeu Covas, Sandra Coccuzzo Sampaio, Maria Carolina Elias, José Salvatore Leister Patané, Vincent Louis Viala, Antonio Jorge Martins, Ricardo Haddad, Claudia Renata dos Santos Barros, Elaine Cristina Marqueze, Raul Machado Neto, Debora Botequio Moretti, Bibiana Santos, João Paulo Kitajima, Erika Freitas, David Schlesinger, Simone Kashima, Evandra Strazza Rodrigues, Svetoslav Nanev Slavov, Elaine Vieira dos Santos, Rafael dos Santos Bezerra, Luiz Carlos Junior de Alcantara, Marta Giovanetti, Vagner Fonseca, Flavia Aburjaile, Rodrigo Tocantins Calado. |
| EPI_ISL_1445214 | UBS JOSE HILARIO DOS SANTOS | Instituto Butantan / Mendelics | Dimas Tadeu Covas, Sandra Coccuzzo Sampaio, Maria Carolina Elias, José Salvatore Leister Patané, Vincent Louis Viala, Antonio Jorge Martins, Ricardo Haddad, Claudia Renata dos Santos Barros, Elaine Cristina Marqueze, Raul Machado Neto, Debora Botequio Moretti, Bibiana Santos, João Paulo Kitajima, Erika Freitas, David Schlesinger, Simone Kashima, Evandra Strazza Rodrigues, Svetoslav Nanev Slavov, Elaine Vieira dos Santos, Rafael dos Santos Bezerra, Luiz Carlos Junior de Alcantara, Marta Giovanetti, Vagner Fonseca, Flavia Aburjaile, Rodrigo Tocantins Calado. |
| EPI_ISL_1445215, EPI_ISL_1445216 | UBS HELENA MARREY | Instituto Butantan / Mendelics | Dimas Tadeu Covas, Sandra Coccuzzo Sampaio, Maria Carolina Elias, José Salvatore Leister Patané, Vincent Louis Viala, Antonio Jorge Martins, Ricardo Haddad, Claudia Renata dos Santos Barros, Elaine Cristina Marqueze, Raul Machado Neto, Debora Botequio Moretti, Bibiana Santos, João Paulo Kitajima, Erika Freitas, David Schlesinger, Simone Kashima, Evandra Strazza Rodrigues, Svetoslav Nanev Slavov, Elaine Vieira dos Santos, Rafael dos Santos Bezerra, Luiz Carlos Junior de Alcantara, Marta Giovanetti, Vagner Fonseca, Flavia Aburjaile, Rodrigo Tocantins Calado. |
| EPI_ISL_1445217 | UBS EMILIA COSME CERQUEIRA | Instituto Butantan / Mendelics | Dimas Tadeu Covas, Sandra Coccuzzo Sampaio, Maria Carolina Elias, José Salvatore Leister Patané, Vincent Louis Viala, Antonio Jorge Martins, Ricardo Haddad, Claudia Renata dos Santos Barros, Elaine Cristina Marqueze, Raul Machado Neto, Debora Botequio Moretti, Bibiana Santos, João Paulo Kitajima, Erika Freitas, David Schlesinger, Simone Kashima, Evandra Strazza Rodrigues, Svetoslav Nanev Slavov, Elaine Vieira dos Santos, Rafael dos Santos Bezerra, Luiz Carlos Junior de Alcantara, Marta Giovanetti, Vagner Fonseca, Flavia Aburjaile, Rodrigo Tocantins Calado. |
| EPI_ISL_1445218 | UBS SYLVIO JOAO L DE LUCIA | Instituto Butantan / Mendelics | Dimas Tadeu Covas, Sandra Coccuzzo Sampaio, Maria Carolina Elias, José Salvatore Leister Patané, Vincent Louis Viala, Antonio Jorge Martins, Ricardo Haddad, Claudia Renata dos Santos Barros, Elaine Cristina Marqueze, Raul Machado Neto, Debora Botequio Moretti, Bibiana Santos, João Paulo Kitajima, Erika Freitas, David Schlesinger, Simone Kashima, Evandra Strazza Rodrigues, Svetoslav Nanev Slavov, Elaine Vieira dos Santos, Rafael dos Santos Bezerra, Luiz Carlos Junior de Alcantara, Marta Giovanetti, Vagner Fonseca, Flavia Aburjaile, Rodrigo Tocantins Calado. |
| EPI_ISL_1445219, EPI_ISL_1445220 | UBS EMILIA COSME CERQUEIRA | Instituto Butantan / Mendelics | Dimas Tadeu Covas, Sandra Coccuzzo Sampaio, Maria Carolina Elias, José Salvatore Leister Patané, Vincent Louis Viala, Antonio Jorge Martins, Ricardo Haddad, Claudia Renata dos Santos Barros, Elaine Cristina Marqueze, Raul Machado Neto, Debora Botequio Moretti, Bibiana Santos, João Paulo Kitajima, Erika Freitas, David Schlesinger, Simone Kashima, Evandra Strazza Rodrigues, Svetoslav Nanev Slavov, Elaine Vieira dos Santos, Rafael dos Santos Bezerra, Luiz Carlos Junior de Alcantara, Marta Giovanetti, Vagner Fonseca, Flavia Aburjaile, Rodrigo Tocantins Calado. |
| EPI_ISL_1445221, EPI_ISL_1445223 | UBS HELENA MARREY | Instituto Butantan / Mendelics | Dimas Tadeu Covas, Sandra Coccuzzo Sampaio, Maria Carolina Elias, José Salvatore Leister Patané, Vincent Louis Viala, Antonio Jorge Martins, Ricardo Haddad, Claudia Renata dos Santos Barros, Elaine Cristina Marqueze, Raul Machado Neto, Debora Botequio Moretti, Bibiana Santos, João Paulo Kitajima, Erika Freitas, David Schlesinger, Simone Kashima, Evandra Strazza Rodrigues, Svetoslav Nanev Slavov, Elaine Vieira dos Santos, Rafael dos Santos Bezerra, Luiz Carlos Junior de Alcantara, Marta Giovanetti, Vagner Fonseca, Flavia Aburjaile, Rodrigo Tocantins Calado. |
| EPI_ISL_1445224 | PPA FENELON GUEDES PEREIRA | Instituto Butantan / Mendelics | Dimas Tadeu Covas, Sandra Coccuzzo Sampaio, Maria Carolina Elias, José Salvatore Leister Patané, Vincent Louis Viala, Antonio Jorge Martins, Ricardo Haddad, Claudia Renata dos Santos Barros, Elaine Cristina Marqueze, Raul Machado Neto, Debora Botequio Moretti, Bibiana Santos, João Paulo Kitajima, Erika Freitas, David Schlesinger, Simone Kashima, Evandra Strazza Rodrigues, Svetoslav Nanev Slavov, Elaine Vieira dos Santos, Rafael dos Santos Bezerra, Luiz Carlos Junior de Alcantara, Marta Giovanetti, Vagner Fonseca, Flavia Aburjaile, Rodrigo Tocantins Calado. |
| EPI_ISL_1445225 | PRONTO SOCORRO DR ANTONIO FLAVIO FRANCA | Instituto Butantan / Mendelics | Dimas Tadeu Covas, Sandra Coccuzzo Sampaio, Maria Carolina Elias, José Salvatore Leister Patané, Vincent Louis Viala, Antonio Jorge Martins, Ricardo Haddad, Claudia Renata dos Santos Barros, Elaine Cristina Marqueze, Raul Machado Neto, Debora Botequio Moretti, Bibiana Santos, João Paulo Kitajima, Erika Freitas, David Schlesinger, Simone Kashima, Evandra Strazza Rodrigues, Svetoslav Nanev Slavov, Elaine Vieira dos Santos, Rafael dos Santos Bezerra, Luiz Carlos Junior de Alcantara, Marta Giovanetti, Vagner Fonseca, Flavia Aburjaile, Rodrigo Tocantins Calado. |
| EPI_ISL_1445226 | UBS SYLVIO JOAO L DE LUCIA | Instituto Butantan / Mendelics | Dimas Tadeu Covas, Sandra Coccuzzo Sampaio, Maria Carolina Elias, José Salvatore Leister Patané, Vincent Louis Viala, Antonio Jorge Martins, Ricardo Haddad, Claudia Renata dos Santos Barros, Elaine Cristina Marqueze, Raul Machado Neto, Debora Botequio Moretti, Bibiana Santos, João Paulo |

| | | | |
|---|---|---|---|
| | | | Kitajima, Erika Freitas, David Schlesinger, Simone Kashima, Evandra Strazza Rodrigues, Svetoslav Nanev Slavov, Elaine Vieira dos Santos, Rafael dos Santos Bezerra, Luiz Carlos Junior de Alcantara, Marta Giovanetti, Vagner Fonseca, Flavia Aburjaile, Rodrigo Tocantins Calado. |
| EPI_ISL_1445227 | UBS EMILIA COSME CERQUEIRA | Instituto Butantan / Mendelics | Dimas Tadeu Covas, Sandra Coccuzzo Sampaio, Maria Carolina Elias, José Salvatore Leister Patané, Vincent Louis Viala, Antonio Jorge Martins, Ricardo Haddad, Claudia Renata dos Santos Barros, Elaine Cristina Marqueze, Raul Machado Neto, Debora Botequio Moretti, Bibiana Santos, João Paulo Kitajima, Erika Freitas, David Schlesinger, Simone Kashima, Evandra Strazza Rodrigues, Svetoslav Nanev Slavov, Elaine Vieira dos Santos, Rafael dos Santos Bezerra, Luiz Carlos Junior de Alcantara, Marta Giovanetti, Vagner Fonseca, Flavia Aburjaile, Rodrigo Tocantins Calado. |
| EPI_ISL_1445228 | UBS IRMA AGUEDA MARIA JAIME | Instituto Butantan / Mendelics | Dimas Tadeu Covas, Sandra Coccuzzo Sampaio, Maria Carolina Elias, José Salvatore Leister Patané, Vincent Louis Viala, Antonio Jorge Martins, Ricardo Haddad, Claudia Renata dos Santos Barros, Elaine Cristina Marqueze, Raul Machado Neto, Debora Botequio Moretti, Bibiana Santos, João Paulo Kitajima, Erika Freitas, David Schlesinger, Simone Kashima, Evandra Strazza Rodrigues, Svetoslav Nanev Slavov, Elaine Vieira dos Santos, Rafael dos Santos Bezerra, Luiz Carlos Junior de Alcantara, Marta Giovanetti, Vagner Fonseca, Flavia Aburjaile, Rodrigo Tocantins Calado. |
| EPI_ISL_1445230, EPI_ISL_1445231, EPI_ISL_1445233, EPI_ISL_1445234, EPI_ISL_1445236, EPI_ISL_1445237 | VIGILANCIA EPIDEMIOLOGICA | Instituto Butantan / Mendelics | Dimas Tadeu Covas, Sandra Coccuzzo Sampaio, Maria Carolina Elias, José Salvatore Leister Patané, Vincent Louis Viala, Antonio Jorge Martins, Ricardo Haddad, Claudia Renata dos Santos Barros, Elaine Cristina Marqueze, Raul Machado Neto, Debora Botequio Moretti, Bibiana Santos, João Paulo Kitajima, Erika Freitas, David Schlesinger, Simone Kashima, Evandra Strazza Rodrigues, Svetoslav Nanev Slavov, Elaine Vieira dos Santos, Rafael dos Santos Bezerra, Luiz Carlos Junior de Alcantara, Marta Giovanetti, Vagner Fonseca, Flavia Aburjaile, Rodrigo Tocantins Calado. |
| EPI_ISL_1445238, EPI_ISL_1445239, EPI_ISL_1445240, EPI_ISL_1445246, EPI_ISL_1445247 | SECAO CENTRO DE DIAGNOSTICO SECEDI | Instituto Butantan / Mendelics | Dimas Tadeu Covas, Sandra Coccuzzo Sampaio, Maria Carolina Elias, José Salvatore Leister Patané, Vincent Louis Viala, Antonio Jorge Martins, Ricardo Haddad, Claudia Renata dos Santos Barros, Elaine Cristina Marqueze, Raul Machado Neto, Debora Botequio Moretti, Bibiana Santos, João Paulo Kitajima, Erika Freitas, David Schlesinger, Simone Kashima, Evandra Strazza Rodrigues, Svetoslav Nanev Slavov, Elaine Vieira dos Santos, Rafael dos Santos Bezerra, Luiz Carlos Junior de Alcantara, Marta Giovanetti, Vagner Fonseca, Flavia Aburjaile, Rodrigo Tocantins Calado. |
| EPI_ISL_1445250, EPI_ISL_1445254, EPI_ISL_1445255, EPI_ISL_1445257, EPI_ISL_1445258, EPI_ISL_1445259, EPI_ISL_1445260, EPI_ISL_1445261, EPI_ISL_1445263 | VIGILANCIA EPIDEMIOLOGICA | Instituto Butantan / Mendelics | Dimas Tadeu Covas, Sandra Coccuzzo Sampaio, Maria Carolina Elias, José Salvatore Leister Patané, Vincent Louis Viala, Antonio Jorge Martins, Ricardo Haddad, Claudia Renata dos Santos Barros, Elaine Cristina Marqueze, Raul Machado Neto, Debora Botequio Moretti, Bibiana Santos, João Paulo Kitajima, Erika Freitas, David Schlesinger, Simone Kashima, Evandra Strazza Rodrigues, Svetoslav Nanev Slavov, Elaine Vieira dos Santos, Rafael dos Santos Bezerra, Luiz Carlos Junior de Alcantara, Marta Giovanetti, Vagner Fonseca, Flavia Aburjaile, Rodrigo Tocantins Calado. |
| EPI_ISL_1445264 | AMBULATORIO MEDICO DE ESPECIALIDADES DE PERUIBE | Instituto Butantan / Mendelics | Dimas Tadeu Covas, Sandra Coccuzzo Sampaio, Maria Carolina Elias, José Salvatore Leister Patané, Vincent Louis Viala, Antonio Jorge Martins, Ricardo Haddad, Claudia Renata dos Santos Barros, Elaine Cristina Marqueze, Raul Machado Neto, Debora Botequio Moretti, Bibiana Santos, João Paulo Kitajima, Erika Freitas, David Schlesinger, Simone Kashima, Evandra Strazza Rodrigues, Svetoslav Nanev Slavov, Elaine Vieira dos Santos, Rafael dos Santos Bezerra, Luiz Carlos Junior de Alcantara, Marta Giovanetti, Vagner Fonseca, Flavia Aburjaile, Rodrigo Tocantins Calado. |
| EPI_ISL_1445265 | VIGILANCIA EPIDEMIOLOGICA | Instituto Butantan / Mendelics | Dimas Tadeu Covas, Sandra Coccuzzo Sampaio, Maria Carolina Elias, José Salvatore Leister Patané, Vincent Louis Viala, Antonio Jorge Martins, Ricardo Haddad, Claudia Renata dos Santos Barros, Elaine Cristina Marqueze, Raul Machado Neto, Debora Botequio Moretti, Bibiana Santos, João Paulo Kitajima, Erika Freitas, David Schlesinger, Simone Kashima, Evandra Strazza Rodrigues, Svetoslav Nanev Slavov, Elaine Vieira dos Santos, Rafael dos Santos Bezerra, Luiz Carlos Junior de Alcantara, Marta Giovanetti, Vagner Fonseca, Flavia Aburjaile, Rodrigo Tocantins Calado. |
| EPI_ISL_1445266, EPI_ISL_1445267 | AMBULATORIO MEDICO DE ESPECIALIDADES DE PERUIBE | Instituto Butantan / Mendelics | Dimas Tadeu Covas, Sandra Coccuzzo Sampaio, Maria Carolina Elias, José Salvatore Leister Patané, Vincent Louis Viala, Antonio Jorge Martins, Ricardo Haddad, Claudia Renata dos Santos Barros, Elaine Cristina Marqueze, Raul Machado Neto, Debora Botequio Moretti, Bibiana Santos, João Paulo Kitajima, Erika Freitas, David Schlesinger, Simone Kashima, Evandra Strazza Rodrigues, Svetoslav Nanev Slavov, Elaine Vieira dos Santos, Rafael dos Santos Bezerra, Luiz Carlos Junior de Alcantara, Marta Giovanetti, Vagner Fonseca, Flavia Aburjaile, Rodrigo Tocantins Calado. |
| EPI_ISL_1445268 | VIGILANCIA EPIDEMIOLOGICA | Instituto Butantan / Mendelics | Dimas Tadeu Covas, Sandra Coccuzzo Sampaio, Maria Carolina Elias, José Salvatore Leister Patané, Vincent Louis Viala, Antonio Jorge Martins, Ricardo Haddad, Claudia Renata dos Santos Barros, Elaine Cristina Marqueze, Raul Machado Neto, Debora Botequio Moretti, Bibiana Santos, João Paulo Kitajima, Erika Freitas, David Schlesinger, Simone Kashima, Evandra Strazza Rodrigues, Svetoslav Nanev Slavov, Elaine Vieira dos Santos, Rafael dos Santos Bezerra, Luiz Carlos Junior de Alcantara, Marta Giovanetti, Vagner Fonseca, Flavia Aburjaile, Rodrigo Tocantins Calado. |
| EPI_ISL_1445269 | SERV DE VIG SANITARIA EPIDEMIO E CTRL DE ZOONOZES GUARUJA | Instituto Butantan / Mendelics | Dimas Tadeu Covas, Sandra Coccuzzo Sampaio, Maria Carolina Elias, José Salvatore Leister Patané, Vincent Louis Viala, Antonio Jorge Martins, Ricardo Haddad, Claudia Renata dos Santos Barros, Elaine Cristina Marqueze, Raul Machado Neto, Debora Botequio Moretti, Bibiana Santos, João Paulo Kitajima, Erika Freitas, David Schlesinger, Simone Kashima, Evandra Strazza Rodrigues, Svetoslav Nanev Slavov, Elaine Vieira dos Santos, Rafael dos Santos Bezerra, Luiz Carlos Junior de Alcantara, Marta Giovanetti, Vagner Fonseca, Flavia Aburjaile, Rodrigo Tocantins Calado. |
| EPI_ISL_1445271 | VIGILANCIA EPIDEMIOLOGICA | Instituto Butantan / Mendelics | Dimas Tadeu Covas, Sandra Coccuzzo Sampaio, Maria Carolina Elias, José Salvatore Leister Patané, Vincent Louis Viala, Antonio Jorge Martins, Ricardo Haddad, Claudia Renata dos Santos Barros, Elaine Cristina Marqueze, Raul Machado Neto, Debora Botequio Moretti, Bibiana Santos, João Paulo Kitajima, Erika Freitas, David Schlesinger, Simone Kashima, Evandra Strazza Rodrigues, Svetoslav Nanev Slavov, Elaine Vieira dos Santos, Rafael dos Santos Bezerra, Luiz Carlos Junior de Alcantara, Marta Giovanetti, Vagner Fonseca, Flavia Aburjaile, Rodrigo Tocantins Calado. |
| EPI_ISL_1445274 | AMBULATORIO MEDICO DE ESPECIALIDADES DE PERUIBE | Instituto Butantan / Mendelics | Dimas Tadeu Covas, Sandra Coccuzzo Sampaio, Maria Carolina Elias, José Salvatore Leister Patané, Vincent Louis Viala, Antonio Jorge Martins, Ricardo Haddad, Claudia Renata dos Santos Barros, Elaine Cristina Marqueze, Raul Machado Neto, Debora Botequio Moretti, Bibiana Santos, João Paulo Kitajima, Erika Freitas, David Schlesinger, Simone Kashima, Evandra Strazza Rodrigues, Svetoslav Nanev Slavov, Elaine Vieira dos Santos, Rafael dos |
| EPI_ISL_1468413, EPI_ISL_1468414, EPI_ISL_1468415 | LACEN do Estado de Goias | Instituto Adolfo Lutz, Interdiciplinary Procedures Center, Strategic Laboratory | Claudio Tavares Sacchi, Claudia Regina Gonçalves, Erica Valessa Ramos Gomes, Karoline Rodrigues Campos, Caio Vinicius Dias Lopes |
| EPI_ISL_1468416 | Secretaria Municipal de Saude de Andradina | Instituto Adolfo Lutz, Interdiciplinary Procedures Center, Strategic Laboratory | Claudio Tavares Sacchi, Claudia Regina Gonçalves, Erica Valessa Ramos Gomes, Karoline Rodrigues Campos, Caio Vinicius Dias Lopes |
| EPI_ISL_1468417 | Santa Casa de Aracatuba Hospital Sagrado Coracao de Jesus | Instituto Adolfo Lutz, Interdiciplinary Procedures Center, Strategic Laboratory | Claudio Tavares Sacchi, Claudia Regina Gonçalves, Erica Valessa Ramos Gomes, Karoline Rodrigues Campos, Caio Vinicius Dias Lopes |
| EPI_ISL_1468418 | Secretaria Municipal de Saude de Birigui | Instituto Adolfo Lutz, Interdiciplinary Procedures Center, Strategic Laboratory | Claudio Tavares Sacchi, Claudia Regina Gonçalves, Erica Valessa Ramos Gomes, Karoline Rodrigues Campos, Caio Vinicius Dias Lopes |
| EPI_ISL_1468419 | Santa Casa de Aracatuba Hospital Sagrado Coracao de Jesus | Instituto Adolfo Lutz, Interdiciplinary Procedures Center, Strategic Laboratory | Claudio Tavares Sacchi, Claudia Regina Gonçalves, Erica Valessa Ramos Gomes, Karoline Rodrigues Campos, Caio Vinicius Dias Lopes |
| EPI_ISL_1468420, EPI_ISL_1468421 | Santa Casa de Birigui | Instituto Adolfo Lutz, Interdiciplinary Procedures Center, Strategic Laboratory | Claudio Tavares Sacchi, Claudia Regina Gonçalves, Erica Valessa Ramos Gomes, Karoline Rodrigues Campos, Caio Vinicius Dias Lopes |
| EPI_ISL_1468424 | Santa Casa de Aracatuba Hospital Sagrado Coracao de Jesus | Instituto Adolfo Lutz, Interdiciplinary Procedures Center, Strategic Laboratory | Claudio Tavares Sacchi, Claudia Regina Gonçalves, Erica Valessa Ramos Gomes, Karoline Rodrigues Campos, Caio Vinicius Dias Lopes |
| EPI_ISL_1468425 | Santa Casa de Birigui | Instituto Adolfo Lutz, Interdiciplinary Procedures Center, Strategic Laboratory | Claudio Tavares Sacchi, Claudia Regina Gonçalves, Erica Valessa Ramos Gomes, Karoline Rodrigues Campos, Caio Vinicius Dias Lopes |
| EPI_ISL_1468426, EPI_ISL_1468427, EPI_ISL_1468429, EPI_ISL_1468430 | Centro de Saude II Matao | Instituto Adolfo Lutz, Interdiciplinary Procedures Center, Strategic Laboratory | Claudio Tavares Sacchi, Claudia Regina Gonçalves, Erica Valessa Ramos Gomes, Karoline Rodrigues Campos, Caio Vinicius Dias Lopes |
| EPI_ISL_1493573, EPI_ISL_1493574, EPI_ISL_1493575, EPI_ISL_1493576, EPI_ISL_1493577 | LACEN do Estado de Goias | Instituto Adolfo Lutz, Interdiciplinary Procedures Center, Strategic Laboratory | Claudio Tavares Sacchi, Claudia Regina Gonçalves, Erica Valessa Ramos Gomes, Karoline Rodrigues Campos, Caio Vinicius Dias Lopes |
| EPI_ISL_1493578, EPI_ISL_1493579 | LACEN do Estado de Rondonia | Instituto Adolfo Lutz, Interdiciplinary Procedures Center, Strategic Laboratory | Claudio Tavares Sacchi, Claudia Regina Gonçalves, Erica Valessa Ramos Gomes, Karoline Rodrigues Campos, Caio Vinicius Dias Lopes |
| EPI_ISL_1493580 | CS II Dr Miguel Vitaliano Orlandia | Instituto Adolfo Lutz, Interdiciplinary Procedures Center, | Claudio Tavares Sacchi, Claudia Regina Gonçalves, Erica Valessa Ramos Gomes, Karoline Rodrigues Campos, Caio Vinicius Dias Lopes |

| | | | |
|---|---|---|---|
| | | Strategic Laboratory | |
| EPI_ISL_1493581 | Sae servico de Atendimento Especializado | Instituto Adolfo Lutz, Interdiciplinary Procedures Center, Strategic Laboratory | Claudio Tavares Sacchi, Claudia Regina Gonçalves, Erica Valessa Ramos Gomes, Karoline Rodrigues Campos, Caio Vinicius Dias Lopes |
| EPI_ISL_1493582 | CS II Dr Miguel Vitaliano Orlandia | Instituto Adolfo Lutz, Interdiciplinary Procedures Center, Strategic Laboratory | Claudio Tavares Sacchi, Claudia Regina Gonçalves, Erica Valessa Ramos Gomes, Karoline Rodrigues Campos, Caio Vinicius Dias Lopes |
| EPI_ISL_1494924 | LACEN do Estado de Rondonia | Instituto Adolfo Lutz, Interdiciplinary Procedures Center, Strategic Laboratory | Claudio Tavares Sacchi, Claudia Regina Gonçalves, Erica Valessa Ramos Gomes, Karoline Rodrigues Campos, Caio Vinicius Dias Lopes |
| EPI_ISL_1494977, EPI_ISL_1494978, EPI_ISL_1494982, EPI_ISL_1494986, EPI_ISL_1494987, EPI_ISL_1494989, EPI_ISL_1494994, EPI_ISL_1494998, EPI_ISL_1495005, EPI_ISL_1495007, EPI_ISL_1495010, EPI_ISL_1495011, EPI_ISL_1495012, EPI_ISL_1495014, EPI_ISL_1495015, EPI_ISL_1495016, EPI_ISL_1495019, EPI_ISL_1495020, EPI_ISL_1495021, EPI_ISL_1495022, EPI_ISL_1495023, EPI_ISL_1495024, EPI_ISL_1495025, EPI_ISL_1495026, EPI_ISL_1495031, EPI_ISL_1495032, EPI_ISL_1495035, EPI_ISL_1495036, EPI_ISL_1495037, EPI_ISL_1495038 | | | |
| see above | Laboratório de Biologia Integrativa | Laboratório de Biologia Integrativa | Filipe Romero Rebello Moreira, Diego Menezes Bonfim, Victor Emmanuel Viana Geddes, Danielle Alves Gomes Zauli, Joice do Prado Silva, Aline Brito de Lima, Frederico Scott Varella Malta, Alessandro Clayton de Souza Ferreira, Victor Cavalcanti Pardini, Daniel Costa Queiroz, Rafael Marques de Souza, Lucyene Miguita Luiz, Paula Luize Camargos Fonseca, Rennan Garcias Moreira, Nuno Rodrigues Faria, Carolina Moreira Voloch, Renan Pedra de Souza, Renato Santana Aguiar |
| EPI_ISL_1498380 | Associação Fundo de Incentivo à Pesquisa (AFIP). | Associação Fundo de Incentivo à Pesquisa (AFIP). | Priscila Farias Tempaku, Juliana Nogueira Martins Rodrigues, Erika Rodrigues de Oliveira, Debora R. Ramadan, Soraya Sgambatti de Andrade, Sergio Tufik. |
| EPI_ISL_1498916 | Associação Fundo de Incentivo à Pesquisa (AFIP) | Associação Fundo de Incentivo à Pesquisa (AFIP) | Priscila Farias Tempaku, Juliana Nogueira Martins Rodrigues, Erika Rodrigues de Oliveira, Debora R. Ramadan, Soraya Sgambatti de Andrade, Sergio Tufik. |
| EPI_ISL_1498917 | Hospital Estadual de Mirandopolis | Instituto Adolfo Lutz, Interdiciplinary Procedures Center, Strategic Laboratory | Claudio Tavares Sacchi, Claudia Regina Gonçalves, Erica Valessa Ramos Gomes, Karoline Rodrigues Campos, Caio Vinicius Dias Lopes |
| EPI_ISL_1499105 | Associação Fundo de Incentivo à Pesquisa (AFIP) | Associação Fundo de Incentivo à Pesquisa (AFIP) | Priscila Farias Tempaku, Juliana Nogueira Martins Rodrigues, Erika Rodrigues de Oliveira, Debora R. Ramadan, Soraya Sgambatti de Andrade, Sergio Tufik. |
| EPI_ISL_1509639 | UBS de Auriflama | Instituto Adolfo Lutz, Interdiciplinary Procedures Center, Strategic Laboratory | Claudio Tavares Sacchi, Claudia Regina Gonçalves, Erica Valessa Ramos Gomes, Karoline Rodrigues Campos, Caio Vinicius Dias Lopes, Leonardo Jose Tadeu de Araujo |
| EPI_ISL_1509720 | UBS de Auriflama | Instituto Adolfo Lutz, Interdiciplinary Procedures Center, Strategic Laboratory | Claudio Tavares Sacchi, Claudia Regina Gonçalves, Erica Valessa Ramos Gomes, Karoline Rodrigues Campos, Caio Vinicius Dias Lopes |
| EPI_ISL_1511643 | Genomic and molecular Biology Group, A.C.Camargo Cancer Center | Laboratory of Bioinformatics and Computational Biology, A.C.Camargo Cancer Center | Giovana Torrezan, Dirce Carraro, Israel Tojal, Alexandre Defelicibus |
| EPI_ISL_1520108, EPI_ISL_1520109 | LACEN do Estado de Rondonia | Instituto Adolfo Lutz, Interdiciplinary Procedures Center, Strategic Laboratory | Claudio Tavares Sacchi, Claudia Regina Gonçalves, Erica Valessa Ramos Gomes, Karoline Rodrigues Campos, Caio Vinicius Dias Lopes |
| EPI_ISL_1520123 | Centro de Saude II Dr Jose de Felipe Espito Santo do Pinhal SP | Instituto Adolfo Lutz, Interdiciplinary Procedures Center, Strategic Laboratory | Claudio Tavares Sacchi, Claudia Regina Gonçalves, Erica Valessa Ramos Gomes, Karoline Rodrigues Campos, Caio Vinicius Dias Lopes |
| EPI_ISL_1520129, EPI_ISL_1520130, EPI_ISL_1520131, EPI_ISL_1520136 | Centro de Saude II Dr Jose Paione Mococa | Instituto Adolfo Lutz, Interdiciplinary Procedures Center, Strategic Laboratory | Claudio Tavares Sacchi, Claudia Regina Gonçalves, Erica Valessa Ramos Gomes, Karoline Rodrigues Campos, Caio Vinicius Dias Lopes |
| EPI_ISL_1533689, EPI_ISL_1533690 | Centro de Saude II Dr. Jose Paione Mococa | Instituto Adolfo Lutz, Interdiciplinary Procedures Center, Strategic Laboratory | Claudio Tavares Sacchi, Claudia Regina Gonçalves, Erica Valessa Ramos Gomes, Karoline Rodrigues Campos, Caio Vinicius Dias Lopes, Leonardo Jose Tadeu de Araujo |
| EPI_ISL_1533695 | Hospital Municipal Dr Mario Gatti | Instituto Adolfo Lutz, Interdiciplinary Procedures Center, Strategic Laboratory | Claudio Tavares Sacchi, Claudia Regina Gonçalves, Erica Valessa Ramos Gomes, Karoline Rodrigues Campos, Caio Vinicius Dias Lopes, Leonardo Jose Tadeu de Araujo |
| EPI_ISL_1533696 | Hospital Municipal de Pedreira | Instituto Adolfo Lutz, Interdiciplinary Procedures Center, Strategic Laboratory | Claudio Tavares Sacchi, Claudia Regina Gonçalves, Erica Valessa Ramos Gomes, Karoline Rodrigues Campos, Caio Vinicius Dias Lopes, Leonardo Jose Tadeu de Araujo |
| EPI_ISL_1533699 | Hospital das Clinicas Luzia de Pinho Melo | Instituto Adolfo Lutz, Interdiciplinary Procedures Center, Strategic Laboratory | Claudio Tavares Sacchi, Claudia Regina Gonçalves, Erica Valessa Ramos Gomes, Karoline Rodrigues Campos, Caio Vinicius Dias Lopes, Leonardo Jose Tadeu de Araujo |
| EPI_ISL_1533701 | Hospital Estadual de Vila Alpina Org Social Seconci Sao Paulo | Instituto Adolfo Lutz, Interdiciplinary Procedures Center, Strategic Laboratory | Claudio Tavares Sacchi, Claudia Regina Gonçalves, Erica Valessa Ramos Gomes, Karoline Rodrigues Campos, Caio Vinicius Dias Lopes, Leonardo Jose Tadeu de Araujo |
| EPI_ISL_1533702 | Hospital Universitario da USP Sao Paulo | Instituto Adolfo Lutz, Interdiciplinary Procedures Center, Strategic Laboratory | Claudio Tavares Sacchi, Claudia Regina Gonçalves, Erica Valessa Ramos Gomes, Karoline Rodrigues Campos, Caio Vinicius Dias Lopes, Leonardo Jose Tadeu de Araujo |
| EPI_ISL_1533703 | Pronto Atendimento Sao Jose | Instituto Adolfo Lutz, Interdiciplinary Procedures Center, Strategic Laboratory | Claudio Tavares Sacchi, Claudia Regina Gonçalves, Erica Valessa Ramos Gomes, Karoline Rodrigues Campos, Caio Vinicius Dias Lopes, Leonardo Jose Tadeu de Araujo |
| EPI_ISL_1533704 | Santa Casa de Atibaia Pro Saude | Instituto Adolfo Lutz, Interdiciplinary Procedures Center, Strategic Laboratory | Claudio Tavares Sacchi, Claudia Regina Gonçalves, Erica Valessa Ramos Gomes, Karoline Rodrigues Campos, Caio Vinicius Dias Lopes, Leonardo Jose Tadeu de Araujo |
| EPI_ISL_1533706 | PS Mun Santana Lauro Ribas Braga | Instituto Adolfo Lutz, Interdiciplinary Procedures Center, Strategic Laboratory | Claudio Tavares Sacchi, Claudia Regina Gonçalves, Erica Valessa Ramos Gomes, Karoline Rodrigues Campos, Caio Vinicius Dias Lopes, Leonardo Jose Tadeu de Araujo |
| EPI_ISL_1533707 | Hosp Mun Planalto Waldomiro de Paula | Instituto Adolfo Lutz, Interdiciplinary Procedures Center, Strategic Laboratory | Claudio Tavares Sacchi, Claudia Regina Gonçalves, Erica Valessa Ramos Gomes, Karoline Rodrigues Campos, Caio Vinicius Dias Lopes, Leonardo Jose Tadeu de Araujo |
| EPI_ISL_1533709, EPI_ISL_1533710 | Hospital Estadual de Vila Alpina Org Social Seconci Sao Paulo | Instituto Adolfo Lutz, Interdiciplinary Procedures Center, Strategic Laboratory | Claudio Tavares Sacchi, Claudia Regina Gonçalves, Erica Valessa Ramos Gomes, Karoline Rodrigues Campos, Caio Vinicius Dias Lopes, Leonardo Jose Tadeu de Araujo |
| EPI_ISL_1533711 | Hospital Santa Clara | Instituto Adolfo Lutz, Interdiciplinary Procedures Center, Strategic Laboratory | Claudio Tavares Sacchi, Claudia Regina Gonçalves, Erica Valessa Ramos Gomes, Karoline Rodrigues Campos, Caio Vinicius Dias Lopes, Leonardo Jose Tadeu de Araujo |
| EPI_ISL_1533712 | Hospital Universitario da USP Sao Paulo | Instituto Adolfo Lutz, Interdiciplinary Procedures Center, Strategic Laboratory | Claudio Tavares Sacchi, Claudia Regina Gonçalves, Erica Valessa Ramos Gomes, Karoline Rodrigues Campos, Caio Vinicius Dias Lopes, Leonardo Jose Tadeu de Araujo |
| EPI_ISL_1533713 | Unidade de Pronto Atendimento Jd Amanda | Instituto Adolfo Lutz, Interdiciplinary Procedures Center, Strategic Laboratory | Claudio Tavares Sacchi, Claudia Regina Gonçalves, Erica Valessa Ramos Gomes, Karoline Rodrigues Campos, Caio Vinicius Dias Lopes, Leonardo Jose Tadeu de Araujo |
| EPI_ISL_1533714 | Pronto Socorro Dr Osmar Mesquita | Instituto Adolfo Lutz, Interdiciplinary Procedures Center, Strategic Laboratory | Claudio Tavares Sacchi, Claudia Regina Gonçalves, Erica Valessa Ramos Gomes, Karoline Rodrigues Campos, Caio Vinicius Dias Lopes, Leonardo Jose Tadeu de Araujo |
| EPI_ISL_1533715 | Santa Casa de Aracatuba Hospital Sagrado Coracao de Jesus | Instituto Adolfo Lutz, Interdiciplinary Procedures Center, Strategic Laboratory | Claudio Tavares Sacchi, Claudia Regina Gonçalves, Erica Valessa Ramos Gomes, Karoline Rodrigues Campos, Caio Vinicius Dias Lopes, Leonardo Jose Tadeu de Araujo |
| EPI_ISL_1533716 | Vigilancia Em Saude | Instituto Adolfo Lutz, Interdiciplinary Procedures Center, Strategic Laboratory | Claudio Tavares Sacchi, Claudia Regina Gonçalves, Erica Valessa Ramos Gomes, Karoline Rodrigues Campos, Caio Vinicius Dias Lopes, Leonardo Jose Tadeu de Araujo |
| EPI_ISL_1533717 | Unidade de Pronto Atendimento Jd Amanda | Instituto Adolfo Lutz, Interdiciplinary Procedures Center, Strategic Laboratory | Claudio Tavares Sacchi, Claudia Regina Gonçalves, Erica Valessa Ramos Gomes, Karoline Rodrigues Campos, Caio Vinicius Dias Lopes, Leonardo Jose Tadeu de Araujo |
| EPI_ISL_1533718 | Servico de Verificacao de Obitos Svo Guarulhos | Instituto Adolfo Lutz, Interdiciplinary Procedures Center, Strategic Laboratory | Claudio Tavares Sacchi, Claudia Regina Gonçalves, Erica Valessa Ramos Gomes, Karoline Rodrigues Campos, Caio Vinicius Dias Lopes, Leonardo Jose Tadeu de Araujo |

| | | | |
|---|---|---|---|
| EPI_ISL_1533719 | Policlinica Munic da Est Turistica de Holambra | Instituto Adolfo Lutz, Interdiciplinary Procedures Center, Strategic Laboratory | Claudio Tavares Sacchi, Claudia Regina Gonçalves, Erica Valessa Ramos Gomes, Karoline Rodrigues Campos, Caio Vinicius Dias Lopes, Leonardo Jose Tadeu de Araujo |
| EPI_ISL_1533720 | Santa Casa de Penapolis | Instituto Adolfo Lutz, Interdiciplinary Procedures Center, Strategic Laboratory | Claudio Tavares Sacchi, Claudia Regina Gonçalves, Erica Valessa Ramos Gomes, Karoline Rodrigues Campos, Caio Vinicius Dias Lopes, Leonardo Jose Tadeu de Araujo |
| EPI_ISL_1533722 | Secretaria Municipal de Saude De Piracaia | Instituto Adolfo Lutz, Interdiciplinary Procedures Center, Strategic Laboratory | Claudio Tavares Sacchi, Claudia Regina Gonçalves, Erica Valessa Ramos Gomes, Karoline Rodrigues Campos, Caio Vinicius Dias Lopes, Leonardo Jose Tadeu de Araujo |
| EPI_ISL_1533723 | Hospital Geral de Itaquaquecetuba | Instituto Adolfo Lutz, Interdiciplinary Procedures Center, Strategic Laboratory | Claudio Tavares Sacchi, Claudia Regina Gonçalves, Erica Valessa Ramos Gomes, Karoline Rodrigues Campos, Caio Vinicius Dias Lopes, Leonardo Jose Tadeu de Araujo |
| EPI_ISL_1533725 | Hospital Geral de Guarulhos | Instituto Adolfo Lutz, Interdiciplinary Procedures Center, Strategic Laboratory | Claudio Tavares Sacchi, Claudia Regina Gonçalves, Erica Valessa Ramos Gomes, Karoline Rodrigues Campos, Caio Vinicius Dias Lopes, Leonardo Jose Tadeu de Araujo |
| EPI_ISL_1533726 | Hospital Estadual de Campanha Covid 19 Barradas | Instituto Adolfo Lutz, Interdiciplinary Procedures Center, Strategic Laboratory | Claudio Tavares Sacchi, Claudia Regina Gonçalves, Erica Valessa Ramos Gomes, Karoline Rodrigues Campos, Caio Vinicius Dias Lopes, Leonardo Jose Tadeu de Araujo |
| EPI_ISL_1533727 | Hospital Universitario da USP Sao Paulo | Instituto Adolfo Lutz, Interdiciplinary Procedures Center, Strategic Laboratory | Claudio Tavares Sacchi, Claudia Regina Gonçalves, Erica Valessa Ramos Gomes, Karoline Rodrigues Campos, Caio Vinicius Dias Lopes, Leonardo Jose Tadeu de Araujo |
| EPI_ISL_1578453 | Laboratório de Biologia Molecular do Hospital das Clínicas da Faculdade de Medicina de Botucatu/SP | Laboratórios Genômica Funcional (FCA/UNESP) e Biologia Molecular (FMB-HC/UNESP) - Rede de Vigilância Genômica (Vigenômica)/UNESP | Patrícia Akemi Assato; Felipe Allan da Silva da Costa; Bianca Cechetto Carlos; Flavia Hebner Barbosa Trovão; Guilherme Targino Valente; Rejane Maria Tommasini Grotto; Jayme A. Souza-Neto. |
| EPI_ISL_1578740, EPI_ISL_1579258, EPI_ISL_1580502 | Laboratório de Biologia Molecular do Hospital das Clínicas da Faculdade de Medicina de Botucatu/SP | Laboratórios de Genômica Funcional (FCA/UNESP) e Biologia Molecular (FMB-HC/UNESP) - Rede de Vigilância Genômica (Vigenômica)/UNESP | Patrícia Akemi Assato; Felipe Allan da Silva da Costa; Bianca Cechetto Carlos; Flavia Hebner Barbosa Trovão; Guilherme Targino Valente; Rejane Maria Tommasini Grotto; Jayme A. Souza-Neto. |
| EPI_ISL_1583650, EPI_ISL_1583656, EPI_ISL_1583663, EPI_ISL_1583665, EPI_ISL_1583672, EPI_ISL_1583673, EPI_ISL_1583674 | Central Public Health Laboratory - LACEN -Bahia, Salvador, Brazil | Central Public Health Laboratory - LACEN -Bahia, Salvador, Brazil | Stephane Tosta, Luciana Oliveira, Vanessa Nardy,Patrícia Cajado,Marcela Gómez, Breno Dominguez, Jaqueline Gomes, Vagner Fonseca,Marta Giovanetti,Luiz Alcantara, Felicidade Pereira, Arabela Leal |
| EPI_ISL_1583675 | DNA Laboratory | Central Public Health Laboratory - LACEN -Bahia, Salvador, Brazil | Stephane Tosta, Luciana Oliveira, Vanessa Nardy,Patrícia Cajado,Marcela Gómez, Breno Dominguez, Jaqueline Gomes, Vagner Fonseca,Marta Giovanetti,Luiz Alcantara, Felicidade Pereira, Arabela Leal |
| EPI_ISL_1583676, EPI_ISL_1583677, EPI_ISL_1583679, EPI_ISL_1583680, EPI_ISL_1583681, EPI_ISL_1583682, EPI_ISL_1583683, EPI_ISL_1583694, EPI_ISL_1583697, EPI_ISL_1583700, EPI_ISL_1583716, EPI_ISL_1583719, EPI_ISL_1583722, EPI_ISL_1583725, EPI_ISL_1583730, EPI_ISL_1583733 | | | |
| see above | Central Public Health Laboratory - LACEN -Bahia, Salvador, Brazil | Central Public Health Laboratory - LACEN -Bahia, Salvador, Brazil | Stephane Tosta, Luciana Oliveira, Vanessa Nardy,Patrícia Cajado,Marcela Gómez, Breno Dominguez, Jaqueline Gomes, Vagner Fonseca,Marta Giovanetti,Luiz Alcantara, Felicidade Pereira, Arabela Leal |
| EPI_ISL_1592227, EPI_ISL_1592229, EPI_ISL_1592230 | Laboratório Hermes Pardini | Laboratório de Biologia Integrativa, Instituto de Ciências Biológicas, Universidade Federal de Minas Gerais | Filipe Romero Rebello Moreira, Diego Menezes Bonfim, Danielle Alves Gomes Zauli, Joice do Prado Silva, Aline Brito de Lima, Frederico Scott Varella Malta, Alessandro Clayton de Souza Ferreira, Victor Cavalcanti Pardini, Daniel Costa Queiroz, Rafael Marques de Souza, Victor Emmanuel Viana Geddes, Walyson Coelho Costa, Wagner Carlos Santos Magalhaes, Rennan Garcias Moreira, Carolina Moreira Voloch, Renan Pedra de Souza, Renato Santana Aguiar. |
| EPI_ISL_1608161, EPI_ISL_1608162, EPI_ISL_1608163, EPI_ISL_1608164, EPI_ISL_1608165, EPI_ISL_1608166, EPI_ISL_1608167, EPI_ISL_1608168, EPI_ISL_1608169, EPI_ISL_1608170, EPI_ISL_1608171 | | | |
| see above | Hospital São Rafael - IDOR | Hospital São Rafael - IDOR | Carolina Kymie Vasques Nonaka, Tiago Gräf, Camila Araújo de Lorenzo Barcia, Vanessa Ferreira Costa, Janderson Lopes de Oliveira, Rogério da Hora Passos, Clarissa Araújo Gurgel Rocha, Iasmin Nogueira Bastos, Maria Clara Brito de Santana, Ian Marinho Santos, Karoline Almeida Felix de Sousa, Isadora Cristina de Siqueira, Thamires Gomes Lopes Weber, Ana Verena Almeida Mendes, Bruno Solano de Freitas Souza |
| EPI_ISL_1620638 | Hospital Municipal Dr. Ignacio Proença de Gouvea | Instituto Butantan / Mendelics | Alexander Roberto Precioso, Dimas Tadeu Covas, Sandra Coccuzzo Sampaio, Maria Carolina Elias, José Salvatore Leister Patané, Vincent Louis Viala, Antonio Jorge Martins, Ricardo Haddad, Claudia Renata dos Santos Barros, Elaine Cristina Marqueze, Raul Machado Neto, Debora Botequio Moretti, Bibiana Santos, João Paulo Kitajima, Erika Freitas, David Schlesinger, Simone Kashima, Evandra Strazza Rodrigues, Svetoslav Nanev Slavov, Elaine Vieira dos Santos, Rafael dos Santos Bezerra, Luiz Carlos Junior de Alcantara, Marta Giovanetti, Vagner Fonseca, Flavia Aburjaile, Rodrigo Tocantins Calado. |
| EPI_ISL_1620639 | Fleury | Instituto Butantan / Mendelics | Alexander Roberto Precioso, Dimas Tadeu Covas, Sandra Coccuzzo Sampaio, Maria Carolina Elias, José Salvatore Leister Patané, Vincent Louis Viala, Antonio Jorge Martins, Ricardo Haddad, Claudia Renata dos Santos Barros, Elaine Cristina Marqueze, Raul Machado Neto, Debora Botequio Moretti, Bibiana Santos, João Paulo Kitajima, Erika Freitas, David Schlesinger, Simone Kashima, Evandra Strazza Rodrigues, Svetoslav Nanev Slavov, Elaine Vieira dos Santos, Rafael dos Santos Bezerra, Luiz Carlos Junior de Alcantara, Marta Giovanetti, Vagner Fonseca, Flavia Aburjaile, Rodrigo Tocantins Calado. |
| EPI_ISL_1625972 | Ama J Angela | Instituto Adolfo Lutz, Interdiciplinary Procedures Center, Strategic Laboratory | Claudio Tavares Sacchi, Claudia Regina Gonçalves, Erica Valessa Ramos Gomes, Karoline Rodrigues Campos, Caio Vinicius Dias Lopes, Leonardo Jose Tadeu de Araujo, Katia Correa de Oliveira Santos |
| EPI_ISL_1625974 | Santa Casa de Aracatuba Hospital Sagrado Coracao de Jesus | Instituto Adolfo Lutz, Interdiciplinary Procedures Center, Strategic Laboratory | Claudio Tavares Sacchi, Claudia Regina Gonçalves, Erica Valessa Ramos Gomes, Karoline Rodrigues Campos, Caio Vinicius Dias Lopes, Leonardo Jose Tadeu de Araujo, Katia Correa de Oliveira Santos |
| EPI_ISL_1625975 | CS II Dr Antonio Vicoso Moreira De Rezende Sumare | Instituto Adolfo Lutz, Interdiciplinary Procedures Center, Strategic Laboratory | Claudio Tavares Sacchi, Claudia Regina Gonçalves, Erica Valessa Ramos Gomes, Karoline Rodrigues Campos, Caio Vinicius Dias Lopes, Leonardo Jose Tadeu de Araujo, Katia Correa de Oliveira Santos |
| EPI_ISL_1625976 | Centro de Saude III de Divinolandia | Instituto Adolfo Lutz, Interdiciplinary Procedures Center, Strategic Laboratory | Claudio Tavares Sacchi, Claudia Regina Gonçalves, Erica Valessa Ramos Gomes, Karoline Rodrigues Campos, Caio Vinicius Dias Lopes, Leonardo Jose Tadeu de Araujo, Katia Correa de Oliveira Santos |
| EPI_ISL_1625978, EPI_ISL_1625984, EPI_ISL_1625986, EPI_ISL_1625987, EPI_ISL_1625988, EPI_ISL_1625989, EPI_ISL_1625990, EPI_ISL_1625991, EPI_ISL_1625992, EPI_ISL_1625993, EPI_ISL_1625994, EPI_ISL_1625995, EPI_ISL_1625997, EPI_ISL_1625998, EPI_ISL_1625999, EPI_ISL_1626001, EPI_ISL_1626003, EPI_ISL_1626004, EPI_ISL_1626006, EPI_ISL_1626606 | | | |
| see above | Instituto Adolfo Lutz - Regional de Rio Claro | Instituto Adolfo Lutz, Interdiciplinary Procedures Center, Strategic Laboratory | Claudio Tavares Sacchi, Claudia Regina Gonçalves, Erica Valessa Ramos Gomes, Karoline Rodrigues Campos, Caio Vinicius Dias Lopes, Leonardo Jose Tadeu de Araujo, Katia Correa de Oliveira Santos |
| EPI_ISL_1628344 | Hospital Geral de Pedreira | Instituto Adolfo Lutz, Interdiciplinary Procedures Center, Strategic Laboratory | Claudio Tavares Sacchi, Claudia Regina Gonçalves, Erica Valessa Ramos Gomes, Karoline Rodrigues Campos, Caio Vinicius Dias Lopes, Leonardo Jose Tadeu de Araujo, Katia Correa de Oliveira Santos |
| EPI_ISL_1628347, EPI_ISL_1628348, EPI_ISL_1628349, EPI_ISL_1628350, EPI_ISL_1628351, EPI_ISL_1628352, EPI_ISL_1628353, EPI_ISL_1628354, EPI_ISL_1628355, EPI_ISL_1628356, EPI_ISL_1628357, EPI_ISL_1628358, EPI_ISL_1628359, EPI_ISL_1628360, EPI_ISL_1628361, EPI_ISL_1628362 | | | |
| see above | Instituto Adolfo Lutz Central | Instituto Adolfo Lutz, Interdiciplinary Procedures Center, Strategic Laboratory | Claudio Tavares Sacchi, Claudia Regina Gonçalves, Erica Valessa Ramos Gomes, Karoline Rodrigues Campos, Caio Vinicius Dias Lopes, Leonardo Jose Tadeu de Araujo, Katia Correa de Oliveira Santos |
| EPI_ISL_1628363, EPI_ISL_1628364, EPI_ISL_1628365 | LACEN do Estado de Goias | Instituto Adolfo Lutz, Interdiciplinary Procedures Center, Strategic Laboratory | Claudio Tavares Sacchi, Claudia Regina Gonçalves, Erica Valessa Ramos Gomes, Karoline Rodrigues Campos, Caio Vinicius Dias Lopes, Leonardo Jose Tadeu de Araujo, Katia Correa de Oliveira Santos |
| EPI_ISL_1628366 | CS III de Patrocinio Paulista | Instituto Adolfo Lutz, Interdiciplinary Procedures Center, Strategic Laboratory | Claudio Tavares Sacchi, Claudia Regina Gonçalves, Erica Valessa Ramos Gomes, Karoline Rodrigues Campos, Caio Vinicius Dias Lopes, Leonardo Jose Tadeu de Araujo, Katia Correa de Oliveira Santos |
| EPI_ISL_1628367 | Santa Casa de Guaira | Instituto Adolfo Lutz, Interdiciplinary Procedures Center, Strategic Laboratory | Claudio Tavares Sacchi, Claudia Regina Gonçalves, Erica Valessa Ramos Gomes, Karoline Rodrigues Campos, Caio Vinicius Dias Lopes, Leonardo Jose Tadeu de Araujo, Katia Correa de Oliveira Santos |
| EPI_ISL_1628368 | Centro Medico Social Comunitario Januario Teodoro de Souza | Instituto Adolfo Lutz, Interdiciplinary Procedures Center, Strategic Laboratory | Claudio Tavares Sacchi, Claudia Regina Gonçalves, Erica Valessa Ramos Gomes, Karoline Rodrigues Campos, Caio Vinicius Dias Lopes, Leonardo Jose Tadeu de Araujo, Katia Correa de Oliveira Santos |

| | | | |
|---|---|---|---|
| EPI_ISL_1628369 | UPA Dr Luis Atilio Losi Viana Ribeirao Preto | Instituto Adolfo Lutz, Interdiciplinary Procedures Center, Strategic Laboratory | Claudio Tavares Sacchi, Claudia Regina Gonçalves, Erica Valessa Ramos Gomes, Karoline Rodrigues Campos, Caio Vinicius Dias Lopes, Leonardo Jose Tadeu de Araujo, Katia Correa de Oliveira Santos |
| EPI_ISL_1628370 | Sae Servico De Atendimento Especializado | Instituto Adolfo Lutz, Interdiciplinary Procedures Center, Strategic Laboratory | Claudio Tavares Sacchi, Claudia Regina Gonçalves, Erica Valessa Ramos Gomes, Karoline Rodrigues Campos, Caio Vinicius Dias Lopes, Leonardo Jose Tadeu de Araujo, Katia Correa de Oliveira Santos |
| EPI_ISL_1628372 | Hospital Sao Marcos da Sama Morro Agudo | Instituto Adolfo Lutz, Interdiciplinary Procedures Center, Strategic Laboratory | Claudio Tavares Sacchi, Claudia Regina Gonçalves, Erica Valessa Ramos Gomes, Karoline Rodrigues Campos, Caio Vinicius Dias Lopes, Leonardo Jose Tadeu de Araujo, Katia Correa de Oliveira Santos |
| EPI_ISL_1628373 | UPA De Bebedouro | Instituto Adolfo Lutz, Interdiciplinary Procedures Center, Strategic Laboratory | Claudio Tavares Sacchi, Claudia Regina Gonçalves, Erica Valessa Ramos Gomes, Karoline Rodrigues Campos, Caio Vinicius Dias Lopes, Leonardo Jose Tadeu de Araujo, Katia Correa de Oliveira Santos |
| EPI_ISL_1628374 | Santa Casa De Cravinhos | Instituto Adolfo Lutz, Interdiciplinary Procedures Center, Strategic Laboratory | Claudio Tavares Sacchi, Claudia Regina Gonçalves, Erica Valessa Ramos Gomes, Karoline Rodrigues Campos, Caio Vinicius Dias Lopes, Leonardo Jose Tadeu de Araujo, Katia Correa de Oliveira Santos |
| EPI_ISL_1628375, EPI_ISL_1628376 | Secretaria Municipal de Saude de Guariba | Instituto Adolfo Lutz, Interdiciplinary Procedures Center, Strategic Laboratory | Claudio Tavares Sacchi, Claudia Regina Gonçalves, Erica Valessa Ramos Gomes, Karoline Rodrigues Campos, Caio Vinicius Dias Lopes, Leonardo Jose Tadeu de Araujo, Katia Correa de Oliveira Santos |
| EPI_ISL_1628377 | Centro De Saude II Ibitinga | Instituto Adolfo Lutz, Interdiciplinary Procedures Center, Strategic Laboratory | Claudio Tavares Sacchi, Claudia Regina Gonçalves, Erica Valessa Ramos Gomes, Karoline Rodrigues Campos, Caio Vinicius Dias Lopes, Leonardo Jose Tadeu de Araujo, Katia Correa de Oliveira Santos |
| EPI_ISL_1628378 | Vigilancia Epidemiologica Jardinopolis | Instituto Adolfo Lutz, Interdiciplinary Procedures Center, Strategic Laboratory | Claudio Tavares Sacchi, Claudia Regina Gonçalves, Erica Valessa Ramos Gomes, Karoline Rodrigues Campos, Caio Vinicius Dias Lopes, Leonardo Jose Tadeu de Araujo, Katia Correa de Oliveira Santos |
| EPI_ISL_1628379 | UPA Dr Luis Atilio Losi Viana Ribeirao Preto | Instituto Adolfo Lutz, Interdiciplinary Procedures Center, Strategic Laboratory | Claudio Tavares Sacchi, Claudia Regina Gonçalves, Erica Valessa Ramos Gomes, Karoline Rodrigues Campos, Caio Vinicius Dias Lopes, Leonardo Jose Tadeu de Araujo, Katia Correa de Oliveira Santos |
| EPI_ISL_1628380 | Santa Casa De Pitangueiras | Instituto Adolfo Lutz, Interdiciplinary Procedures Center, Strategic Laboratory | Claudio Tavares Sacchi, Claudia Regina Gonçalves, Erica Valessa Ramos Gomes, Karoline Rodrigues Campos, Caio Vinicius Dias Lopes, Leonardo Jose Tadeu de Araujo, Katia Correa de Oliveira Santos |
| EPI_ISL_1630158 | Laboratório de Microbiologia Molecular - Universidade FEEVALE | Laboratório de Microbiologia Molecular - Universidade FEEVALE | Alana Witt Hansen, Fágner Henrique Heldt, Fernando Rosado Spilki, Flávio Silveira, Juliana Schons Gularte, Juliane Deise Fleck, Mariana Soares da Silva, Meriane Demoliner, Matheus Nunes Weber, Paula Rodrigues de Almeida, Micheli Filippi. |
| EPI_ISL_1652237 | Fleury | Instituto Butantan / Mendelics | Alexander Roberto Precioso, Dimas Tadeu Covas, Sandra Coccuzzo Sampaio, Maria Carolina Elias, José Salvatore Leister Patané, Vincent Louis Viala, Antonio Jorge Martins, Ricardo Haddad, Claudia Renata dos Santos Barros, Elaine Cristina Marqueze, Raul Machado Neto, Debora Botequio Moretti, Bibiana Santos, João Paulo Kitajima, Erika Freitas, David Schlesinger, Simone Kashima, Evandra Strazza Rodrigues, Svetoslav Nanev Slavov, Elaine Vieira dos Santos, Rafael dos Santos Bezerra, Luiz Carlos Junior de Alcantara, Marta Giovanetti, Vagner Fonseca, Flavia Aburjaile, Rodrigo Tocantins Calado. |
| EPI_ISL_1664113, EPI_ISL_1664114, EPI_ISL_1664115, EPI_ISL_1664116, EPI_ISL_1664117, EPI_ISL_1664118, EPI_ISL_1664119, EPI_ISL_1664120, EPI_ISL_1664121, EPI_ISL_1664122, EPI_ISL_1664123, EPI_ISL_1664124, EPI_ISL_1664125, EPI_ISL_1664127, EPI_ISL_1664128, EPI_ISL_1664129, EPI_ISL_1664130, EPI_ISL_1664131, EPI_ISL_1664132, EPI_ISL_1664133, EPI_ISL_1664134, EPI_ISL_1664135, EPI_ISL_1664136, EPI_ISL_1664137, EPI_ISL_1664139, EPI_ISL_1664140, EPI_ISL_1664141, EPI_ISL_1664142, EPI_ISL_1664143, EPI_ISL_1664144, EPI_ISL_1664145, EPI_ISL_1664146, EPI_ISL_1664147, EPI_ISL_1664148, EPI_ISL_1664149, EPI_ISL_1664150, EPI_ISL_1664151, EPI_ISL_1664152, EPI_ISL_1664153, EPI_ISL_1664154, EPI_ISL_1664155, EPI_ISL_1664156, EPI_ISL_1664157, EPI_ISL_1664158, EPI_ISL_1664159, EPI_ISL_1664160, EPI_ISL_1664161, EPI_ISL_1664162, EPI_ISL_1664163, EPI_ISL_1664164, EPI_ISL_1664165, EPI_ISL_1664166, EPI_ISL_1664167, EPI_ISL_1664168, EPI_ISL_1664169, EPI_ISL_1664170, EPI_ISL_1664171, EPI_ISL_1664172, EPI_ISL_1664173, EPI_ISL_1664174, EPI_ISL_1664176, EPI_ISL_1664177, EPI_ISL_1664178, EPI_ISL_1664179, EPI_ISL_1664180, EPI_ISL_1664181, EPI_ISL_1664182, EPI_ISL_1664183, EPI_ISL_1664186, EPI_ISL_1664187, EPI_ISL_1664188, EPI_ISL_1664189, EPI_ISL_1664190, EPI_ISL_1664191, EPI_ISL_1664192, EPI_ISL_1664193, EPI_ISL_1664194, EPI_ISL_1664195, EPI_ISL_1664196, EPI_ISL_1664197, EPI_ISL_1664198, EPI_ISL_1664199, EPI_ISL_1664200, EPI_ISL_1664201, EPI_ISL_1664202 | | | |
| see above | Laboratorio Central Noel Nutels | Bioinformatics Laboratory / LNCC | Luiz G P de Almeida, Alessandra P Lamarca, Ronaldo da Silva F Jr, Liliane Cavalcante, Alexandra L Gerber, Ana Paula de C Guimarães, Douglas Terra Machado, Cassia Alves, Diana Mariani, Thais Felix Cruz, Mario Sergio Ribeiro, Silvia Carvalho, Flávio Dias da Silva, Marcio Henrique de Oliveira Garcia, Leandro Magalhães de Souza, Cristiane Gomes da Silva, Caio Luiz Pereira Ribeiro, Andréa Cony Cavalcanti, Claudia Maria Braga de Mello, Amilcar Tanuri, Ana Tereza R Vasconcelos |
| EPI_ISL_1708318 | Fundação Hospitalar de Hematologia e Hemoterapia do Amazonas | Laboratório de Estudos de Vírus Emergentes | William M. Souza; Mariene R. Amorim, Renata Sesti-Costa, Lais D. Coimbra, Daniel A. Toledo-Teixeira, Pierina L. Parise, Priscilla P. Barbosa, Karina Bispo-dos-Santos, Luciana S. Mofatto, Camila L. Simeoni, Natalia S. Brunetti, Ingra M. Claro, Adriana S. S. Duarte, Thais M. Coletti, Audrey B. Zangirolami, Carolina Costa-Lima, Arilson Bernardo S. P. Gomes, Lucas I. Buscaratti, Flavia C. Sales, Vitor A. Costa, Lucas A.M. Franco, Darlan S. Candido, Oliver G. Pybus, Jaqueline G. de Jesus, Camila A.M. Silva, Mariana S. Ramundo, Giulia M. Ferreira, Mariana C. Pinho, Leandro M. Souza, Esmenia C. Rocha, Pamela S. Andrade, Myuki A.E. Crispim, Grazielle C. Maktura, Erika R. Manuli, Magnun N.N. Santos, Cecilia C. Camilo, Rodrigo N. Angerami, Maria L. Moretti, Fernando R. Spilki, Clarice W. Arns, Marcelo Addas-Carvalho, Bruno D. Benites, Marcelo A.S. Mori, Nelson Gaburo, Christopher Dye, Chieh-Hsi Wu, Henrique Marques-Souza, Rafael E. Marques, Alessandro S. Farias, Michael S. Diamond, Nuno R. Faria, Ester C. Sabino, Fabia |
| EPI_ISL_1715134 | Centro de Saude II Ibitinga | Instituto Adolfo Lutz, Interdiciplinary Procedures Center, Strategic Laboratory | Claudio Tavares Sacchi, Claudia Regina Gonçalves, Erica Valessa Ramos Gomes, Karoline Rodrigues Campos, Caio Vinicius Dias Lopes, Leonardo Jose Tadeu de Araujo, Katia Correa de Oliveira Santos |
| EPI_ISL_1715135, EPI_ISL_1715136 | CS II Dr Jose Ferreira Telles | Instituto Adolfo Lutz, Interdiciplinary Procedures Center, Strategic Laboratory | Claudio Tavares Sacchi, Claudia Regina Gonçalves, Erica Valessa Ramos Gomes, Karoline Rodrigues Campos, Caio Vinicius Dias Lopes, Leonardo Jose Tadeu de Araujo, Katia Correa de Oliveira Santos |
| EPI_ISL_1715137 | Secretaria Municipal De Saude | Instituto Adolfo Lutz, Interdiciplinary Procedures Center, Strategic Laboratory | Claudio Tavares Sacchi, Claudia Regina Gonçalves, Erica Valessa Ramos Gomes, Karoline Rodrigues Campos, Caio Vinicius Dias Lopes, Leonardo Jose Tadeu de Araujo, Katia Correa de Oliveira Santos |
| EPI_ISL_1715138 | Santa Casa De Cravinhos | Instituto Adolfo Lutz, Interdiciplinary Procedures Center, Strategic Laboratory | Claudio Tavares Sacchi, Claudia Regina Gonçalves, Erica Valessa Ramos Gomes, Karoline Rodrigues Campos, Caio Vinicius Dias Lopes, Leonardo Jose Tadeu de Araujo, Katia Correa de Oliveira Santos |
| EPI_ISL_1715139 | Sae Servico De Atendimento Especializado | Instituto Adolfo Lutz, Interdiciplinary Procedures Center, Strategic Laboratory | Claudio Tavares Sacchi, Claudia Regina Gonçalves, Erica Valessa Ramos Gomes, Karoline Rodrigues Campos, Caio Vinicius Dias Lopes, Leonardo Jose Tadeu de Araujo, Katia Correa de Oliveira Santos |
| EPI_ISL_1715140 | UPA Dr Luis Atilio Losi Viana Ribeirao Preto | Instituto Adolfo Lutz, Interdiciplinary Procedures Center, Strategic Laboratory | Claudio Tavares Sacchi, Claudia Regina Gonçalves, Erica Valessa Ramos Gomes, Karoline Rodrigues Campos, Caio Vinicius Dias Lopes, Leonardo Jose Tadeu de Araujo, Katia Correa de Oliveira Santos |
| EPI_ISL_1715141 | Centro De Saude II Ibitinga | Instituto Adolfo Lutz, Interdiciplinary Procedures Center, Strategic Laboratory | Claudio Tavares Sacchi, Claudia Regina Gonçalves, Erica Valessa Ramos Gomes, Karoline Rodrigues Campos, Caio Vinicius Dias Lopes, Leonardo Jose Tadeu de Araujo, Katia Correa de Oliveira Santos |
| EPI_ISL_1715142 | CS III de Patrocinio Paulista | Instituto Adolfo Lutz, Interdiciplinary Procedures Center, Strategic Laboratory | Claudio Tavares Sacchi, Claudia Regina Gonçalves, Erica Valessa Ramos Gomes, Karoline Rodrigues Campos, Caio Vinicius Dias Lopes, Leonardo Jose Tadeu de Araujo, Katia Correa de Oliveira Santos |
| EPI_ISL_1715143 | Secretaria Municipal de Saude De Guariba | Instituto Adolfo Lutz, Interdiciplinary Procedures Center, Strategic Laboratory | Claudio Tavares Sacchi, Claudia Regina Gonçalves, Erica Valessa Ramos Gomes, Karoline Rodrigues Campos, Caio Vinicius Dias Lopes, Leonardo Jose Tadeu de Araujo, Katia Correa de Oliveira Santos |
| EPI_ISL_1715144 | UBDS Dr Joao Baptista Quartim Central | Instituto Adolfo Lutz, Interdiciplinary Procedures Center, Strategic Laboratory | Claudio Tavares Sacchi, Claudia Regina Gonçalves, Erica Valessa Ramos Gomes, Karoline Rodrigues Campos, Caio Vinicius Dias Lopes, Leonardo Jose Tadeu de Araujo, Katia Correa de Oliveira Santos |
| EPI_ISL_1715145 | UPA de Bebedouro | Instituto Adolfo Lutz, Interdiciplinary Procedures Center, Strategic Laboratory | Claudio Tavares Sacchi, Claudia Regina Gonçalves, Erica Valessa Ramos Gomes, Karoline Rodrigues Campos, Caio Vinicius Dias Lopes, Leonardo Jose Tadeu de Araujo, Katia Correa de Oliveira Santos |
| EPI_ISL_1715146, EPI_ISL_1715147 | Instituto Adolfo Lutz - Regional de Aracatuba | Instituto Adolfo Lutz, Interdiciplinary Procedures Center, Strategic Laboratory | Claudio Tavares Sacchi, Claudia Regina Gonçalves, Erica Valessa Ramos Gomes, Karoline Rodrigues Campos, Caio Vinicius Dias Lopes, Leonardo Jose Tadeu de Araujo, Katia Correa de Oliveira Santos |
| EPI_ISL_1715148 | Instituto Adolfo Lutz - Regional de Ribeirao Preto | Instituto Adolfo Lutz, Interdiciplinary Procedures Center, Strategic Laboratory | Claudio Tavares Sacchi, Claudia Regina Gonçalves, Erica Valessa Ramos Gomes, Karoline Rodrigues Campos, Caio Vinicius Dias Lopes, Leonardo Jose Tadeu de Araujo, Katia Correa de Oliveira Santos |
| EPI_ISL_1715149, EPI_ISL_1715150, EPI_ISL_1715151, EPI_ISL_1715152, EPI_ISL_1715153, EPI_ISL_1715154, EPI_ISL_1715155, EPI_ISL_1715156, EPI_ISL_1715157, EPI_ISL_1715158, EPI_ISL_1715159 | | | |
| see above | Instituto Adolfo Lutz Central | Instituto Adolfo Lutz, Interdiciplinary Procedures Center, Strategic Laboratory | Claudio Tavares Sacchi, Claudia Regina Gonçalves, Erica Valessa Ramos Gomes, Karoline Rodrigues Campos, Caio Vinicius Dias Lopes, Leonardo Jose Tadeu de Araujo, Katia Correa de Oliveira Santos |

| | | | |
|---|---|---|---|
| EPI_ISL_1716397, EPI_ISL_1716398, EPI_ISL_1716400, EPI_ISL_1716401, EPI_ISL_1716402, EPI_ISL_1716403, EPI_ISL_1716404, EPI_ISL_1716405, EPI_ISL_1716406, EPI_ISL_1716407, EPI_ISL_1716408, EPI_ISL_1716409, EPI_ISL_1716410, EPI_ISL_1716411, EPI_ISL_1716412, EPI_ISL_1716413, EPI_ISL_1716414, EPI_ISL_1716416, EPI_ISL_1716417, EPI_ISL_1716418, EPI_ISL_1716419, EPI_ISL_1716420, EPI_ISL_1716421, EPI_ISL_1716422, EPI_ISL_1716423, EPI_ISL_1716424, EPI_ISL_1716425, EPI_ISL_1716427, EPI_ISL_1716428, EPI_ISL_1716429, EPI_ISL_1716430, EPI_ISL_1716431, EPI_ISL_1716432, EPI_ISL_1716433, EPI_ISL_1716434, EPI_ISL_1716435, EPI_ISL_1716436, EPI_ISL_1716437, EPI_ISL_1716438, EPI_ISL_1716439, EPI_ISL_1716440, EPI_ISL_1716441, EPI_ISL_1716442, EPI_ISL_1716443, EPI_ISL_1716444, EPI_ISL_1716445, EPI_ISL_1716446, EPI_ISL_1716447, EPI_ISL_1716448, EPI_ISL_1716449, EPI_ISL_1716450, EPI_ISL_1716451, EPI_ISL_1716452, EPI_ISL_1716453, EPI_ISL_1716454, EPI_ISL_1716455, EPI_ISL_1716456 | | | |
| see above | Laboratorio Saude | LGBio (Laboratorio de Genetica & Biodiversidade) | Mariana Pires de Campos Telles, Daniela de Melo e Silva, Elisangela de Paula Silveira Lacerda, Renata de Oliveira Dias, Rhewter Nunes, Cintia Pelegrineti Targueta de Azevedo Brito, Ramilla dos Santos Braga, Thais Guimarães Castro, Thays Millena Alves Pedroso, Amanda Alves de Melo, Francyelli Mello Andrade |
| EPI_ISL_1716457, EPI_ISL_1716458, EPI_ISL_1716459, EPI_ISL_1716460 | LACEN (Laboratorio de Saude Publica Dr. Giovanni Cysneiros) | LGBio (Laboratorio de Genetica & Biodiversidade) | Mariana Pires de Campos Telles, Daniela de Melo e Silva, Elisangela de Paula Silveira Lacerda, Renata de Oliveira Dias, Rhewter Nunes, Cintia Pelegrineti Targueta de Azevedo Brito, Ramilla dos Santos Braga, Thais Guimarães Castro, Thays Millena Alves Pedroso, Amanda Alves de Melo, Francyelli Mello Andrade |
| EPI_ISL_1716461, EPI_ISL_1716462, EPI_ISL_1716463, EPI_ISL_1716464, EPI_ISL_1716465, EPI_ISL_1716466, EPI_ISL_1716467, EPI_ISL_1716468, EPI_ISL_1716471, EPI_ISL_1716472, EPI_ISL_1716474, EPI_ISL_1716480, EPI_ISL_1716481, EPI_ISL_1716482, EPI_ISL_1716483, EPI_ISL_1716484, EPI_ISL_1716485, EPI_ISL_1716487, EPI_ISL_1716488, EPI_ISL_1716490, EPI_ISL_1716491, EPI_ISL_1716495, EPI_ISL_1716496, EPI_ISL_1716497, EPI_ISL_1716499, EPI_ISL_1716501, EPI_ISL_1716503, EPI_ISL_1716504, EPI_ISL_1716505, EPI_ISL_1716506, EPI_ISL_1716507, EPI_ISL_1716508, EPI_ISL_1716509, EPI_ISL_1716510, EPI_ISL_1716511, EPI_ISL_1716512, EPI_ISL_1716513 | | | |
| see above | LACEN (Laboratorio de Saude Publica Dr. Giovanni Cysneiros) | LGBio (Laboratorio de Genetica & Biodiversidade) | Mariana Pires de Campos Telles, Daniela de Melo e Silva, Elisangela de Paula Silveira Lacerda, Renata de Oliveira Dias, Rhewter Nunes, Cintia Pelegrineti Targueta de Azevedo Brito, Ramilla dos Santos Braga, Thais Guimarães Castro, Thays Millena Alves Pedroso, Amanda Alves de Melo, Aparecido Divino da Cruz, Luiz Augusto Pereira, Thais Cidália Vieira Gigonzac, Marc Alexandre Duarte Gigonzac, Alex Honda Bernardes, Francyelli Mello Andrade |
| EPI_ISL_1731574 | UBDS Dr Marco Antonio Sahao Vila Virginia | Instituto Adolfo Lutz, Interdiciplinary Procedures Center, Strategic Laboratory | Claudio Tavares Sacchi, Claudia Regina Gonçalves, Erica Valessa Ramos Gomes, Karoline Rodrigues Campos, Caio Vinicius Dias Lopes, Leonardo Jose Tadeu de Araujo, Katia Correa de Oliveira Santos |
| EPI_ISL_1731575 | Casa de Caridade Sao Vicente de Paulo Cajuru | Instituto Adolfo Lutz, Interdiciplinary Procedures Center, Strategic Laboratory | Claudio Tavares Sacchi, Claudia Regina Gonçalves, Erica Valessa Ramos Gomes, Karoline Rodrigues Campos, Caio Vinicius Dias Lopes, Leonardo Jose Tadeu de Araujo, Katia Correa de Oliveira Santos |
| EPI_ISL_1731576 | UPA Dr Luis Atilio Losi Viana Ribeirao Preto | Instituto Adolfo Lutz, Interdiciplinary Procedures Center, Strategic Laboratory | Claudio Tavares Sacchi, Claudia Regina Gonçalves, Erica Valessa Ramos Gomes, Karoline Rodrigues Campos, Caio Vinicius Dias Lopes, Leonardo Jose Tadeu de Araujo, Katia Correa de Oliveira Santos |
| EPI_ISL_1731577 | Hospital Municipal Dr Mario Gatti Campinas | Instituto Adolfo Lutz, Interdiciplinary Procedures Center, Strategic Laboratory | Claudio Tavares Sacchi, Claudia Regina Gonçalves, Erica Valessa Ramos Gomes, Karoline Rodrigues Campos, Caio Vinicius Dias Lopes, Leonardo Jose Tadeu de Araujo, Katia Correa de Oliveira Santos |
| EPI_ISL_1731578, EPI_ISL_1731579, EPI_ISL_1731580 | Instituto Adolfo Lutz - Regional de Aracatuba | Instituto Adolfo Lutz, Interdiciplinary Procedures Center, Strategic Laboratory | Claudio Tavares Sacchi, Claudia Regina Gonçalves, Erica Valessa Ramos Gomes, Karoline Rodrigues Campos, Caio Vinicius Dias Lopes, Leonardo Jose Tadeu de Araujo, Katia Correa de Oliveira Santos |
| EPI_ISL_1731581, EPI_ISL_1731582 | Instituto Adolfo Lutz - Regional de Ribeirao Preto | Instituto Adolfo Lutz, Interdiciplinary Procedures Center, Strategic Laboratory | Claudio Tavares Sacchi, Claudia Regina Gonçalves, Erica Valessa Ramos Gomes, Karoline Rodrigues Campos, Caio Vinicius Dias Lopes, Leonardo Jose Tadeu de Araujo, Katia Correa de Oliveira Santos |
| EPI_ISL_1731583, EPI_ISL_1731584, EPI_ISL_1731585, EPI_ISL_1731586, EPI_ISL_1731587, EPI_ISL_1731588, EPI_ISL_1731589, EPI_ISL_1731590, EPI_ISL_1731591, EPI_ISL_1731592, EPI_ISL_1731594, EPI_ISL_1731595, EPI_ISL_1731596, EPI_ISL_1731597, EPI_ISL_1731598, EPI_ISL_1731599, EPI_ISL_1731600, EPI_ISL_1731601, EPI_ISL_1731602, EPI_ISL_1731603, EPI_ISL_1731604, EPI_ISL_1731605, EPI_ISL_1731607, EPI_ISL_1731608, EPI_ISL_1731609, EPI_ISL_1731610, EPI_ISL_1731611, EPI_ISL_1731612 | | | |
| see above | Instituto Adolfo Lutz Central | Instituto Adolfo Lutz, Interdiciplinary Procedures Center, Strategic Laboratory | Claudio Tavares Sacchi, Claudia Regina Gonçalves, Erica Valessa Ramos Gomes, Karoline Rodrigues Campos, Caio Vinicius Dias Lopes, Leonardo Jose Tadeu de Araujo, Katia Correa de Oliveira Santos |
| EPI_ISL_1734843, EPI_ISL_1734844, EPI_ISL_1734846, EPI_ISL_1734847, EPI_ISL_1734849, EPI_ISL_1734850, EPI_ISL_1734852, EPI_ISL_1734853, EPI_ISL_1734855, EPI_ISL_1734857, EPI_ISL_1734858, EPI_ISL_1734860, EPI_ISL_1734861, EPI_ISL_1734863, EPI_ISL_1734866, EPI_ISL_1734868, EPI_ISL_1734869, EPI_ISL_1734871, EPI_ISL_1734872, EPI_ISL_1734874, EPI_ISL_1734877, EPI_ISL_1734878, EPI_ISL_1734880, EPI_ISL_1734881, EPI_ISL_1734883, EPI_ISL_1734884 | | | |
| see above | Instituto de Biotecnologia - UNESP-Botucatu-SP | Instituto de Biotecnologia - UNESP-Botucatu-SP | Fábio Sossai Possebon; Leila Sabrina Ullmann; Cecília Artico Banho; Cíntia Bittar; Guilherme Campos; Helena Lage Ferreira; Jorge A. Petroli Marchesi; Lívia Sacchetto; Maisa C. Pereira Parra; Marília Moraes; Maurício L. Nogueira; Paula Rahal; Paulo Inacio da Costa; João Pessoa Araújo Jr. |
| EPI_ISL_1752637 | UBDS Dr Marco Antonio Sahao Vila Virginia | Instituto Adolfo Lutz, Interdiciplinary Procedures Center, Strategic Laboratory | Claudio Tavares Sacchi, Claudia Regina Gonçalves, Erica Valessa Ramos Gomes, Karoline Rodrigues Campos, Caio Vinicius Dias Lopes, Leonardo Jose Tadeu de Araujo, Katia Correa de Oliveira Santos |
| EPI_ISL_1752638 | Casa de Caridade Sao Vicente de Paulo Cajuru | Instituto Adolfo Lutz, Interdiciplinary Procedures Center, Strategic Laboratory | Claudio Tavares Sacchi, Claudia Regina Gonçalves, Erica Valessa Ramos Gomes, Karoline Rodrigues Campos, Caio Vinicius Dias Lopes, Leonardo Jose Tadeu de Araujo, Katia Correa de Oliveira Santos |
| EPI_ISL_1752639 | UPA Dr Luis Atilio Losi Viana Ribeirao Preto | Instituto Adolfo Lutz, Interdiciplinary Procedures Center, Strategic Laboratory | Claudio Tavares Sacchi, Claudia Regina Gonçalves, Erica Valessa Ramos Gomes, Karoline Rodrigues Campos, Caio Vinicius Dias Lopes, Leonardo Jose Tadeu de Araujo, Katia Correa de Oliveira Santos |
| EPI_ISL_1752640 | Hospital Municipal Dr Mario Gatti Campinas | Instituto Adolfo Lutz, Interdiciplinary Procedures Center, Strategic Laboratory | Claudio Tavares Sacchi, Claudia Regina Gonçalves, Erica Valessa Ramos Gomes, Karoline Rodrigues Campos, Caio Vinicius Dias Lopes, Leonardo Jose Tadeu de Araujo, Katia Correa de Oliveira Santos |
| EPI_ISL_1752641, EPI_ISL_1752642, EPI_ISL_1752643 | Instituto Adolfo Lutz - Regional de Aracatuba | Instituto Adolfo Lutz, Interdiciplinary Procedures Center, Strategic Laboratory | Claudio Tavares Sacchi, Claudia Regina Gonçalves, Erica Valessa Ramos Gomes, Karoline Rodrigues Campos, Caio Vinicius Dias Lopes, Leonardo Jose Tadeu de Araujo, Katia Correa de Oliveira Santos |
| EPI_ISL_1752645 | Instituto Adolfo Lutz - Regional de Ribeirao Preto | Instituto Adolfo Lutz, Interdiciplinary Procedures Center, Strategic Laboratory | Claudio Tavares Sacchi, Claudia Regina Gonçalves, Erica Valessa Ramos Gomes, Karoline Rodrigues Campos, Caio Vinicius Dias Lopes, Leonardo Jose Tadeu de Araujo, Katia Correa de Oliveira Santos |
| EPI_ISL_1752646, EPI_ISL_1752647, EPI_ISL_1752648, EPI_ISL_1752649, EPI_ISL_1752650, EPI_ISL_1752651, EPI_ISL_1752652, EPI_ISL_1752653, EPI_ISL_1752654, EPI_ISL_1752655, EPI_ISL_1752656, EPI_ISL_1752657, EPI_ISL_1752658, EPI_ISL_1752659, EPI_ISL_1752660, EPI_ISL_1752661, EPI_ISL_1752662, EPI_ISL_1752663, EPI_ISL_1752664, EPI_ISL_1752665, EPI_ISL_1752666, EPI_ISL_1752667, EPI_ISL_1752668 | | | |
| see above | Instituto Adolfo Lutz Central | Instituto Adolfo Lutz, Interdiciplinary Procedures Center, Strategic Laboratory | Claudio Tavares Sacchi, Claudia Regina Gonçalves, Erica Valessa Ramos Gomes, Karoline Rodrigues Campos, Caio Vinicius Dias Lopes, Leonardo Jose Tadeu de Araujo, Katia Correa de Oliveira Santos |
| EPI_ISL_1754186 | Laboratório de Pesquisa em Virologia, FAMERP, SJRP | Laboratório de Pesquisa em Virologia, FAMERP, SJRP | Cecília Artico Banho; Fábio Sossai Possebon; Leila Sabrina Ullmann; Cíntia Bittar; Guilherme Campos; Helena Lage Ferreira; Jorge A. Petroli Marchesi; Lívia Sacchetto; Maisa C. Pereira Parra; Marília Moraes; Paula Rahal; Paulo Inacio da Costa; João Pessoa Araújo Jr.; Maurício L. Nogueira |
| EPI_ISL_1786560, EPI_ISL_1786561, EPI_ISL_1786562, EPI_ISL_1786563, EPI_ISL_1786564, EPI_ISL_1786565, EPI_ISL_1786566, EPI_ISL_1786567, EPI_ISL_1786568 | Laboratorio de Virologia Clinica do HCFMRP-USP | Hocentro de Ribeirao Preto - Laboratório de Biologia Molecular | Svetoslav Nanev Slavov, Marta Giovanetti, Rafael dos Santos Bezerra, Vagner Fonseca, Elaine Vieira Santos, Evandra Strazza Rodrigues, Talita Adelino, Joilson Xavier, Josiane Serrano Borges, Mariane Evaristo, Glauco de Carvalho Pereira, Aparecida Yulie Yamamoto, Diego Villa Clé, Rodrigo Tocantins Calado, Dimas Tadeu Covas, Luiz Carlos Junior Alcantara, Simone Kashima |
| EPI_ISL_1795081 | CENTRO DE SAUDE III AFFONSO LUZZI SANTA CRUZ DAS PALMEIRAS | Instituto Butantan / ESALQ-Piracicaba | Instituto Butantan: Alexander Roberto Precioso, Dimas Tadeu Covas, Sandra Coccuzzo Sampaio, Maria Carolina Elias, José Salvatore Leister Patané, Vincent Louis Viala, Antonio Jorge Martins, Ricardo Haddad, Claudia Renata dos Santos Barros, Elaine Cristina Marqueze, Raul Machado Neto, Debora Botequio Moretti. Centro de Genômica Funcional da ESALQ: Luiz Lehmann Coutinho, Ricardo Augusto Brassaloti, Raquel de Lello Rocha Campos Cassano. NGS Soluções Genômicas: Pilar Drummond Sampaio Corrêa Mariani. FZEA-USP PirassunUnga: Mirele Daiana Poleti, Jessika Cristina Chagas Lesbon, Elisangela Chicaroni Mattos, Heidge Fukumasu. USP-Botucatu: Rejane Maria Tommasini Grotto, Jayme A. Souza-Neto, Guilherme Targino Valente, Patricia Akemi Assato, Felipe Allan da Silva da Costa, Bianca Cechetto Carlos. Mendelics: Bibiana Santos, João Paulo Kitajima, Erika Freitas, David Schlesinger. Hemocentro Ribeirão Preto: Simone Kashima, Evandra Strazza Rodrigues, Svetoslav Nanev Slavov, Elaine Vieira dos Santos, Rafael dos Santos Bezerra, Luiz Carlos Junior de Alcantara, Marta Giovanetti, Vagner Fonseca, Flavia Aburjaile, Rodrigo Tocantins Calado. |
| EPI_ISL_1795082, EPI_ISL_1795083 | POLICLINICA HORTOLANDIA | Instituto Butantan / ESALQ-Piracicaba | Instituto Butantan: Alexander Roberto Precioso, Dimas Tadeu Covas, Sandra Coccuzzo Sampaio, Maria Carolina Elias, José Salvatore Leister Patané, Vincent Louis Viala, Antonio Jorge Martins, Ricardo Haddad, Claudia Renata dos Santos Barros, Elaine Cristina Marqueze, Raul Machado Neto, Debora Botequio Moretti. Centro de Genômica Funcional da ESALQ: Luiz Lehmann Coutinho, Ricardo Augusto Brassaloti, Raquel de Lello Rocha Campos Cassano. NGS Soluções Genômicas: Pilar Drummond Sampaio Corrêa Mariani. FZEA-USP Pirassununga: Mirele Daiana Poleti, Jessika Cristina Chagas Lesbon, Elisangela Chicaroni Mattos, Heidge Fukumasu. USP-Botucatu: Rejane Maria Tommasini Grotto, Jayme A. Souza-Neto, Guilherme Targino Valente, Patricia Akemi Assato, Felipe Allan da Silva da Costa, Bianca Cechetto Carlos. Mendelics: Bibiana Santos, João Paulo Kitajima, Erika Freitas, |

| | | | |
|---|---|---|---|
| | | | David Schlesinger. Hemocentro Ribeirão Preto: Simone Kashima, Evandra Strazza Rodrigues, Svetoslav Nanev Slavov, Elaine Vieira dos Santos, Rafael dos Santos Bezerra, Luiz Carlos Junior de Alcantara, Marta Giovanetti, Vagner Fonseca, Flavia Aburjaile, Rodrigo Tocantins Calado. |
| EPI_ISL_1795093 | CENTRO INTEGRADO DE SAUDE | Instituto Butantan / ESALQ-Piracicaba | Instituto Butantan: Alexander Roberto Precioso, Dimas Tadeu Covas, Sandra Coccuzzo Sampaio, Maria Carolina Elias, José Salvatore Leister Patané, Vincent Louis Viala, Antonio Jorge Martins, Ricardo Haddad, Claudia Renata dos Santos Barros, Elaine Cristina Marqueze, Raul Machado Neto, Debora Botequio Moretti. Centro de Genômica Funcional da ESALQ: Luiz Lehmann Coutinho, Ricardo Augusto Brassaloti, Raquel de Lello Rocha Campos Cassano. NGS Soluções Genômicas: Pilar Drummond Sampaio Corrêa Mariani. FZEA-USP Pirassununga: Mirele Daiana Poleti, Jessika Cristina Chagas Lesbon, Elisangela Chicaroni Mattos, Heidge Fukumasu. USP-Botucatu: Rejane Maria Tommasini Grotto, Jayme A. Souza-Neto, Guilherme Targino Valente, Patricia Akemi Assato, Felipe Allan da Silva da Costa, Bianca Cechetto Carlos. Mendelics: Bibiana Santos, João Paulo Kitajima, Erika Freitas, David Schlesinger. Hemocentro Ribeirão Preto: Simone Kashima, Evandra Strazza Rodrigues, Svetoslav Nanev Slavov, Elaine Vieira dos Santos, Rafael dos Santos Bezerra, Luiz Carlos Junior de Alcantara, Marta Giovanetti, Vagner Fonseca, Flavia Aburjaile, Rodrigo Tocantins Calado. |
| EPI_ISL_1795094 | VIGILANCIA EPIDEMIOLOGICA DE LEME | Instituto Butantan / ESALQ-Piracicaba | Instituto Butantan: Alexander Roberto Precioso, Dimas Tadeu Covas, Sandra Coccuzzo Sampaio, Maria Carolina Elias, José Salvatore Leister Patané, Vincent Louis Viala, Antonio Jorge Martins, Ricardo Haddad, Claudia Renata dos Santos Barros, Elaine Cristina Marqueze, Raul Machado Neto, Debora Botequio Moretti. Centro de Genômica Funcional da ESALQ: Luiz Lehmann Coutinho, Ricardo Augusto Brassaloti, Raquel de Lello Rocha Campos Cassano. NGS Soluções Genômicas: Pilar Drummond Sampaio Corrêa Mariani. FZEA-USP Pirassununga: Mirele Daiana Poleti, Jessika Cristina Chagas Lesbon, Elisangela Chicaroni Mattos, Heidge Fukumasu. USP-Botucatu: Rejane Maria Tommasini Grotto, Jayme A. Souza-Neto, Guilherme Targino Valente, Patricia Akemi Assato, Felipe Allan da Silva da Costa, Bianca Cechetto Carlos. Mendelics: Bibiana Santos, João Paulo Kitajima, Erika Freitas, David Schlesinger. Hemocentro Ribeirão Preto: Simone Kashima, Evandra Strazza Rodrigues, Svetoslav Nanev Slavov, Elaine Vieira dos Santos, Rafael dos Santos Bezerra, Luiz Carlos Junior de Alcantara, Marta Giovanetti, Vagner Fonseca, Flavia Aburjaile, Rodrigo Tocantins Calado. |
| EPI_ISL_1795097 | LABORATORIO DR PAULO EMILIO DALESSANDRO PINDAMONHANGABA | Instituto Butantan / ESALQ-Piracicaba | Instituto Butantan: Alexander Roberto Precioso, Dimas Tadeu Covas, Sandra Coccuzzo Sampaio, Maria Carolina Elias, José Salvatore Leister Patané, Vincent Louis Viala, Antonio Jorge Martins, Ricardo Haddad, Claudia Renata dos Santos Barros, Elaine Cristina Marqueze, Raul Machado Neto, Debora Botequio Moretti. Centro de Genômica Funcional da ESALQ: Luiz Lehmann Coutinho, Ricardo Augusto Brassaloti, Raquel de Lello Rocha Campos Cassano. NGS Soluções Genômicas: Pilar Drummond Sampaio Corrêa Mariani. FZEA-USP Pirassununga: Mirele Daiana Poleti, Jessika Cristina Chagas Lesbon, Elisangela Chicaroni Mattos, Heidge Fukumasu. USP-Botucatu: Rejane Maria Tommasini Grotto, Jayme A. Souza-Neto, Guilherme Targino Valente, Patricia Akemi Assato, Felipe Allan da Silva da Costa, Bianca Cechetto Carlos. Mendelics: Bibiana Santos, João Paulo Kitajima, Erika Freitas, David Schlesinger. Hemocentro Ribeirão Preto: Simone Kashima, Evandra Strazza Rodrigues, Svetoslav Nanev Slavov, Elaine Vieira dos Santos, Rafael dos Santos Bezerra, Luiz Carlos Junior de Alcantara, Marta Giovanetti, Vagner Fonseca, Flavia Aburjaile, Rodrigo Tocantins Calado. |
| EPI_ISL_1795098, EPI_ISL_1795099 | ESALQ | Instituto Butantan / ESALQ-Piracicaba | Instituto Butantan: Alexander Roberto Precioso, Dimas Tadeu Covas, Sandra Coccuzzo Sampaio, Maria Carolina Elias, José Salvatore Leister Patané, Vincent Louis Viala, Antonio Jorge Martins, Ricardo Haddad, Claudia Renata dos Santos Barros, Elaine Cristina Marqueze, Raul Machado Neto, Debora Botequio Moretti. Centro de Genômica Funcional da ESALQ: Luiz Lehmann Coutinho, Ricardo Augusto Brassaloti, Raquel de Lello Rocha Campos Cassano. NGS Soluções Genômicas: Pilar Drummond Sampaio Corrêa Mariani. FZEA-USP Pirassununga: Mirele Daiana Poleti, Jessika Cristina Chagas Lesbon, Elisangela Chicaroni Mattos, Heidge Fukumasu. USP-Botucatu: Rejane Maria Tommasini Grotto, Jayme A. Souza-Neto, Guilherme Targino Valente, Patricia Akemi Assato, Felipe Allan da Silva da Costa, Bianca Cechetto Carlos. Mendelics: Bibiana Santos, João Paulo Kitajima, Erika Freitas, David Schlesinger. Hemocentro Ribeirão Preto: Simone Kashima, Evandra Strazza Rodrigues, Svetoslav Nanev Slavov, Elaine Vieira dos Santos, Rafael dos Santos Bezerra, Luiz Carlos Junior de Alcantara, Marta Giovanetti, Vagner Fonseca, Flavia Aburjaile, Rodrigo Tocantins Calado. |
| EPI_ISL_1795100 | LABORATORIO MUNICIPAL DE PIRACICABA | Instituto Butantan / ESALQ-Piracicaba | Instituto Butantan: Alexander Roberto Precioso, Dimas Tadeu Covas, Sandra Coccuzzo Sampaio, Maria Carolina Elias, José Salvatore Leister Patané, Vincent Louis Viala, Antonio Jorge Martins, Ricardo Haddad, Claudia Renata dos Santos Barros, Elaine Cristina Marqueze, Raul Machado Neto, Debora Botequio Moretti. Centro de Genômica Funcional da ESALQ: Luiz Lehmann Coutinho, Ricardo Augusto Brassaloti, Raquel de Lello Rocha Campos Cassano. NGS Soluções Genômicas: Pilar Drummond Sampaio Corrêa Mariani. FZEA-USP Pirassununga: Mirele Daiana Poleti, Jessika Cristina Chagas Lesbon, Elisangela Chicaroni Mattos, Heidge Fukumasu. USP-Botucatu: Rejane Maria Tommasini Grotto, Jayme A. Souza-Neto, Guilherme Targino Valente, Patricia Akemi Assato, Felipe Allan da Silva da Costa, Bianca Cechetto Carlos. Mendelics: Bibiana Santos, João Paulo Kitajima, Erika Freitas, David Schlesinger. Hemocentro Ribeirão Preto: Simone Kashima, Evandra Strazza Rodrigues, Svetoslav Nanev Slavov, Elaine Vieira dos Santos, Rafael dos Santos Bezerra, Luiz Carlos Junior de Alcantara, Marta Giovanetti, Vagner Fonseca, Flavia Aburjaile, Rodrigo Tocantins Calado. |
| EPI_ISL_1795101 | HOSPITAL DOS FORNCEDORES | Instituto Butantan / ESALQ-Piracicaba | Instituto Butantan: Alexander Roberto Precioso, Dimas Tadeu Covas, Sandra Coccuzzo Sampaio, Maria Carolina Elias, José Salvatore Leister Patané, Vincent Louis Viala, Antonio Jorge Martins, Ricardo Haddad, Claudia Renata dos Santos Barros, Elaine Cristina Marqueze, Raul Machado Neto, Debora Botequio Moretti. Centro de Genômica Funcional da ESALQ: Luiz Lehmann Coutinho, Ricardo Augusto Brassaloti, Raquel de Lello Rocha Campos Cassano. NGS Soluções Genômicas: Pilar Drummond Sampaio Corrêa Mariani. FZEA-USP Pirassununga: Mirele Daiana Poleti, Jessika Cristina Chagas Lesbon, Elisangela Chicaroni Mattos, Heidge Fukumasu. USP-Botucatu: Rejane Maria Tommasini Grotto, Jayme A. Souza-Neto, Guilherme Targino Valente, Patricia Akemi Assato, Felipe Allan da Silva da Costa, Bianca Cechetto Carlos. Mendelics: Bibiana Santos, João Paulo Kitajima, Erika Freitas, David Schlesinger. Hemocentro Ribeirão Preto: Simone Kashima, Evandra Strazza Rodrigues, Svetoslav Nanev Slavov, Elaine Vieira dos Santos, Rafael dos Santos Bezerra, Luiz Carlos Junior de Alcantara, Marta Giovanetti, Vagner Fonseca, Flavia Aburjaile, Rodrigo Tocantins Calado. |
| EPI_ISL_1795102 | LABORATORIO MUNICIPAL DE PIRACICABA | Instituto Butantan / ESALQ-Piracicaba | Instituto Butantan: Alexander Roberto Precioso, Dimas Tadeu Covas, Sandra Coccuzzo Sampaio, Maria Carolina Elias, José Salvatore Leister Patané, Vincent Louis Viala, Antonio Jorge Martins, Ricardo Haddad, Claudia Renata dos Santos Barros, Elaine Cristina Marqueze, Raul Machado Neto, Debora Botequio Moretti. Centro de Genômica Funcional da ESALQ: Luiz Lehmann Coutinho, Ricardo Augusto Brassaloti, Raquel de Lello Rocha Campos Cassano. NGS Soluções Genômicas: Pilar Drummond Sampaio Corrêa Mariani. FZEA-USP Pirassununga: Mirele Daiana Poleti, Jessika Cristina Chagas Lesbon, Elisangela Chicaroni Mattos, Heidge Fukumasu. USP-Botucatu: Rejane Maria Tommasini Grotto, Jayme A. Souza-Neto, Guilherme Targino Valente, Patricia Akemi Assato, Felipe Allan da Silva da Costa, Bianca Cechetto Carlos. Mendelics: Bibiana Santos, João Paulo Kitajima, Erika Freitas, David Schlesinger. Hemocentro Ribeirão Preto: Simone Kashima, Evandra Strazza Rodrigues, Svetoslav Nanev Slavov, Elaine Vieira dos Santos, Rafael dos Santos Bezerra, Luiz Carlos Junior de Alcantara, Marta Giovanetti, Vagner Fonseca, Flavia Aburjaile, Rodrigo Tocantins Calado. |
| EPI_ISL_1795103 | HOSPITAL DOS FORNCEDORES | Instituto Butantan / ESALQ-Piracicaba | Instituto Butantan: Alexander Roberto Precioso, Dimas Tadeu Covas, Sandra Coccuzzo Sampaio, Maria Carolina Elias, José Salvatore Leister Patané, Vincent Louis Viala, Antonio Jorge Martins, Ricardo Haddad, Claudia Renata dos Santos Barros, Elaine Cristina Marqueze, Raul Machado Neto, Debora Botequio Moretti. Centro de Genômica Funcional da ESALQ: Luiz Lehmann Coutinho, Ricardo Augusto Brassaloti, Raquel de Lello Rocha Campos Cassano. NGS Soluções Genômicas: Pilar Drummond Sampaio Corrêa Mariani. FZEA-USP Pirassununga: Mirele Daiana Poleti, Jessika Cristina Chagas Lesbon, Elisangela Chicaroni Mattos, Heidge Fukumasu. USP-Botucatu: Rejane Maria Tommasini Grotto, Jayme A. Souza-Neto, Guilherme Targino Valente, Patricia Akemi Assato, Felipe Allan da Silva da Costa, Bianca Cechetto Carlos. Mendelics: Bibiana Santos, João Paulo Kitajima, Erika Freitas, David Schlesinger. Hemocentro Ribeirão Preto: Simone Kashima, Evandra Strazza Rodrigues, Svetoslav Nanev Slavov, Elaine Vieira dos Santos, Rafael dos Santos Bezerra, Luiz Carlos Junior de Alcantara, Marta Giovanetti, Vagner Fonseca, Flavia Aburjaile, Rodrigo Tocantins Calado. |
| EPI_ISL_1795104, EPI_ISL_1795105 | LABORATORIO MUNICIPAL DE PIRACICABA | Instituto Butantan / ESALQ-Piracicaba | Instituto Butantan: Alexander Roberto Precioso, Dimas Tadeu Covas, Sandra Coccuzzo Sampaio, Maria Carolina Elias, José Salvatore Leister Patané, Vincent Louis Viala, Antonio Jorge Martins, Ricardo Haddad, Claudia Renata dos Santos Barros, Elaine Cristina Marqueze, Raul Machado Neto, Debora Botequio Moretti. Centro de Genômica Funcional da ESALQ: Luiz Lehmann Coutinho, Ricardo Augusto Brassaloti, Raquel de Lello Rocha Campos Cassano. NGS Soluções Genômicas: Pilar Drummond Sampaio Corrêa Mariani. FZEA-USP Pirassununga: Mirele Daiana Poleti, Jessika Cristina Chagas Lesbon, Elisangela Chicaroni Mattos, Heidge Fukumasu. USP-Botucatu: Rejane Maria Tommasini Grotto, Jayme A. Souza-Neto, Guilherme Targino Valente, Patricia Akemi Assato, Felipe Allan da Silva da Costa, Bianca Cechetto Carlos. Mendelics: Bibiana Santos, João Paulo Kitajima, Erika Freitas, David Schlesinger. Hemocentro Ribeirão Preto: Simone Kashima, Evandra Strazza Rodrigues, Svetoslav Nanev Slavov, Elaine Vieira dos Santos, Rafael dos Santos Bezerra, Luiz Carlos Junior de Alcantara, Marta Giovanetti, Vagner Fonseca, Flavia Aburjaile, Rodrigo Tocantins Calado. |
| EPI_ISL_1795106 | HOSPITAL DOS FORNCEDORES | Instituto Butantan / ESALQ-Piracicaba | Instituto Butantan: Alexander Roberto Precioso, Dimas Tadeu Covas, Sandra Coccuzzo Sampaio, Maria Carolina Elias, José Salvatore Leister Patané, Vincent Louis Viala, Antonio Jorge Martins, Ricardo Haddad, Claudia Renata dos Santos Barros, Elaine Cristina Marqueze, Raul Machado Neto, Debora Botequio Moretti. Centro de Genômica Funcional da ESALQ: Luiz Lehmann Coutinho, Ricardo Augusto Brassaloti, Raquel de Lello Rocha Campos Cassano. NGS Soluções Genômicas: Pilar Drummond Sampaio Corrêa Mariani. FZEA-USP Pirassununga: Mirele Daiana Poleti, Jessika Cristina Chagas Lesbon, Elisangela Chicaroni Mattos, Heidge Fukumasu. USP-Botucatu: Rejane Maria Tommasini Grotto, Jayme A. Souza-Neto, Guilherme Targino Valente, Patricia Akemi Assato, Felipe Allan da Silva da Costa, Bianca Cechetto Carlos. Mendelics: Bibiana Santos, João Paulo Kitajima, Erika Freitas, David Schlesinger. Hemocentro Ribeirão Preto: Simone Kashima, Evandra Strazza Rodrigues, Svetoslav Nanev Slavov, Elaine Vieira dos Santos, Rafael dos Santos Bezerra, Luiz Carlos Junior de Alcantara, Marta Giovanetti, Vagner Fonseca, Flavia Aburjaile, Rodrigo Tocantins Calado. |

| | | | |
|---|---|---|---|
| EPI_ISL_1795107 | COORDENADORIA MUNICIPAL DE SAUDE DE IRACEMAPOLIS | Instituto Butantan / ESALQ-Piracicaba | Instituto Butantan: Alexander Roberto Precioso, Dimas Tadeu Covas, Sandra Coccuzzo Sampaio, Maria Carolina Elias, José Salvatore Leister Patané, Vincent Louis Viala, Antonio Jorge Martins, Ricardo Haddad, Claudia Renata dos Santos Barros, Elaine Cristina Marqueze, Raul Machado Neto, Debora Botequio Moretti. Centro de Genômica Funcional da ESALQ: Luiz Lehmann Coutinho, Ricardo Augusto Brassaloti, Raquel de Lello Rocha Campos Cassano. NGS Soluções Genômicas: Pilar Drummond Sampaio Corrêa Mariani. FZEA-USP Pirassununga: Mirele Daiana Poleti, Jessika Cristina Chagas Lesbon, Elisangela Chicaroni Mattos, Heidge Fukumasu. USP-Botucatu: Rejane Maria Tommasini Grotto, Jayme A. Souza-Neto, Guilherme Targino Valente, Patricia Akemi Assato, Felipe Allan da Silva da Costa, Bianca Cechetto Carlos. Mendelics: Bibiana Santos, João Paulo Kitajima, Erika Freitas, David Schlesinger. Hemocentro Ribeirão Preto: Simone Kashima, Evandra Strazza Rodrigues, Svetoslav Nanev Slavov, Elaine Vieira dos Santos, Rafael dos Santos Bezerra, Luiz Carlos Junior de Alcantara, Marta Giovanetti, Vagner Fonseca, Flavia Aburjaile, Rodrigo Tocantins Calado. |
| EPI_ISL_1795108 | SECRETARIA DE SAUDE DE SAO PEDRO | Instituto Butantan / ESALQ-Piracicaba | Instituto Butantan: Alexander Roberto Precioso, Dimas Tadeu Covas, Sandra Coccuzzo Sampaio, Maria Carolina Elias, José Salvatore Leister Patané, Vincent Louis Viala, Antonio Jorge Martins, Ricardo Haddad, Claudia Renata dos Santos Barros, Elaine Cristina Marqueze, Raul Machado Neto, Debora Botequio Moretti. Centro de Genômica Funcional da ESALQ: Luiz Lehmann Coutinho, Ricardo Augusto Brassaloti, Raquel de Lello Rocha Campos Cassano. NGS Soluções Genômicas: Pilar Drummond Sampaio Corrêa Mariani. FZEA-USP Pirassununga: Mirele Daiana Poleti, Jessika Cristina Chagas Lesbon, Elisangela Chicaroni Mattos, Heidge Fukumasu. USP-Botucatu: Rejane Maria Tommasini Grotto, Jayme A. Souza-Neto, Guilherme Targino Valente, Patricia Akemi Assato, Felipe Allan da Silva da Costa, Bianca Cechetto Carlos. Mendelics: Bibiana Santos, João Paulo Kitajima, Erika Freitas, David Schlesinger. Hemocentro Ribeirão Preto: Simone Kashima, Evandra Strazza Rodrigues, Svetoslav Nanev Slavov, Elaine Vieira dos Santos, Rafael dos Santos Bezerra, Luiz Carlos Junior de Alcantara, Marta Giovanetti, Vagner Fonseca, Flavia Aburjaile, Rodrigo Tocantins Calado. |
| EPI_ISL_1795109 | CENTRO DE ESPECIALIDDS MEDICAS IRMA LEOPOLDINA PIRASSUNUNGA | Instituto Butantan / ESALQ-Piracicaba | Instituto Butantan: Alexander Roberto Precioso, Dimas Tadeu Covas, Sandra Coccuzzo Sampaio, Maria Carolina Elias, José Salvatore Leister Patané, Vincent Louis Viala, Antonio Jorge Martins, Ricardo Haddad, Claudia Renata dos Santos Barros, Elaine Cristina Marqueze, Raul Machado Neto, Debora Botequio Moretti. Centro de Genômica Funcional da ESALQ: Luiz Lehmann Coutinho, Ricardo Augusto Brassaloti, Raquel de Lello Rocha Campos Cassano. NGS Soluções Genômicas: Pilar Drummond Sampaio Corrêa Mariani. FZEA-USP Pirassununga: Mirele Daiana Poleti, Jessika Cristina Chagas Lesbon, Elisangela Chicaroni Mattos, Heidge Fukumasu. USP-Botucatu: Rejane Maria Tommasini Grotto, Jayme A. Souza-Neto, Guilherme Targino Valente, Patricia Akemi Assato, Felipe Allan da Silva da Costa, Bianca Cechetto Carlos. Mendelics: Bibiana Santos, João Paulo Kitajima, Erika Freitas, David Schlesinger. Hemocentro Ribeirão Preto: Simone Kashima, Evandra Strazza Rodrigues, Svetoslav Nanev Slavov, Elaine Vieira dos Santos, Rafael dos Santos Bezerra, Luiz Carlos Junior de Alcantara, Marta Giovanetti, Vagner Fonseca, Flavia Aburjaile, Rodrigo Tocantins Calado. |
| EPI_ISL_1795110 | UNIDADE ESF DR LUIZ SPINA | Instituto Butantan / ESALQ-Piracicaba | Instituto Butantan: Alexander Roberto Precioso, Dimas Tadeu Covas, Sandra Coccuzzo Sampaio, Maria Carolina Elias, José Salvatore Leister Patané, Vincent Louis Viala, Antonio Jorge Martins, Ricardo Haddad, Claudia Renata dos Santos Barros, Elaine Cristina Marqueze, Raul Machado Neto, Debora Botequio Moretti. Centro de Genômica Funcional da ESALQ: Luiz Lehmann Coutinho, Ricardo Augusto Brassaloti, Raquel de Lello Rocha Campos Cassano. NGS Soluções Genômicas: Pilar Drummond Sampaio Corrêa Mariani. FZEA-USP Pirassununga: Mirele Daiana Poleti, Jessika Cristina Chagas Lesbon, Elisangela Chicaroni Mattos, Heidge Fukumasu. USP-Botucatu: Rejane Maria Tommasini Grotto, Jayme A. Souza-Neto, Guilherme Targino Valente, Patricia Akemi Assato, Felipe Allan da Silva da Costa, Bianca Cechetto Carlos. Mendelics: Bibiana Santos, João Paulo Kitajima, Erika Freitas, David Schlesinger. Hemocentro Ribeirão Preto: Simone Kashima, Evandra Strazza Rodrigues, Svetoslav Nanev Slavov, Elaine Vieira dos Santos, Rafael dos Santos Bezerra, Luiz Carlos Junior de Alcantara, Marta Giovanetti, Vagner Fonseca, Flavia Aburjaile, Rodrigo Tocantins Calado. |
| EPI_ISL_1795111, EPI_ISL_1795112 | SECRETARIA MUNICIPAL DA SAUDE DE GUARIBA | Instituto Butantan / ESALQ-Piracicaba | Instituto Butantan: Alexander Roberto Precioso, Dimas Tadeu Covas, Sandra Coccuzzo Sampaio, Maria Carolina Elias, José Salvatore Leister Patané, Vincent Louis Viala, Antonio Jorge Martins, Ricardo Haddad, Claudia Renata dos Santos Barros, Elaine Cristina Marqueze, Raul Machado Neto, Debora Botequio Moretti. Centro de Genômica Funcional da ESALQ: Luiz Lehmann Coutinho, Ricardo Augusto Brassaloti, Raquel de Lello Rocha Campos Cassano. NGS Soluções Genômicas: Pilar Drummond Sampaio Corrêa Mariani. FZEA-USP Pirassununga: Mirele Daiana Poleti, Jessika Cristina Chagas Lesbon, Elisangela Chicaroni Mattos, Heidge Fukumasu. USP-Botucatu: Rejane Maria Tommasini Grotto, Jayme A. Souza-Neto, Guilherme Targino Valente, Patricia Akemi Assato, Felipe Allan da Silva da Costa, Bianca Cechetto Carlos. Mendelics: Bibiana Santos, João Paulo Kitajima, Erika Freitas, David Schlesinger. Hemocentro Ribeirão Preto: Simone Kashima, Evandra Strazza Rodrigues, Svetoslav Nanev Slavov, Elaine Vieira dos Santos, Rafael dos Santos Bezerra, Luiz Carlos Junior de Alcantara, Marta Giovanetti, Vagner Fonseca, Flavia Aburjaile, Rodrigo Tocantins Calado. |
| EPI_ISL_1795113 | AMBULATORIO DE ATENDIMENTO DE DST DE GUARIBA | Instituto Butantan / ESALQ-Piracicaba | Instituto Butantan: Alexander Roberto Precioso, Dimas Tadeu Covas, Sandra Coccuzzo Sampaio, Maria Carolina Elias, José Salvatore Leister Patané, Vincent Louis Viala, Antonio Jorge Martins, Ricardo Haddad, Claudia Renata dos Santos Barros, Elaine Cristina Marqueze, Raul Machado Neto, Debora Botequio Moretti. Centro de Genômica Funcional da ESALQ: Luiz Lehmann Coutinho, Ricardo Augusto Brassaloti, Raquel de Lello Rocha Campos Cassano. NGS Soluções Genômicas: Pilar Drummond Sampaio Corrêa Mariani. FZEA-USP Pirassununga: Mirele Daiana Poleti, Jessika Cristina Chagas Lesbon, Elisangela Chicaroni Mattos, Heidge Fukumasu. USP-Botucatu: Rejane Maria Tommasini Grotto, Jayme A. Souza-Neto, Guilherme Targino Valente, Patricia Akemi Assato, Felipe Allan da Silva da Costa, Bianca Cechetto Carlos. Mendelics: Bibiana Santos, João Paulo Kitajima, Erika Freitas, David Schlesinger. Hemocentro Ribeirão Preto: Simone Kashima, Evandra Strazza Rodrigues, Svetoslav Nanev Slavov, Elaine Vieira dos Santos, Rafael dos Santos Bezerra, Luiz Carlos Junior de Alcantara, Marta Giovanetti, Vagner Fonseca, Flavia Aburjaile, Rodrigo Tocantins Calado. |
| EPI_ISL_1795114, EPI_ISL_1795115 | CENTRO DE SAUDE III BORBOREMA | Instituto Butantan / ESALQ-Piracicaba | Instituto Butantan: Alexander Roberto Precioso, Dimas Tadeu Covas, Sandra Coccuzzo Sampaio, Maria Carolina Elias, José Salvatore Leister Patané, Vincent Louis Viala, Antonio Jorge Martins, Ricardo Haddad, Claudia Renata dos Santos Barros, Elaine Cristina Marqueze, Raul Machado Neto, Debora Botequio Moretti. Centro de Genômica Funcional da ESALQ: Luiz Lehmann Coutinho, Ricardo Augusto Brassaloti, Raquel de Lello Rocha Campos Cassano. NGS Soluções Genômicas: Pilar Drummond Sampaio Corrêa Mariani. FZEA-USP Pirassununga: Mirele Daiana Poleti, Jessika Cristina Chagas Lesbon, Elisangela Chicaroni Mattos, Heidge Fukumasu. USP-Botucatu: Rejane Maria Tommasini Grotto, Jayme A. Souza-Neto, Guilherme Targino Valente, Patricia Akemi Assato, Felipe Allan da Silva da Costa, Bianca Cechetto Carlos. Mendelics: Bibiana Santos, João Paulo Kitajima, Erika Freitas, David Schlesinger. Hemocentro Ribeirão Preto: Simone Kashima, Evandra Strazza Rodrigues, Svetoslav Nanev Slavov, Elaine Vieira dos Santos, Rafael dos Santos Bezerra, Luiz Carlos Junior de Alcantara, Marta Giovanetti, Vagner Fonseca, Flavia Aburjaile, Rodrigo Tocantins Calado. |
| EPI_ISL_1795116, EPI_ISL_1795117, EPI_ISL_1795118, EPI_ISL_1795119 | UNIDADE MISTA DE LUIZ ANTONIO | Instituto Butantan / ESALQ-Piracicaba | Instituto Butantan: Alexander Roberto Precioso, Dimas Tadeu Covas, Sandra Coccuzzo Sampaio, Maria Carolina Elias, José Salvatore Leister Patané, Vincent Louis Viala, Antonio Jorge Martins, Ricardo Haddad, Claudia Renata dos Santos Barros, Elaine Cristina Marqueze, Raul Machado Neto, Debora Botequio Moretti. Centro de Genômica Funcional da ESALQ: Luiz Lehmann Coutinho, Ricardo Augusto Brassaloti, Raquel de Lello Rocha Campos Cassano. NGS Soluções Genômicas: Pilar Drummond Sampaio Corrêa Mariani. FZEA-USP Pirassununga: Mirele Daiana Poleti, Jessika Cristina Chagas Lesbon, Elisangela Chicaroni Mattos, Heidge Fukumasu. USP-Botucatu: Rejane Maria Tommasini Grotto, Jayme A. Souza-Neto, Guilherme Targino Valente, Patricia Akemi Assato, Felipe Allan da Silva da Costa, Bianca Cechetto Carlos. Mendelics: Bibiana Santos, João Paulo Kitajima, Erika Freitas, David Schlesinger. Hemocentro Ribeirão Preto: Simone Kashima, Evandra Strazza Rodrigues, Svetoslav Nanev Slavov, Elaine Vieira dos Santos, Rafael dos Santos Bezerra, Luiz Carlos Junior de Alcantara, Marta Giovanetti, Vagner Fonseca, Flavia Aburjaile, Rodrigo Tocantins Calado. |
| EPI_ISL_1795120, EPI_ISL_1795121 | CENTRO DE SAUDE DE CAJOBI | Instituto Butantan / ESALQ-Piracicaba | Instituto Butantan: Alexander Roberto Precioso, Dimas Tadeu Covas, Sandra Coccuzzo Sampaio, Maria Carolina Elias, José Salvatore Leister Patané, Vincent Louis Viala, Antonio Jorge Martins, Ricardo Haddad, Claudia Renata dos Santos Barros, Elaine Cristina Marqueze, Raul Machado Neto, Debora Botequio Moretti. Centro de Genômica Funcional da ESALQ: Luiz Lehmann Coutinho, Ricardo Augusto Brassaloti, Raquel de Lello Rocha Campos Cassano. NGS Soluções Genômicas: Pilar Drummond Sampaio Corrêa Mariani. FZEA-USP Pirassununga: Mirele Daiana Poleti, Jessika Cristina Chagas Lesbon, Elisangela Chicaroni Mattos, Heidge Fukumasu. USP-Botucatu: Rejane Maria Tommasini Grotto, Jayme A. Souza-Neto, Guilherme Targino Valente, Patricia Akemi Assato, Felipe Allan da Silva da Costa, Bianca Cechetto Carlos. Mendelics: Bibiana Santos, João Paulo Kitajima, Erika Freitas, David Schlesinger. Hemocentro Ribeirão Preto: Simone Kashima, Evandra Strazza Rodrigues, Svetoslav Nanev Slavov, Elaine Vieira dos Santos, Rafael dos Santos Bezerra, Luiz Carlos Junior de Alcantara, Marta Giovanetti, Vagner Fonseca, Flavia Aburjaile, Rodrigo Tocantins Calado. |
| EPI_ISL_1795122, EPI_ISL_1795123, EPI_ISL_1795124 | UNIDADE DE VIGILANCIA EM SAUDE | Instituto Butantan / ESALQ-Piracicaba | Instituto Butantan: Alexander Roberto Precioso, Dimas Tadeu Covas, Sandra Coccuzzo Sampaio, Maria Carolina Elias, José Salvatore Leister Patané, Vincent Louis Viala, Antonio Jorge Martins, Ricardo Haddad, Claudia Renata dos Santos Barros, Elaine Cristina Marqueze, Raul Machado Neto, Debora Botequio Moretti. Centro de Genômica Funcional da ESALQ: Luiz Lehmann Coutinho, Ricardo Augusto Brassaloti, Raquel de Lello Rocha Campos Cassano. NGS Soluções Genômicas: Pilar Drummond Sampaio Corrêa Mariani. FZEA-USP Pirassununga: Mirele Daiana Poleti, Jessika Cristina Chagas Lesbon, Elisangela Chicaroni Mattos, Heidge Fukumasu. USP-Botucatu: Rejane Maria Tommasini Grotto, Jayme A. Souza-Neto, Guilherme Targino Valente, Patricia Akemi Assato, Felipe Allan da Silva da Costa, Bianca Cechetto Carlos. Mendelics: Bibiana Santos, João Paulo Kitajima, Erika Freitas, David Schlesinger. Hemocentro Ribeirão Preto: Simone Kashima, Evandra Strazza Rodrigues, Svetoslav Nanev Slavov, Elaine Vieira dos Santos, Rafael dos Santos Bezerra, Luiz Carlos Junior de Alcantara, Marta Giovanetti, Vagner Fonseca, Flavia Aburjaile, Rodrigo Tocantins Calado. |
| EPI_ISL_1795125 | CENTRO DE SAUDE III CANDIDO RODRIGUES | Instituto Butantan / ESALQ-Piracicaba | Instituto Butantan: Alexander Roberto Precioso, Dimas Tadeu Covas, Sandra Coccuzzo Sampaio, Maria Carolina Elias, José Salvatore Leister Patané, Vincent Louis Viala, Antonio Jorge Martins, Ricardo Haddad, Claudia Renata dos Santos Barros, Elaine Cristina Marqueze, Raul Machado Neto, Debora |

| | | | |
|---|---|---|---|
| | | | Botequio Moretti. Centro de Genômica Funcional da ESALQ: Luiz Lehmann Coutinho, Ricardo Augusto Brassaloti, Raquel de Lello Rocha Campos Cassano. NGS Soluções Genômicas: Pilar Drummond Sampaio Corrêa Mariani. FZEA-USP Pirassununga: Mirele Daiana Poleti, Jessika Cristina Chagas Lesbon, Elisangela Chicaroni Mattos, Heidge Fukumasu. USP-Botucatu: Rejane Maria Tommasini Grotto, Jayme A. Souza-Neto, Guilherme Targino Valente, Patricia Akemi Assato, Felipe Allan da Silva da Costa, Bianca Cechetto Carlos. Mendelics: Bibiana Santos, João Paulo Kitajima, Erika Freitas, David Schlesinger. Hemocentro Ribeirão Preto: Simone Kashima, Evandra Strazza Rodrigues, Svetoslav Nanev Slavov, Elaine Vieira dos Santos, Rafael dos Santos Bezerra, Luiz Carlos Junior de Alcantara, Marta Giovanetti, Vagner Fonseca, Flavia Aburjaile, Rodrigo Tocantins Calado. |
| EPI_ISL_1795126 | CENTRO DE SAUDE III SALES OLIVEIRA | Instituto Butantan / ESALQ-Piracicaba | Instituto Butantan: Alexander Roberto Precioso, Dimas Tadeu Covas, Sandra Coccuzzo Sampaio, Maria Carolina Elias, José Salvatore Leister Patané, Vincent Louis Viala, Antonio Jorge Martins, Ricardo Haddad, Claudia Renata dos Santos Barros, Elaine Cristina Marqueze, Raul Machado Neto, Debora Botequio Moretti. Centro de Genômica Funcional da ESALQ: Luiz Lehmann Coutinho, Ricardo Augusto Brassaloti, Raquel de Lello Rocha Campos Cassano. NGS Soluções Genômicas: Pilar Drummond Sampaio Corrêa Mariani. FZEA-USP Pirassununga: Mirele Daiana Poleti, Jessika Cristina Chagas Lesbon, Elisangela Chicaroni Mattos, Heidge Fukumasu. USP-Botucatu: Rejane Maria Tommasini Grotto, Jayme A. Souza-Neto, Guilherme Targino Valente, Patricia Akemi Assato, Felipe Allan da Silva da Costa, Bianca Cechetto Carlos. Mendelics: Bibiana Santos, João Paulo Kitajima, Erika Freitas, David Schlesinger. Hemocentro Ribeirão Preto: Simone Kashima, Evandra Strazza Rodrigues, Svetoslav Nanev Slavov, Elaine Vieira dos Santos, Rafael dos Santos Bezerra, Luiz Carlos Junior de Alcantara, Marta Giovanetti, Vagner Fonseca, Flavia Aburjaile, Rodrigo Tocantins Calado. |
| EPI_ISL_1795127 | CENTRO DE SAUDE III CANDIDO RODRIGUES | Instituto Butantan / ESALQ-Piracicaba | Instituto Butantan: Alexander Roberto Precioso, Dimas Tadeu Covas, Sandra Coccuzzo Sampaio, Maria Carolina Elias, José Salvatore Leister Patané, Vincent Louis Viala, Antonio Jorge Martins, Ricardo Haddad, Claudia Renata dos Santos Barros, Elaine Cristina Marqueze, Raul Machado Neto, Debora Botequio Moretti. Centro de Genômica Funcional da ESALQ: Luiz Lehmann Coutinho, Ricardo Augusto Brassaloti, Raquel de Lello Rocha Campos Cassano. NGS Soluções Genômicas: Pilar Drummond Sampaio Corrêa Mariani. FZEA-USP Pirassununga: Mirele Daiana Poleti, Jessika Cristina Chagas Lesbon, Elisangela Chicaroni Mattos, Heidge Fukumasu. USP-Botucatu: Rejane Maria Tommasini Grotto, Jayme A. Souza-Neto, Guilherme Targino Valente, Patricia Akemi Assato, Felipe Allan da Silva da Costa, Bianca Cechetto Carlos. Mendelics: Bibiana Santos, João Paulo Kitajima, Erika Freitas, David Schlesinger. Hemocentro Ribeirão Preto: Simone Kashima, Evandra Strazza Rodrigues, Svetoslav Nanev Slavov, Elaine Vieira dos Santos, Rafael dos Santos Bezerra, Luiz Carlos Junior de Alcantara, Marta Giovanetti, Vagner Fonseca, Flavia Aburjaile, Rodrigo Tocantins Calado. |
| EPI_ISL_1795128, EPI_ISL_1795129, EPI_ISL_1795130, EPI_ISL_1795131, EPI_ISL_1795132, EPI_ISL_1795133, EPI_ISL_1795134, EPI_ISL_1795135 | UNIDADE DE VIGILANCIA EM SAUDE | Instituto Butantan / ESALQ-Piracicaba | Instituto Butantan: Alexander Roberto Precioso, Dimas Tadeu Covas, Sandra Coccuzzo Sampaio, Maria Carolina Elias, José Salvatore Leister Patané, Vincent Louis Viala, Antonio Jorge Martins, Ricardo Haddad, Claudia Renata dos Santos Barros, Elaine Cristina Marqueze, Raul Machado Neto, Debora Botequio Moretti. Centro de Genômica Funcional da ESALQ: Luiz Lehmann Coutinho, Ricardo Augusto Brassaloti, Raquel de Lello Rocha Campos Cassano. NGS Soluções Genômicas: Pilar Drummond Sampaio Corrêa Mariani. FZEA-USP Pirassununga: Mirele Daiana Poleti, Jessika Cristina Chagas Lesbon, Elisangela Chicaroni Mattos, Heidge Fukumasu. USP-Botucatu: Rejane Maria Tommasini Grotto, Jayme A. Souza-Neto, Guilherme Targino Valente, Patricia Akemi Assato, Felipe Allan da Silva da Costa, Bianca Cechetto Carlos. Mendelics: Bibiana Santos, João Paulo Kitajima, Erika Freitas, David Schlesinger. Hemocentro Ribeirão Preto: Simone Kashima, Evandra Strazza Rodrigues, Svetoslav Nanev Slavov, Elaine Vieira dos Santos, Rafael dos Santos Bezerra, Luiz Carlos Junior de Alcantara, Marta Giovanetti, Vagner Fonseca, Flavia Aburjaile, Rodrigo Tocantins Calado. |
| EPI_ISL_1795136, EPI_ISL_1795137, EPI_ISL_1795138, EPI_ISL_1795139, EPI_ISL_1795140 | UNIDADE MISTA DE VISTA ALEGRE DO ALTO VISTA ALEGRE DO ALTO | Instituto Butantan / ESALQ-Piracicaba | Instituto Butantan: Alexander Roberto Precioso, Dimas Tadeu Covas, Sandra Coccuzzo Sampaio, Maria Carolina Elias, José Salvatore Leister Patané, Vincent Louis Viala, Antonio Jorge Martins, Ricardo Haddad, Claudia Renata dos Santos Barros, Elaine Cristina Marqueze, Raul Machado Neto, Debora Botequio Moretti. Centro de Genômica Funcional da ESALQ: Luiz Lehmann Coutinho, Ricardo Augusto Brassaloti, Raquel de Lello Rocha Campos Cassano. NGS Soluções Genômicas: Pilar Drummond Sampaio Corrêa Mariani. FZEA-USP Pirassununga: Mirele Daiana Poleti, Jessika Cristina Chagas Lesbon, Elisangela Chicaroni Mattos, Heidge Fukumasu. USP-Botucatu: Rejane Maria Tommasini Grotto, Jayme A. Souza-Neto, Guilherme Targino Valente, Patricia Akemi Assato, Felipe Allan da Silva da Costa, Bianca Cechetto Carlos. Mendelics: Bibiana Santos, João Paulo Kitajima, Erika Freitas, David Schlesinger. Hemocentro Ribeirão Preto: Simone Kashima, Evandra Strazza Rodrigues, Svetoslav Nanev Slavov, Elaine Vieira dos Santos, Rafael dos Santos Bezerra, Luiz Carlos Junior de Alcantara, Marta Giovanetti, Vagner Fonseca, Flavia Aburjaile, Rodrigo Tocantins Calado. |
| EPI_ISL_1795141, EPI_ISL_1795142, EPI_ISL_1795143, EPI_ISL_1795144, EPI_ISL_1795145, EPI_ISL_1795146, EPI_ISL_1795147, EPI_ISL_1795148 | POLICLINICA HORTOLANDIA | Instituto Butantan / ESALQ-Piracicaba | Instituto Butantan: Alexander Roberto Precioso, Dimas Tadeu Covas, Sandra Coccuzzo Sampaio, Maria Carolina Elias, José Salvatore Leister Patané, Vincent Louis Viala, Antonio Jorge Martins, Ricardo Haddad, Claudia Renata dos Santos Barros, Elaine Cristina Marqueze, Raul Machado Neto, Debora Botequio Moretti. Centro de Genômica Funcional da ESALQ: Luiz Lehmann Coutinho, Ricardo Augusto Brassaloti, Raquel de Lello Rocha Campos Cassano. NGS Soluções Genômicas: Pilar Drummond Sampaio Corrêa Mariani. FZEA-USP Pirassununga: Mirele Daiana Poleti, Jessika Cristina Chagas Lesbon, Elisangela Chicaroni Mattos, Heidge Fukumasu. USP-Botucatu: Rejane Maria Tommasini Grotto, Jayme A. Souza-Neto, Guilherme Targino Valente, Patricia Akemi Assato, Felipe Allan da Silva da Costa, Bianca Cechetto Carlos. Mendelics: Bibiana Santos, João Paulo Kitajima, Erika Freitas, David Schlesinger. Hemocentro Ribeirão Preto: Simone Kashima, Evandra Strazza Rodrigues, Svetoslav Nanev Slavov, Elaine Vieira dos Santos, Rafael dos Santos Bezerra, Luiz Carlos Junior de Alcantara, Marta Giovanetti, Vagner Fonseca, Flavia Aburjaile, Rodrigo Tocantins Calado. |
| EPI_ISL_1795149, EPI_ISL_1795150 | UNIDADE MISTA DE VISTA ALEGRE DO ALTO VISTA ALEGRE DO ALTO | Instituto Butantan / ESALQ-Piracicaba | Instituto Butantan: Alexander Roberto Precioso, Dimas Tadeu Covas, Sandra Coccuzzo Sampaio, Maria Carolina Elias, José Salvatore Leister Patané, Vincent Louis Viala, Antonio Jorge Martins, Ricardo Haddad, Claudia Renata dos Santos Barros, Elaine Cristina Marqueze, Raul Machado Neto, Debora Botequio Moretti. Centro de Genômica Funcional da ESALQ: Luiz Lehmann Coutinho, Ricardo Augusto Brassaloti, Raquel de Lello Rocha Campos Cassano. NGS Soluções Genômicas: Pilar Drummond Sampaio Corrêa Mariani. FZEA-USP Pirassununga: Mirele Daiana Poleti, Jessika Cristina Chagas Lesbon, Elisangela Chicaroni Mattos, Heidge Fukumasu. USP-Botucatu: Rejane Maria Tommasini Grotto, Jayme A. Souza-Neto, Guilherme Targino Valente, Patricia Akemi Assato, Felipe Allan da Silva da Costa, Bianca Cechetto Carlos. Mendelics: Bibiana Santos, João Paulo Kitajima, Erika Freitas, David Schlesinger. Hemocentro Ribeirão Preto: Simone Kashima, Evandra Strazza Rodrigues, Svetoslav Nanev Slavov, Elaine Vieira dos Santos, Rafael dos Santos Bezerra, Luiz Carlos Junior de Alcantara, Marta Giovanetti, Vagner Fonseca, Flavia Aburjaile, Rodrigo Tocantins Calado. |
| EPI_ISL_1795151 | CENTRO DE SAUDE DE CAJOBI | Instituto Butantan / ESALQ-Piracicaba | Instituto Butantan: Alexander Roberto Precioso, Dimas Tadeu Covas, Sandra Coccuzzo Sampaio, Maria Carolina Elias, José Salvatore Leister Patané, Vincent Louis Viala, Antonio Jorge Martins, Ricardo Haddad, Claudia Renata dos Santos Barros, Elaine Cristina Marqueze, Raul Machado Neto, Debora Botequio Moretti. Centro de Genômica Funcional da ESALQ: Luiz Lehmann Coutinho, Ricardo Augusto Brassaloti, Raquel de Lello Rocha Campos Cassano. NGS Soluções Genômicas: Pilar Drummond Sampaio Corrêa Mariani. FZEA-USP Pirassununga: Mirele Daiana Poleti, Jessika Cristina Chagas Lesbon, Elisangela Chicaroni Mattos, Heidge Fukumasu. USP-Botucatu: Rejane Maria Tommasini Grotto, Jayme A. Souza-Neto, Guilherme Targino Valente, Patricia Akemi Assato, Felipe Allan da Silva da Costa, Bianca Cechetto Carlos. Mendelics: Bibiana Santos, João Paulo Kitajima, Erika Freitas, David Schlesinger. Hemocentro Ribeirão Preto: Simone Kashima, Evandra Strazza Rodrigues, Svetoslav Nanev Slavov, Elaine Vieira dos Santos, Rafael dos Santos Bezerra, Luiz Carlos Junior de Alcantara, Marta Giovanetti, Vagner Fonseca, Flavia Aburjaile, Rodrigo Tocantins Calado. |
| EPI_ISL_1795152, EPI_ISL_1795153, EPI_ISL_1795154, EPI_ISL_1795155, EPI_ISL_1795156, EPI_ISL_1795157, EPI_ISL_1795158, EPI_ISL_1795159 | POLICLINICA HORTOLANDIA | Instituto Butantan / ESALQ-Piracicaba | Instituto Butantan: Alexander Roberto Precioso, Dimas Tadeu Covas, Sandra Coccuzzo Sampaio, Maria Carolina Elias, José Salvatore Leister Patané, Vincent Louis Viala, Antonio Jorge Martins, Ricardo Haddad, Claudia Renata dos Santos Barros, Elaine Cristina Marqueze, Raul Machado Neto, Debora Botequio Moretti. Centro de Genômica Funcional da ESALQ: Luiz Lehmann Coutinho, Ricardo Augusto Brassaloti, Raquel de Lello Rocha Campos Cassano. NGS Soluções Genômicas: Pilar Drummond Sampaio Corrêa Mariani. FZEA-USP Pirassununga: Mirele Daiana Poleti, Jessika Cristina Chagas Lesbon, Elisangela Chicaroni Mattos, Heidge Fukumasu. USP-Botucatu: Rejane Maria Tommasini Grotto, Jayme A. Souza-Neto, Guilherme Targino Valente, Patricia Akemi Assato, Felipe Allan da Silva da Costa, Bianca Cechetto Carlos. Mendelics: Bibiana Santos, João Paulo Kitajima, Erika Freitas, David Schlesinger. Hemocentro Ribeirão Preto: Simone Kashima, Evandra Strazza Rodrigues, Svetoslav Nanev Slavov, Elaine Vieira dos Santos, Rafael dos Santos Bezerra, Luiz Carlos Junior de Alcantara, Marta Giovanetti, Vagner Fonseca, Flavia Aburjaile, Rodrigo Tocantins Calado. |
| EPI_ISL_1795160 | UBS JARDIM ITAMARATY | Instituto Butantan / ESALQ-Piracicaba | Instituto Butantan: Alexander Roberto Precioso, Dimas Tadeu Covas, Sandra Coccuzzo Sampaio, Maria Carolina Elias, José Salvatore Leister Patané, Vincent Louis Viala, Antonio Jorge Martins, Ricardo Haddad, Claudia Renata dos Santos Barros, Elaine Cristina Marqueze, Raul Machado Neto, Debora Botequio Moretti. Centro de Genômica Funcional da ESALQ: Luiz Lehmann Coutinho, Ricardo Augusto Brassaloti, Raquel de Lello Rocha Campos Cassano. NGS Soluções Genômicas: Pilar Drummond Sampaio Corrêa Mariani. FZEA-USP Pirassununga: Mirele Daiana Poleti, Jessika Cristina Chagas Lesbon, Elisangela Chicaroni Mattos, Heidge Fukumasu. USP-Botucatu: Rejane Maria Tommasini Grotto, Jayme A. Souza-Neto, Guilherme Targino Valente, Patricia Akemi Assato, Felipe Allan da Silva da Costa, Bianca Cechetto Carlos. Mendelics: Bibiana Santos, João Paulo Kitajima, Erika Freitas, David Schlesinger. Hemocentro Ribeirão Preto: Simone Kashima, Evandra Strazza Rodrigues, Svetoslav Nanev Slavov, Elaine Vieira dos Santos, Rafael dos Santos Bezerra, Luiz Carlos Junior de Alcantara, Marta Giovanetti, Vagner Fonseca, Flavia Aburjaile, Rodrigo Tocantins Calado. |
| EPI_ISL_1795161 | CS III VILA ODILON | Instituto Butantan / ESALQ-Piracicaba | Instituto Butantan: Alexander Roberto Precioso, Dimas Tadeu Covas, Sandra Coccuzzo Sampaio, Maria Carolina Elias, José Salvatore Leister Patané, Vincent Louis Viala, Antonio Jorge Martins, Ricardo Haddad, Claudia Renata dos Santos Barros, Elaine Cristina Marqueze, Raul Machado Neto, Debora Botequio Moretti. Centro de Genômica Funcional da ESALQ: Luiz Lehmann Coutinho, Ricardo Augusto Brassaloti, Raquel de Lello Rocha Campos Cassano. NGS Soluções Genômicas: Pilar Drummond Sampaio Corrêa Mariani. FZEA-USP Pirassununga: Mirele Daiana Poleti, Jessika Cristina Chagas |

| | | | |
|---|---|---|---|
| | | | Lesbon, Elisangela Chicaroni Mattos, Heidge Fukumasu. USP-Botucatu: Rejane Maria Tommasini Grotto, Jayme A. Souza-Neto, Guilherme Targino Valente, Patricia Akemi Assato, Felipe Allan da Silva da Costa, Bianca Cechetto Carlos. Mendelics: Bibiana Santos, João Paulo Kitajima, Erika Freitas, David Schlesinger. Hemocentro Ribeirão Preto: Simone Kashima, Evandra Strazza Rodrigues, Svetoslav Nanev Slavov, Elaine Vieira dos Santos, Rafael dos Santos Bezerra, Luiz Carlos Junior de Alcantara, Marta Giovanetti, Vagner Fonseca, Flavia Aburjaile, Rodrigo Tocantins Calado. |
| EPI_ISL_1795162, EPI_ISL_1795163, EPI_ISL_1795164 | CENTRO DE SAUDE DR PLINIO ALBERS DE GALIA | Instituto Butantan / ESALQ-Piracicaba | Instituto Butantan: Alexander Roberto Precioso, Dimas Tadeu Covas, Sandra Coccuzzo Sampaio, Maria Carolina Elias, José Salvatore Leister Patané, Vincent Louis Viala, Antonio Jorge Martins, Ricardo Haddad, Claudia Renata dos Santos Barros, Elaine Cristina Marqueze, Raul Machado Neto, Debora Botequio Moretti. Centro de Genômica Funcional da ESALQ: Luiz Lehmann Coutinho, Ricardo Augusto Brassaloti, Raquel de Lello Rocha Campos Cassano. NGS Soluções Genômicas: Pilar Drummond Sampaio Corrêa Mariani. FZEA-USP Pirassununga: Mirele Daiana Poleti, Jessika Cristina Chagas Lesbon, Elisangela Chicaroni Mattos, Heidge Fukumasu. USP-Botucatu: Rejane Maria Tommasini Grotto, Jayme A. Souza-Neto, Guilherme Targino Valente, Patricia Akemi Assato, Felipe Allan da Silva da Costa, Bianca Cechetto Carlos. Mendelics: Bibiana Santos, João Paulo Kitajima, Erika Freitas, David Schlesinger. Hemocentro Ribeirão Preto: Simone Kashima, Evandra Strazza Rodrigues, Svetoslav Nanev Slavov, Elaine Vieira dos Santos, Rafael dos Santos Bezerra, Luiz Carlos Junior de Alcantara, Marta Giovanetti, Vagner Fonseca, Flavia Aburjaile, Rodrigo Tocantins Calado. |
| EPI_ISL_1795165 | CS DE ALVARO CARVALHO | Instituto Butantan / ESALQ-Piracicaba | Instituto Butantan: Alexander Roberto Precioso, Dimas Tadeu Covas, Sandra Coccuzzo Sampaio, Maria Carolina Elias, José Salvatore Leister Patané, Vincent Louis Viala, Antonio Jorge Martins, Ricardo Haddad, Claudia Renata dos Santos Barros, Elaine Cristina Marqueze, Raul Machado Neto, Debora Botequio Moretti. Centro de Genômica Funcional da ESALQ: Luiz Lehmann Coutinho, Ricardo Augusto Brassaloti, Raquel de Lello Rocha Campos Cassano. NGS Soluções Genômicas: Pilar Drummond Sampaio Corrêa Mariani. FZEA-USP Pirassununga: Mirele Daiana Poleti, Jessika Cristina Chagas Lesbon, Elisangela Chicaroni Mattos, Heidge Fukumasu. USP-Botucatu: Rejane Maria Tommasini Grotto, Jayme A. Souza-Neto, Guilherme Targino Valente, Patricia Akemi Assato, Felipe Allan da Silva da Costa, Bianca Cechetto Carlos. Mendelics: Bibiana Santos, João Paulo Kitajima, Erika Freitas, David Schlesinger. Hemocentro Ribeirão Preto: Simone Kashima, Evandra Strazza Rodrigues, Svetoslav Nanev Slavov, Elaine Vieira dos Santos, Rafael dos Santos Bezerra, Luiz Carlos Junior de Alcantara, Marta Giovanetti, Vagner Fonseca, Flavia Aburjaile, Rodrigo Tocantins Calado. |
| EPI_ISL_1795166 | CENTRO DE SAUDE DR PLINIO ALBERS DE GALIA | Instituto Butantan / ESALQ-Piracicaba | Instituto Butantan: Alexander Roberto Precioso, Dimas Tadeu Covas, Sandra Coccuzzo Sampaio, Maria Carolina Elias, José Salvatore Leister Patané, Vincent Louis Viala, Antonio Jorge Martins, Ricardo Haddad, Claudia Renata dos Santos Barros, Elaine Cristina Marqueze, Raul Machado Neto, Debora Botequio Moretti. Centro de Genômica Funcional da ESALQ: Luiz Lehmann Coutinho, Ricardo Augusto Brassaloti, Raquel de Lello Rocha Campos Cassano. NGS Soluções Genômicas: Pilar Drummond Sampaio Corrêa Mariani. FZEA-USP Pirassununga: Mirele Daiana Poleti, Jessika Cristina Chagas Lesbon, Elisangela Chicaroni Mattos, Heidge Fukumasu. USP-Botucatu: Rejane Maria Tommasini Grotto, Jayme A. Souza-Neto, Guilherme Targino Valente, Patricia Akemi Assato, Felipe Allan da Silva da Costa, Bianca Cechetto Carlos. Mendelics: Bibiana Santos, João Paulo Kitajima, Erika Freitas, David Schlesinger. Hemocentro Ribeirão Preto: Simone Kashima, Evandra Strazza Rodrigues, Svetoslav Nanev Slavov, Elaine Vieira dos Santos, Rafael dos Santos Bezerra, Luiz Carlos Junior de Alcantara, Marta Giovanetti, Vagner Fonseca, Flavia Aburjaile, Rodrigo Tocantins Calado. |
| EPI_ISL_1795167, EPI_ISL_1795168, EPI_ISL_1795169 | CENTRO DE SAUDE DE FLORINEA | Instituto Butantan / ESALQ-Piracicaba | Instituto Butantan: Alexander Roberto Precioso, Dimas Tadeu Covas, Sandra Coccuzzo Sampaio, Maria Carolina Elias, José Salvatore Leister Patané, Vincent Louis Viala, Antonio Jorge Martins, Ricardo Haddad, Claudia Renata dos Santos Barros, Elaine Cristina Marqueze, Raul Machado Neto, Debora Botequio Moretti. Centro de Genômica Funcional da ESALQ: Luiz Lehmann Coutinho, Ricardo Augusto Brassaloti, Raquel de Lello Rocha Campos Cassano. NGS Soluções Genômicas: Pilar Drummond Sampaio Corrêa Mariani. FZEA-USP Pirassununga: Mirele Daiana Poleti, Jessika Cristina Chagas Lesbon, Elisangela Chicaroni Mattos, Heidge Fukumasu. USP-Botucatu: Rejane Maria Tommasini Grotto, Jayme A. Souza-Neto, Guilherme Targino Valente, Patricia Akemi Assato, Felipe Allan da Silva da Costa, Bianca Cechetto Carlos. Mendelics: Bibiana Santos, João Paulo Kitajima, Erika Freitas, David Schlesinger. Hemocentro Ribeirão Preto: Simone Kashima, Evandra Strazza Rodrigues, Svetoslav Nanev Slavov, Elaine Vieira dos Santos, Rafael dos Santos Bezerra, Luiz Carlos Junior de Alcantara, Marta Giovanetti, Vagner Fonseca, Flavia Aburjaile, Rodrigo Tocantins Calado. |
| EPI_ISL_1795170 | CENTRO DE SAUDE DE MARACAI | Instituto Butantan / ESALQ-Piracicaba | Instituto Butantan: Alexander Roberto Precioso, Dimas Tadeu Covas, Sandra Coccuzzo Sampaio, Maria Carolina Elias, José Salvatore Leister Patané, Vincent Louis Viala, Antonio Jorge Martins, Ricardo Haddad, Claudia Renata dos Santos Barros, Elaine Cristina Marqueze, Raul Machado Neto, Debora Botequio Moretti. Centro de Genômica Funcional da ESALQ: Luiz Lehmann Coutinho, Ricardo Augusto Brassaloti, Raquel de Lello Rocha Campos Cassano. NGS Soluções Genômicas: Pilar Drummond Sampaio Corrêa Mariani. FZEA-USP Pirassununga: Mirele Daiana Poleti, Jessika Cristina Chagas Lesbon, Elisangela Chicaroni Mattos, Heidge Fukumasu. USP-Botucatu: Rejane Maria Tommasini Grotto, Jayme A. Souza-Neto, Guilherme Targino Valente, Patricia Akemi Assato, Felipe Allan da Silva da Costa, Bianca Cechetto Carlos. Mendelics: Bibiana Santos, João Paulo Kitajima, Erika Freitas, David Schlesinger. Hemocentro Ribeirão Preto: Simone Kashima, Evandra Strazza Rodrigues, Svetoslav Nanev Slavov, Elaine Vieira dos Santos, Rafael dos Santos Bezerra, Luiz Carlos Junior de Alcantara, Marta Giovanetti, Vagner Fonseca, Flavia Aburjaile, Rodrigo Tocantins Calado. |
| EPI_ISL_1795171 | CS DE OSCAR BRESSANE PSF | Instituto Butantan / ESALQ-Piracicaba | Instituto Butantan: Alexander Roberto Precioso, Dimas Tadeu Covas, Sandra Coccuzzo Sampaio, Maria Carolina Elias, José Salvatore Leister Patané, Vincent Louis Viala, Antonio Jorge Martins, Ricardo Haddad, Claudia Renata dos Santos Barros, Elaine Cristina Marqueze, Raul Machado Neto, Debora Botequio Moretti. Centro de Genômica Funcional da ESALQ: Luiz Lehmann Coutinho, Ricardo Augusto Brassaloti, Raquel de Lello Rocha Campos Cassano. NGS Soluções Genômicas: Pilar Drummond Sampaio Corrêa Mariani. FZEA-USP Pirassununga: Mirele Daiana Poleti, Jessika Cristina Chagas Lesbon, Elisangela Chicaroni Mattos, Heidge Fukumasu. USP-Botucatu: Rejane Maria Tommasini Grotto, Jayme A. Souza-Neto, Guilherme Targino Valente, Patricia Akemi Assato, Felipe Allan da Silva da Costa, Bianca Cechetto Carlos. Mendelics: Bibiana Santos, João Paulo Kitajima, Erika Freitas, David Schlesinger. Hemocentro Ribeirão Preto: Simone Kashima, Evandra Strazza Rodrigues, Svetoslav Nanev Slavov, Elaine Vieira dos Santos, Rafael dos Santos Bezerra, Luiz Carlos Junior de Alcantara, Marta Giovanetti, Vagner Fonseca, Flavia Aburjaile, Rodrigo Tocantins Calado. |
| EPI_ISL_1795172 | CENTRO DE SAUDE DE MARACAI | Instituto Butantan / ESALQ-Piracicaba | Instituto Butantan: Alexander Roberto Precioso, Dimas Tadeu Covas, Sandra Coccuzzo Sampaio, Maria Carolina Elias, José Salvatore Leister Patané, Vincent Louis Viala, Antonio Jorge Martins, Ricardo Haddad, Claudia Renata dos Santos Barros, Elaine Cristina Marqueze, Raul Machado Neto, Debora Botequio Moretti. Centro de Genômica Funcional da ESALQ: Luiz Lehmann Coutinho, Ricardo Augusto Brassaloti, Raquel de Lello Rocha Campos Cassano. NGS Soluções Genômicas: Pilar Drummond Sampaio Corrêa Mariani. FZEA-USP Pirassununga: Mirele Daiana Poleti, Jessika Cristina Chagas Lesbon, Elisangela Chicaroni Mattos, Heidge Fukumasu. USP-Botucatu: Rejane Maria Tommasini Grotto, Jayme A. Souza-Neto, Guilherme Targino Valente, Patricia Akemi Assato, Felipe Allan da Silva da Costa, Bianca Cechetto Carlos. Mendelics: Bibiana Santos, João Paulo Kitajima, Erika Freitas, David Schlesinger. Hemocentro Ribeirão Preto: Simone Kashima, Evandra Strazza Rodrigues, Svetoslav Nanev Slavov, Elaine Vieira dos Santos, Rafael dos Santos Bezerra, Luiz Carlos Junior de Alcantara, Marta Giovanetti, Vagner Fonseca, Flavia Aburjaile, Rodrigo Tocantins Calado. |
| EPI_ISL_1795173 | CS DE OSCAR BRESSANE PSF | Instituto Butantan / ESALQ-Piracicaba | Instituto Butantan: Alexander Roberto Precioso, Dimas Tadeu Covas, Sandra Coccuzzo Sampaio, Maria Carolina Elias, José Salvatore Leister Patané, Vincent Louis Viala, Antonio Jorge Martins, Ricardo Haddad, Claudia Renata dos Santos Barros, Elaine Cristina Marqueze, Raul Machado Neto, Debora Botequio Moretti. Centro de Genômica Funcional da ESALQ: Luiz Lehmann Coutinho, Ricardo Augusto Brassaloti, Raquel de Lello Rocha Campos Cassano. NGS Soluções Genômicas: Pilar Drummond Sampaio Corrêa Mariani. FZEA-USP Pirassununga: Mirele Daiana Poleti, Jessika Cristina Chagas Lesbon, Elisangela Chicaroni Mattos, Heidge Fukumasu. USP-Botucatu: Rejane Maria Tommasini Grotto, Jayme A. Souza-Neto, Guilherme Targino Valente, Patricia Akemi Assato, Felipe Allan da Silva da Costa, Bianca Cechetto Carlos. Mendelics: Bibiana Santos, João Paulo Kitajima, Erika Freitas, David Schlesinger. Hemocentro Ribeirão Preto: Simone Kashima, Evandra Strazza Rodrigues, Svetoslav Nanev Slavov, Elaine Vieira dos Santos, Rafael dos Santos Bezerra, Luiz Carlos Junior de Alcantara, Marta Giovanetti, Vagner Fonseca, Flavia Aburjaile, Rodrigo Tocantins Calado. |
| EPI_ISL_1795174, EPI_ISL_1795175, EPI_ISL_1795176 | USF SALERNO | Instituto Butantan / ESALQ-Piracicaba | Instituto Butantan: Alexander Roberto Precioso, Dimas Tadeu Covas, Sandra Coccuzzo Sampaio, Maria Carolina Elias, José Salvatore Leister Patané, Vincent Louis Viala, Antonio Jorge Martins, Ricardo Haddad, Claudia Renata dos Santos Barros, Elaine Cristina Marqueze, Raul Machado Neto, Debora Botequio Moretti. Centro de Genômica Funcional da ESALQ: Luiz Lehmann Coutinho, Ricardo Augusto Brassaloti, Raquel de Lello Rocha Campos Cassano. NGS Soluções Genômicas: Pilar Drummond Sampaio Corrêa Mariani. FZEA-USP Pirassununga: Mirele Daiana Poleti, Jessika Cristina Chagas Lesbon, Elisangela Chicaroni Mattos, Heidge Fukumasu. USP-Botucatu: Rejane Maria Tommasini Grotto, Jayme A. Souza-Neto, Guilherme Targino Valente, Patricia Akemi Assato, Felipe Allan da Silva da Costa, Bianca Cechetto Carlos. Mendelics: Bibiana Santos, João Paulo Kitajima, Erika Freitas, David Schlesinger. Hemocentro Ribeirão Preto: Simone Kashima, Evandra Strazza Rodrigues, Svetoslav Nanev Slavov, Elaine Vieira dos Santos, Rafael dos Santos Bezerra, Luiz Carlos Junior de Alcantara, Marta Giovanetti, Vagner Fonseca, Flavia Aburjaile, Rodrigo Tocantins Calado. |
| EPI_ISL_1795177, EPI_ISL_1795178, EPI_ISL_1795179, EPI_ISL_1795180, EPI_ISL_1795181 | POLICLINICA HORTOLANDIA | Instituto Butantan / ESALQ-Piracicaba | Instituto Butantan: Alexander Roberto Precioso, Dimas Tadeu Covas, Sandra Coccuzzo Sampaio, Maria Carolina Elias, José Salvatore Leister Patané, Vincent Louis Viala, Antonio Jorge Martins, Ricardo Haddad, Claudia Renata dos Santos Barros, Elaine Cristina Marqueze, Raul Machado Neto, Debora Botequio Moretti. Centro de Genômica Funcional da ESALQ: Luiz Lehmann Coutinho, Ricardo Augusto Brassaloti, Raquel de Lello Rocha Campos Cassano. NGS Soluções Genômicas: Pilar Drummond Sampaio Corrêa Mariani. FZEA-USP Pirassununga: Mirele Daiana Poleti, Jessika Cristina Chagas Lesbon, Elisangela Chicaroni Mattos, Heidge Fukumasu. USP-Botucatu: Rejane Maria Tommasini Grotto, Jayme A. Souza-Neto, Guilherme Targino Valente, Patricia Akemi Assato, Felipe Allan da Silva da Costa, Bianca Cechetto Carlos. Mendelics: Bibiana Santos, João Paulo Kitajima, Erika Freitas, |

| | | | |
|---|---|---|---|
| | | | David Schlesinger. Hemocentro Ribeirão Preto: Simone Kashima, Evandra Strazza Rodrigues, Svetoslav Nanev Slavov, Elaine Vieira dos Santos, Rafael dos Santos Bezerra, Luiz Carlos Junior de Alcantara, Marta Giovanetti, Vagner Fonseca, Flavia Aburjaile, Rodrigo Tocantins Calado. |
| EPI_ISL_1795182 | DEPARTAMENTO DE SAUDE MUNICIPAL SOCORRO SP | Instituto Butantan / ESALQ-Piracicaba | Instituto Butantan: Alexander Roberto Precioso, Dimas Tadeu Covas, Sandra Coccuzzo Sampaio, Maria Carolina Elias, José Salvatore Leister Patané, Vincent Louis Viala, Antonio Jorge Martins, Ricardo Haddad, Claudia Renata dos Santos Barros, Elaine Cristina Marqueze, Raul Machado Neto, Debora Botequio Moretti. Centro de Genômica Funcional da ESALQ: Luiz Lehmann Coutinho, Ricardo Augusto Brassaloti, Raquel de Lello Rocha Campos Cassano. NGS Soluções Genômicas: Pilar Drummond Sampaio Corrêa Mariani. FZEA-USP Pirassununga: Mirele Daiana Poleti, Jessika Cristina Chagas Lesbon, Elisangela Chicaroni Mattos, Heidge Fukumasu. USP-Botucatu: Rejane Maria Tommasini Grotto, Jayme A. Souza-Neto, Guilherme Targino Valente, Patricia Akemi Assato, Felipe Allan da Silva da Costa, Bianca Cechetto Carlos. Mendelics: Bibiana Santos, João Paulo Kitajima, Erika Freitas, David Schlesinger. Hemocentro Ribeirão Preto: Simone Kashima, Evandra Strazza Rodrigues, Svetoslav Nanev Slavov, Elaine Vieira dos Santos, Rafael dos Santos Bezerra, Luiz Carlos Junior de Alcantara, Marta Giovanetti, Vagner Fonseca, Flavia Aburjaile, Rodrigo Tocantins Calado. |
| EPI_ISL_1795183, EPI_ISL_1795184, EPI_ISL_1795185, EPI_ISL_1795186 | POLICLINICA HORTOLANDIA | Instituto Butantan / ESALQ-Piracicaba | Instituto Butantan: Alexander Roberto Precioso, Dimas Tadeu Covas, Sandra Coccuzzo Sampaio, Maria Carolina Elias, José Salvatore Leister Patané, Vincent Louis Viala, Antonio Jorge Martins, Ricardo Haddad, Claudia Renata dos Santos Barros, Elaine Cristina Marqueze, Raul Machado Neto, Debora Botequio Moretti. Centro de Genômica Funcional da ESALQ: Luiz Lehmann Coutinho, Ricardo Augusto Brassaloti, Raquel de Lello Rocha Campos Cassano. NGS Soluções Genômicas: Pilar Drummond Sampaio Corrêa Mariani. FZEA-USP Pirassununga: Mirele Daiana Poleti, Jessika Cristina Chagas Lesbon, Elisangela Chicaroni Mattos, Heidge Fukumasu. USP-Botucatu: Rejane Maria Tommasini Grotto, Jayme A. Souza-Neto, Guilherme Targino Valente, Patricia Akemi Assato, Felipe Allan da Silva da Costa, Bianca Cechetto Carlos. Mendelics: Bibiana Santos, João Paulo Kitajima, Erika Freitas, David Schlesinger. Hemocentro Ribeirão Preto: Simone Kashima, Evandra Strazza Rodrigues, Svetoslav Nanev Slavov, Elaine Vieira dos Santos, Rafael dos Santos Bezerra, Luiz Carlos Junior de Alcantara, Marta Giovanetti, Vagner Fonseca, Flavia Aburjaile, Rodrigo Tocantins Calado. |
| EPI_ISL_1795187 | USF SALERNO | Instituto Butantan / ESALQ-Piracicaba | Instituto Butantan: Alexander Roberto Precioso, Dimas Tadeu Covas, Sandra Coccuzzo Sampaio, Maria Carolina Elias, José Salvatore Leister Patané, Vincent Louis Viala, Antonio Jorge Martins, Ricardo Haddad, Claudia Renata dos Santos Barros, Elaine Cristina Marqueze, Raul Machado Neto, Debora Botequio Moretti. Centro de Genômica Funcional da ESALQ: Luiz Lehmann Coutinho, Ricardo Augusto Brassaloti, Raquel de Lello Rocha Campos Cassano. NGS Soluções Genômicas: Pilar Drummond Sampaio Corrêa Mariani. FZEA-USP Pirassununga: Mirele Daiana Poleti, Jessika Cristina Chagas Lesbon, Elisangela Chicaroni Mattos, Heidge Fukumasu. USP-Botucatu: Rejane Maria Tommasini Grotto, Jayme A. Souza-Neto, Guilherme Targino Valente, Patricia Akemi Assato, Felipe Allan da Silva da Costa, Bianca Cechetto Carlos. Mendelics: Bibiana Santos, João Paulo Kitajima, Erika Freitas, David Schlesinger. Hemocentro Ribeirão Preto: Simone Kashima, Evandra Strazza Rodrigues, Svetoslav Nanev Slavov, Elaine Vieira dos Santos, Rafael dos Santos Bezerra, Luiz Carlos Junior de Alcantara, Marta Giovanetti, Vagner Fonseca, Flavia Aburjaile, Rodrigo Tocantins Calado. |
| EPI_ISL_1795188 | POLICLINICA HORTOLANDIA | Instituto Butantan / ESALQ-Piracicaba | Instituto Butantan: Alexander Roberto Precioso, Dimas Tadeu Covas, Sandra Coccuzzo Sampaio, Maria Carolina Elias, José Salvatore Leister Patané, Vincent Louis Viala, Antonio Jorge Martins, Ricardo Haddad, Claudia Renata dos Santos Barros, Elaine Cristina Marqueze, Raul Machado Neto, Debora Botequio Moretti. Centro de Genômica Funcional da ESALQ: Luiz Lehmann Coutinho, Ricardo Augusto Brassaloti, Raquel de Lello Rocha Campos Cassano. NGS Soluções Genômicas: Pilar Drummond Sampaio Corrêa Mariani. FZEA-USP Pirassununga: Mirele Daiana Poleti, Jessika Cristina Chagas Lesbon, Elisangela Chicaroni Mattos, Heidge Fukumasu. USP-Botucatu: Rejane Maria Tommasini Grotto, Jayme A. Souza-Neto, Guilherme Targino Valente, Patricia Akemi Assato, Felipe Allan da Silva da Costa, Bianca Cechetto Carlos. Mendelics: Bibiana Santos, João Paulo Kitajima, Erika Freitas, David Schlesinger. Hemocentro Ribeirão Preto: Simone Kashima, Evandra Strazza Rodrigues, Svetoslav Nanev Slavov, Elaine Vieira dos Santos, Rafael dos Santos Bezerra, Luiz Carlos Junior de Alcantara, Marta Giovanetti, Vagner Fonseca, Flavia Aburjaile, Rodrigo Tocantins Calado. |
| EPI_ISL_1795189 | USF SALERNO | Instituto Butantan / ESALQ-Piracicaba | Instituto Butantan: Alexander Roberto Precioso, Dimas Tadeu Covas, Sandra Coccuzzo Sampaio, Maria Carolina Elias, José Salvatore Leister Patané, Vincent Louis Viala, Antonio Jorge Martins, Ricardo Haddad, Claudia Renata dos Santos Barros, Elaine Cristina Marqueze, Raul Machado Neto, Debora Botequio Moretti. Centro de Genômica Funcional da ESALQ: Luiz Lehmann Coutinho, Ricardo Augusto Brassaloti, Raquel de Lello Rocha Campos Cassano. NGS Soluções Genômicas: Pilar Drummond Sampaio Corrêa Mariani. FZEA-USP Pirassununga: Mirele Daiana Poleti, Jessika Cristina Chagas Lesbon, Elisangela Chicaroni Mattos, Heidge Fukumasu. USP-Botucatu: Rejane Maria Tommasini Grotto, Jayme A. Souza-Neto, Guilherme Targino Valente, Patricia Akemi Assato, Felipe Allan da Silva da Costa, Bianca Cechetto Carlos. Mendelics: Bibiana Santos, João Paulo Kitajima, Erika Freitas, David Schlesinger. Hemocentro Ribeirão Preto: Simone Kashima, Evandra Strazza Rodrigues, Svetoslav Nanev Slavov, Elaine Vieira dos Santos, Rafael dos Santos Bezerra, Luiz Carlos Junior de Alcantara, Marta Giovanetti, Vagner Fonseca, Flavia Aburjaile, Rodrigo Tocantins Calado. |
| EPI_ISL_1795190 | POLICLINICA HORTOLANDIA | Instituto Butantan / ESALQ-Piracicaba | Instituto Butantan: Alexander Roberto Precioso, Dimas Tadeu Covas, Sandra Coccuzzo Sampaio, Maria Carolina Elias, José Salvatore Leister Patané, Vincent Louis Viala, Antonio Jorge Martins, Ricardo Haddad, Claudia Renata dos Santos Barros, Elaine Cristina Marqueze, Raul Machado Neto, Debora Botequio Moretti. Centro de Genômica Funcional da ESALQ: Luiz Lehmann Coutinho, Ricardo Augusto Brassaloti, Raquel de Lello Rocha Campos Cassano. NGS Soluções Genômicas: Pilar Drummond Sampaio Corrêa Mariani. FZEA-USP Pirassununga: Mirele Daiana Poleti, Jessika Cristina Chagas Lesbon, Elisangela Chicaroni Mattos, Heidge Fukumasu. USP-Botucatu: Rejane Maria Tommasini Grotto, Jayme A. Souza-Neto, Guilherme Targino Valente, Patricia Akemi Assato, Felipe Allan da Silva da Costa, Bianca Cechetto Carlos. Mendelics: Bibiana Santos, João Paulo Kitajima, Erika Freitas, David Schlesinger. Hemocentro Ribeirão Preto: Simone Kashima, Evandra Strazza Rodrigues, Svetoslav Nanev Slavov, Elaine Vieira dos Santos, Rafael dos Santos Bezerra, Luiz Carlos Junior de Alcantara, Marta Giovanetti, Vagner Fonseca, Flavia Aburjaile, Rodrigo Tocantins Calado. |
| EPI_ISL_1795192, EPI_ISL_1795193 | USF SALERNO | Instituto Butantan / ESALQ-Piracicaba | Instituto Butantan: Alexander Roberto Precioso, Dimas Tadeu Covas, Sandra Coccuzzo Sampaio, Maria Carolina Elias, José Salvatore Leister Patané, Vincent Louis Viala, Antonio Jorge Martins, Ricardo Haddad, Claudia Renata dos Santos Barros, Elaine Cristina Marqueze, Raul Machado Neto, Debora Botequio Moretti. Centro de Genômica Funcional da ESALQ: Luiz Lehmann Coutinho, Ricardo Augusto Brassaloti, Raquel de Lello Rocha Campos Cassano. NGS Soluções Genômicas: Pilar Drummond Sampaio Corrêa Mariani. FZEA-USP Pirassununga: Mirele Daiana Poleti, Jessika Cristina Chagas Lesbon, Elisangela Chicaroni Mattos, Heidge Fukumasu. USP-Botucatu: Rejane Maria Tommasini Grotto, Jayme A. Souza-Neto, Guilherme Targino Valente, Patricia Akemi Assato, Felipe Allan da Silva da Costa, Bianca Cechetto Carlos. Mendelics: Bibiana Santos, João Paulo Kitajima, Erika Freitas, David Schlesinger. Hemocentro Ribeirão Preto: Simone Kashima, Evandra Strazza Rodrigues, Svetoslav Nanev Slavov, Elaine Vieira dos Santos, Rafael dos Santos Bezerra, Luiz Carlos Junior de Alcantara, Marta Giovanetti, Vagner Fonseca, Flavia Aburjaile, Rodrigo Tocantins Calado. |
| EPI_ISL_1795194, EPI_ISL_1795195, EPI_ISL_1795196, EPI_ISL_1795197, EPI_ISL_1795198, EPI_ISL_1795199, EPI_ISL_1795200, EPI_ISL_1795201, EPI_ISL_1795202, EPI_ISL_1795203, EPI_ISL_1795204, EPI_ISL_1795205, EPI_ISL_1795206, EPI_ISL_1795207 | | | |
| see above | AMBULATORIO DE ESPECIALIDADE V E MOGI MIRIM | Instituto Butantan / ESALQ-Piracicaba | Instituto Butantan: Alexander Roberto Precioso, Dimas Tadeu Covas, Sandra Coccuzzo Sampaio, Maria Carolina Elias, José Salvatore Leister Patané, Vincent Louis Viala, Antonio Jorge Martins, Ricardo Haddad, Claudia Renata dos Santos Barros, Elaine Cristina Marqueze, Raul Machado Neto, Debora Botequio Moretti. Centro de Genômica Funcional da ESALQ: Luiz Lehmann Coutinho, Ricardo Augusto Brassaloti, Raquel de Lello Rocha Campos Cassano. NGS Soluções Genômicas: Pilar Drummond Sampaio Corrêa Mariani. FZEA-USP Pirassununga: Mirele Daiana Poleti, Jessika Cristina Chagas Lesbon, Elisangela Chicaroni Mattos, Heidge Fukumasu. USP-Botucatu: Rejane Maria Tommasini Grotto, Jayme A. Souza-Neto, Guilherme Targino Valente, Patricia Akemi Assato, Felipe Allan da Silva da Costa, Bianca Cechetto Carlos. Mendelics: Bibiana Santos, João Paulo Kitajima, Erika Freitas, David Schlesinger. Hemocentro Ribeirão Preto: Simone Kashima, Evandra Strazza Rodrigues, Svetoslav Nanev Slavov, Elaine Vieira dos Santos, Rafael dos Santos Bezerra, Luiz Carlos Junior de Alcantara, Marta Giovanetti, Vagner Fonseca, Flavia Aburjaile, Rodrigo Tocantins Calado. |
| EPI_ISL_1795208, EPI_ISL_1795209 | PRONTO SOCORRO MUNICIPAL TAMBAU | Instituto Butantan / ESALQ-Piracicaba | Instituto Butantan: Alexander Roberto Precioso, Dimas Tadeu Covas, Sandra Coccuzzo Sampaio, Maria Carolina Elias, José Salvatore Leister Patané, Vincent Louis Viala, Antonio Jorge Martins, Ricardo Haddad, Claudia Renata dos Santos Barros, Elaine Cristina Marqueze, Raul Machado Neto, Debora Botequio Moretti. Centro de Genômica Funcional da ESALQ: Luiz Lehmann Coutinho, Ricardo Augusto Brassaloti, Raquel de Lello Rocha Campos Cassano. NGS Soluções Genômicas: Pilar Drummond Sampaio Corrêa Mariani. FZEA-USP Pirassununga: Mirele Daiana Poleti, Jessika Cristina Chagas Lesbon, Elisangela Chicaroni Mattos, Heidge Fukumasu. USP-Botucatu: Rejane Maria Tommasini Grotto, Jayme A. Souza-Neto, Guilherme Targino Valente, Patricia Akemi Assato, Felipe Allan da Silva da Costa, Bianca Cechetto Carlos. Mendelics: Bibiana Santos, João Paulo Kitajima, Erika Freitas, David Schlesinger. Hemocentro Ribeirão Preto: Simone Kashima, Evandra Strazza Rodrigues, Svetoslav Nanev Slavov, Elaine Vieira dos Santos, Rafael dos Santos Bezerra, Luiz Carlos Junior de Alcantara, Marta Giovanetti, Vagner Fonseca, Flavia Aburjaile, Rodrigo Tocantins Calado. |
| EPI_ISL_1795210 | AMBULATORIO DE ESPECIALIDADE V E MOGI MIRIM | Instituto Butantan / ESALQ-Piracicaba | Instituto Butantan: Alexander Roberto Precioso, Dimas Tadeu Covas, Sandra Coccuzzo Sampaio, Maria Carolina Elias, José Salvatore Leister Patané, Vincent Louis Viala, Antonio Jorge Martins, Ricardo Haddad, Claudia Renata dos Santos Barros, Elaine Cristina Marqueze, Raul Machado Neto, Debora Botequio Moretti. Centro de Genômica Funcional da ESALQ: Luiz Lehmann Coutinho, Ricardo Augusto Brassaloti, Raquel de Lello Rocha Campos Cassano. NGS Soluções Genômicas: Pilar Drummond Sampaio Corrêa Mariani. FZEA-USP Pirassununga: Mirele Daiana Poleti, Jessika Cristina Chagas Lesbon, Elisangela Chicaroni Mattos, Heidge Fukumasu. USP-Botucatu: Rejane Maria Tommasini Grotto, Jayme A. Souza-Neto, Guilherme Targino |

| | | | |
|---|---|---|---|
| | | | Valente, Patricia Akemi Assato, Felipe Allan da Silva da Costa, Bianca Cechetto Carlos. Mendelics: Bibiana Santos, João Paulo Kitajima, Erika Freitas, David Schlesinger. Hemocentro Ribeirão Preto: Simone Kashima, Evandra Strazza Rodrigues, Svetoslav Nanev Slavov, Elaine Vieira dos Santos, Rafael dos Santos Bezerra, Luiz Carlos Junior de Alcantara, Marta Giovanetti, Vagner Fonseca, Flavia Aburjaile, Rodrigo Tocantins Calado. |
| EPI_ISL_1795211 | HOSPITAL MUNICIPAL DR MARIO GATTI CAMPINAS | Instituto Butantan / ESALQ-Piracicaba | Instituto Butantan: Alexander Roberto Precioso, Dimas Tadeu Covas, Sandra Coccuzzo Sampaio, Maria Carolina Elias, José Salvatore Leister Patané, Vincent Louis Viala, Antonio Jorge Martins, Ricardo Haddad, Claudia Renata dos Santos Barros, Elaine Cristina Marqueze, Raul Machado Neto, Debora Botequio Moretti. Centro de Genômica Funcional da ESALQ: Luiz Lehmann Coutinho, Ricardo Augusto Brassaloti, Raquel de Lello Rocha Campos Cassano. NGS Soluções Genômicas: Pilar Drummond Sampaio Corrêa Mariani. FZEA-USP Pirassununga: Mirele Daiana Poleti, Jessika Cristina Chagas Lesbon, Elisangela Chicaroni Mattos, Heidge Fukumasu. USP-Botucatu: Rejane Maria Tommasini Grotto, Jayme A. Souza-Neto, Guilherme Targino Valente, Patricia Akemi Assato, Felipe Allan da Silva da Costa, Bianca Cechetto Carlos. Mendelics: Bibiana Santos, João Paulo Kitajima, Erika Freitas, David Schlesinger. Hemocentro Ribeirão Preto: Simone Kashima, Evandra Strazza Rodrigues, Svetoslav Nanev Slavov, Elaine Vieira dos Santos, Rafael dos Santos Bezerra, Luiz Carlos Junior de Alcantara, Marta Giovanetti, Vagner Fonseca, Flavia Aburjaile, Rodrigo Tocantins Calado. |
| EPI_ISL_1795212 | CENTRO DE SAUDE II DR ALCIDES FACUNDO ARROYO | Instituto Butantan / ESALQ-Piracicaba | Instituto Butantan: Alexander Roberto Precioso, Dimas Tadeu Covas, Sandra Coccuzzo Sampaio, Maria Carolina Elias, José Salvatore Leister Patané, Vincent Louis Viala, Antonio Jorge Martins, Ricardo Haddad, Claudia Renata dos Santos Barros, Elaine Cristina Marqueze, Raul Machado Neto, Debora Botequio Moretti. Centro de Genômica Funcional da ESALQ: Luiz Lehmann Coutinho, Ricardo Augusto Brassaloti, Raquel de Lello Rocha Campos Cassano. NGS Soluções Genômicas: Pilar Drummond Sampaio Corrêa Mariani. FZEA-USP Pirassununga: Mirele Daiana Poleti, Jessika Cristina Chagas Lesbon, Elisangela Chicaroni Mattos, Heidge Fukumasu. USP-Botucatu: Rejane Maria Tommasini Grotto, Jayme A. Souza-Neto, Guilherme Targino Valente, Patricia Akemi Assato, Felipe Allan da Silva da Costa, Bianca Cechetto Carlos. Mendelics: Bibiana Santos, João Paulo Kitajima, Erika Freitas, David Schlesinger. Hemocentro Ribeirão Preto: Simone Kashima, Evandra Strazza Rodrigues, Svetoslav Nanev Slavov, Elaine Vieira dos Santos, Rafael dos Santos Bezerra, Luiz Carlos Junior de Alcantara, Marta Giovanetti, Vagner Fonseca, Flavia Aburjaile, Rodrigo Tocantins Calado. |
| EPI_ISL_1795213, EPI_ISL_1795214, EPI_ISL_1795215, EPI_ISL_1795216 | PRONTO SOCORRO MUNICIPAL DE SEVERINIA | Instituto Butantan / ESALQ-Piracicaba | Instituto Butantan: Alexander Roberto Precioso, Dimas Tadeu Covas, Sandra Coccuzzo Sampaio, Maria Carolina Elias, José Salvatore Leister Patané, Vincent Louis Viala, Antonio Jorge Martins, Ricardo Haddad, Claudia Renata dos Santos Barros, Elaine Cristina Marqueze, Raul Machado Neto, Debora Botequio Moretti. Centro de Genômica Funcional da ESALQ: Luiz Lehmann Coutinho, Ricardo Augusto Brassaloti, Raquel de Lello Rocha Campos Cassano. NGS Soluções Genômicas: Pilar Drummond Sampaio Corrêa Mariani. FZEA-USP Pirassununga: Mirele Daiana Poleti, Jessika Cristina Chagas Lesbon, Elisangela Chicaroni Mattos, Heidge Fukumasu. USP-Botucatu: Rejane Maria Tommasini Grotto, Jayme A. Souza-Neto, Guilherme Targino Valente, Patricia Akemi Assato, Felipe Allan da Silva da Costa, Bianca Cechetto Carlos. Mendelics: Bibiana Santos, João Paulo Kitajima, Erika Freitas, David Schlesinger. Hemocentro Ribeirão Preto: Simone Kashima, Evandra Strazza Rodrigues, Svetoslav Nanev Slavov, Elaine Vieira dos Santos, Rafael dos Santos Bezerra, Luiz Carlos Junior de Alcantara, Marta Giovanetti, Vagner Fonseca, Flavia Aburjaile, Rodrigo Tocantins Calado. |
| EPI_ISL_1795217 | VIGILANCIA EPIDEMIOLOGICA JARDINOPOLIS SP | Instituto Butantan / ESALQ-Piracicaba | Instituto Butantan: Alexander Roberto Precioso, Dimas Tadeu Covas, Sandra Coccuzzo Sampaio, Maria Carolina Elias, José Salvatore Leister Patané, Vincent Louis Viala, Antonio Jorge Martins, Ricardo Haddad, Claudia Renata dos Santos Barros, Elaine Cristina Marqueze, Raul Machado Neto, Debora Botequio Moretti. Centro de Genômica Funcional da ESALQ: Luiz Lehmann Coutinho, Ricardo Augusto Brassaloti, Raquel de Lello Rocha Campos Cassano. NGS Soluções Genômicas: Pilar Drummond Sampaio Corrêa Mariani. FZEA-USP Pirassununga: Mirele Daiana Poleti, Jessika Cristina Chagas Lesbon, Elisangela Chicaroni Mattos, Heidge Fukumasu. USP-Botucatu: Rejane Maria Tommasini Grotto, Jayme A. Souza-Neto, Guilherme Targino Valente, Patricia Akemi Assato, Felipe Allan da Silva da Costa, Bianca Cechetto Carlos. Mendelics: Bibiana Santos, João Paulo Kitajima, Erika Freitas, David Schlesinger. Hemocentro Ribeirão Preto: Simone Kashima, Evandra Strazza Rodrigues, Svetoslav Nanev Slavov, Elaine Vieira dos Santos, Rafael dos Santos Bezerra, Luiz Carlos Junior de Alcantara, Marta Giovanetti, Vagner Fonseca, Flavia Aburjaile, Rodrigo Tocantins Calado. |
| EPI_ISL_1795218 | VIGILANCIA EPIDIMIOLOGICA DE ARACARIGUAMA | Instituto Butantan / ESALQ-Piracicaba | Instituto Butantan: Alexander Roberto Precioso, Dimas Tadeu Covas, Sandra Coccuzzo Sampaio, Maria Carolina Elias, José Salvatore Leister Patané, Vincent Louis Viala, Antonio Jorge Martins, Ricardo Haddad, Claudia Renata dos Santos Barros, Elaine Cristina Marqueze, Raul Machado Neto, Debora Botequio Moretti. Centro de Genômica Funcional da ESALQ: Luiz Lehmann Coutinho, Ricardo Augusto Brassaloti, Raquel de Lello Rocha Campos Cassano. NGS Soluções Genômicas: Pilar Drummond Sampaio Corrêa Mariani. FZEA-USP Pirassununga: Mirele Daiana Poleti, Jessika Cristina Chagas Lesbon, Elisangela Chicaroni Mattos, Heidge Fukumasu. USP-Botucatu: Rejane Maria Tommasini Grotto, Jayme A. Souza-Neto, Guilherme Targino Valente, Patricia Akemi Assato, Felipe Allan da Silva da Costa, Bianca Cechetto Carlos. Mendelics: Bibiana Santos, João Paulo Kitajima, Erika Freitas, David Schlesinger. Hemocentro Ribeirão Preto: Simone Kashima, Evandra Strazza Rodrigues, Svetoslav Nanev Slavov, Elaine Vieira dos Santos, Rafael dos Santos Bezerra, Luiz Carlos Junior de Alcantara, Marta Giovanetti, Vagner Fonseca, Flavia Aburjaile, Rodrigo Tocantins Calado. |
| EPI_ISL_1795219 | PRONTO ATENDIMENTO UNIDADE SAUDE ADALBERTO ROCHA GUAREI | Instituto Butantan / ESALQ-Piracicaba | Instituto Butantan: Alexander Roberto Precioso, Dimas Tadeu Covas, Sandra Coccuzzo Sampaio, Maria Carolina Elias, José Salvatore Leister Patané, Vincent Louis Viala, Antonio Jorge Martins, Ricardo Haddad, Claudia Renata dos Santos Barros, Elaine Cristina Marqueze, Raul Machado Neto, Debora Botequio Moretti. Centro de Genômica Funcional da ESALQ: Luiz Lehmann Coutinho, Ricardo Augusto Brassaloti, Raquel de Lello Rocha Campos Cassano. NGS Soluções Genômicas: Pilar Drummond Sampaio Corrêa Mariani. FZEA-USP Pirassununga: Mirele Daiana Poleti, Jessika Cristina Chagas Lesbon, Elisangela Chicaroni Mattos, Heidge Fukumasu. USP-Botucatu: Rejane Maria Tommasini Grotto, Jayme A. Souza-Neto, Guilherme Targino Valente, Patricia Akemi Assato, Felipe Allan da Silva da Costa, Bianca Cechetto Carlos. Mendelics: Bibiana Santos, João Paulo Kitajima, Erika Freitas, David Schlesinger. Hemocentro Ribeirão Preto: Simone Kashima, Evandra Strazza Rodrigues, Svetoslav Nanev Slavov, Elaine Vieira dos Santos, Rafael dos Santos Bezerra, Luiz Carlos Junior de Alcantara, Marta Giovanetti, Vagner Fonseca, Flavia Aburjaile, Rodrigo Tocantins Calado. |
| EPI_ISL_1795220 | UNIDADE SENTINELA COVID19 | Instituto Butantan / ESALQ-Piracicaba | Instituto Butantan: Alexander Roberto Precioso, Dimas Tadeu Covas, Sandra Coccuzzo Sampaio, Maria Carolina Elias, José Salvatore Leister Patané, Vincent Louis Viala, Antonio Jorge Martins, Ricardo Haddad, Claudia Renata dos Santos Barros, Elaine Cristina Marqueze, Raul Machado Neto, Debora Botequio Moretti. Centro de Genômica Funcional da ESALQ: Luiz Lehmann Coutinho, Ricardo Augusto Brassaloti, Raquel de Lello Rocha Campos Cassano. NGS Soluções Genômicas: Pilar Drummond Sampaio Corrêa Mariani. FZEA-USP Pirassununga: Mirele Daiana Poleti, Jessika Cristina Chagas Lesbon, Elisangela Chicaroni Mattos, Heidge Fukumasu. USP-Botucatu: Rejane Maria Tommasini Grotto, Jayme A. Souza-Neto, Guilherme Targino Valente, Patricia Akemi Assato, Felipe Allan da Silva da Costa, Bianca Cechetto Carlos. Mendelics: Bibiana Santos, João Paulo Kitajima, Erika Freitas, David Schlesinger. Hemocentro Ribeirão Preto: Simone Kashima, Evandra Strazza Rodrigues, Svetoslav Nanev Slavov, Elaine Vieira dos Santos, Rafael dos Santos Bezerra, Luiz Carlos Junior de Alcantara, Marta Giovanetti, Vagner Fonseca, Flavia Aburjaile, Rodrigo Tocantins Calado. |
| EPI_ISL_1795221 | HOSPITAL REGIONAL DE ITAPETININGA | Instituto Butantan / ESALQ-Piracicaba | Instituto Butantan: Alexander Roberto Precioso, Dimas Tadeu Covas, Sandra Coccuzzo Sampaio, Maria Carolina Elias, José Salvatore Leister Patané, Vincent Louis Viala, Antonio Jorge Martins, Ricardo Haddad, Claudia Renata dos Santos Barros, Elaine Cristina Marqueze, Raul Machado Neto, Debora Botequio Moretti. Centro de Genômica Funcional da ESALQ: Luiz Lehmann Coutinho, Ricardo Augusto Brassaloti, Raquel de Lello Rocha Campos Cassano. NGS Soluções Genômicas: Pilar Drummond Sampaio Corrêa Mariani. FZEA-USP Pirassununga: Mirele Daiana Poleti, Jessika Cristina Chagas Lesbon, Elisangela Chicaroni Mattos, Heidge Fukumasu. USP-Botucatu: Rejane Maria Tommasini Grotto, Jayme A. Souza-Neto, Guilherme Targino Valente, Patricia Akemi Assato, Felipe Allan da Silva da Costa, Bianca Cechetto Carlos. Mendelics: Bibiana Santos, João Paulo Kitajima, Erika Freitas, David Schlesinger. Hemocentro Ribeirão Preto: Simone Kashima, Evandra Strazza Rodrigues, Svetoslav Nanev Slavov, Elaine Vieira dos Santos, Rafael dos Santos Bezerra, Luiz Carlos Junior de Alcantara, Marta Giovanetti, Vagner Fonseca, Flavia Aburjaile, Rodrigo Tocantins Calado. |
| EPI_ISL_1795222 | UNIDADE MISTA ARACOIABA DA SERRA | Instituto Butantan / ESALQ-Piracicaba | Instituto Butantan: Alexander Roberto Precioso, Dimas Tadeu Covas, Sandra Coccuzzo Sampaio, Maria Carolina Elias, José Salvatore Leister Patané, Vincent Louis Viala, Antonio Jorge Martins, Ricardo Haddad, Claudia Renata dos Santos Barros, Elaine Cristina Marqueze, Raul Machado Neto, Debora Botequio Moretti. Centro de Genômica Funcional da ESALQ: Luiz Lehmann Coutinho, Ricardo Augusto Brassaloti, Raquel de Lello Rocha Campos Cassano. NGS Soluções Genômicas: Pilar Drummond Sampaio Corrêa Mariani. FZEA-USP Pirassununga: Mirele Daiana Poleti, Jessika Cristina Chagas Lesbon, Elisangela Chicaroni Mattos, Heidge Fukumasu. USP-Botucatu: Rejane Maria Tommasini Grotto, Jayme A. Souza-Neto, Guilherme Targino Valente, Patricia Akemi Assato, Felipe Allan da Silva da Costa, Bianca Cechetto Carlos. Mendelics: Bibiana Santos, João Paulo Kitajima, Erika Freitas, David Schlesinger. Hemocentro Ribeirão Preto: Simone Kashima, Evandra Strazza Rodrigues, Svetoslav Nanev Slavov, Elaine Vieira dos Santos, Rafael dos Santos Bezerra, Luiz Carlos Junior de Alcantara, Marta Giovanetti, Vagner Fonseca, Flavia Aburjaile, Rodrigo Tocantins Calado. |
| EPI_ISL_1795223 | HOSPITAL REGIONAL DE ITAPETININGA | Instituto Butantan / ESALQ-Piracicaba | Instituto Butantan: Alexander Roberto Precioso, Dimas Tadeu Covas, Sandra Coccuzzo Sampaio, Maria Carolina Elias, José Salvatore Leister Patané, Vincent Louis Viala, Antonio Jorge Martins, Ricardo Haddad, Claudia Renata dos Santos Barros, Elaine Cristina Marqueze, Raul Machado Neto, Debora Botequio Moretti. Centro de Genômica Funcional da ESALQ: Luiz Lehmann Coutinho, Ricardo Augusto Brassaloti, Raquel de Lello Rocha Campos Cassano. NGS Soluções Genômicas: Pilar Drummond Sampaio Corrêa Mariani. FZEA-USP Pirassununga: Mirele Daiana Poleti, Jessika Cristina Chagas Lesbon, Elisangela Chicaroni Mattos, Heidge Fukumasu. USP-Botucatu: Rejane Maria Tommasini Grotto, Jayme A. Souza-Neto, Guilherme Targino |

| | | | |
|---|---|---|---|
| | | | Valente, Patricia Akemi Assato, Felipe Allan da Silva da Costa, Bianca Cechetto Carlos. Mendelics: Bibiana Santos, João Paulo Kitajima, Erika Freitas, David Schlesinger. Hemocentro Ribeirão Preto: Simone Kashima, Evandra Strazza Rodrigues, Svetoslav Nanev Slavov, Elaine Vieira dos Santos, Rafael dos Santos Bezerra, Luiz Carlos Junior de Alcantara, Marta Giovanetti, Vagner Fonseca, Flavia Aburjaile, Rodrigo Tocantins Calado. |
| EPI_ISL_1795224 | UNIDADE BASICA DE SAUDE DR APOLONIO MORAES E SOUZA | Instituto Butantan / ESALQ-Piracicaba | Instituto Butantan: Alexander Roberto Precioso, Dimas Tadeu Covas, Sandra Coccuzzo Sampaio, Maria Carolina Elias, José Salvatore Leister Patané, Vincent Louis Viala, Antonio Jorge Martins, Ricardo Haddad, Claudia Renata dos Santos Barros, Elaine Cristina Marqueze, Raul Machado Neto, Debora Botequio Moretti. Centro de Genômica Funcional da ESALQ: Luiz Lehmann Coutinho, Ricardo Augusto Brassaloti, Raquel de Lello Rocha Campos Cassano. NGS Soluções Genômicas: Pilar Drummond Sampaio Corrêa Mariani. FZEA-USP Pirassununga: Mirele Daiana Poleti, Jessika Cristina Chagas Lesbon, Elisangela Chicaroni Mattos, Heidge Fukumasu. USP-Botucatu: Rejane Maria Tommasini Grotto, Jayme A. Souza-Neto, Guilherme Targino Valente, Patricia Akemi Assato, Felipe Allan da Silva da Costa, Bianca Cechetto Carlos. Mendelics: Bibiana Santos, João Paulo Kitajima, Erika Freitas, David Schlesinger. Hemocentro Ribeirão Preto: Simone Kashima, Evandra Strazza Rodrigues, Svetoslav Nanev Slavov, Elaine Vieira dos Santos, Rafael dos Santos Bezerra, Luiz Carlos Junior de Alcantara, Marta Giovanetti, Vagner Fonseca, Flavia Aburjaile, Rodrigo Tocantins Calado. |
| EPI_ISL_1795225 | UNIDADE BASICA DE SAUDE DR WILSON HAYEK SAIHG | Instituto Butantan / ESALQ-Piracicaba | Instituto Butantan: Alexander Roberto Precioso, Dimas Tadeu Covas, Sandra Coccuzzo Sampaio, Maria Carolina Elias, José Salvatore Leister Patané, Vincent Louis Viala, Antonio Jorge Martins, Ricardo Haddad, Claudia Renata dos Santos Barros, Elaine Cristina Marqueze, Raul Machado Neto, Debora Botequio Moretti. Centro de Genômica Funcional da ESALQ: Luiz Lehmann Coutinho, Ricardo Augusto Brassaloti, Raquel de Lello Rocha Campos Cassano. NGS Soluções Genômicas: Pilar Drummond Sampaio Corrêa Mariani. FZEA-USP Pirassununga: Mirele Daiana Poleti, Jessika Cristina Chagas Lesbon, Elisangela Chicaroni Mattos, Heidge Fukumasu. USP-Botucatu: Rejane Maria Tommasini Grotto, Jayme A. Souza-Neto, Guilherme Targino Valente, Patricia Akemi Assato, Felipe Allan da Silva da Costa, Bianca Cechetto Carlos. Mendelics: Bibiana Santos, João Paulo Kitajima, Erika Freitas, David Schlesinger. Hemocentro Ribeirão Preto: Simone Kashima, Evandra Strazza Rodrigues, Svetoslav Nanev Slavov, Elaine Vieira dos Santos, Rafael dos Santos Bezerra, Luiz Carlos Junior de Alcantara, Marta Giovanetti, Vagner Fonseca, Flavia Aburjaile, Rodrigo Tocantins Calado. |
| EPI_ISL_1795226 | UNIDADE BASICA DR JOSE PARASSU CARVALHO | Instituto Butantan / ESALQ-Piracicaba | Instituto Butantan: Alexander Roberto Precioso, Dimas Tadeu Covas, Sandra Coccuzzo Sampaio, Maria Carolina Elias, José Salvatore Leister Patané, Vincent Louis Viala, Antonio Jorge Martins, Ricardo Haddad, Claudia Renata dos Santos Barros, Elaine Cristina Marqueze, Raul Machado Neto, Debora Botequio Moretti. Centro de Genômica Funcional da ESALQ: Luiz Lehmann Coutinho, Ricardo Augusto Brassaloti, Raquel de Lello Rocha Campos Cassano. NGS Soluções Genômicas: Pilar Drummond Sampaio Corrêa Mariani. FZEA-USP Pirassununga: Mirele Daiana Poleti, Jessika Cristina Chagas Lesbon, Elisangela Chicaroni Mattos, Heidge Fukumasu. USP-Botucatu: Rejane Maria Tommasini Grotto, Jayme A. Souza-Neto, Guilherme Targino Valente, Patricia Akemi Assato, Felipe Allan da Silva da Costa, Bianca Cechetto Carlos. Mendelics: Bibiana Santos, João Paulo Kitajima, Erika Freitas, David Schlesinger. Hemocentro Ribeirão Preto: Simone Kashima, Evandra Strazza Rodrigues, Svetoslav Nanev Slavov, Elaine Vieira dos Santos, Rafael dos Santos Bezerra, Luiz Carlos Junior de Alcantara, Marta Giovanetti, Vagner Fonseca, Flavia Aburjaile, Rodrigo Tocantins Calado. |
| EPI_ISL_1795227 | UNIDADE BASICA DE SAUDE DR LOTFALLAH MIZIARA | Instituto Butantan / ESALQ-Piracicaba | Instituto Butantan: Alexander Roberto Precioso, Dimas Tadeu Covas, Sandra Coccuzzo Sampaio, Maria Carolina Elias, José Salvatore Leister Patané, Vincent Louis Viala, Antonio Jorge Martins, Ricardo Haddad, Claudia Renata dos Santos Barros, Elaine Cristina Marqueze, Raul Machado Neto, Debora Botequio Moretti. Centro de Genômica Funcional da ESALQ: Luiz Lehmann Coutinho, Ricardo Augusto Brassaloti, Raquel de Lello Rocha Campos Cassano. NGS Soluções Genômicas: Pilar Drummond Sampaio Corrêa Mariani. FZEA-USP Pirassununga: Mirele Daiana Poleti, Jessika Cristina Chagas Lesbon, Elisangela Chicaroni Mattos, Heidge Fukumasu. USP-Botucatu: Rejane Maria Tommasini Grotto, Jayme A. Souza-Neto, Guilherme Targino Valente, Patricia Akemi Assato, Felipe Allan da Silva da Costa, Bianca Cechetto Carlos. Mendelics: Bibiana Santos, João Paulo Kitajima, Erika Freitas, David Schlesinger. Hemocentro Ribeirão Preto: Simone Kashima, Evandra Strazza Rodrigues, Svetoslav Nanev Slavov, Elaine Vieira dos Santos, Rafael dos Santos Bezerra, Luiz Carlos Junior de Alcantara, Marta Giovanetti, Vagner Fonseca, Flavia Aburjaile, Rodrigo Tocantins Calado. |
| EPI_ISL_1795228, EPI_ISL_1795229 | CENTRO DE REFERENCIA DO IDOSO DR HUMBERTO MENDES DE CARVALHO | Instituto Butantan / ESALQ-Piracicaba | Instituto Butantan: Alexander Roberto Precioso, Dimas Tadeu Covas, Sandra Coccuzzo Sampaio, Maria Carolina Elias, José Salvatore Leister Patané, Vincent Louis Viala, Antonio Jorge Martins, Ricardo Haddad, Claudia Renata dos Santos Barros, Elaine Cristina Marqueze, Raul Machado Neto, Debora Botequio Moretti. Centro de Genômica Funcional da ESALQ: Luiz Lehmann Coutinho, Ricardo Augusto Brassaloti, Raquel de Lello Rocha Campos Cassano. NGS Soluções Genômicas: Pilar Drummond Sampaio Corrêa Mariani. FZEA-USP Pirassununga: Mirele Daiana Poleti, Jessika Cristina Chagas Lesbon, Elisangela Chicaroni Mattos, Heidge Fukumasu. USP-Botucatu: Rejane Maria Tommasini Grotto, Jayme A. Souza-Neto, Guilherme Targino Valente, Patricia Akemi Assato, Felipe Allan da Silva da Costa, Bianca Cechetto Carlos. Mendelics: Bibiana Santos, João Paulo Kitajima, Erika Freitas, David Schlesinger. Hemocentro Ribeirão Preto: Simone Kashima, Evandra Strazza Rodrigues, Svetoslav Nanev Slavov, Elaine Vieira dos Santos, Rafael dos Santos Bezerra, Luiz Carlos Junior de Alcantara, Marta Giovanetti, Vagner Fonseca, Flavia Aburjaile, Rodrigo Tocantins Calado. |
| EPI_ISL_1795230, EPI_ISL_1795231 | UNIDADE DE SAUDE DA FAMILIA JOSE ADALBERTO LELLIS GARCIA | Instituto Butantan / ESALQ-Piracicaba | Instituto Butantan: Alexander Roberto Precioso, Dimas Tadeu Covas, Sandra Coccuzzo Sampaio, Maria Carolina Elias, José Salvatore Leister Patané, Vincent Louis Viala, Antonio Jorge Martins, Ricardo Haddad, Claudia Renata dos Santos Barros, Elaine Cristina Marqueze, Raul Machado Neto, Debora Botequio Moretti. Centro de Genômica Funcional da ESALQ: Luiz Lehmann Coutinho, Ricardo Augusto Brassaloti, Raquel de Lello Rocha Campos Cassano. NGS Soluções Genômicas: Pilar Drummond Sampaio Corrêa Mariani. FZEA-USP Pirassununga: Mirele Daiana Poleti, Jessika Cristina Chagas Lesbon, Elisangela Chicaroni Mattos, Heidge Fukumasu. USP-Botucatu: Rejane Maria Tommasini Grotto, Jayme A. Souza-Neto, Guilherme Targino Valente, Patricia Akemi Assato, Felipe Allan da Silva da Costa, Bianca Cechetto Carlos. Mendelics: Bibiana Santos, João Paulo Kitajima, Erika Freitas, David Schlesinger. Hemocentro Ribeirão Preto: Simone Kashima, Evandra Strazza Rodrigues, Svetoslav Nanev Slavov, Elaine Vieira dos Santos, Rafael dos Santos Bezerra, Luiz Carlos Junior de Alcantara, Marta Giovanetti, Vagner Fonseca, Flavia Aburjaile, Rodrigo Tocantins Calado. |
| EPI_ISL_1795232, EPI_ISL_1795233 | CS DE URUPES | Instituto Butantan / ESALQ-Piracicaba | Instituto Butantan: Alexander Roberto Precioso, Dimas Tadeu Covas, Sandra Coccuzzo Sampaio, Maria Carolina Elias, José Salvatore Leister Patané, Vincent Louis Viala, Antonio Jorge Martins, Ricardo Haddad, Claudia Renata dos Santos Barros, Elaine Cristina Marqueze, Raul Machado Neto, Debora Botequio Moretti. Centro de Genômica Funcional da ESALQ: Luiz Lehmann Coutinho, Ricardo Augusto Brassaloti, Raquel de Lello Rocha Campos Cassano. NGS Soluções Genômicas: Pilar Drummond Sampaio Corrêa Mariani. FZEA-USP Pirassununga: Mirele Daiana Poleti, Jessika Cristina Chagas Lesbon, Elisangela Chicaroni Mattos, Heidge Fukumasu. USP-Botucatu: Rejane Maria Tommasini Grotto, Jayme A. Souza-Neto, Guilherme Targino Valente, Patricia Akemi Assato, Felipe Allan da Silva da Costa, Bianca Cechetto Carlos. Mendelics: Bibiana Santos, João Paulo Kitajima, Erika Freitas, David Schlesinger. Hemocentro Ribeirão Preto: Simone Kashima, Evandra Strazza Rodrigues, Svetoslav Nanev Slavov, Elaine Vieira dos Santos, Rafael dos Santos Bezerra, Luiz Carlos Junior de Alcantara, Marta Giovanetti, Vagner Fonseca, Flavia Aburjaile, Rodrigo Tocantins Calado. |
| EPI_ISL_1795234, EPI_ISL_1795235, EPI_ISL_1795236, EPI_ISL_1795237, EPI_ISL_1795238 | CS DE BALSAMO | Instituto Butantan / ESALQ-Piracicaba | Instituto Butantan: Alexander Roberto Precioso, Dimas Tadeu Covas, Sandra Coccuzzo Sampaio, Maria Carolina Elias, José Salvatore Leister Patané, Vincent Louis Viala, Antonio Jorge Martins, Ricardo Haddad, Claudia Renata dos Santos Barros, Elaine Cristina Marqueze, Raul Machado Neto, Debora Botequio Moretti. Centro de Genômica Funcional da ESALQ: Luiz Lehmann Coutinho, Ricardo Augusto Brassaloti, Raquel de Lello Rocha Campos Cassano. NGS Soluções Genômicas: Pilar Drummond Sampaio Corrêa Mariani. FZEA-USP Pirassununga: Mirele Daiana Poleti, Jessika Cristina Chagas Lesbon, Elisangela Chicaroni Mattos, Heidge Fukumasu. USP-Botucatu: Rejane Maria Tommasini Grotto, Jayme A. Souza-Neto, Guilherme Targino Valente, Patricia Akemi Assato, Felipe Allan da Silva da Costa, Bianca Cechetto Carlos. Mendelics: Bibiana Santos, João Paulo Kitajima, Erika Freitas, David Schlesinger. Hemocentro Ribeirão Preto: Simone Kashima, Evandra Strazza Rodrigues, Svetoslav Nanev Slavov, Elaine Vieira dos Santos, Rafael dos Santos Bezerra, Luiz Carlos Junior de Alcantara, Marta Giovanetti, Vagner Fonseca, Flavia Aburjaile, Rodrigo Tocantins Calado. |
| EPI_ISL_1795239 | CS DE URUPES | Instituto Butantan / ESALQ-Piracicaba | Instituto Butantan: Alexander Roberto Precioso, Dimas Tadeu Covas, Sandra Coccuzzo Sampaio, Maria Carolina Elias, José Salvatore Leister Patané, Vincent Louis Viala, Antonio Jorge Martins, Ricardo Haddad, Claudia Renata dos Santos Barros, Elaine Cristina Marqueze, Raul Machado Neto, Debora Botequio Moretti. Centro de Genômica Funcional da ESALQ: Luiz Lehmann Coutinho, Ricardo Augusto Brassaloti, Raquel de Lello Rocha Campos Cassano. NGS Soluções Genômicas: Pilar Drummond Sampaio Corrêa Mariani. FZEA-USP Pirassununga: Mirele Daiana Poleti, Jessika Cristina Chagas Lesbon, Elisangela Chicaroni Mattos, Heidge Fukumasu. USP-Botucatu: Rejane Maria Tommasini Grotto, Jayme A. Souza-Neto, Guilherme Targino Valente, Patricia Akemi Assato, Felipe Allan da Silva da Costa, Bianca Cechetto Carlos. Mendelics: Bibiana Santos, João Paulo Kitajima, Erika Freitas, David Schlesinger. Hemocentro Ribeirão Preto: Simone Kashima, Evandra Strazza Rodrigues, Svetoslav Nanev Slavov, Elaine Vieira dos Santos, Rafael dos Santos Bezerra, Luiz Carlos Junior de Alcantara, Marta Giovanetti, Vagner Fonseca, Flavia Aburjaile, Rodrigo Tocantins Calado. |
| EPI_ISL_1795240 | CS DE SEBASTIANOPOLIS DO SUL | Instituto Butantan / ESALQ-Piracicaba | Instituto Butantan: Alexander Roberto Precioso, Dimas Tadeu Covas, Sandra Coccuzzo Sampaio, Maria Carolina Elias, José Salvatore Leister Patané, Vincent Louis Viala, Antonio Jorge Martins, Ricardo Haddad, Claudia Renata dos Santos Barros, Elaine Cristina Marqueze, Raul Machado Neto, Debora Botequio Moretti. Centro de Genômica Funcional da ESALQ: Luiz Lehmann Coutinho, Ricardo Augusto Brassaloti, Raquel de Lello Rocha Campos Cassano. NGS Soluções Genômicas: Pilar Drummond Sampaio Corrêa Mariani. FZEA-USP Pirassununga: Mirele Daiana Poleti, Jessika Cristina Chagas Lesbon, Elisangela Chicaroni Mattos, Heidge Fukumasu. USP-Botucatu: Rejane Maria Tommasini Grotto, Jayme A. Souza-Neto, Guilherme Targino |

| | | | |
|---|---|---|---|
| | | | Valente, Patricia Akemi Assato, Felipe Allan da Silva da Costa, Bianca Cechetto Carlos. Mendelics: Bibiana Santos, João Paulo Kitajima, Erika Freitas, David Schlesinger. Hemocentro Ribeirão Preto: Simone Kashima, Evandra Strazza Rodrigues, Svetoslav Nanev Slavov, Elaine Vieira dos Santos, Rafael dos Santos Bezerra, Luiz Carlos Junior de Alcantara, Marta Giovanetti, Vagner Fonseca, Flavia Aburjaile, Rodrigo Tocantins Calado. |
| EPI_ISL_1795241 | CS DE BALSAMO | Instituto Butantan / ESALQ-Piracicaba | Instituto Butantan: Alexander Roberto Precioso, Dimas Tadeu Covas, Sandra Coccuzzo Sampaio, Maria Carolina Elias, José Salvatore Leister Patané, Vincent Louis Viala, Antonio Jorge Martins, Ricardo Haddad, Claudia Renata dos Santos Barros, Elaine Cristina Marqueze, Raul Machado Neto, Debora Botequio Moretti. Centro de Genômica Funcional da ESALQ: Luiz Lehmann Coutinho, Ricardo Augusto Brassaloti, Raquel de Lello Rocha Campos Cassano. NGS Soluções Genômicas: Pilar Drummond Sampaio Corrêa Mariani. FZEA-USP Pirassununga: Mirele Daiana Poleti, Jessika Cristina Chagas Lesbon, Elisangela Chicaroni Mattos, Heidge Fukumasu. USP-Botucatu: Rejane Maria Tommasini Grotto, Jayme A. Souza-Neto, Guilherme Targino Valente, Patricia Akemi Assato, Felipe Allan da Silva da Costa, Bianca Cechetto Carlos. Mendelics: Bibiana Santos, João Paulo Kitajima, Erika Freitas, David Schlesinger. Hemocentro Ribeirão Preto: Simone Kashima, Evandra Strazza Rodrigues, Svetoslav Nanev Slavov, Elaine Vieira dos Santos, Rafael dos Santos Bezerra, Luiz Carlos Junior de Alcantara, Marta Giovanetti, Vagner Fonseca, Flavia Aburjaile, Rodrigo Tocantins Calado. |
| EPI_ISL_1795242, EPI_ISL_1795243, EPI_ISL_1795244 | CS DE URUPES | Instituto Butantan / ESALQ-Piracicaba | Instituto Butantan: Alexander Roberto Precioso, Dimas Tadeu Covas, Sandra Coccuzzo Sampaio, Maria Carolina Elias, José Salvatore Leister Patané, Vincent Louis Viala, Antonio Jorge Martins, Ricardo Haddad, Claudia Renata dos Santos Barros, Elaine Cristina Marqueze, Raul Machado Neto, Debora Botequio Moretti. Centro de Genômica Funcional da ESALQ: Luiz Lehmann Coutinho, Ricardo Augusto Brassaloti, Raquel de Lello Rocha Campos Cassano. NGS Soluções Genômicas: Pilar Drummond Sampaio Corrêa Mariani. FZEA-USP Pirassununga: Mirele Daiana Poleti, Jessika Cristina Chagas Lesbon, Elisangela Chicaroni Mattos, Heidge Fukumasu. USP-Botucatu: Rejane Maria Tommasini Grotto, Jayme A. Souza-Neto, Guilherme Targino Valente, Patricia Akemi Assato, Felipe Allan da Silva da Costa, Bianca Cechetto Carlos. Mendelics: Bibiana Santos, João Paulo Kitajima, Erika Freitas, David Schlesinger. Hemocentro Ribeirão Preto: Simone Kashima, Evandra Strazza Rodrigues, Svetoslav Nanev Slavov, Elaine Vieira dos Santos, Rafael dos Santos Bezerra, Luiz Carlos Junior de Alcantara, Marta Giovanetti, Vagner Fonseca, Flavia Aburjaile, Rodrigo Tocantins Calado. |
| EPI_ISL_1795245, EPI_ISL_1795246, EPI_ISL_1795247 | EMERGENCIA RESPIRATORIA DE NOVA GRANADA | Instituto Butantan / ESALQ-Piracicaba | Instituto Butantan: Alexander Roberto Precioso, Dimas Tadeu Covas, Sandra Coccuzzo Sampaio, Maria Carolina Elias, José Salvatore Leister Patané, Vincent Louis Viala, Antonio Jorge Martins, Ricardo Haddad, Claudia Renata dos Santos Barros, Elaine Cristina Marqueze, Raul Machado Neto, Debora Botequio Moretti. Centro de Genômica Funcional da ESALQ: Luiz Lehmann Coutinho, Ricardo Augusto Brassaloti, Raquel de Lello Rocha Campos Cassano. NGS Soluções Genômicas: Pilar Drummond Sampaio Corrêa Mariani. FZEA-USP Pirassununga: Mirele Daiana Poleti, Jessika Cristina Chagas Lesbon, Elisangela Chicaroni Mattos, Heidge Fukumasu. USP-Botucatu: Rejane Maria Tommasini Grotto, Jayme A. Souza-Neto, Guilherme Targino Valente, Patricia Akemi Assato, Felipe Allan da Silva da Costa, Bianca Cechetto Carlos. Mendelics: Bibiana Santos, João Paulo Kitajima, Erika Freitas, David Schlesinger. Hemocentro Ribeirão Preto: Simone Kashima, Evandra Strazza Rodrigues, Svetoslav Nanev Slavov, Elaine Vieira dos Santos, Rafael dos Santos Bezerra, Luiz Carlos Junior de Alcantara, Marta Giovanetti, Vagner Fonseca, Flavia Aburjaile, Rodrigo Tocantins Calado. |
| EPI_ISL_1795248, EPI_ISL_1795249, EPI_ISL_1795250 | ESF NOVA TANABI II | Instituto Butantan / ESALQ-Piracicaba | Instituto Butantan: Alexander Roberto Precioso, Dimas Tadeu Covas, Sandra Coccuzzo Sampaio, Maria Carolina Elias, José Salvatore Leister Patané, Vincent Louis Viala, Antonio Jorge Martins, Ricardo Haddad, Claudia Renata dos Santos Barros, Elaine Cristina Marqueze, Raul Machado Neto, Debora Botequio Moretti. Centro de Genômica Funcional da ESALQ: Luiz Lehmann Coutinho, Ricardo Augusto Brassaloti, Raquel de Lello Rocha Campos Cassano. NGS Soluções Genômicas: Pilar Drummond Sampaio Corrêa Mariani. FZEA-USP Pirassununga: Mirele Daiana Poleti, Jessika Cristina Chagas Lesbon, Elisangela Chicaroni Mattos, Heidge Fukumasu. USP-Botucatu: Rejane Maria Tommasini Grotto, Jayme A. Souza-Neto, Guilherme Targino Valente, Patricia Akemi Assato, Felipe Allan da Silva da Costa, Bianca Cechetto Carlos. Mendelics: Bibiana Santos, João Paulo Kitajima, Erika Freitas, David Schlesinger. Hemocentro Ribeirão Preto: Simone Kashima, Evandra Strazza Rodrigues, Svetoslav Nanev Slavov, Elaine Vieira dos Santos, Rafael dos Santos Bezerra, Luiz Carlos Junior de Alcantara, Marta Giovanetti, Vagner Fonseca, Flavia Aburjaile, Rodrigo Tocantins Calado. |
| EPI_ISL_1795251, EPI_ISL_1795252, EPI_ISL_1795253, EPI_ISL_1795254, EPI_ISL_1795255, EPI_ISL_1795256, EPI_ISL_1795257 | UBS II DE TANABI MILTON MARTINS PERCHES | Instituto Butantan / ESALQ-Piracicaba | Instituto Butantan: Alexander Roberto Precioso, Dimas Tadeu Covas, Sandra Coccuzzo Sampaio, Maria Carolina Elias, José Salvatore Leister Patané, Vincent Louis Viala, Antonio Jorge Martins, Ricardo Haddad, Claudia Renata dos Santos Barros, Elaine Cristina Marqueze, Raul Machado Neto, Debora Botequio Moretti. Centro de Genômica Funcional da ESALQ: Luiz Lehmann Coutinho, Ricardo Augusto Brassaloti, Raquel de Lello Rocha Campos Cassano. NGS Soluções Genômicas: Pilar Drummond Sampaio Corrêa Mariani. FZEA-USP Pirassununga: Mirele Daiana Poleti, Jessika Cristina Chagas Lesbon, Elisangela Chicaroni Mattos, Heidge Fukumasu. USP-Botucatu: Rejane Maria Tommasini Grotto, Jayme A. Souza-Neto, Guilherme Targino Valente, Patricia Akemi Assato, Felipe Allan da Silva da Costa, Bianca Cechetto Carlos. Mendelics: Bibiana Santos, João Paulo Kitajima, Erika Freitas, David Schlesinger. Hemocentro Ribeirão Preto: Simone Kashima, Evandra Strazza Rodrigues, Svetoslav Nanev Slavov, Elaine Vieira dos Santos, Rafael dos Santos Bezerra, Luiz Carlos Junior de Alcantara, Marta Giovanetti, Vagner Fonseca, Flavia Aburjaile, Rodrigo Tocantins Calado. |
| EPI_ISL_1795258 | CS DE PLANALTO | Instituto Butantan / ESALQ-Piracicaba | Instituto Butantan: Alexander Roberto Precioso, Dimas Tadeu Covas, Sandra Coccuzzo Sampaio, Maria Carolina Elias, José Salvatore Leister Patané, Vincent Louis Viala, Antonio Jorge Martins, Ricardo Haddad, Claudia Renata dos Santos Barros, Elaine Cristina Marqueze, Raul Machado Neto, Debora Botequio Moretti. Centro de Genômica Funcional da ESALQ: Luiz Lehmann Coutinho, Ricardo Augusto Brassaloti, Raquel de Lello Rocha Campos Cassano. NGS Soluções Genômicas: Pilar Drummond Sampaio Corrêa Mariani. FZEA-USP Pirassununga: Mirele Daiana Poleti, Jessika Cristina Chagas Lesbon, Elisangela Chicaroni Mattos, Heidge Fukumasu. USP-Botucatu: Rejane Maria Tommasini Grotto, Jayme A. Souza-Neto, Guilherme Targino Valente, Patricia Akemi Assato, Felipe Allan da Silva da Costa, Bianca Cechetto Carlos. Mendelics: Bibiana Santos, João Paulo Kitajima, Erika Freitas, David Schlesinger. Hemocentro Ribeirão Preto: Simone Kashima, Evandra Strazza Rodrigues, Svetoslav Nanev Slavov, Elaine Vieira dos Santos, Rafael dos Santos Bezerra, Luiz Carlos Junior de Alcantara, Marta Giovanetti, Vagner Fonseca, Flavia Aburjaile, Rodrigo Tocantins Calado. |
| EPI_ISL_1795259, EPI_ISL_1795260, EPI_ISL_1795261, EPI_ISL_1795262, EPI_ISL_1795263 | CS DE ICEM | Instituto Butantan / ESALQ-Piracicaba | Instituto Butantan: Alexander Roberto Precioso, Dimas Tadeu Covas, Sandra Coccuzzo Sampaio, Maria Carolina Elias, José Salvatore Leister Patané, Vincent Louis Viala, Antonio Jorge Martins, Ricardo Haddad, Claudia Renata dos Santos Barros, Elaine Cristina Marqueze, Raul Machado Neto, Debora Botequio Moretti. Centro de Genômica Funcional da ESALQ: Luiz Lehmann Coutinho, Ricardo Augusto Brassaloti, Raquel de Lello Rocha Campos Cassano. NGS Soluções Genômicas: Pilar Drummond Sampaio Corrêa Mariani. FZEA-USP Pirassununga: Mirele Daiana Poleti, Jessika Cristina Chagas Lesbon, Elisangela Chicaroni Mattos, Heidge Fukumasu. USP-Botucatu: Rejane Maria Tommasini Grotto, Jayme A. Souza-Neto, Guilherme Targino Valente, Patricia Akemi Assato, Felipe Allan da Silva da Costa, Bianca Cechetto Carlos. Mendelics: Bibiana Santos, João Paulo Kitajima, Erika Freitas, David Schlesinger. Hemocentro Ribeirão Preto: Simone Kashima, Evandra Strazza Rodrigues, Svetoslav Nanev Slavov, Elaine Vieira dos Santos, Rafael dos Santos Bezerra, Luiz Carlos Junior de Alcantara, Marta Giovanetti, Vagner Fonseca, Flavia Aburjaile, Rodrigo Tocantins Calado. |
| EPI_ISL_1795264 | CS DE PARANAPUA | Instituto Butantan / ESALQ-Piracicaba | Instituto Butantan: Alexander Roberto Precioso, Dimas Tadeu Covas, Sandra Coccuzzo Sampaio, Maria Carolina Elias, José Salvatore Leister Patané, Vincent Louis Viala, Antonio Jorge Martins, Ricardo Haddad, Claudia Renata dos Santos Barros, Elaine Cristina Marqueze, Raul Machado Neto, Debora Botequio Moretti. Centro de Genômica Funcional da ESALQ: Luiz Lehmann Coutinho, Ricardo Augusto Brassaloti, Raquel de Lello Rocha Campos Cassano. NGS Soluções Genômicas: Pilar Drummond Sampaio Corrêa Mariani. FZEA-USP Pirassununga: Mirele Daiana Poleti, Jessika Cristina Chagas Lesbon, Elisangela Chicaroni Mattos, Heidge Fukumasu. USP-Botucatu: Rejane Maria Tommasini Grotto, Jayme A. Souza-Neto, Guilherme Targino Valente, Patricia Akemi Assato, Felipe Allan da Silva da Costa, Bianca Cechetto Carlos. Mendelics: Bibiana Santos, João Paulo Kitajima, Erika Freitas, David Schlesinger. Hemocentro Ribeirão Preto: Simone Kashima, Evandra Strazza Rodrigues, Svetoslav Nanev Slavov, Elaine Vieira dos Santos, Rafael dos Santos Bezerra, Luiz Carlos Junior de Alcantara, Marta Giovanetti, Vagner Fonseca, Flavia Aburjaile, Rodrigo Tocantins Calado. |
| EPI_ISL_1795265 | CS DE ICEM | Instituto Butantan / ESALQ-Piracicaba | Instituto Butantan: Alexander Roberto Precioso, Dimas Tadeu Covas, Sandra Coccuzzo Sampaio, Maria Carolina Elias, José Salvatore Leister Patané, Vincent Louis Viala, Antonio Jorge Martins, Ricardo Haddad, Claudia Renata dos Santos Barros, Elaine Cristina Marqueze, Raul Machado Neto, Debora Botequio Moretti. Centro de Genômica Funcional da ESALQ: Luiz Lehmann Coutinho, Ricardo Augusto Brassaloti, Raquel de Lello Rocha Campos Cassano. NGS Soluções Genômicas: Pilar Drummond Sampaio Corrêa Mariani. FZEA-USP Pirassununga: Mirele Daiana Poleti, Jessika Cristina Chagas Lesbon, Elisangela Chicaroni Mattos, Heidge Fukumasu. USP-Botucatu: Rejane Maria Tommasini Grotto, Jayme A. Souza-Neto, Guilherme Targino Valente, Patricia Akemi Assato, Felipe Allan da Silva da Costa, Bianca Cechetto Carlos. Mendelics: Bibiana Santos, João Paulo Kitajima, Erika Freitas, David Schlesinger. Hemocentro Ribeirão Preto: Simone Kashima, Evandra Strazza Rodrigues, Svetoslav Nanev Slavov, Elaine Vieira dos Santos, Rafael dos Santos Bezerra, Luiz Carlos Junior de Alcantara, Marta Giovanetti, Vagner Fonseca, Flavia Aburjaile, Rodrigo Tocantins Calado. |
| EPI_ISL_1795266, EPI_ISL_1795267, EPI_ISL_1795268 | UBS LUIS FACHIN IPIGUA | Instituto Butantan / ESALQ-Piracicaba | Instituto Butantan: Alexander Roberto Precioso, Dimas Tadeu Covas, Sandra Coccuzzo Sampaio, Maria Carolina Elias, José Salvatore Leister Patané, Vincent Louis Viala, Antonio Jorge Martins, Ricardo Haddad, Claudia Renata dos Santos Barros, Elaine Cristina Marqueze, Raul Machado Neto, Debora Botequio Moretti. Centro de Genômica Funcional da ESALQ: Luiz Lehmann Coutinho, Ricardo Augusto Brassaloti, Raquel de Lello Rocha Campos Cassano. NGS Soluções Genômicas: Pilar Drummond Sampaio Corrêa Mariani. FZEA-USP Pirassununga: Mirele Daiana Poleti, Jessika Cristina Chagas Lesbon, Elisangela Chicaroni Mattos, Heidge Fukumasu. USP-Botucatu: Rejane Maria Tommasini Grotto, Jayme A. Souza-Neto, Guilherme Targino |

| | | | |
|---|---|---|---|
| | | | Valente, Patricia Akemi Assato, Felipe Allan da Silva da Costa, Bianca Cechetto Carlos. Mendelics: Bibiana Santos, João Paulo Kitajima, Erika Freitas, David Schlesinger. Hemocentro Ribeirão Preto: Simone Kashima, Evandra Strazza Rodrigues, Svetoslav Nanev Slavov, Elaine Vieira dos Santos, Rafael dos Santos Bezerra, Luiz Carlos Junior de Alcantara, Marta Giovanetti, Vagner Fonseca, Flavia Aburjaile, Rodrigo Tocantins Calado. |
| EPI_ISL_1795269, EPI_ISL_1795270 | UBS TEREZA GALLO UCHOA | Instituto Butantan / ESALQ-Piracicaba | Instituto Butantan: Alexander Roberto Precioso, Dimas Tadeu Covas, Sandra Coccuzzo Sampaio, Maria Carolina Elias, José Salvatore Leister Patané, Vincent Louis Viala, Antonio Jorge Martins, Ricardo Haddad, Claudia Renata dos Santos Barros, Elaine Cristina Marqueze, Raul Machado Neto, Debora Botequio Moretti. Centro de Genômica Funcional da ESALQ: Luiz Lehmann Coutinho, Ricardo Augusto Brassaloti, Raquel de Lello Rocha Campos Cassano. NGS Soluções Genômicas: Pilar Drummond Sampaio Corrêa Mariani. FZEA-USP Pirassununga: Mirele Daiana Poleti, Jessika Cristina Chagas Lesbon, Elisangela Chicaroni Mattos, Heidge Fukumasu. USP-Botucatu: Rejane Maria Tommasini Grotto, Jayme A. Souza-Neto, Guilherme Targino Valente, Patricia Akemi Assato, Felipe Allan da Silva da Costa, Bianca Cechetto Carlos. Mendelics: Bibiana Santos, João Paulo Kitajima, Erika Freitas, David Schlesinger. Hemocentro Ribeirão Preto: Simone Kashima, Evandra Strazza Rodrigues, Svetoslav Nanev Slavov, Elaine Vieira dos Santos, Rafael dos Santos Bezerra, Luiz Carlos Junior de Alcantara, Marta Giovanetti, Vagner Fonseca, Flavia Aburjaile, Rodrigo Tocantins Calado. |
| EPI_ISL_1795271 | CS DE PARANAPUA | Instituto Butantan / ESALQ-Piracicaba | Instituto Butantan: Alexander Roberto Precioso, Dimas Tadeu Covas, Sandra Coccuzzo Sampaio, Maria Carolina Elias, José Salvatore Leister Patané, Vincent Louis Viala, Antonio Jorge Martins, Ricardo Haddad, Claudia Renata dos Santos Barros, Elaine Cristina Marqueze, Raul Machado Neto, Debora Botequio Moretti. Centro de Genômica Funcional da ESALQ: Luiz Lehmann Coutinho, Ricardo Augusto Brassaloti, Raquel de Lello Rocha Campos Cassano. NGS Soluções Genômicas: Pilar Drummond Sampaio Corrêa Mariani. FZEA-USP Pirassununga: Mirele Daiana Poleti, Jessika Cristina Chagas Lesbon, Elisangela Chicaroni Mattos, Heidge Fukumasu. USP-Botucatu: Rejane Maria Tommasini Grotto, Jayme A. Souza-Neto, Guilherme Targino Valente, Patricia Akemi Assato, Felipe Allan da Silva da Costa, Bianca Cechetto Carlos. Mendelics: Bibiana Santos, João Paulo Kitajima, Erika Freitas, David Schlesinger. Hemocentro Ribeirão Preto: Simone Kashima, Evandra Strazza Rodrigues, Svetoslav Nanev Slavov, Elaine Vieira dos Santos, Rafael dos Santos Bezerra, Luiz Carlos Junior de Alcantara, Marta Giovanetti, Vagner Fonseca, Flavia Aburjaile, Rodrigo Tocantins Calado. |
| EPI_ISL_1795272, EPI_ISL_1795273 | UBS TEREZA GALLO UCHOA | Instituto Butantan / ESALQ-Piracicaba | Instituto Butantan: Alexander Roberto Precioso, Dimas Tadeu Covas, Sandra Coccuzzo Sampaio, Maria Carolina Elias, José Salvatore Leister Patané, Vincent Louis Viala, Antonio Jorge Martins, Ricardo Haddad, Claudia Renata dos Santos Barros, Elaine Cristina Marqueze, Raul Machado Neto, Debora Botequio Moretti. Centro de Genômica Funcional da ESALQ: Luiz Lehmann Coutinho, Ricardo Augusto Brassaloti, Raquel de Lello Rocha Campos Cassano. NGS Soluções Genômicas: Pilar Drummond Sampaio Corrêa Mariani. FZEA-USP Pirassununga: Mirele Daiana Poleti, Jessika Cristina Chagas Lesbon, Elisangela Chicaroni Mattos, Heidge Fukumasu. USP-Botucatu: Rejane Maria Tommasini Grotto, Jayme A. Souza-Neto, Guilherme Targino Valente, Patricia Akemi Assato, Felipe Allan da Silva da Costa, Bianca Cechetto Carlos. Mendelics: Bibiana Santos, João Paulo Kitajima, Erika Freitas, David Schlesinger. Hemocentro Ribeirão Preto: Simone Kashima, Evandra Strazza Rodrigues, Svetoslav Nanev Slavov, Elaine Vieira dos Santos, Rafael dos Santos Bezerra, Luiz Carlos Junior de Alcantara, Marta Giovanetti, Vagner Fonseca, Flavia Aburjaile, Rodrigo Tocantins Calado. |
| EPI_ISL_1795274, EPI_ISL_1795275 | CENTRO MEDICO PMESP | Instituto Butantan / ESALQ-Piracicaba | Instituto Butantan: Alexander Roberto Precioso, Dimas Tadeu Covas, Sandra Coccuzzo Sampaio, Maria Carolina Elias, José Salvatore Leister Patané, Vincent Louis Viala, Antonio Jorge Martins, Ricardo Haddad, Claudia Renata dos Santos Barros, Elaine Cristina Marqueze, Raul Machado Neto, Debora Botequio Moretti. Centro de Genômica Funcional da ESALQ: Luiz Lehmann Coutinho, Ricardo Augusto Brassaloti, Raquel de Lello Rocha Campos Cassano. NGS Soluções Genômicas: Pilar Drummond Sampaio Corrêa Mariani. FZEA-USP Pirassununga: Mirele Daiana Poleti, Jessika Cristina Chagas Lesbon, Elisangela Chicaroni Mattos, Heidge Fukumasu. USP-Botucatu: Rejane Maria Tommasini Grotto, Jayme A. Souza-Neto, Guilherme Targino Valente, Patricia Akemi Assato, Felipe Allan da Silva da Costa, Bianca Cechetto Carlos. Mendelics: Bibiana Santos, João Paulo Kitajima, Erika Freitas, David Schlesinger. Hemocentro Ribeirão Preto: Simone Kashima, Evandra Strazza Rodrigues, Svetoslav Nanev Slavov, Elaine Vieira dos Santos, Rafael dos Santos Bezerra, Luiz Carlos Junior de Alcantara, Marta Giovanetti, Vagner Fonseca, Flavia Aburjaile, Rodrigo Tocantins Calado. |
| EPI_ISL_1795276, EPI_ISL_1795277, EPI_ISL_1795278, EPI_ISL_1795279, EPI_ISL_1795280, EPI_ISL_1795281, EPI_ISL_1795282, EPI_ISL_1795283, EPI_ISL_1795284, EPI_ISL_1795285, EPI_ISL_1795286 | | | |
| see above | CS DE PALESTINA | Instituto Butantan / ESALQ-Piracicaba | Instituto Butantan: Alexander Roberto Precioso, Dimas Tadeu Covas, Sandra Coccuzzo Sampaio, Maria Carolina Elias, José Salvatore Leister Patané, Vincent Louis Viala, Antonio Jorge Martins, Ricardo Haddad, Claudia Renata dos Santos Barros, Elaine Cristina Marqueze, Raul Machado Neto, Debora Botequio Moretti. Centro de Genômica Funcional da ESALQ: Luiz Lehmann Coutinho, Ricardo Augusto Brassaloti, Raquel de Lello Rocha Campos Cassano. NGS Soluções Genômicas: Pilar Drummond Sampaio Corrêa Mariani. FZEA-USP Pirassununga: Mirele Daiana Poleti, Jessika Cristina Chagas Lesbon, Elisangela Chicaroni Mattos, Heidge Fukumasu. USP-Botucatu: Rejane Maria Tommasini Grotto, Jayme A. Souza-Neto, Guilherme Targino Valente, Patricia Akemi Assato, Felipe Allan da Silva da Costa, Bianca Cechetto Carlos. Mendelics: Bibiana Santos, João Paulo Kitajima, Erika Freitas, David Schlesinger. Hemocentro Ribeirão Preto: Simone Kashima, Evandra Strazza Rodrigues, Svetoslav Nanev Slavov, Elaine Vieira dos Santos, Rafael dos Santos Bezerra, Luiz Carlos Junior de Alcantara, Marta Giovanetti, Vagner Fonseca, Flavia Aburjaile, Rodrigo Tocantins Calado. |
| EPI_ISL_1795287 | UBS ALCIMINIO DE ASSIS LOURENCO BADY BASSITT | Instituto Butantan / ESALQ-Piracicaba | Instituto Butantan: Alexander Roberto Precioso, Dimas Tadeu Covas, Sandra Coccuzzo Sampaio, Maria Carolina Elias, José Salvatore Leister Patané, Vincent Louis Viala, Antonio Jorge Martins, Ricardo Haddad, Claudia Renata dos Santos Barros, Elaine Cristina Marqueze, Raul Machado Neto, Debora Botequio Moretti. Centro de Genômica Funcional da ESALQ: Luiz Lehmann Coutinho, Ricardo Augusto Brassaloti, Raquel de Lello Rocha Campos Cassano. NGS Soluções Genômicas: Pilar Drummond Sampaio Corrêa Mariani. FZEA-USP Pirassununga: Mirele Daiana Poleti, Jessika Cristina Chagas Lesbon, Elisangela Chicaroni Mattos, Heidge Fukumasu. USP-Botucatu: Rejane Maria Tommasini Grotto, Jayme A. Souza-Neto, Guilherme Targino Valente, Patricia Akemi Assato, Felipe Allan da Silva da Costa, Bianca Cechetto Carlos. Mendelics: Bibiana Santos, João Paulo Kitajima, Erika Freitas, David Schlesinger. Hemocentro Ribeirão Preto: Simone Kashima, Evandra Strazza Rodrigues, Svetoslav Nanev Slavov, Elaine Vieira dos Santos, Rafael dos Santos Bezerra, Luiz Carlos Junior de Alcantara, Marta Giovanetti, Vagner Fonseca, Flavia Aburjaile, Rodrigo Tocantins Calado. |
| EPI_ISL_1795288 | CS DE NIPOA | Instituto Butantan / ESALQ-Piracicaba | Instituto Butantan: Alexander Roberto Precioso, Dimas Tadeu Covas, Sandra Coccuzzo Sampaio, Maria Carolina Elias, José Salvatore Leister Patané, Vincent Louis Viala, Antonio Jorge Martins, Ricardo Haddad, Claudia Renata dos Santos Barros, Elaine Cristina Marqueze, Raul Machado Neto, Debora Botequio Moretti. Centro de Genômica Funcional da ESALQ: Luiz Lehmann Coutinho, Ricardo Augusto Brassaloti, Raquel de Lello Rocha Campos Cassano. NGS Soluções Genômicas: Pilar Drummond Sampaio Corrêa Mariani. FZEA-USP Pirassununga: Mirele Daiana Poleti, Jessika Cristina Chagas Lesbon, Elisangela Chicaroni Mattos, Heidge Fukumasu. USP-Botucatu: Rejane Maria Tommasini Grotto, Jayme A. Souza-Neto, Guilherme Targino Valente, Patricia Akemi Assato, Felipe Allan da Silva da Costa, Bianca Cechetto Carlos. Mendelics: Bibiana Santos, João Paulo Kitajima, Erika Freitas, David Schlesinger. Hemocentro Ribeirão Preto: Simone Kashima, Evandra Strazza Rodrigues, Svetoslav Nanev Slavov, Elaine Vieira dos Santos, Rafael dos Santos Bezerra, Luiz Carlos Junior de Alcantara, Marta Giovanetti, Vagner Fonseca, Flavia Aburjaile, Rodrigo Tocantins Calado. |
| EPI_ISL_1795289 | UBS ALCIMINIO DE ASSIS LOURENCO BADY BASSITT | Instituto Butantan / ESALQ-Piracicaba | Instituto Butantan: Alexander Roberto Precioso, Dimas Tadeu Covas, Sandra Coccuzzo Sampaio, Maria Carolina Elias, José Salvatore Leister Patané, Vincent Louis Viala, Antonio Jorge Martins, Ricardo Haddad, Claudia Renata dos Santos Barros, Elaine Cristina Marqueze, Raul Machado Neto, Debora Botequio Moretti. Centro de Genômica Funcional da ESALQ: Luiz Lehmann Coutinho, Ricardo Augusto Brassaloti, Raquel de Lello Rocha Campos Cassano. NGS Soluções Genômicas: Pilar Drummond Sampaio Corrêa Mariani. FZEA-USP Pirassununga: Mirele Daiana Poleti, Jessika Cristina Chagas Lesbon, Elisangela Chicaroni Mattos, Heidge Fukumasu. USP-Botucatu: Rejane Maria Tommasini Grotto, Jayme A. Souza-Neto, Guilherme Targino Valente, Patricia Akemi Assato, Felipe Allan da Silva da Costa, Bianca Cechetto Carlos. Mendelics: Bibiana Santos, João Paulo Kitajima, Erika Freitas, David Schlesinger. Hemocentro Ribeirão Preto: Simone Kashima, Evandra Strazza Rodrigues, Svetoslav Nanev Slavov, Elaine Vieira dos Santos, Rafael dos Santos Bezerra, Luiz Carlos Junior de Alcantara, Marta Giovanetti, Vagner Fonseca, Flavia Aburjaile, Rodrigo Tocantins Calado. |
| EPI_ISL_1795290 | USF EUCALIPTOS | Instituto Butantan / ESALQ-Piracicaba | Instituto Butantan: Alexander Roberto Precioso, Dimas Tadeu Covas, Sandra Coccuzzo Sampaio, Maria Carolina Elias, José Salvatore Leister Patané, Vincent Louis Viala, Antonio Jorge Martins, Ricardo Haddad, Claudia Renata dos Santos Barros, Elaine Cristina Marqueze, Raul Machado Neto, Debora Botequio Moretti. Centro de Genômica Funcional da ESALQ: Luiz Lehmann Coutinho, Ricardo Augusto Brassaloti, Raquel de Lello Rocha Campos Cassano. NGS Soluções Genômicas: Pilar Drummond Sampaio Corrêa Mariani. FZEA-USP Pirassununga: Mirele Daiana Poleti, Jessika Cristina Chagas Lesbon, Elisangela Chicaroni Mattos, Heidge Fukumasu. USP-Botucatu: Rejane Maria Tommasini Grotto, Jayme A. Souza-Neto, Guilherme Targino Valente, Patricia Akemi Assato, Felipe Allan da Silva da Costa, Bianca Cechetto Carlos. Mendelics: Bibiana Santos, João Paulo Kitajima, Erika Freitas, David Schlesinger. Hemocentro Ribeirão Preto: Simone Kashima, Evandra Strazza Rodrigues, Svetoslav Nanev Slavov, Elaine Vieira dos Santos, Rafael dos Santos Bezerra, Luiz Carlos Junior de Alcantara, Marta Giovanetti, Vagner Fonseca, Flavia Aburjaile, Rodrigo Tocantins Calado. |
| EPI_ISL_1795291, EPI_ISL_1795292 | AMBULATORIO DE ESPECIALIDADE V E MOGI MIRIM | Instituto Butantan / ESALQ-Piracicaba | Instituto Butantan: Alexander Roberto Precioso, Dimas Tadeu Covas, Sandra Coccuzzo Sampaio, Maria Carolina Elias, José Salvatore Leister Patané, Vincent Louis Viala, Antonio Jorge Martins, Ricardo Haddad, Claudia Renata dos Santos Barros, Elaine Cristina Marqueze, Raul Machado Neto, Debora Botequio Moretti. Centro de Genômica Funcional da ESALQ: Luiz Lehmann Coutinho, Ricardo Augusto Brassaloti, Raquel de Lello Rocha Campos Cassano. NGS Soluções Genômicas: Pilar Drummond Sampaio Corrêa Mariani. FZEA-USP Pirassununga: Mirele Daiana Poleti, Jessika Cristina Chagas Lesbon, Elisangela Chicaroni Mattos, Heidge Fukumasu. USP-Botucatu: Rejane Maria Tommasini Grotto, Jayme A. Souza-Neto, Guilherme Targino Valente, Patricia Akemi Assato, Felipe Allan da Silva da Costa, Bianca Cechetto Carlos. Mendelics: Bibiana Santos, João Paulo Kitajima, Erika Freitas, |

| | | | |
|---|---|---|---|
| | | | David Schlesinger. Hemocentro Ribeirão Preto: Simone Kashima, Evandra Strazza Rodrigues, Svetoslav Nanev Slavov, Elaine Vieira dos Santos, Rafael dos Santos Bezerra, Luiz Carlos Junior de Alcantara, Marta Giovanetti, Vagner Fonseca, Flavia Aburjaile, Rodrigo Tocantins Calado. |
| EPI_ISL_1795293, EPI_ISL_1795294 | UNIDADE DE SAUDE DE ITOBI ALCIBIADES PIRES | Instituto Butantan / ESALQ-Piracicaba | Instituto Butantan: Alexander Roberto Precioso, Dimas Tadeu Covas, Sandra Coccuzzo Sampaio, Maria Carolina Elias, José Salvatore Leister Patané, Vincent Louis Viala, Antonio Jorge Martins, Ricardo Haddad, Claudia Renata dos Santos Barros, Elaine Cristina Marqueze, Raul Machado Neto, Debora Botequio Moretti. Centro de Genômica Funcional da ESALQ: Luiz Lehmann Coutinho, Ricardo Augusto Brassaloti, Raquel de Lello Rocha Campos Cassano. NGS Soluções Genômicas: Pilar Drummond Sampaio Corrêa Mariani. FZEA-USP Pirassununga: Mirele Daiana Poleti, Jessika Cristina Chagas Lesbon, Elisangela Chicaroni Mattos, Heidge Fukumasu. USP-Botucatu: Rejane Maria Tommasini Grotto, Jayme A. Souza-Neto, Guilherme Targino Valente, Patricia Akemi Assato, Felipe Allan da Silva da Costa, Bianca Cechetto Carlos. Mendelics: Bibiana Santos, João Paulo Kitajima, Erika Freitas, David Schlesinger. Hemocentro Ribeirão Preto: Simone Kashima, Evandra Strazza Rodrigues, Svetoslav Nanev Slavov, Elaine Vieira dos Santos, Rafael dos Santos Bezerra, Luiz Carlos Junior de Alcantara, Marta Giovanetti, Vagner Fonseca, Flavia Aburjaile, Rodrigo Tocantins Calado. |
| EPI_ISL_1795295 | USF GUACUANO | Instituto Butantan / ESALQ-Piracicaba | Instituto Butantan: Alexander Roberto Precioso, Dimas Tadeu Covas, Sandra Coccuzzo Sampaio, Maria Carolina Elias, José Salvatore Leister Patané, Vincent Louis Viala, Antonio Jorge Martins, Ricardo Haddad, Claudia Renata dos Santos Barros, Elaine Cristina Marqueze, Raul Machado Neto, Debora Botequio Moretti. Centro de Genômica Funcional da ESALQ: Luiz Lehmann Coutinho, Ricardo Augusto Brassaloti, Raquel de Lello Rocha Campos Cassano. NGS Soluções Genômicas: Pilar Drummond Sampaio Corrêa Mariani. FZEA-USP Pirassununga: Mirele Daiana Poleti, Jessika Cristina Chagas Lesbon, Elisangela Chicaroni Mattos, Heidge Fukumasu. USP-Botucatu: Rejane Maria Tommasini Grotto, Jayme A. Souza-Neto, Guilherme Targino Valente, Patricia Akemi Assato, Felipe Allan da Silva da Costa, Bianca Cechetto Carlos. Mendelics: Bibiana Santos, João Paulo Kitajima, Erika Freitas, David Schlesinger. Hemocentro Ribeirão Preto: Simone Kashima, Evandra Strazza Rodrigues, Svetoslav Nanev Slavov, Elaine Vieira dos Santos, Rafael dos Santos Bezerra, Luiz Carlos Junior de Alcantara, Marta Giovanetti, Vagner Fonseca, Flavia Aburjaile, Rodrigo Tocantins Calado. |
| EPI_ISL_1795296, EPI_ISL_1795297 | CENTRO DE SAUDE III AFFONSO LUZZI SANTA CRUZ DAS PALMEIRAS | Instituto Butantan / ESALQ-Piracicaba | Instituto Butantan: Alexander Roberto Precioso, Dimas Tadeu Covas, Sandra Coccuzzo Sampaio, Maria Carolina Elias, José Salvatore Leister Patané, Vincent Louis Viala, Antonio Jorge Martins, Ricardo Haddad, Claudia Renata dos Santos Barros, Elaine Cristina Marqueze, Raul Machado Neto, Debora Botequio Moretti. Centro de Genômica Funcional da ESALQ: Luiz Lehmann Coutinho, Ricardo Augusto Brassaloti, Raquel de Lello Rocha Campos Cassano. NGS Soluções Genômicas: Pilar Drummond Sampaio Corrêa Mariani. FZEA-USP Pirassununga: Mirele Daiana Poleti, Jessika Cristina Chagas Lesbon, Elisangela Chicaroni Mattos, Heidge Fukumasu. USP-Botucatu: Rejane Maria Tommasini Grotto, Jayme A. Souza-Neto, Guilherme Targino Valente, Patricia Akemi Assato, Felipe Allan da Silva da Costa, Bianca Cechetto Carlos. Mendelics: Bibiana Santos, João Paulo Kitajima, Erika Freitas, David Schlesinger. Hemocentro Ribeirão Preto: Simone Kashima, Evandra Strazza Rodrigues, Svetoslav Nanev Slavov, Elaine Vieira dos Santos, Rafael dos Santos Bezerra, Luiz Carlos Junior de Alcantara, Marta Giovanetti, Vagner Fonseca, Flavia Aburjaile, Rodrigo Tocantins Calado. |
| EPI_ISL_1795298 | UNIDADE DE SAUDE DE ITOBI ALCIBIADES PIRES | Instituto Butantan / ESALQ-Piracicaba | Instituto Butantan: Alexander Roberto Precioso, Dimas Tadeu Covas, Sandra Coccuzzo Sampaio, Maria Carolina Elias, José Salvatore Leister Patané, Vincent Louis Viala, Antonio Jorge Martins, Ricardo Haddad, Claudia Renata dos Santos Barros, Elaine Cristina Marqueze, Raul Machado Neto, Debora Botequio Moretti. Centro de Genômica Funcional da ESALQ: Luiz Lehmann Coutinho, Ricardo Augusto Brassaloti, Raquel de Lello Rocha Campos Cassano. NGS Soluções Genômicas: Pilar Drummond Sampaio Corrêa Mariani. FZEA-USP Pirassununga: Mirele Daiana Poleti, Jessika Cristina Chagas Lesbon, Elisangela Chicaroni Mattos, Heidge Fukumasu. USP-Botucatu: Rejane Maria Tommasini Grotto, Jayme A. Souza-Neto, Guilherme Targino Valente, Patricia Akemi Assato, Felipe Allan da Silva da Costa, Bianca Cechetto Carlos. Mendelics: Bibiana Santos, João Paulo Kitajima, Erika Freitas, David Schlesinger. Hemocentro Ribeirão Preto: Simone Kashima, Evandra Strazza Rodrigues, Svetoslav Nanev Slavov, Elaine Vieira dos Santos, Rafael dos Santos Bezerra, Luiz Carlos Junior de Alcantara, Marta Giovanetti, Vagner Fonseca, Flavia Aburjaile, Rodrigo Tocantins Calado. |
| EPI_ISL_1795299 | CENTRO DE SAUDE III AFFONSO LUZZI SANTA CRUZ DAS PALMEIRAS | Instituto Butantan / ESALQ-Piracicaba | Instituto Butantan: Alexander Roberto Precioso, Dimas Tadeu Covas, Sandra Coccuzzo Sampaio, Maria Carolina Elias, José Salvatore Leister Patané, Vincent Louis Viala, Antonio Jorge Martins, Ricardo Haddad, Claudia Renata dos Santos Barros, Elaine Cristina Marqueze, Raul Machado Neto, Debora Botequio Moretti. Centro de Genômica Funcional da ESALQ: Luiz Lehmann Coutinho, Ricardo Augusto Brassaloti, Raquel de Lello Rocha Campos Cassano. NGS Soluções Genômicas: Pilar Drummond Sampaio Corrêa Mariani. FZEA-USP Pirassununga: Mirele Daiana Poleti, Jessika Cristina Chagas Lesbon, Elisangela Chicaroni Mattos, Heidge Fukumasu. USP-Botucatu: Rejane Maria Tommasini Grotto, Jayme A. Souza-Neto, Guilherme Targino Valente, Patricia Akemi Assato, Felipe Allan da Silva da Costa, Bianca Cechetto Carlos. Mendelics: Bibiana Santos, João Paulo Kitajima, Erika Freitas, David Schlesinger. Hemocentro Ribeirão Preto: Simone Kashima, Evandra Strazza Rodrigues, Svetoslav Nanev Slavov, Elaine Vieira dos Santos, Rafael dos Santos Bezerra, Luiz Carlos Junior de Alcantara, Marta Giovanetti, Vagner Fonseca, Flavia Aburjaile, Rodrigo Tocantins Calado. |
| EPI_ISL_1795300, EPI_ISL_1795301, EPI_ISL_1795302 | PRONTO SOCORRO MUNICIPAL TAMBAU | Instituto Butantan / ESALQ-Piracicaba | Instituto Butantan: Alexander Roberto Precioso, Dimas Tadeu Covas, Sandra Coccuzzo Sampaio, Maria Carolina Elias, José Salvatore Leister Patané, Vincent Louis Viala, Antonio Jorge Martins, Ricardo Haddad, Claudia Renata dos Santos Barros, Elaine Cristina Marqueze, Raul Machado Neto, Debora Botequio Moretti. Centro de Genômica Funcional da ESALQ: Luiz Lehmann Coutinho, Ricardo Augusto Brassaloti, Raquel de Lello Rocha Campos Cassano. NGS Soluções Genômicas: Pilar Drummond Sampaio Corrêa Mariani. FZEA-USP Pirassununga: Mirele Daiana Poleti, Jessika Cristina Chagas Lesbon, Elisangela Chicaroni Mattos, Heidge Fukumasu. USP-Botucatu: Rejane Maria Tommasini Grotto, Jayme A. Souza-Neto, Guilherme Targino Valente, Patricia Akemi Assato, Felipe Allan da Silva da Costa, Bianca Cechetto Carlos. Mendelics: Bibiana Santos, João Paulo Kitajima, Erika Freitas, David Schlesinger. Hemocentro Ribeirão Preto: Simone Kashima, Evandra Strazza Rodrigues, Svetoslav Nanev Slavov, Elaine Vieira dos Santos, Rafael dos Santos Bezerra, Luiz Carlos Junior de Alcantara, Marta Giovanetti, Vagner Fonseca, Flavia Aburjaile, Rodrigo Tocantins Calado. |
| EPI_ISL_1795303 | LABORATORIO DE FRANCA | Instituto Butantan / ESALQ-Piracicaba | Instituto Butantan: Alexander Roberto Precioso, Dimas Tadeu Covas, Sandra Coccuzzo Sampaio, Maria Carolina Elias, José Salvatore Leister Patané, Vincent Louis Viala, Antonio Jorge Martins, Ricardo Haddad, Claudia Renata dos Santos Barros, Elaine Cristina Marqueze, Raul Machado Neto, Debora Botequio Moretti. Centro de Genômica Funcional da ESALQ: Luiz Lehmann Coutinho, Ricardo Augusto Brassaloti, Raquel de Lello Rocha Campos Cassano. NGS Soluções Genômicas: Pilar Drummond Sampaio Corrêa Mariani. FZEA-USP Pirassununga: Mirele Daiana Poleti, Jessika Cristina Chagas Lesbon, Elisangela Chicaroni Mattos, Heidge Fukumasu. USP-Botucatu: Rejane Maria Tommasini Grotto, Jayme A. Souza-Neto, Guilherme Targino Valente, Patricia Akemi Assato, Felipe Allan da Silva da Costa, Bianca Cechetto Carlos. Mendelics: Bibiana Santos, João Paulo Kitajima, Erika Freitas, David Schlesinger. Hemocentro Ribeirão Preto: Simone Kashima, Evandra Strazza Rodrigues, Svetoslav Nanev Slavov, Elaine Vieira dos Santos, Rafael dos Santos Bezerra, Luiz Carlos Junior de Alcantara, Marta Giovanetti, Vagner Fonseca, Flavia Aburjaile, Rodrigo Tocantins Calado. |
| EPI_ISL_1795304 | PRONTO SOCORRO MUNICIPAL TAMBAU | Instituto Butantan / ESALQ-Piracicaba | Instituto Butantan: Alexander Roberto Precioso, Dimas Tadeu Covas, Sandra Coccuzzo Sampaio, Maria Carolina Elias, José Salvatore Leister Patané, Vincent Louis Viala, Antonio Jorge Martins, Ricardo Haddad, Claudia Renata dos Santos Barros, Elaine Cristina Marqueze, Raul Machado Neto, Debora Botequio Moretti. Centro de Genômica Funcional da ESALQ: Luiz Lehmann Coutinho, Ricardo Augusto Brassaloti, Raquel de Lello Rocha Campos Cassano. NGS Soluções Genômicas: Pilar Drummond Sampaio Corrêa Mariani. FZEA-USP Pirassununga: Mirele Daiana Poleti, Jessika Cristina Chagas Lesbon, Elisangela Chicaroni Mattos, Heidge Fukumasu. USP-Botucatu: Rejane Maria Tommasini Grotto, Jayme A. Souza-Neto, Guilherme Targino Valente, Patricia Akemi Assato, Felipe Allan da Silva da Costa, Bianca Cechetto Carlos. Mendelics: Bibiana Santos, João Paulo Kitajima, Erika Freitas, David Schlesinger. Hemocentro Ribeirão Preto: Simone Kashima, Evandra Strazza Rodrigues, Svetoslav Nanev Slavov, Elaine Vieira dos Santos, Rafael dos Santos Bezerra, Luiz Carlos Junior de Alcantara, Marta Giovanetti, Vagner Fonseca, Flavia Aburjaile, Rodrigo Tocantins Calado. |
| EPI_ISL_1795305 | UNIDADE DA SAUDE DO ADULTO CASA BRANCA PREFEITURA | Instituto Butantan / ESALQ-Piracicaba | Instituto Butantan: Alexander Roberto Precioso, Dimas Tadeu Covas, Sandra Coccuzzo Sampaio, Maria Carolina Elias, José Salvatore Leister Patané, Vincent Louis Viala, Antonio Jorge Martins, Ricardo Haddad, Claudia Renata dos Santos Barros, Elaine Cristina Marqueze, Raul Machado Neto, Debora Botequio Moretti. Centro de Genômica Funcional da ESALQ: Luiz Lehmann Coutinho, Ricardo Augusto Brassaloti, Raquel de Lello Rocha Campos Cassano. NGS Soluções Genômicas: Pilar Drummond Sampaio Corrêa Mariani. FZEA-USP Pirassununga: Mirele Daiana Poleti, Jessika Cristina Chagas Lesbon, Elisangela Chicaroni Mattos, Heidge Fukumasu. USP-Botucatu: Rejane Maria Tommasini Grotto, Jayme A. Souza-Neto, Guilherme Targino Valente, Patricia Akemi Assato, Felipe Allan da Silva da Costa, Bianca Cechetto Carlos. Mendelics: Bibiana Santos, João Paulo Kitajima, Erika Freitas, David Schlesinger. Hemocentro Ribeirão Preto: Simone Kashima, Evandra Strazza Rodrigues, Svetoslav Nanev Slavov, Elaine Vieira dos Santos, Rafael dos Santos Bezerra, Luiz Carlos Junior de Alcantara, Marta Giovanetti, Vagner Fonseca, Flavia Aburjaile, Rodrigo Tocantins Calado. |

EPI_ISL_1795306, EPI_ISL_1795307, EPI_ISL_1795308, EPI_ISL_1795309, EPI_ISL_1795310, EPI_ISL_1795311, EPI_ISL_1795312, EPI_ISL_1795313, EPI_ISL_1795314, EPI_ISL_1795315, EPI_ISL_1795316, EPI_ISL_1795317, EPI_ISL_1795318, EPI_ISL_1795319, EPI_ISL_1795320, EPI_ISL_1795321, EPI_ISL_1795322, EPI_ISL_1795323

| | | | |
|---|---|---|---|
| see above | LABORATORIO DE FRANCA | Instituto Butantan / ESALQ-Piracicaba | Instituto Butantan: Alexander Roberto Precioso, Dimas Tadeu Covas, Sandra Coccuzzo Sampaio, Maria Carolina Elias, José Salvatore Leister Patané, Vincent Louis Viala, Antonio Jorge Martins, Ricardo Haddad, Claudia Renata dos Santos Barros, Elaine Cristina Marqueze, Raul Machado Neto, Debora Botequio Moretti. Centro de Genômica Funcional da ESALQ: Luiz Lehmann Coutinho, Ricardo Augusto Brassaloti, Raquel de Lello Rocha Campos Cassano. NGS Soluções Genômicas: Pilar Drummond Sampaio Corrêa Mariani. FZEA-USP Pirassununga: Mirele Daiana Poleti, Jessika Cristina Chagas Lesbon, Elisangela Chicaroni Mattos, Heidge Fukumasu. USP-Botucatu: Rejane Maria Tommasini Grotto, Jayme A. Souza-Neto, Guilherme Targino Valente, Patricia Akemi Assato, Felipe Allan da Silva da Costa, Bianca Cechetto Carlos. Mendelics: Bibiana Santos, João Paulo Kitajima, Erika Freitas, |

| | | | |
|---|---|---|---|
| | | | David Schlesinger. Hemocentro Ribeirão Preto: Simone Kashima, Evandra Strazza Rodrigues, Svetoslav Nanev Slavov, Elaine Vieira dos Santos, Rafael dos Santos Bezerra, Luiz Carlos Junior de Alcantara, Marta Giovanetti, Vagner Fonseca, Flavia Aburjaile, Rodrigo Tocantins Calado. |
| EPI_ISL_1795324 | UBS II DE NARANDIBA | Instituto Butantan / ESALQ-Piracicaba | Instituto Butantan: Alexander Roberto Precioso, Dimas Tadeu Covas, Sandra Coccuzzo Sampaio, Maria Carolina Elias, José Salvatore Leister Patané, Vincent Louis Viala, Antonio Jorge Martins, Ricardo Haddad, Claudia Renata dos Santos Barros, Elaine Cristina Marqueze, Raul Machado Neto, Debora Botequio Moretti. Centro de Genômica Funcional da ESALQ: Luiz Lehmann Coutinho, Ricardo Augusto Brassaloti, Raquel de Lello Rocha Campos Cassano. NGS Soluções Genômicas: Pilar Drummond Sampaio Corrêa Mariani. FZEA-USP Pirassununga: Mirele Daiana Poleti, Jessika Cristina Chagas Lesbon, Elisangela Chicaroni Mattos, Heidge Fukumasu. USP-Botucatu: Rejane Maria Tommasini Grotto, Jayme A. Souza-Neto, Guilherme Targino Valente, Patricia Akemi Assato, Felipe Allan da Silva da Costa, Bianca Cechetto Carlos. Mendelics: Bibiana Santos, João Paulo Kitajima, Erika Freitas, David Schlesinger. Hemocentro Ribeirão Preto: Simone Kashima, Evandra Strazza Rodrigues, Svetoslav Nanev Slavov, Elaine Vieira dos Santos, Rafael dos Santos Bezerra, Luiz Carlos Junior de Alcantara, Marta Giovanetti, Vagner Fonseca, Flavia Aburjaile, Rodrigo Tocantins Calado. |
| EPI_ISL_1795325 | CENTRO DE ESPECIALIDADES DE PRIMAVERA | Instituto Butantan / ESALQ-Piracicaba | Instituto Butantan: Alexander Roberto Precioso, Dimas Tadeu Covas, Sandra Coccuzzo Sampaio, Maria Carolina Elias, José Salvatore Leister Patané, Vincent Louis Viala, Antonio Jorge Martins, Ricardo Haddad, Claudia Renata dos Santos Barros, Elaine Cristina Marqueze, Raul Machado Neto, Debora Botequio Moretti. Centro de Genômica Funcional da ESALQ: Luiz Lehmann Coutinho, Ricardo Augusto Brassaloti, Raquel de Lello Rocha Campos Cassano. NGS Soluções Genômicas: Pilar Drummond Sampaio Corrêa Mariani. FZEA-USP Pirassununga: Mirele Daiana Poleti, Jessika Cristina Chagas Lesbon, Elisangela Chicaroni Mattos, Heidge Fukumasu. USP-Botucatu: Rejane Maria Tommasini Grotto, Jayme A. Souza-Neto, Guilherme Targino Valente, Patricia Akemi Assato, Felipe Allan da Silva da Costa, Bianca Cechetto Carlos. Mendelics: Bibiana Santos, João Paulo Kitajima, Erika Freitas, David Schlesinger. Hemocentro Ribeirão Preto: Simone Kashima, Evandra Strazza Rodrigues, Svetoslav Nanev Slavov, Elaine Vieira dos Santos, Rafael dos Santos Bezerra, Luiz Carlos Junior de Alcantara, Marta Giovanetti, Vagner Fonseca, Flavia Aburjaile, Rodrigo Tocantins Calado. |
| EPI_ISL_1795326 | UBS II DE REGENTE FEIJO | Instituto Butantan / ESALQ-Piracicaba | Instituto Butantan: Alexander Roberto Precioso, Dimas Tadeu Covas, Sandra Coccuzzo Sampaio, Maria Carolina Elias, José Salvatore Leister Patané, Vincent Louis Viala, Antonio Jorge Martins, Ricardo Haddad, Claudia Renata dos Santos Barros, Elaine Cristina Marqueze, Raul Machado Neto, Debora Botequio Moretti. Centro de Genômica Funcional da ESALQ: Luiz Lehmann Coutinho, Ricardo Augusto Brassaloti, Raquel de Lello Rocha Campos Cassano. NGS Soluções Genômicas: Pilar Drummond Sampaio Corrêa Mariani. FZEA-USP Pirassununga: Mirele Daiana Poleti, Jessika Cristina Chagas Lesbon, Elisangela Chicaroni Mattos, Heidge Fukumasu. USP-Botucatu: Rejane Maria Tommasini Grotto, Jayme A. Souza-Neto, Guilherme Targino Valente, Patricia Akemi Assato, Felipe Allan da Silva da Costa, Bianca Cechetto Carlos. Mendelics: Bibiana Santos, João Paulo Kitajima, Erika Freitas, David Schlesinger. Hemocentro Ribeirão Preto: Simone Kashima, Evandra Strazza Rodrigues, Svetoslav Nanev Slavov, Elaine Vieira dos Santos, Rafael dos Santos Bezerra, Luiz Carlos Junior de Alcantara, Marta Giovanetti, Vagner Fonseca, Flavia Aburjaile, Rodrigo Tocantins Calado. |
| EPI_ISL_1795327 | CENTRO DE SAUDE II JUNQUEIROPOLIS | Instituto Butantan / ESALQ-Piracicaba | Instituto Butantan: Alexander Roberto Precioso, Dimas Tadeu Covas, Sandra Coccuzzo Sampaio, Maria Carolina Elias, José Salvatore Leister Patané, Vincent Louis Viala, Antonio Jorge Martins, Ricardo Haddad, Claudia Renata dos Santos Barros, Elaine Cristina Marqueze, Raul Machado Neto, Debora Botequio Moretti. Centro de Genômica Funcional da ESALQ: Luiz Lehmann Coutinho, Ricardo Augusto Brassaloti, Raquel de Lello Rocha Campos Cassano. NGS Soluções Genômicas: Pilar Drummond Sampaio Corrêa Mariani. FZEA-USP Pirassununga: Mirele Daiana Poleti, Jessika Cristina Chagas Lesbon, Elisangela Chicaroni Mattos, Heidge Fukumasu. USP-Botucatu: Rejane Maria Tommasini Grotto, Jayme A. Souza-Neto, Guilherme Targino Valente, Patricia Akemi Assato, Felipe Allan da Silva da Costa, Bianca Cechetto Carlos. Mendelics: Bibiana Santos, João Paulo Kitajima, Erika Freitas, David Schlesinger. Hemocentro Ribeirão Preto: Simone Kashima, Evandra Strazza Rodrigues, Svetoslav Nanev Slavov, Elaine Vieira dos Santos, Rafael dos Santos Bezerra, Luiz Carlos Junior de Alcantara, Marta Giovanetti, Vagner Fonseca, Flavia Aburjaile, Rodrigo Tocantins Calado. |
| EPI_ISL_1795328 | UBS II DR EXPEDITO SHIZUO KUROCE | Instituto Butantan / ESALQ-Piracicaba | Instituto Butantan: Alexander Roberto Precioso, Dimas Tadeu Covas, Sandra Coccuzzo Sampaio, Maria Carolina Elias, José Salvatore Leister Patané, Vincent Louis Viala, Antonio Jorge Martins, Ricardo Haddad, Claudia Renata dos Santos Barros, Elaine Cristina Marqueze, Raul Machado Neto, Debora Botequio Moretti. Centro de Genômica Funcional da ESALQ: Luiz Lehmann Coutinho, Ricardo Augusto Brassaloti, Raquel de Lello Rocha Campos Cassano. NGS Soluções Genômicas: Pilar Drummond Sampaio Corrêa Mariani. FZEA-USP Pirassununga: Mirele Daiana Poleti, Jessika Cristina Chagas Lesbon, Elisangela Chicaroni Mattos, Heidge Fukumasu. USP-Botucatu: Rejane Maria Tommasini Grotto, Jayme A. Souza-Neto, Guilherme Targino Valente, Patricia Akemi Assato, Felipe Allan da Silva da Costa, Bianca Cechetto Carlos. Mendelics: Bibiana Santos, João Paulo Kitajima, Erika Freitas, David Schlesinger. Hemocentro Ribeirão Preto: Simone Kashima, Evandra Strazza Rodrigues, Svetoslav Nanev Slavov, Elaine Vieira dos Santos, Rafael dos Santos Bezerra, Luiz Carlos Junior de Alcantara, Marta Giovanetti, Vagner Fonseca, Flavia Aburjaile, Rodrigo Tocantins Calado. |
| EPI_ISL_1795329 | CENTRO DE ESPECIALIDADES DE PRIMAVERA | Instituto Butantan / ESALQ-Piracicaba | Instituto Butantan: Alexander Roberto Precioso, Dimas Tadeu Covas, Sandra Coccuzzo Sampaio, Maria Carolina Elias, José Salvatore Leister Patané, Vincent Louis Viala, Antonio Jorge Martins, Ricardo Haddad, Claudia Renata dos Santos Barros, Elaine Cristina Marqueze, Raul Machado Neto, Debora Botequio Moretti. Centro de Genômica Funcional da ESALQ: Luiz Lehmann Coutinho, Ricardo Augusto Brassaloti, Raquel de Lello Rocha Campos Cassano. NGS Soluções Genômicas: Pilar Drummond Sampaio Corrêa Mariani. FZEA-USP Pirassununga: Mirele Daiana Poleti, Jessika Cristina Chagas Lesbon, Elisangela Chicaroni Mattos, Heidge Fukumasu. USP-Botucatu: Rejane Maria Tommasini Grotto, Jayme A. Souza-Neto, Guilherme Targino Valente, Patricia Akemi Assato, Felipe Allan da Silva da Costa, Bianca Cechetto Carlos. Mendelics: Bibiana Santos, João Paulo Kitajima, Erika Freitas, David Schlesinger. Hemocentro Ribeirão Preto: Simone Kashima, Evandra Strazza Rodrigues, Svetoslav Nanev Slavov, Elaine Vieira dos Santos, Rafael dos Santos Bezerra, Luiz Carlos Junior de Alcantara, Marta Giovanetti, Vagner Fonseca, Flavia Aburjaile, Rodrigo Tocantins Calado. |
| EPI_ISL_1795330 | UBS II DR EXPEDITO SHIZUO KUROCE | Instituto Butantan / ESALQ-Piracicaba | Instituto Butantan: Alexander Roberto Precioso, Dimas Tadeu Covas, Sandra Coccuzzo Sampaio, Maria Carolina Elias, José Salvatore Leister Patané, Vincent Louis Viala, Antonio Jorge Martins, Ricardo Haddad, Claudia Renata dos Santos Barros, Elaine Cristina Marqueze, Raul Machado Neto, Debora Botequio Moretti. Centro de Genômica Funcional da ESALQ: Luiz Lehmann Coutinho, Ricardo Augusto Brassaloti, Raquel de Lello Rocha Campos Cassano. NGS Soluções Genômicas: Pilar Drummond Sampaio Corrêa Mariani. FZEA-USP Pirassununga: Mirele Daiana Poleti, Jessika Cristina Chagas Lesbon, Elisangela Chicaroni Mattos, Heidge Fukumasu. USP-Botucatu: Rejane Maria Tommasini Grotto, Jayme A. Souza-Neto, Guilherme Targino Valente, Patricia Akemi Assato, Felipe Allan da Silva da Costa, Bianca Cechetto Carlos. Mendelics: Bibiana Santos, João Paulo Kitajima, Erika Freitas, David Schlesinger. Hemocentro Ribeirão Preto: Simone Kashima, Evandra Strazza Rodrigues, Svetoslav Nanev Slavov, Elaine Vieira dos Santos, Rafael dos Santos Bezerra, Luiz Carlos Junior de Alcantara, Marta Giovanetti, Vagner Fonseca, Flavia Aburjaile, Rodrigo Tocantins Calado. |
| EPI_ISL_1795331 | CENTRO DE SAUDE II JUNQUEIROPOLIS | Instituto Butantan / ESALQ-Piracicaba | Instituto Butantan: Alexander Roberto Precioso, Dimas Tadeu Covas, Sandra Coccuzzo Sampaio, Maria Carolina Elias, José Salvatore Leister Patané, Vincent Louis Viala, Antonio Jorge Martins, Ricardo Haddad, Claudia Renata dos Santos Barros, Elaine Cristina Marqueze, Raul Machado Neto, Debora Botequio Moretti. Centro de Genômica Funcional da ESALQ: Luiz Lehmann Coutinho, Ricardo Augusto Brassaloti, Raquel de Lello Rocha Campos Cassano. NGS Soluções Genômicas: Pilar Drummond Sampaio Corrêa Mariani. FZEA-USP Pirassununga: Mirele Daiana Poleti, Jessika Cristina Chagas Lesbon, Elisangela Chicaroni Mattos, Heidge Fukumasu. USP-Botucatu: Rejane Maria Tommasini Grotto, Jayme A. Souza-Neto, Guilherme Targino Valente, Patricia Akemi Assato, Felipe Allan da Silva da Costa, Bianca Cechetto Carlos. Mendelics: Bibiana Santos, João Paulo Kitajima, Erika Freitas, David Schlesinger. Hemocentro Ribeirão Preto: Simone Kashima, Evandra Strazza Rodrigues, Svetoslav Nanev Slavov, Elaine Vieira dos Santos, Rafael dos Santos Bezerra, Luiz Carlos Junior de Alcantara, Marta Giovanetti, Vagner Fonseca, Flavia Aburjaile, Rodrigo Tocantins Calado. |
| EPI_ISL_1795332 | UBS III DE RANCHARIA | Instituto Butantan / ESALQ-Piracicaba | Instituto Butantan: Alexander Roberto Precioso, Dimas Tadeu Covas, Sandra Coccuzzo Sampaio, Maria Carolina Elias, José Salvatore Leister Patané, Vincent Louis Viala, Antonio Jorge Martins, Ricardo Haddad, Claudia Renata dos Santos Barros, Elaine Cristina Marqueze, Raul Machado Neto, Debora Botequio Moretti. Centro de Genômica Funcional da ESALQ: Luiz Lehmann Coutinho, Ricardo Augusto Brassaloti, Raquel de Lello Rocha Campos Cassano. NGS Soluções Genômicas: Pilar Drummond Sampaio Corrêa Mariani. FZEA-USP Pirassununga: Mirele Daiana Poleti, Jessika Cristina Chagas Lesbon, Elisangela Chicaroni Mattos, Heidge Fukumasu. USP-Botucatu: Rejane Maria Tommasini Grotto, Jayme A. Souza-Neto, Guilherme Targino Valente, Patricia Akemi Assato, Felipe Allan da Silva da Costa, Bianca Cechetto Carlos. Mendelics: Bibiana Santos, João Paulo Kitajima, Erika Freitas, David Schlesinger. Hemocentro Ribeirão Preto: Simone Kashima, Evandra Strazza Rodrigues, Svetoslav Nanev Slavov, Elaine Vieira dos Santos, Rafael dos Santos Bezerra, Luiz Carlos Junior de Alcantara, Marta Giovanetti, Vagner Fonseca, Flavia Aburjaile, Rodrigo Tocantins Calado. |
| EPI_ISL_1795333 | CENTRO DE ESPECIALIDADES DE PRIMAVERA | Instituto Butantan / ESALQ-Piracicaba | Instituto Butantan: Alexander Roberto Precioso, Dimas Tadeu Covas, Sandra Coccuzzo Sampaio, Maria Carolina Elias, José Salvatore Leister Patané, Vincent Louis Viala, Antonio Jorge Martins, Ricardo Haddad, Claudia Renata dos Santos Barros, Elaine Cristina Marqueze, Raul Machado Neto, Debora Botequio Moretti. Centro de Genômica Funcional da ESALQ: Luiz Lehmann Coutinho, Ricardo Augusto Brassaloti, Raquel de Lello Rocha Campos Cassano. NGS Soluções Genômicas: Pilar Drummond Sampaio Corrêa Mariani. FZEA-USP Pirassununga: Mirele Daiana Poleti, Jessika Cristina Chagas Lesbon, Elisangela Chicaroni Mattos, Heidge Fukumasu. USP-Botucatu: Rejane Maria Tommasini Grotto, Jayme A. Souza-Neto, Guilherme Targino Valente, Patricia Akemi Assato, Felipe Allan da Silva da Costa, Bianca Cechetto Carlos. Mendelics: Bibiana Santos, João Paulo Kitajima, Erika Freitas, David Schlesinger. Hemocentro Ribeirão Preto: Simone Kashima, Evandra Strazza Rodrigues, Svetoslav Nanev Slavov, Elaine Vieira dos Santos, Rafael dos Santos Bezerra, Luiz Carlos Junior de Alcantara, Marta Giovanetti, Vagner Fonseca, Flavia Aburjaile, Rodrigo Tocantins Calado. |

| | | | |
|---|---|---|---|
| EPI_ISL_1795334 | UBS MARCIA CRISTIANE DA SILVA DE OURO VERDE | Instituto Butantan / ESALQ-Piracicaba | Instituto Butantan: Alexander Roberto Precioso, Dimas Tadeu Covas, Sandra Coccuzzo Sampaio, Maria Carolina Elias, José Salvatore Leister Patané, Vincent Louis Viala, Antonio Jorge Martins, Ricardo Haddad, Claudia Renata dos Santos Barros, Elaine Cristina Marqueze, Raul Machado Neto, Debora Botequio Moretti. Centro de Genômica Funcional da ESALQ: Luiz Lehmann Coutinho, Ricardo Augusto Brassaloti, Raquel de Lello Rocha Campos Cassano. NGS Soluções Genômicas: Pilar Drummond Sampaio Corrêa Mariani. FZEA-USP Pirassununga: Mirele Daiana Poleti, Jessika Cristina Chagas Lesbon, Elisangela Chicaroni Mattos, Heidge Fukumasu. USP-Botucatu: Rejane Maria Tommasini Grotto, Jayme A. Souza-Neto, Guilherme Targino Valente, Patricia Akemi Assato, Felipe Allan da Silva da Costa, Bianca Cechetto Carlos. Mendelics: Bibiana Santos, João Paulo Kitajima, Erika Freitas, David Schlesinger. Hemocentro Ribeirão Preto: Simone Kashima, Evandra Strazza Rodrigues, Svetoslav Nanev Slavov, Elaine Vieira dos Santos, Rafael dos Santos Bezerra, Luiz Carlos Junior de Alcantara, Marta Giovanetti, Vagner Fonseca, Flavia Aburjaile, Rodrigo Tocantins Calado. |
| EPI_ISL_1795335 | UBS III DE RANCHARIA | Instituto Butantan / ESALQ-Piracicaba | Instituto Butantan: Alexander Roberto Precioso, Dimas Tadeu Covas, Sandra Coccuzzo Sampaio, Maria Carolina Elias, José Salvatore Leister Patané, Vincent Louis Viala, Antonio Jorge Martins, Ricardo Haddad, Claudia Renata dos Santos Barros, Elaine Cristina Marqueze, Raul Machado Neto, Debora Botequio Moretti. Centro de Genômica Funcional da ESALQ: Luiz Lehmann Coutinho, Ricardo Augusto Brassaloti, Raquel de Lello Rocha Campos Cassano. NGS Soluções Genômicas: Pilar Drummond Sampaio Corrêa Mariani. FZEA-USP Pirassununga: Mirele Daiana Poleti, Jessika Cristina Chagas Lesbon, Elisangela Chicaroni Mattos, Heidge Fukumasu. USP-Botucatu: Rejane Maria Tommasini Grotto, Jayme A. Souza-Neto, Guilherme Targino Valente, Patricia Akemi Assato, Felipe Allan da Silva da Costa, Bianca Cechetto Carlos. Mendelics: Bibiana Santos, João Paulo Kitajima, Erika Freitas, David Schlesinger. Hemocentro Ribeirão Preto: Simone Kashima, Evandra Strazza Rodrigues, Svetoslav Nanev Slavov, Elaine Vieira dos Santos, Rafael dos Santos Bezerra, Luiz Carlos Junior de Alcantara, Marta Giovanetti, Vagner Fonseca, Flavia Aburjaile, Rodrigo Tocantins Calado. |
| EPI_ISL_1795336 | PAS JOAO ANTONIO DO NASCIMENTO | Instituto Butantan / ESALQ-Piracicaba | Instituto Butantan: Alexander Roberto Precioso, Dimas Tadeu Covas, Sandra Coccuzzo Sampaio, Maria Carolina Elias, José Salvatore Leister Patané, Vincent Louis Viala, Antonio Jorge Martins, Ricardo Haddad, Claudia Renata dos Santos Barros, Elaine Cristina Marqueze, Raul Machado Neto, Debora Botequio Moretti. Centro de Genômica Funcional da ESALQ: Luiz Lehmann Coutinho, Ricardo Augusto Brassaloti, Raquel de Lello Rocha Campos Cassano. NGS Soluções Genômicas: Pilar Drummond Sampaio Corrêa Mariani. FZEA-USP Pirassununga: Mirele Daiana Poleti, Jessika Cristina Chagas Lesbon, Elisangela Chicaroni Mattos, Heidge Fukumasu. USP-Botucatu: Rejane Maria Tommasini Grotto, Jayme A. Souza-Neto, Guilherme Targino Valente, Patricia Akemi Assato, Felipe Allan da Silva da Costa, Bianca Cechetto Carlos. Mendelics: Bibiana Santos, João Paulo Kitajima, Erika Freitas, David Schlesinger. Hemocentro Ribeirão Preto: Simone Kashima, Evandra Strazza Rodrigues, Svetoslav Nanev Slavov, Elaine Vieira dos Santos, Rafael dos Santos Bezerra, Luiz Carlos Junior de Alcantara, Marta Giovanetti, Vagner Fonseca, Flavia Aburjaile, Rodrigo Tocantins Calado. |
| EPI_ISL_1795337, EPI_ISL_1795338 | HOSPITAL MUNICIPAL REYNALDO GUERRA CAJATI | Instituto Butantan / ESALQ-Piracicaba | Instituto Butantan: Alexander Roberto Precioso, Dimas Tadeu Covas, Sandra Coccuzzo Sampaio, Maria Carolina Elias, José Salvatore Leister Patané, Vincent Louis Viala, Antonio Jorge Martins, Ricardo Haddad, Claudia Renata dos Santos Barros, Elaine Cristina Marqueze, Raul Machado Neto, Debora Botequio Moretti. Centro de Genômica Funcional da ESALQ: Luiz Lehmann Coutinho, Ricardo Augusto Brassaloti, Raquel de Lello Rocha Campos Cassano. NGS Soluções Genômicas: Pilar Drummond Sampaio Corrêa Mariani. FZEA-USP Pirassununga: Mirele Daiana Poleti, Jessika Cristina Chagas Lesbon, Elisangela Chicaroni Mattos, Heidge Fukumasu. USP-Botucatu: Rejane Maria Tommasini Grotto, Jayme A. Souza-Neto, Guilherme Targino Valente, Patricia Akemi Assato, Felipe Allan da Silva da Costa, Bianca Cechetto Carlos. Mendelics: Bibiana Santos, João Paulo Kitajima, Erika Freitas, David Schlesinger. Hemocentro Ribeirão Preto: Simone Kashima, Evandra Strazza Rodrigues, Svetoslav Nanev Slavov, Elaine Vieira dos Santos, Rafael dos Santos Bezerra, Luiz Carlos Junior de Alcantara, Marta Giovanetti, Vagner Fonseca, Flavia Aburjaile, Rodrigo Tocantins Calado. |
| EPI_ISL_1795339, EPI_ISL_1795340 | VIGILANCIA EM SAUDE | Instituto Butantan / ESALQ-Piracicaba | Instituto Butantan: Alexander Roberto Precioso, Dimas Tadeu Covas, Sandra Coccuzzo Sampaio, Maria Carolina Elias, José Salvatore Leister Patané, Vincent Louis Viala, Antonio Jorge Martins, Ricardo Haddad, Claudia Renata dos Santos Barros, Elaine Cristina Marqueze, Raul Machado Neto, Debora Botequio Moretti. Centro de Genômica Funcional da ESALQ: Luiz Lehmann Coutinho, Ricardo Augusto Brassaloti, Raquel de Lello Rocha Campos Cassano. NGS Soluções Genômicas: Pilar Drummond Sampaio Corrêa Mariani. FZEA-USP Pirassununga: Mirele Daiana Poleti, Jessika Cristina Chagas Lesbon, Elisangela Chicaroni Mattos, Heidge Fukumasu. USP-Botucatu: Rejane Maria Tommasini Grotto, Jayme A. Souza-Neto, Guilherme Targino Valente, Patricia Akemi Assato, Felipe Allan da Silva da Costa, Bianca Cechetto Carlos. Mendelics: Bibiana Santos, João Paulo Kitajima, Erika Freitas, David Schlesinger. Hemocentro Ribeirão Preto: Simone Kashima, Evandra Strazza Rodrigues, Svetoslav Nanev Slavov, Elaine Vieira dos Santos, Rafael dos Santos Bezerra, Luiz Carlos Junior de Alcantara, Marta Giovanetti, Vagner Fonseca, Flavia Aburjaile, Rodrigo Tocantins Calado. |
| EPI_ISL_1795341 | HOSPITAL MUNICIPAL REYNALDO GUERRA CAJATI | Instituto Butantan / ESALQ-Piracicaba | Instituto Butantan: Alexander Roberto Precioso, Dimas Tadeu Covas, Sandra Coccuzzo Sampaio, Maria Carolina Elias, José Salvatore Leister Patané, Vincent Louis Viala, Antonio Jorge Martins, Ricardo Haddad, Claudia Renata dos Santos Barros, Elaine Cristina Marqueze, Raul Machado Neto, Debora Botequio Moretti. Centro de Genômica Funcional da ESALQ: Luiz Lehmann Coutinho, Ricardo Augusto Brassaloti, Raquel de Lello Rocha Campos Cassano. NGS Soluções Genômicas: Pilar Drummond Sampaio Corrêa Mariani. FZEA-USP Pirassununga: Mirele Daiana Poleti, Jessika Cristina Chagas Lesbon, Elisangela Chicaroni Mattos, Heidge Fukumasu. USP-Botucatu: Rejane Maria Tommasini Grotto, Jayme A. Souza-Neto, Guilherme Targino Valente, Patricia Akemi Assato, Felipe Allan da Silva da Costa, Bianca Cechetto Carlos. Mendelics: Bibiana Santos, João Paulo Kitajima, Erika Freitas, David Schlesinger. Hemocentro Ribeirão Preto: Simone Kashima, Evandra Strazza Rodrigues, Svetoslav Nanev Slavov, Elaine Vieira dos Santos, Rafael dos Santos Bezerra, Luiz Carlos Junior de Alcantara, Marta Giovanetti, Vagner Fonseca, Flavia Aburjaile, Rodrigo Tocantins Calado. |
| EPI_ISL_1795342, EPI_ISL_1795343 | VIGILANCIA EM SAUDE | Instituto Butantan / ESALQ-Piracicaba | Instituto Butantan: Alexander Roberto Precioso, Dimas Tadeu Covas, Sandra Coccuzzo Sampaio, Maria Carolina Elias, José Salvatore Leister Patané, Vincent Louis Viala, Antonio Jorge Martins, Ricardo Haddad, Claudia Renata dos Santos Barros, Elaine Cristina Marqueze, Raul Machado Neto, Debora Botequio Moretti. Centro de Genômica Funcional da ESALQ: Luiz Lehmann Coutinho, Ricardo Augusto Brassaloti, Raquel de Lello Rocha Campos Cassano. NGS Soluções Genômicas: Pilar Drummond Sampaio Corrêa Mariani. FZEA-USP Pirassununga: Mirele Daiana Poleti, Jessika Cristina Chagas Lesbon, Elisangela Chicaroni Mattos, Heidge Fukumasu. USP-Botucatu: Rejane Maria Tommasini Grotto, Jayme A. Souza-Neto, Guilherme Targino Valente, Patricia Akemi Assato, Felipe Allan da Silva da Costa, Bianca Cechetto Carlos. Mendelics: Bibiana Santos, João Paulo Kitajima, Erika Freitas, David Schlesinger. Hemocentro Ribeirão Preto: Simone Kashima, Evandra Strazza Rodrigues, Svetoslav Nanev Slavov, Elaine Vieira dos Santos, Rafael dos Santos Bezerra, Luiz Carlos Junior de Alcantara, Marta Giovanetti, Vagner Fonseca, Flavia Aburjaile, Rodrigo Tocantins Calado. |
| EPI_ISL_1795344 | HOSPITAL E MATERNIDADE NOSSA SENHORA DA AJUDA | Instituto Butantan / ESALQ-Piracicaba | Instituto Butantan: Alexander Roberto Precioso, Dimas Tadeu Covas, Sandra Coccuzzo Sampaio, Maria Carolina Elias, José Salvatore Leister Patané, Vincent Louis Viala, Antonio Jorge Martins, Ricardo Haddad, Claudia Renata dos Santos Barros, Elaine Cristina Marqueze, Raul Machado Neto, Debora Botequio Moretti. Centro de Genômica Funcional da ESALQ: Luiz Lehmann Coutinho, Ricardo Augusto Brassaloti, Raquel de Lello Rocha Campos Cassano. NGS Soluções Genômicas: Pilar Drummond Sampaio Corrêa Mariani. FZEA-USP Pirassununga: Mirele Daiana Poleti, Jessika Cristina Chagas Lesbon, Elisangela Chicaroni Mattos, Heidge Fukumasu. USP-Botucatu: Rejane Maria Tommasini Grotto, Jayme A. Souza-Neto, Guilherme Targino Valente, Patricia Akemi Assato, Felipe Allan da Silva da Costa, Bianca Cechetto Carlos. Mendelics: Bibiana Santos, João Paulo Kitajima, Erika Freitas, David Schlesinger. Hemocentro Ribeirão Preto: Simone Kashima, Evandra Strazza Rodrigues, Svetoslav Nanev Slavov, Elaine Vieira dos Santos, Rafael dos Santos Bezerra, Luiz Carlos Junior de Alcantara, Marta Giovanetti, Vagner Fonseca, Flavia Aburjaile, Rodrigo Tocantins Calado. |
| EPI_ISL_1795345 | HOSPITAL DE CAMPANHA COVID 19 MUNICIPIO DE TAUBATE | Instituto Butantan / ESALQ-Piracicaba | Instituto Butantan: Alexander Roberto Precioso, Dimas Tadeu Covas, Sandra Coccuzzo Sampaio, Maria Carolina Elias, José Salvatore Leister Patané, Vincent Louis Viala, Antonio Jorge Martins, Ricardo Haddad, Claudia Renata dos Santos Barros, Elaine Cristina Marqueze, Raul Machado Neto, Debora Botequio Moretti. Centro de Genômica Funcional da ESALQ: Luiz Lehmann Coutinho, Ricardo Augusto Brassaloti, Raquel de Lello Rocha Campos Cassano. NGS Soluções Genômicas: Pilar Drummond Sampaio Corrêa Mariani. FZEA-USP Pirassununga: Mirele Daiana Poleti, Jessika Cristina Chagas Lesbon, Elisangela Chicaroni Mattos, Heidge Fukumasu. USP-Botucatu: Rejane Maria Tommasini Grotto, Jayme A. Souza-Neto, Guilherme Targino Valente, Patricia Akemi Assato, Felipe Allan da Silva da Costa, Bianca Cechetto Carlos. Mendelics: Bibiana Santos, João Paulo Kitajima, Erika Freitas, David Schlesinger. Hemocentro Ribeirão Preto: Simone Kashima, Evandra Strazza Rodrigues, Svetoslav Nanev Slavov, Elaine Vieira dos Santos, Rafael dos Santos Bezerra, Luiz Carlos Junior de Alcantara, Marta Giovanetti, Vagner Fonseca, Flavia Aburjaile, Rodrigo Tocantins Calado. |
| EPI_ISL_1795346, EPI_ISL_1795347, EPI_ISL_1795348, EPI_ISL_1795349, EPI_ISL_1795350, EPI_ISL_1795351, EPI_ISL_1795352, EPI_ISL_1795353, EPI_ISL_1795354, EPI_ISL_1795355, EPI_ISL_1795356, EPI_ISL_1795357, EPI_ISL_1795358, EPI_ISL_1795359, EPI_ISL_1795360 | | | |
| see above | LABORATORIO DR PAULO EMILIO DALESSANDRO PINDAMONHANGABA | Instituto Butantan / ESALQ-Piracicaba | Instituto Butantan: Alexander Roberto Precioso, Dimas Tadeu Covas, Sandra Coccuzzo Sampaio, Maria Carolina Elias, José Salvatore Leister Patané, Vincent Louis Viala, Antonio Jorge Martins, Ricardo Haddad, Claudia Renata dos Santos Barros, Elaine Cristina Marqueze, Raul Machado Neto, Debora Botequio Moretti. Centro de Genômica Funcional da ESALQ: Luiz Lehmann Coutinho, Ricardo Augusto Brassaloti, Raquel de Lello Rocha Campos Cassano. NGS Soluções Genômicas: Pilar Drummond Sampaio Corrêa Mariani. FZEA-USP Pirassununga: Mirele Daiana Poleti, Jessika Cristina Chagas Lesbon, Elisangela Chicaroni Mattos, Heidge Fukumasu. USP-Botucatu: Rejane Maria Tommasini Grotto, Jayme A. Souza-Neto, Guilherme Targino Valente, Patricia Akemi Assato, Felipe Allan da Silva da Costa, Bianca Cechetto Carlos. Mendelics: Bibiana Santos, João Paulo Kitajima, Erika Freitas, David Schlesinger. Hemocentro Ribeirão Preto: Simone Kashima, Evandra Strazza Rodrigues, Svetoslav Nanev Slavov, Elaine Vieira dos Santos, Rafael dos Santos Bezerra, Luiz Carlos Junior de Alcantara, Marta Giovanetti, Vagner Fonseca, Flavia Aburjaile, Rodrigo Tocantins Calado. |
| EPI_ISL_1795361 | SANTA CASA DE MISERICORDIA DE UBATUBA | Instituto Butantan / ESALQ-Piracicaba | Instituto Butantan: Alexander Roberto Precioso, Dimas Tadeu Covas, Sandra Coccuzzo Sampaio, Maria Carolina Elias, José Salvatore Leister Patané, |

| | | | |
|---|---|---|---|
| | | | Vincent Louis Viala, Antonio Jorge Martins, Ricardo Haddad, Claudia Renata dos Santos Barros, Elaine Cristina Marqueze, Raul Machado Neto, Debora Botequio Moretti. Centro de Genômica Funcional da ESALQ: Luiz Lehmann Coutinho, Ricardo Augusto Brassaloti, Raquel de Lello Rocha Campos Cassano. NGS Soluções Genômicas: Pilar Drummond Sampaio Corrêa Mariani. FZEA-USP Pirassununga: Mirele Daiana Poleti, Jessika Cristina Chagas Lesbon, Elisangela Chicaroni Mattos, Heidge Fukumasu. USP-Botucatu: Rejane Maria Tommasini Grotto, Jayme A. Souza-Neto, Guilherme Targino Valente, Patricia Akemi Assato, Felipe Allan da Silva da Costa, Bianca Cechetto Carlos. Mendelics: Bibiana Santos, João Paulo Kitajima, Erika Freitas, David Schlesinger. Hemocentro Ribeirão Preto: Simone Kashima, Evandra Strazza Rodrigues, Svetoslav Nanev Slavov, Elaine Vieira dos Santos, Rafael dos Santos Bezerra, Luiz Carlos Junior de Alcantara, Marta Giovanetti, Vagner Fonseca, Flavia Aburjaile, Rodrigo Tocantins Calado. |
| EPI_ISL_1795362, EPI_ISL_1795363 | UNIDADE BASICA DE SAUDE DE PIQUETE | Instituto Butantan / ESALQ-Piracicaba | Instituto Butantan: Alexander Roberto Precioso, Dimas Tadeu Covas, Sandra Coccuzzo Sampaio, Maria Carolina Elias, José Salvatore Leister Patané, Vincent Louis Viala, Antonio Jorge Martins, Ricardo Haddad, Claudia Renata dos Santos Barros, Elaine Cristina Marqueze, Raul Machado Neto, Debora Botequio Moretti. Centro de Genômica Funcional da ESALQ: Luiz Lehmann Coutinho, Ricardo Augusto Brassaloti, Raquel de Lello Rocha Campos Cassano. NGS Soluções Genômicas: Pilar Drummond Sampaio Corrêa Mariani. FZEA-USP Pirassununga: Mirele Daiana Poleti, Jessika Cristina Chagas Lesbon, Elisangela Chicaroni Mattos, Heidge Fukumasu. USP-Botucatu: Rejane Maria Tommasini Grotto, Jayme A. Souza-Neto, Guilherme Targino Valente, Patricia Akemi Assato, Felipe Allan da Silva da Costa, Bianca Cechetto Carlos. Mendelics: Bibiana Santos, João Paulo Kitajima, Erika Freitas, David Schlesinger. Hemocentro Ribeirão Preto: Simone Kashima, Evandra Strazza Rodrigues, Svetoslav Nanev Slavov, Elaine Vieira dos Santos, Rafael dos Santos Bezerra, Luiz Carlos Junior de Alcantara, Marta Giovanetti, Vagner Fonseca, Flavia Aburjaile, Rodrigo Tocantins Calado. |
| EPI_ISL_1795364 | SECRETARIA DE SAUDE | Instituto Butantan / ESALQ-Piracicaba | Instituto Butantan: Alexander Roberto Precioso, Dimas Tadeu Covas, Sandra Coccuzzo Sampaio, Maria Carolina Elias, José Salvatore Leister Patané, Vincent Louis Viala, Antonio Jorge Martins, Ricardo Haddad, Claudia Renata dos Santos Barros, Elaine Cristina Marqueze, Raul Machado Neto, Debora Botequio Moretti. Centro de Genômica Funcional da ESALQ: Luiz Lehmann Coutinho, Ricardo Augusto Brassaloti, Raquel de Lello Rocha Campos Cassano. NGS Soluções Genômicas: Pilar Drummond Sampaio Corrêa Mariani. FZEA-USP Pirassununga: Mirele Daiana Poleti, Jessika Cristina Chagas Lesbon, Elisangela Chicaroni Mattos, Heidge Fukumasu. USP-Botucatu: Rejane Maria Tommasini Grotto, Jayme A. Souza-Neto, Guilherme Targino Valente, Patricia Akemi Assato, Felipe Allan da Silva da Costa, Bianca Cechetto Carlos. Mendelics: Bibiana Santos, João Paulo Kitajima, Erika Freitas, David Schlesinger. Hemocentro Ribeirão Preto: Simone Kashima, Evandra Strazza Rodrigues, Svetoslav Nanev Slavov, Elaine Vieira dos Santos, Rafael dos Santos Bezerra, Luiz Carlos Junior de Alcantara, Marta Giovanetti, Vagner Fonseca, Flavia Aburjaile, Rodrigo Tocantins Calado. |
| EPI_ISL_1795365, EPI_ISL_1795366 | LABORATORIO DR PAULO EMILIO DALESSANDRO PINDAMONHANGABA | Instituto Butantan / ESALQ-Piracicaba | Instituto Butantan: Alexander Roberto Precioso, Dimas Tadeu Covas, Sandra Coccuzzo Sampaio, Maria Carolina Elias, José Salvatore Leister Patané, Vincent Louis Viala, Antonio Jorge Martins, Ricardo Haddad, Claudia Renata dos Santos Barros, Elaine Cristina Marqueze, Raul Machado Neto, Debora Botequio Moretti. Centro de Genômica Funcional da ESALQ: Luiz Lehmann Coutinho, Ricardo Augusto Brassaloti, Raquel de Lello Rocha Campos Cassano. NGS Soluções Genômicas: Pilar Drummond Sampaio Corrêa Mariani. FZEA-USP Pirassununga: Mirele Daiana Poleti, Jessika Cristina Chagas Lesbon, Elisangela Chicaroni Mattos, Heidge Fukumasu. USP-Botucatu: Rejane Maria Tommasini Grotto, Jayme A. Souza-Neto, Guilherme Targino Valente, Patricia Akemi Assato, Felipe Allan da Silva da Costa, Bianca Cechetto Carlos. Mendelics: Bibiana Santos, João Paulo Kitajima, Erika Freitas, David Schlesinger. Hemocentro Ribeirão Preto: Simone Kashima, Evandra Strazza Rodrigues, Svetoslav Nanev Slavov, Elaine Vieira dos Santos, Rafael dos Santos Bezerra, Luiz Carlos Junior de Alcantara, Marta Giovanetti, Vagner Fonseca, Flavia Aburjaile, Rodrigo Tocantins Calado. |
| EPI_ISL_1795367 | AMBULATORIO MEDICO MUNICIPAL DE AGUDOS | Instituto Butantan / ESALQ-Piracicaba | Instituto Butantan: Alexander Roberto Precioso, Dimas Tadeu Covas, Sandra Coccuzzo Sampaio, Maria Carolina Elias, José Salvatore Leister Patané, Vincent Louis Viala, Antonio Jorge Martins, Ricardo Haddad, Claudia Renata dos Santos Barros, Elaine Cristina Marqueze, Raul Machado Neto, Debora Botequio Moretti. Centro de Genômica Funcional da ESALQ: Luiz Lehmann Coutinho, Ricardo Augusto Brassaloti, Raquel de Lello Rocha Campos Cassano. NGS Soluções Genômicas: Pilar Drummond Sampaio Corrêa Mariani. FZEA-USP Pirassununga: Mirele Daiana Poleti, Jessika Cristina Chagas Lesbon, Elisangela Chicaroni Mattos, Heidge Fukumasu. USP-Botucatu: Rejane Maria Tommasini Grotto, Jayme A. Souza-Neto, Guilherme Targino Valente, Patricia Akemi Assato, Felipe Allan da Silva da Costa, Bianca Cechetto Carlos. Mendelics: Bibiana Santos, João Paulo Kitajima, Erika Freitas, David Schlesinger. Hemocentro Ribeirão Preto: Simone Kashima, Evandra Strazza Rodrigues, Svetoslav Nanev Slavov, Elaine Vieira dos Santos, Rafael dos Santos Bezerra, Luiz Carlos Junior de Alcantara, Marta Giovanetti, Vagner Fonseca, Flavia Aburjaile, Rodrigo Tocantins Calado. |
| EPI_ISL_1795368 | CENTRO INTEGRADO DE SAUDE | Instituto Butantan / ESALQ-Piracicaba | Instituto Butantan: Alexander Roberto Precioso, Dimas Tadeu Covas, Sandra Coccuzzo Sampaio, Maria Carolina Elias, José Salvatore Leister Patané, Vincent Louis Viala, Antonio Jorge Martins, Ricardo Haddad, Claudia Renata dos Santos Barros, Elaine Cristina Marqueze, Raul Machado Neto, Debora Botequio Moretti. Centro de Genômica Funcional da ESALQ: Luiz Lehmann Coutinho, Ricardo Augusto Brassaloti, Raquel de Lello Rocha Campos Cassano. NGS Soluções Genômicas: Pilar Drummond Sampaio Corrêa Mariani. FZEA-USP Pirassununga: Mirele Daiana Poleti, Jessika Cristina Chagas Lesbon, Elisangela Chicaroni Mattos, Heidge Fukumasu. USP-Botucatu: Rejane Maria Tommasini Grotto, Jayme A. Souza-Neto, Guilherme Targino Valente, Patricia Akemi Assato, Felipe Allan da Silva da Costa, Bianca Cechetto Carlos. Mendelics: Bibiana Santos, João Paulo Kitajima, Erika Freitas, David Schlesinger. Hemocentro Ribeirão Preto: Simone Kashima, Evandra Strazza Rodrigues, Svetoslav Nanev Slavov, Elaine Vieira dos Santos, Rafael dos Santos Bezerra, Luiz Carlos Junior de Alcantara, Marta Giovanetti, Vagner Fonseca, Flavia Aburjaile, Rodrigo Tocantins Calado. |
| EPI_ISL_1795369 | AMBULATORIO MEDICO MUNICIPAL DE AGUDOS | Instituto Butantan / ESALQ-Piracicaba | Instituto Butantan: Alexander Roberto Precioso, Dimas Tadeu Covas, Sandra Coccuzzo Sampaio, Maria Carolina Elias, José Salvatore Leister Patané, Vincent Louis Viala, Antonio Jorge Martins, Ricardo Haddad, Claudia Renata dos Santos Barros, Elaine Cristina Marqueze, Raul Machado Neto, Debora Botequio Moretti. Centro de Genômica Funcional da ESALQ: Luiz Lehmann Coutinho, Ricardo Augusto Brassaloti, Raquel de Lello Rocha Campos Cassano. NGS Soluções Genômicas: Pilar Drummond Sampaio Corrêa Mariani. FZEA-USP Pirassununga: Mirele Daiana Poleti, Jessika Cristina Chagas Lesbon, Elisangela Chicaroni Mattos, Heidge Fukumasu. USP-Botucatu: Rejane Maria Tommasini Grotto, Jayme A. Souza-Neto, Guilherme Targino Valente, Patricia Akemi Assato, Felipe Allan da Silva da Costa, Bianca Cechetto Carlos. Mendelics: Bibiana Santos, João Paulo Kitajima, Erika Freitas, David Schlesinger. Hemocentro Ribeirão Preto: Simone Kashima, Evandra Strazza Rodrigues, Svetoslav Nanev Slavov, Elaine Vieira dos Santos, Rafael dos Santos Bezerra, Luiz Carlos Junior de Alcantara, Marta Giovanetti, Vagner Fonseca, Flavia Aburjaile, Rodrigo Tocantins Calado. |
| EPI_ISL_1795370 | CENTRO INTEGRADO DE SAUDE | Instituto Butantan / ESALQ-Piracicaba | Instituto Butantan: Alexander Roberto Precioso, Dimas Tadeu Covas, Sandra Coccuzzo Sampaio, Maria Carolina Elias, José Salvatore Leister Patané, Vincent Louis Viala, Antonio Jorge Martins, Ricardo Haddad, Claudia Renata dos Santos Barros, Elaine Cristina Marqueze, Raul Machado Neto, Debora Botequio Moretti. Centro de Genômica Funcional da ESALQ: Luiz Lehmann Coutinho, Ricardo Augusto Brassaloti, Raquel de Lello Rocha Campos Cassano. NGS Soluções Genômicas: Pilar Drummond Sampaio Corrêa Mariani. FZEA-USP Pirassununga: Mirele Daiana Poleti, Jessika Cristina Chagas Lesbon, Elisangela Chicaroni Mattos, Heidge Fukumasu. USP-Botucatu: Rejane Maria Tommasini Grotto, Jayme A. Souza-Neto, Guilherme Targino Valente, Patricia Akemi Assato, Felipe Allan da Silva da Costa, Bianca Cechetto Carlos. Mendelics: Bibiana Santos, João Paulo Kitajima, Erika Freitas, David Schlesinger. Hemocentro Ribeirão Preto: Simone Kashima, Evandra Strazza Rodrigues, Svetoslav Nanev Slavov, Elaine Vieira dos Santos, Rafael dos Santos Bezerra, Luiz Carlos Junior de Alcantara, Marta Giovanetti, Vagner Fonseca, Flavia Aburjaile, Rodrigo Tocantins Calado. |
| EPI_ISL_1795371, EPI_ISL_1795372 | HOSPITAL SANTA THEREZINHA BROTAS | Instituto Butantan / ESALQ-Piracicaba | Instituto Butantan: Alexander Roberto Precioso, Dimas Tadeu Covas, Sandra Coccuzzo Sampaio, Maria Carolina Elias, José Salvatore Leister Patané, Vincent Louis Viala, Antonio Jorge Martins, Ricardo Haddad, Claudia Renata dos Santos Barros, Elaine Cristina Marqueze, Raul Machado Neto, Debora Botequio Moretti. Centro de Genômica Funcional da ESALQ: Luiz Lehmann Coutinho, Ricardo Augusto Brassaloti, Raquel de Lello Rocha Campos Cassano. NGS Soluções Genômicas: Pilar Drummond Sampaio Corrêa Mariani. FZEA-USP Pirassununga: Mirele Daiana Poleti, Jessika Cristina Chagas Lesbon, Elisangela Chicaroni Mattos, Heidge Fukumasu. USP-Botucatu: Rejane Maria Tommasini Grotto, Jayme A. Souza-Neto, Guilherme Targino Valente, Patricia Akemi Assato, Felipe Allan da Silva da Costa, Bianca Cechetto Carlos. Mendelics: Bibiana Santos, João Paulo Kitajima, Erika Freitas, David Schlesinger. Hemocentro Ribeirão Preto: Simone Kashima, Evandra Strazza Rodrigues, Svetoslav Nanev Slavov, Elaine Vieira dos Santos, Rafael dos Santos Bezerra, Luiz Carlos Junior de Alcantara, Marta Giovanetti, Vagner Fonseca, Flavia Aburjaile, Rodrigo Tocantins Calado. |
| EPI_ISL_1795373 | SECRETARIA MUNICIPAL DE SAUDE DE PEDERNEIRAS | Instituto Butantan / ESALQ-Piracicaba | Instituto Butantan: Alexander Roberto Precioso, Dimas Tadeu Covas, Sandra Coccuzzo Sampaio, Maria Carolina Elias, José Salvatore Leister Patané, Vincent Louis Viala, Antonio Jorge Martins, Ricardo Haddad, Claudia Renata dos Santos Barros, Elaine Cristina Marqueze, Raul Machado Neto, Debora Botequio Moretti. Centro de Genômica Funcional da ESALQ: Luiz Lehmann Coutinho, Ricardo Augusto Brassaloti, Raquel de Lello Rocha Campos Cassano. NGS Soluções Genômicas: Pilar Drummond Sampaio Corrêa Mariani. FZEA-USP Pirassununga: Mirele Daiana Poleti, Jessika Cristina Chagas Lesbon, Elisangela Chicaroni Mattos, Heidge Fukumasu. USP-Botucatu: Rejane Maria Tommasini Grotto, Jayme A. Souza-Neto, Guilherme Targino Valente, Patricia Akemi Assato, Felipe Allan da Silva da Costa, Bianca Cechetto Carlos. Mendelics: Bibiana Santos, João Paulo Kitajima, Erika Freitas, David Schlesinger. Hemocentro Ribeirão Preto: Simone Kashima, Evandra Strazza Rodrigues, Svetoslav Nanev Slavov, Elaine Vieira dos Santos, Rafael dos Santos Bezerra, Luiz Carlos Junior de Alcantara, Marta Giovanetti, Vagner Fonseca, Flavia Aburjaile, Rodrigo Tocantins Calado. |
| EPI_ISL_1795374, EPI_ISL_1795375 | SANTA CASA DE DOIS CORREGOS | Instituto Butantan / ESALQ-Piracicaba | Instituto Butantan: Alexander Roberto Precioso, Dimas Tadeu Covas, Sandra Coccuzzo Sampaio, Maria Carolina Elias, José Salvatore Leister Patané, Vincent Louis Viala, Antonio Jorge Martins, Ricardo Haddad, Claudia Renata dos Santos Barros, Elaine Cristina Marqueze, Raul Machado Neto, Debora Botequio Moretti. Centro de Genômica Funcional da ESALQ: Luiz Lehmann Coutinho, Ricardo Augusto Brassaloti, Raquel de Lello Rocha Campos |

| | | | |
|---|---|---|---|
| | | | Cassano. NGS Soluções Genômicas: Pilar Drummond Sampaio Corrêa Mariani. FZEA-USP Pirassununga: Mirele Daiana Poleti, Jessika Cristina Chagas Lesbon, Elisangela Chicaroni Mattos, Heidge Fukumasu. USP-Botucatu: Rejane Maria Tommasini Grotto, Jayme A. Souza-Neto, Guilherme Targino Valente, Patricia Akemi Assato, Felipe Allan da Silva da Costa, Bianca Cechetto Carlos. Mendelics: Bibiana Santos, João Paulo Kitajima, Erika Freitas, David Schlesinger. Hemocentro Ribeirão Preto: Simone Kashima, Evandra Strazza Rodrigues, Svetoslav Nanev Slavov, Elaine Vieira dos Santos, Rafael dos Santos Bezerra, Luiz Carlos Junior de Alcantara, Marta Giovanetti, Vagner Fonseca, Flavia Aburjaile, Rodrigo Tocantins Calado. |
| EPI_ISL_1795376, EPI_ISL_1795377, EPI_ISL_1795378 | ESF MINEIROS DO TIETE | Instituto Butantan / ESALQ-Piracicaba | Instituto Butantan: Alexander Roberto Precioso, Dimas Tadeu Covas, Sandra Coccuzzo Sampaio, Maria Carolina Elias, José Salvatore Leister Patané, Vincent Louis Viala, Antonio Jorge Martins, Ricardo Haddad, Claudia Renata dos Santos Barros, Elaine Cristina Marqueze, Raul Machado Neto, Debora Botequio Moretti. Centro de Genômica Funcional da ESALQ: Luiz Lehmann Coutinho, Ricardo Augusto Brassaloti, Raquel de Lello Rocha Campos Cassano. NGS Soluções Genômicas: Pilar Drummond Sampaio Corrêa Mariani. FZEA-USP Pirassununga: Mirele Daiana Poleti, Jessika Cristina Chagas Lesbon, Elisangela Chicaroni Mattos, Heidge Fukumasu. USP-Botucatu: Rejane Maria Tommasini Grotto, Jayme A. Souza-Neto, Guilherme Targino Valente, Patricia Akemi Assato, Felipe Allan da Silva da Costa, Bianca Cechetto Carlos. Mendelics: Bibiana Santos, João Paulo Kitajima, Erika Freitas, David Schlesinger. Hemocentro Ribeirão Preto: Simone Kashima, Evandra Strazza Rodrigues, Svetoslav Nanev Slavov, Elaine Vieira dos Santos, Rafael dos Santos Bezerra, Luiz Carlos Junior de Alcantara, Marta Giovanetti, Vagner Fonseca, Flavia Aburjaile, Rodrigo Tocantins Calado. |
| EPI_ISL_1795379 | NUCLEO DE SAUDE VILA FALCAO DE BAURU | Instituto Butantan / ESALQ-Piracicaba | Instituto Butantan: Alexander Roberto Precioso, Dimas Tadeu Covas, Sandra Coccuzzo Sampaio, Maria Carolina Elias, José Salvatore Leister Patané, Vincent Louis Viala, Antonio Jorge Martins, Ricardo Haddad, Claudia Renata dos Santos Barros, Elaine Cristina Marqueze, Raul Machado Neto, Debora Botequio Moretti. Centro de Genômica Funcional da ESALQ: Luiz Lehmann Coutinho, Ricardo Augusto Brassaloti, Raquel de Lello Rocha Campos Cassano. NGS Soluções Genômicas: Pilar Drummond Sampaio Corrêa Mariani. FZEA-USP Pirassununga: Mirele Daiana Poleti, Jessika Cristina Chagas Lesbon, Elisangela Chicaroni Mattos, Heidge Fukumasu. USP-Botucatu: Rejane Maria Tommasini Grotto, Jayme A. Souza-Neto, Guilherme Targino Valente, Patricia Akemi Assato, Felipe Allan da Silva da Costa, Bianca Cechetto Carlos. Mendelics: Bibiana Santos, João Paulo Kitajima, Erika Freitas, David Schlesinger. Hemocentro Ribeirão Preto: Simone Kashima, Evandra Strazza Rodrigues, Svetoslav Nanev Slavov, Elaine Vieira dos Santos, Rafael dos Santos Bezerra, Luiz Carlos Junior de Alcantara, Marta Giovanetti, Vagner Fonseca, Flavia Aburjaile, Rodrigo Tocantins Calado. |
| EPI_ISL_1795380 | HOSPITAL MATERNIDADE SAO JOSE ITAPUI | Instituto Butantan / ESALQ-Piracicaba | Instituto Butantan: Alexander Roberto Precioso, Dimas Tadeu Covas, Sandra Coccuzzo Sampaio, Maria Carolina Elias, José Salvatore Leister Patané, Vincent Louis Viala, Antonio Jorge Martins, Ricardo Haddad, Claudia Renata dos Santos Barros, Elaine Cristina Marqueze, Raul Machado Neto, Debora Botequio Moretti. Centro de Genômica Funcional da ESALQ: Luiz Lehmann Coutinho, Ricardo Augusto Brassaloti, Raquel de Lello Rocha Campos Cassano. NGS Soluções Genômicas: Pilar Drummond Sampaio Corrêa Mariani. FZEA-USP Pirassununga: Mirele Daiana Poleti, Jessika Cristina Chagas Lesbon, Elisangela Chicaroni Mattos, Heidge Fukumasu. USP-Botucatu: Rejane Maria Tommasini Grotto, Jayme A. Souza-Neto, Guilherme Targino Valente, Patricia Akemi Assato, Felipe Allan da Silva da Costa, Bianca Cechetto Carlos. Mendelics: Bibiana Santos, João Paulo Kitajima, Erika Freitas, David Schlesinger. Hemocentro Ribeirão Preto: Simone Kashima, Evandra Strazza Rodrigues, Svetoslav Nanev Slavov, Elaine Vieira dos Santos, Rafael dos Santos Bezerra, Luiz Carlos Junior de Alcantara, Marta Giovanetti, Vagner Fonseca, Flavia Aburjaile, Rodrigo Tocantins Calado. |
| EPI_ISL_1795381 | SECRETARIA MUNICIPAL DE SAUDE DE MACATUBA | Instituto Butantan / ESALQ-Piracicaba | Instituto Butantan: Alexander Roberto Precioso, Dimas Tadeu Covas, Sandra Coccuzzo Sampaio, Maria Carolina Elias, José Salvatore Leister Patané, Vincent Louis Viala, Antonio Jorge Martins, Ricardo Haddad, Claudia Renata dos Santos Barros, Elaine Cristina Marqueze, Raul Machado Neto, Debora Botequio Moretti. Centro de Genômica Funcional da ESALQ: Luiz Lehmann Coutinho, Ricardo Augusto Brassaloti, Raquel de Lello Rocha Campos Cassano. NGS Soluções Genômicas: Pilar Drummond Sampaio Corrêa Mariani. FZEA-USP Pirassununga: Mirele Daiana Poleti, Jessika Cristina Chagas Lesbon, Elisangela Chicaroni Mattos, Heidge Fukumasu. USP-Botucatu: Rejane Maria Tommasini Grotto, Jayme A. Souza-Neto, Guilherme Targino Valente, Patricia Akemi Assato, Felipe Allan da Silva da Costa, Bianca Cechetto Carlos. Mendelics: Bibiana Santos, João Paulo Kitajima, Erika Freitas, David Schlesinger. Hemocentro Ribeirão Preto: Simone Kashima, Evandra Strazza Rodrigues, Svetoslav Nanev Slavov, Elaine Vieira dos Santos, Rafael dos Santos Bezerra, Luiz Carlos Junior de Alcantara, Marta Giovanetti, Vagner Fonseca, Flavia Aburjaile, Rodrigo Tocantins Calado. |
| EPI_ISL_1795382, EPI_ISL_1795383 | LABORATORIO DR PAULO EMILIO DALESSANDRO PINDAMONHANGABA | Instituto Butantan / ESALQ-Piracicaba | Instituto Butantan: Alexander Roberto Precioso, Dimas Tadeu Covas, Sandra Coccuzzo Sampaio, Maria Carolina Elias, José Salvatore Leister Patané, Vincent Louis Viala, Antonio Jorge Martins, Ricardo Haddad, Claudia Renata dos Santos Barros, Elaine Cristina Marqueze, Raul Machado Neto, Debora Botequio Moretti. Centro de Genômica Funcional da ESALQ: Luiz Lehmann Coutinho, Ricardo Augusto Brassaloti, Raquel de Lello Rocha Campos Cassano. NGS Soluções Genômicas: Pilar Drummond Sampaio Corrêa Mariani. FZEA-USP Pirassununga: Mirele Daiana Poleti, Jessika Cristina Chagas Lesbon, Elisangela Chicaroni Mattos, Heidge Fukumasu. USP-Botucatu: Rejane Maria Tommasini Grotto, Jayme A. Souza-Neto, Guilherme Targino Valente, Patricia Akemi Assato, Felipe Allan da Silva da Costa, Bianca Cechetto Carlos. Mendelics: Bibiana Santos, João Paulo Kitajima, Erika Freitas, David Schlesinger. Hemocentro Ribeirão Preto: Simone Kashima, Evandra Strazza Rodrigues, Svetoslav Nanev Slavov, Elaine Vieira dos Santos, Rafael dos Santos Bezerra, Luiz Carlos Junior de Alcantara, Marta Giovanetti, Vagner Fonseca, Flavia Aburjaile, Rodrigo Tocantins Calado. |
| EPI_ISL_1795384 | USF SALERNO | Instituto Butantan / ESALQ-Piracicaba | Instituto Butantan: Alexander Roberto Precioso, Dimas Tadeu Covas, Sandra Coccuzzo Sampaio, Maria Carolina Elias, José Salvatore Leister Patané, Vincent Louis Viala, Antonio Jorge Martins, Ricardo Haddad, Claudia Renata dos Santos Barros, Elaine Cristina Marqueze, Raul Machado Neto, Debora Botequio Moretti. Centro de Genômica Funcional da ESALQ: Luiz Lehmann Coutinho, Ricardo Augusto Brassaloti, Raquel de Lello Rocha Campos Cassano. NGS Soluções Genômicas: Pilar Drummond Sampaio Corrêa Mariani. FZEA-USP Pirassununga: Mirele Daiana Poleti, Jessika Cristina Chagas Lesbon, Elisangela Chicaroni Mattos, Heidge Fukumasu. USP-Botucatu: Rejane Maria Tommasini Grotto, Jayme A. Souza-Neto, Guilherme Targino Valente, Patricia Akemi Assato, Felipe Allan da Silva da Costa, Bianca Cechetto Carlos. Mendelics: Bibiana Santos, João Paulo Kitajima, Erika Freitas, David Schlesinger. Hemocentro Ribeirão Preto: Simone Kashima, Evandra Strazza Rodrigues, Svetoslav Nanev Slavov, Elaine Vieira dos Santos, Rafael dos Santos Bezerra, Luiz Carlos Junior de Alcantara, Marta Giovanetti, Vagner Fonseca, Flavia Aburjaile, Rodrigo Tocantins Calado. |
| EPI_ISL_1795385, EPI_ISL_1795386 | UNIDADE DE VIGILANCIA EM SAUDE | Instituto Butantan / ESALQ-Piracicaba | Instituto Butantan: Alexander Roberto Precioso, Dimas Tadeu Covas, Sandra Coccuzzo Sampaio, Maria Carolina Elias, José Salvatore Leister Patané, Vincent Louis Viala, Antonio Jorge Martins, Ricardo Haddad, Claudia Renata dos Santos Barros, Elaine Cristina Marqueze, Raul Machado Neto, Debora Botequio Moretti. Centro de Genômica Funcional da ESALQ: Luiz Lehmann Coutinho, Ricardo Augusto Brassaloti, Raquel de Lello Rocha Campos Cassano. NGS Soluções Genômicas: Pilar Drummond Sampaio Corrêa Mariani. FZEA-USP Pirassununga: Mirele Daiana Poleti, Jessika Cristina Chagas Lesbon, Elisangela Chicaroni Mattos, Heidge Fukumasu. USP-Botucatu: Rejane Maria Tommasini Grotto, Jayme A. Souza-Neto, Guilherme Targino Valente, Patricia Akemi Assato, Felipe Allan da Silva da Costa, Bianca Cechetto Carlos. Mendelics: Bibiana Santos, João Paulo Kitajima, Erika Freitas, David Schlesinger. Hemocentro Ribeirão Preto: Simone Kashima, Evandra Strazza Rodrigues, Svetoslav Nanev Slavov, Elaine Vieira dos Santos, Rafael dos Santos Bezerra, Luiz Carlos Junior de Alcantara, Marta Giovanetti, Vagner Fonseca, Flavia Aburjaile, Rodrigo Tocantins Calado. |
| EPI_ISL_1795387, EPI_ISL_1795388 | POLICLINICA HORTOLANDIA | Instituto Butantan / ESALQ-Piracicaba | Instituto Butantan: Alexander Roberto Precioso, Dimas Tadeu Covas, Sandra Coccuzzo Sampaio, Maria Carolina Elias, José Salvatore Leister Patané, Vincent Louis Viala, Antonio Jorge Martins, Ricardo Haddad, Claudia Renata dos Santos Barros, Elaine Cristina Marqueze, Raul Machado Neto, Debora Botequio Moretti. Centro de Genômica Funcional da ESALQ: Luiz Lehmann Coutinho, Ricardo Augusto Brassaloti, Raquel de Lello Rocha Campos Cassano. NGS Soluções Genômicas: Pilar Drummond Sampaio Corrêa Mariani. FZEA-USP Pirassununga: Mirele Daiana Poleti, Jessika Cristina Chagas Lesbon, Elisangela Chicaroni Mattos, Heidge Fukumasu. USP-Botucatu: Rejane Maria Tommasini Grotto, Jayme A. Souza-Neto, Guilherme Targino Valente, Patricia Akemi Assato, Felipe Allan da Silva da Costa, Bianca Cechetto Carlos. Mendelics: Bibiana Santos, João Paulo Kitajima, Erika Freitas, David Schlesinger. Hemocentro Ribeirão Preto: Simone Kashima, Evandra Strazza Rodrigues, Svetoslav Nanev Slavov, Elaine Vieira dos Santos, Rafael dos Santos Bezerra, Luiz Carlos Junior de Alcantara, Marta Giovanetti, Vagner Fonseca, Flavia Aburjaile, Rodrigo Tocantins Calado. |
| EPI_ISL_1795389 | CS DE BALSAMO | Instituto Butantan / ESALQ-Piracicaba | Instituto Butantan: Alexander Roberto Precioso, Dimas Tadeu Covas, Sandra Coccuzzo Sampaio, Maria Carolina Elias, José Salvatore Leister Patané, Vincent Louis Viala, Antonio Jorge Martins, Ricardo Haddad, Claudia Renata dos Santos Barros, Elaine Cristina Marqueze, Raul Machado Neto, Debora Botequio Moretti. Centro de Genômica Funcional da ESALQ: Luiz Lehmann Coutinho, Ricardo Augusto Brassaloti, Raquel de Lello Rocha Campos Cassano. NGS Soluções Genômicas: Pilar Drummond Sampaio Corrêa Mariani. FZEA-USP Pirassununga: Mirele Daiana Poleti, Jessika Cristina Chagas Lesbon, Elisangela Chicaroni Mattos, Heidge Fukumasu. USP-Botucatu: Rejane Maria Tommasini Grotto, Jayme A. Souza-Neto, Guilherme Targino Valente, Patricia Akemi Assato, Felipe Allan da Silva da Costa, Bianca Cechetto Carlos. Mendelics: Bibiana Santos, João Paulo Kitajima, Erika Freitas, David Schlesinger. Hemocentro Ribeirão Preto: Simone Kashima, Evandra Strazza Rodrigues, Svetoslav Nanev Slavov, Elaine Vieira dos Santos, Rafael dos Santos Bezerra, Luiz Carlos Junior de Alcantara, Marta Giovanetti, Vagner Fonseca, Flavia Aburjaile, Rodrigo Tocantins Calado. |
| EPI_ISL_1795390 | CS DE OSCAR BRESSANE PSF | Instituto Butantan / ESALQ-Piracicaba | Instituto Butantan: Alexander Roberto Precioso, Dimas Tadeu Covas, Sandra Coccuzzo Sampaio, Maria Carolina Elias, José Salvatore Leister Patané, Vincent Louis Viala, Antonio Jorge Martins, Ricardo Haddad, Claudia Renata dos Santos Barros, Elaine Cristina Marqueze, Raul Machado Neto, Debora Botequio Moretti. Centro de Genômica Funcional da ESALQ: Luiz Lehmann Coutinho, Ricardo Augusto Brassaloti, Raquel de Lello Rocha Campos Cassano. NGS Soluções Genômicas: Pilar Drummond Sampaio Corrêa Mariani. FZEA-USP Pirassununga: Mirele Daiana Poleti, Jessika Cristina Chagas Lesbon, Elisangela Chicaroni Mattos, Heidge Fukumasu. USP-Botucatu: Rejane Maria Tommasini Grotto, Jayme A. Souza-Neto, Guilherme Targino |

| | | | |
|---|---|---|---|
| | | | Valente, Patricia Akemi Assato, Felipe Allan da Silva da Costa, Bianca Cechetto Carlos. Mendelics: Bibiana Santos, João Paulo Kitajima, Erika Freitas, David Schlesinger. Hemocentro Ribeirão Preto: Simone Kashima, Evandra Strazza Rodrigues, Svetoslav Nanev Slavov, Elaine Vieira dos Santos, Rafael dos Santos Bezerra, Luiz Carlos Junior de Alcantara, Marta Giovanetti, Vagner Fonseca, Flavia Aburjaile, Rodrigo Tocantins Calado. |
| EPI_ISL_1795391 | USF ROSA CRUZ | Instituto Butantan / ESALQ-Piracicaba | Instituto Butantan: Alexander Roberto Precioso, Dimas Tadeu Covas, Sandra Coccuzzo Sampaio, Maria Carolina Elias, José Salvatore Leister Patané, Vincent Louis Viala, Antonio Jorge Martins, Ricardo Haddad, Claudia Renata dos Santos Barros, Elaine Cristina Marqueze, Raul Machado Neto, Debora Botequio Moretti. Centro de Genômica Funcional da ESALQ: Luiz Lehmann Coutinho, Ricardo Augusto Brassaloti, Raquel de Lello Rocha Campos Cassano. NGS Soluções Genômicas: Pilar Drummond Sampaio Corrêa Mariani. FZEA-USP Pirassununga: Mirele Daiana Poleti, Jessika Cristina Chagas Lesbon, Elisangela Chicaroni Mattos, Heidge Fukumasu. USP-Botucatu: Rejane Maria Tommasini Grotto, Jayme A. Souza-Neto, Guilherme Targino Valente, Patricia Akemi Assato, Felipe Allan da Silva da Costa, Bianca Cechetto Carlos. Mendelics: Bibiana Santos, João Paulo Kitajima, Erika Freitas, David Schlesinger. Hemocentro Ribeirão Preto: Simone Kashima, Evandra Strazza Rodrigues, Svetoslav Nanev Slavov, Elaine Vieira dos Santos, Rafael dos Santos Bezerra, Luiz Carlos Junior de Alcantara, Marta Giovanetti, Vagner Fonseca, Flavia Aburjaile, Rodrigo Tocantins Calado. |
| EPI_ISL_1821204 | Santa Casa de Cravinhos | Instituto Adolfo Lutz, Interdiciplinary Procedures Center, Strategic Laboratory | Claudio Tavares Sacchi, Claudia Regina Gonçalves, Erica Valessa Ramos Gomes, Karoline Rodrigues Campos, Caio Vinicius Dias Lopes, Leonardo Jose Tadeu de Araujo |
| EPI_ISL_1821207 | Secretaria Municipal de Saúde de Lins | Instituto Adolfo Lutz, Interdiciplinary Procedures Center, Strategic Laboratory | Claudio Tavares Sacchi, Claudia Regina Gonçalves, Erica Valessa Ramos Gomes, Karoline Rodrigues Campos, Caio Vinicius Dias Lopes, Leonardo Jose Tadeu de Araujo |
| EPI_ISL_1821208 | Hospital de Base de Bauru Hospital de Base Setima Regiao | Instituto Adolfo Lutz, Interdiciplinary Procedures Center, Strategic Laboratory | Claudio Tavares Sacchi, Claudia Regina Gonçalves, Erica Valessa Ramos Gomes, Karoline Rodrigues Campos, Caio Vinicius Dias Lopes, Leonardo Jose Tadeu de Araujo |
| EPI_ISL_1821209 | CS de Paulo de Faria | Instituto Adolfo Lutz, Interdiciplinary Procedures Center, Strategic Laboratory | Claudio Tavares Sacchi, Claudia Regina Gonçalves, Erica Valessa Ramos Gomes, Karoline Rodrigues Campos, Caio Vinicius Dias Lopes, Leonardo Jose Tadeu de Araujo |
| EPI_ISL_1821210 | Hospital Universitario de Marilia | Instituto Adolfo Lutz, Interdiciplinary Procedures Center, Strategic Laboratory | Claudio Tavares Sacchi, Claudia Regina Gonçalves, Erica Valessa Ramos Gomes, Karoline Rodrigues Campos, Caio Vinicius Dias Lopes, Leonardo Jose Tadeu de Araujo |
| EPI_ISL_1821211, EPI_ISL_1821212, EPI_ISL_1821213 | Diagnóstico da America S/A | Instituto Adolfo Lutz, Interdiciplinary Procedures Center, Strategic Laboratory | Claudio Tavares Sacchi, Claudia Regina Gonçalves, Erica Valessa Ramos Gomes, Karoline Rodrigues Campos, Caio Vinicius Dias Lopes, Leonardo Jose Tadeu de Araujo |
| EPI_ISL_1821214, EPI_ISL_1821215, EPI_ISL_1821216, EPI_ISL_1821217, EPI_ISL_1821218, EPI_ISL_1821219, EPI_ISL_1821220, EPI_ISL_1821221, EPI_ISL_1821222, EPI_ISL_1821223, EPI_ISL_1821224 | | | |
| see above | Instituto Adolfo Lutz - Regional de Aracatuba | Instituto Adolfo Lutz, Interdiciplinary Procedures Center, Strategic Laboratory | Claudio Tavares Sacchi, Claudia Regina Gonçalves, Erica Valessa Ramos Gomes, Karoline Rodrigues Campos, Caio Vinicius Dias Lopes, Leonardo Jose Tadeu de Araujo |
| EPI_ISL_1821234, EPI_ISL_1821235, EPI_ISL_1821236, EPI_ISL_1821237, EPI_ISL_1821239, EPI_ISL_1821240, EPI_ISL_1821241, EPI_ISL_1821242, EPI_ISL_1821244, EPI_ISL_1821245 | Instituto Adolfo Lutz - Regional de Marilia | Instituto Adolfo Lutz, Interdiciplinary Procedures Center, Strategic Laboratory | Claudio Tavares Sacchi, Claudia Regina Gonçalves, Erica Valessa Ramos Gomes, Karoline Rodrigues Campos, Caio Vinicius Dias Lopes, Leonardo Jose Tadeu de Araujo |
| EPI_ISL_1821246, EPI_ISL_1821247, EPI_ISL_1821248, EPI_ISL_1821249, EPI_ISL_1821250, EPI_ISL_1821251, EPI_ISL_1821252, EPI_ISL_1821253, EPI_ISL_1821254, EPI_ISL_1821255, EPI_ISL_1821256, EPI_ISL_1821257, EPI_ISL_1821258, EPI_ISL_1821259, EPI_ISL_1821260, EPI_ISL_1821261, EPI_ISL_1821262, EPI_ISL_1821263, EPI_ISL_1821264, EPI_ISL_1821265 | | | |
| see above | Instituto Adolfo Lutz - Regional de Ribeirao Preto | Instituto Adolfo Lutz, Interdiciplinary Procedures Center, Strategic Laboratory | Claudio Tavares Sacchi, Claudia Regina Gonçalves, Erica Valessa Ramos Gomes, Karoline Rodrigues Campos, Caio Vinicius Dias Lopes, Leonardo Jose Tadeu de Araujo |
| EPI_ISL_1821266, EPI_ISL_1821267, EPI_ISL_1821268, EPI_ISL_1821269, EPI_ISL_1821270, EPI_ISL_1821271, EPI_ISL_1821272, EPI_ISL_1821273 | Instituto Adolfo Lutz Central | Instituto Adolfo Lutz, Interdiciplinary Procedures Center, Strategic Laboratory | Claudio Tavares Sacchi, Claudia Regina Gonçalves, Erica Valessa Ramos Gomes, Karoline Rodrigues Campos, Caio Vinicius Dias Lopes, Leonardo Jose Tadeu de Araujo |
| EPI_ISL_1858267, EPI_ISL_1858269, EPI_ISL_1858271, EPI_ISL_1858273, EPI_ISL_1858275, EPI_ISL_1858277, EPI_ISL_1858279, EPI_ISL_1858282, EPI_ISL_1858284, EPI_ISL_1858286, EPI_ISL_1858288, EPI_ISL_1858290, EPI_ISL_1858292, EPI_ISL_1858294, EPI_ISL_1858295, EPI_ISL_1858297, EPI_ISL_1858299, EPI_ISL_1858301, EPI_ISL_1858304, EPI_ISL_1858306, EPI_ISL_1858308, EPI_ISL_1858310, EPI_ISL_1858312, EPI_ISL_1858314, EPI_ISL_1858315, EPI_ISL_1858317, EPI_ISL_1858319, EPI_ISL_1858321, EPI_ISL_1858323, EPI_ISL_1858325, EPI_ISL_1858327, EPI_ISL_1858329, EPI_ISL_1858331, EPI_ISL_1858333, EPI_ISL_1858334, EPI_ISL_1858336, EPI_ISL_1858338, EPI_ISL_1858340, EPI_ISL_1858342, EPI_ISL_1858344, EPI_ISL_1858346, EPI_ISL_1858348, EPI_ISL_1858350, EPI_ISL_1858352, EPI_ISL_1858354, EPI_ISL_1858356, EPI_ISL_1858359, EPI_ISL_1858361, EPI_ISL_1858363, EPI_ISL_1858365, EPI_ISL_1858367, EPI_ISL_1858369, EPI_ISL_1858371, EPI_ISL_1858373, EPI_ISL_1858375, EPI_ISL_1858377, EPI_ISL_1858379, EPI_ISL_1858380, EPI_ISL_1858383, EPI_ISL_1858384, EPI_ISL_1858387, EPI_ISL_1858389, EPI_ISL_1858391, EPI_ISL_1858392, EPI_ISL_1858395, EPI_ISL_1858397, EPI_ISL_1858399, EPI_ISL_1858401, EPI_ISL_1858403, EPI_ISL_1858405, EPI_ISL_1858409, EPI_ISL_1858411, EPI_ISL_1858414, EPI_ISL_1858416, EPI_ISL_1858418, EPI_ISL_1858420, EPI_ISL_1858422, EPI_ISL_1858423, EPI_ISL_1858426, EPI_ISL_1858428, EPI_ISL_1858430, EPI_ISL_1858432, EPI_ISL_1858434, EPI_ISL_1858436, EPI_ISL_1858438, EPI_ISL_1858440, EPI_ISL_1858442, EPI_ISL_1858444, EPI_ISL_1858446, EPI_ISL_1858449, EPI_ISL_1858451, EPI_ISL_1858453, EPI_ISL_1858454, EPI_ISL_1858456, EPI_ISL_1858458, EPI_ISL_1858460, EPI_ISL_1858462, EPI_ISL_1858464, EPI_ISL_1858466, EPI_ISL_1858468, EPI_ISL_1858470, EPI_ISL_1858473, EPI_ISL_1858475, EPI_ISL_1858477, EPI_ISL_1858479, EPI_ISL_1858481, EPI_ISL_1858483, EPI_ISL_1858486, EPI_ISL_1858488, EPI_ISL_1858490, EPI_ISL_1858492, EPI_ISL_1858494, EPI_ISL_1858496, EPI_ISL_1858498, EPI_ISL_1858500, EPI_ISL_1858504, EPI_ISL_1858506, EPI_ISL_1858508, EPI_ISL_1858510, EPI_ISL_1858512, EPI_ISL_1858514, EPI_ISL_1858516, EPI_ISL_1858518, EPI_ISL_1858520, EPI_ISL_1858522, EPI_ISL_1858524, EPI_ISL_1858526, EPI_ISL_1858528, EPI_ISL_1858530, EPI_ISL_1858532, EPI_ISL_1858534, EPI_ISL_1858536, EPI_ISL_1858538, EPI_ISL_1858540, EPI_ISL_1858542, EPI_ISL_1858546, EPI_ISL_1858548, EPI_ISL_1858549, EPI_ISL_1858551, EPI_ISL_1858554, EPI_ISL_1858555, EPI_ISL_1858557, EPI_ISL_1858559, EPI_ISL_1858561, EPI_ISL_1858563, EPI_ISL_1858565, EPI_ISL_1858567, EPI_ISL_1858569, EPI_ISL_1858571, EPI_ISL_1858572, EPI_ISL_1858575, EPI_ISL_1858577, EPI_ISL_1858578, EPI_ISL_1858580, EPI_ISL_1858582, EPI_ISL_1858584, EPI_ISL_1858586, EPI_ISL_1858590, EPI_ISL_1858592, EPI_ISL_1858594, EPI_ISL_1858596, EPI_ISL_1858598, EPI_ISL_1858599, EPI_ISL_1858602, EPI_ISL_1858603, EPI_ISL_1858606, EPI_ISL_1858607, EPI_ISL_1858610, EPI_ISL_1858611, EPI_ISL_1858613, EPI_ISL_1858615, EPI_ISL_1858617, EPI_ISL_1858619, EPI_ISL_1858621, EPI_ISL_1858623, EPI_ISL_1858625, EPI_ISL_1858627, EPI_ISL_1858629, EPI_ISL_1858631, EPI_ISL_1858633, EPI_ISL_1858636, EPI_ISL_1858637, EPI_ISL_1858639, EPI_ISL_1858641, EPI_ISL_1858643, EPI_ISL_1858645, EPI_ISL_1858648, EPI_ISL_1858650, EPI_ISL_1858651, EPI_ISL_1858653, EPI_ISL_1858655, EPI_ISL_1858656, EPI_ISL_1858658, EPI_ISL_1858660, EPI_ISL_1858662, EPI_ISL_1858664, EPI_ISL_1858666, EPI_ISL_1858668, EPI_ISL_1858670, EPI_ISL_1858672, EPI_ISL_1858674, EPI_ISL_1858675, EPI_ISL_1858677, EPI_ISL_1858680, EPI_ISL_1858682, EPI_ISL_1858684, EPI_ISL_1858686, EPI_ISL_1858688, EPI_ISL_1858691, EPI_ISL_1858693, EPI_ISL_1858694, EPI_ISL_1858697, EPI_ISL_1858699, EPI_ISL_1858701, EPI_ISL_1858703, EPI_ISL_1858705, EPI_ISL_1858707, EPI_ISL_1858709, EPI_ISL_1858711, EPI_ISL_1858713, EPI_ISL_1858715, EPI_ISL_1858717, EPI_ISL_1858719, EPI_ISL_1858721, EPI_ISL_1858722, EPI_ISL_1858724, EPI_ISL_1858728, EPI_ISL_1858730, EPI_ISL_1858732, EPI_ISL_1858734, EPI_ISL_1858736, EPI_ISL_1858738, EPI_ISL_1858740, EPI_ISL_1858741, EPI_ISL_1858743, EPI_ISL_1858745, EPI_ISL_1858747, EPI_ISL_1858749, EPI_ISL_1858751, EPI_ISL_1858752, EPI_ISL_1858754, EPI_ISL_1858756 | | | |
| see above | Laboratorio Central Noel Nutels | Bioinformatics Laboratory / LNCC | Luiz G P de Almeida, Alessandra P Lamarca, Ronaldo da Silva F Jr, Liliane Cavalcante, Alexandra L Gerber, Ana Paula de C Guimarães, Douglas Terra Machado, Cassia Alves, Diana Mariani, Thais Felix Cruz, Mario Sergio Ribeiro, Silvia Carvalho, Flávio Dias da Silva, Marcio Henrique de Oliveira Garcia, Leandro Magalhães de Souza, Cristiane Gomes da Silva, Caio Luiz Pereira Ribeiro, Andréa Cony Cavalcanti, Claudia Maria Braga de Mello, Amilcar Tanuri, Ana Tereza R Vasconcelos |
| EPI_ISL_1858758, EPI_ISL_1858761, EPI_ISL_1858763, EPI_ISL_1858765, EPI_ISL_1858767, EPI_ISL_1858769, EPI_ISL_1858771, EPI_ISL_1858775, EPI_ISL_1858777, EPI_ISL_1858779, EPI_ISL_1858781, EPI_ISL_1858783, EPI_ISL_1858786, EPI_ISL_1858788, EPI_ISL_1858790, EPI_ISL_1858792, EPI_ISL_1858794, EPI_ISL_1858796, EPI_ISL_1858798, EPI_ISL_1858800, EPI_ISL_1858802, EPI_ISL_1858803, EPI_ISL_1858805, EPI_ISL_1858807, EPI_ISL_1858809, EPI_ISL_1858811, EPI_ISL_1858814, EPI_ISL_1858816, EPI_ISL_1858818, EPI_ISL_1858819, EPI_ISL_1858822, EPI_ISL_1858824, EPI_ISL_1858826, EPI_ISL_1858828, EPI_ISL_1858830, EPI_ISL_1858832, EPI_ISL_1858834, EPI_ISL_1858836, EPI_ISL_1858838, EPI_ISL_1858840, EPI_ISL_1858842, EPI_ISL_1858843, EPI_ISL_1858846, EPI_ISL_1858847, EPI_ISL_1858850, EPI_ISL_1858852, EPI_ISL_1858853, EPI_ISL_1858856, EPI_ISL_1858858, EPI_ISL_1858860, EPI_ISL_1858862, EPI_ISL_1858864, EPI_ISL_1858866, EPI_ISL_1858868, EPI_ISL_1858870, EPI_ISL_1858872, EPI_ISL_1858874, EPI_ISL_1858876, EPI_ISL_1858878, EPI_ISL_1858880, EPI_ISL_1858882, EPI_ISL_1858884, EPI_ISL_1858886, EPI_ISL_1858888, EPI_ISL_1858890, EPI_ISL_1858892, EPI_ISL_1858894, EPI_ISL_1858896, EPI_ISL_1858898, EPI_ISL_1858900, EPI_ISL_1858902, EPI_ISL_1858904, EPI_ISL_1858906, EPI_ISL_1858908, EPI_ISL_1858910, EPI_ISL_1858912, EPI_ISL_1858914, EPI_ISL_1858916, EPI_ISL_1858918, EPI_ISL_1858920, EPI_ISL_1858922, EPI_ISL_1858924, EPI_ISL_1858926, EPI_ISL_1858928, EPI_ISL_1858930, EPI_ISL_1858933, EPI_ISL_1858934, EPI_ISL_1858936, EPI_ISL_1858938, EPI_ISL_1858940, EPI_ISL_1858942, EPI_ISL_1858944, EPI_ISL_1858946, EPI_ISL_1858948, EPI_ISL_1858950, EPI_ISL_1858952, EPI_ISL_1858954, EPI_ISL_1858957, EPI_ISL_1858958, EPI_ISL_1858960, EPI_ISL_1858963, EPI_ISL_1858964, EPI_ISL_1858966, EPI_ISL_1858968, EPI_ISL_1858970, EPI_ISL_1858973, EPI_ISL_1858975, EPI_ISL_1858977, EPI_ISL_1858979, EPI_ISL_1858981, EPI_ISL_1858984, EPI_ISL_1858986, EPI_ISL_1858988, EPI_ISL_1858990, EPI_ISL_1858992, EPI_ISL_1858994, EPI_ISL_1858996, EPI_ISL_1858998, EPI_ISL_1859000, EPI_ISL_1859002, EPI_ISL_1859005, EPI_ISL_1859007, EPI_ISL_1859008 | | | |
| see above | Unidade de apoio ao diagnóstico da COVID - UNADIG | Bioinformatics Laboratory / LNCC | Luiz G P de Almeida, Alessandra P Lamarca, Ronaldo da Silva F Jr, Liliane Cavalcante, Alexandra L Gerber, Ana Paula de C Guimarães, Douglas Terra Machado, Cassia Alves, Diana Mariani, Thais Felix Cruz, Mario Sergio Ribeiro, Silvia Carvalho, Flávio Dias da Silva, Marcio Henrique de Oliveira Garcia, Leandro Magalhães de Souza, Cristiane Gomes da Silva, Caio Luiz Pereira Ribeiro, Andréa Cony Cavalcanti, Claudia Maria Braga de Mello, Amilcar Tanuri, Ana Tereza R Vasconcelos |
| EPI_ISL_1911958, EPI_ISL_1911959, EPI_ISL_1911960, EPI_ISL_1911961, EPI_ISL_1911962, EPI_ISL_1911964 | Instituto de Biotecnologia - UNESP-Botucatu-SP | Instituto de Biotecnologia - UNESP-Botucatu-SP | Fábio Sossai Possebon; Leila Sabrina Ullmann; Cecília Artico Banho; Cíntia Bittar; Guilherme Campos; Helena Lage Ferreira; Jorge A. Petroli Marchesi; Lívia Sacchetto; Maisa C. Pereira Parra; Marília Moraes; Maurício L. Nogueira; Paula Rahal; Paulo Inacio da Costa; João Pessoa Araújo Jr. |
| EPI_ISL_1941583 | Laboratorio de Pesquisa em Virologia | Laboratorio de Pesquisa em Virologia | Cecília Artico Banho, Lívia Sacchetto, Fábio Sossai Possebon, Leila Sabrina Ullmann, Cíntia Bittar, Guilherme Campos, Helena Lage Ferreira, Jorge A. Petroli Marchesi, Maisa C. Pereira Parra, Marília Moraes, Paula Rahal, Paulo Inacio da Costa, João Pessoa Araújo Jr., Maurício Lacerda Nogueira. |
| EPI_ISL_1966060, EPI_ISL_1966061, EPI_ISL_1966063, EPI_ISL_1966064, | VIGILANCIA EPIDEMIOLOGICA | Instituto Butantan / Mendelics | Instituto Butantan: Dimas Tadeu Covas, Sandra Coccuzzo Sampaio, Maria Carolina Elias, José Salvatore Leister Patané, Vincent Louis Viala, Antonio Jorge Martins, Ricardo Haddad, Claudia Renata dos Santos Barros, Elaine Cristina Marqueze, Raul Machado Neto, Debora Botequio Moretti, Jardelina de |

| | | | |
|---|---|---|---|
| EPI_ISL_1966065, EPI_ISL_1966066, EPI_ISL_1966068 | | | Souza Todao Bernardino, Loyze Paola Oliveira de Lima, Luiz Aurelio de Campos Crispin. Centro de Genômica Funcional da ESALQ: Luiz Lehmann Coutinho, Ricardo Augusto Brassaloti, Raquel de Lello Rocha Campos Cassano. NGS Soluções Genômicas: Pilar Drummond Sampaio Corrêa Mariani. FZEA-USP Pirassununga: Mirele Daiana Poleti, Jessika Cristina Chagas Lesbon, Elisangela Chicaroni Mattos, Heidge Fukumasu. USP-Botucatu: Rejane Maria Tommasini Grotto, Jayme A. Souza-Neto, Guilherme Targino Valente, Patricia Akemi Assato, Felipe Allan da Silva da Costa, Bianca Cechetto Carlos. Mendelics: Bibiana Santos, João Paulo Kitajima, Erika Freitas, David Schlesinger. Hemocentro Ribeirão Preto: Simone Kashima, Evandra Strazza Rodrigues, Svetoslav Nanev Slavov, Elaine Vieira dos Santos, Rafael dos Santos Bezerra, Luiz Carlos Junior de Alcantara, Marta Giovanetti, Vagner Fonseca, Flavia Aburjaile, Rodrigo Tocantins Calado. FAMERP-SJRP: Cecília Artico Banho, Lívia Sacchetto, Fábio Sossai Possebon, Leila Sabrina Ullmann, Cíntia Bittar, Guilherme Campos, Helena Lage Ferreira, Jorge A. Petroli Marchesi, Maisa C. Pereira Parra, Marília Moraes, Paula Rahal, Paulo Inacio da Costa, João Pessoa Araújo Jr., Maurício Lacerda Nogueira. Prefeitura de Sao Paulo: Melissa Palmieri. |
| EPI_ISL_1966070, EPI_ISL_1966071, EPI_ISL_1966072, EPI_ISL_1966077, EPI_ISL_1966078 | SECAO CENTRO DE DIAGNOSTICO SECEDI | Instituto Butantan / Mendelics | Instituto Butantan: Dimas Tadeu Covas, Sandra Coccuzzo Sampaio, Maria Carolina Elias, José Salvatore Leister Patané, Vincent Louis Viala, Antonio Jorge Martins, Ricardo Haddad, Claudia Renata dos Santos Barros, Elaine Cristina Marqueze, Raul Machado Neto, Debora Botequio Moretti, Jardelina de Souza Todao Bernardino, Loyze Paola Oliveira de Lima, Luiz Aurelio de Campos Crispin. Centro de Genômica Funcional da ESALQ: Luiz Lehmann Coutinho, Ricardo Augusto Brassaloti, Raquel de Lello Rocha Campos Cassano. NGS Soluções Genômicas: Pilar Drummond Sampaio Corrêa Mariani. FZEA-USP Pirassununga: Mirele Daiana Poleti, Jessika Cristina Chagas Lesbon, Elisangela Chicaroni Mattos, Heidge Fukumasu. USP-Botucatu: Rejane Maria Tommasini Grotto, Jayme A. Souza-Neto, Guilherme Targino Valente, Patricia Akemi Assato, Felipe Allan da Silva da Costa, Bianca Cechetto Carlos. Mendelics: Bibiana Santos, João Paulo Kitajima, Erika Freitas, David Schlesinger. Hemocentro Ribeirão Preto: Simone Kashima, Evandra Strazza Rodrigues, Svetoslav Nanev Slavov, Elaine Vieira dos Santos, Rafael dos Santos Bezerra, Luiz Carlos Junior de Alcantara, Marta Giovanetti, Vagner Fonseca, Flavia Aburjaile, Rodrigo Tocantins Calado. FAMERP-SJRP: Cecília Artico Banho, Lívia Sacchetto, Fábio Sossai Possebon, Leila Sabrina Ullmann, Cíntia Bittar, Guilherme Campos, Helena Lage Ferreira, Jorge A. Petroli Marchesi, Maisa C. Pereira Parra, Marília Moraes, Paula Rahal, Paulo Inacio da Costa, João Pessoa Araújo Jr., Maurício Lacerda Nogueira. Prefeitura de Sao Paulo: Melissa Palmieri. |
| EPI_ISL_1966080, EPI_ISL_1966081, EPI_ISL_1966083, EPI_ISL_1966084, EPI_ISL_1966085, EPI_ISL_1966086, EPI_ISL_1966087, EPI_ISL_1966088, EPI_ISL_1966089, EPI_ISL_1966090, EPI_ISL_1966092 | | | |
| see above | VIGILANCIA EPIDEMIOLOGICA | Instituto Butantan / Mendelics | Instituto Butantan: Dimas Tadeu Covas, Sandra Coccuzzo Sampaio, Maria Carolina Elias, José Salvatore Leister Patané, Vincent Louis Viala, Antonio Jorge Martins, Ricardo Haddad, Claudia Renata dos Santos Barros, Elaine Cristina Marqueze, Raul Machado Neto, Debora Botequio Moretti, Jardelina de Souza Todao Bernardino, Loyze Paola Oliveira de Lima, Luiz Aurelio de Campos Crispin. Centro de Genômica Funcional da ESALQ: Luiz Lehmann Coutinho, Ricardo Augusto Brassaloti, Raquel de Lello Rocha Campos Cassano. NGS Soluções Genômicas: Pilar Drummond Sampaio Corrêa Mariani. FZEA-USP Pirassununga: Mirele Daiana Poleti, Jessika Cristina Chagas Lesbon, Elisangela Chicaroni Mattos, Heidge Fukumasu. USP-Botucatu: Rejane Maria Tommasini Grotto, Jayme A. Souza-Neto, Guilherme Targino Valente, Patricia Akemi Assato, Felipe Allan da Silva da Costa, Bianca Cechetto Carlos. Mendelics: Bibiana Santos, João Paulo Kitajima, Erika Freitas, David Schlesinger. Hemocentro Ribeirão Preto: Simone Kashima, Evandra Strazza Rodrigues, Svetoslav Nanev Slavov, Elaine Vieira dos Santos, Rafael dos Santos Bezerra, Luiz Carlos Junior de Alcantara, Marta Giovanetti, Vagner Fonseca, Flavia Aburjaile, Rodrigo Tocantins Calado. FAMERP-SJRP: Cecília Artico Banho, Lívia Sacchetto, Fábio Sossai Possebon, Leila Sabrina Ullmann, Cíntia Bittar, Guilherme Campos, Helena Lage Ferreira, Jorge A. Petroli Marchesi, Maisa C. Pereira Parra, Marília Moraes, Paula Rahal, Paulo Inacio da Costa, João Pessoa Araújo Jr., Maurício Lacerda Nogueira. Prefeitura de Sao Paulo: Melissa Palmieri. |
| EPI_ISL_1966093 | SERV DE VIG SANITARIA EPIDEMIO E CTRL DE ZOONOZES GUARUJA | Instituto Butantan / Mendelics | Instituto Butantan: Dimas Tadeu Covas, Sandra Coccuzzo Sampaio, Maria Carolina Elias, José Salvatore Leister Patané, Vincent Louis Viala, Antonio Jorge Martins, Ricardo Haddad, Claudia Renata dos Santos Barros, Elaine Cristina Marqueze, Raul Machado Neto, Debora Botequio Moretti, Jardelina de Souza Todao Bernardino, Loyze Paola Oliveira de Lima, Luiz Aurelio de Campos Crispin. Centro de Genômica Funcional da ESALQ: Luiz Lehmann Coutinho, Ricardo Augusto Brassaloti, Raquel de Lello Rocha Campos Cassano. NGS Soluções Genômicas: Pilar Drummond Sampaio Corrêa Mariani. FZEA-USP Pirassununga: Mirele Daiana Poleti, Jessika Cristina Chagas Lesbon, Elisangela Chicaroni Mattos, Heidge Fukumasu. USP-Botucatu: Rejane Maria Tommasini Grotto, Jayme A. Souza-Neto, Guilherme Targino Valente, Patricia Akemi Assato, Felipe Allan da Silva da Costa, Bianca Cechetto Carlos. Mendelics: Bibiana Santos, João Paulo Kitajima, Erika Freitas, David Schlesinger. Hemocentro Ribeirão Preto: Simone Kashima, Evandra Strazza Rodrigues, Svetoslav Nanev Slavov, Elaine Vieira dos Santos, Rafael dos Santos Bezerra, Luiz Carlos Junior de Alcantara, Marta Giovanetti, Vagner Fonseca, Flavia Aburjaile, Rodrigo Tocantins Calado. FAMERP-SJRP: Cecília Artico Banho, Lívia Sacchetto, Fábio Sossai Possebon, Leila Sabrina Ullmann, Cíntia Bittar, Guilherme Campos, Helena Lage Ferreira, Jorge A. Petroli Marchesi, Maisa C. Pereira Parra, Marília Moraes, Paula Rahal, Paulo Inacio da Costa, João Pessoa Araújo Jr., Maurício Lacerda Nogueira. Prefeitura de Sao Paulo: Melissa Palmieri. |
| EPI_ISL_1966095, EPI_ISL_1966096, EPI_ISL_1966097, EPI_ISL_1966098 | AMBULATORIO MEDICO DE ESPECIALIDADES DE PERUIBE | Instituto Butantan / Mendelics | Instituto Butantan: Dimas Tadeu Covas, Sandra Coccuzzo Sampaio, Maria Carolina Elias, José Salvatore Leister Patané, Vincent Louis Viala, Antonio Jorge Martins, Ricardo Haddad, Claudia Renata dos Santos Barros, Elaine Cristina Marqueze, Raul Machado Neto, Debora Botequio Moretti, Jardelina de Souza Todao Bernardino, Loyze Paola Oliveira de Lima, Luiz Aurelio de Campos Crispin. Centro de Genômica Funcional da ESALQ: Luiz Lehmann Coutinho, Ricardo Augusto Brassaloti, Raquel de Lello Rocha Campos Cassano. NGS Soluções Genômicas: Pilar Drummond Sampaio Corrêa Mariani. FZEA-USP Pirassununga: Mirele Daiana Poleti, Jessika Cristina Chagas Lesbon, Elisangela Chicaroni Mattos, Heidge Fukumasu. USP-Botucatu: Rejane Maria Tommasini Grotto, Jayme A. Souza-Neto, Guilherme Targino Valente, Patricia Akemi Assato, Felipe Allan da Silva da Costa, Bianca Cechetto Carlos. Mendelics: Bibiana Santos, João Paulo Kitajima, Erika Freitas, David Schlesinger. Hemocentro Ribeirão Preto: Simone Kashima, Evandra Strazza Rodrigues, Svetoslav Nanev Slavov, Elaine Vieira dos Santos, Rafael dos Santos Bezerra, Luiz Carlos Junior de Alcantara, Marta Giovanetti, Vagner Fonseca, Flavia Aburjaile, Rodrigo Tocantins Calado. FAMERP-SJRP: Cecília Artico Banho, Lívia Sacchetto, Fábio Sossai Possebon, Leila Sabrina Ullmann, Cíntia Bittar, Guilherme Campos, Helena Lage Ferreira, Jorge A. Petroli Marchesi, Maisa C. Pereira Parra, Marília Moraes, Paula Rahal, Paulo Inacio da Costa, João Pessoa Araújo Jr., Maurício Lacerda Nogueira. Prefeitura de Sao Paulo: Melissa Palmieri. |
| EPI_ISL_1966102 | USF TRES PONTES | Instituto Butantan / Mendelics | Instituto Butantan: Dimas Tadeu Covas, Sandra Coccuzzo Sampaio, Maria Carolina Elias, José Salvatore Leister Patané, Vincent Louis Viala, Antonio Jorge Martins, Ricardo Haddad, Claudia Renata dos Santos Barros, Elaine Cristina Marqueze, Raul Machado Neto, Debora Botequio Moretti, Jardelina de Souza Todao Bernardino, Loyze Paola Oliveira de Lima, Luiz Aurelio de Campos Crispin. Centro de Genômica Funcional da ESALQ: Luiz Lehmann Coutinho, Ricardo Augusto Brassaloti, Raquel de Lello Rocha Campos Cassano. NGS Soluções Genômicas: Pilar Drummond Sampaio Corrêa Mariani. FZEA-USP Pirassununga: Mirele Daiana Poleti, Jessika Cristina Chagas Lesbon, Elisangela Chicaroni Mattos, Heidge Fukumasu. USP-Botucatu: Rejane Maria Tommasini Grotto, Jayme A. Souza-Neto, Guilherme Targino Valente, Patricia Akemi Assato, Felipe Allan da Silva da Costa, Bianca Cechetto Carlos. Mendelics: Bibiana Santos, João Paulo Kitajima, Erika Freitas, David Schlesinger. Hemocentro Ribeirão Preto: Simone Kashima, Evandra Strazza Rodrigues, Svetoslav Nanev Slavov, Elaine Vieira dos Santos, Rafael dos Santos Bezerra, Luiz Carlos Junior de Alcantara, Marta Giovanetti, Vagner Fonseca, Flavia Aburjaile, Rodrigo Tocantins Calado. FAMERP-SJRP: Cecília Artico Banho, Lívia Sacchetto, Fábio Sossai Possebon, Leila Sabrina Ullmann, Cíntia Bittar, Guilherme Campos, Helena Lage Ferreira, Jorge A. Petroli Marchesi, Maisa C. Pereira Parra, Marília Moraes, Paula Rahal, Paulo Inacio da Costa, João Pessoa Araújo Jr., Maurício Lacerda Nogueira. Prefeitura de Sao Paulo: Melissa Palmieri. |
| EPI_ISL_1966103 | SMS SECRETARIA MUNICIPAL DE SAUDE DE BOITUVA | Instituto Butantan / Mendelics | Instituto Butantan: Dimas Tadeu Covas, Sandra Coccuzzo Sampaio, Maria Carolina Elias, José Salvatore Leister Patané, Vincent Louis Viala, Antonio Jorge Martins, Ricardo Haddad, Claudia Renata dos Santos Barros, Elaine Cristina Marqueze, Raul Machado Neto, Debora Botequio Moretti, Jardelina de Souza Todao Bernardino, Loyze Paola Oliveira de Lima, Luiz Aurelio de Campos Crispin. Centro de Genômica Funcional da ESALQ: Luiz Lehmann Coutinho, Ricardo Augusto Brassaloti, Raquel de Lello Rocha Campos Cassano. NGS Soluções Genômicas: Pilar Drummond Sampaio Corrêa Mariani. FZEA-USP Pirassununga: Mirele Daiana Poleti, Jessika Cristina Chagas Lesbon, Elisangela Chicaroni Mattos, Heidge Fukumasu. USP-Botucatu: Rejane Maria Tommasini Grotto, Jayme A. Souza-Neto, Guilherme Targino Valente, Patricia Akemi Assato, Felipe Allan da Silva da Costa, Bianca Cechetto Carlos. Mendelics: Bibiana Santos, João Paulo Kitajima, Erika Freitas, David Schlesinger. Hemocentro Ribeirão Preto: Simone Kashima, Evandra Strazza Rodrigues, Svetoslav Nanev Slavov, Elaine Vieira dos Santos, Rafael dos Santos Bezerra, Luiz Carlos Junior de Alcantara, Marta Giovanetti, Vagner Fonseca, Flavia Aburjaile, Rodrigo Tocantins Calado. FAMERP-SJRP: Cecília Artico Banho, Lívia Sacchetto, Fábio Sossai Possebon, Leila Sabrina Ullmann, Cíntia Bittar, Guilherme Campos, Helena Lage Ferreira, Jorge A. Petroli Marchesi, Maisa C. Pereira Parra, Marília Moraes, Paula Rahal, Paulo Inacio da Costa, João Pessoa Araújo Jr., Maurício Lacerda Nogueira. Prefeitura de Sao Paulo: Melissa Palmieri. |
| EPI_ISL_1966107 | SECRETARIA MUNICIPAL DE SAUDE SOROCABA | Instituto Butantan / Mendelics | Instituto Butantan: Dimas Tadeu Covas, Sandra Coccuzzo Sampaio, Maria Carolina Elias, José Salvatore Leister Patané, Vincent Louis Viala, Antonio Jorge Martins, Ricardo Haddad, Claudia Renata dos Santos Barros, Elaine Cristina Marqueze, Raul Machado Neto, Debora Botequio Moretti, Jardelina de Souza Todao Bernardino, Loyze Paola Oliveira de Lima, Luiz Aurelio de Campos Crispin. Centro de Genômica Funcional da ESALQ: Luiz Lehmann Coutinho, Ricardo Augusto Brassaloti, Raquel de Lello Rocha Campos Cassano. NGS Soluções Genômicas: Pilar Drummond Sampaio Corrêa Mariani. FZEA-USP Pirassununga: Mirele Daiana Poleti, Jessika Cristina Chagas Lesbon, Elisangela Chicaroni Mattos, Heidge Fukumasu. USP-Botucatu: Rejane Maria Tommasini Grotto, Jayme A. Souza-Neto, Guilherme Targino Valente, Patricia Akemi Assato, Felipe Allan da Silva da Costa, Bianca Cechetto Carlos. Mendelics: Bibiana Santos, João Paulo Kitajima, Erika Freitas, David Schlesinger. Hemocentro Ribeirão Preto: Simone Kashima, Evandra Strazza |

| | | | |
|---|---|---|---|
| | | | Rodrigues, Svetoslav Nanev Slavov, Elaine Vieira dos Santos, Rafael dos Santos Bezerra, Luiz Carlos Junior de Alcantara, Marta Giovanetti, Vagner Fonseca, Flavia Aburjaile, Rodrigo Tocantins Calado. FAMERP-SJRP: Cecília Artico Banho, Lívia Sacchetto, Fábio Sossai Possebon, Leila Sabrina Ullmann, Cíntia Bittar, Guilherme Campos, Helena Lage Ferreira, Jorge A. Petroli Marchesi, Maisa C. Pereira Parra, Marília Moraes, Paula Rahal, Paulo Inacio da Costa, João Pessoa Araújo Jr., Maurício Lacerda Nogueira. Prefeitura de Sao Paulo: Melissa Palmieri. |
| EPI_ISL_1966109 | CENTRO DE SAUDE II DR GABRIEL MESQUITA VARGEM GDE DO SUL | Instituto Butantan / Mendelics | Instituto Butantan: Dimas Tadeu Covas, Sandra Coccuzzo Sampaio, Maria Carolina Elias, José Salvatore Leister Patané, Vincent Louis Viala, Antonio Jorge Martins, Ricardo Haddad, Claudia Renata dos Santos Barros, Elaine Cristina Marqueze, Raul Machado Neto, Debora Botequio Moretti, Jardelina de Souza Todao Bernardino, Loyze Paola Oliveira de Lima, Luiz Aurelio de Campos Crispin. Centro de Genômica Funcional da ESALQ: Luiz Lehmann Coutinho, Ricardo Augusto Brassaloti, Raquel de Lello Rocha Campos Cassano. NGS Soluções Genômicas: Pilar Drummond Sampaio Corrêa Mariani. FZEA-USP Pirassununga: Mirele Daiana Poleti, Jessika Cristina Chagas Lesbon, Elisangela Chicaroni Mattos, Heidge Fukumasu. USP-Botucatu: Rejane Maria Tommasini Grotto, Jayme A. Souza-Neto, Guilherme Targino Valente, Patricia Akemi Assato, Felipe Allan da Silva da Costa, Bianca Cechetto Carlos. Mendelics: Bibiana Santos, João Paulo Kitajima, Erika Freitas, David Schlesinger. Hemocentro Ribeirão Preto: Simone Kashima, Evandra Strazza Rodrigues, Svetoslav Nanev Slavov, Elaine Vieira dos Santos, Rafael dos Santos Bezerra, Luiz Carlos Junior de Alcantara, Marta Giovanetti, Vagner Fonseca, Flavia Aburjaile, Rodrigo Tocantins Calado. FAMERP-SJRP: Cecília Artico Banho, Lívia Sacchetto, Fábio Sossai Possebon, Leila Sabrina Ullmann, Cíntia Bittar, Guilherme Campos, Helena Lage Ferreira, Jorge A. Petroli Marchesi, Maisa C. Pereira Parra, Marília Moraes, Paula Rahal, Paulo Inacio da Costa, João Pessoa Araújo Jr., Maurício Lacerda Nogueira. Prefeitura de Sao Paulo: Melissa Palmieri. |
| EPI_ISL_1966113, EPI_ISL_1966114, EPI_ISL_1966116 | CS II EGIDIO BRUNHARA MORRO AGUDO | Instituto Butantan / Mendelics | Instituto Butantan: Dimas Tadeu Covas, Sandra Coccuzzo Sampaio, Maria Carolina Elias, José Salvatore Leister Patané, Vincent Louis Viala, Antonio Jorge Martins, Ricardo Haddad, Claudia Renata dos Santos Barros, Elaine Cristina Marqueze, Raul Machado Neto, Debora Botequio Moretti, Jardelina de Souza Todao Bernardino, Loyze Paola Oliveira de Lima, Luiz Aurelio de Campos Crispin. Centro de Genômica Funcional da ESALQ: Luiz Lehmann Coutinho, Ricardo Augusto Brassaloti, Raquel de Lello Rocha Campos Cassano. NGS Soluções Genômicas: Pilar Drummond Sampaio Corrêa Mariani. FZEA-USP Pirassununga: Mirele Daiana Poleti, Jessika Cristina Chagas Lesbon, Elisangela Chicaroni Mattos, Heidge Fukumasu. USP-Botucatu: Rejane Maria Tommasini Grotto, Jayme A. Souza-Neto, Guilherme Targino Valente, Patricia Akemi Assato, Felipe Allan da Silva da Costa, Bianca Cechetto Carlos. Mendelics: Bibiana Santos, João Paulo Kitajima, Erika Freitas, David Schlesinger. Hemocentro Ribeirão Preto: Simone Kashima, Evandra Strazza Rodrigues, Svetoslav Nanev Slavov, Elaine Vieira dos Santos, Rafael dos Santos Bezerra, Luiz Carlos Junior de Alcantara, Marta Giovanetti, Vagner Fonseca, Flavia Aburjaile, Rodrigo Tocantins Calado. FAMERP-SJRP: Cecília Artico Banho, Lívia Sacchetto, Fábio Sossai Possebon, Leila Sabrina Ullmann, Cíntia Bittar, Guilherme Campos, Helena Lage Ferreira, Jorge A. Petroli Marchesi, Maisa C. Pereira Parra, Marília Moraes, Paula Rahal, Paulo Inacio da Costa, João Pessoa Araújo Jr., Maurício Lacerda Nogueira. Prefeitura de Sao Paulo: Melissa Palmieri. |
| EPI_ISL_1966118, EPI_ISL_1966119, EPI_ISL_1966121, EPI_ISL_1966122, EPI_ISL_1966123 | CENTRO DE REFERENCIA DO IDOSO DR HUMBERTO MENDES DE CARVALHO | Instituto Butantan / Mendelics | Instituto Butantan: Dimas Tadeu Covas, Sandra Coccuzzo Sampaio, Maria Carolina Elias, José Salvatore Leister Patané, Vincent Louis Viala, Antonio Jorge Martins, Ricardo Haddad, Claudia Renata dos Santos Barros, Elaine Cristina Marqueze, Raul Machado Neto, Debora Botequio Moretti, Jardelina de Souza Todao Bernardino, Loyze Paola Oliveira de Lima, Luiz Aurelio de Campos Crispin. Centro de Genômica Funcional da ESALQ: Luiz Lehmann Coutinho, Ricardo Augusto Brassaloti, Raquel de Lello Rocha Campos Cassano. NGS Soluções Genômicas: Pilar Drummond Sampaio Corrêa Mariani. FZEA-USP Pirassununga: Mirele Daiana Poleti, Jessika Cristina Chagas Lesbon, Elisangela Chicaroni Mattos, Heidge Fukumasu. USP-Botucatu: Rejane Maria Tommasini Grotto, Jayme A. Souza-Neto, Guilherme Targino Valente, Patricia Akemi Assato, Felipe Allan da Silva da Costa, Bianca Cechetto Carlos. Mendelics: Bibiana Santos, João Paulo Kitajima, Erika Freitas, David Schlesinger. Hemocentro Ribeirão Preto: Simone Kashima, Evandra Strazza Rodrigues, Svetoslav Nanev Slavov, Elaine Vieira dos Santos, Rafael dos Santos Bezerra, Luiz Carlos Junior de Alcantara, Marta Giovanetti, Vagner Fonseca, Flavia Aburjaile, Rodrigo Tocantins Calado. FAMERP-SJRP: Cecília Artico Banho, Lívia Sacchetto, Fábio Sossai Possebon, Leila Sabrina Ullmann, Cíntia Bittar, Guilherme Campos, Helena Lage Ferreira, Jorge A. Petroli Marchesi, Maisa C. Pereira Parra, Marília Moraes, Paula Rahal, Paulo Inacio da Costa, João Pessoa Araújo Jr., Maurício Lacerda Nogueira. Prefeitura de Sao Paulo: Melissa Palmieri. |
| EPI_ISL_1966125 | SECRETARIA MUNICIPAL DE SAUDE SOROCABA | Instituto Butantan / Mendelics | Instituto Butantan: Dimas Tadeu Covas, Sandra Coccuzzo Sampaio, Maria Carolina Elias, José Salvatore Leister Patané, Vincent Louis Viala, Antonio Jorge Martins, Ricardo Haddad, Claudia Renata dos Santos Barros, Elaine Cristina Marqueze, Raul Machado Neto, Debora Botequio Moretti, Jardelina de Souza Todao Bernardino, Loyze Paola Oliveira de Lima, Luiz Aurelio de Campos Crispin. Centro de Genômica Funcional da ESALQ: Luiz Lehmann Coutinho, Ricardo Augusto Brassaloti, Raquel de Lello Rocha Campos Cassano. NGS Soluções Genômicas: Pilar Drummond Sampaio Corrêa Mariani. FZEA-USP Pirassununga: Mirele Daiana Poleti, Jessika Cristina Chagas Lesbon, Elisangela Chicaroni Mattos, Heidge Fukumasu. USP-Botucatu: Rejane Maria Tommasini Grotto, Jayme A. Souza-Neto, Guilherme Targino Valente, Patricia Akemi Assato, Felipe Allan da Silva da Costa, Bianca Cechetto Carlos. Mendelics: Bibiana Santos, João Paulo Kitajima, Erika Freitas, David Schlesinger. Hemocentro Ribeirão Preto: Simone Kashima, Evandra Strazza Rodrigues, Svetoslav Nanev Slavov, Elaine Vieira dos Santos, Rafael dos Santos Bezerra, Luiz Carlos Junior de Alcantara, Marta Giovanetti, Vagner Fonseca, Flavia Aburjaile, Rodrigo Tocantins Calado. FAMERP-SJRP: Cecília Artico Banho, Lívia Sacchetto, Fábio Sossai Possebon, Leila Sabrina Ullmann, Cíntia Bittar, Guilherme Campos, Helena Lage Ferreira, Jorge A. Petroli Marchesi, Maisa C. Pereira Parra, Marília Moraes, Paula Rahal, Paulo Inacio da Costa, João Pessoa Araújo Jr., Maurício Lacerda Nogueira. Prefeitura de Sao Paulo: Melissa Palmieri. |
| EPI_ISL_1966126, EPI_ISL_1966128 | SAE SERVICO DE ATENDIMENTO ESPECIALIZADO | Instituto Butantan / Mendelics | Instituto Butantan: Dimas Tadeu Covas, Sandra Coccuzzo Sampaio, Maria Carolina Elias, José Salvatore Leister Patané, Vincent Louis Viala, Antonio Jorge Martins, Ricardo Haddad, Claudia Renata dos Santos Barros, Elaine Cristina Marqueze, Raul Machado Neto, Debora Botequio Moretti, Jardelina de Souza Todao Bernardino, Loyze Paola Oliveira de Lima, Luiz Aurelio de Campos Crispin. Centro de Genômica Funcional da ESALQ: Luiz Lehmann Coutinho, Ricardo Augusto Brassaloti, Raquel de Lello Rocha Campos Cassano. NGS Soluções Genômicas: Pilar Drummond Sampaio Corrêa Mariani. FZEA-USP Pirassununga: Mirele Daiana Poleti, Jessika Cristina Chagas Lesbon, Elisangela Chicaroni Mattos, Heidge Fukumasu. USP-Botucatu: Rejane Maria Tommasini Grotto, Jayme A. Souza-Neto, Guilherme Targino Valente, Patricia Akemi Assato, Felipe Allan da Silva da Costa, Bianca Cechetto Carlos. Mendelics: Bibiana Santos, João Paulo Kitajima, Erika Freitas, David Schlesinger. Hemocentro Ribeirão Preto: Simone Kashima, Evandra Strazza Rodrigues, Svetoslav Nanev Slavov, Elaine Vieira dos Santos, Rafael dos Santos Bezerra, Luiz Carlos Junior de Alcantara, Marta Giovanetti, Vagner Fonseca, Flavia Aburjaile, Rodrigo Tocantins Calado. FAMERP-SJRP: Cecília Artico Banho, Lívia Sacchetto, Fábio Sossai Possebon, Leila Sabrina Ullmann, Cíntia Bittar, Guilherme Campos, Helena Lage Ferreira, Jorge A. Petroli Marchesi, Maisa C. Pereira Parra, Marília Moraes, Paula Rahal, Paulo Inacio da Costa, João Pessoa Araújo Jr., Maurício Lacerda Nogueira. Prefeitura de Sao Paulo: Melissa Palmieri. |
| EPI_ISL_1966129 | PRONTO SOCORRO MUNICIPAL DE TATUI | Instituto Butantan / Mendelics | Instituto Butantan: Dimas Tadeu Covas, Sandra Coccuzzo Sampaio, Maria Carolina Elias, José Salvatore Leister Patané, Vincent Louis Viala, Antonio Jorge Martins, Ricardo Haddad, Claudia Renata dos Santos Barros, Elaine Cristina Marqueze, Raul Machado Neto, Debora Botequio Moretti, Jardelina de Souza Todao Bernardino, Loyze Paola Oliveira de Lima, Luiz Aurelio de Campos Crispin. Centro de Genômica Funcional da ESALQ: Luiz Lehmann Coutinho, Ricardo Augusto Brassaloti, Raquel de Lello Rocha Campos Cassano. NGS Soluções Genômicas: Pilar Drummond Sampaio Corrêa Mariani. FZEA-USP Pirassununga: Mirele Daiana Poleti, Jessika Cristina Chagas Lesbon, Elisangela Chicaroni Mattos, Heidge Fukumasu. USP-Botucatu: Rejane Maria Tommasini Grotto, Jayme A. Souza-Neto, Guilherme Targino Valente, Patricia Akemi Assato, Felipe Allan da Silva da Costa, Bianca Cechetto Carlos. Mendelics: Bibiana Santos, João Paulo Kitajima, Erika Freitas, David Schlesinger. Hemocentro Ribeirão Preto: Simone Kashima, Evandra Strazza Rodrigues, Svetoslav Nanev Slavov, Elaine Vieira dos Santos, Rafael dos Santos Bezerra, Luiz Carlos Junior de Alcantara, Marta Giovanetti, Vagner Fonseca, Flavia Aburjaile, Rodrigo Tocantins Calado. FAMERP-SJRP: Cecília Artico Banho, Lívia Sacchetto, Fábio Sossai Possebon, Leila Sabrina Ullmann, Cíntia Bittar, Guilherme Campos, Helena Lage Ferreira, Jorge A. Petroli Marchesi, Maisa C. Pereira Parra, Marília Moraes, Paula Rahal, Paulo Inacio da Costa, João Pessoa Araújo Jr., Maurício Lacerda Nogueira. Prefeitura de Sao Paulo: Melissa Palmieri. |
| EPI_ISL_1966133, EPI_ISL_1966134, EPI_ISL_1966135, EPI_ISL_1966137, EPI_ISL_1966138, EPI_ISL_1966139, EPI_ISL_1966140, EPI_ISL_1966142 | PRONTO ATENDIMENTO VILA PADRE ANCHIETA | Instituto Butantan / Mendelics | Instituto Butantan: Dimas Tadeu Covas, Sandra Coccuzzo Sampaio, Maria Carolina Elias, José Salvatore Leister Patané, Vincent Louis Viala, Antonio Jorge Martins, Ricardo Haddad, Claudia Renata dos Santos Barros, Elaine Cristina Marqueze, Raul Machado Neto, Debora Botequio Moretti, Jardelina de Souza Todao Bernardino, Loyze Paola Oliveira de Lima, Luiz Aurelio de Campos Crispin. Centro de Genômica Funcional da ESALQ: Luiz Lehmann Coutinho, Ricardo Augusto Brassaloti, Raquel de Lello Rocha Campos Cassano. NGS Soluções Genômicas: Pilar Drummond Sampaio Corrêa Mariani. FZEA-USP Pirassununga: Mirele Daiana Poleti, Jessika Cristina Chagas Lesbon, Elisangela Chicaroni Mattos, Heidge Fukumasu. USP-Botucatu: Rejane Maria Tommasini Grotto, Jayme A. Souza-Neto, Guilherme Targino Valente, Patricia Akemi Assato, Felipe Allan da Silva da Costa, Bianca Cechetto Carlos. Mendelics: Bibiana Santos, João Paulo Kitajima, Erika Freitas, David Schlesinger. Hemocentro Ribeirão Preto: Simone Kashima, Evandra Strazza Rodrigues, Svetoslav Nanev Slavov, Elaine Vieira dos Santos, Rafael dos Santos Bezerra, Luiz Carlos Junior de Alcantara, Marta Giovanetti, Vagner Fonseca, Flavia Aburjaile, Rodrigo Tocantins Calado. FAMERP-SJRP: Cecília Artico Banho, Lívia Sacchetto, Fábio Sossai Possebon, Leila Sabrina Ullmann, Cíntia Bittar, Guilherme Campos, Helena Lage Ferreira, Jorge A. Petroli Marchesi, Maisa C. Pereira Parra, Marília Moraes, Paula Rahal, Paulo Inacio da Costa, João Pessoa Araújo Jr., Maurício Lacerda Nogueira. Prefeitura de Sao Paulo: Melissa Palmieri. |
| EPI_ISL_1966145, EPI_ISL_1966146 | COMPLEXO HOSPITALAR OURO VERDE DE CAMPINAS | Instituto Butantan / Mendelics | Instituto Butantan: Dimas Tadeu Covas, Sandra Coccuzzo Sampaio, Maria Carolina Elias, José Salvatore Leister Patané, Vincent Louis Viala, Antonio Jorge Martins, Ricardo Haddad, Claudia Renata dos Santos Barros, Elaine Cristina Marqueze, Raul Machado Neto, Debora Botequio Moretti, Jardelina de |

| | | | |
|---|---|---|---|
| | | | Souza Todao Bernardino, Loyze Paola Oliveira de Lima, Luiz Aurelio de Campos Crispin. Centro de Genômica Funcional da ESALQ: Luiz Lehmann Coutinho, Ricardo Augusto Brassaloti, Raquel de Lello Rocha Campos Cassano. NGS Soluções Genômicas: Pilar Drummond Sampaio Corrêa Mariani. FZEA-USP Pirassununga: Mirele Daiana Poleti, Jessika Cristina Chagas Lesbon, Elisangela Chicaroni Mattos, Heidge Fukumasu. USP-Botucatu: Rejane Maria Tommasini Grotto, Jayme A. Souza-Neto, Guilherme Targino Valente, Patricia Akemi Assato, Felipe Allan da Silva da Costa, Bianca Cechetto Carlos. Mendelics: Bibiana Santos, João Paulo Kitajima, Erika Freitas, David Schlesinger. Hemocentro Ribeirão Preto: Simone Kashima, Evandra Strazza Rodrigues, Svetoslav Nanev Slavov, Elaine Vieira dos Santos, Rafael dos Santos Bezerra, Luiz Carlos Junior de Alcantara, Marta Giovanetti, Vagner Fonseca, Flavia Aburjaile, Rodrigo Tocantins Calado. FAMERP-SJRP: Cecília Artico Banho, Lívia Sacchetto, Fábio Sossai Possebon, Leila Sabrina Ullmann, Cíntia Bittar, Guilherme Campos, Helena Lage Ferreira, Jorge A. Petroli Marchesi, Maisa C. Pereira Parra, Marília Moraes, Paula Rahal, Paulo Inacio da Costa, João Pessoa Araújo Jr., Maurício Lacerda Nogueira. Prefeitura de Sao Paulo: Melissa Palmieri. |
| EPI_ISL_1966147 | HOSPITAL REGIONAL DE ITAPETININGA | Instituto Butantan / Mendelics | Instituto Butantan: Dimas Tadeu Covas, Sandra Coccuzzo Sampaio, Maria Carolina Elias, José Salvatore Leister Patané, Vincent Louis Viala, Antonio Jorge Martins, Ricardo Haddad, Claudia Renata dos Santos Barros, Elaine Cristina Marqueze, Raul Machado Neto, Debora Botequio Moretti, Jardelina de Souza Todao Bernardino, Loyze Paola Oliveira de Lima, Luiz Aurelio de Campos Crispin. Centro de Genômica Funcional da ESALQ: Luiz Lehmann Coutinho, Ricardo Augusto Brassaloti, Raquel de Lello Rocha Campos Cassano. NGS Soluções Genômicas: Pilar Drummond Sampaio Corrêa Mariani. FZEA-USP Pirassununga: Mirele Daiana Poleti, Jessika Cristina Chagas Lesbon, Elisangela Chicaroni Mattos, Heidge Fukumasu. USP-Botucatu: Rejane Maria Tommasini Grotto, Jayme A. Souza-Neto, Guilherme Targino Valente, Patricia Akemi Assato, Felipe Allan da Silva da Costa, Bianca Cechetto Carlos. Mendelics: Bibiana Santos, João Paulo Kitajima, Erika Freitas, David Schlesinger. Hemocentro Ribeirão Preto: Simone Kashima, Evandra Strazza Rodrigues, Svetoslav Nanev Slavov, Elaine Vieira dos Santos, Rafael dos Santos Bezerra, Luiz Carlos Junior de Alcantara, Marta Giovanetti, Vagner Fonseca, Flavia Aburjaile, Rodrigo Tocantins Calado. FAMERP-SJRP: Cecília Artico Banho, Lívia Sacchetto, Fábio Sossai Possebon, Leila Sabrina Ullmann, Cíntia Bittar, Guilherme Campos, Helena Lage Ferreira, Jorge A. Petroli Marchesi, Maisa C. Pereira Parra, Marília Moraes, Paula Rahal, Paulo Inacio da Costa, João Pessoa Araújo Jr., Maurício Lacerda Nogueira. Prefeitura de Sao Paulo: Melissa Palmieri. |
| EPI_ISL_1966148, EPI_ISL_1966149, EPI_ISL_1966150, EPI_ISL_1966151, EPI_ISL_1966152, EPI_ISL_1966153 | SECRETARIA MUNICIPAL DE SAUDE DE PIRACAIA | Instituto Butantan / Mendelics | Instituto Butantan: Dimas Tadeu Covas, Sandra Coccuzzo Sampaio, Maria Carolina Elias, José Salvatore Leister Patané, Vincent Louis Viala, Antonio Jorge Martins, Ricardo Haddad, Claudia Renata dos Santos Barros, Elaine Cristina Marqueze, Raul Machado Neto, Debora Botequio Moretti, Jardelina de Souza Todao Bernardino, Loyze Paola Oliveira de Lima, Luiz Aurelio de Campos Crispin. Centro de Genômica Funcional da ESALQ: Luiz Lehmann Coutinho, Ricardo Augusto Brassaloti, Raquel de Lello Rocha Campos Cassano. NGS Soluções Genômicas: Pilar Drummond Sampaio Corrêa Mariani. FZEA-USP Pirassununga: Mirele Daiana Poleti, Jessika Cristina Chagas Lesbon, Elisangela Chicaroni Mattos, Heidge Fukumasu. USP-Botucatu: Rejane Maria Tommasini Grotto, Jayme A. Souza-Neto, Guilherme Targino Valente, Patricia Akemi Assato, Felipe Allan da Silva da Costa, Bianca Cechetto Carlos. Mendelics: Bibiana Santos, João Paulo Kitajima, Erika Freitas, David Schlesinger. Hemocentro Ribeirão Preto: Simone Kashima, Evandra Strazza Rodrigues, Svetoslav Nanev Slavov, Elaine Vieira dos Santos, Rafael dos Santos Bezerra, Luiz Carlos Junior de Alcantara, Marta Giovanetti, Vagner Fonseca, Flavia Aburjaile, Rodrigo Tocantins Calado. FAMERP-SJRP: Cecília Artico Banho, Lívia Sacchetto, Fábio Sossai Possebon, Leila Sabrina Ullmann, Cíntia Bittar, Guilherme Campos, Helena Lage Ferreira, Jorge A. Petroli Marchesi, Maisa C. Pereira Parra, Marília Moraes, Paula Rahal, Paulo Inacio da Costa, João Pessoa Araújo Jr., Maurício Lacerda Nogueira. Prefeitura de Sao Paulo: Melissa Palmieri. |
| EPI_ISL_1966154, EPI_ISL_1966155 | SECRETARIA MUNICIPAL DE SAUDE SOROCABA | Instituto Butantan / Mendelics | Instituto Butantan: Dimas Tadeu Covas, Sandra Coccuzzo Sampaio, Maria Carolina Elias, José Salvatore Leister Patané, Vincent Louis Viala, Antonio Jorge Martins, Ricardo Haddad, Claudia Renata dos Santos Barros, Elaine Cristina Marqueze, Raul Machado Neto, Debora Botequio Moretti, Jardelina de Souza Todao Bernardino, Loyze Paola Oliveira de Lima, Luiz Aurelio de Campos Crispin. Centro de Genômica Funcional da ESALQ: Luiz Lehmann Coutinho, Ricardo Augusto Brassaloti, Raquel de Lello Rocha Campos Cassano. NGS Soluções Genômicas: Pilar Drummond Sampaio Corrêa Mariani. FZEA-USP Pirassununga: Mirele Daiana Poleti, Jessika Cristina Chagas Lesbon, Elisangela Chicaroni Mattos, Heidge Fukumasu. USP-Botucatu: Rejane Maria Tommasini Grotto, Jayme A. Souza-Neto, Guilherme Targino Valente, Patricia Akemi Assato, Felipe Allan da Silva da Costa, Bianca Cechetto Carlos. Mendelics: Bibiana Santos, João Paulo Kitajima, Erika Freitas, David Schlesinger. Hemocentro Ribeirão Preto: Simone Kashima, Evandra Strazza Rodrigues, Svetoslav Nanev Slavov, Elaine Vieira dos Santos, Rafael dos Santos Bezerra, Luiz Carlos Junior de Alcantara, Marta Giovanetti, Vagner Fonseca, Flavia Aburjaile, Rodrigo Tocantins Calado. FAMERP-SJRP: Cecília Artico Banho, Lívia Sacchetto, Fábio Sossai Possebon, Leila Sabrina Ullmann, Cíntia Bittar, Guilherme Campos, Helena Lage Ferreira, Jorge A. Petroli Marchesi, Maisa C. Pereira Parra, Marília Moraes, Paula Rahal, Paulo Inacio da Costa, João Pessoa Araújo Jr., Maurício Lacerda Nogueira. Prefeitura de Sao Paulo: Melissa Palmieri. |
| EPI_ISL_1966156 | PRONTO ATENDIMENTO VILA PADRE ANCHIETA | Instituto Butantan / Mendelics | Instituto Butantan: Dimas Tadeu Covas, Sandra Coccuzzo Sampaio, Maria Carolina Elias, José Salvatore Leister Patané, Vincent Louis Viala, Antonio Jorge Martins, Ricardo Haddad, Claudia Renata dos Santos Barros, Elaine Cristina Marqueze, Raul Machado Neto, Debora Botequio Moretti, Jardelina de Souza Todao Bernardino, Loyze Paola Oliveira de Lima, Luiz Aurelio de Campos Crispin. Centro de Genômica Funcional da ESALQ: Luiz Lehmann Coutinho, Ricardo Augusto Brassaloti, Raquel de Lello Rocha Campos Cassano. NGS Soluções Genômicas: Pilar Drummond Sampaio Corrêa Mariani. FZEA-USP Pirassununga: Mirele Daiana Poleti, Jessika Cristina Chagas Lesbon, Elisangela Chicaroni Mattos, Heidge Fukumasu. USP-Botucatu: Rejane Maria Tommasini Grotto, Jayme A. Souza-Neto, Guilherme Targino Valente, Patricia Akemi Assato, Felipe Allan da Silva da Costa, Bianca Cechetto Carlos. Mendelics: Bibiana Santos, João Paulo Kitajima, Erika Freitas, David Schlesinger. Hemocentro Ribeirão Preto: Simone Kashima, Evandra Strazza Rodrigues, Svetoslav Nanev Slavov, Elaine Vieira dos Santos, Rafael dos Santos Bezerra, Luiz Carlos Junior de Alcantara, Marta Giovanetti, Vagner Fonseca, Flavia Aburjaile, Rodrigo Tocantins Calado. FAMERP-SJRP: Cecília Artico Banho, Lívia Sacchetto, Fábio Sossai Possebon, Leila Sabrina Ullmann, Cíntia Bittar, Guilherme Campos, Helena Lage Ferreira, Jorge A. Petroli Marchesi, Maisa C. Pereira Parra, Marília Moraes, Paula Rahal, Paulo Inacio da Costa, João Pessoa Araújo Jr., Maurício Lacerda Nogueira. Prefeitura de Sao Paulo: Melissa Palmieri. |
| EPI_ISL_1966158, EPI_ISL_1966159, EPI_ISL_1966160, EPI_ISL_1966161 | COMPLEXO HOSPITALAR OURO VERDE DE CAMPINAS | Instituto Butantan / Mendelics | Instituto Butantan: Dimas Tadeu Covas, Sandra Coccuzzo Sampaio, Maria Carolina Elias, José Salvatore Leister Patané, Vincent Louis Viala, Antonio Jorge Martins, Ricardo Haddad, Claudia Renata dos Santos Barros, Elaine Cristina Marqueze, Raul Machado Neto, Debora Botequio Moretti, Jardelina de Souza Todao Bernardino, Loyze Paola Oliveira de Lima, Luiz Aurelio de Campos Crispin. Centro de Genômica Funcional da ESALQ: Luiz Lehmann Coutinho, Ricardo Augusto Brassaloti, Raquel de Lello Rocha Campos Cassano. NGS Soluções Genômicas: Pilar Drummond Sampaio Corrêa Mariani. FZEA-USP Pirassununga: Mirele Daiana Poleti, Jessika Cristina Chagas Lesbon, Elisangela Chicaroni Mattos, Heidge Fukumasu. USP-Botucatu: Rejane Maria Tommasini Grotto, Jayme A. Souza-Neto, Guilherme Targino Valente, Patricia Akemi Assato, Felipe Allan da Silva da Costa, Bianca Cechetto Carlos. Mendelics: Bibiana Santos, João Paulo Kitajima, Erika Freitas, David Schlesinger. Hemocentro Ribeirão Preto: Simone Kashima, Evandra Strazza Rodrigues, Svetoslav Nanev Slavov, Elaine Vieira dos Santos, Rafael dos Santos Bezerra, Luiz Carlos Junior de Alcantara, Marta Giovanetti, Vagner Fonseca, Flavia Aburjaile, Rodrigo Tocantins Calado. FAMERP-SJRP: Cecília Artico Banho, Lívia Sacchetto, Fábio Sossai Possebon, Leila Sabrina Ullmann, Cíntia Bittar, Guilherme Campos, Helena Lage Ferreira, Jorge A. Petroli Marchesi, Maisa C. Pereira Parra, Marília Moraes, Paula Rahal, Paulo Inacio da Costa, João Pessoa Araújo Jr., Maurício Lacerda Nogueira. Prefeitura de Sao Paulo: Melissa Palmieri. |
| EPI_ISL_1966164, EPI_ISL_1966165, EPI_ISL_1966166 | UNIDADE RESPIRATORIA NOVA HORTOLANDIA | Instituto Butantan / Mendelics | Instituto Butantan: Dimas Tadeu Covas, Sandra Coccuzzo Sampaio, Maria Carolina Elias, José Salvatore Leister Patané, Vincent Louis Viala, Antonio Jorge Martins, Ricardo Haddad, Claudia Renata dos Santos Barros, Elaine Cristina Marqueze, Raul Machado Neto, Debora Botequio Moretti, Jardelina de Souza Todao Bernardino, Loyze Paola Oliveira de Lima, Luiz Aurelio de Campos Crispin. Centro de Genômica Funcional da ESALQ: Luiz Lehmann Coutinho, Ricardo Augusto Brassaloti, Raquel de Lello Rocha Campos Cassano. NGS Soluções Genômicas: Pilar Drummond Sampaio Corrêa Mariani. FZEA-USP Pirassununga: Mirele Daiana Poleti, Jessika Cristina Chagas Lesbon, Elisangela Chicaroni Mattos, Heidge Fukumasu. USP-Botucatu: Rejane Maria Tommasini Grotto, Jayme A. Souza-Neto, Guilherme Targino Valente, Patricia Akemi Assato, Felipe Allan da Silva da Costa, Bianca Cechetto Carlos. Mendelics: Bibiana Santos, João Paulo Kitajima, Erika Freitas, David Schlesinger. Hemocentro Ribeirão Preto: Simone Kashima, Evandra Strazza Rodrigues, Svetoslav Nanev Slavov, Elaine Vieira dos Santos, Rafael dos Santos Bezerra, Luiz Carlos Junior de Alcantara, Marta Giovanetti, Vagner Fonseca, Flavia Aburjaile, Rodrigo Tocantins Calado. FAMERP-SJRP: Cecília Artico Banho, Lívia Sacchetto, Fábio Sossai Possebon, Leila Sabrina Ullmann, Cíntia Bittar, Guilherme Campos, Helena Lage Ferreira, Jorge A. Petroli Marchesi, Maisa C. Pereira Parra, Marília Moraes, Paula Rahal, Paulo Inacio da Costa, João Pessoa Araújo Jr., Maurício Lacerda Nogueira. Prefeitura de Sao Paulo: Melissa Palmieri. |
| EPI_ISL_1966167, EPI_ISL_1966168, EPI_ISL_1966169, EPI_ISL_1966170, EPI_ISL_1966171, EPI_ISL_1966173 | UNIDADE DE PRONTO ATENDIMENTO UPA DRA ANA OLIVIA BENTIVOGLIO | Instituto Butantan / Mendelics | Instituto Butantan: Dimas Tadeu Covas, Sandra Coccuzzo Sampaio, Maria Carolina Elias, José Salvatore Leister Patané, Vincent Louis Viala, Antonio Jorge Martins, Ricardo Haddad, Claudia Renata dos Santos Barros, Elaine Cristina Marqueze, Raul Machado Neto, Debora Botequio Moretti, Jardelina de Souza Todao Bernardino, Loyze Paola Oliveira de Lima, Luiz Aurelio de Campos Crispin. Centro de Genômica Funcional da ESALQ: Luiz Lehmann Coutinho, Ricardo Augusto Brassaloti, Raquel de Lello Rocha Campos Cassano. NGS Soluções Genômicas: Pilar Drummond Sampaio Corrêa Mariani. FZEA-USP Pirassununga: Mirele Daiana Poleti, Jessika Cristina Chagas Lesbon, Elisangela Chicaroni Mattos, Heidge Fukumasu. USP-Botucatu: Rejane Maria Tommasini Grotto, Jayme A. Souza-Neto, Guilherme Targino Valente, Patricia Akemi Assato, Felipe Allan da Silva da Costa, Bianca Cechetto Carlos. Mendelics: Bibiana Santos, João Paulo Kitajima, Erika Freitas, David Schlesinger. Hemocentro Ribeirão Preto: Simone Kashima, Evandra Strazza Rodrigues, Svetoslav Nanev Slavov, Elaine Vieira dos Santos, Rafael dos Santos Bezerra, Luiz Carlos Junior de Alcantara, Marta Giovanetti, Vagner Fonseca, Flavia Aburjaile, Rodrigo Tocantins Calado. FAMERP-SJRP: Cecília Artico Banho, Lívia Sacchetto, Fábio Sossai Possebon, Leila Sabrina |

| | | | |
|---|---|---|---|
| | | | Ullmann, Cíntia Bittar, Guilherme Campos, Helena Lage Ferreira, Jorge A. Petroli Marchesi, Maisa C. Pereira Parra, Marília Moraes, Paula Rahal, Paulo Inacio da Costa, João Pessoa Araújo Jr., Maurício Lacerda Nogueira. Prefeitura de Sao Paulo: Melissa Palmieri. |
| EPI_ISL_1966176 | PRONTO ATENDIMENTO DA PONTE DE SAO JOAO | Instituto Butantan / Mendelics | Instituto Butantan: Dimas Tadeu Covas, Sandra Coccuzzo Sampaio, Maria Carolina Elias, José Salvatore Leister Patané, Vincent Louis Viala, Antonio Jorge Martins, Ricardo Haddad, Claudia Renata dos Santos Barros, Elaine Cristina Marqueze, Raul Machado Neto, Debora Botequio Moretti, Jardelina de Souza Todao Bernardino, Loyze Paola Oliveira de Lima, Luiz Aurelio de Campos Crispin. Centro de Genômica Funcional da ESALQ: Luiz Lehmann Coutinho, Ricardo Augusto Brassaloti, Raquel de Lello Rocha Campos Cassano. NGS Soluções Genômicas: Pilar Drummond Sampaio Corrêa Mariani. FZEA-USP Pirassununga: Mirele Daiana Poleti, Jessika Cristina Chagas Lesbon, Elisangela Chicaroni Mattos, Heidge Fukumasu. USP-Botucatu: Rejane Maria Tommasini Grotto, Jayme A. Souza-Neto, Guilherme Targino Valente, Patricia Akemi Assato, Felipe Allan da Silva da Costa, Bianca Cechetto Carlos. Mendelics: Bibiana Santos, João Paulo Kitajima, Erika Freitas, David Schlesinger. Hemocentro Ribeirão Preto: Simone Kashima, Evandra Strazza Rodrigues, Svetoslav Nanev Slavov, Elaine Vieira dos Santos, Rafael dos Santos Bezerra, Luiz Carlos Junior de Alcantara, Marta Giovanetti, Vagner Fonseca, Flavia Aburjaile, Rodrigo Tocantins Calado. FAMERP-SJRP: Cecília Artico Banho, Lívia Sacchetto, Fábio Sossai Possebon, Leila Sabrina Ullmann, Cíntia Bittar, Guilherme Campos, Helena Lage Ferreira, Jorge A. Petroli Marchesi, Maisa C. Pereira Parra, Marília Moraes, Paula Rahal, Paulo Inacio da Costa, João Pessoa Araújo Jr., Maurício Lacerda Nogueira. Prefeitura de Sao Paulo: Melissa Palmieri. |
| EPI_ISL_1966177 | SECRETARIA MUNICIPAL DE SAUDE SOROCABA | Instituto Butantan / Mendelics | Instituto Butantan: Dimas Tadeu Covas, Sandra Coccuzzo Sampaio, Maria Carolina Elias, José Salvatore Leister Patané, Vincent Louis Viala, Antonio Jorge Martins, Ricardo Haddad, Claudia Renata dos Santos Barros, Elaine Cristina Marqueze, Raul Machado Neto, Debora Botequio Moretti, Jardelina de Souza Todao Bernardino, Loyze Paola Oliveira de Lima, Luiz Aurelio de Campos Crispin. Centro de Genômica Funcional da ESALQ: Luiz Lehmann Coutinho, Ricardo Augusto Brassaloti, Raquel de Lello Rocha Campos Cassano. NGS Soluções Genômicas: Pilar Drummond Sampaio Corrêa Mariani. FZEA-USP Pirassununga: Mirele Daiana Poleti, Jessika Cristina Chagas Lesbon, Elisangela Chicaroni Mattos, Heidge Fukumasu. USP-Botucatu: Rejane Maria Tommasini Grotto, Jayme A. Souza-Neto, Guilherme Targino Valente, Patricia Akemi Assato, Felipe Allan da Silva da Costa, Bianca Cechetto Carlos. Mendelics: Bibiana Santos, João Paulo Kitajima, Erika Freitas, David Schlesinger. Hemocentro Ribeirão Preto: Simone Kashima, Evandra Strazza Rodrigues, Svetoslav Nanev Slavov, Elaine Vieira dos Santos, Rafael dos Santos Bezerra, Luiz Carlos Junior de Alcantara, Marta Giovanetti, Vagner Fonseca, Flavia Aburjaile, Rodrigo Tocantins Calado. FAMERP-SJRP: Cecília Artico Banho, Lívia Sacchetto, Fábio Sossai Possebon, Leila Sabrina Ullmann, Cíntia Bittar, Guilherme Campos, Helena Lage Ferreira, Jorge A. Petroli Marchesi, Maisa C. Pereira Parra, Marília Moraes, Paula Rahal, Paulo Inacio da Costa, João Pessoa Araújo Jr., Maurício Lacerda Nogueira. Prefeitura de Sao Paulo: Melissa Palmieri. |
| EPI_ISL_1966179 | PRONTO ATENDIMENTO VILA PADRE ANCHIETA | Instituto Butantan / Mendelics | Instituto Butantan: Dimas Tadeu Covas, Sandra Coccuzzo Sampaio, Maria Carolina Elias, José Salvatore Leister Patané, Vincent Louis Viala, Antonio Jorge Martins, Ricardo Haddad, Claudia Renata dos Santos Barros, Elaine Cristina Marqueze, Raul Machado Neto, Debora Botequio Moretti, Jardelina de Souza Todao Bernardino, Loyze Paola Oliveira de Lima, Luiz Aurelio de Campos Crispin. Centro de Genômica Funcional da ESALQ: Luiz Lehmann Coutinho, Ricardo Augusto Brassaloti, Raquel de Lello Rocha Campos Cassano. NGS Soluções Genômicas: Pilar Drummond Sampaio Corrêa Mariani. FZEA-USP Pirassununga: Mirele Daiana Poleti, Jessika Cristina Chagas Lesbon, Elisangela Chicaroni Mattos, Heidge Fukumasu. USP-Botucatu: Rejane Maria Tommasini Grotto, Jayme A. Souza-Neto, Guilherme Targino Valente, Patricia Akemi Assato, Felipe Allan da Silva da Costa, Bianca Cechetto Carlos. Mendelics: Bibiana Santos, João Paulo Kitajima, Erika Freitas, David Schlesinger. Hemocentro Ribeirão Preto: Simone Kashima, Evandra Strazza Rodrigues, Svetoslav Nanev Slavov, Elaine Vieira dos Santos, Rafael dos Santos Bezerra, Luiz Carlos Junior de Alcantara, Marta Giovanetti, Vagner Fonseca, Flavia Aburjaile, Rodrigo Tocantins Calado. FAMERP-SJRP: Cecília Artico Banho, Lívia Sacchetto, Fábio Sossai Possebon, Leila Sabrina Ullmann, Cíntia Bittar, Guilherme Campos, Helena Lage Ferreira, Jorge A. Petroli Marchesi, Maisa C. Pereira Parra, Marília Moraes, Paula Rahal, Paulo Inacio da Costa, João Pessoa Araújo Jr., Maurício Lacerda Nogueira. Prefeitura de Sao Paulo: Melissa Palmieri. |
| EPI_ISL_1966181, EPI_ISL_1966182, EPI_ISL_1966183, EPI_ISL_1966184, EPI_ISL_1966185, EPI_ISL_1966186 | SERV DE VIG SANITARIA EPIDEMIO E CTRL DE ZOONOZES GUARUJA | Instituto Butantan / Mendelics | Instituto Butantan: Dimas Tadeu Covas, Sandra Coccuzzo Sampaio, Maria Carolina Elias, José Salvatore Leister Patané, Vincent Louis Viala, Antonio Jorge Martins, Ricardo Haddad, Claudia Renata dos Santos Barros, Elaine Cristina Marqueze, Raul Machado Neto, Debora Botequio Moretti, Jardelina de Souza Todao Bernardino, Loyze Paola Oliveira de Lima, Luiz Aurelio de Campos Crispin. Centro de Genômica Funcional da ESALQ: Luiz Lehmann Coutinho, Ricardo Augusto Brassaloti, Raquel de Lello Rocha Campos Cassano. NGS Soluções Genômicas: Pilar Drummond Sampaio Corrêa Mariani. FZEA-USP Pirassununga: Mirele Daiana Poleti, Jessika Cristina Chagas Lesbon, Elisangela Chicaroni Mattos, Heidge Fukumasu. USP-Botucatu: Rejane Maria Tommasini Grotto, Jayme A. Souza-Neto, Guilherme Targino Valente, Patricia Akemi Assato, Felipe Allan da Silva da Costa, Bianca Cechetto Carlos. Mendelics: Bibiana Santos, João Paulo Kitajima, Erika Freitas, David Schlesinger. Hemocentro Ribeirão Preto: Simone Kashima, Evandra Strazza Rodrigues, Svetoslav Nanev Slavov, Elaine Vieira dos Santos, Rafael dos Santos Bezerra, Luiz Carlos Junior de Alcantara, Marta Giovanetti, Vagner Fonseca, Flavia Aburjaile, Rodrigo Tocantins Calado. FAMERP-SJRP: Cecília Artico Banho, Lívia Sacchetto, Fábio Sossai Possebon, Leila Sabrina Ullmann, Cíntia Bittar, Guilherme Campos, Helena Lage Ferreira, Jorge A. Petroli Marchesi, Maisa C. Pereira Parra, Marília Moraes, Paula Rahal, Paulo Inacio da Costa, João Pessoa Araújo Jr., Maurício Lacerda Nogueira. Prefeitura de Sao Paulo: Melissa Palmieri. |
| EPI_ISL_1966187 | SMS IPERO | Instituto Butantan / Mendelics | Instituto Butantan: Dimas Tadeu Covas, Sandra Coccuzzo Sampaio, Maria Carolina Elias, José Salvatore Leister Patané, Vincent Louis Viala, Antonio Jorge Martins, Ricardo Haddad, Claudia Renata dos Santos Barros, Elaine Cristina Marqueze, Raul Machado Neto, Debora Botequio Moretti, Jardelina de Souza Todao Bernardino, Loyze Paola Oliveira de Lima, Luiz Aurelio de Campos Crispin. Centro de Genômica Funcional da ESALQ: Luiz Lehmann Coutinho, Ricardo Augusto Brassaloti, Raquel de Lello Rocha Campos Cassano. NGS Soluções Genômicas: Pilar Drummond Sampaio Corrêa Mariani. FZEA-USP Pirassununga: Mirele Daiana Poleti, Jessika Cristina Chagas Lesbon, Elisangela Chicaroni Mattos, Heidge Fukumasu. USP-Botucatu: Rejane Maria Tommasini Grotto, Jayme A. Souza-Neto, Guilherme Targino Valente, Patricia Akemi Assato, Felipe Allan da Silva da Costa, Bianca Cechetto Carlos. Mendelics: Bibiana Santos, João Paulo Kitajima, Erika Freitas, David Schlesinger. Hemocentro Ribeirão Preto: Simone Kashima, Evandra Strazza Rodrigues, Svetoslav Nanev Slavov, Elaine Vieira dos Santos, Rafael dos Santos Bezerra, Luiz Carlos Junior de Alcantara, Marta Giovanetti, Vagner Fonseca, Flavia Aburjaile, Rodrigo Tocantins Calado. FAMERP-SJRP: Cecília Artico Banho, Lívia Sacchetto, Fábio Sossai Possebon, Leila Sabrina Ullmann, Cíntia Bittar, Guilherme Campos, Helena Lage Ferreira, Jorge A. Petroli Marchesi, Maisa C. Pereira Parra, Marília Moraes, Paula Rahal, Paulo Inacio da Costa, João Pessoa Araújo Jr., Maurício Lacerda Nogueira. Prefeitura de Sao Paulo: Melissa Palmieri. |
| EPI_ISL_1966188 | UBS SALTO DE SAO JOSE | Instituto Butantan / Mendelics | Instituto Butantan: Dimas Tadeu Covas, Sandra Coccuzzo Sampaio, Maria Carolina Elias, José Salvatore Leister Patané, Vincent Louis Viala, Antonio Jorge Martins, Ricardo Haddad, Claudia Renata dos Santos Barros, Elaine Cristina Marqueze, Raul Machado Neto, Debora Botequio Moretti, Jardelina de Souza Todao Bernardino, Loyze Paola Oliveira de Lima, Luiz Aurelio de Campos Crispin. Centro de Genômica Funcional da ESALQ: Luiz Lehmann Coutinho, Ricardo Augusto Brassaloti, Raquel de Lello Rocha Campos Cassano. NGS Soluções Genômicas: Pilar Drummond Sampaio Corrêa Mariani. FZEA-USP Pirassununga: Mirele Daiana Poleti, Jessika Cristina Chagas Lesbon, Elisangela Chicaroni Mattos, Heidge Fukumasu. USP-Botucatu: Rejane Maria Tommasini Grotto, Jayme A. Souza-Neto, Guilherme Targino Valente, Patricia Akemi Assato, Felipe Allan da Silva da Costa, Bianca Cechetto Carlos. Mendelics: Bibiana Santos, João Paulo Kitajima, Erika Freitas, David Schlesinger. Hemocentro Ribeirão Preto: Simone Kashima, Evandra Strazza Rodrigues, Svetoslav Nanev Slavov, Elaine Vieira dos Santos, Rafael dos Santos Bezerra, Luiz Carlos Junior de Alcantara, Marta Giovanetti, Vagner Fonseca, Flavia Aburjaile, Rodrigo Tocantins Calado. FAMERP-SJRP: Cecília Artico Banho, Lívia Sacchetto, Fábio Sossai Possebon, Leila Sabrina Ullmann, Cíntia Bittar, Guilherme Campos, Helena Lage Ferreira, Jorge A. Petroli Marchesi, Maisa C. Pereira Parra, Marília Moraes, Paula Rahal, Paulo Inacio da Costa, João Pessoa Araújo Jr., Maurício Lacerda Nogueira. Prefeitura de Sao Paulo: Melissa Palmieri. |
| EPI_ISL_1966189, EPI_ISL_1966190, EPI_ISL_1966191, EPI_ISL_1966192, EPI_ISL_1966195, EPI_ISL_1966196 | SECAO CENTRO DE DIAGNOSTICO SECEDI | Instituto Butantan / Mendelics | Instituto Butantan: Dimas Tadeu Covas, Sandra Coccuzzo Sampaio, Maria Carolina Elias, José Salvatore Leister Patané, Vincent Louis Viala, Antonio Jorge Martins, Ricardo Haddad, Claudia Renata dos Santos Barros, Elaine Cristina Marqueze, Raul Machado Neto, Debora Botequio Moretti, Jardelina de Souza Todao Bernardino, Loyze Paola Oliveira de Lima, Luiz Aurelio de Campos Crispin. Centro de Genômica Funcional da ESALQ: Luiz Lehmann Coutinho, Ricardo Augusto Brassaloti, Raquel de Lello Rocha Campos Cassano. NGS Soluções Genômicas: Pilar Drummond Sampaio Corrêa Mariani. FZEA-USP Pirassununga: Mirele Daiana Poleti, Jessika Cristina Chagas Lesbon, Elisangela Chicaroni Mattos, Heidge Fukumasu. USP-Botucatu: Rejane Maria Tommasini Grotto, Jayme A. Souza-Neto, Guilherme Targino Valente, Patricia Akemi Assato, Felipe Allan da Silva da Costa, Bianca Cechetto Carlos. Mendelics: Bibiana Santos, João Paulo Kitajima, Erika Freitas, David Schlesinger. Hemocentro Ribeirão Preto: Simone Kashima, Evandra Strazza Rodrigues, Svetoslav Nanev Slavov, Elaine Vieira dos Santos, Rafael dos Santos Bezerra, Luiz Carlos Junior de Alcantara, Marta Giovanetti, Vagner Fonseca, Flavia Aburjaile, Rodrigo Tocantins Calado. FAMERP-SJRP: Cecília Artico Banho, Lívia Sacchetto, Fábio Sossai Possebon, Leila Sabrina Ullmann, Cíntia Bittar, Guilherme Campos, Helena Lage Ferreira, Jorge A. Petroli Marchesi, Maisa C. Pereira Parra, Marília Moraes, Paula Rahal, Paulo Inacio da Costa, João Pessoa Araújo Jr., Maurício Lacerda Nogueira. Prefeitura de Sao Paulo: Melissa Palmieri. |
| EPI_ISL_1966198, EPI_ISL_1966199, EPI_ISL_1966200 | SECRETARIA MUNICIPAL DE SAUDE SOROCABA | Instituto Butantan / Mendelics | Instituto Butantan: Dimas Tadeu Covas, Sandra Coccuzzo Sampaio, Maria Carolina Elias, José Salvatore Leister Patané, Vincent Louis Viala, Antonio Jorge Martins, Ricardo Haddad, Claudia Renata dos Santos Barros, Elaine Cristina Marqueze, Raul Machado Neto, Debora Botequio Moretti, Jardelina de Souza Todao Bernardino, Loyze Paola Oliveira de Lima, Luiz Aurelio de Campos Crispin. Centro de Genômica Funcional da ESALQ: Luiz Lehmann Coutinho, Ricardo Augusto Brassaloti, Raquel de Lello Rocha Campos Cassano. NGS Soluções Genômicas: Pilar Drummond Sampaio Corrêa Mariani. |

| | | | |
|---|---|---|---|
| | | | FZEA-USP Pirassununga: Mirele Daiana Poleti, Jessika Cristina Chagas Lesbon, Elisangela Chicaroni Mattos, Heidge Fukumasu. USP-Botucatu: Rejane Maria Tommasini Grotto, Jayme A. Souza-Neto, Guilherme Targino Valente, Patricia Akemi Assato, Felipe Allan da Silva da Costa, Bianca Cechetto Carlos. Mendelics: Bibiana Santos, João Paulo Kitajima, Erika Freitas, David Schlesinger. Hemocentro Ribeirão Preto: Simone Kashima, Evandra Strazza Rodrigues, Svetoslav Nanev Slavov, Elaine Vieira dos Santos, Rafael dos Santos Bezerra, Luiz Carlos Junior de Alcantara, Marta Giovanetti, Vagner Fonseca, Flavia Aburjaile, Rodrigo Tocantins Calado. FAMERP-SJRP: Cecília Artico Banho, Lívia Sacchetto, Fábio Sossai Possebon, Leila Sabrina Ullmann, Cíntia Bittar, Guilherme Campos, Helena Lage Ferreira, Jorge A. Petroli Marchesi, Maisa C. Pereira Parra, Marília Moraes, Paula Rahal, Paulo Inacio da Costa, João Pessoa Araújo Jr., Maurício Lacerda Nogueira. Prefeitura de Sao Paulo: Melissa Palmieri. |
| EPI_ISL_1966201 | UNIDADE BASICA DE SAUDE DA FAMILIA BILAC | Instituto Butantan / Mendelics | Instituto Butantan: Dimas Tadeu Covas, Sandra Coccuzzo Sampaio, Maria Carolina Elias, José Salvatore Leister Patané, Vincent Louis Viala, Antonio Jorge Martins, Ricardo Haddad, Claudia Renata dos Santos Barros, Elaine Cristina Marqueze, Raul Machado Neto, Debora Botequio Moretti, Jardelina de Souza Todao Bernardino, Loyze Paola Oliveira de Lima, Luiz Aurelio de Campos Crispin. Centro de Genômica Funcional da ESALQ: Luiz Lehmann Coutinho, Ricardo Augusto Brassaloti, Raquel de Lello Rocha Campos Cassano. NGS Soluções Genômicas: Pilar Drummond Sampaio Corrêa Mariani. FZEA-USP Pirassununga: Mirele Daiana Poleti, Jessika Cristina Chagas Lesbon, Elisangela Chicaroni Mattos, Heidge Fukumasu. USP-Botucatu: Rejane Maria Tommasini Grotto, Jayme A. Souza-Neto, Guilherme Targino Valente, Patricia Akemi Assato, Felipe Allan da Silva da Costa, Bianca Cechetto Carlos. Mendelics: Bibiana Santos, João Paulo Kitajima, Erika Freitas, David Schlesinger. Hemocentro Ribeirão Preto: Simone Kashima, Evandra Strazza Rodrigues, Svetoslav Nanev Slavov, Elaine Vieira dos Santos, Rafael dos Santos Bezerra, Luiz Carlos Junior de Alcantara, Marta Giovanetti, Vagner Fonseca, Flavia Aburjaile, Rodrigo Tocantins Calado. FAMERP-SJRP: Cecília Artico Banho, Lívia Sacchetto, Fábio Sossai Possebon, Leila Sabrina Ullmann, Cíntia Bittar, Guilherme Campos, Helena Lage Ferreira, Jorge A. Petroli Marchesi, Maisa C. Pereira Parra, Marília Moraes, Paula Rahal, Paulo Inacio da Costa, João Pessoa Araújo Jr., Maurício Lacerda Nogueira. Prefeitura de Sao Paulo: Melissa Palmieri. |
| EPI_ISL_1966202 | UBS JOSE SABINO FERREIRA | Instituto Butantan / Mendelics | Instituto Butantan: Dimas Tadeu Covas, Sandra Coccuzzo Sampaio, Maria Carolina Elias, José Salvatore Leister Patané, Vincent Louis Viala, Antonio Jorge Martins, Ricardo Haddad, Claudia Renata dos Santos Barros, Elaine Cristina Marqueze, Raul Machado Neto, Debora Botequio Moretti, Jardelina de Souza Todao Bernardino, Loyze Paola Oliveira de Lima, Luiz Aurelio de Campos Crispin. Centro de Genômica Funcional da ESALQ: Luiz Lehmann Coutinho, Ricardo Augusto Brassaloti, Raquel de Lello Rocha Campos Cassano. NGS Soluções Genômicas: Pilar Drummond Sampaio Corrêa Mariani. FZEA-USP Pirassununga: Mirele Daiana Poleti, Jessika Cristina Chagas Lesbon, Elisangela Chicaroni Mattos, Heidge Fukumasu. USP-Botucatu: Rejane Maria Tommasini Grotto, Jayme A. Souza-Neto, Guilherme Targino Valente, Patricia Akemi Assato, Felipe Allan da Silva da Costa, Bianca Cechetto Carlos. Mendelics: Bibiana Santos, João Paulo Kitajima, Erika Freitas, David Schlesinger. Hemocentro Ribeirão Preto: Simone Kashima, Evandra Strazza Rodrigues, Svetoslav Nanev Slavov, Elaine Vieira dos Santos, Rafael dos Santos Bezerra, Luiz Carlos Junior de Alcantara, Marta Giovanetti, Vagner Fonseca, Flavia Aburjaile, Rodrigo Tocantins Calado. FAMERP-SJRP: Cecília Artico Banho, Lívia Sacchetto, Fábio Sossai Possebon, Leila Sabrina Ullmann, Cíntia Bittar, Guilherme Campos, Helena Lage Ferreira, Jorge A. Petroli Marchesi, Maisa C. Pereira Parra, Marília Moraes, Paula Rahal, Paulo Inacio da Costa, João Pessoa Araújo Jr., Maurício Lacerda Nogueira. Prefeitura de Sao Paulo: Melissa Palmieri. |
| EPI_ISL_1966203 | UBS IRMA AGUEDA MARIA JAIME | Instituto Butantan / Mendelics | Instituto Butantan: Dimas Tadeu Covas, Sandra Coccuzzo Sampaio, Maria Carolina Elias, José Salvatore Leister Patané, Vincent Louis Viala, Antonio Jorge Martins, Ricardo Haddad, Claudia Renata dos Santos Barros, Elaine Cristina Marqueze, Raul Machado Neto, Debora Botequio Moretti, Jardelina de Souza Todao Bernardino, Loyze Paola Oliveira de Lima, Luiz Aurelio de Campos Crispin. Centro de Genômica Funcional da ESALQ: Luiz Lehmann Coutinho, Ricardo Augusto Brassaloti, Raquel de Lello Rocha Campos Cassano. NGS Soluções Genômicas: Pilar Drummond Sampaio Corrêa Mariani. FZEA-USP Pirassununga: Mirele Daiana Poleti, Jessika Cristina Chagas Lesbon, Elisangela Chicaroni Mattos, Heidge Fukumasu. USP-Botucatu: Rejane Maria Tommasini Grotto, Jayme A. Souza-Neto, Guilherme Targino Valente, Patricia Akemi Assato, Felipe Allan da Silva da Costa, Bianca Cechetto Carlos. Mendelics: Bibiana Santos, João Paulo Kitajima, Erika Freitas, David Schlesinger. Hemocentro Ribeirão Preto: Simone Kashima, Evandra Strazza Rodrigues, Svetoslav Nanev Slavov, Elaine Vieira dos Santos, Rafael dos Santos Bezerra, Luiz Carlos Junior de Alcantara, Marta Giovanetti, Vagner Fonseca, Flavia Aburjaile, Rodrigo Tocantins Calado. FAMERP-SJRP: Cecília Artico Banho, Lívia Sacchetto, Fábio Sossai Possebon, Leila Sabrina Ullmann, Cíntia Bittar, Guilherme Campos, Helena Lage Ferreira, Jorge A. Petroli Marchesi, Maisa C. Pereira Parra, Marília Moraes, Paula Rahal, Paulo Inacio da Costa, João Pessoa Araújo Jr., Maurício Lacerda Nogueira. Prefeitura de Sao Paulo: Melissa Palmieri. |
| EPI_ISL_1966204 | UBS JOSE HILARIO DOS SANTOS | Instituto Butantan / Mendelics | Instituto Butantan: Dimas Tadeu Covas, Sandra Coccuzzo Sampaio, Maria Carolina Elias, José Salvatore Leister Patané, Vincent Louis Viala, Antonio Jorge Martins, Ricardo Haddad, Claudia Renata dos Santos Barros, Elaine Cristina Marqueze, Raul Machado Neto, Debora Botequio Moretti, Jardelina de Souza Todao Bernardino, Loyze Paola Oliveira de Lima, Luiz Aurelio de Campos Crispin. Centro de Genômica Funcional da ESALQ: Luiz Lehmann Coutinho, Ricardo Augusto Brassaloti, Raquel de Lello Rocha Campos Cassano. NGS Soluções Genômicas: Pilar Drummond Sampaio Corrêa Mariani. FZEA-USP Pirassununga: Mirele Daiana Poleti, Jessika Cristina Chagas Lesbon, Elisangela Chicaroni Mattos, Heidge Fukumasu. USP-Botucatu: Rejane Maria Tommasini Grotto, Jayme A. Souza-Neto, Guilherme Targino Valente, Patricia Akemi Assato, Felipe Allan da Silva da Costa, Bianca Cechetto Carlos. Mendelics: Bibiana Santos, João Paulo Kitajima, Erika Freitas, David Schlesinger. Hemocentro Ribeirão Preto: Simone Kashima, Evandra Strazza Rodrigues, Svetoslav Nanev Slavov, Elaine Vieira dos Santos, Rafael dos Santos Bezerra, Luiz Carlos Junior de Alcantara, Marta Giovanetti, Vagner Fonseca, Flavia Aburjaile, Rodrigo Tocantins Calado. FAMERP-SJRP: Cecília Artico Banho, Lívia Sacchetto, Fábio Sossai Possebon, Leila Sabrina Ullmann, Cíntia Bittar, Guilherme Campos, Helena Lage Ferreira, Jorge A. Petroli Marchesi, Maisa C. Pereira Parra, Marília Moraes, Paula Rahal, Paulo Inacio da Costa, João Pessoa Araújo Jr., Maurício Lacerda Nogueira. Prefeitura de Sao Paulo: Melissa Palmieri. |
| EPI_ISL_1966205 | UBS DARCY ALVES E ROBALINHO | Instituto Butantan / Mendelics | Instituto Butantan: Dimas Tadeu Covas, Sandra Coccuzzo Sampaio, Maria Carolina Elias, José Salvatore Leister Patané, Vincent Louis Viala, Antonio Jorge Martins, Ricardo Haddad, Claudia Renata dos Santos Barros, Elaine Cristina Marqueze, Raul Machado Neto, Debora Botequio Moretti, Jardelina de Souza Todao Bernardino, Loyze Paola Oliveira de Lima, Luiz Aurelio de Campos Crispin. Centro de Genômica Funcional da ESALQ: Luiz Lehmann Coutinho, Ricardo Augusto Brassaloti, Raquel de Lello Rocha Campos Cassano. NGS Soluções Genômicas: Pilar Drummond Sampaio Corrêa Mariani. FZEA-USP Pirassununga: Mirele Daiana Poleti, Jessika Cristina Chagas Lesbon, Elisangela Chicaroni Mattos, Heidge Fukumasu. USP-Botucatu: Rejane Maria Tommasini Grotto, Jayme A. Souza-Neto, Guilherme Targino Valente, Patricia Akemi Assato, Felipe Allan da Silva da Costa, Bianca Cechetto Carlos. Mendelics: Bibiana Santos, João Paulo Kitajima, Erika Freitas, David Schlesinger. Hemocentro Ribeirão Preto: Simone Kashima, Evandra Strazza Rodrigues, Svetoslav Nanev Slavov, Elaine Vieira dos Santos, Rafael dos Santos Bezerra, Luiz Carlos Junior de Alcantara, Marta Giovanetti, Vagner Fonseca, Flavia Aburjaile, Rodrigo Tocantins Calado. FAMERP-SJRP: Cecília Artico Banho, Lívia Sacchetto, Fábio Sossai Possebon, Leila Sabrina Ullmann, Cíntia Bittar, Guilherme Campos, Helena Lage Ferreira, Jorge A. Petroli Marchesi, Maisa C. Pereira Parra, Marília Moraes, Paula Rahal, Paulo Inacio da Costa, João Pessoa Araújo Jr., Maurício Lacerda Nogueira. Prefeitura de Sao Paulo: Melissa Palmieri. |
| EPI_ISL_1966206 | UBS OTACILIO FIRMINO LOPES | Instituto Butantan / Mendelics | Instituto Butantan: Dimas Tadeu Covas, Sandra Coccuzzo Sampaio, Maria Carolina Elias, José Salvatore Leister Patané, Vincent Louis Viala, Antonio Jorge Martins, Ricardo Haddad, Claudia Renata dos Santos Barros, Elaine Cristina Marqueze, Raul Machado Neto, Debora Botequio Moretti, Jardelina de Souza Todao Bernardino, Loyze Paola Oliveira de Lima, Luiz Aurelio de Campos Crispin. Centro de Genômica Funcional da ESALQ: Luiz Lehmann Coutinho, Ricardo Augusto Brassaloti, Raquel de Lello Rocha Campos Cassano. NGS Soluções Genômicas: Pilar Drummond Sampaio Corrêa Mariani. FZEA-USP Pirassununga: Mirele Daiana Poleti, Jessika Cristina Chagas Lesbon, Elisangela Chicaroni Mattos, Heidge Fukumasu. USP-Botucatu: Rejane Maria Tommasini Grotto, Jayme A. Souza-Neto, Guilherme Targino Valente, Patricia Akemi Assato, Felipe Allan da Silva da Costa, Bianca Cechetto Carlos. Mendelics: Bibiana Santos, João Paulo Kitajima, Erika Freitas, David Schlesinger. Hemocentro Ribeirão Preto: Simone Kashima, Evandra Strazza Rodrigues, Svetoslav Nanev Slavov, Elaine Vieira dos Santos, Rafael dos Santos Bezerra, Luiz Carlos Junior de Alcantara, Marta Giovanetti, Vagner Fonseca, Flavia Aburjaile, Rodrigo Tocantins Calado. FAMERP-SJRP: Cecília Artico Banho, Lívia Sacchetto, Fábio Sossai Possebon, Leila Sabrina Ullmann, Cíntia Bittar, Guilherme Campos, Helena Lage Ferreira, Jorge A. Petroli Marchesi, Maisa C. Pereira Parra, Marília Moraes, Paula Rahal, Paulo Inacio da Costa, João Pessoa Araújo Jr., Maurício Lacerda Nogueira. Prefeitura de Sao Paulo: Melissa Palmieri. |
| EPI_ISL_1966207 | UBS FRANCISCA LIMA DE LIRA | Instituto Butantan / Mendelics | Instituto Butantan: Dimas Tadeu Covas, Sandra Coccuzzo Sampaio, Maria Carolina Elias, José Salvatore Leister Patané, Vincent Louis Viala, Antonio Jorge Martins, Ricardo Haddad, Claudia Renata dos Santos Barros, Elaine Cristina Marqueze, Raul Machado Neto, Debora Botequio Moretti, Jardelina de Souza Todao Bernardino, Loyze Paola Oliveira de Lima, Luiz Aurelio de Campos Crispin. Centro de Genômica Funcional da ESALQ: Luiz Lehmann Coutinho, Ricardo Augusto Brassaloti, Raquel de Lello Rocha Campos Cassano. NGS Soluções Genômicas: Pilar Drummond Sampaio Corrêa Mariani. FZEA-USP Pirassununga: Mirele Daiana Poleti, Jessika Cristina Chagas Lesbon, Elisangela Chicaroni Mattos, Heidge Fukumasu. USP-Botucatu: Rejane Maria Tommasini Grotto, Jayme A. Souza-Neto, Guilherme Targino Valente, Patricia Akemi Assato, Felipe Allan da Silva da Costa, Bianca Cechetto Carlos. Mendelics: Bibiana Santos, João Paulo Kitajima, Erika Freitas, David Schlesinger. Hemocentro Ribeirão Preto: Simone Kashima, Evandra Strazza Rodrigues, Svetoslav Nanev Slavov, Elaine Vieira dos Santos, Rafael dos Santos Bezerra, Luiz Carlos Junior de Alcantara, Marta Giovanetti, Vagner Fonseca, Flavia Aburjaile, Rodrigo Tocantins Calado. FAMERP-SJRP: Cecília Artico Banho, Lívia Sacchetto, Fábio Sossai Possebon, Leila Sabrina Ullmann, Cíntia Bittar, Guilherme Campos, Helena Lage Ferreira, Jorge A. Petroli Marchesi, Maisa C. Pereira Parra, Marília Moraes, Paula Rahal, Paulo Inacio da Costa, João Pessoa Araújo Jr., Maurício Lacerda Nogueira. Prefeitura de Sao Paulo: Melissa Palmieri. |

| | | | |
|---|---|---|---|
| EPI_ISL_1966208 | UBS OTACILIO FIRMINO LOPES | Instituto Butantan / Mendelics | Instituto Butantan: Dimas Tadeu Covas, Sandra Coccuzzo Sampaio, Maria Carolina Elias, José Salvatore Leister Patané, Vincent Louis Viala, Antonio Jorge Martins, Ricardo Haddad, Claudia Renata dos Santos Barros, Elaine Cristina Marqueze, Raul Machado Neto, Debora Botequio Moretti, Jardelina de Souza Todao Bernardino, Loyze Paola Oliveira de Lima, Luiz Aurelio de Campos Crispin. Centro de Genômica Funcional da ESALQ: Luiz Lehmann Coutinho, Ricardo Augusto Brassaloti, Raquel de Lello Rocha Campos Cassano. NGS Soluções Genômicas: Pilar Drummond Sampaio Corrêa Mariani. FZEA-USP Pirassununga: Mirele Daiana Poleti, Jessika Cristina Chagas Lesbon, Elisangela Chicaroni Mattos, Heidge Fukumasu. USP-Botucatu: Rejane Maria Tommasini Grotto, Jayme A. Souza-Neto, Guilherme Targino Valente, Patricia Akemi Assato, Felipe Allan da Silva da Costa, Bianca Cechetto Carlos. Mendelics: Bibiana Santos, João Paulo Kitajima, Erika Freitas, David Schlesinger. Hemocentro Ribeirão Preto: Simone Kashima, Evandra Strazza Rodrigues, Svetoslav Nanev Slavov, Elaine Vieira dos Santos, Rafael dos Santos Bezerra, Luiz Carlos Junior de Alcantara, Marta Giovanetti, Vagner Fonseca, Flavia Aburjaile, Rodrigo Tocantins Calado. FAMERP-SJRP: Cecília Artico Banho, Lívia Sacchetto, Fábio Sossai Possebon, Leila Sabrina Ullmann, Cíntia Bittar, Guilherme Campos, Helena Lage Ferreira, Jorge A. Petroli Marchesi, Maisa C. Pereira Parra, Marília Moraes, Paula Rahal, Paulo Inacio da Costa, João Pessoa Araújo Jr., Maurício Lacerda Nogueira. Prefeitura de Sao Paulo: Melissa Palmieri. |
| EPI_ISL_1966209, EPI_ISL_1966210 | UBS HELENA MARREY | Instituto Butantan / Mendelics | Instituto Butantan: Dimas Tadeu Covas, Sandra Coccuzzo Sampaio, Maria Carolina Elias, José Salvatore Leister Patané, Vincent Louis Viala, Antonio Jorge Martins, Ricardo Haddad, Claudia Renata dos Santos Barros, Elaine Cristina Marqueze, Raul Machado Neto, Debora Botequio Moretti, Jardelina de Souza Todao Bernardino, Loyze Paola Oliveira de Lima, Luiz Aurelio de Campos Crispin. Centro de Genômica Funcional da ESALQ: Luiz Lehmann Coutinho, Ricardo Augusto Brassaloti, Raquel de Lello Rocha Campos Cassano. NGS Soluções Genômicas: Pilar Drummond Sampaio Corrêa Mariani. FZEA-USP Pirassununga: Mirele Daiana Poleti, Jessika Cristina Chagas Lesbon, Elisangela Chicaroni Mattos, Heidge Fukumasu. USP-Botucatu: Rejane Maria Tommasini Grotto, Jayme A. Souza-Neto, Guilherme Targino Valente, Patricia Akemi Assato, Felipe Allan da Silva da Costa, Bianca Cechetto Carlos. Mendelics: Bibiana Santos, João Paulo Kitajima, Erika Freitas, David Schlesinger. Hemocentro Ribeirão Preto: Simone Kashima, Evandra Strazza Rodrigues, Svetoslav Nanev Slavov, Elaine Vieira dos Santos, Rafael dos Santos Bezerra, Luiz Carlos Junior de Alcantara, Marta Giovanetti, Vagner Fonseca, Flavia Aburjaile, Rodrigo Tocantins Calado. FAMERP-SJRP: Cecília Artico Banho, Lívia Sacchetto, Fábio Sossai Possebon, Leila Sabrina Ullmann, Cíntia Bittar, Guilherme Campos, Helena Lage Ferreira, Jorge A. Petroli Marchesi, Maisa C. Pereira Parra, Marília Moraes, Paula Rahal, Paulo Inacio da Costa, João Pessoa Araújo Jr., Maurício Lacerda Nogueira. Prefeitura de Sao Paulo: Melissa Palmieri. |
| EPI_ISL_1966211 | UBS EMILIA COSME CERQUEIRA | Instituto Butantan / Mendelics | Instituto Butantan: Dimas Tadeu Covas, Sandra Coccuzzo Sampaio, Maria Carolina Elias, José Salvatore Leister Patané, Vincent Louis Viala, Antonio Jorge Martins, Ricardo Haddad, Claudia Renata dos Santos Barros, Elaine Cristina Marqueze, Raul Machado Neto, Debora Botequio Moretti, Jardelina de Souza Todao Bernardino, Loyze Paola Oliveira de Lima, Luiz Aurelio de Campos Crispin. Centro de Genômica Funcional da ESALQ: Luiz Lehmann Coutinho, Ricardo Augusto Brassaloti, Raquel de Lello Rocha Campos Cassano. NGS Soluções Genômicas: Pilar Drummond Sampaio Corrêa Mariani. FZEA-USP Pirassununga: Mirele Daiana Poleti, Jessika Cristina Chagas Lesbon, Elisangela Chicaroni Mattos, Heidge Fukumasu. USP-Botucatu: Rejane Maria Tommasini Grotto, Jayme A. Souza-Neto, Guilherme Targino Valente, Patricia Akemi Assato, Felipe Allan da Silva da Costa, Bianca Cechetto Carlos. Mendelics: Bibiana Santos, João Paulo Kitajima, Erika Freitas, David Schlesinger. Hemocentro Ribeirão Preto: Simone Kashima, Evandra Strazza Rodrigues, Svetoslav Nanev Slavov, Elaine Vieira dos Santos, Rafael dos Santos Bezerra, Luiz Carlos Junior de Alcantara, Marta Giovanetti, Vagner Fonseca, Flavia Aburjaile, Rodrigo Tocantins Calado. FAMERP-SJRP: Cecília Artico Banho, Lívia Sacchetto, Fábio Sossai Possebon, Leila Sabrina Ullmann, Cíntia Bittar, Guilherme Campos, Helena Lage Ferreira, Jorge A. Petroli Marchesi, Maisa C. Pereira Parra, Marília Moraes, Paula Rahal, Paulo Inacio da Costa, João Pessoa Araújo Jr., Maurício Lacerda Nogueira. Prefeitura de Sao Paulo: Melissa Palmieri. |
| EPI_ISL_1966212 | UBS SYLVIO JOAO L DE LUCIA | Instituto Butantan / Mendelics | Instituto Butantan: Dimas Tadeu Covas, Sandra Coccuzzo Sampaio, Maria Carolina Elias, José Salvatore Leister Patané, Vincent Louis Viala, Antonio Jorge Martins, Ricardo Haddad, Claudia Renata dos Santos Barros, Elaine Cristina Marqueze, Raul Machado Neto, Debora Botequio Moretti, Jardelina de Souza Todao Bernardino, Loyze Paola Oliveira de Lima, Luiz Aurelio de Campos Crispin. Centro de Genômica Funcional da ESALQ: Luiz Lehmann Coutinho, Ricardo Augusto Brassaloti, Raquel de Lello Rocha Campos Cassano. NGS Soluções Genômicas: Pilar Drummond Sampaio Corrêa Mariani. FZEA-USP Pirassununga: Mirele Daiana Poleti, Jessika Cristina Chagas Lesbon, Elisangela Chicaroni Mattos, Heidge Fukumasu. USP-Botucatu: Rejane Maria Tommasini Grotto, Jayme A. Souza-Neto, Guilherme Targino Valente, Patricia Akemi Assato, Felipe Allan da Silva da Costa, Bianca Cechetto Carlos. Mendelics: Bibiana Santos, João Paulo Kitajima, Erika Freitas, David Schlesinger. Hemocentro Ribeirão Preto: Simone Kashima, Evandra Strazza Rodrigues, Svetoslav Nanev Slavov, Elaine Vieira dos Santos, Rafael dos Santos Bezerra, Luiz Carlos Junior de Alcantara, Marta Giovanetti, Vagner Fonseca, Flavia Aburjaile, Rodrigo Tocantins Calado. FAMERP-SJRP: Cecília Artico Banho, Lívia Sacchetto, Fábio Sossai Possebon, Leila Sabrina Ullmann, Cíntia Bittar, Guilherme Campos, Helena Lage Ferreira, Jorge A. Petroli Marchesi, Maisa C. Pereira Parra, Marília Moraes, Paula Rahal, Paulo Inacio da Costa, João Pessoa Araújo Jr., Maurício Lacerda Nogueira. Prefeitura de Sao Paulo: Melissa Palmieri. |
| EPI_ISL_1966213, EPI_ISL_1966214 | UBS EMILIA COSME CERQUEIRA | Instituto Butantan / Mendelics | Instituto Butantan: Dimas Tadeu Covas, Sandra Coccuzzo Sampaio, Maria Carolina Elias, José Salvatore Leister Patané, Vincent Louis Viala, Antonio Jorge Martins, Ricardo Haddad, Claudia Renata dos Santos Barros, Elaine Cristina Marqueze, Raul Machado Neto, Debora Botequio Moretti, Jardelina de Souza Todao Bernardino, Loyze Paola Oliveira de Lima, Luiz Aurelio de Campos Crispin. Centro de Genômica Funcional da ESALQ: Luiz Lehmann Coutinho, Ricardo Augusto Brassaloti, Raquel de Lello Rocha Campos Cassano. NGS Soluções Genômicas: Pilar Drummond Sampaio Corrêa Mariani. FZEA-USP Pirassununga: Mirele Daiana Poleti, Jessika Cristina Chagas Lesbon, Elisangela Chicaroni Mattos, Heidge Fukumasu. USP-Botucatu: Rejane Maria Tommasini Grotto, Jayme A. Souza-Neto, Guilherme Targino Valente, Patricia Akemi Assato, Felipe Allan da Silva da Costa, Bianca Cechetto Carlos. Mendelics: Bibiana Santos, João Paulo Kitajima, Erika Freitas, David Schlesinger. Hemocentro Ribeirão Preto: Simone Kashima, Evandra Strazza Rodrigues, Svetoslav Nanev Slavov, Elaine Vieira dos Santos, Rafael dos Santos Bezerra, Luiz Carlos Junior de Alcantara, Marta Giovanetti, Vagner Fonseca, Flavia Aburjaile, Rodrigo Tocantins Calado. FAMERP-SJRP: Cecília Artico Banho, Lívia Sacchetto, Fábio Sossai Possebon, Leila Sabrina Ullmann, Cíntia Bittar, Guilherme Campos, Helena Lage Ferreira, Jorge A. Petroli Marchesi, Maisa C. Pereira Parra, Marília Moraes, Paula Rahal, Paulo Inacio da Costa, João Pessoa Araújo Jr., Maurício Lacerda Nogueira. Prefeitura de Sao Paulo: Melissa Palmieri. |
| EPI_ISL_1966215, EPI_ISL_1966217 | UBS HELENA MARREY | Instituto Butantan / Mendelics | Instituto Butantan: Dimas Tadeu Covas, Sandra Coccuzzo Sampaio, Maria Carolina Elias, José Salvatore Leister Patané, Vincent Louis Viala, Antonio Jorge Martins, Ricardo Haddad, Claudia Renata dos Santos Barros, Elaine Cristina Marqueze, Raul Machado Neto, Debora Botequio Moretti, Jardelina de Souza Todao Bernardino, Loyze Paola Oliveira de Lima, Luiz Aurelio de Campos Crispin. Centro de Genômica Funcional da ESALQ: Luiz Lehmann Coutinho, Ricardo Augusto Brassaloti, Raquel de Lello Rocha Campos Cassano. NGS Soluções Genômicas: Pilar Drummond Sampaio Corrêa Mariani. FZEA-USP Pirassununga: Mirele Daiana Poleti, Jessika Cristina Chagas Lesbon, Elisangela Chicaroni Mattos, Heidge Fukumasu. USP-Botucatu: Rejane Maria Tommasini Grotto, Jayme A. Souza-Neto, Guilherme Targino Valente, Patricia Akemi Assato, Felipe Allan da Silva da Costa, Bianca Cechetto Carlos. Mendelics: Bibiana Santos, João Paulo Kitajima, Erika Freitas, David Schlesinger. Hemocentro Ribeirão Preto: Simone Kashima, Evandra Strazza Rodrigues, Svetoslav Nanev Slavov, Elaine Vieira dos Santos, Rafael dos Santos Bezerra, Luiz Carlos Junior de Alcantara, Marta Giovanetti, Vagner Fonseca, Flavia Aburjaile, Rodrigo Tocantins Calado. FAMERP-SJRP: Cecília Artico Banho, Lívia Sacchetto, Fábio Sossai Possebon, Leila Sabrina Ullmann, Cíntia Bittar, Guilherme Campos, Helena Lage Ferreira, Jorge A. Petroli Marchesi, Maisa C. Pereira Parra, Marília Moraes, Paula Rahal, Paulo Inacio da Costa, João Pessoa Araújo Jr., Maurício Lacerda Nogueira. Prefeitura de Sao Paulo: Melissa Palmieri. |
| EPI_ISL_1966218 | PPA FENELON GUEDES PEREIRA | Instituto Butantan / Mendelics | Instituto Butantan: Dimas Tadeu Covas, Sandra Coccuzzo Sampaio, Maria Carolina Elias, José Salvatore Leister Patané, Vincent Louis Viala, Antonio Jorge Martins, Ricardo Haddad, Claudia Renata dos Santos Barros, Elaine Cristina Marqueze, Raul Machado Neto, Debora Botequio Moretti, Jardelina de Souza Todao Bernardino, Loyze Paola Oliveira de Lima, Luiz Aurelio de Campos Crispin. Centro de Genômica Funcional da ESALQ: Luiz Lehmann Coutinho, Ricardo Augusto Brassaloti, Raquel de Lello Rocha Campos Cassano. NGS Soluções Genômicas: Pilar Drummond Sampaio Corrêa Mariani. FZEA-USP Pirassununga: Mirele Daiana Poleti, Jessika Cristina Chagas Lesbon, Elisangela Chicaroni Mattos, Heidge Fukumasu. USP-Botucatu: Rejane Maria Tommasini Grotto, Jayme A. Souza-Neto, Guilherme Targino Valente, Patricia Akemi Assato, Felipe Allan da Silva da Costa, Bianca Cechetto Carlos. Mendelics: Bibiana Santos, João Paulo Kitajima, Erika Freitas, David Schlesinger. Hemocentro Ribeirão Preto: Simone Kashima, Evandra Strazza Rodrigues, Svetoslav Nanev Slavov, Elaine Vieira dos Santos, Rafael dos Santos Bezerra, Luiz Carlos Junior de Alcantara, Marta Giovanetti, Vagner Fonseca, Flavia Aburjaile, Rodrigo Tocantins Calado. FAMERP-SJRP: Cecília Artico Banho, Lívia Sacchetto, Fábio Sossai Possebon, Leila Sabrina Ullmann, Cíntia Bittar, Guilherme Campos, Helena Lage Ferreira, Jorge A. Petroli Marchesi, Maisa C. Pereira Parra, Marília Moraes, Paula Rahal, Paulo Inacio da Costa, João Pessoa Araújo Jr., Maurício Lacerda Nogueira. Prefeitura de Sao Paulo: Melissa Palmieri. |
| EPI_ISL_1966222 | PRONTO ATENDIMENTO SAO JOSE | Instituto Butantan / Mendelics | Instituto Butantan: Dimas Tadeu Covas, Sandra Coccuzzo Sampaio, Maria Carolina Elias, José Salvatore Leister Patané, Vincent Louis Viala, Antonio Jorge Martins, Ricardo Haddad, Claudia Renata dos Santos Barros, Elaine Cristina Marqueze, Raul Machado Neto, Debora Botequio Moretti, Jardelina de Souza Todao Bernardino, Loyze Paola Oliveira de Lima, Luiz Aurelio de Campos Crispin. Centro de Genômica Funcional da ESALQ: Luiz Lehmann Coutinho, Ricardo Augusto Brassaloti, Raquel de Lello Rocha Campos Cassano. NGS Soluções Genômicas: Pilar Drummond Sampaio Corrêa Mariani. FZEA-USP Pirassununga: Mirele Daiana Poleti, Jessika Cristina Chagas Lesbon, Elisangela Chicaroni Mattos, Heidge Fukumasu. USP-Botucatu: Rejane Maria Tommasini Grotto, Jayme A. Souza-Neto, Guilherme Targino Valente, Patricia Akemi Assato, Felipe Allan da Silva da Costa, Bianca Cechetto Carlos. Mendelics: Bibiana Santos, João Paulo Kitajima, Erika Freitas, David Schlesinger. Hemocentro Ribeirão Preto: Simone Kashima, Evandra Strazza |

| | | | |
|---|---|---|---|
| | | | Rodrigues, Svetoslav Nanev Slavov, Elaine Vieira dos Santos, Rafael dos Santos Bezerra, Luiz Carlos Junior de Alcantara, Marta Giovanetti, Vagner Fonseca, Flavia Aburjaile, Rodrigo Tocantins Calado. FAMERP-SJRP: Cecília Artico Banho, Lívia Sacchetto, Fábio Sossai Possebon, Leila Sabrina Ullmann, Cíntia Bittar, Guilherme Campos, Helena Lage Ferreira, Jorge A. Petroli Marchesi, Maisa C. Pereira Parra, Marília Moraes, Paula Rahal, Paulo Inacio da Costa, João Pessoa Araújo Jr., Maurício Lacerda Nogueira. Prefeitura de Sao Paulo: Melissa Palmieri. |
| EPI_ISL_1966223, EPI_ISL_1966224, EPI_ISL_1966225 | PRONTO ATENDIMENTO VILA PADRE ANCHIETA | Instituto Butantan / Mendelics | Instituto Butantan: Dimas Tadeu Covas, Sandra Coccuzzo Sampaio, Maria Carolina Elias, José Salvatore Leister Patané, Vincent Louis Viala, Antonio Jorge Martins, Ricardo Haddad, Claudia Renata dos Santos Barros, Elaine Cristina Marqueze, Raul Machado Neto, Debora Botequio Moretti, Jardelina de Souza Todao Bernardino, Loyze Paola Oliveira de Lima, Luiz Aurelio de Campos Crispin. Centro de Genômica Funcional da ESALQ: Luiz Lehmann Coutinho, Ricardo Augusto Brassaloti, Raquel de Lello Rocha Campos Cassano. NGS Soluções Genômicas: Pilar Drummond Sampaio Corrêa Mariani. FZEA-USP Pirassununga: Mirele Daiana Poleti, Jessika Cristina Chagas Lesbon, Elisangela Chicaroni Mattos, Heidge Fukumasu. USP-Botucatu: Rejane Maria Tommasini Grotto, Jayme A. Souza-Neto, Guilherme Targino Valente, Patricia Akemi Assato, Felipe Allan da Silva da Costa, Bianca Cechetto Carlos. Mendelics: Bibiana Santos, João Paulo Kitajima, Erika Freitas, David Schlesinger. Hemocentro Ribeirão Preto: Simone Kashima, Evandra Strazza Rodrigues, Svetoslav Nanev Slavov, Elaine Vieira dos Santos, Rafael dos Santos Bezerra, Luiz Carlos Junior de Alcantara, Marta Giovanetti, Vagner Fonseca, Flavia Aburjaile, Rodrigo Tocantins Calado. FAMERP-SJRP: Cecília Artico Banho, Lívia Sacchetto, Fábio Sossai Possebon, Leila Sabrina Ullmann, Cíntia Bittar, Guilherme Campos, Helena Lage Ferreira, Jorge A. Petroli Marchesi, Maisa C. Pereira Parra, Marília Moraes, Paula Rahal, Paulo Inacio da Costa, João Pessoa Araújo Jr., Maurício Lacerda Nogueira. Prefeitura de Sao Paulo: Melissa Palmieri. |
| EPI_ISL_1966226 | PRONTO ATENDIMENTO SAO JOSE | Instituto Butantan / Mendelics | Instituto Butantan: Dimas Tadeu Covas, Sandra Coccuzzo Sampaio, Maria Carolina Elias, José Salvatore Leister Patané, Vincent Louis Viala, Antonio Jorge Martins, Ricardo Haddad, Claudia Renata dos Santos Barros, Elaine Cristina Marqueze, Raul Machado Neto, Debora Botequio Moretti, Jardelina de Souza Todao Bernardino, Loyze Paola Oliveira de Lima, Luiz Aurelio de Campos Crispin. Centro de Genômica Funcional da ESALQ: Luiz Lehmann Coutinho, Ricardo Augusto Brassaloti, Raquel de Lello Rocha Campos Cassano. NGS Soluções Genômicas: Pilar Drummond Sampaio Corrêa Mariani. FZEA-USP Pirassununga: Mirele Daiana Poleti, Jessika Cristina Chagas Lesbon, Elisangela Chicaroni Mattos, Heidge Fukumasu. USP-Botucatu: Rejane Maria Tommasini Grotto, Jayme A. Souza-Neto, Guilherme Targino Valente, Patricia Akemi Assato, Felipe Allan da Silva da Costa, Bianca Cechetto Carlos. Mendelics: Bibiana Santos, João Paulo Kitajima, Erika Freitas, David Schlesinger. Hemocentro Ribeirão Preto: Simone Kashima, Evandra Strazza Rodrigues, Svetoslav Nanev Slavov, Elaine Vieira dos Santos, Rafael dos Santos Bezerra, Luiz Carlos Junior de Alcantara, Marta Giovanetti, Vagner Fonseca, Flavia Aburjaile, Rodrigo Tocantins Calado. FAMERP-SJRP: Cecília Artico Banho, Lívia Sacchetto, Fábio Sossai Possebon, Leila Sabrina Ullmann, Cíntia Bittar, Guilherme Campos, Helena Lage Ferreira, Jorge A. Petroli Marchesi, Maisa C. Pereira Parra, Marília Moraes, Paula Rahal, Paulo Inacio da Costa, João Pessoa Araújo Jr., Maurício Lacerda Nogueira. Prefeitura de Sao Paulo: Melissa Palmieri. |
| EPI_ISL_1966227 | UBS SAO JUDAS | Instituto Butantan / Mendelics | Instituto Butantan: Dimas Tadeu Covas, Sandra Coccuzzo Sampaio, Maria Carolina Elias, José Salvatore Leister Patané, Vincent Louis Viala, Antonio Jorge Martins, Ricardo Haddad, Claudia Renata dos Santos Barros, Elaine Cristina Marqueze, Raul Machado Neto, Debora Botequio Moretti, Jardelina de Souza Todao Bernardino, Loyze Paola Oliveira de Lima, Luiz Aurelio de Campos Crispin. Centro de Genômica Funcional da ESALQ: Luiz Lehmann Coutinho, Ricardo Augusto Brassaloti, Raquel de Lello Rocha Campos Cassano. NGS Soluções Genômicas: Pilar Drummond Sampaio Corrêa Mariani. FZEA-USP Pirassununga: Mirele Daiana Poleti, Jessika Cristina Chagas Lesbon, Elisangela Chicaroni Mattos, Heidge Fukumasu. USP-Botucatu: Rejane Maria Tommasini Grotto, Jayme A. Souza-Neto, Guilherme Targino Valente, Patricia Akemi Assato, Felipe Allan da Silva da Costa, Bianca Cechetto Carlos. Mendelics: Bibiana Santos, João Paulo Kitajima, Erika Freitas, David Schlesinger. Hemocentro Ribeirão Preto: Simone Kashima, Evandra Strazza Rodrigues, Svetoslav Nanev Slavov, Elaine Vieira dos Santos, Rafael dos Santos Bezerra, Luiz Carlos Junior de Alcantara, Marta Giovanetti, Vagner Fonseca, Flavia Aburjaile, Rodrigo Tocantins Calado. FAMERP-SJRP: Cecília Artico Banho, Lívia Sacchetto, Fábio Sossai Possebon, Leila Sabrina Ullmann, Cíntia Bittar, Guilherme Campos, Helena Lage Ferreira, Jorge A. Petroli Marchesi, Maisa C. Pereira Parra, Marília Moraes, Paula Rahal, Paulo Inacio da Costa, João Pessoa Araújo Jr., Maurício Lacerda Nogueira. Prefeitura de Sao Paulo: Melissa Palmieri. |
| EPI_ISL_1966229 | UBS SYLVIO JOAO L DE LUCIA | Instituto Butantan / Mendelics | Instituto Butantan: Dimas Tadeu Covas, Sandra Coccuzzo Sampaio, Maria Carolina Elias, José Salvatore Leister Patané, Vincent Louis Viala, Antonio Jorge Martins, Ricardo Haddad, Claudia Renata dos Santos Barros, Elaine Cristina Marqueze, Raul Machado Neto, Debora Botequio Moretti, Jardelina de Souza Todao Bernardino, Loyze Paola Oliveira de Lima, Luiz Aurelio de Campos Crispin. Centro de Genômica Funcional da ESALQ: Luiz Lehmann Coutinho, Ricardo Augusto Brassaloti, Raquel de Lello Rocha Campos Cassano. NGS Soluções Genômicas: Pilar Drummond Sampaio Corrêa Mariani. FZEA-USP Pirassununga: Mirele Daiana Poleti, Jessika Cristina Chagas Lesbon, Elisangela Chicaroni Mattos, Heidge Fukumasu. USP-Botucatu: Rejane Maria Tommasini Grotto, Jayme A. Souza-Neto, Guilherme Targino Valente, Patricia Akemi Assato, Felipe Allan da Silva da Costa, Bianca Cechetto Carlos. Mendelics: Bibiana Santos, João Paulo Kitajima, Erika Freitas, David Schlesinger. Hemocentro Ribeirão Preto: Simone Kashima, Evandra Strazza Rodrigues, Svetoslav Nanev Slavov, Elaine Vieira dos Santos, Rafael dos Santos Bezerra, Luiz Carlos Junior de Alcantara, Marta Giovanetti, Vagner Fonseca, Flavia Aburjaile, Rodrigo Tocantins Calado. FAMERP-SJRP: Cecília Artico Banho, Lívia Sacchetto, Fábio Sossai Possebon, Leila Sabrina Ullmann, Cíntia Bittar, Guilherme Campos, Helena Lage Ferreira, Jorge A. Petroli Marchesi, Maisa C. Pereira Parra, Marília Moraes, Paula Rahal, Paulo Inacio da Costa, João Pessoa Araújo Jr., Maurício Lacerda Nogueira. Prefeitura de Sao Paulo: Melissa Palmieri. |
| EPI_ISL_1966230 | UBS EMILIA COSME CERQUEIRA | Instituto Butantan / Mendelics | Instituto Butantan: Dimas Tadeu Covas, Sandra Coccuzzo Sampaio, Maria Carolina Elias, José Salvatore Leister Patané, Vincent Louis Viala, Antonio Jorge Martins, Ricardo Haddad, Claudia Renata dos Santos Barros, Elaine Cristina Marqueze, Raul Machado Neto, Debora Botequio Moretti, Jardelina de Souza Todao Bernardino, Loyze Paola Oliveira de Lima, Luiz Aurelio de Campos Crispin. Centro de Genômica Funcional da ESALQ: Luiz Lehmann Coutinho, Ricardo Augusto Brassaloti, Raquel de Lello Rocha Campos Cassano. NGS Soluções Genômicas: Pilar Drummond Sampaio Corrêa Mariani. FZEA-USP Pirassununga: Mirele Daiana Poleti, Jessika Cristina Chagas Lesbon, Elisangela Chicaroni Mattos, Heidge Fukumasu. USP-Botucatu: Rejane Maria Tommasini Grotto, Jayme A. Souza-Neto, Guilherme Targino Valente, Patricia Akemi Assato, Felipe Allan da Silva da Costa, Bianca Cechetto Carlos. Mendelics: Bibiana Santos, João Paulo Kitajima, Erika Freitas, David Schlesinger. Hemocentro Ribeirão Preto: Simone Kashima, Evandra Strazza Rodrigues, Svetoslav Nanev Slavov, Elaine Vieira dos Santos, Rafael dos Santos Bezerra, Luiz Carlos Junior de Alcantara, Marta Giovanetti, Vagner Fonseca, Flavia Aburjaile, Rodrigo Tocantins Calado. FAMERP-SJRP: Cecília Artico Banho, Lívia Sacchetto, Fábio Sossai Possebon, Leila Sabrina Ullmann, Cíntia Bittar, Guilherme Campos, Helena Lage Ferreira, Jorge A. Petroli Marchesi, Maisa C. Pereira Parra, Marília Moraes, Paula Rahal, Paulo Inacio da Costa, João Pessoa Araújo Jr., Maurício Lacerda Nogueira. Prefeitura de Sao Paulo: Melissa Palmieri. |
| EPI_ISL_1966231 | UBS IRMA AGUEDA MARIA JAIME | Instituto Butantan / Mendelics | Instituto Butantan: Dimas Tadeu Covas, Sandra Coccuzzo Sampaio, Maria Carolina Elias, José Salvatore Leister Patané, Vincent Louis Viala, Antonio Jorge Martins, Ricardo Haddad, Claudia Renata dos Santos Barros, Elaine Cristina Marqueze, Raul Machado Neto, Debora Botequio Moretti, Jardelina de Souza Todao Bernardino, Loyze Paola Oliveira de Lima, Luiz Aurelio de Campos Crispin. Centro de Genômica Funcional da ESALQ: Luiz Lehmann Coutinho, Ricardo Augusto Brassaloti, Raquel de Lello Rocha Campos Cassano. NGS Soluções Genômicas: Pilar Drummond Sampaio Corrêa Mariani. FZEA-USP Pirassununga: Mirele Daiana Poleti, Jessika Cristina Chagas Lesbon, Elisangela Chicaroni Mattos, Heidge Fukumasu. USP-Botucatu: Rejane Maria Tommasini Grotto, Jayme A. Souza-Neto, Guilherme Targino Valente, Patricia Akemi Assato, Felipe Allan da Silva da Costa, Bianca Cechetto Carlos. Mendelics: Bibiana Santos, João Paulo Kitajima, Erika Freitas, David Schlesinger. Hemocentro Ribeirão Preto: Simone Kashima, Evandra Strazza Rodrigues, Svetoslav Nanev Slavov, Elaine Vieira dos Santos, Rafael dos Santos Bezerra, Luiz Carlos Junior de Alcantara, Marta Giovanetti, Vagner Fonseca, Flavia Aburjaile, Rodrigo Tocantins Calado. FAMERP-SJRP: Cecília Artico Banho, Lívia Sacchetto, Fábio Sossai Possebon, Leila Sabrina Ullmann, Cíntia Bittar, Guilherme Campos, Helena Lage Ferreira, Jorge A. Petroli Marchesi, Maisa C. Pereira Parra, Marília Moraes, Paula Rahal, Paulo Inacio da Costa, João Pessoa Araújo Jr., Maurício Lacerda Nogueira. Prefeitura de Sao Paulo: Melissa Palmieri. |
| EPI_ISL_1966232 | USAFA GUILHERMINA | Instituto Butantan / Mendelics | Instituto Butantan: Dimas Tadeu Covas, Sandra Coccuzzo Sampaio, Maria Carolina Elias, José Salvatore Leister Patané, Vincent Louis Viala, Antonio Jorge Martins, Ricardo Haddad, Claudia Renata dos Santos Barros, Elaine Cristina Marqueze, Raul Machado Neto, Debora Botequio Moretti, Jardelina de Souza Todao Bernardino, Loyze Paola Oliveira de Lima, Luiz Aurelio de Campos Crispin. Centro de Genômica Funcional da ESALQ: Luiz Lehmann Coutinho, Ricardo Augusto Brassaloti, Raquel de Lello Rocha Campos Cassano. NGS Soluções Genômicas: Pilar Drummond Sampaio Corrêa Mariani. FZEA-USP Pirassununga: Mirele Daiana Poleti, Jessika Cristina Chagas Lesbon, Elisangela Chicaroni Mattos, Heidge Fukumasu. USP-Botucatu: Rejane Maria Tommasini Grotto, Jayme A. Souza-Neto, Guilherme Targino Valente, Patricia Akemi Assato, Felipe Allan da Silva da Costa, Bianca Cechetto Carlos. Mendelics: Bibiana Santos, João Paulo Kitajima, Erika Freitas, David Schlesinger. Hemocentro Ribeirão Preto: Simone Kashima, Evandra Strazza Rodrigues, Svetoslav Nanev Slavov, Elaine Vieira dos Santos, Rafael dos Santos Bezerra, Luiz Carlos Junior de Alcantara, Marta Giovanetti, Vagner Fonseca, Flavia Aburjaile, Rodrigo Tocantins Calado. FAMERP-SJRP: Cecília Artico Banho, Lívia Sacchetto, Fábio Sossai Possebon, Leila Sabrina Ullmann, Cíntia Bittar, Guilherme Campos, Helena Lage Ferreira, Jorge A. Petroli Marchesi, Maisa C. Pereira Parra, Marília Moraes, Paula Rahal, Paulo Inacio da Costa, João Pessoa Araújo Jr., Maurício Lacerda Nogueira. Prefeitura de Sao Paulo: Melissa Palmieri. |
| EPI_ISL_1966233 | USAFA MARACANA | Instituto Butantan / Mendelics | Instituto Butantan: Dimas Tadeu Covas, Sandra Coccuzzo Sampaio, Maria Carolina Elias, José Salvatore Leister Patané, Vincent Louis Viala, Antonio Jorge Martins, Ricardo Haddad, Claudia Renata dos Santos Barros, Elaine Cristina Marqueze, Raul Machado Neto, Debora Botequio Moretti, Jardelina de |

| | | | |
|---|---|---|---|
| | | | Souza Todao Bernardino, Loyze Paola Oliveira de Lima, Luiz Aurelio de Campos Crispin. Centro de Genômica Funcional da ESALQ: Luiz Lehmann Coutinho, Ricardo Augusto Brassaloti, Raquel de Lello Rocha Campos Cassano. NGS Soluções Genômicas: Pilar Drummond Sampaio Corrêa Mariani. FZEA-USP Pirassununga: Mirele Daiana Poleti, Jessika Cristina Chagas Lesbon, Elisangela Chicaroni Mattos, Heidge Fukumasu. USP-Botucatu: Rejane Maria Tommasini Grotto, Jayme A. Souza-Neto, Guilherme Targino Valente, Patricia Akemi Assato, Felipe Allan da Silva da Costa, Bianca Cechetto Carlos. Mendelics: Bibiana Santos, João Paulo Kitajima, Erika Freitas, David Schlesinger. Hemocentro Ribeirão Preto: Simone Kashima, Evandra Strazza Rodrigues, Svetoslav Nanev Slavov, Elaine Vieira dos Santos, Rafael dos Santos Bezerra, Luiz Carlos Junior de Alcantara, Marta Giovanetti, Vagner Fonseca, Flavia Aburjaile, Rodrigo Tocantins Calado. FAMERP-SJRP: Cecília Artico Banho, Lívia Sacchetto, Fábio Sossai Possebon, Leila Sabrina Ullmann, Cíntia Bittar, Guilherme Campos, Helena Lage Ferreira, Jorge A. Petroli Marchesi, Maisa C. Pereira Parra, Marília Moraes, Paula Rahal, Paulo Inacio da Costa, João Pessoa Araújo Jr., Maurício Lacerda Nogueira. Prefeitura de Sao Paulo: Melissa Palmieri. |
| EPI_ISL_1966236 | UNIDADE MISTA DE TAIACU | Instituto Butantan / Mendelics | Instituto Butantan: Dimas Tadeu Covas, Sandra Coccuzzo Sampaio, Maria Carolina Elias, José Salvatore Leister Patané, Vincent Louis Viala, Antonio Jorge Martins, Ricardo Haddad, Claudia Renata dos Santos Barros, Elaine Cristina Marqueze, Raul Machado Neto, Debora Botequio Moretti, Jardelina de Souza Todao Bernardino, Loyze Paola Oliveira de Lima, Luiz Aurelio de Campos Crispin. Centro de Genômica Funcional da ESALQ: Luiz Lehmann Coutinho, Ricardo Augusto Brassaloti, Raquel de Lello Rocha Campos Cassano. NGS Soluções Genômicas: Pilar Drummond Sampaio Corrêa Mariani. FZEA-USP Pirassununga: Mirele Daiana Poleti, Jessika Cristina Chagas Lesbon, Elisangela Chicaroni Mattos, Heidge Fukumasu. USP-Botucatu: Rejane Maria Tommasini Grotto, Jayme A. Souza-Neto, Guilherme Targino Valente, Patricia Akemi Assato, Felipe Allan da Silva da Costa, Bianca Cechetto Carlos. Mendelics: Bibiana Santos, João Paulo Kitajima, Erika Freitas, David Schlesinger. Hemocentro Ribeirão Preto: Simone Kashima, Evandra Strazza Rodrigues, Svetoslav Nanev Slavov, Elaine Vieira dos Santos, Rafael dos Santos Bezerra, Luiz Carlos Junior de Alcantara, Marta Giovanetti, Vagner Fonseca, Flavia Aburjaile, Rodrigo Tocantins Calado. FAMERP-SJRP: Cecília Artico Banho, Lívia Sacchetto, Fábio Sossai Possebon, Leila Sabrina Ullmann, Cíntia Bittar, Guilherme Campos, Helena Lage Ferreira, Jorge A. Petroli Marchesi, Maisa C. Pereira Parra, Marília Moraes, Paula Rahal, Paulo Inacio da Costa, João Pessoa Araújo Jr., Maurício Lacerda Nogueira. Prefeitura de Sao Paulo: Melissa Palmieri. |
| EPI_ISL_1966240 | CENTRO DE SAUDE DR MARIO DIAS DE AGUIAR CAPIVARI | Instituto Butantan / Mendelics | Instituto Butantan: Dimas Tadeu Covas, Sandra Coccuzzo Sampaio, Maria Carolina Elias, José Salvatore Leister Patané, Vincent Louis Viala, Antonio Jorge Martins, Ricardo Haddad, Claudia Renata dos Santos Barros, Elaine Cristina Marqueze, Raul Machado Neto, Debora Botequio Moretti, Jardelina de Souza Todao Bernardino, Loyze Paola Oliveira de Lima, Luiz Aurelio de Campos Crispin. Centro de Genômica Funcional da ESALQ: Luiz Lehmann Coutinho, Ricardo Augusto Brassaloti, Raquel de Lello Rocha Campos Cassano. NGS Soluções Genômicas: Pilar Drummond Sampaio Corrêa Mariani. FZEA-USP Pirassununga: Mirele Daiana Poleti, Jessika Cristina Chagas Lesbon, Elisangela Chicaroni Mattos, Heidge Fukumasu. USP-Botucatu: Rejane Maria Tommasini Grotto, Jayme A. Souza-Neto, Guilherme Targino Valente, Patricia Akemi Assato, Felipe Allan da Silva da Costa, Bianca Cechetto Carlos. Mendelics: Bibiana Santos, João Paulo Kitajima, Erika Freitas, David Schlesinger. Hemocentro Ribeirão Preto: Simone Kashima, Evandra Strazza Rodrigues, Svetoslav Nanev Slavov, Elaine Vieira dos Santos, Rafael dos Santos Bezerra, Luiz Carlos Junior de Alcantara, Marta Giovanetti, Vagner Fonseca, Flavia Aburjaile, Rodrigo Tocantins Calado. FAMERP-SJRP: Cecília Artico Banho, Lívia Sacchetto, Fábio Sossai Possebon, Leila Sabrina Ullmann, Cíntia Bittar, Guilherme Campos, Helena Lage Ferreira, Jorge A. Petroli Marchesi, Maisa C. Pereira Parra, Marília Moraes, Paula Rahal, Paulo Inacio da Costa, João Pessoa Araújo Jr., Maurício Lacerda Nogueira. Prefeitura de Sao Paulo: Melissa Palmieri. |
| EPI_ISL_1966242 | SANTA CASA DE MIS E MATERNIDADE N SRA DA CONCEICAO CUNHA | Instituto Butantan / Mendelics | Instituto Butantan: Dimas Tadeu Covas, Sandra Coccuzzo Sampaio, Maria Carolina Elias, José Salvatore Leister Patané, Vincent Louis Viala, Antonio Jorge Martins, Ricardo Haddad, Claudia Renata dos Santos Barros, Elaine Cristina Marqueze, Raul Machado Neto, Debora Botequio Moretti, Jardelina de Souza Todao Bernardino, Loyze Paola Oliveira de Lima, Luiz Aurelio de Campos Crispin. Centro de Genômica Funcional da ESALQ: Luiz Lehmann Coutinho, Ricardo Augusto Brassaloti, Raquel de Lello Rocha Campos Cassano. NGS Soluções Genômicas: Pilar Drummond Sampaio Corrêa Mariani. FZEA-USP Pirassununga: Mirele Daiana Poleti, Jessika Cristina Chagas Lesbon, Elisangela Chicaroni Mattos, Heidge Fukumasu. USP-Botucatu: Rejane Maria Tommasini Grotto, Jayme A. Souza-Neto, Guilherme Targino Valente, Patricia Akemi Assato, Felipe Allan da Silva da Costa, Bianca Cechetto Carlos. Mendelics: Bibiana Santos, João Paulo Kitajima, Erika Freitas, David Schlesinger. Hemocentro Ribeirão Preto: Simone Kashima, Evandra Strazza Rodrigues, Svetoslav Nanev Slavov, Elaine Vieira dos Santos, Rafael dos Santos Bezerra, Luiz Carlos Junior de Alcantara, Marta Giovanetti, Vagner Fonseca, Flavia Aburjaile, Rodrigo Tocantins Calado. FAMERP-SJRP: Cecília Artico Banho, Lívia Sacchetto, Fábio Sossai Possebon, Leila Sabrina Ullmann, Cíntia Bittar, Guilherme Campos, Helena Lage Ferreira, Jorge A. Petroli Marchesi, Maisa C. Pereira Parra, Marília Moraes, Paula Rahal, Paulo Inacio da Costa, João Pessoa Araújo Jr., Maurício Lacerda Nogueira. Prefeitura de Sao Paulo: Melissa Palmieri. |
| EPI_ISL_1966243 | UNIDADE MISTA DE SAUDE DE DUMONT | Instituto Butantan / Mendelics | Instituto Butantan: Dimas Tadeu Covas, Sandra Coccuzzo Sampaio, Maria Carolina Elias, José Salvatore Leister Patané, Vincent Louis Viala, Antonio Jorge Martins, Ricardo Haddad, Claudia Renata dos Santos Barros, Elaine Cristina Marqueze, Raul Machado Neto, Debora Botequio Moretti, Jardelina de Souza Todao Bernardino, Loyze Paola Oliveira de Lima, Luiz Aurelio de Campos Crispin. Centro de Genômica Funcional da ESALQ: Luiz Lehmann Coutinho, Ricardo Augusto Brassaloti, Raquel de Lello Rocha Campos Cassano. NGS Soluções Genômicas: Pilar Drummond Sampaio Corrêa Mariani. FZEA-USP Pirassununga: Mirele Daiana Poleti, Jessika Cristina Chagas Lesbon, Elisangela Chicaroni Mattos, Heidge Fukumasu. USP-Botucatu: Rejane Maria Tommasini Grotto, Jayme A. Souza-Neto, Guilherme Targino Valente, Patricia Akemi Assato, Felipe Allan da Silva da Costa, Bianca Cechetto Carlos. Mendelics: Bibiana Santos, João Paulo Kitajima, Erika Freitas, David Schlesinger. Hemocentro Ribeirão Preto: Simone Kashima, Evandra Strazza Rodrigues, Svetoslav Nanev Slavov, Elaine Vieira dos Santos, Rafael dos Santos Bezerra, Luiz Carlos Junior de Alcantara, Marta Giovanetti, Vagner Fonseca, Flavia Aburjaile, Rodrigo Tocantins Calado. FAMERP-SJRP: Cecília Artico Banho, Lívia Sacchetto, Fábio Sossai Possebon, Leila Sabrina Ullmann, Cíntia Bittar, Guilherme Campos, Helena Lage Ferreira, Jorge A. Petroli Marchesi, Maisa C. Pereira Parra, Marília Moraes, Paula Rahal, Paulo Inacio da Costa, João Pessoa Araújo Jr., Maurício Lacerda Nogueira. Prefeitura de Sao Paulo: Melissa Palmieri. |
| EPI_ISL_1966244 | POSTO MEDICO NATAL DIEGUES DE ESTIVA GERBI | Instituto Butantan / Mendelics | Instituto Butantan: Dimas Tadeu Covas, Sandra Coccuzzo Sampaio, Maria Carolina Elias, José Salvatore Leister Patané, Vincent Louis Viala, Antonio Jorge Martins, Ricardo Haddad, Claudia Renata dos Santos Barros, Elaine Cristina Marqueze, Raul Machado Neto, Debora Botequio Moretti, Jardelina de Souza Todao Bernardino, Loyze Paola Oliveira de Lima, Luiz Aurelio de Campos Crispin. Centro de Genômica Funcional da ESALQ: Luiz Lehmann Coutinho, Ricardo Augusto Brassaloti, Raquel de Lello Rocha Campos Cassano. NGS Soluções Genômicas: Pilar Drummond Sampaio Corrêa Mariani. FZEA-USP Pirassununga: Mirele Daiana Poleti, Jessika Cristina Chagas Lesbon, Elisangela Chicaroni Mattos, Heidge Fukumasu. USP-Botucatu: Rejane Maria Tommasini Grotto, Jayme A. Souza-Neto, Guilherme Targino Valente, Patricia Akemi Assato, Felipe Allan da Silva da Costa, Bianca Cechetto Carlos. Mendelics: Bibiana Santos, João Paulo Kitajima, Erika Freitas, David Schlesinger. Hemocentro Ribeirão Preto: Simone Kashima, Evandra Strazza Rodrigues, Svetoslav Nanev Slavov, Elaine Vieira dos Santos, Rafael dos Santos Bezerra, Luiz Carlos Junior de Alcantara, Marta Giovanetti, Vagner Fonseca, Flavia Aburjaile, Rodrigo Tocantins Calado. FAMERP-SJRP: Cecília Artico Banho, Lívia Sacchetto, Fábio Sossai Possebon, Leila Sabrina Ullmann, Cíntia Bittar, Guilherme Campos, Helena Lage Ferreira, Jorge A. Petroli Marchesi, Maisa C. Pereira Parra, Marília Moraes, Paula Rahal, Paulo Inacio da Costa, João Pessoa Araújo Jr., Maurício Lacerda Nogueira. Prefeitura de Sao Paulo: Melissa Palmieri. |
| EPI_ISL_1966245 | UNIDADE BASICA DE SAUDE DR ORESTES DE MOURA PINTO | Instituto Butantan / Mendelics | Instituto Butantan: Dimas Tadeu Covas, Sandra Coccuzzo Sampaio, Maria Carolina Elias, José Salvatore Leister Patané, Vincent Louis Viala, Antonio Jorge Martins, Ricardo Haddad, Claudia Renata dos Santos Barros, Elaine Cristina Marqueze, Raul Machado Neto, Debora Botequio Moretti, Jardelina de Souza Todao Bernardino, Loyze Paola Oliveira de Lima, Luiz Aurelio de Campos Crispin. Centro de Genômica Funcional da ESALQ: Luiz Lehmann Coutinho, Ricardo Augusto Brassaloti, Raquel de Lello Rocha Campos Cassano. NGS Soluções Genômicas: Pilar Drummond Sampaio Corrêa Mariani. FZEA-USP Pirassununga: Mirele Daiana Poleti, Jessika Cristina Chagas Lesbon, Elisangela Chicaroni Mattos, Heidge Fukumasu. USP-Botucatu: Rejane Maria Tommasini Grotto, Jayme A. Souza-Neto, Guilherme Targino Valente, Patricia Akemi Assato, Felipe Allan da Silva da Costa, Bianca Cechetto Carlos. Mendelics: Bibiana Santos, João Paulo Kitajima, Erika Freitas, David Schlesinger. Hemocentro Ribeirão Preto: Simone Kashima, Evandra Strazza Rodrigues, Svetoslav Nanev Slavov, Elaine Vieira dos Santos, Rafael dos Santos Bezerra, Luiz Carlos Junior de Alcantara, Marta Giovanetti, Vagner Fonseca, Flavia Aburjaile, Rodrigo Tocantins Calado. FAMERP-SJRP: Cecília Artico Banho, Lívia Sacchetto, Fábio Sossai Possebon, Leila Sabrina Ullmann, Cíntia Bittar, Guilherme Campos, Helena Lage Ferreira, Jorge A. Petroli Marchesi, Maisa C. Pereira Parra, Marília Moraes, Paula Rahal, Paulo Inacio da Costa, João Pessoa Araújo Jr., Maurício Lacerda Nogueira. Prefeitura de Sao Paulo: Melissa Palmieri. |
| EPI_ISL_1966246 | UNIDADE MISTA DE IGUAPE | Instituto Butantan / Mendelics | Instituto Butantan: Dimas Tadeu Covas, Sandra Coccuzzo Sampaio, Maria Carolina Elias, José Salvatore Leister Patané, Vincent Louis Viala, Antonio Jorge Martins, Ricardo Haddad, Claudia Renata dos Santos Barros, Elaine Cristina Marqueze, Raul Machado Neto, Debora Botequio Moretti, Jardelina de Souza Todao Bernardino, Loyze Paola Oliveira de Lima, Luiz Aurelio de Campos Crispin. Centro de Genômica Funcional da ESALQ: Luiz Lehmann Coutinho, Ricardo Augusto Brassaloti, Raquel de Lello Rocha Campos Cassano. NGS Soluções Genômicas: Pilar Drummond Sampaio Corrêa Mariani. FZEA-USP Pirassununga: Mirele Daiana Poleti, Jessika Cristina Chagas Lesbon, Elisangela Chicaroni Mattos, Heidge Fukumasu. USP-Botucatu: Rejane Maria Tommasini Grotto, Jayme A. Souza-Neto, Guilherme Targino Valente, Patricia Akemi Assato, Felipe Allan da Silva da Costa, Bianca Cechetto Carlos. Mendelics: Bibiana Santos, João Paulo Kitajima, Erika Freitas, David Schlesinger. Hemocentro Ribeirão Preto: Simone Kashima, Evandra Strazza Rodrigues, Svetoslav Nanev Slavov, Elaine Vieira dos Santos, Rafael dos Santos Bezerra, Luiz Carlos Junior de Alcantara, Marta Giovanetti, Vagner Fonseca, Flavia Aburjaile, Rodrigo Tocantins Calado. FAMERP-SJRP: Cecília Artico Banho, Lívia Sacchetto, Fábio Sossai Possebon, Leila Sabrina |

| | | | |
|---|---|---|---|
| | | | Ullmann, Cíntia Bittar, Guilherme Campos, Helena Lage Ferreira, Jorge A. Petroli Marchesi, Maisa C. Pereira Parra, Marília Moraes, Paula Rahal, Paulo Inacio da Costa, João Pessoa Araújo Jr., Maurício Lacerda Nogueira. Prefeitura de Sao Paulo: Melissa Palmieri. |
| EPI_ISL_1966247 | SECRETARIA DE SAUDE DO MUNICIPIO DE IPUA | Instituto Butantan / Mendelics | Instituto Butantan: Dimas Tadeu Covas, Sandra Coccuzzo Sampaio, Maria Carolina Elias, José Salvatore Leister Patané, Vincent Louis Viala, Antonio Jorge Martins, Ricardo Haddad, Claudia Renata dos Santos Barros, Elaine Cristina Marqueze, Raul Machado Neto, Debora Botequio Moretti, Jardelina de Souza Todao Bernardino, Loyze Paola Oliveira de Lima, Luiz Aurelio de Campos Crispin. Centro de Genômica Funcional da ESALQ: Luiz Lehmann Coutinho, Ricardo Augusto Brassaloti, Raquel de Lello Rocha Campos Cassano. NGS Soluções Genômicas: Pilar Drummond Sampaio Corrêa Mariani. FZEA-USP Pirassununga: Mirele Daiana Poleti, Jessika Cristina Chagas Lesbon, Elisangela Chicaroni Mattos, Heidge Fukumasu. USP-Botucatu: Rejane Maria Tommasini Grotto, Jayme A. Souza-Neto, Guilherme Targino Valente, Patricia Akemi Assato, Felipe Allan da Silva da Costa, Bianca Cechetto Carlos. Mendelics: Bibiana Santos, João Paulo Kitajima, Erika Freitas, David Schlesinger. Hemocentro Ribeirão Preto: Simone Kashima, Evandra Strazza Rodrigues, Svetoslav Nanev Slavov, Elaine Vieira dos Santos, Rafael dos Santos Bezerra, Luiz Carlos Junior de Alcantara, Marta Giovanetti, Vagner Fonseca, Flavia Aburjaile, Rodrigo Tocantins Calado. FAMERP-SJRP: Cecília Artico Banho, Lívia Sacchetto, Fábio Sossai Possebon, Leila Sabrina Ullmann, Cíntia Bittar, Guilherme Campos, Helena Lage Ferreira, Jorge A. Petroli Marchesi, Maisa C. Pereira Parra, Marília Moraes, Paula Rahal, Paulo Inacio da Costa, João Pessoa Araújo Jr., Maurício Lacerda Nogueira. Prefeitura de Sao Paulo: Melissa Palmieri. |
| EPI_ISL_1966248, EPI_ISL_1966249 | UNIDADE DE PRONTO ATENDIMENTO DE JALES | Instituto Butantan / Mendelics | Instituto Butantan: Dimas Tadeu Covas, Sandra Coccuzzo Sampaio, Maria Carolina Elias, José Salvatore Leister Patané, Vincent Louis Viala, Antonio Jorge Martins, Ricardo Haddad, Claudia Renata dos Santos Barros, Elaine Cristina Marqueze, Raul Machado Neto, Debora Botequio Moretti, Jardelina de Souza Todao Bernardino, Loyze Paola Oliveira de Lima, Luiz Aurelio de Campos Crispin. Centro de Genômica Funcional da ESALQ: Luiz Lehmann Coutinho, Ricardo Augusto Brassaloti, Raquel de Lello Rocha Campos Cassano. NGS Soluções Genômicas: Pilar Drummond Sampaio Corrêa Mariani. FZEA-USP Pirassununga: Mirele Daiana Poleti, Jessika Cristina Chagas Lesbon, Elisangela Chicaroni Mattos, Heidge Fukumasu. USP-Botucatu: Rejane Maria Tommasini Grotto, Jayme A. Souza-Neto, Guilherme Targino Valente, Patricia Akemi Assato, Felipe Allan da Silva da Costa, Bianca Cechetto Carlos. Mendelics: Bibiana Santos, João Paulo Kitajima, Erika Freitas, David Schlesinger. Hemocentro Ribeirão Preto: Simone Kashima, Evandra Strazza Rodrigues, Svetoslav Nanev Slavov, Elaine Vieira dos Santos, Rafael dos Santos Bezerra, Luiz Carlos Junior de Alcantara, Marta Giovanetti, Vagner Fonseca, Flavia Aburjaile, Rodrigo Tocantins Calado. FAMERP-SJRP: Cecília Artico Banho, Lívia Sacchetto, Fábio Sossai Possebon, Leila Sabrina Ullmann, Cíntia Bittar, Guilherme Campos, Helena Lage Ferreira, Jorge A. Petroli Marchesi, Maisa C. Pereira Parra, Marília Moraes, Paula Rahal, Paulo Inacio da Costa, João Pessoa Araújo Jr., Maurício Lacerda Nogueira. Prefeitura de Sao Paulo: Melissa Palmieri. |
| EPI_ISL_1966250 | UNIDADE SAUDE DA FAMILIA DRA ZILDA ARNS NEUMANN JALES | Instituto Butantan / Mendelics | Instituto Butantan: Dimas Tadeu Covas, Sandra Coccuzzo Sampaio, Maria Carolina Elias, José Salvatore Leister Patané, Vincent Louis Viala, Antonio Jorge Martins, Ricardo Haddad, Claudia Renata dos Santos Barros, Elaine Cristina Marqueze, Raul Machado Neto, Debora Botequio Moretti, Jardelina de Souza Todao Bernardino, Loyze Paola Oliveira de Lima, Luiz Aurelio de Campos Crispin. Centro de Genômica Funcional da ESALQ: Luiz Lehmann Coutinho, Ricardo Augusto Brassaloti, Raquel de Lello Rocha Campos Cassano. NGS Soluções Genômicas: Pilar Drummond Sampaio Corrêa Mariani. FZEA-USP Pirassununga: Mirele Daiana Poleti, Jessika Cristina Chagas Lesbon, Elisangela Chicaroni Mattos, Heidge Fukumasu. USP-Botucatu: Rejane Maria Tommasini Grotto, Jayme A. Souza-Neto, Guilherme Targino Valente, Patricia Akemi Assato, Felipe Allan da Silva da Costa, Bianca Cechetto Carlos. Mendelics: Bibiana Santos, João Paulo Kitajima, Erika Freitas, David Schlesinger. Hemocentro Ribeirão Preto: Simone Kashima, Evandra Strazza Rodrigues, Svetoslav Nanev Slavov, Elaine Vieira dos Santos, Rafael dos Santos Bezerra, Luiz Carlos Junior de Alcantara, Marta Giovanetti, Vagner Fonseca, Flavia Aburjaile, Rodrigo Tocantins Calado. FAMERP-SJRP: Cecília Artico Banho, Lívia Sacchetto, Fábio Sossai Possebon, Leila Sabrina Ullmann, Cíntia Bittar, Guilherme Campos, Helena Lage Ferreira, Jorge A. Petroli Marchesi, Maisa C. Pereira Parra, Marília Moraes, Paula Rahal, Paulo Inacio da Costa, João Pessoa Araújo Jr., Maurício Lacerda Nogueira. Prefeitura de Sao Paulo: Melissa Palmieri. |
| EPI_ISL_1966251 | ESF DR LUIS ERNESTO SANDI MORI JALES | Instituto Butantan / Mendelics | Instituto Butantan: Dimas Tadeu Covas, Sandra Coccuzzo Sampaio, Maria Carolina Elias, José Salvatore Leister Patané, Vincent Louis Viala, Antonio Jorge Martins, Ricardo Haddad, Claudia Renata dos Santos Barros, Elaine Cristina Marqueze, Raul Machado Neto, Debora Botequio Moretti, Jardelina de Souza Todao Bernardino, Loyze Paola Oliveira de Lima, Luiz Aurelio de Campos Crispin. Centro de Genômica Funcional da ESALQ: Luiz Lehmann Coutinho, Ricardo Augusto Brassaloti, Raquel de Lello Rocha Campos Cassano. NGS Soluções Genômicas: Pilar Drummond Sampaio Corrêa Mariani. FZEA-USP Pirassununga: Mirele Daiana Poleti, Jessika Cristina Chagas Lesbon, Elisangela Chicaroni Mattos, Heidge Fukumasu. USP-Botucatu: Rejane Maria Tommasini Grotto, Jayme A. Souza-Neto, Guilherme Targino Valente, Patricia Akemi Assato, Felipe Allan da Silva da Costa, Bianca Cechetto Carlos. Mendelics: Bibiana Santos, João Paulo Kitajima, Erika Freitas, David Schlesinger. Hemocentro Ribeirão Preto: Simone Kashima, Evandra Strazza Rodrigues, Svetoslav Nanev Slavov, Elaine Vieira dos Santos, Rafael dos Santos Bezerra, Luiz Carlos Junior de Alcantara, Marta Giovanetti, Vagner Fonseca, Flavia Aburjaile, Rodrigo Tocantins Calado. FAMERP-SJRP: Cecília Artico Banho, Lívia Sacchetto, Fábio Sossai Possebon, Leila Sabrina Ullmann, Cíntia Bittar, Guilherme Campos, Helena Lage Ferreira, Jorge A. Petroli Marchesi, Maisa C. Pereira Parra, Marília Moraes, Paula Rahal, Paulo Inacio da Costa, João Pessoa Araújo Jr., Maurício Lacerda Nogueira. Prefeitura de Sao Paulo: Melissa Palmieri. |
| EPI_ISL_1966252, EPI_ISL_1966253, EPI_ISL_1966255 | UBS DR MOHANNA ADAS | Instituto Butantan / Mendelics | Instituto Butantan: Dimas Tadeu Covas, Sandra Coccuzzo Sampaio, Maria Carolina Elias, José Salvatore Leister Patané, Vincent Louis Viala, Antonio Jorge Martins, Ricardo Haddad, Claudia Renata dos Santos Barros, Elaine Cristina Marqueze, Raul Machado Neto, Debora Botequio Moretti, Jardelina de Souza Todao Bernardino, Loyze Paola Oliveira de Lima, Luiz Aurelio de Campos Crispin. Centro de Genômica Funcional da ESALQ: Luiz Lehmann Coutinho, Ricardo Augusto Brassaloti, Raquel de Lello Rocha Campos Cassano. NGS Soluções Genômicas: Pilar Drummond Sampaio Corrêa Mariani. FZEA-USP Pirassununga: Mirele Daiana Poleti, Jessika Cristina Chagas Lesbon, Elisangela Chicaroni Mattos, Heidge Fukumasu. USP-Botucatu: Rejane Maria Tommasini Grotto, Jayme A. Souza-Neto, Guilherme Targino Valente, Patricia Akemi Assato, Felipe Allan da Silva da Costa, Bianca Cechetto Carlos. Mendelics: Bibiana Santos, João Paulo Kitajima, Erika Freitas, David Schlesinger. Hemocentro Ribeirão Preto: Simone Kashima, Evandra Strazza Rodrigues, Svetoslav Nanev Slavov, Elaine Vieira dos Santos, Rafael dos Santos Bezerra, Luiz Carlos Junior de Alcantara, Marta Giovanetti, Vagner Fonseca, Flavia Aburjaile, Rodrigo Tocantins Calado. FAMERP-SJRP: Cecília Artico Banho, Lívia Sacchetto, Fábio Sossai Possebon, Leila Sabrina Ullmann, Cíntia Bittar, Guilherme Campos, Helena Lage Ferreira, Jorge A. Petroli Marchesi, Maisa C. Pereira Parra, Marília Moraes, Paula Rahal, Paulo Inacio da Costa, João Pessoa Araújo Jr., Maurício Lacerda Nogueira. Prefeitura de Sao Paulo: Melissa Palmieri. |
| EPI_ISL_1966256 | CENTRO INTEGRADO DE SAUDE | Instituto Butantan / Mendelics | Instituto Butantan: Dimas Tadeu Covas, Sandra Coccuzzo Sampaio, Maria Carolina Elias, José Salvatore Leister Patané, Vincent Louis Viala, Antonio Jorge Martins, Ricardo Haddad, Claudia Renata dos Santos Barros, Elaine Cristina Marqueze, Raul Machado Neto, Debora Botequio Moretti, Jardelina de Souza Todao Bernardino, Loyze Paola Oliveira de Lima, Luiz Aurelio de Campos Crispin. Centro de Genômica Funcional da ESALQ: Luiz Lehmann Coutinho, Ricardo Augusto Brassaloti, Raquel de Lello Rocha Campos Cassano. NGS Soluções Genômicas: Pilar Drummond Sampaio Corrêa Mariani. FZEA-USP Pirassununga: Mirele Daiana Poleti, Jessika Cristina Chagas Lesbon, Elisangela Chicaroni Mattos, Heidge Fukumasu. USP-Botucatu: Rejane Maria Tommasini Grotto, Jayme A. Souza-Neto, Guilherme Targino Valente, Patricia Akemi Assato, Felipe Allan da Silva da Costa, Bianca Cechetto Carlos. Mendelics: Bibiana Santos, João Paulo Kitajima, Erika Freitas, David Schlesinger. Hemocentro Ribeirão Preto: Simone Kashima, Evandra Strazza Rodrigues, Svetoslav Nanev Slavov, Elaine Vieira dos Santos, Rafael dos Santos Bezerra, Luiz Carlos Junior de Alcantara, Marta Giovanetti, Vagner Fonseca, Flavia Aburjaile, Rodrigo Tocantins Calado. FAMERP-SJRP: Cecília Artico Banho, Lívia Sacchetto, Fábio Sossai Possebon, Leila Sabrina Ullmann, Cíntia Bittar, Guilherme Campos, Helena Lage Ferreira, Jorge A. Petroli Marchesi, Maisa C. Pereira Parra, Marília Moraes, Paula Rahal, Paulo Inacio da Costa, João Pessoa Araújo Jr., Maurício Lacerda Nogueira. Prefeitura de Sao Paulo: Melissa Palmieri. |
| EPI_ISL_1966257, EPI_ISL_1966258 | CS DE RUBINEIA | Instituto Butantan / Mendelics | Instituto Butantan: Dimas Tadeu Covas, Sandra Coccuzzo Sampaio, Maria Carolina Elias, José Salvatore Leister Patané, Vincent Louis Viala, Antonio Jorge Martins, Ricardo Haddad, Claudia Renata dos Santos Barros, Elaine Cristina Marqueze, Raul Machado Neto, Debora Botequio Moretti, Jardelina de Souza Todao Bernardino, Loyze Paola Oliveira de Lima, Luiz Aurelio de Campos Crispin. Centro de Genômica Funcional da ESALQ: Luiz Lehmann Coutinho, Ricardo Augusto Brassaloti, Raquel de Lello Rocha Campos Cassano. NGS Soluções Genômicas: Pilar Drummond Sampaio Corrêa Mariani. FZEA-USP Pirassununga: Mirele Daiana Poleti, Jessika Cristina Chagas Lesbon, Elisangela Chicaroni Mattos, Heidge Fukumasu. USP-Botucatu: Rejane Maria Tommasini Grotto, Jayme A. Souza-Neto, Guilherme Targino Valente, Patricia Akemi Assato, Felipe Allan da Silva da Costa, Bianca Cechetto Carlos. Mendelics: Bibiana Santos, João Paulo Kitajima, Erika Freitas, David Schlesinger. Hemocentro Ribeirão Preto: Simone Kashima, Evandra Strazza Rodrigues, Svetoslav Nanev Slavov, Elaine Vieira dos Santos, Rafael dos Santos Bezerra, Luiz Carlos Junior de Alcantara, Marta Giovanetti, Vagner Fonseca, Flavia Aburjaile, Rodrigo Tocantins Calado. FAMERP-SJRP: Cecília Artico Banho, Lívia Sacchetto, Fábio Sossai Possebon, Leila Sabrina Ullmann, Cíntia Bittar, Guilherme Campos, Helena Lage Ferreira, Jorge A. Petroli Marchesi, Maisa C. Pereira Parra, Marília Moraes, Paula Rahal, Paulo Inacio da Costa, João Pessoa Araújo Jr., Maurício Lacerda Nogueira. Prefeitura de Sao Paulo: Melissa Palmieri. |
| EPI_ISL_1966259 | SANTA CASA DE MISERICORDIA DE UBATUBA | Instituto Butantan / Mendelics | Instituto Butantan: Dimas Tadeu Covas, Sandra Coccuzzo Sampaio, Maria Carolina Elias, José Salvatore Leister Patané, Vincent Louis Viala, Antonio Jorge Martins, Ricardo Haddad, Claudia Renata dos Santos Barros, Elaine Cristina Marqueze, Raul Machado Neto, Debora Botequio Moretti, Jardelina de Souza Todao Bernardino, Loyze Paola Oliveira de Lima, Luiz Aurelio de Campos Crispin. Centro de Genômica Funcional da ESALQ: Luiz Lehmann Coutinho, Ricardo Augusto Brassaloti, Raquel de Lello Rocha Campos Cassano. NGS Soluções Genômicas: Pilar Drummond Sampaio Corrêa Mariani. |

| | | | |
|---|---|---|---|
| | | | FZEA-USP Pirassununga: Mirele Daiana Poleti, Jessika Cristina Chagas Lesbon, Elisangela Chicaroni Mattos, Heidge Fukumasu. USP-Botucatu: Rejane Maria Tommasini Grotto, Jayme A. Souza-Neto, Guilherme Targino Valente, Patricia Akemi Assato, Felipe Allan da Silva da Costa, Bianca Cechetto Carlos. Mendelics: Bibiana Santos, João Paulo Kitajima, Erika Freitas, David Schlesinger. Hemocentro Ribeirão Preto: Simone Kashima, Evandra Strazza Rodrigues, Svetoslav Nanev Slavov, Elaine Vieira dos Santos, Rafael dos Santos Bezerra, Luiz Carlos Junior de Alcantara, Marta Giovanetti, Vagner Fonseca, Flavia Aburjaile, Rodrigo Tocantins Calado. FAMERP-SJRP: Cecília Artico Banho, Lívia Sacchetto, Fábio Sossai Possebon, Leila Sabrina Ullmann, Cíntia Bittar, Guilherme Campos, Helena Lage Ferreira, Jorge A. Petroli Marchesi, Maisa C. Pereira Parra, Marília Moraes, Paula Rahal, Paulo Inacio da Costa, João Pessoa Araújo Jr., Maurício Lacerda Nogueira. Prefeitura de Sao Paulo: Melissa Palmieri. |
| EPI_ISL_1966260, EPI_ISL_1966262, EPI_ISL_1966263, EPI_ISL_1966264 | SMS SECRETARIA MUNICIPAL DE SAUDE DE BOITUVA | Instituto Butantan / Mendelics | Instituto Butantan: Dimas Tadeu Covas, Sandra Coccuzzo Sampaio, Maria Carolina Elias, José Salvatore Leister Patané, Vincent Louis Viala, Antonio Jorge Martins, Ricardo Haddad, Claudia Renata dos Santos Barros, Elaine Cristina Marqueze, Raul Machado Neto, Debora Botequio Moretti, Jardelina de Souza Todao Bernardino, Loyze Paola Oliveira de Lima, Luiz Aurelio de Campos Crispin. Centro de Genômica Funcional da ESALQ: Luiz Lehmann Coutinho, Ricardo Augusto Brassaloti, Raquel de Lello Rocha Campos Cassano. NGS Soluções Genômicas: Pilar Drummond Sampaio Corrêa Mariani. FZEA-USP Pirassununga: Mirele Daiana Poleti, Jessika Cristina Chagas Lesbon, Elisangela Chicaroni Mattos, Heidge Fukumasu. USP-Botucatu: Rejane Maria Tommasini Grotto, Jayme A. Souza-Neto, Guilherme Targino Valente, Patricia Akemi Assato, Felipe Allan da Silva da Costa, Bianca Cechetto Carlos. Mendelics: Bibiana Santos, João Paulo Kitajima, Erika Freitas, David Schlesinger. Hemocentro Ribeirão Preto: Simone Kashima, Evandra Strazza Rodrigues, Svetoslav Nanev Slavov, Elaine Vieira dos Santos, Rafael dos Santos Bezerra, Luiz Carlos Junior de Alcantara, Marta Giovanetti, Vagner Fonseca, Flavia Aburjaile, Rodrigo Tocantins Calado. FAMERP-SJRP: Cecília Artico Banho, Lívia Sacchetto, Fábio Sossai Possebon, Leila Sabrina Ullmann, Cíntia Bittar, Guilherme Campos, Helena Lage Ferreira, Jorge A. Petroli Marchesi, Maisa C. Pereira Parra, Marília Moraes, Paula Rahal, Paulo Inacio da Costa, João Pessoa Araújo Jr., Maurício Lacerda Nogueira. Prefeitura de Sao Paulo: Melissa Palmieri. |
| EPI_ISL_1966265 | UBS II DE NARANDIBA | Instituto Butantan / Mendelics | Instituto Butantan: Dimas Tadeu Covas, Sandra Coccuzzo Sampaio, Maria Carolina Elias, José Salvatore Leister Patané, Vincent Louis Viala, Antonio Jorge Martins, Ricardo Haddad, Claudia Renata dos Santos Barros, Elaine Cristina Marqueze, Raul Machado Neto, Debora Botequio Moretti, Jardelina de Souza Todao Bernardino, Loyze Paola Oliveira de Lima, Luiz Aurelio de Campos Crispin. Centro de Genômica Funcional da ESALQ: Luiz Lehmann Coutinho, Ricardo Augusto Brassaloti, Raquel de Lello Rocha Campos Cassano. NGS Soluções Genômicas: Pilar Drummond Sampaio Corrêa Mariani. FZEA-USP Pirassununga: Mirele Daiana Poleti, Jessika Cristina Chagas Lesbon, Elisangela Chicaroni Mattos, Heidge Fukumasu. USP-Botucatu: Rejane Maria Tommasini Grotto, Jayme A. Souza-Neto, Guilherme Targino Valente, Patricia Akemi Assato, Felipe Allan da Silva da Costa, Bianca Cechetto Carlos. Mendelics: Bibiana Santos, João Paulo Kitajima, Erika Freitas, David Schlesinger. Hemocentro Ribeirão Preto: Simone Kashima, Evandra Strazza Rodrigues, Svetoslav Nanev Slavov, Elaine Vieira dos Santos, Rafael dos Santos Bezerra, Luiz Carlos Junior de Alcantara, Marta Giovanetti, Vagner Fonseca, Flavia Aburjaile, Rodrigo Tocantins Calado. FAMERP-SJRP: Cecília Artico Banho, Lívia Sacchetto, Fábio Sossai Possebon, Leila Sabrina Ullmann, Cíntia Bittar, Guilherme Campos, Helena Lage Ferreira, Jorge A. Petroli Marchesi, Maisa C. Pereira Parra, Marília Moraes, Paula Rahal, Paulo Inacio da Costa, João Pessoa Araújo Jr., Maurício Lacerda Nogueira. Prefeitura de Sao Paulo: Melissa Palmieri. |
| EPI_ISL_1966266 | UBS IRMA AGUEDA MARIA JAIME | Instituto Butantan / Mendelics | Instituto Butantan: Dimas Tadeu Covas, Sandra Coccuzzo Sampaio, Maria Carolina Elias, José Salvatore Leister Patané, Vincent Louis Viala, Antonio Jorge Martins, Ricardo Haddad, Claudia Renata dos Santos Barros, Elaine Cristina Marqueze, Raul Machado Neto, Debora Botequio Moretti, Jardelina de Souza Todao Bernardino, Loyze Paola Oliveira de Lima, Luiz Aurelio de Campos Crispin. Centro de Genômica Funcional da ESALQ: Luiz Lehmann Coutinho, Ricardo Augusto Brassaloti, Raquel de Lello Rocha Campos Cassano. NGS Soluções Genômicas: Pilar Drummond Sampaio Corrêa Mariani. FZEA-USP Pirassununga: Mirele Daiana Poleti, Jessika Cristina Chagas Lesbon, Elisangela Chicaroni Mattos, Heidge Fukumasu. USP-Botucatu: Rejane Maria Tommasini Grotto, Jayme A. Souza-Neto, Guilherme Targino Valente, Patricia Akemi Assato, Felipe Allan da Silva da Costa, Bianca Cechetto Carlos. Mendelics: Bibiana Santos, João Paulo Kitajima, Erika Freitas, David Schlesinger. Hemocentro Ribeirão Preto: Simone Kashima, Evandra Strazza Rodrigues, Svetoslav Nanev Slavov, Elaine Vieira dos Santos, Rafael dos Santos Bezerra, Luiz Carlos Junior de Alcantara, Marta Giovanetti, Vagner Fonseca, Flavia Aburjaile, Rodrigo Tocantins Calado. FAMERP-SJRP: Cecília Artico Banho, Lívia Sacchetto, Fábio Sossai Possebon, Leila Sabrina Ullmann, Cíntia Bittar, Guilherme Campos, Helena Lage Ferreira, Jorge A. Petroli Marchesi, Maisa C. Pereira Parra, Marília Moraes, Paula Rahal, Paulo Inacio da Costa, João Pessoa Araújo Jr., Maurício Lacerda Nogueira. Prefeitura de Sao Paulo: Melissa Palmieri. |
| EPI_ISL_1966267, EPI_ISL_1966268, EPI_ISL_1966269 | VIGILANCIA EM SAUDE SAE VIGILANCIA EPIDEMIOLOGICA | Instituto Butantan / Mendelics | Instituto Butantan: Dimas Tadeu Covas, Sandra Coccuzzo Sampaio, Maria Carolina Elias, José Salvatore Leister Patané, Vincent Louis Viala, Antonio Jorge Martins, Ricardo Haddad, Claudia Renata dos Santos Barros, Elaine Cristina Marqueze, Raul Machado Neto, Debora Botequio Moretti, Jardelina de Souza Todao Bernardino, Loyze Paola Oliveira de Lima, Luiz Aurelio de Campos Crispin. Centro de Genômica Funcional da ESALQ: Luiz Lehmann Coutinho, Ricardo Augusto Brassaloti, Raquel de Lello Rocha Campos Cassano. NGS Soluções Genômicas: Pilar Drummond Sampaio Corrêa Mariani. FZEA-USP Pirassununga: Mirele Daiana Poleti, Jessika Cristina Chagas Lesbon, Elisangela Chicaroni Mattos, Heidge Fukumasu. USP-Botucatu: Rejane Maria Tommasini Grotto, Jayme A. Souza-Neto, Guilherme Targino Valente, Patricia Akemi Assato, Felipe Allan da Silva da Costa, Bianca Cechetto Carlos. Mendelics: Bibiana Santos, João Paulo Kitajima, Erika Freitas, David Schlesinger. Hemocentro Ribeirão Preto: Simone Kashima, Evandra Strazza Rodrigues, Svetoslav Nanev Slavov, Elaine Vieira dos Santos, Rafael dos Santos Bezerra, Luiz Carlos Junior de Alcantara, Marta Giovanetti, Vagner Fonseca, Flavia Aburjaile, Rodrigo Tocantins Calado. FAMERP-SJRP: Cecília Artico Banho, Lívia Sacchetto, Fábio Sossai Possebon, Leila Sabrina Ullmann, Cíntia Bittar, Guilherme Campos, Helena Lage Ferreira, Jorge A. Petroli Marchesi, Maisa C. Pereira Parra, Marília Moraes, Paula Rahal, Paulo Inacio da Costa, João Pessoa Araújo Jr., Maurício Lacerda Nogueira. Prefeitura de Sao Paulo: Melissa Palmieri. |
| EPI_ISL_1966270, EPI_ISL_1966271, EPI_ISL_1966272, EPI_ISL_1966273, EPI_ISL_1966274, EPI_ISL_1966275, EPI_ISL_1966276, EPI_ISL_1966277 | SECRETARIA MUNICIPAL DE SAUDE DE ANDRADINA | Instituto Butantan / Mendelics | Instituto Butantan: Dimas Tadeu Covas, Sandra Coccuzzo Sampaio, Maria Carolina Elias, José Salvatore Leister Patané, Vincent Louis Viala, Antonio Jorge Martins, Ricardo Haddad, Claudia Renata dos Santos Barros, Elaine Cristina Marqueze, Raul Machado Neto, Debora Botequio Moretti, Jardelina de Souza Todao Bernardino, Loyze Paola Oliveira de Lima, Luiz Aurelio de Campos Crispin. Centro de Genômica Funcional da ESALQ: Luiz Lehmann Coutinho, Ricardo Augusto Brassaloti, Raquel de Lello Rocha Campos Cassano. NGS Soluções Genômicas: Pilar Drummond Sampaio Corrêa Mariani. FZEA-USP Pirassununga: Mirele Daiana Poleti, Jessika Cristina Chagas Lesbon, Elisangela Chicaroni Mattos, Heidge Fukumasu. USP-Botucatu: Rejane Maria Tommasini Grotto, Jayme A. Souza-Neto, Guilherme Targino Valente, Patricia Akemi Assato, Felipe Allan da Silva da Costa, Bianca Cechetto Carlos. Mendelics: Bibiana Santos, João Paulo Kitajima, Erika Freitas, David Schlesinger. Hemocentro Ribeirão Preto: Simone Kashima, Evandra Strazza Rodrigues, Svetoslav Nanev Slavov, Elaine Vieira dos Santos, Rafael dos Santos Bezerra, Luiz Carlos Junior de Alcantara, Marta Giovanetti, Vagner Fonseca, Flavia Aburjaile, Rodrigo Tocantins Calado. FAMERP-SJRP: Cecília Artico Banho, Lívia Sacchetto, Fábio Sossai Possebon, Leila Sabrina Ullmann, Cíntia Bittar, Guilherme Campos, Helena Lage Ferreira, Jorge A. Petroli Marchesi, Maisa C. Pereira Parra, Marília Moraes, Paula Rahal, Paulo Inacio da Costa, João Pessoa Araújo Jr., Maurício Lacerda Nogueira. Prefeitura de Sao Paulo: Melissa Palmieri. |
| EPI_ISL_1966278, EPI_ISL_1966279, EPI_ISL_1966280, EPI_ISL_1966281, EPI_ISL_1966282, EPI_ISL_1966283, EPI_ISL_1966284, EPI_ISL_1966285, EPI_ISL_1966286 | UBS DR ALFREDO DANTAS DE SOUZA UMUARAMA | Instituto Butantan / Mendelics | Instituto Butantan: Dimas Tadeu Covas, Sandra Coccuzzo Sampaio, Maria Carolina Elias, José Salvatore Leister Patané, Vincent Louis Viala, Antonio Jorge Martins, Ricardo Haddad, Claudia Renata dos Santos Barros, Elaine Cristina Marqueze, Raul Machado Neto, Debora Botequio Moretti, Jardelina de Souza Todao Bernardino, Loyze Paola Oliveira de Lima, Luiz Aurelio de Campos Crispin. Centro de Genômica Funcional da ESALQ: Luiz Lehmann Coutinho, Ricardo Augusto Brassaloti, Raquel de Lello Rocha Campos Cassano. NGS Soluções Genômicas: Pilar Drummond Sampaio Corrêa Mariani. FZEA-USP Pirassununga: Mirele Daiana Poleti, Jessika Cristina Chagas Lesbon, Elisangela Chicaroni Mattos, Heidge Fukumasu. USP-Botucatu: Rejane Maria Tommasini Grotto, Jayme A. Souza-Neto, Guilherme Targino Valente, Patricia Akemi Assato, Felipe Allan da Silva da Costa, Bianca Cechetto Carlos. Mendelics: Bibiana Santos, João Paulo Kitajima, Erika Freitas, David Schlesinger. Hemocentro Ribeirão Preto: Simone Kashima, Evandra Strazza Rodrigues, Svetoslav Nanev Slavov, Elaine Vieira dos Santos, Rafael dos Santos Bezerra, Luiz Carlos Junior de Alcantara, Marta Giovanetti, Vagner Fonseca, Flavia Aburjaile, Rodrigo Tocantins Calado. FAMERP-SJRP: Cecília Artico Banho, Lívia Sacchetto, Fábio Sossai Possebon, Leila Sabrina Ullmann, Cíntia Bittar, Guilherme Campos, Helena Lage Ferreira, Jorge A. Petroli Marchesi, Maisa C. Pereira Parra, Marília Moraes, Paula Rahal, Paulo Inacio da Costa, João Pessoa Araújo Jr., Maurício Lacerda Nogueira. Prefeitura de Sao Paulo: Melissa Palmieri. |
| EPI_ISL_1966287 | UBS DE AURIFLAMA | Instituto Butantan / Mendelics | Instituto Butantan: Dimas Tadeu Covas, Sandra Coccuzzo Sampaio, Maria Carolina Elias, José Salvatore Leister Patané, Vincent Louis Viala, Antonio Jorge Martins, Ricardo Haddad, Claudia Renata dos Santos Barros, Elaine Cristina Marqueze, Raul Machado Neto, Debora Botequio Moretti, Jardelina de Souza Todao Bernardino, Loyze Paola Oliveira de Lima, Luiz Aurelio de Campos Crispin. Centro de Genômica Funcional da ESALQ: Luiz Lehmann Coutinho, Ricardo Augusto Brassaloti, Raquel de Lello Rocha Campos Cassano. NGS Soluções Genômicas: Pilar Drummond Sampaio Corrêa Mariani. FZEA-USP Pirassununga: Mirele Daiana Poleti, Jessika Cristina Chagas Lesbon, Elisangela Chicaroni Mattos, Heidge Fukumasu. USP-Botucatu: Rejane Maria Tommasini Grotto, Jayme A. Souza-Neto, Guilherme Targino Valente, Patricia Akemi Assato, Felipe Allan da Silva da Costa, Bianca Cechetto Carlos. Mendelics: Bibiana Santos, João Paulo Kitajima, Erika Freitas, David Schlesinger. Hemocentro Ribeirão Preto: Simone Kashima, Evandra Strazza Rodrigues, Svetoslav Nanev Slavov, Elaine Vieira dos Santos, Rafael dos Santos Bezerra, Luiz Carlos Junior de Alcantara, Marta Giovanetti, Vagner Fonseca, Flavia Aburjaile, Rodrigo Tocantins Calado. FAMERP-SJRP: Cecília Artico Banho, Lívia Sacchetto, Fábio Sossai Possebon, Leila Sabrina Ullmann, Cíntia Bittar, Guilherme Campos, Helena Lage Ferreira, Jorge A. Petroli Marchesi, Maisa C. Pereira Parra, Marília Moraes, Paula Rahal, Paulo Inacio da Costa, João Pessoa Araújo Jr., Maurício Lacerda Nogueira. Prefeitura de Sao Paulo: Melissa Palmieri. |

| | | | |
|---|---|---|---|
| EPI_ISL_1966288 | UNIDADE BASICA DE SAUDE DR BARTOLOMEU MARAGLIANO VENERE | Instituto Butantan / Mendelics | Instituto Butantan: Dimas Tadeu Covas, Sandra Coccuzzo Sampaio, Maria Carolina Elias, José Salvatore Leister Patané, Vincent Louis Viala, Antonio Jorge Martins, Ricardo Haddad, Claudia Renata dos Santos Barros, Elaine Cristina Marqueze, Raul Machado Neto, Debora Botequio Moretti, Jardelina de Souza Todao Bernardino, Loyze Paola Oliveira de Lima, Luiz Aurelio de Campos Crispin. Centro de Genômica Funcional da ESALQ: Luiz Lehmann Coutinho, Ricardo Augusto Brassaloti, Raquel de Lello Rocha Campos Cassano. NGS Soluções Genômicas: Pilar Drummond Sampaio Corrêa Mariani. FZEA-USP Pirassununga: Mirele Daiana Poleti, Jessika Cristina Chagas Lesbon, Elisangela Chicaroni Mattos, Heidge Fukumasu. USP-Botucatu: Rejane Maria Tommasini Grotto, Jayme A. Souza-Neto, Guilherme Targino Valente, Patricia Akemi Assato, Felipe Allan da Silva da Costa, Bianca Cechetto Carlos. Mendelics: Bibiana Santos, João Paulo Kitajima, Erika Freitas, David Schlesinger. Hemocentro Ribeirão Preto: Simone Kashima, Evandra Strazza Rodrigues, Svetoslav Nanev Slavov, Elaine Vieira dos Santos, Rafael dos Santos Bezerra, Luiz Carlos Junior de Alcantara, Marta Giovanetti, Vagner Fonseca, Flavia Aburjaile, Rodrigo Tocantins Calado. FAMERP-SJRP: Cecília Artico Banho, Lívia Sacchetto, Fábio Sossai Possebon, Leila Sabrina Ullmann, Cíntia Bittar, Guilherme Campos, Helena Lage Ferreira, Jorge A. Petroli Marchesi, Maisa C. Pereira Parra, Marília Moraes, Paula Rahal, Paulo Inacio da Costa, João Pessoa Araújo Jr., Maurício Lacerda Nogueira. Prefeitura de Sao Paulo: Melissa Palmieri. |
| EPI_ISL_1966289 | UBS DR ALLY ALAHMAR DE BARRETOS | Instituto Butantan / Mendelics | Instituto Butantan: Dimas Tadeu Covas, Sandra Coccuzzo Sampaio, Maria Carolina Elias, José Salvatore Leister Patané, Vincent Louis Viala, Antonio Jorge Martins, Ricardo Haddad, Claudia Renata dos Santos Barros, Elaine Cristina Marqueze, Raul Machado Neto, Debora Botequio Moretti, Jardelina de Souza Todao Bernardino, Loyze Paola Oliveira de Lima, Luiz Aurelio de Campos Crispin. Centro de Genômica Funcional da ESALQ: Luiz Lehmann Coutinho, Ricardo Augusto Brassaloti, Raquel de Lello Rocha Campos Cassano. NGS Soluções Genômicas: Pilar Drummond Sampaio Corrêa Mariani. FZEA-USP Pirassununga: Mirele Daiana Poleti, Jessika Cristina Chagas Lesbon, Elisangela Chicaroni Mattos, Heidge Fukumasu. USP-Botucatu: Rejane Maria Tommasini Grotto, Jayme A. Souza-Neto, Guilherme Targino Valente, Patricia Akemi Assato, Felipe Allan da Silva da Costa, Bianca Cechetto Carlos. Mendelics: Bibiana Santos, João Paulo Kitajima, Erika Freitas, David Schlesinger. Hemocentro Ribeirão Preto: Simone Kashima, Evandra Strazza Rodrigues, Svetoslav Nanev Slavov, Elaine Vieira dos Santos, Rafael dos Santos Bezerra, Luiz Carlos Junior de Alcantara, Marta Giovanetti, Vagner Fonseca, Flavia Aburjaile, Rodrigo Tocantins Calado. FAMERP-SJRP: Cecília Artico Banho, Lívia Sacchetto, Fábio Sossai Possebon, Leila Sabrina Ullmann, Cíntia Bittar, Guilherme Campos, Helena Lage Ferreira, Jorge A. Petroli Marchesi, Maisa C. Pereira Parra, Marília Moraes, Paula Rahal, Paulo Inacio da Costa, João Pessoa Araújo Jr., Maurício Lacerda Nogueira. Prefeitura de Sao Paulo: Melissa Palmieri. |
| EPI_ISL_1966290, EPI_ISL_1966291 | UNIDADE BASICA DE SAUDE DA FAMILIA BILAC | Instituto Butantan / Mendelics | Instituto Butantan: Dimas Tadeu Covas, Sandra Coccuzzo Sampaio, Maria Carolina Elias, José Salvatore Leister Patané, Vincent Louis Viala, Antonio Jorge Martins, Ricardo Haddad, Claudia Renata dos Santos Barros, Elaine Cristina Marqueze, Raul Machado Neto, Debora Botequio Moretti, Jardelina de Souza Todao Bernardino, Loyze Paola Oliveira de Lima, Luiz Aurelio de Campos Crispin. Centro de Genômica Funcional da ESALQ: Luiz Lehmann Coutinho, Ricardo Augusto Brassaloti, Raquel de Lello Rocha Campos Cassano. NGS Soluções Genômicas: Pilar Drummond Sampaio Corrêa Mariani. FZEA-USP Pirassununga: Mirele Daiana Poleti, Jessika Cristina Chagas Lesbon, Elisangela Chicaroni Mattos, Heidge Fukumasu. USP-Botucatu: Rejane Maria Tommasini Grotto, Jayme A. Souza-Neto, Guilherme Targino Valente, Patricia Akemi Assato, Felipe Allan da Silva da Costa, Bianca Cechetto Carlos. Mendelics: Bibiana Santos, João Paulo Kitajima, Erika Freitas, David Schlesinger. Hemocentro Ribeirão Preto: Simone Kashima, Evandra Strazza Rodrigues, Svetoslav Nanev Slavov, Elaine Vieira dos Santos, Rafael dos Santos Bezerra, Luiz Carlos Junior de Alcantara, Marta Giovanetti, Vagner Fonseca, Flavia Aburjaile, Rodrigo Tocantins Calado. FAMERP-SJRP: Cecília Artico Banho, Lívia Sacchetto, Fábio Sossai Possebon, Leila Sabrina Ullmann, Cíntia Bittar, Guilherme Campos, Helena Lage Ferreira, Jorge A. Petroli Marchesi, Maisa C. Pereira Parra, Marília Moraes, Paula Rahal, Paulo Inacio da Costa, João Pessoa Araújo Jr., Maurício Lacerda Nogueira. Prefeitura de Sao Paulo: Melissa Palmieri. |
| EPI_ISL_1966292 | CENTRO DE SAUDE DE FLORINEA | Instituto Butantan / Mendelics | Instituto Butantan: Dimas Tadeu Covas, Sandra Coccuzzo Sampaio, Maria Carolina Elias, José Salvatore Leister Patané, Vincent Louis Viala, Antonio Jorge Martins, Ricardo Haddad, Claudia Renata dos Santos Barros, Elaine Cristina Marqueze, Raul Machado Neto, Debora Botequio Moretti, Jardelina de Souza Todao Bernardino, Loyze Paola Oliveira de Lima, Luiz Aurelio de Campos Crispin. Centro de Genômica Funcional da ESALQ: Luiz Lehmann Coutinho, Ricardo Augusto Brassaloti, Raquel de Lello Rocha Campos Cassano. NGS Soluções Genômicas: Pilar Drummond Sampaio Corrêa Mariani. FZEA-USP Pirassununga: Mirele Daiana Poleti, Jessika Cristina Chagas Lesbon, Elisangela Chicaroni Mattos, Heidge Fukumasu. USP-Botucatu: Rejane Maria Tommasini Grotto, Jayme A. Souza-Neto, Guilherme Targino Valente, Patricia Akemi Assato, Felipe Allan da Silva da Costa, Bianca Cechetto Carlos. Mendelics: Bibiana Santos, João Paulo Kitajima, Erika Freitas, David Schlesinger. Hemocentro Ribeirão Preto: Simone Kashima, Evandra Strazza Rodrigues, Svetoslav Nanev Slavov, Elaine Vieira dos Santos, Rafael dos Santos Bezerra, Luiz Carlos Junior de Alcantara, Marta Giovanetti, Vagner Fonseca, Flavia Aburjaile, Rodrigo Tocantins Calado. FAMERP-SJRP: Cecília Artico Banho, Lívia Sacchetto, Fábio Sossai Possebon, Leila Sabrina Ullmann, Cíntia Bittar, Guilherme Campos, Helena Lage Ferreira, Jorge A. Petroli Marchesi, Maisa C. Pereira Parra, Marília Moraes, Paula Rahal, Paulo Inacio da Costa, João Pessoa Araújo Jr., Maurício Lacerda Nogueira. Prefeitura de Sao Paulo: Melissa Palmieri. |
| EPI_ISL_1966293 | CENTRO DE SAUDE III GLICERIO GIMAIEL ELIAS GIMAIEL | Instituto Butantan / Mendelics | Instituto Butantan: Dimas Tadeu Covas, Sandra Coccuzzo Sampaio, Maria Carolina Elias, José Salvatore Leister Patané, Vincent Louis Viala, Antonio Jorge Martins, Ricardo Haddad, Claudia Renata dos Santos Barros, Elaine Cristina Marqueze, Raul Machado Neto, Debora Botequio Moretti, Jardelina de Souza Todao Bernardino, Loyze Paola Oliveira de Lima, Luiz Aurelio de Campos Crispin. Centro de Genômica Funcional da ESALQ: Luiz Lehmann Coutinho, Ricardo Augusto Brassaloti, Raquel de Lello Rocha Campos Cassano. NGS Soluções Genômicas: Pilar Drummond Sampaio Corrêa Mariani. FZEA-USP Pirassununga: Mirele Daiana Poleti, Jessika Cristina Chagas Lesbon, Elisangela Chicaroni Mattos, Heidge Fukumasu. USP-Botucatu: Rejane Maria Tommasini Grotto, Jayme A. Souza-Neto, Guilherme Targino Valente, Patricia Akemi Assato, Felipe Allan da Silva da Costa, Bianca Cechetto Carlos. Mendelics: Bibiana Santos, João Paulo Kitajima, Erika Freitas, David Schlesinger. Hemocentro Ribeirão Preto: Simone Kashima, Evandra Strazza Rodrigues, Svetoslav Nanev Slavov, Elaine Vieira dos Santos, Rafael dos Santos Bezerra, Luiz Carlos Junior de Alcantara, Marta Giovanetti, Vagner Fonseca, Flavia Aburjaile, Rodrigo Tocantins Calado. FAMERP-SJRP: Cecília Artico Banho, Lívia Sacchetto, Fábio Sossai Possebon, Leila Sabrina Ullmann, Cíntia Bittar, Guilherme Campos, Helena Lage Ferreira, Jorge A. Petroli Marchesi, Maisa C. Pereira Parra, Marília Moraes, Paula Rahal, Paulo Inacio da Costa, João Pessoa Araújo Jr., Maurício Lacerda Nogueira. Prefeitura de Sao Paulo: Melissa Palmieri. |
| EPI_ISL_1966294, EPI_ISL_1966295, EPI_ISL_1966296, EPI_ISL_1966297, EPI_ISL_1966298, EPI_ISL_1966299, EPI_ISL_1966300, EPI_ISL_1966301 | VIGILANCIA EPIDEMIOLOGICA DE IBATE | Instituto Butantan / Mendelics | Instituto Butantan: Dimas Tadeu Covas, Sandra Coccuzzo Sampaio, Maria Carolina Elias, José Salvatore Leister Patané, Vincent Louis Viala, Antonio Jorge Martins, Ricardo Haddad, Claudia Renata dos Santos Barros, Elaine Cristina Marqueze, Raul Machado Neto, Debora Botequio Moretti, Jardelina de Souza Todao Bernardino, Loyze Paola Oliveira de Lima, Luiz Aurelio de Campos Crispin. Centro de Genômica Funcional da ESALQ: Luiz Lehmann Coutinho, Ricardo Augusto Brassaloti, Raquel de Lello Rocha Campos Cassano. NGS Soluções Genômicas: Pilar Drummond Sampaio Corrêa Mariani. FZEA-USP Pirassununga: Mirele Daiana Poleti, Jessika Cristina Chagas Lesbon, Elisangela Chicaroni Mattos, Heidge Fukumasu. USP-Botucatu: Rejane Maria Tommasini Grotto, Jayme A. Souza-Neto, Guilherme Targino Valente, Patricia Akemi Assato, Felipe Allan da Silva da Costa, Bianca Cechetto Carlos. Mendelics: Bibiana Santos, João Paulo Kitajima, Erika Freitas, David Schlesinger. Hemocentro Ribeirão Preto: Simone Kashima, Evandra Strazza Rodrigues, Svetoslav Nanev Slavov, Elaine Vieira dos Santos, Rafael dos Santos Bezerra, Luiz Carlos Junior de Alcantara, Marta Giovanetti, Vagner Fonseca, Flavia Aburjaile, Rodrigo Tocantins Calado. FAMERP-SJRP: Cecília Artico Banho, Lívia Sacchetto, Fábio Sossai Possebon, Leila Sabrina Ullmann, Cíntia Bittar, Guilherme Campos, Helena Lage Ferreira, Jorge A. Petroli Marchesi, Maisa C. Pereira Parra, Marília Moraes, Paula Rahal, Paulo Inacio da Costa, João Pessoa Araújo Jr., Maurício Lacerda Nogueira. Prefeitura de Sao Paulo: Melissa Palmieri. |
| EPI_ISL_1966302, EPI_ISL_1966303, EPI_ISL_1966304, EPI_ISL_1966305 | PRONTO SOCORRO DE AGENOR DE CAMPOS MONGAGUA | Instituto Butantan / Mendelics | Instituto Butantan: Dimas Tadeu Covas, Sandra Coccuzzo Sampaio, Maria Carolina Elias, José Salvatore Leister Patané, Vincent Louis Viala, Antonio Jorge Martins, Ricardo Haddad, Claudia Renata dos Santos Barros, Elaine Cristina Marqueze, Raul Machado Neto, Debora Botequio Moretti, Jardelina de Souza Todao Bernardino, Loyze Paola Oliveira de Lima, Luiz Aurelio de Campos Crispin. Centro de Genômica Funcional da ESALQ: Luiz Lehmann Coutinho, Ricardo Augusto Brassaloti, Raquel de Lello Rocha Campos Cassano. NGS Soluções Genômicas: Pilar Drummond Sampaio Corrêa Mariani. FZEA-USP Pirassununga: Mirele Daiana Poleti, Jessika Cristina Chagas Lesbon, Elisangela Chicaroni Mattos, Heidge Fukumasu. USP-Botucatu: Rejane Maria Tommasini Grotto, Jayme A. Souza-Neto, Guilherme Targino Valente, Patricia Akemi Assato, Felipe Allan da Silva da Costa, Bianca Cechetto Carlos. Mendelics: Bibiana Santos, João Paulo Kitajima, Erika Freitas, David Schlesinger. Hemocentro Ribeirão Preto: Simone Kashima, Evandra Strazza Rodrigues, Svetoslav Nanev Slavov, Elaine Vieira dos Santos, Rafael dos Santos Bezerra, Luiz Carlos Junior de Alcantara, Marta Giovanetti, Vagner Fonseca, Flavia Aburjaile, Rodrigo Tocantins Calado. FAMERP-SJRP: Cecília Artico Banho, Lívia Sacchetto, Fábio Sossai Possebon, Leila Sabrina Ullmann, Cíntia Bittar, Guilherme Campos, Helena Lage Ferreira, Jorge A. Petroli Marchesi, Maisa C. Pereira Parra, Marília Moraes, Paula Rahal, Paulo Inacio da Costa, João Pessoa Araújo Jr., Maurício Lacerda Nogueira. Prefeitura de Sao Paulo: Melissa Palmieri. |
| EPI_ISL_1966306, EPI_ISL_1966307 | UNIDADE DE PRONTO ATENDIMENTO DE AGENOR DE CAMPOS | Instituto Butantan / Mendelics | Instituto Butantan: Dimas Tadeu Covas, Sandra Coccuzzo Sampaio, Maria Carolina Elias, José Salvatore Leister Patané, Vincent Louis Viala, Antonio Jorge Martins, Ricardo Haddad, Claudia Renata dos Santos Barros, Elaine Cristina Marqueze, Raul Machado Neto, Debora Botequio Moretti, Jardelina de Souza Todao Bernardino, Loyze Paola Oliveira de Lima, Luiz Aurelio de Campos Crispin. Centro de Genômica Funcional da ESALQ: Luiz Lehmann Coutinho, Ricardo Augusto Brassaloti, Raquel de Lello Rocha Campos Cassano. NGS Soluções Genômicas: Pilar Drummond Sampaio Corrêa Mariani. FZEA-USP Pirassununga: Mirele Daiana Poleti, Jessika Cristina Chagas Lesbon, Elisangela Chicaroni Mattos, Heidge Fukumasu. USP-Botucatu: Rejane Maria Tommasini Grotto, Jayme A. Souza-Neto, Guilherme Targino Valente, Patricia Akemi Assato, Felipe Allan da Silva da Costa, Bianca Cechetto Carlos. Mendelics: Bibiana Santos, João Paulo Kitajima, Erika Freitas, David Schlesinger. Hemocentro Ribeirão Preto: Simone Kashima, Evandra Strazza |

| | | | |
|---|---|---|---|
| | | | Rodrigues, Svetoslav Nanev Slavov, Elaine Vieira dos Santos, Rafael dos Santos Bezerra, Luiz Carlos Junior de Alcantara, Marta Giovanetti, Vagner Fonseca, Flavia Aburjaile, Rodrigo Tocantins Calado. FAMERP-SJRP: Cecília Artico Banho, Lívia Sacchetto, Fábio Sossai Possebon, Leila Sabrina Ullmann, Cíntia Bittar, Guilherme Campos, Helena Lage Ferreira, Jorge A. Petroli Marchesi, Maisa C. Pereira Parra, Marília Moraes, Paula Rahal, Paulo Inacio da Costa, João Pessoa Araújo Jr., Maurício Lacerda Nogueira. Prefeitura de Sao Paulo: Melissa Palmieri. |
| EPI_ISL_1966308 | PRONTO SOCORRO DE AGENOR DE CAMPOS MONGAGUA | Instituto Butantan / Mendelics | Instituto Butantan: Dimas Tadeu Covas, Sandra Coccuzzo Sampaio, Maria Carolina Elias, José Salvatore Leister Patané, Vincent Louis Viala, Antonio Jorge Martins, Ricardo Haddad, Claudia Renata dos Santos Barros, Elaine Cristina Marqueze, Raul Machado Neto, Debora Botequio Moretti, Jardelina de Souza Todao Bernardino, Loyze Paola Oliveira de Lima, Luiz Aurelio de Campos Crispin. Centro de Genômica Funcional da ESALQ: Luiz Lehmann Coutinho, Ricardo Augusto Brassaloti, Raquel de Lello Rocha Campos Cassano. NGS Soluções Genômicas: Pilar Drummond Sampaio Corrêa Mariani. FZEA-USP Pirassununga: Mirele Daiana Poleti, Jessika Cristina Chagas Lesbon, Elisangela Chicaroni Mattos, Heidge Fukumasu. USP-Botucatu: Rejane Maria Tommasini Grotto, Jayme A. Souza-Neto, Guilherme Targino Valente, Patricia Akemi Assato, Felipe Allan da Silva da Costa, Bianca Cechetto Carlos. Mendelics: Bibiana Santos, João Paulo Kitajima, Erika Freitas, David Schlesinger. Hemocentro Ribeirão Preto: Simone Kashima, Evandra Strazza Rodrigues, Svetoslav Nanev Slavov, Elaine Vieira dos Santos, Rafael dos Santos Bezerra, Luiz Carlos Junior de Alcantara, Marta Giovanetti, Vagner Fonseca, Flavia Aburjaile, Rodrigo Tocantins Calado. FAMERP-SJRP: Cecília Artico Banho, Lívia Sacchetto, Fábio Sossai Possebon, Leila Sabrina Ullmann, Cíntia Bittar, Guilherme Campos, Helena Lage Ferreira, Jorge A. Petroli Marchesi, Maisa C. Pereira Parra, Marília Moraes, Paula Rahal, Paulo Inacio da Costa, João Pessoa Araújo Jr., Maurício Lacerda Nogueira. Prefeitura de Sao Paulo: Melissa Palmieri. |
| EPI_ISL_1966310 | UNIDADE DE PRONTO ATENDIMENTO DE AGENOR DE CAMPOS | Instituto Butantan / Mendelics | Instituto Butantan: Dimas Tadeu Covas, Sandra Coccuzzo Sampaio, Maria Carolina Elias, José Salvatore Leister Patané, Vincent Louis Viala, Antonio Jorge Martins, Ricardo Haddad, Claudia Renata dos Santos Barros, Elaine Cristina Marqueze, Raul Machado Neto, Debora Botequio Moretti, Jardelina de Souza Todao Bernardino, Loyze Paola Oliveira de Lima, Luiz Aurelio de Campos Crispin. Centro de Genômica Funcional da ESALQ: Luiz Lehmann Coutinho, Ricardo Augusto Brassaloti, Raquel de Lello Rocha Campos Cassano. NGS Soluções Genômicas: Pilar Drummond Sampaio Corrêa Mariani. FZEA-USP Pirassununga: Mirele Daiana Poleti, Jessika Cristina Chagas Lesbon, Elisangela Chicaroni Mattos, Heidge Fukumasu. USP-Botucatu: Rejane Maria Tommasini Grotto, Jayme A. Souza-Neto, Guilherme Targino Valente, Patricia Akemi Assato, Felipe Allan da Silva da Costa, Bianca Cechetto Carlos. Mendelics: Bibiana Santos, João Paulo Kitajima, Erika Freitas, David Schlesinger. Hemocentro Ribeirão Preto: Simone Kashima, Evandra Strazza Rodrigues, Svetoslav Nanev Slavov, Elaine Vieira dos Santos, Rafael dos Santos Bezerra, Luiz Carlos Junior de Alcantara, Marta Giovanetti, Vagner Fonseca, Flavia Aburjaile, Rodrigo Tocantins Calado. FAMERP-SJRP: Cecília Artico Banho, Lívia Sacchetto, Fábio Sossai Possebon, Leila Sabrina Ullmann, Cíntia Bittar, Guilherme Campos, Helena Lage Ferreira, Jorge A. Petroli Marchesi, Maisa C. Pereira Parra, Marília Moraes, Paula Rahal, Paulo Inacio da Costa, João Pessoa Araújo Jr., Maurício Lacerda Nogueira. Prefeitura de Sao Paulo: Melissa Palmieri. |
| EPI_ISL_1966311, EPI_ISL_1966312, EPI_ISL_1966313, EPI_ISL_1966314, EPI_ISL_1966315 | PRONTO SOCORRO DE AGENOR DE CAMPOS MONGAGUA | Instituto Butantan / Mendelics | Instituto Butantan: Dimas Tadeu Covas, Sandra Coccuzzo Sampaio, Maria Carolina Elias, José Salvatore Leister Patané, Vincent Louis Viala, Antonio Jorge Martins, Ricardo Haddad, Claudia Renata dos Santos Barros, Elaine Cristina Marqueze, Raul Machado Neto, Debora Botequio Moretti, Jardelina de Souza Todao Bernardino, Loyze Paola Oliveira de Lima, Luiz Aurelio de Campos Crispin. Centro de Genômica Funcional da ESALQ: Luiz Lehmann Coutinho, Ricardo Augusto Brassaloti, Raquel de Lello Rocha Campos Cassano. NGS Soluções Genômicas: Pilar Drummond Sampaio Corrêa Mariani. FZEA-USP Pirassununga: Mirele Daiana Poleti, Jessika Cristina Chagas Lesbon, Elisangela Chicaroni Mattos, Heidge Fukumasu. USP-Botucatu: Rejane Maria Tommasini Grotto, Jayme A. Souza-Neto, Guilherme Targino Valente, Patricia Akemi Assato, Felipe Allan da Silva da Costa, Bianca Cechetto Carlos. Mendelics: Bibiana Santos, João Paulo Kitajima, Erika Freitas, David Schlesinger. Hemocentro Ribeirão Preto: Simone Kashima, Evandra Strazza Rodrigues, Svetoslav Nanev Slavov, Elaine Vieira dos Santos, Rafael dos Santos Bezerra, Luiz Carlos Junior de Alcantara, Marta Giovanetti, Vagner Fonseca, Flavia Aburjaile, Rodrigo Tocantins Calado. FAMERP-SJRP: Cecília Artico Banho, Lívia Sacchetto, Fábio Sossai Possebon, Leila Sabrina Ullmann, Cíntia Bittar, Guilherme Campos, Helena Lage Ferreira, Jorge A. Petroli Marchesi, Maisa C. Pereira Parra, Marília Moraes, Paula Rahal, Paulo Inacio da Costa, João Pessoa Araújo Jr., Maurício Lacerda Nogueira. Prefeitura de Sao Paulo: Melissa Palmieri. |
| EPI_ISL_1966316 | UNIDADE DE PRONTO ATENDIMENTO DE AGENOR DE CAMPOS | Instituto Butantan / Mendelics | Instituto Butantan: Dimas Tadeu Covas, Sandra Coccuzzo Sampaio, Maria Carolina Elias, José Salvatore Leister Patané, Vincent Louis Viala, Antonio Jorge Martins, Ricardo Haddad, Claudia Renata dos Santos Barros, Elaine Cristina Marqueze, Raul Machado Neto, Debora Botequio Moretti, Jardelina de Souza Todao Bernardino, Loyze Paola Oliveira de Lima, Luiz Aurelio de Campos Crispin. Centro de Genômica Funcional da ESALQ: Luiz Lehmann Coutinho, Ricardo Augusto Brassaloti, Raquel de Lello Rocha Campos Cassano. NGS Soluções Genômicas: Pilar Drummond Sampaio Corrêa Mariani. FZEA-USP Pirassununga: Mirele Daiana Poleti, Jessika Cristina Chagas Lesbon, Elisangela Chicaroni Mattos, Heidge Fukumasu. USP-Botucatu: Rejane Maria Tommasini Grotto, Jayme A. Souza-Neto, Guilherme Targino Valente, Patricia Akemi Assato, Felipe Allan da Silva da Costa, Bianca Cechetto Carlos. Mendelics: Bibiana Santos, João Paulo Kitajima, Erika Freitas, David Schlesinger. Hemocentro Ribeirão Preto: Simone Kashima, Evandra Strazza Rodrigues, Svetoslav Nanev Slavov, Elaine Vieira dos Santos, Rafael dos Santos Bezerra, Luiz Carlos Junior de Alcantara, Marta Giovanetti, Vagner Fonseca, Flavia Aburjaile, Rodrigo Tocantins Calado. FAMERP-SJRP: Cecília Artico Banho, Lívia Sacchetto, Fábio Sossai Possebon, Leila Sabrina Ullmann, Cíntia Bittar, Guilherme Campos, Helena Lage Ferreira, Jorge A. Petroli Marchesi, Maisa C. Pereira Parra, Marília Moraes, Paula Rahal, Paulo Inacio da Costa, João Pessoa Araújo Jr., Maurício Lacerda Nogueira. Prefeitura de Sao Paulo: Melissa Palmieri. |
| EPI_ISL_1966317, EPI_ISL_1966318 | PRONTO SOCORRO DE AGENOR DE CAMPOS MONGAGUA | Instituto Butantan / Mendelics | Instituto Butantan: Dimas Tadeu Covas, Sandra Coccuzzo Sampaio, Maria Carolina Elias, José Salvatore Leister Patané, Vincent Louis Viala, Antonio Jorge Martins, Ricardo Haddad, Claudia Renata dos Santos Barros, Elaine Cristina Marqueze, Raul Machado Neto, Debora Botequio Moretti, Jardelina de Souza Todao Bernardino, Loyze Paola Oliveira de Lima, Luiz Aurelio de Campos Crispin. Centro de Genômica Funcional da ESALQ: Luiz Lehmann Coutinho, Ricardo Augusto Brassaloti, Raquel de Lello Rocha Campos Cassano. NGS Soluções Genômicas: Pilar Drummond Sampaio Corrêa Mariani. FZEA-USP Pirassununga: Mirele Daiana Poleti, Jessika Cristina Chagas Lesbon, Elisangela Chicaroni Mattos, Heidge Fukumasu. USP-Botucatu: Rejane Maria Tommasini Grotto, Jayme A. Souza-Neto, Guilherme Targino Valente, Patricia Akemi Assato, Felipe Allan da Silva da Costa, Bianca Cechetto Carlos. Mendelics: Bibiana Santos, João Paulo Kitajima, Erika Freitas, David Schlesinger. Hemocentro Ribeirão Preto: Simone Kashima, Evandra Strazza Rodrigues, Svetoslav Nanev Slavov, Elaine Vieira dos Santos, Rafael dos Santos Bezerra, Luiz Carlos Junior de Alcantara, Marta Giovanetti, Vagner Fonseca, Flavia Aburjaile, Rodrigo Tocantins Calado. FAMERP-SJRP: Cecília Artico Banho, Lívia Sacchetto, Fábio Sossai Possebon, Leila Sabrina Ullmann, Cíntia Bittar, Guilherme Campos, Helena Lage Ferreira, Jorge A. Petroli Marchesi, Maisa C. Pereira Parra, Marília Moraes, Paula Rahal, Paulo Inacio da Costa, João Pessoa Araújo Jr., Maurício Lacerda Nogueira. Prefeitura de Sao Paulo: Melissa Palmieri. |
| EPI_ISL_1966319 | UNIDADE DE PRONTO ATENDIMENTO DE AGENOR DE CAMPOS | Instituto Butantan / Mendelics | Instituto Butantan: Dimas Tadeu Covas, Sandra Coccuzzo Sampaio, Maria Carolina Elias, José Salvatore Leister Patané, Vincent Louis Viala, Antonio Jorge Martins, Ricardo Haddad, Claudia Renata dos Santos Barros, Elaine Cristina Marqueze, Raul Machado Neto, Debora Botequio Moretti, Jardelina de Souza Todao Bernardino, Loyze Paola Oliveira de Lima, Luiz Aurelio de Campos Crispin. Centro de Genômica Funcional da ESALQ: Luiz Lehmann Coutinho, Ricardo Augusto Brassaloti, Raquel de Lello Rocha Campos Cassano. NGS Soluções Genômicas: Pilar Drummond Sampaio Corrêa Mariani. FZEA-USP Pirassununga: Mirele Daiana Poleti, Jessika Cristina Chagas Lesbon, Elisangela Chicaroni Mattos, Heidge Fukumasu. USP-Botucatu: Rejane Maria Tommasini Grotto, Jayme A. Souza-Neto, Guilherme Targino Valente, Patricia Akemi Assato, Felipe Allan da Silva da Costa, Bianca Cechetto Carlos. Mendelics: Bibiana Santos, João Paulo Kitajima, Erika Freitas, David Schlesinger. Hemocentro Ribeirão Preto: Simone Kashima, Evandra Strazza Rodrigues, Svetoslav Nanev Slavov, Elaine Vieira dos Santos, Rafael dos Santos Bezerra, Luiz Carlos Junior de Alcantara, Marta Giovanetti, Vagner Fonseca, Flavia Aburjaile, Rodrigo Tocantins Calado. FAMERP-SJRP: Cecília Artico Banho, Lívia Sacchetto, Fábio Sossai Possebon, Leila Sabrina Ullmann, Cíntia Bittar, Guilherme Campos, Helena Lage Ferreira, Jorge A. Petroli Marchesi, Maisa C. Pereira Parra, Marília Moraes, Paula Rahal, Paulo Inacio da Costa, João Pessoa Araújo Jr., Maurício Lacerda Nogueira. Prefeitura de Sao Paulo: Melissa Palmieri. |
| EPI_ISL_1966320 | PRONTO SOCORRO DE AGENOR DE CAMPOS MONGAGUA | Instituto Butantan / Mendelics | Instituto Butantan: Dimas Tadeu Covas, Sandra Coccuzzo Sampaio, Maria Carolina Elias, José Salvatore Leister Patané, Vincent Louis Viala, Antonio Jorge Martins, Ricardo Haddad, Claudia Renata dos Santos Barros, Elaine Cristina Marqueze, Raul Machado Neto, Debora Botequio Moretti, Jardelina de Souza Todao Bernardino, Loyze Paola Oliveira de Lima, Luiz Aurelio de Campos Crispin. Centro de Genômica Funcional da ESALQ: Luiz Lehmann Coutinho, Ricardo Augusto Brassaloti, Raquel de Lello Rocha Campos Cassano. NGS Soluções Genômicas: Pilar Drummond Sampaio Corrêa Mariani. FZEA-USP Pirassununga: Mirele Daiana Poleti, Jessika Cristina Chagas Lesbon, Elisangela Chicaroni Mattos, Heidge Fukumasu. USP-Botucatu: Rejane Maria Tommasini Grotto, Jayme A. Souza-Neto, Guilherme Targino Valente, Patricia Akemi Assato, Felipe Allan da Silva da Costa, Bianca Cechetto Carlos. Mendelics: Bibiana Santos, João Paulo Kitajima, Erika Freitas, David Schlesinger. Hemocentro Ribeirão Preto: Simone Kashima, Evandra Strazza Rodrigues, Svetoslav Nanev Slavov, Elaine Vieira dos Santos, Rafael dos Santos Bezerra, Luiz Carlos Junior de Alcantara, Marta Giovanetti, Vagner Fonseca, Flavia Aburjaile, Rodrigo Tocantins Calado. FAMERP-SJRP: Cecília Artico Banho, Lívia Sacchetto, Fábio Sossai Possebon, Leila Sabrina Ullmann, Cíntia Bittar, Guilherme Campos, Helena Lage Ferreira, Jorge A. Petroli Marchesi, Maisa C. Pereira Parra, Marília Moraes, Paula Rahal, Paulo Inacio da Costa, João Pessoa Araújo Jr., Maurício Lacerda Nogueira. Prefeitura de Sao Paulo: Melissa Palmieri. |
| EPI_ISL_1966321, EPI_ISL_1966322 | UPA SILVINA | Instituto Butantan / Mendelics | Instituto Butantan: Dimas Tadeu Covas, Sandra Coccuzzo Sampaio, Maria Carolina Elias, José Salvatore Leister Patané, Vincent Louis Viala, Antonio Jorge Martins, Ricardo Haddad, Claudia Renata dos Santos Barros, Elaine Cristina Marqueze, Raul Machado Neto, Debora Botequio Moretti, Jardelina de |

| | | | |
|---|---|---|---|
| | | | Souza Todao Bernardino, Loyze Paola Oliveira de Lima, Luiz Aurelio de Campos Crispin. Centro de Genômica Funcional da ESALQ: Luiz Lehmann Coutinho, Ricardo Augusto Brassaloti, Raquel de Lello Rocha Campos Cassano. NGS Soluções Genômicas: Pilar Drummond Sampaio Corrêa Mariani. FZEA-USP Pirassununga: Mirele Daiana Poleti, Jessika Cristina Chagas Lesbon, Elisangela Chicaroni Mattos, Heidge Fukumasu. USP-Botucatu: Rejane Maria Tommasini Grotto, Jayme A. Souza-Neto, Guilherme Targino Valente, Patricia Akemi Assato, Felipe Allan da Silva da Costa, Bianca Cechetto Carlos. Mendelics: Bibiana Santos, João Paulo Kitajima, Erika Freitas, David Schlesinger. Hemocentro Ribeirão Preto: Simone Kashima, Evandra Strazza Rodrigues, Svetoslav Nanev Slavov, Elaine Vieira dos Santos, Rafael dos Santos Bezerra, Luiz Carlos Junior de Alcantara, Marta Giovanetti, Vagner Fonseca, Flavia Aburjaile, Rodrigo Tocantins Calado. FAMERP-SJRP: Cecília Artico Banho, Lívia Sacchetto, Fábio Sossai Possebon, Leila Sabrina Ullmann, Cíntia Bittar, Guilherme Campos, Helena Lage Ferreira, Jorge A. Petroli Marchesi, Maisa C. Pereira Parra, Marília Moraes, Paula Rahal, Paulo Inacio da Costa, João Pessoa Araújo Jr., Maurício Lacerda Nogueira. Prefeitura de Sao Paulo: Melissa Palmieri. |
| EPI_ISL_1966323, EPI_ISL_1966324, EPI_ISL_1966325, EPI_ISL_1966326, EPI_ISL_1966328, EPI_ISL_1966331, EPI_ISL_1966332, EPI_ISL_1966333, EPI_ISL_1966334, EPI_ISL_1966335, EPI_ISL_1966336, EPI_ISL_1966337, EPI_ISL_1966338, EPI_ISL_1966339, EPI_ISL_1966340, EPI_ISL_1966341 | | | |
| see above | Ambulatório IB | Instituto Butantan / Mendelics | Instituto Butantan: Dimas Tadeu Covas, Sandra Coccuzzo Sampaio, Maria Carolina Elias, José Salvatore Leister Patané, Vincent Louis Viala, Antonio Jorge Martins, Ricardo Haddad, Claudia Renata dos Santos Barros, Elaine Cristina Marqueze, Raul Machado Neto, Debora Botequio Moretti, Jardelina de Souza Todao Bernardino, Loyze Paola Oliveira de Lima, Luiz Aurelio de Campos Crispin. Centro de Genômica Funcional da ESALQ: Luiz Lehmann Coutinho, Ricardo Augusto Brassaloti, Raquel de Lello Rocha Campos Cassano. NGS Soluções Genômicas: Pilar Drummond Sampaio Corrêa Mariani. FZEA-USP Pirassununga: Mirele Daiana Poleti, Jessika Cristina Chagas Lesbon, Elisangela Chicaroni Mattos, Heidge Fukumasu. USP-Botucatu: Rejane Maria Tommasini Grotto, Jayme A. Souza-Neto, Guilherme Targino Valente, Patricia Akemi Assato, Felipe Allan da Silva da Costa, Bianca Cechetto Carlos. Mendelics: Bibiana Santos, João Paulo Kitajima, Erika Freitas, David Schlesinger. Hemocentro Ribeirão Preto: Simone Kashima, Evandra Strazza Rodrigues, Svetoslav Nanev Slavov, Elaine Vieira dos Santos, Rafael dos Santos Bezerra, Luiz Carlos Junior de Alcantara, Marta Giovanetti, Vagner Fonseca, Flavia Aburjaile, Rodrigo Tocantins Calado. FAMERP-SJRP: Cecília Artico Banho, Lívia Sacchetto, Fábio Sossai Possebon, Leila Sabrina Ullmann, Cíntia Bittar, Guilherme Campos, Helena Lage Ferreira, Jorge A. Petroli Marchesi, Maisa C. Pereira Parra, Marília Moraes, Paula Rahal, Paulo Inacio da Costa, João Pessoa Araújo Jr., Maurício Lacerda Nogueira. Prefeitura de Sao Paulo: Melissa Palmieri. |
| EPI_ISL_1966343, EPI_ISL_1966344, EPI_ISL_1966345 | CENTRO DE SAUDE III TABATINGA | Instituto Butantan / Mendelics | Instituto Butantan: Dimas Tadeu Covas, Sandra Coccuzzo Sampaio, Maria Carolina Elias, José Salvatore Leister Patané, Vincent Louis Viala, Antonio Jorge Martins, Ricardo Haddad, Claudia Renata dos Santos Barros, Elaine Cristina Marqueze, Raul Machado Neto, Debora Botequio Moretti, Jardelina de Souza Todao Bernardino, Loyze Paola Oliveira de Lima, Luiz Aurelio de Campos Crispin. Centro de Genômica Funcional da ESALQ: Luiz Lehmann Coutinho, Ricardo Augusto Brassaloti, Raquel de Lello Rocha Campos Cassano. NGS Soluções Genômicas: Pilar Drummond Sampaio Corrêa Mariani. FZEA-USP Pirassununga: Mirele Daiana Poleti, Jessika Cristina Chagas Lesbon, Elisangela Chicaroni Mattos, Heidge Fukumasu. USP-Botucatu: Rejane Maria Tommasini Grotto, Jayme A. Souza-Neto, Guilherme Targino Valente, Patricia Akemi Assato, Felipe Allan da Silva da Costa, Bianca Cechetto Carlos. Mendelics: Bibiana Santos, João Paulo Kitajima, Erika Freitas, David Schlesinger. Hemocentro Ribeirão Preto: Simone Kashima, Evandra Strazza Rodrigues, Svetoslav Nanev Slavov, Elaine Vieira dos Santos, Rafael dos Santos Bezerra, Luiz Carlos Junior de Alcantara, Marta Giovanetti, Vagner Fonseca, Flavia Aburjaile, Rodrigo Tocantins Calado. FAMERP-SJRP: Cecília Artico Banho, Lívia Sacchetto, Fábio Sossai Possebon, Leila Sabrina Ullmann, Cíntia Bittar, Guilherme Campos, Helena Lage Ferreira, Jorge A. Petroli Marchesi, Maisa C. Pereira Parra, Marília Moraes, Paula Rahal, Paulo Inacio da Costa, João Pessoa Araújo Jr., Maurício Lacerda Nogueira. Prefeitura de Sao Paulo: Melissa Palmieri. |
| EPI_ISL_1966346, EPI_ISL_1966347, EPI_ISL_1966348, EPI_ISL_1966349 | SECRETARIA MUNICIPAL DE SAUDE DE VALPARAISO SP | Instituto Butantan / Mendelics | Instituto Butantan: Dimas Tadeu Covas, Sandra Coccuzzo Sampaio, Maria Carolina Elias, José Salvatore Leister Patané, Vincent Louis Viala, Antonio Jorge Martins, Ricardo Haddad, Claudia Renata dos Santos Barros, Elaine Cristina Marqueze, Raul Machado Neto, Debora Botequio Moretti, Jardelina de Souza Todao Bernardino, Loyze Paola Oliveira de Lima, Luiz Aurelio de Campos Crispin. Centro de Genômica Funcional da ESALQ: Luiz Lehmann Coutinho, Ricardo Augusto Brassaloti, Raquel de Lello Rocha Campos Cassano. NGS Soluções Genômicas: Pilar Drummond Sampaio Corrêa Mariani. FZEA-USP Pirassununga: Mirele Daiana Poleti, Jessika Cristina Chagas Lesbon, Elisangela Chicaroni Mattos, Heidge Fukumasu. USP-Botucatu: Rejane Maria Tommasini Grotto, Jayme A. Souza-Neto, Guilherme Targino Valente, Patricia Akemi Assato, Felipe Allan da Silva da Costa, Bianca Cechetto Carlos. Mendelics: Bibiana Santos, João Paulo Kitajima, Erika Freitas, David Schlesinger. Hemocentro Ribeirão Preto: Simone Kashima, Evandra Strazza Rodrigues, Svetoslav Nanev Slavov, Elaine Vieira dos Santos, Rafael dos Santos Bezerra, Luiz Carlos Junior de Alcantara, Marta Giovanetti, Vagner Fonseca, Flavia Aburjaile, Rodrigo Tocantins Calado. FAMERP-SJRP: Cecília Artico Banho, Lívia Sacchetto, Fábio Sossai Possebon, Leila Sabrina Ullmann, Cíntia Bittar, Guilherme Campos, Helena Lage Ferreira, Jorge A. Petroli Marchesi, Maisa C. Pereira Parra, Marília Moraes, Paula Rahal, Paulo Inacio da Costa, João Pessoa Araújo Jr., Maurício Lacerda Nogueira. Prefeitura de Sao Paulo: Melissa Palmieri. |
| EPI_ISL_1966351 | ESF VILA GLORIA I | Instituto Butantan / Mendelics | Instituto Butantan: Dimas Tadeu Covas, Sandra Coccuzzo Sampaio, Maria Carolina Elias, José Salvatore Leister Patané, Vincent Louis Viala, Antonio Jorge Martins, Ricardo Haddad, Claudia Renata dos Santos Barros, Elaine Cristina Marqueze, Raul Machado Neto, Debora Botequio Moretti, Jardelina de Souza Todao Bernardino, Loyze Paola Oliveira de Lima, Luiz Aurelio de Campos Crispin. Centro de Genômica Funcional da ESALQ: Luiz Lehmann Coutinho, Ricardo Augusto Brassaloti, Raquel de Lello Rocha Campos Cassano. NGS Soluções Genômicas: Pilar Drummond Sampaio Corrêa Mariani. FZEA-USP Pirassununga: Mirele Daiana Poleti, Jessika Cristina Chagas Lesbon, Elisangela Chicaroni Mattos, Heidge Fukumasu. USP-Botucatu: Rejane Maria Tommasini Grotto, Jayme A. Souza-Neto, Guilherme Targino Valente, Patricia Akemi Assato, Felipe Allan da Silva da Costa, Bianca Cechetto Carlos. Mendelics: Bibiana Santos, João Paulo Kitajima, Erika Freitas, David Schlesinger. Hemocentro Ribeirão Preto: Simone Kashima, Evandra Strazza Rodrigues, Svetoslav Nanev Slavov, Elaine Vieira dos Santos, Rafael dos Santos Bezerra, Luiz Carlos Junior de Alcantara, Marta Giovanetti, Vagner Fonseca, Flavia Aburjaile, Rodrigo Tocantins Calado. FAMERP-SJRP: Cecília Artico Banho, Lívia Sacchetto, Fábio Sossai Possebon, Leila Sabrina Ullmann, Cíntia Bittar, Guilherme Campos, Helena Lage Ferreira, Jorge A. Petroli Marchesi, Maisa C. Pereira Parra, Marília Moraes, Paula Rahal, Paulo Inacio da Costa, João Pessoa Araújo Jr., Maurício Lacerda Nogueira. Prefeitura de Sao Paulo: Melissa Palmieri. |
| EPI_ISL_1966352, EPI_ISL_1966353, EPI_ISL_1966354, EPI_ISL_1966355, EPI_ISL_1966356 | UNIDADE REFERENCIAL SUDOESTE | Instituto Butantan / Mendelics | Instituto Butantan: Dimas Tadeu Covas, Sandra Coccuzzo Sampaio, Maria Carolina Elias, José Salvatore Leister Patané, Vincent Louis Viala, Antonio Jorge Martins, Ricardo Haddad, Claudia Renata dos Santos Barros, Elaine Cristina Marqueze, Raul Machado Neto, Debora Botequio Moretti, Jardelina de Souza Todao Bernardino, Loyze Paola Oliveira de Lima, Luiz Aurelio de Campos Crispin. Centro de Genômica Funcional da ESALQ: Luiz Lehmann Coutinho, Ricardo Augusto Brassaloti, Raquel de Lello Rocha Campos Cassano. NGS Soluções Genômicas: Pilar Drummond Sampaio Corrêa Mariani. FZEA-USP Pirassununga: Mirele Daiana Poleti, Jessika Cristina Chagas Lesbon, Elisangela Chicaroni Mattos, Heidge Fukumasu. USP-Botucatu: Rejane Maria Tommasini Grotto, Jayme A. Souza-Neto, Guilherme Targino Valente, Patricia Akemi Assato, Felipe Allan da Silva da Costa, Bianca Cechetto Carlos. Mendelics: Bibiana Santos, João Paulo Kitajima, Erika Freitas, David Schlesinger. Hemocentro Ribeirão Preto: Simone Kashima, Evandra Strazza Rodrigues, Svetoslav Nanev Slavov, Elaine Vieira dos Santos, Rafael dos Santos Bezerra, Luiz Carlos Junior de Alcantara, Marta Giovanetti, Vagner Fonseca, Flavia Aburjaile, Rodrigo Tocantins Calado. FAMERP-SJRP: Cecília Artico Banho, Lívia Sacchetto, Fábio Sossai Possebon, Leila Sabrina Ullmann, Cíntia Bittar, Guilherme Campos, Helena Lage Ferreira, Jorge A. Petroli Marchesi, Maisa C. Pereira Parra, Marília Moraes, Paula Rahal, Paulo Inacio da Costa, João Pessoa Araújo Jr., Maurício Lacerda Nogueira. Prefeitura de Sao Paulo: Melissa Palmieri. |
| EPI_ISL_1966357, EPI_ISL_1966358, EPI_ISL_1966359 | SMS SECRETARIA MUNICIPAL DE SAUDE DE BOITUVA | Instituto Butantan / Mendelics | Instituto Butantan: Dimas Tadeu Covas, Sandra Coccuzzo Sampaio, Maria Carolina Elias, José Salvatore Leister Patané, Vincent Louis Viala, Antonio Jorge Martins, Ricardo Haddad, Claudia Renata dos Santos Barros, Elaine Cristina Marqueze, Raul Machado Neto, Debora Botequio Moretti, Jardelina de Souza Todao Bernardino, Loyze Paola Oliveira de Lima, Luiz Aurelio de Campos Crispin. Centro de Genômica Funcional da ESALQ: Luiz Lehmann Coutinho, Ricardo Augusto Brassaloti, Raquel de Lello Rocha Campos Cassano. NGS Soluções Genômicas: Pilar Drummond Sampaio Corrêa Mariani. FZEA-USP Pirassununga: Mirele Daiana Poleti, Jessika Cristina Chagas Lesbon, Elisangela Chicaroni Mattos, Heidge Fukumasu. USP-Botucatu: Rejane Maria Tommasini Grotto, Jayme A. Souza-Neto, Guilherme Targino Valente, Patricia Akemi Assato, Felipe Allan da Silva da Costa, Bianca Cechetto Carlos. Mendelics: Bibiana Santos, João Paulo Kitajima, Erika Freitas, David Schlesinger. Hemocentro Ribeirão Preto: Simone Kashima, Evandra Strazza Rodrigues, Svetoslav Nanev Slavov, Elaine Vieira dos Santos, Rafael dos Santos Bezerra, Luiz Carlos Junior de Alcantara, Marta Giovanetti, Vagner Fonseca, Flavia Aburjaile, Rodrigo Tocantins Calado. FAMERP-SJRP: Cecília Artico Banho, Lívia Sacchetto, Fábio Sossai Possebon, Leila Sabrina Ullmann, Cíntia Bittar, Guilherme Campos, Helena Lage Ferreira, Jorge A. Petroli Marchesi, Maisa C. Pereira Parra, Marília Moraes, Paula Rahal, Paulo Inacio da Costa, João Pessoa Araújo Jr., Maurício Lacerda Nogueira. Prefeitura de Sao Paulo: Melissa Palmieri. |
| EPI_ISL_1966360 | SECRETARIA DE SAUDE CABREUVA | Instituto Butantan / Mendelics | Instituto Butantan: Dimas Tadeu Covas, Sandra Coccuzzo Sampaio, Maria Carolina Elias, José Salvatore Leister Patané, Vincent Louis Viala, Antonio Jorge Martins, Ricardo Haddad, Claudia Renata dos Santos Barros, Elaine Cristina Marqueze, Raul Machado Neto, Debora Botequio Moretti, Jardelina de Souza Todao Bernardino, Loyze Paola Oliveira de Lima, Luiz Aurelio de Campos Crispin. Centro de Genômica Funcional da ESALQ: Luiz Lehmann Coutinho, Ricardo Augusto Brassaloti, Raquel de Lello Rocha Campos Cassano. NGS Soluções Genômicas: Pilar Drummond Sampaio Corrêa Mariani. FZEA-USP Pirassununga: Mirele Daiana Poleti, Jessika Cristina Chagas Lesbon, Elisangela Chicaroni Mattos, Heidge Fukumasu. USP-Botucatu: Rejane Maria Tommasini Grotto, Jayme A. Souza-Neto, Guilherme Targino Valente, Patricia Akemi Assato, Felipe Allan da Silva da Costa, Bianca Cechetto Carlos. Mendelics: Bibiana Santos, João Paulo Kitajima, Erika Freitas, David Schlesinger. Hemocentro Ribeirão Preto: Simone Kashima, Evandra Strazza |

| | | | |
|---|---|---|---|
| | | | Rodrigues, Svetoslav Nanev Slavov, Elaine Vieira dos Santos, Rafael dos Santos Bezerra, Luiz Carlos Junior de Alcantara, Marta Giovanetti, Vagner Fonseca, Flavia Aburjaile, Rodrigo Tocantins Calado. FAMERP-SJRP: Cecília Artico Banho, Lívia Sacchetto, Fábio Sossai Possebon, Leila Sabrina Ullmann, Cíntia Bittar, Guilherme Campos, Helena Lage Ferreira, Jorge A. Petroli Marchesi, Maisa C. Pereira Parra, Marília Moraes, Paula Rahal, Paulo Inacio da Costa, João Pessoa Araújo Jr., Maurício Lacerda Nogueira. Prefeitura de Sao Paulo: Melissa Palmieri. |
| EPI_ISL_1966361, EPI_ISL_1966362, EPI_ISL_1966363 | PRONTO ATENDIMENTO SAO JOSE | Instituto Butantan / Mendelics | Instituto Butantan: Dimas Tadeu Covas, Sandra Coccuzzo Sampaio, Maria Carolina Elias, José Salvatore Leister Patané, Vincent Louis Viala, Antonio Jorge Martins, Ricardo Haddad, Claudia Renata dos Santos Barros, Elaine Cristina Marqueze, Raul Machado Neto, Debora Botequio Moretti, Jardelina de Souza Todao Bernardino, Loyze Paola Oliveira de Lima, Luiz Aurelio de Campos Crispin. Centro de Genômica Funcional da ESALQ: Luiz Lehmann Coutinho, Ricardo Augusto Brassaloti, Raquel de Lello Rocha Campos Cassano. NGS Soluções Genômicas: Pilar Drummond Sampaio Corrêa Mariani. FZEA-USP Pirassununga: Mirele Daiana Poleti, Jessika Cristina Chagas Lesbon, Elisangela Chicaroni Mattos, Heidge Fukumasu. USP-Botucatu: Rejane Maria Tommasini Grotto, Jayme A. Souza-Neto, Guilherme Targino Valente, Patricia Akemi Assato, Felipe Allan da Silva da Costa, Bianca Cechetto Carlos. Mendelics: Bibiana Santos, João Paulo Kitajima, Erika Freitas, David Schlesinger. Hemocentro Ribeirão Preto: Simone Kashima, Evandra Strazza Rodrigues, Svetoslav Nanev Slavov, Elaine Vieira dos Santos, Rafael dos Santos Bezerra, Luiz Carlos Junior de Alcantara, Marta Giovanetti, Vagner Fonseca, Flavia Aburjaile, Rodrigo Tocantins Calado. FAMERP-SJRP: Cecília Artico Banho, Lívia Sacchetto, Fábio Sossai Possebon, Leila Sabrina Ullmann, Cíntia Bittar, Guilherme Campos, Helena Lage Ferreira, Jorge A. Petroli Marchesi, Maisa C. Pereira Parra, Marília Moraes, Paula Rahal, Paulo Inacio da Costa, João Pessoa Araújo Jr., Maurício Lacerda Nogueira. Prefeitura de Sao Paulo: Melissa Palmieri. |
| EPI_ISL_1966364 | HOSPITAL MUNICIPAL DR MARIO GATTI CAMPINAS | Instituto Butantan / Mendelics | Instituto Butantan: Dimas Tadeu Covas, Sandra Coccuzzo Sampaio, Maria Carolina Elias, José Salvatore Leister Patané, Vincent Louis Viala, Antonio Jorge Martins, Ricardo Haddad, Claudia Renata dos Santos Barros, Elaine Cristina Marqueze, Raul Machado Neto, Debora Botequio Moretti, Jardelina de Souza Todao Bernardino, Loyze Paola Oliveira de Lima, Luiz Aurelio de Campos Crispin. Centro de Genômica Funcional da ESALQ: Luiz Lehmann Coutinho, Ricardo Augusto Brassaloti, Raquel de Lello Rocha Campos Cassano. NGS Soluções Genômicas: Pilar Drummond Sampaio Corrêa Mariani. FZEA-USP Pirassununga: Mirele Daiana Poleti, Jessika Cristina Chagas Lesbon, Elisangela Chicaroni Mattos, Heidge Fukumasu. USP-Botucatu: Rejane Maria Tommasini Grotto, Jayme A. Souza-Neto, Guilherme Targino Valente, Patricia Akemi Assato, Felipe Allan da Silva da Costa, Bianca Cechetto Carlos. Mendelics: Bibiana Santos, João Paulo Kitajima, Erika Freitas, David Schlesinger. Hemocentro Ribeirão Preto: Simone Kashima, Evandra Strazza Rodrigues, Svetoslav Nanev Slavov, Elaine Vieira dos Santos, Rafael dos Santos Bezerra, Luiz Carlos Junior de Alcantara, Marta Giovanetti, Vagner Fonseca, Flavia Aburjaile, Rodrigo Tocantins Calado. FAMERP-SJRP: Cecília Artico Banho, Lívia Sacchetto, Fábio Sossai Possebon, Leila Sabrina Ullmann, Cíntia Bittar, Guilherme Campos, Helena Lage Ferreira, Jorge A. Petroli Marchesi, Maisa C. Pereira Parra, Marília Moraes, Paula Rahal, Paulo Inacio da Costa, João Pessoa Araújo Jr., Maurício Lacerda Nogueira. Prefeitura de Sao Paulo: Melissa Palmieri. |
| EPI_ISL_1966365, EPI_ISL_1966366 | CENTRO DE SAUDE DE CANITAR | Instituto Butantan / Mendelics | Instituto Butantan: Dimas Tadeu Covas, Sandra Coccuzzo Sampaio, Maria Carolina Elias, José Salvatore Leister Patané, Vincent Louis Viala, Antonio Jorge Martins, Ricardo Haddad, Claudia Renata dos Santos Barros, Elaine Cristina Marqueze, Raul Machado Neto, Debora Botequio Moretti, Jardelina de Souza Todao Bernardino, Loyze Paola Oliveira de Lima, Luiz Aurelio de Campos Crispin. Centro de Genômica Funcional da ESALQ: Luiz Lehmann Coutinho, Ricardo Augusto Brassaloti, Raquel de Lello Rocha Campos Cassano. NGS Soluções Genômicas: Pilar Drummond Sampaio Corrêa Mariani. FZEA-USP Pirassununga: Mirele Daiana Poleti, Jessika Cristina Chagas Lesbon, Elisangela Chicaroni Mattos, Heidge Fukumasu. USP-Botucatu: Rejane Maria Tommasini Grotto, Jayme A. Souza-Neto, Guilherme Targino Valente, Patricia Akemi Assato, Felipe Allan da Silva da Costa, Bianca Cechetto Carlos. Mendelics: Bibiana Santos, João Paulo Kitajima, Erika Freitas, David Schlesinger. Hemocentro Ribeirão Preto: Simone Kashima, Evandra Strazza Rodrigues, Svetoslav Nanev Slavov, Elaine Vieira dos Santos, Rafael dos Santos Bezerra, Luiz Carlos Junior de Alcantara, Marta Giovanetti, Vagner Fonseca, Flavia Aburjaile, Rodrigo Tocantins Calado. FAMERP-SJRP: Cecília Artico Banho, Lívia Sacchetto, Fábio Sossai Possebon, Leila Sabrina Ullmann, Cíntia Bittar, Guilherme Campos, Helena Lage Ferreira, Jorge A. Petroli Marchesi, Maisa C. Pereira Parra, Marília Moraes, Paula Rahal, Paulo Inacio da Costa, João Pessoa Araújo Jr., Maurício Lacerda Nogueira. Prefeitura de Sao Paulo: Melissa Palmieri. |
| EPI_ISL_1966368 | CENTRO DE SAUDE DE CHAVANTES | Instituto Butantan / Mendelics | Instituto Butantan: Dimas Tadeu Covas, Sandra Coccuzzo Sampaio, Maria Carolina Elias, José Salvatore Leister Patané, Vincent Louis Viala, Antonio Jorge Martins, Ricardo Haddad, Claudia Renata dos Santos Barros, Elaine Cristina Marqueze, Raul Machado Neto, Debora Botequio Moretti, Jardelina de Souza Todao Bernardino, Loyze Paola Oliveira de Lima, Luiz Aurelio de Campos Crispin. Centro de Genômica Funcional da ESALQ: Luiz Lehmann Coutinho, Ricardo Augusto Brassaloti, Raquel de Lello Rocha Campos Cassano. NGS Soluções Genômicas: Pilar Drummond Sampaio Corrêa Mariani. FZEA-USP Pirassununga: Mirele Daiana Poleti, Jessika Cristina Chagas Lesbon, Elisangela Chicaroni Mattos, Heidge Fukumasu. USP-Botucatu: Rejane Maria Tommasini Grotto, Jayme A. Souza-Neto, Guilherme Targino Valente, Patricia Akemi Assato, Felipe Allan da Silva da Costa, Bianca Cechetto Carlos. Mendelics: Bibiana Santos, João Paulo Kitajima, Erika Freitas, David Schlesinger. Hemocentro Ribeirão Preto: Simone Kashima, Evandra Strazza Rodrigues, Svetoslav Nanev Slavov, Elaine Vieira dos Santos, Rafael dos Santos Bezerra, Luiz Carlos Junior de Alcantara, Marta Giovanetti, Vagner Fonseca, Flavia Aburjaile, Rodrigo Tocantins Calado. FAMERP-SJRP: Cecília Artico Banho, Lívia Sacchetto, Fábio Sossai Possebon, Leila Sabrina Ullmann, Cíntia Bittar, Guilherme Campos, Helena Lage Ferreira, Jorge A. Petroli Marchesi, Maisa C. Pereira Parra, Marília Moraes, Paula Rahal, Paulo Inacio da Costa, João Pessoa Araújo Jr., Maurício Lacerda Nogueira. Prefeitura de Sao Paulo: Melissa Palmieri. |
| EPI_ISL_1966369, EPI_ISL_1966370, EPI_ISL_1966371, EPI_ISL_1966372, EPI_ISL_1966373 | UPA DR FRANCO DA ROCHA | Instituto Butantan / Mendelics | Instituto Butantan: Dimas Tadeu Covas, Sandra Coccuzzo Sampaio, Maria Carolina Elias, José Salvatore Leister Patané, Vincent Louis Viala, Antonio Jorge Martins, Ricardo Haddad, Claudia Renata dos Santos Barros, Elaine Cristina Marqueze, Raul Machado Neto, Debora Botequio Moretti, Jardelina de Souza Todao Bernardino, Loyze Paola Oliveira de Lima, Luiz Aurelio de Campos Crispin. Centro de Genômica Funcional da ESALQ: Luiz Lehmann Coutinho, Ricardo Augusto Brassaloti, Raquel de Lello Rocha Campos Cassano. NGS Soluções Genômicas: Pilar Drummond Sampaio Corrêa Mariani. FZEA-USP Pirassununga: Mirele Daiana Poleti, Jessika Cristina Chagas Lesbon, Elisangela Chicaroni Mattos, Heidge Fukumasu. USP-Botucatu: Rejane Maria Tommasini Grotto, Jayme A. Souza-Neto, Guilherme Targino Valente, Patricia Akemi Assato, Felipe Allan da Silva da Costa, Bianca Cechetto Carlos. Mendelics: Bibiana Santos, João Paulo Kitajima, Erika Freitas, David Schlesinger. Hemocentro Ribeirão Preto: Simone Kashima, Evandra Strazza Rodrigues, Svetoslav Nanev Slavov, Elaine Vieira dos Santos, Rafael dos Santos Bezerra, Luiz Carlos Junior de Alcantara, Marta Giovanetti, Vagner Fonseca, Flavia Aburjaile, Rodrigo Tocantins Calado. FAMERP-SJRP: Cecília Artico Banho, Lívia Sacchetto, Fábio Sossai Possebon, Leila Sabrina Ullmann, Cíntia Bittar, Guilherme Campos, Helena Lage Ferreira, Jorge A. Petroli Marchesi, Maisa C. Pereira Parra, Marília Moraes, Paula Rahal, Paulo Inacio da Costa, João Pessoa Araújo Jr., Maurício Lacerda Nogueira. Prefeitura de Sao Paulo: Melissa Palmieri. |
| EPI_ISL_1966374 | USF II DE HERCULANDIA | Instituto Butantan / Mendelics | Instituto Butantan: Dimas Tadeu Covas, Sandra Coccuzzo Sampaio, Maria Carolina Elias, José Salvatore Leister Patané, Vincent Louis Viala, Antonio Jorge Martins, Ricardo Haddad, Claudia Renata dos Santos Barros, Elaine Cristina Marqueze, Raul Machado Neto, Debora Botequio Moretti, Jardelina de Souza Todao Bernardino, Loyze Paola Oliveira de Lima, Luiz Aurelio de Campos Crispin. Centro de Genômica Funcional da ESALQ: Luiz Lehmann Coutinho, Ricardo Augusto Brassaloti, Raquel de Lello Rocha Campos Cassano. NGS Soluções Genômicas: Pilar Drummond Sampaio Corrêa Mariani. FZEA-USP Pirassununga: Mirele Daiana Poleti, Jessika Cristina Chagas Lesbon, Elisangela Chicaroni Mattos, Heidge Fukumasu. USP-Botucatu: Rejane Maria Tommasini Grotto, Jayme A. Souza-Neto, Guilherme Targino Valente, Patricia Akemi Assato, Felipe Allan da Silva da Costa, Bianca Cechetto Carlos. Mendelics: Bibiana Santos, João Paulo Kitajima, Erika Freitas, David Schlesinger. Hemocentro Ribeirão Preto: Simone Kashima, Evandra Strazza Rodrigues, Svetoslav Nanev Slavov, Elaine Vieira dos Santos, Rafael dos Santos Bezerra, Luiz Carlos Junior de Alcantara, Marta Giovanetti, Vagner Fonseca, Flavia Aburjaile, Rodrigo Tocantins Calado. FAMERP-SJRP: Cecília Artico Banho, Lívia Sacchetto, Fábio Sossai Possebon, Leila Sabrina Ullmann, Cíntia Bittar, Guilherme Campos, Helena Lage Ferreira, Jorge A. Petroli Marchesi, Maisa C. Pereira Parra, Marília Moraes, Paula Rahal, Paulo Inacio da Costa, João Pessoa Araújo Jr., Maurício Lacerda Nogueira. Prefeitura de Sao Paulo: Melissa Palmieri. |
| EPI_ISL_1966375, EPI_ISL_1966376 | UNIDADE RESPIRATORIA NOVA HORTOLANDIA | Instituto Butantan / Mendelics | Instituto Butantan: Dimas Tadeu Covas, Sandra Coccuzzo Sampaio, Maria Carolina Elias, José Salvatore Leister Patané, Vincent Louis Viala, Antonio Jorge Martins, Ricardo Haddad, Claudia Renata dos Santos Barros, Elaine Cristina Marqueze, Raul Machado Neto, Debora Botequio Moretti, Jardelina de Souza Todao Bernardino, Loyze Paola Oliveira de Lima, Luiz Aurelio de Campos Crispin. Centro de Genômica Funcional da ESALQ: Luiz Lehmann Coutinho, Ricardo Augusto Brassaloti, Raquel de Lello Rocha Campos Cassano. NGS Soluções Genômicas: Pilar Drummond Sampaio Corrêa Mariani. FZEA-USP Pirassununga: Mirele Daiana Poleti, Jessika Cristina Chagas Lesbon, Elisangela Chicaroni Mattos, Heidge Fukumasu. USP-Botucatu: Rejane Maria Tommasini Grotto, Jayme A. Souza-Neto, Guilherme Targino Valente, Patricia Akemi Assato, Felipe Allan da Silva da Costa, Bianca Cechetto Carlos. Mendelics: Bibiana Santos, João Paulo Kitajima, Erika Freitas, David Schlesinger. Hemocentro Ribeirão Preto: Simone Kashima, Evandra Strazza Rodrigues, Svetoslav Nanev Slavov, Elaine Vieira dos Santos, Rafael dos Santos Bezerra, Luiz Carlos Junior de Alcantara, Marta Giovanetti, Vagner Fonseca, Flavia Aburjaile, Rodrigo Tocantins Calado. FAMERP-SJRP: Cecília Artico Banho, Lívia Sacchetto, Fábio Sossai Possebon, Leila Sabrina Ullmann, Cíntia Bittar, Guilherme Campos, Helena Lage Ferreira, Jorge A. Petroli Marchesi, Maisa C. Pereira Parra, Marília Moraes, Paula Rahal, Paulo Inacio da Costa, João Pessoa Araújo Jr., Maurício Lacerda Nogueira. Prefeitura de Sao Paulo: Melissa Palmieri. |
| EPI_ISL_1966377, EPI_ISL_1966378 | SMS IPERO | Instituto Butantan / Mendelics | Instituto Butantan: Dimas Tadeu Covas, Sandra Coccuzzo Sampaio, Maria Carolina Elias, José Salvatore Leister Patané, Vincent Louis Viala, Antonio Jorge Martins, Ricardo Haddad, Claudia Renata dos Santos Barros, Elaine Cristina Marqueze, Raul Machado Neto, Debora Botequio Moretti, Jardelina de |

| | | | |
|---|---|---|---|
| | | | Souza Todao Bernardino, Loyze Paola Oliveira de Lima, Luiz Aurelio de Campos Crispin. Centro de Genômica Funcional da ESALQ: Luiz Lehmann Coutinho, Ricardo Augusto Brassaloti, Raquel de Lello Rocha Campos Cassano. NGS Soluções Genômicas: Pilar Drummond Sampaio Corrêa Mariani. FZEA-USP Pirassununga: Mirele Daiana Poleti, Jessika Cristina Chagas Lesbon, Elisangela Chicaroni Mattos, Heidge Fukumasu. USP-Botucatu: Rejane Maria Tommasini Grotto, Jayme A. Souza-Neto, Guilherme Targino Valente, Patricia Akemi Assato, Felipe Allan da Silva da Costa, Bianca Cechetto Carlos. Mendelics: Bibiana Santos, João Paulo Kitajima, Erika Freitas, David Schlesinger. Hemocentro Ribeirão Preto: Simone Kashima, Evandra Strazza Rodrigues, Svetoslav Nanev Slavov, Elaine Vieira dos Santos, Rafael dos Santos Bezerra, Luiz Carlos Junior de Alcantara, Marta Giovanetti, Vagner Fonseca, Flavia Aburjaile, Rodrigo Tocantins Calado. FAMERP-SJRP: Cecília Artico Banho, Lívia Sacchetto, Fábio Sossai Possebon, Leila Sabrina Ullmann, Cíntia Bittar, Guilherme Campos, Helena Lage Ferreira, Jorge A. Petroli Marchesi, Maisa C. Pereira Parra, Marília Moraes, Paula Rahal, Paulo Inacio da Costa, João Pessoa Araújo Jr., Maurício Lacerda Nogueira. Prefeitura de Sao Paulo: Melissa Palmieri. |
| EPI_ISL_1966379 | POLICLINICA COVID 19 ITAPETININGA | Instituto Butantan / Mendelics | Instituto Butantan: Dimas Tadeu Covas, Sandra Coccuzzo Sampaio, Maria Carolina Elias, José Salvatore Leister Patané, Vincent Louis Viala, Antonio Jorge Martins, Ricardo Haddad, Claudia Renata dos Santos Barros, Elaine Cristina Marqueze, Raul Machado Neto, Debora Botequio Moretti, Jardelina de Souza Todao Bernardino, Loyze Paola Oliveira de Lima, Luiz Aurelio de Campos Crispin. Centro de Genômica Funcional da ESALQ: Luiz Lehmann Coutinho, Ricardo Augusto Brassaloti, Raquel de Lello Rocha Campos Cassano. NGS Soluções Genômicas: Pilar Drummond Sampaio Corrêa Mariani. FZEA-USP Pirassununga: Mirele Daiana Poleti, Jessika Cristina Chagas Lesbon, Elisangela Chicaroni Mattos, Heidge Fukumasu. USP-Botucatu: Rejane Maria Tommasini Grotto, Jayme A. Souza-Neto, Guilherme Targino Valente, Patricia Akemi Assato, Felipe Allan da Silva da Costa, Bianca Cechetto Carlos. Mendelics: Bibiana Santos, João Paulo Kitajima, Erika Freitas, David Schlesinger. Hemocentro Ribeirão Preto: Simone Kashima, Evandra Strazza Rodrigues, Svetoslav Nanev Slavov, Elaine Vieira dos Santos, Rafael dos Santos Bezerra, Luiz Carlos Junior de Alcantara, Marta Giovanetti, Vagner Fonseca, Flavia Aburjaile, Rodrigo Tocantins Calado. FAMERP-SJRP: Cecília Artico Banho, Lívia Sacchetto, Fábio Sossai Possebon, Leila Sabrina Ullmann, Cíntia Bittar, Guilherme Campos, Helena Lage Ferreira, Jorge A. Petroli Marchesi, Maisa C. Pereira Parra, Marília Moraes, Paula Rahal, Paulo Inacio da Costa, João Pessoa Araújo Jr., Maurício Lacerda Nogueira. Prefeitura de Sao Paulo: Melissa Palmieri. |
| EPI_ISL_1966380 | HOSPITAL REGIONAL DE ITAPETININGA | Instituto Butantan / Mendelics | Instituto Butantan: Dimas Tadeu Covas, Sandra Coccuzzo Sampaio, Maria Carolina Elias, José Salvatore Leister Patané, Vincent Louis Viala, Antonio Jorge Martins, Ricardo Haddad, Claudia Renata dos Santos Barros, Elaine Cristina Marqueze, Raul Machado Neto, Debora Botequio Moretti, Jardelina de Souza Todao Bernardino, Loyze Paola Oliveira de Lima, Luiz Aurelio de Campos Crispin. Centro de Genômica Funcional da ESALQ: Luiz Lehmann Coutinho, Ricardo Augusto Brassaloti, Raquel de Lello Rocha Campos Cassano. NGS Soluções Genômicas: Pilar Drummond Sampaio Corrêa Mariani. FZEA-USP Pirassununga: Mirele Daiana Poleti, Jessika Cristina Chagas Lesbon, Elisangela Chicaroni Mattos, Heidge Fukumasu. USP-Botucatu: Rejane Maria Tommasini Grotto, Jayme A. Souza-Neto, Guilherme Targino Valente, Patricia Akemi Assato, Felipe Allan da Silva da Costa, Bianca Cechetto Carlos. Mendelics: Bibiana Santos, João Paulo Kitajima, Erika Freitas, David Schlesinger. Hemocentro Ribeirão Preto: Simone Kashima, Evandra Strazza Rodrigues, Svetoslav Nanev Slavov, Elaine Vieira dos Santos, Rafael dos Santos Bezerra, Luiz Carlos Junior de Alcantara, Marta Giovanetti, Vagner Fonseca, Flavia Aburjaile, Rodrigo Tocantins Calado. FAMERP-SJRP: Cecília Artico Banho, Lívia Sacchetto, Fábio Sossai Possebon, Leila Sabrina Ullmann, Cíntia Bittar, Guilherme Campos, Helena Lage Ferreira, Jorge A. Petroli Marchesi, Maisa C. Pereira Parra, Marília Moraes, Paula Rahal, Paulo Inacio da Costa, João Pessoa Araújo Jr., Maurício Lacerda Nogueira. Prefeitura de Sao Paulo: Melissa Palmieri. |
| EPI_ISL_1966381, EPI_ISL_1966382, EPI_ISL_1966383, EPI_ISL_1966384 | POLICLINICA COVID 19 ITAPETININGA | Instituto Butantan / Mendelics | Instituto Butantan: Dimas Tadeu Covas, Sandra Coccuzzo Sampaio, Maria Carolina Elias, José Salvatore Leister Patané, Vincent Louis Viala, Antonio Jorge Martins, Ricardo Haddad, Claudia Renata dos Santos Barros, Elaine Cristina Marqueze, Raul Machado Neto, Debora Botequio Moretti, Jardelina de Souza Todao Bernardino, Loyze Paola Oliveira de Lima, Luiz Aurelio de Campos Crispin. Centro de Genômica Funcional da ESALQ: Luiz Lehmann Coutinho, Ricardo Augusto Brassaloti, Raquel de Lello Rocha Campos Cassano. NGS Soluções Genômicas: Pilar Drummond Sampaio Corrêa Mariani. FZEA-USP Pirassununga: Mirele Daiana Poleti, Jessika Cristina Chagas Lesbon, Elisangela Chicaroni Mattos, Heidge Fukumasu. USP-Botucatu: Rejane Maria Tommasini Grotto, Jayme A. Souza-Neto, Guilherme Targino Valente, Patricia Akemi Assato, Felipe Allan da Silva da Costa, Bianca Cechetto Carlos. Mendelics: Bibiana Santos, João Paulo Kitajima, Erika Freitas, David Schlesinger. Hemocentro Ribeirão Preto: Simone Kashima, Evandra Strazza Rodrigues, Svetoslav Nanev Slavov, Elaine Vieira dos Santos, Rafael dos Santos Bezerra, Luiz Carlos Junior de Alcantara, Marta Giovanetti, Vagner Fonseca, Flavia Aburjaile, Rodrigo Tocantins Calado. FAMERP-SJRP: Cecília Artico Banho, Lívia Sacchetto, Fábio Sossai Possebon, Leila Sabrina Ullmann, Cíntia Bittar, Guilherme Campos, Helena Lage Ferreira, Jorge A. Petroli Marchesi, Maisa C. Pereira Parra, Marília Moraes, Paula Rahal, Paulo Inacio da Costa, João Pessoa Araújo Jr., Maurício Lacerda Nogueira. Prefeitura de Sao Paulo: Melissa Palmieri. |
| EPI_ISL_1966385, EPI_ISL_1966386, EPI_ISL_1966387, EPI_ISL_1966388 | UNIDADE DE PRONTO ATENDIMENTO UPA DRA ANA OLIVIA BENTIVOGLIO | Instituto Butantan / Mendelics | Instituto Butantan: Dimas Tadeu Covas, Sandra Coccuzzo Sampaio, Maria Carolina Elias, José Salvatore Leister Patané, Vincent Louis Viala, Antonio Jorge Martins, Ricardo Haddad, Claudia Renata dos Santos Barros, Elaine Cristina Marqueze, Raul Machado Neto, Debora Botequio Moretti, Jardelina de Souza Todao Bernardino, Loyze Paola Oliveira de Lima, Luiz Aurelio de Campos Crispin. Centro de Genômica Funcional da ESALQ: Luiz Lehmann Coutinho, Ricardo Augusto Brassaloti, Raquel de Lello Rocha Campos Cassano. NGS Soluções Genômicas: Pilar Drummond Sampaio Corrêa Mariani. FZEA-USP Pirassununga: Mirele Daiana Poleti, Jessika Cristina Chagas Lesbon, Elisangela Chicaroni Mattos, Heidge Fukumasu. USP-Botucatu: Rejane Maria Tommasini Grotto, Jayme A. Souza-Neto, Guilherme Targino Valente, Patricia Akemi Assato, Felipe Allan da Silva da Costa, Bianca Cechetto Carlos. Mendelics: Bibiana Santos, João Paulo Kitajima, Erika Freitas, David Schlesinger. Hemocentro Ribeirão Preto: Simone Kashima, Evandra Strazza Rodrigues, Svetoslav Nanev Slavov, Elaine Vieira dos Santos, Rafael dos Santos Bezerra, Luiz Carlos Junior de Alcantara, Marta Giovanetti, Vagner Fonseca, Flavia Aburjaile, Rodrigo Tocantins Calado. FAMERP-SJRP: Cecília Artico Banho, Lívia Sacchetto, Fábio Sossai Possebon, Leila Sabrina Ullmann, Cíntia Bittar, Guilherme Campos, Helena Lage Ferreira, Jorge A. Petroli Marchesi, Maisa C. Pereira Parra, Marília Moraes, Paula Rahal, Paulo Inacio da Costa, João Pessoa Araújo Jr., Maurício Lacerda Nogueira. Prefeitura de Sao Paulo: Melissa Palmieri. |
| EPI_ISL_1966389, EPI_ISL_1966390, EPI_ISL_1966391, EPI_ISL_1966392 | SECRETARIA MUNICIPAL DA SAUDE DE JOANOPOLIS | Instituto Butantan / Mendelics | Instituto Butantan: Dimas Tadeu Covas, Sandra Coccuzzo Sampaio, Maria Carolina Elias, José Salvatore Leister Patané, Vincent Louis Viala, Antonio Jorge Martins, Ricardo Haddad, Claudia Renata dos Santos Barros, Elaine Cristina Marqueze, Raul Machado Neto, Debora Botequio Moretti, Jardelina de Souza Todao Bernardino, Loyze Paola Oliveira de Lima, Luiz Aurelio de Campos Crispin. Centro de Genômica Funcional da ESALQ: Luiz Lehmann Coutinho, Ricardo Augusto Brassaloti, Raquel de Lello Rocha Campos Cassano. NGS Soluções Genômicas: Pilar Drummond Sampaio Corrêa Mariani. FZEA-USP Pirassununga: Mirele Daiana Poleti, Jessika Cristina Chagas Lesbon, Elisangela Chicaroni Mattos, Heidge Fukumasu. USP-Botucatu: Rejane Maria Tommasini Grotto, Jayme A. Souza-Neto, Guilherme Targino Valente, Patricia Akemi Assato, Felipe Allan da Silva da Costa, Bianca Cechetto Carlos. Mendelics: Bibiana Santos, João Paulo Kitajima, Erika Freitas, David Schlesinger. Hemocentro Ribeirão Preto: Simone Kashima, Evandra Strazza Rodrigues, Svetoslav Nanev Slavov, Elaine Vieira dos Santos, Rafael dos Santos Bezerra, Luiz Carlos Junior de Alcantara, Marta Giovanetti, Vagner Fonseca, Flavia Aburjaile, Rodrigo Tocantins Calado. FAMERP-SJRP: Cecília Artico Banho, Lívia Sacchetto, Fábio Sossai Possebon, Leila Sabrina Ullmann, Cíntia Bittar, Guilherme Campos, Helena Lage Ferreira, Jorge A. Petroli Marchesi, Maisa C. Pereira Parra, Marília Moraes, Paula Rahal, Paulo Inacio da Costa, João Pessoa Araújo Jr., Maurício Lacerda Nogueira. Prefeitura de Sao Paulo: Melissa Palmieri. |
| EPI_ISL_1966394 | ADECIO POMPEU CARVALHO | Instituto Butantan / Mendelics | Instituto Butantan: Dimas Tadeu Covas, Sandra Coccuzzo Sampaio, Maria Carolina Elias, José Salvatore Leister Patané, Vincent Louis Viala, Antonio Jorge Martins, Ricardo Haddad, Claudia Renata dos Santos Barros, Elaine Cristina Marqueze, Raul Machado Neto, Debora Botequio Moretti, Jardelina de Souza Todao Bernardino, Loyze Paola Oliveira de Lima, Luiz Aurelio de Campos Crispin. Centro de Genômica Funcional da ESALQ: Luiz Lehmann Coutinho, Ricardo Augusto Brassaloti, Raquel de Lello Rocha Campos Cassano. NGS Soluções Genômicas: Pilar Drummond Sampaio Corrêa Mariani. FZEA-USP Pirassununga: Mirele Daiana Poleti, Jessika Cristina Chagas Lesbon, Elisangela Chicaroni Mattos, Heidge Fukumasu. USP-Botucatu: Rejane Maria Tommasini Grotto, Jayme A. Souza-Neto, Guilherme Targino Valente, Patricia Akemi Assato, Felipe Allan da Silva da Costa, Bianca Cechetto Carlos. Mendelics: Bibiana Santos, João Paulo Kitajima, Erika Freitas, David Schlesinger. Hemocentro Ribeirão Preto: Simone Kashima, Evandra Strazza Rodrigues, Svetoslav Nanev Slavov, Elaine Vieira dos Santos, Rafael dos Santos Bezerra, Luiz Carlos Junior de Alcantara, Marta Giovanetti, Vagner Fonseca, Flavia Aburjaile, Rodrigo Tocantins Calado. FAMERP-SJRP: Cecília Artico Banho, Lívia Sacchetto, Fábio Sossai Possebon, Leila Sabrina Ullmann, Cíntia Bittar, Guilherme Campos, Helena Lage Ferreira, Jorge A. Petroli Marchesi, Maisa C. Pereira Parra, Marília Moraes, Paula Rahal, Paulo Inacio da Costa, João Pessoa Araújo Jr., Maurício Lacerda Nogueira. Prefeitura de Sao Paulo: Melissa Palmieri. |
| EPI_ISL_1966395 | SAE CTA SERV ATEND ESPEC TESTAGEM E ACONSELHAMENTO | Instituto Butantan / Mendelics | Instituto Butantan: Dimas Tadeu Covas, Sandra Coccuzzo Sampaio, Maria Carolina Elias, José Salvatore Leister Patané, Vincent Louis Viala, Antonio Jorge Martins, Ricardo Haddad, Claudia Renata dos Santos Barros, Elaine Cristina Marqueze, Raul Machado Neto, Debora Botequio Moretti, Jardelina de Souza Todao Bernardino, Loyze Paola Oliveira de Lima, Luiz Aurelio de Campos Crispin. Centro de Genômica Funcional da ESALQ: Luiz Lehmann Coutinho, Ricardo Augusto Brassaloti, Raquel de Lello Rocha Campos Cassano. NGS Soluções Genômicas: Pilar Drummond Sampaio Corrêa Mariani. FZEA-USP Pirassununga: Mirele Daiana Poleti, Jessika Cristina Chagas Lesbon, Elisangela Chicaroni Mattos, Heidge Fukumasu. USP-Botucatu: Rejane Maria Tommasini Grotto, Jayme A. Souza-Neto, Guilherme Targino Valente, Patricia Akemi Assato, Felipe Allan da Silva da Costa, Bianca Cechetto Carlos. Mendelics: Bibiana Santos, João Paulo Kitajima, Erika Freitas, David Schlesinger. Hemocentro Ribeirão Preto: Simone Kashima, Evandra Strazza Rodrigues, Svetoslav Nanev Slavov, Elaine Vieira dos Santos, Rafael dos Santos Bezerra, Luiz Carlos Junior de Alcantara, Marta Giovanetti, Vagner Fonseca, Flavia Aburjaile, Rodrigo Tocantins Calado. FAMERP-SJRP: Cecília Artico Banho, Lívia Sacchetto, Fábio Sossai Possebon, Leila Sabrina |

| | | | |
|---|---|---|---|
| | | | Ullmann, Cíntia Bittar, Guilherme Campos, Helena Lage Ferreira, Jorge A. Petroli Marchesi, Maisa C. Pereira Parra, Marília Moraes, Paula Rahal, Paulo Inacio da Costa, João Pessoa Araújo Jr., Maurício Lacerda Nogueira. Prefeitura de Sao Paulo: Melissa Palmieri. |
| EPI_ISL_1966396, EPI_ISL_1966397 | PSF DR ANTONIO PIRES DE ALMEIDA PORTO FELIZ | Instituto Butantan / Mendelics | Instituto Butantan: Dimas Tadeu Covas, Sandra Coccuzzo Sampaio, Maria Carolina Elias, José Salvatore Leister Patané, Vincent Louis Viala, Antonio Jorge Martins, Ricardo Haddad, Claudia Renata dos Santos Barros, Elaine Cristina Marqueze, Raul Machado Neto, Debora Botequio Moretti, Jardelina de Souza Todao Bernardino, Loyze Paola Oliveira de Lima, Luiz Aurelio de Campos Crispin. Centro de Genômica Funcional da ESALQ: Luiz Lehmann Coutinho, Ricardo Augusto Brassaloti, Raquel de Lello Rocha Campos Cassano. NGS Soluções Genômicas: Pilar Drummond Sampaio Corrêa Mariani. FZEA-USP Pirassununga: Mirele Daiana Poleti, Jessika Cristina Chagas Lesbon, Elisangela Chicaroni Mattos, Heidge Fukumasu. USP-Botucatu: Rejane Maria Tommasini Grotto, Jayme A. Souza-Neto, Guilherme Targino Valente, Patricia Akemi Assato, Felipe Allan da Silva da Costa, Bianca Cechetto Carlos. Mendelics: Bibiana Santos, João Paulo Kitajima, Erika Freitas, David Schlesinger. Hemocentro Ribeirão Preto: Simone Kashima, Evandra Strazza Rodrigues, Svetoslav Nanev Slavov, Elaine Vieira dos Santos, Rafael dos Santos Bezerra, Luiz Carlos Junior de Alcantara, Marta Giovanetti, Vagner Fonseca, Flavia Aburjaile, Rodrigo Tocantins Calado. FAMERP-SJRP: Cecília Artico Banho, Lívia Sacchetto, Fábio Sossai Possebon, Leila Sabrina Ullmann, Cíntia Bittar, Guilherme Campos, Helena Lage Ferreira, Jorge A. Petroli Marchesi, Maisa C. Pereira Parra, Marília Moraes, Paula Rahal, Paulo Inacio da Costa, João Pessoa Araújo Jr., Maurício Lacerda Nogueira. Prefeitura de Sao Paulo: Melissa Palmieri. |
| EPI_ISL_1966398 | CENTRO DE SAUDE DE SALMOURAO | Instituto Butantan / Mendelics | Instituto Butantan: Dimas Tadeu Covas, Sandra Coccuzzo Sampaio, Maria Carolina Elias, José Salvatore Leister Patané, Vincent Louis Viala, Antonio Jorge Martins, Ricardo Haddad, Claudia Renata dos Santos Barros, Elaine Cristina Marqueze, Raul Machado Neto, Debora Botequio Moretti, Jardelina de Souza Todao Bernardino, Loyze Paola Oliveira de Lima, Luiz Aurelio de Campos Crispin. Centro de Genômica Funcional da ESALQ: Luiz Lehmann Coutinho, Ricardo Augusto Brassaloti, Raquel de Lello Rocha Campos Cassano. NGS Soluções Genômicas: Pilar Drummond Sampaio Corrêa Mariani. FZEA-USP Pirassununga: Mirele Daiana Poleti, Jessika Cristina Chagas Lesbon, Elisangela Chicaroni Mattos, Heidge Fukumasu. USP-Botucatu: Rejane Maria Tommasini Grotto, Jayme A. Souza-Neto, Guilherme Targino Valente, Patricia Akemi Assato, Felipe Allan da Silva da Costa, Bianca Cechetto Carlos. Mendelics: Bibiana Santos, João Paulo Kitajima, Erika Freitas, David Schlesinger. Hemocentro Ribeirão Preto: Simone Kashima, Evandra Strazza Rodrigues, Svetoslav Nanev Slavov, Elaine Vieira dos Santos, Rafael dos Santos Bezerra, Luiz Carlos Junior de Alcantara, Marta Giovanetti, Vagner Fonseca, Flavia Aburjaile, Rodrigo Tocantins Calado. FAMERP-SJRP: Cecília Artico Banho, Lívia Sacchetto, Fábio Sossai Possebon, Leila Sabrina Ullmann, Cíntia Bittar, Guilherme Campos, Helena Lage Ferreira, Jorge A. Petroli Marchesi, Maisa C. Pereira Parra, Marília Moraes, Paula Rahal, Paulo Inacio da Costa, João Pessoa Araújo Jr., Maurício Lacerda Nogueira. Prefeitura de Sao Paulo: Melissa Palmieri. |
| EPI_ISL_1966399 | CENTRO DE SAUDE II | Instituto Butantan / Mendelics | Instituto Butantan: Dimas Tadeu Covas, Sandra Coccuzzo Sampaio, Maria Carolina Elias, José Salvatore Leister Patané, Vincent Louis Viala, Antonio Jorge Martins, Ricardo Haddad, Claudia Renata dos Santos Barros, Elaine Cristina Marqueze, Raul Machado Neto, Debora Botequio Moretti, Jardelina de Souza Todao Bernardino, Loyze Paola Oliveira de Lima, Luiz Aurelio de Campos Crispin. Centro de Genômica Funcional da ESALQ: Luiz Lehmann Coutinho, Ricardo Augusto Brassaloti, Raquel de Lello Rocha Campos Cassano. NGS Soluções Genômicas: Pilar Drummond Sampaio Corrêa Mariani. FZEA-USP Pirassununga: Mirele Daiana Poleti, Jessika Cristina Chagas Lesbon, Elisangela Chicaroni Mattos, Heidge Fukumasu. USP-Botucatu: Rejane Maria Tommasini Grotto, Jayme A. Souza-Neto, Guilherme Targino Valente, Patricia Akemi Assato, Felipe Allan da Silva da Costa, Bianca Cechetto Carlos. Mendelics: Bibiana Santos, João Paulo Kitajima, Erika Freitas, David Schlesinger. Hemocentro Ribeirão Preto: Simone Kashima, Evandra Strazza Rodrigues, Svetoslav Nanev Slavov, Elaine Vieira dos Santos, Rafael dos Santos Bezerra, Luiz Carlos Junior de Alcantara, Marta Giovanetti, Vagner Fonseca, Flavia Aburjaile, Rodrigo Tocantins Calado. FAMERP-SJRP: Cecília Artico Banho, Lívia Sacchetto, Fábio Sossai Possebon, Leila Sabrina Ullmann, Cíntia Bittar, Guilherme Campos, Helena Lage Ferreira, Jorge A. Petroli Marchesi, Maisa C. Pereira Parra, Marília Moraes, Paula Rahal, Paulo Inacio da Costa, João Pessoa Araújo Jr., Maurício Lacerda Nogueira. Prefeitura de Sao Paulo: Melissa Palmieri. |
| EPI_ISL_1966400 | CLINICA SALTO SAUDE SANTA CRUZ | Instituto Butantan / Mendelics | Instituto Butantan: Dimas Tadeu Covas, Sandra Coccuzzo Sampaio, Maria Carolina Elias, José Salvatore Leister Patané, Vincent Louis Viala, Antonio Jorge Martins, Ricardo Haddad, Claudia Renata dos Santos Barros, Elaine Cristina Marqueze, Raul Machado Neto, Debora Botequio Moretti, Jardelina de Souza Todao Bernardino, Loyze Paola Oliveira de Lima, Luiz Aurelio de Campos Crispin. Centro de Genômica Funcional da ESALQ: Luiz Lehmann Coutinho, Ricardo Augusto Brassaloti, Raquel de Lello Rocha Campos Cassano. NGS Soluções Genômicas: Pilar Drummond Sampaio Corrêa Mariani. FZEA-USP Pirassununga: Mirele Daiana Poleti, Jessika Cristina Chagas Lesbon, Elisangela Chicaroni Mattos, Heidge Fukumasu. USP-Botucatu: Rejane Maria Tommasini Grotto, Jayme A. Souza-Neto, Guilherme Targino Valente, Patricia Akemi Assato, Felipe Allan da Silva da Costa, Bianca Cechetto Carlos. Mendelics: Bibiana Santos, João Paulo Kitajima, Erika Freitas, David Schlesinger. Hemocentro Ribeirão Preto: Simone Kashima, Evandra Strazza Rodrigues, Svetoslav Nanev Slavov, Elaine Vieira dos Santos, Rafael dos Santos Bezerra, Luiz Carlos Junior de Alcantara, Marta Giovanetti, Vagner Fonseca, Flavia Aburjaile, Rodrigo Tocantins Calado. FAMERP-SJRP: Cecília Artico Banho, Lívia Sacchetto, Fábio Sossai Possebon, Leila Sabrina Ullmann, Cíntia Bittar, Guilherme Campos, Helena Lage Ferreira, Jorge A. Petroli Marchesi, Maisa C. Pereira Parra, Marília Moraes, Paula Rahal, Paulo Inacio da Costa, João Pessoa Araújo Jr., Maurício Lacerda Nogueira. Prefeitura de Sao Paulo: Melissa Palmieri. |
| EPI_ISL_1966401 | HOSP E MATERNIDADE MUNICIPAL N SRA MONTE SERRAT | Instituto Butantan / Mendelics | Instituto Butantan: Dimas Tadeu Covas, Sandra Coccuzzo Sampaio, Maria Carolina Elias, José Salvatore Leister Patané, Vincent Louis Viala, Antonio Jorge Martins, Ricardo Haddad, Claudia Renata dos Santos Barros, Elaine Cristina Marqueze, Raul Machado Neto, Debora Botequio Moretti, Jardelina de Souza Todao Bernardino, Loyze Paola Oliveira de Lima, Luiz Aurelio de Campos Crispin. Centro de Genômica Funcional da ESALQ: Luiz Lehmann Coutinho, Ricardo Augusto Brassaloti, Raquel de Lello Rocha Campos Cassano. NGS Soluções Genômicas: Pilar Drummond Sampaio Corrêa Mariani. FZEA-USP Pirassununga: Mirele Daiana Poleti, Jessika Cristina Chagas Lesbon, Elisangela Chicaroni Mattos, Heidge Fukumasu. USP-Botucatu: Rejane Maria Tommasini Grotto, Jayme A. Souza-Neto, Guilherme Targino Valente, Patricia Akemi Assato, Felipe Allan da Silva da Costa, Bianca Cechetto Carlos. Mendelics: Bibiana Santos, João Paulo Kitajima, Erika Freitas, David Schlesinger. Hemocentro Ribeirão Preto: Simone Kashima, Evandra Strazza Rodrigues, Svetoslav Nanev Slavov, Elaine Vieira dos Santos, Rafael dos Santos Bezerra, Luiz Carlos Junior de Alcantara, Marta Giovanetti, Vagner Fonseca, Flavia Aburjaile, Rodrigo Tocantins Calado. FAMERP-SJRP: Cecília Artico Banho, Lívia Sacchetto, Fábio Sossai Possebon, Leila Sabrina Ullmann, Cíntia Bittar, Guilherme Campos, Helena Lage Ferreira, Jorge A. Petroli Marchesi, Maisa C. Pereira Parra, Marília Moraes, Paula Rahal, Paulo Inacio da Costa, João Pessoa Araújo Jr., Maurício Lacerda Nogueira. Prefeitura de Sao Paulo: Melissa Palmieri. |
| EPI_ISL_1966403 | UNIDADE DE PRONTO ATENDIMENTO UPA I SANTA ISABEL | Instituto Butantan / Mendelics | Instituto Butantan: Dimas Tadeu Covas, Sandra Coccuzzo Sampaio, Maria Carolina Elias, José Salvatore Leister Patané, Vincent Louis Viala, Antonio Jorge Martins, Ricardo Haddad, Claudia Renata dos Santos Barros, Elaine Cristina Marqueze, Raul Machado Neto, Debora Botequio Moretti, Jardelina de Souza Todao Bernardino, Loyze Paola Oliveira de Lima, Luiz Aurelio de Campos Crispin. Centro de Genômica Funcional da ESALQ: Luiz Lehmann Coutinho, Ricardo Augusto Brassaloti, Raquel de Lello Rocha Campos Cassano. NGS Soluções Genômicas: Pilar Drummond Sampaio Corrêa Mariani. FZEA-USP Pirassununga: Mirele Daiana Poleti, Jessika Cristina Chagas Lesbon, Elisangela Chicaroni Mattos, Heidge Fukumasu. USP-Botucatu: Rejane Maria Tommasini Grotto, Jayme A. Souza-Neto, Guilherme Targino Valente, Patricia Akemi Assato, Felipe Allan da Silva da Costa, Bianca Cechetto Carlos. Mendelics: Bibiana Santos, João Paulo Kitajima, Erika Freitas, David Schlesinger. Hemocentro Ribeirão Preto: Simone Kashima, Evandra Strazza Rodrigues, Svetoslav Nanev Slavov, Elaine Vieira dos Santos, Rafael dos Santos Bezerra, Luiz Carlos Junior de Alcantara, Marta Giovanetti, Vagner Fonseca, Flavia Aburjaile, Rodrigo Tocantins Calado. FAMERP-SJRP: Cecília Artico Banho, Lívia Sacchetto, Fábio Sossai Possebon, Leila Sabrina Ullmann, Cíntia Bittar, Guilherme Campos, Helena Lage Ferreira, Jorge A. Petroli Marchesi, Maisa C. Pereira Parra, Marília Moraes, Paula Rahal, Paulo Inacio da Costa, João Pessoa Araújo Jr., Maurício Lacerda Nogueira. Prefeitura de Sao Paulo: Melissa Palmieri. |
| EPI_ISL_1966406 | HOSPITAL GERAL DE VILA NOVA CACHOEIRINHA SAO PAULO | Instituto Butantan / Mendelics | Instituto Butantan: Dimas Tadeu Covas, Sandra Coccuzzo Sampaio, Maria Carolina Elias, José Salvatore Leister Patané, Vincent Louis Viala, Antonio Jorge Martins, Ricardo Haddad, Claudia Renata dos Santos Barros, Elaine Cristina Marqueze, Raul Machado Neto, Debora Botequio Moretti, Jardelina de Souza Todao Bernardino, Loyze Paola Oliveira de Lima, Luiz Aurelio de Campos Crispin. Centro de Genômica Funcional da ESALQ: Luiz Lehmann Coutinho, Ricardo Augusto Brassaloti, Raquel de Lello Rocha Campos Cassano. NGS Soluções Genômicas: Pilar Drummond Sampaio Corrêa Mariani. FZEA-USP Pirassununga: Mirele Daiana Poleti, Jessika Cristina Chagas Lesbon, Elisangela Chicaroni Mattos, Heidge Fukumasu. USP-Botucatu: Rejane Maria Tommasini Grotto, Jayme A. Souza-Neto, Guilherme Targino Valente, Patricia Akemi Assato, Felipe Allan da Silva da Costa, Bianca Cechetto Carlos. Mendelics: Bibiana Santos, João Paulo Kitajima, Erika Freitas, David Schlesinger. Hemocentro Ribeirão Preto: Simone Kashima, Evandra Strazza Rodrigues, Svetoslav Nanev Slavov, Elaine Vieira dos Santos, Rafael dos Santos Bezerra, Luiz Carlos Junior de Alcantara, Marta Giovanetti, Vagner Fonseca, Flavia Aburjaile, Rodrigo Tocantins Calado. FAMERP-SJRP: Cecília Artico Banho, Lívia Sacchetto, Fábio Sossai Possebon, Leila Sabrina Ullmann, Cíntia Bittar, Guilherme Campos, Helena Lage Ferreira, Jorge A. Petroli Marchesi, Maisa C. Pereira Parra, Marília Moraes, Paula Rahal, Paulo Inacio da Costa, João Pessoa Araújo Jr., Maurício Lacerda Nogueira. Prefeitura de Sao Paulo: Melissa Palmieri. |
| EPI_ISL_1966407 | CENTRO MEDICO PMESP | Instituto Butantan / Mendelics | Instituto Butantan: Dimas Tadeu Covas, Sandra Coccuzzo Sampaio, Maria Carolina Elias, José Salvatore Leister Patané, Vincent Louis Viala, Antonio Jorge Martins, Ricardo Haddad, Claudia Renata dos Santos Barros, Elaine Cristina Marqueze, Raul Machado Neto, Debora Botequio Moretti, Jardelina de Souza Todao Bernardino, Loyze Paola Oliveira de Lima, Luiz Aurelio de Campos Crispin. Centro de Genômica Funcional da ESALQ: Luiz Lehmann Coutinho, Ricardo Augusto Brassaloti, Raquel de Lello Rocha Campos Cassano. NGS Soluções Genômicas: Pilar Drummond Sampaio Corrêa Mariani. |

| | | | |
|---|---|---|---|
| | | | FZEA-USP Pirassununga: Mirele Daiana Poleti, Jessika Cristina Chagas Lesbon, Elisangela Chicaroni Mattos, Heidge Fukumasu. USP-Botucatu: Rejane Maria Tommasini Grotto, Jayme A. Souza-Neto, Guilherme Targino Valente, Patricia Akemi Assato, Felipe Allan da Silva da Costa, Bianca Cechetto Carlos. Mendelics: Bibiana Santos, João Paulo Kitajima, Erika Freitas, David Schlesinger. Hemocentro Ribeirão Preto: Simone Kashima, Evandra Strazza Rodrigues, Svetoslav Nanev Slavov, Elaine Vieira dos Santos, Rafael dos Santos Bezerra, Luiz Carlos Junior de Alcantara, Marta Giovanetti, Vagner Fonseca, Flavia Aburjaile, Rodrigo Tocantins Calado. FAMERP-SJRP: Cecília Artico Banho, Lívia Sacchetto, Fábio Sossai Possebon, Leila Sabrina Ullmann, Cíntia Bittar, Guilherme Campos, Helena Lage Ferreira, Jorge A. Petroli Marchesi, Maisa C. Pereira Parra, Marília Moraes, Paula Rahal, Paulo Inacio da Costa, João Pessoa Araújo Jr., Maurício Lacerda Nogueira. Prefeitura de Sao Paulo: Melissa Palmieri. |
| EPI_ISL_1966408 | HOSP STA VIRGINIA | Instituto Butantan / Mendelics | Instituto Butantan: Dimas Tadeu Covas, Sandra Coccuzzo Sampaio, Maria Carolina Elias, José Salvatore Leister Patané, Vincent Louis Viala, Antonio Jorge Martins, Ricardo Haddad, Claudia Renata dos Santos Barros, Elaine Cristina Marqueze, Raul Machado Neto, Debora Botequio Moretti, Jardelina de Souza Todao Bernardino, Loyze Paola Oliveira de Lima, Luiz Aurelio de Campos Crispin. Centro de Genômica Funcional da ESALQ: Luiz Lehmann Coutinho, Ricardo Augusto Brassaloti, Raquel de Lello Rocha Campos Cassano. NGS Soluções Genômicas: Pilar Drummond Sampaio Corrêa Mariani. FZEA-USP Pirassununga: Mirele Daiana Poleti, Jessika Cristina Chagas Lesbon, Elisangela Chicaroni Mattos, Heidge Fukumasu. USP-Botucatu: Rejane Maria Tommasini Grotto, Jayme A. Souza-Neto, Guilherme Targino Valente, Patricia Akemi Assato, Felipe Allan da Silva da Costa, Bianca Cechetto Carlos. Mendelics: Bibiana Santos, João Paulo Kitajima, Erika Freitas, David Schlesinger. Hemocentro Ribeirão Preto: Simone Kashima, Evandra Strazza Rodrigues, Svetoslav Nanev Slavov, Elaine Vieira dos Santos, Rafael dos Santos Bezerra, Luiz Carlos Junior de Alcantara, Marta Giovanetti, Vagner Fonseca, Flavia Aburjaile, Rodrigo Tocantins Calado. FAMERP-SJRP: Cecília Artico Banho, Lívia Sacchetto, Fábio Sossai Possebon, Leila Sabrina Ullmann, Cíntia Bittar, Guilherme Campos, Helena Lage Ferreira, Jorge A. Petroli Marchesi, Maisa C. Pereira Parra, Marília Moraes, Paula Rahal, Paulo Inacio da Costa, João Pessoa Araújo Jr., Maurício Lacerda Nogueira. Prefeitura de Sao Paulo: Melissa Palmieri. |
| EPI_ISL_1966409, EPI_ISL_1966410 | HOSPITAL GERAL DE VILA NOVA CACHOEIRINHA SAO PAULO | Instituto Butantan / Mendelics | Instituto Butantan: Dimas Tadeu Covas, Sandra Coccuzzo Sampaio, Maria Carolina Elias, José Salvatore Leister Patané, Vincent Louis Viala, Antonio Jorge Martins, Ricardo Haddad, Claudia Renata dos Santos Barros, Elaine Cristina Marqueze, Raul Machado Neto, Debora Botequio Moretti, Jardelina de Souza Todao Bernardino, Loyze Paola Oliveira de Lima, Luiz Aurelio de Campos Crispin. Centro de Genômica Funcional da ESALQ: Luiz Lehmann Coutinho, Ricardo Augusto Brassaloti, Raquel de Lello Rocha Campos Cassano. NGS Soluções Genômicas: Pilar Drummond Sampaio Corrêa Mariani. FZEA-USP Pirassununga: Mirele Daiana Poleti, Jessika Cristina Chagas Lesbon, Elisangela Chicaroni Mattos, Heidge Fukumasu. USP-Botucatu: Rejane Maria Tommasini Grotto, Jayme A. Souza-Neto, Guilherme Targino Valente, Patricia Akemi Assato, Felipe Allan da Silva da Costa, Bianca Cechetto Carlos. Mendelics: Bibiana Santos, João Paulo Kitajima, Erika Freitas, David Schlesinger. Hemocentro Ribeirão Preto: Simone Kashima, Evandra Strazza Rodrigues, Svetoslav Nanev Slavov, Elaine Vieira dos Santos, Rafael dos Santos Bezerra, Luiz Carlos Junior de Alcantara, Marta Giovanetti, Vagner Fonseca, Flavia Aburjaile, Rodrigo Tocantins Calado. FAMERP-SJRP: Cecília Artico Banho, Lívia Sacchetto, Fábio Sossai Possebon, Leila Sabrina Ullmann, Cíntia Bittar, Guilherme Campos, Helena Lage Ferreira, Jorge A. Petroli Marchesi, Maisa C. Pereira Parra, Marília Moraes, Paula Rahal, Paulo Inacio da Costa, João Pessoa Araújo Jr., Maurício Lacerda Nogueira. Prefeitura de Sao Paulo: Melissa Palmieri. |
| EPI_ISL_1966412 | CENTRO DE SAUDE DE SPEDRO DO TURVO | Instituto Butantan / Mendelics | Instituto Butantan: Dimas Tadeu Covas, Sandra Coccuzzo Sampaio, Maria Carolina Elias, José Salvatore Leister Patané, Vincent Louis Viala, Antonio Jorge Martins, Ricardo Haddad, Claudia Renata dos Santos Barros, Elaine Cristina Marqueze, Raul Machado Neto, Debora Botequio Moretti, Jardelina de Souza Todao Bernardino, Loyze Paola Oliveira de Lima, Luiz Aurelio de Campos Crispin. Centro de Genômica Funcional da ESALQ: Luiz Lehmann Coutinho, Ricardo Augusto Brassaloti, Raquel de Lello Rocha Campos Cassano. NGS Soluções Genômicas: Pilar Drummond Sampaio Corrêa Mariani. FZEA-USP Pirassununga: Mirele Daiana Poleti, Jessika Cristina Chagas Lesbon, Elisangela Chicaroni Mattos, Heidge Fukumasu. USP-Botucatu: Rejane Maria Tommasini Grotto, Jayme A. Souza-Neto, Guilherme Targino Valente, Patricia Akemi Assato, Felipe Allan da Silva da Costa, Bianca Cechetto Carlos. Mendelics: Bibiana Santos, João Paulo Kitajima, Erika Freitas, David Schlesinger. Hemocentro Ribeirão Preto: Simone Kashima, Evandra Strazza Rodrigues, Svetoslav Nanev Slavov, Elaine Vieira dos Santos, Rafael dos Santos Bezerra, Luiz Carlos Junior de Alcantara, Marta Giovanetti, Vagner Fonseca, Flavia Aburjaile, Rodrigo Tocantins Calado. FAMERP-SJRP: Cecília Artico Banho, Lívia Sacchetto, Fábio Sossai Possebon, Leila Sabrina Ullmann, Cíntia Bittar, Guilherme Campos, Helena Lage Ferreira, Jorge A. Petroli Marchesi, Maisa C. Pereira Parra, Marília Moraes, Paula Rahal, Paulo Inacio da Costa, João Pessoa Araújo Jr., Maurício Lacerda Nogueira. Prefeitura de Sao Paulo: Melissa Palmieri. |
| EPI_ISL_1966413, EPI_ISL_1966415, EPI_ISL_1966416, EPI_ISL_1966417, EPI_ISL_1966418, EPI_ISL_1966419, EPI_ISL_1966420, EPI_ISL_1966421, EPI_ISL_1966422, EPI_ISL_1966423, EPI_ISL_1966424, EPI_ISL_1966425, EPI_ISL_1966426, EPI_ISL_1966427, EPI_ISL_1966428, EPI_ISL_1966429, EPI_ISL_1966430, EPI_ISL_1966431, EPI_ISL_1966432 | | | |
| see above | PSF PARQUE EMILIA | Instituto Butantan / Mendelics | Instituto Butantan: Dimas Tadeu Covas, Sandra Coccuzzo Sampaio, Maria Carolina Elias, José Salvatore Leister Patané, Vincent Louis Viala, Antonio Jorge Martins, Ricardo Haddad, Claudia Renata dos Santos Barros, Elaine Cristina Marqueze, Raul Machado Neto, Debora Botequio Moretti, Jardelina de Souza Todao Bernardino, Loyze Paola Oliveira de Lima, Luiz Aurelio de Campos Crispin. Centro de Genômica Funcional da ESALQ: Luiz Lehmann Coutinho, Ricardo Augusto Brassaloti, Raquel de Lello Rocha Campos Cassano. NGS Soluções Genômicas: Pilar Drummond Sampaio Corrêa Mariani. FZEA-USP Pirassununga: Mirele Daiana Poleti, Jessika Cristina Chagas Lesbon, Elisangela Chicaroni Mattos, Heidge Fukumasu. USP-Botucatu: Rejane Maria Tommasini Grotto, Jayme A. Souza-Neto, Guilherme Targino Valente, Patricia Akemi Assato, Felipe Allan da Silva da Costa, Bianca Cechetto Carlos. Mendelics: Bibiana Santos, João Paulo Kitajima, Erika Freitas, David Schlesinger. Hemocentro Ribeirão Preto: Simone Kashima, Evandra Strazza Rodrigues, Svetoslav Nanev Slavov, Elaine Vieira dos Santos, Rafael dos Santos Bezerra, Luiz Carlos Junior de Alcantara, Marta Giovanetti, Vagner Fonseca, Flavia Aburjaile, Rodrigo Tocantins Calado. FAMERP-SJRP: Cecília Artico Banho, Lívia Sacchetto, Fábio Sossai Possebon, Leila Sabrina Ullmann, Cíntia Bittar, Guilherme Campos, Helena Lage Ferreira, Jorge A. Petroli Marchesi, Maisa C. Pereira Parra, Marília Moraes, Paula Rahal, Paulo Inacio da Costa, João Pessoa Araújo Jr., Maurício Lacerda Nogueira. Prefeitura de Sao Paulo: Melissa Palmieri. |
| EPI_ISL_1966434, EPI_ISL_1966435, EPI_ISL_1966436, EPI_ISL_1966437, EPI_ISL_1966438 | UPA DR AKIRA TADA | Instituto Butantan / Mendelics | Instituto Butantan: Dimas Tadeu Covas, Sandra Coccuzzo Sampaio, Maria Carolina Elias, José Salvatore Leister Patané, Vincent Louis Viala, Antonio Jorge Martins, Ricardo Haddad, Claudia Renata dos Santos Barros, Elaine Cristina Marqueze, Raul Machado Neto, Debora Botequio Moretti, Jardelina de Souza Todao Bernardino, Loyze Paola Oliveira de Lima, Luiz Aurelio de Campos Crispin. Centro de Genômica Funcional da ESALQ: Luiz Lehmann Coutinho, Ricardo Augusto Brassaloti, Raquel de Lello Rocha Campos Cassano. NGS Soluções Genômicas: Pilar Drummond Sampaio Corrêa Mariani. FZEA-USP Pirassununga: Mirele Daiana Poleti, Jessika Cristina Chagas Lesbon, Elisangela Chicaroni Mattos, Heidge Fukumasu. USP-Botucatu: Rejane Maria Tommasini Grotto, Jayme A. Souza-Neto, Guilherme Targino Valente, Patricia Akemi Assato, Felipe Allan da Silva da Costa, Bianca Cechetto Carlos. Mendelics: Bibiana Santos, João Paulo Kitajima, Erika Freitas, David Schlesinger. Hemocentro Ribeirão Preto: Simone Kashima, Evandra Strazza Rodrigues, Svetoslav Nanev Slavov, Elaine Vieira dos Santos, Rafael dos Santos Bezerra, Luiz Carlos Junior de Alcantara, Marta Giovanetti, Vagner Fonseca, Flavia Aburjaile, Rodrigo Tocantins Calado. FAMERP-SJRP: Cecília Artico Banho, Lívia Sacchetto, Fábio Sossai Possebon, Leila Sabrina Ullmann, Cíntia Bittar, Guilherme Campos, Helena Lage Ferreira, Jorge A. Petroli Marchesi, Maisa C. Pereira Parra, Marília Moraes, Paula Rahal, Paulo Inacio da Costa, João Pessoa Araújo Jr., Maurício Lacerda Nogueira. Prefeitura de Sao Paulo: Melissa Palmieri. |
| EPI_ISL_1966439 | DEPARTAMENTO DE SAUDE COLETIVA | Instituto Butantan / Mendelics | Instituto Butantan: Dimas Tadeu Covas, Sandra Coccuzzo Sampaio, Maria Carolina Elias, José Salvatore Leister Patané, Vincent Louis Viala, Antonio Jorge Martins, Ricardo Haddad, Claudia Renata dos Santos Barros, Elaine Cristina Marqueze, Raul Machado Neto, Debora Botequio Moretti, Jardelina de Souza Todao Bernardino, Loyze Paola Oliveira de Lima, Luiz Aurelio de Campos Crispin. Centro de Genômica Funcional da ESALQ: Luiz Lehmann Coutinho, Ricardo Augusto Brassaloti, Raquel de Lello Rocha Campos Cassano. NGS Soluções Genômicas: Pilar Drummond Sampaio Corrêa Mariani. FZEA-USP Pirassununga: Mirele Daiana Poleti, Jessika Cristina Chagas Lesbon, Elisangela Chicaroni Mattos, Heidge Fukumasu. USP-Botucatu: Rejane Maria Tommasini Grotto, Jayme A. Souza-Neto, Guilherme Targino Valente, Patricia Akemi Assato, Felipe Allan da Silva da Costa, Bianca Cechetto Carlos. Mendelics: Bibiana Santos, João Paulo Kitajima, Erika Freitas, David Schlesinger. Hemocentro Ribeirão Preto: Simone Kashima, Evandra Strazza Rodrigues, Svetoslav Nanev Slavov, Elaine Vieira dos Santos, Rafael dos Santos Bezerra, Luiz Carlos Junior de Alcantara, Marta Giovanetti, Vagner Fonseca, Flavia Aburjaile, Rodrigo Tocantins Calado. FAMERP-SJRP: Cecília Artico Banho, Lívia Sacchetto, Fábio Sossai Possebon, Leila Sabrina Ullmann, Cíntia Bittar, Guilherme Campos, Helena Lage Ferreira, Jorge A. Petroli Marchesi, Maisa C. Pereira Parra, Marília Moraes, Paula Rahal, Paulo Inacio da Costa, João Pessoa Araújo Jr., Maurício Lacerda Nogueira. Prefeitura de Sao Paulo: Melissa Palmieri. |
| EPI_ISL_1966440, EPI_ISL_1966441, EPI_ISL_1966443 | UBS DR ALFREDO DANTAS DE SOUZA UMUARAMA | Instituto Butantan / Mendelics | Instituto Butantan: Dimas Tadeu Covas, Sandra Coccuzzo Sampaio, Maria Carolina Elias, José Salvatore Leister Patané, Vincent Louis Viala, Antonio Jorge Martins, Ricardo Haddad, Claudia Renata dos Santos Barros, Elaine Cristina Marqueze, Raul Machado Neto, Debora Botequio Moretti, Jardelina de Souza Todao Bernardino, Loyze Paola Oliveira de Lima, Luiz Aurelio de Campos Crispin. Centro de Genômica Funcional da ESALQ: Luiz Lehmann Coutinho, Ricardo Augusto Brassaloti, Raquel de Lello Rocha Campos Cassano. NGS Soluções Genômicas: Pilar Drummond Sampaio Corrêa Mariani. FZEA-USP Pirassununga: Mirele Daiana Poleti, Jessika Cristina Chagas Lesbon, Elisangela Chicaroni Mattos, Heidge Fukumasu. USP-Botucatu: Rejane Maria Tommasini Grotto, Jayme A. Souza-Neto, Guilherme Targino Valente, Patricia Akemi Assato, Felipe Allan da Silva da Costa, Bianca Cechetto Carlos. Mendelics: Bibiana Santos, João Paulo Kitajima, Erika Freitas, David Schlesinger. Hemocentro Ribeirão Preto: Simone Kashima, Evandra Strazza Rodrigues, Svetoslav Nanev Slavov, Elaine Vieira dos Santos, Rafael dos Santos Bezerra, Luiz Carlos Junior de Alcantara, Marta Giovanetti, Vagner |

| | | | |
|---|---|---|---|
| | | | Fonseca, Flavia Aburjaile, Rodrigo Tocantins Calado. FAMERP-SJRP: Cecília Artico Banho, Lívia Sacchetto, Fábio Sossai Possebon, Leila Sabrina Ullmann, Cíntia Bittar, Guilherme Campos, Helena Lage Ferreira, Jorge A. Petroli Marchesi, Maisa C. Pereira Parra, Marília Moraes, Paula Rahal, Paulo Inacio da Costa, João Pessoa Araújo Jr., Maurício Lacerda Nogueira. Prefeitura de Sao Paulo: Melissa Palmieri. |
| EPI_ISL_1966444 | UBS DE AURIFLAMA | Instituto Butantan / Mendelics | Instituto Butantan: Dimas Tadeu Covas, Sandra Coccuzzo Sampaio, Maria Carolina Elias, José Salvatore Leister Patané, Vincent Louis Viala, Antonio Jorge Martins, Ricardo Haddad, Claudia Renata dos Santos Barros, Elaine Cristina Marqueze, Raul Machado Neto, Debora Botequio Moretti, Jardelina de Souza Todao Bernardino, Loyze Paola Oliveira de Lima, Luiz Aurelio de Campos Crispin. Centro de Genômica Funcional da ESALQ: Luiz Lehmann Coutinho, Ricardo Augusto Brassaloti, Raquel de Lello Rocha Campos Cassano. NGS Soluções Genômicas: Pilar Drummond Sampaio Corrêa Mariani. FZEA-USP Pirassununga: Mirele Daiana Poleti, Jessika Cristina Chagas Lesbon, Elisangela Chicaroni Mattos, Heidge Fukumasu. USP-Botucatu: Rejane Maria Tommasini Grotto, Jayme A. Souza-Neto, Guilherme Targino Valente, Patricia Akemi Assato, Felipe Allan da Silva da Costa, Bianca Cechetto Carlos. Mendelics: Bibiana Santos, João Paulo Kitajima, Erika Freitas, David Schlesinger. Hemocentro Ribeirão Preto: Simone Kashima, Evandra Strazza Rodrigues, Svetoslav Nanev Slavov, Elaine Vieira dos Santos, Rafael dos Santos Bezerra, Luiz Carlos Junior de Alcantara, Marta Giovanetti, Vagner Fonseca, Flavia Aburjaile, Rodrigo Tocantins Calado. FAMERP-SJRP: Cecília Artico Banho, Lívia Sacchetto, Fábio Sossai Possebon, Leila Sabrina Ullmann, Cíntia Bittar, Guilherme Campos, Helena Lage Ferreira, Jorge A. Petroli Marchesi, Maisa C. Pereira Parra, Marília Moraes, Paula Rahal, Paulo Inacio da Costa, João Pessoa Araújo Jr., Maurício Lacerda Nogueira. Prefeitura de Sao Paulo: Melissa Palmieri. |
| EPI_ISL_1966445 | UBS DR ALLY ALAHMAR DE BARRETOS | Instituto Butantan / Mendelics | Instituto Butantan: Dimas Tadeu Covas, Sandra Coccuzzo Sampaio, Maria Carolina Elias, José Salvatore Leister Patané, Vincent Louis Viala, Antonio Jorge Martins, Ricardo Haddad, Claudia Renata dos Santos Barros, Elaine Cristina Marqueze, Raul Machado Neto, Debora Botequio Moretti, Jardelina de Souza Todao Bernardino, Loyze Paola Oliveira de Lima, Luiz Aurelio de Campos Crispin. Centro de Genômica Funcional da ESALQ: Luiz Lehmann Coutinho, Ricardo Augusto Brassaloti, Raquel de Lello Rocha Campos Cassano. NGS Soluções Genômicas: Pilar Drummond Sampaio Corrêa Mariani. FZEA-USP Pirassununga: Mirele Daiana Poleti, Jessika Cristina Chagas Lesbon, Elisangela Chicaroni Mattos, Heidge Fukumasu. USP-Botucatu: Rejane Maria Tommasini Grotto, Jayme A. Souza-Neto, Guilherme Targino Valente, Patricia Akemi Assato, Felipe Allan da Silva da Costa, Bianca Cechetto Carlos. Mendelics: Bibiana Santos, João Paulo Kitajima, Erika Freitas, David Schlesinger. Hemocentro Ribeirão Preto: Simone Kashima, Evandra Strazza Rodrigues, Svetoslav Nanev Slavov, Elaine Vieira dos Santos, Rafael dos Santos Bezerra, Luiz Carlos Junior de Alcantara, Marta Giovanetti, Vagner Fonseca, Flavia Aburjaile, Rodrigo Tocantins Calado. FAMERP-SJRP: Cecília Artico Banho, Lívia Sacchetto, Fábio Sossai Possebon, Leila Sabrina Ullmann, Cíntia Bittar, Guilherme Campos, Helena Lage Ferreira, Jorge A. Petroli Marchesi, Maisa C. Pereira Parra, Marília Moraes, Paula Rahal, Paulo Inacio da Costa, João Pessoa Araújo Jr., Maurício Lacerda Nogueira. Prefeitura de Sao Paulo: Melissa Palmieri. |
| EPI_ISL_1966446, EPI_ISL_1966448, EPI_ISL_1966456, EPI_ISL_1966458 | NUCLEO DE SAUDE VILA FALCAO DE BAURU | Instituto Butantan / Mendelics | Instituto Butantan: Dimas Tadeu Covas, Sandra Coccuzzo Sampaio, Maria Carolina Elias, José Salvatore Leister Patané, Vincent Louis Viala, Antonio Jorge Martins, Ricardo Haddad, Claudia Renata dos Santos Barros, Elaine Cristina Marqueze, Raul Machado Neto, Debora Botequio Moretti, Jardelina de Souza Todao Bernardino, Loyze Paola Oliveira de Lima, Luiz Aurelio de Campos Crispin. Centro de Genômica Funcional da ESALQ: Luiz Lehmann Coutinho, Ricardo Augusto Brassaloti, Raquel de Lello Rocha Campos Cassano. NGS Soluções Genômicas: Pilar Drummond Sampaio Corrêa Mariani. FZEA-USP Pirassununga: Mirele Daiana Poleti, Jessika Cristina Chagas Lesbon, Elisangela Chicaroni Mattos, Heidge Fukumasu. USP-Botucatu: Rejane Maria Tommasini Grotto, Jayme A. Souza-Neto, Guilherme Targino Valente, Patricia Akemi Assato, Felipe Allan da Silva da Costa, Bianca Cechetto Carlos. Mendelics: Bibiana Santos, João Paulo Kitajima, Erika Freitas, David Schlesinger. Hemocentro Ribeirão Preto: Simone Kashima, Evandra Strazza Rodrigues, Svetoslav Nanev Slavov, Elaine Vieira dos Santos, Rafael dos Santos Bezerra, Luiz Carlos Junior de Alcantara, Marta Giovanetti, Vagner Fonseca, Flavia Aburjaile, Rodrigo Tocantins Calado. FAMERP-SJRP: Cecília Artico Banho, Lívia Sacchetto, Fábio Sossai Possebon, Leila Sabrina Ullmann, Cíntia Bittar, Guilherme Campos, Helena Lage Ferreira, Jorge A. Petroli Marchesi, Maisa C. Pereira Parra, Marília Moraes, Paula Rahal, Paulo Inacio da Costa, João Pessoa Araújo Jr., Maurício Lacerda Nogueira. Prefeitura de Sao Paulo: Melissa Palmieri. |
| EPI_ISL_1966460, EPI_ISL_1966461, EPI_ISL_1966462 | UBS IV GUARARAPES | Instituto Butantan / Mendelics | Instituto Butantan: Dimas Tadeu Covas, Sandra Coccuzzo Sampaio, Maria Carolina Elias, José Salvatore Leister Patané, Vincent Louis Viala, Antonio Jorge Martins, Ricardo Haddad, Claudia Renata dos Santos Barros, Elaine Cristina Marqueze, Raul Machado Neto, Debora Botequio Moretti, Jardelina de Souza Todao Bernardino, Loyze Paola Oliveira de Lima, Luiz Aurelio de Campos Crispin. Centro de Genômica Funcional da ESALQ: Luiz Lehmann Coutinho, Ricardo Augusto Brassaloti, Raquel de Lello Rocha Campos Cassano. NGS Soluções Genômicas: Pilar Drummond Sampaio Corrêa Mariani. FZEA-USP Pirassununga: Mirele Daiana Poleti, Jessika Cristina Chagas Lesbon, Elisangela Chicaroni Mattos, Heidge Fukumasu. USP-Botucatu: Rejane Maria Tommasini Grotto, Jayme A. Souza-Neto, Guilherme Targino Valente, Patricia Akemi Assato, Felipe Allan da Silva da Costa, Bianca Cechetto Carlos. Mendelics: Bibiana Santos, João Paulo Kitajima, Erika Freitas, David Schlesinger. Hemocentro Ribeirão Preto: Simone Kashima, Evandra Strazza Rodrigues, Svetoslav Nanev Slavov, Elaine Vieira dos Santos, Rafael dos Santos Bezerra, Luiz Carlos Junior de Alcantara, Marta Giovanetti, Vagner Fonseca, Flavia Aburjaile, Rodrigo Tocantins Calado. FAMERP-SJRP: Cecília Artico Banho, Lívia Sacchetto, Fábio Sossai Possebon, Leila Sabrina Ullmann, Cíntia Bittar, Guilherme Campos, Helena Lage Ferreira, Jorge A. Petroli Marchesi, Maisa C. Pereira Parra, Marília Moraes, Paula Rahal, Paulo Inacio da Costa, João Pessoa Araújo Jr., Maurício Lacerda Nogueira. Prefeitura de Sao Paulo: Melissa Palmieri. |
| EPI_ISL_1966463 | CENTRO DE SAUDE II IBITINGA | Instituto Butantan / Mendelics | Instituto Butantan: Dimas Tadeu Covas, Sandra Coccuzzo Sampaio, Maria Carolina Elias, José Salvatore Leister Patané, Vincent Louis Viala, Antonio Jorge Martins, Ricardo Haddad, Claudia Renata dos Santos Barros, Elaine Cristina Marqueze, Raul Machado Neto, Debora Botequio Moretti, Jardelina de Souza Todao Bernardino, Loyze Paola Oliveira de Lima, Luiz Aurelio de Campos Crispin. Centro de Genômica Funcional da ESALQ: Luiz Lehmann Coutinho, Ricardo Augusto Brassaloti, Raquel de Lello Rocha Campos Cassano. NGS Soluções Genômicas: Pilar Drummond Sampaio Corrêa Mariani. FZEA-USP Pirassununga: Mirele Daiana Poleti, Jessika Cristina Chagas Lesbon, Elisangela Chicaroni Mattos, Heidge Fukumasu. USP-Botucatu: Rejane Maria Tommasini Grotto, Jayme A. Souza-Neto, Guilherme Targino Valente, Patricia Akemi Assato, Felipe Allan da Silva da Costa, Bianca Cechetto Carlos. Mendelics: Bibiana Santos, João Paulo Kitajima, Erika Freitas, David Schlesinger. Hemocentro Ribeirão Preto: Simone Kashima, Evandra Strazza Rodrigues, Svetoslav Nanev Slavov, Elaine Vieira dos Santos, Rafael dos Santos Bezerra, Luiz Carlos Junior de Alcantara, Marta Giovanetti, Vagner Fonseca, Flavia Aburjaile, Rodrigo Tocantins Calado. FAMERP-SJRP: Cecília Artico Banho, Lívia Sacchetto, Fábio Sossai Possebon, Leila Sabrina Ullmann, Cíntia Bittar, Guilherme Campos, Helena Lage Ferreira, Jorge A. Petroli Marchesi, Maisa C. Pereira Parra, Marília Moraes, Paula Rahal, Paulo Inacio da Costa, João Pessoa Araújo Jr., Maurício Lacerda Nogueira. Prefeitura de Sao Paulo: Melissa Palmieri. |
| EPI_ISL_1966464, EPI_ISL_1966465 | POLICLINICA COVID 19 ITAPETININGA | Instituto Butantan / Mendelics | Instituto Butantan: Dimas Tadeu Covas, Sandra Coccuzzo Sampaio, Maria Carolina Elias, José Salvatore Leister Patané, Vincent Louis Viala, Antonio Jorge Martins, Ricardo Haddad, Claudia Renata dos Santos Barros, Elaine Cristina Marqueze, Raul Machado Neto, Debora Botequio Moretti, Jardelina de Souza Todao Bernardino, Loyze Paola Oliveira de Lima, Luiz Aurelio de Campos Crispin. Centro de Genômica Funcional da ESALQ: Luiz Lehmann Coutinho, Ricardo Augusto Brassaloti, Raquel de Lello Rocha Campos Cassano. NGS Soluções Genômicas: Pilar Drummond Sampaio Corrêa Mariani. FZEA-USP Pirassununga: Mirele Daiana Poleti, Jessika Cristina Chagas Lesbon, Elisangela Chicaroni Mattos, Heidge Fukumasu. USP-Botucatu: Rejane Maria Tommasini Grotto, Jayme A. Souza-Neto, Guilherme Targino Valente, Patricia Akemi Assato, Felipe Allan da Silva da Costa, Bianca Cechetto Carlos. Mendelics: Bibiana Santos, João Paulo Kitajima, Erika Freitas, David Schlesinger. Hemocentro Ribeirão Preto: Simone Kashima, Evandra Strazza Rodrigues, Svetoslav Nanev Slavov, Elaine Vieira dos Santos, Rafael dos Santos Bezerra, Luiz Carlos Junior de Alcantara, Marta Giovanetti, Vagner Fonseca, Flavia Aburjaile, Rodrigo Tocantins Calado. FAMERP-SJRP: Cecília Artico Banho, Lívia Sacchetto, Fábio Sossai Possebon, Leila Sabrina Ullmann, Cíntia Bittar, Guilherme Campos, Helena Lage Ferreira, Jorge A. Petroli Marchesi, Maisa C. Pereira Parra, Marília Moraes, Paula Rahal, Paulo Inacio da Costa, João Pessoa Araújo Jr., Maurício Lacerda Nogueira. Prefeitura de Sao Paulo: Melissa Palmieri. |
| EPI_ISL_1966466 | HOSPITAL REGIONAL DE ITAPETININGA | Instituto Butantan / Mendelics | Instituto Butantan: Dimas Tadeu Covas, Sandra Coccuzzo Sampaio, Maria Carolina Elias, José Salvatore Leister Patané, Vincent Louis Viala, Antonio Jorge Martins, Ricardo Haddad, Claudia Renata dos Santos Barros, Elaine Cristina Marqueze, Raul Machado Neto, Debora Botequio Moretti, Jardelina de Souza Todao Bernardino, Loyze Paola Oliveira de Lima, Luiz Aurelio de Campos Crispin. Centro de Genômica Funcional da ESALQ: Luiz Lehmann Coutinho, Ricardo Augusto Brassaloti, Raquel de Lello Rocha Campos Cassano. NGS Soluções Genômicas: Pilar Drummond Sampaio Corrêa Mariani. FZEA-USP Pirassununga: Mirele Daiana Poleti, Jessika Cristina Chagas Lesbon, Elisangela Chicaroni Mattos, Heidge Fukumasu. USP-Botucatu: Rejane Maria Tommasini Grotto, Jayme A. Souza-Neto, Guilherme Targino Valente, Patricia Akemi Assato, Felipe Allan da Silva da Costa, Bianca Cechetto Carlos. Mendelics: Bibiana Santos, João Paulo Kitajima, Erika Freitas, David Schlesinger. Hemocentro Ribeirão Preto: Simone Kashima, Evandra Strazza Rodrigues, Svetoslav Nanev Slavov, Elaine Vieira dos Santos, Rafael dos Santos Bezerra, Luiz Carlos Junior de Alcantara, Marta Giovanetti, Vagner Fonseca, Flavia Aburjaile, Rodrigo Tocantins Calado. FAMERP-SJRP: Cecília Artico Banho, Lívia Sacchetto, Fábio Sossai Possebon, Leila Sabrina Ullmann, Cíntia Bittar, Guilherme Campos, Helena Lage Ferreira, Jorge A. Petroli Marchesi, Maisa C. Pereira Parra, Marília Moraes, Paula Rahal, Paulo Inacio da Costa, João Pessoa Araújo Jr., Maurício Lacerda Nogueira. Prefeitura de Sao Paulo: Melissa Palmieri. |
| EPI_ISL_1966467 | POLICLINICA COVID 19 ITAPETININGA | Instituto Butantan / Mendelics | Instituto Butantan: Dimas Tadeu Covas, Sandra Coccuzzo Sampaio, Maria Carolina Elias, José Salvatore Leister Patané, Vincent Louis Viala, Antonio Jorge Martins, Ricardo Haddad, Claudia Renata dos Santos Barros, Elaine Cristina Marqueze, Raul Machado Neto, Debora Botequio Moretti, Jardelina de Souza Todao Bernardino, Loyze Paola Oliveira de Lima, Luiz Aurelio de Campos Crispin. Centro de Genômica Funcional da ESALQ: Luiz Lehmann |

| | | | |
|---|---|---|---|
| | | | Coutinho, Ricardo Augusto Brassaloti, Raquel de Lello Rocha Campos Cassano. NGS Soluções Genômicas: Pilar Drummond Sampaio Corrêa Mariani. FZEA-USP Pirassununga: Mirele Daiana Poleti, Jessika Cristina Chagas Lesbon, Elisangela Chicaroni Mattos, Heidge Fukumasu. USP-Botucatu: Rejane Maria Tommasini Grotto, Jayme A. Souza-Neto, Guilherme Targino Valente, Patricia Akemi Assato, Felipe Allan da Silva da Costa, Bianca Cechetto Carlos. Mendelics: Bibiana Santos, João Paulo Kitajima, Erika Freitas, David Schlesinger. Hemocentro Ribeirão Preto: Simone Kashima, Evandra Strazza Rodrigues, Svetoslav Nanev Slavov, Elaine Vieira dos Santos, Rafael dos Santos Bezerra, Luiz Carlos Junior de Alcantara, Marta Giovanetti, Vagner Fonseca, Flavia Aburjaile, Rodrigo Tocantins Calado. FAMERP-SJRP: Cecília Artico Banho, Lívia Sacchetto, Fábio Sossai Possebon, Leila Sabrina Ullmann, Cíntia Bittar, Guilherme Campos, Helena Lage Ferreira, Jorge A. Petroli Marchesi, Maisa C. Pereira Parra, Marília Moraes, Paula Rahal, Paulo Inacio da Costa, João Pessoa Araújo Jr., Maurício Lacerda Nogueira. Prefeitura de Sao Paulo: Melissa Palmieri. |
| EPI_ISL_1966469, EPI_ISL_1966470 | UBS II DA FAMILIA SEGISFREDO MAMEDE PINTO LOURDES | Instituto Butantan / Mendelics | Instituto Butantan: Dimas Tadeu Covas, Sandra Coccuzzo Sampaio, Maria Carolina Elias, José Salvatore Leister Patané, Vincent Louis Viala, Antonio Jorge Martins, Ricardo Haddad, Claudia Renata dos Santos Barros, Elaine Cristina Marqueze, Raul Machado Neto, Debora Botequio Moretti, Jardelina de Souza Todao Bernardino, Loyze Paola Oliveira de Lima, Luiz Aurelio de Campos Crispin. Centro de Genômica Funcional da ESALQ: Luiz Lehmann Coutinho, Ricardo Augusto Brassaloti, Raquel de Lello Rocha Campos Cassano. NGS Soluções Genômicas: Pilar Drummond Sampaio Corrêa Mariani. FZEA-USP Pirassununga: Mirele Daiana Poleti, Jessika Cristina Chagas Lesbon, Elisangela Chicaroni Mattos, Heidge Fukumasu. USP-Botucatu: Rejane Maria Tommasini Grotto, Jayme A. Souza-Neto, Guilherme Targino Valente, Patricia Akemi Assato, Felipe Allan da Silva da Costa, Bianca Cechetto Carlos. Mendelics: Bibiana Santos, João Paulo Kitajima, Erika Freitas, David Schlesinger. Hemocentro Ribeirão Preto: Simone Kashima, Evandra Strazza Rodrigues, Svetoslav Nanev Slavov, Elaine Vieira dos Santos, Rafael dos Santos Bezerra, Luiz Carlos Junior de Alcantara, Marta Giovanetti, Vagner Fonseca, Flavia Aburjaile, Rodrigo Tocantins Calado. FAMERP-SJRP: Cecília Artico Banho, Lívia Sacchetto, Fábio Sossai Possebon, Leila Sabrina Ullmann, Cíntia Bittar, Guilherme Campos, Helena Lage Ferreira, Jorge A. Petroli Marchesi, Maisa C. Pereira Parra, Marília Moraes, Paula Rahal, Paulo Inacio da Costa, João Pessoa Araújo Jr., Maurício Lacerda Nogueira. Prefeitura de Sao Paulo: Melissa Palmieri. |
| EPI_ISL_1966471 | CENTRO DE SAUDE II MAIRINQUE MAIRINQUE | Instituto Butantan / Mendelics | Instituto Butantan: Dimas Tadeu Covas, Sandra Coccuzzo Sampaio, Maria Carolina Elias, José Salvatore Leister Patané, Vincent Louis Viala, Antonio Jorge Martins, Ricardo Haddad, Claudia Renata dos Santos Barros, Elaine Cristina Marqueze, Raul Machado Neto, Debora Botequio Moretti, Jardelina de Souza Todao Bernardino, Loyze Paola Oliveira de Lima, Luiz Aurelio de Campos Crispin. Centro de Genômica Funcional da ESALQ: Luiz Lehmann Coutinho, Ricardo Augusto Brassaloti, Raquel de Lello Rocha Campos Cassano. NGS Soluções Genômicas: Pilar Drummond Sampaio Corrêa Mariani. FZEA-USP Pirassununga: Mirele Daiana Poleti, Jessika Cristina Chagas Lesbon, Elisangela Chicaroni Mattos, Heidge Fukumasu. USP-Botucatu: Rejane Maria Tommasini Grotto, Jayme A. Souza-Neto, Guilherme Targino Valente, Patricia Akemi Assato, Felipe Allan da Silva da Costa, Bianca Cechetto Carlos. Mendelics: Bibiana Santos, João Paulo Kitajima, Erika Freitas, David Schlesinger. Hemocentro Ribeirão Preto: Simone Kashima, Evandra Strazza Rodrigues, Svetoslav Nanev Slavov, Elaine Vieira dos Santos, Rafael dos Santos Bezerra, Luiz Carlos Junior de Alcantara, Marta Giovanetti, Vagner Fonseca, Flavia Aburjaile, Rodrigo Tocantins Calado. FAMERP-SJRP: Cecília Artico Banho, Lívia Sacchetto, Fábio Sossai Possebon, Leila Sabrina Ullmann, Cíntia Bittar, Guilherme Campos, Helena Lage Ferreira, Jorge A. Petroli Marchesi, Maisa C. Pereira Parra, Marília Moraes, Paula Rahal, Paulo Inacio da Costa, João Pessoa Araújo Jr., Maurício Lacerda Nogueira. Prefeitura de Sao Paulo: Melissa Palmieri. |
| EPI_ISL_1966472, EPI_ISL_1966473 | CENTRO DE SAUDE II DR ALCIDES FACUNDO ARROYO | Instituto Butantan / Mendelics | Instituto Butantan: Dimas Tadeu Covas, Sandra Coccuzzo Sampaio, Maria Carolina Elias, José Salvatore Leister Patané, Vincent Louis Viala, Antonio Jorge Martins, Ricardo Haddad, Claudia Renata dos Santos Barros, Elaine Cristina Marqueze, Raul Machado Neto, Debora Botequio Moretti, Jardelina de Souza Todao Bernardino, Loyze Paola Oliveira de Lima, Luiz Aurelio de Campos Crispin. Centro de Genômica Funcional da ESALQ: Luiz Lehmann Coutinho, Ricardo Augusto Brassaloti, Raquel de Lello Rocha Campos Cassano. NGS Soluções Genômicas: Pilar Drummond Sampaio Corrêa Mariani. FZEA-USP Pirassununga: Mirele Daiana Poleti, Jessika Cristina Chagas Lesbon, Elisangela Chicaroni Mattos, Heidge Fukumasu. USP-Botucatu: Rejane Maria Tommasini Grotto, Jayme A. Souza-Neto, Guilherme Targino Valente, Patricia Akemi Assato, Felipe Allan da Silva da Costa, Bianca Cechetto Carlos. Mendelics: Bibiana Santos, João Paulo Kitajima, Erika Freitas, David Schlesinger. Hemocentro Ribeirão Preto: Simone Kashima, Evandra Strazza Rodrigues, Svetoslav Nanev Slavov, Elaine Vieira dos Santos, Rafael dos Santos Bezerra, Luiz Carlos Junior de Alcantara, Marta Giovanetti, Vagner Fonseca, Flavia Aburjaile, Rodrigo Tocantins Calado. FAMERP-SJRP: Cecília Artico Banho, Lívia Sacchetto, Fábio Sossai Possebon, Leila Sabrina Ullmann, Cíntia Bittar, Guilherme Campos, Helena Lage Ferreira, Jorge A. Petroli Marchesi, Maisa C. Pereira Parra, Marília Moraes, Paula Rahal, Paulo Inacio da Costa, João Pessoa Araújo Jr., Maurício Lacerda Nogueira. Prefeitura de Sao Paulo: Melissa Palmieri. |
| EPI_ISL_1966474 | PAS SILVIA COVAS | Instituto Butantan / Mendelics | Instituto Butantan: Dimas Tadeu Covas, Sandra Coccuzzo Sampaio, Maria Carolina Elias, José Salvatore Leister Patané, Vincent Louis Viala, Antonio Jorge Martins, Ricardo Haddad, Claudia Renata dos Santos Barros, Elaine Cristina Marqueze, Raul Machado Neto, Debora Botequio Moretti, Jardelina de Souza Todao Bernardino, Loyze Paola Oliveira de Lima, Luiz Aurelio de Campos Crispin. Centro de Genômica Funcional da ESALQ: Luiz Lehmann Coutinho, Ricardo Augusto Brassaloti, Raquel de Lello Rocha Campos Cassano. NGS Soluções Genômicas: Pilar Drummond Sampaio Corrêa Mariani. FZEA-USP Pirassununga: Mirele Daiana Poleti, Jessika Cristina Chagas Lesbon, Elisangela Chicaroni Mattos, Heidge Fukumasu. USP-Botucatu: Rejane Maria Tommasini Grotto, Jayme A. Souza-Neto, Guilherme Targino Valente, Patricia Akemi Assato, Felipe Allan da Silva da Costa, Bianca Cechetto Carlos. Mendelics: Bibiana Santos, João Paulo Kitajima, Erika Freitas, David Schlesinger. Hemocentro Ribeirão Preto: Simone Kashima, Evandra Strazza Rodrigues, Svetoslav Nanev Slavov, Elaine Vieira dos Santos, Rafael dos Santos Bezerra, Luiz Carlos Junior de Alcantara, Marta Giovanetti, Vagner Fonseca, Flavia Aburjaile, Rodrigo Tocantins Calado. FAMERP-SJRP: Cecília Artico Banho, Lívia Sacchetto, Fábio Sossai Possebon, Leila Sabrina Ullmann, Cíntia Bittar, Guilherme Campos, Helena Lage Ferreira, Jorge A. Petroli Marchesi, Maisa C. Pereira Parra, Marília Moraes, Paula Rahal, Paulo Inacio da Costa, João Pessoa Araújo Jr., Maurício Lacerda Nogueira. Prefeitura de Sao Paulo: Melissa Palmieri. |
| EPI_ISL_1966475 | PAS CIDADE JARDIM | Instituto Butantan / Mendelics | Instituto Butantan: Dimas Tadeu Covas, Sandra Coccuzzo Sampaio, Maria Carolina Elias, José Salvatore Leister Patané, Vincent Louis Viala, Antonio Jorge Martins, Ricardo Haddad, Claudia Renata dos Santos Barros, Elaine Cristina Marqueze, Raul Machado Neto, Debora Botequio Moretti, Jardelina de Souza Todao Bernardino, Loyze Paola Oliveira de Lima, Luiz Aurelio de Campos Crispin. Centro de Genômica Funcional da ESALQ: Luiz Lehmann Coutinho, Ricardo Augusto Brassaloti, Raquel de Lello Rocha Campos Cassano. NGS Soluções Genômicas: Pilar Drummond Sampaio Corrêa Mariani. FZEA-USP Pirassununga: Mirele Daiana Poleti, Jessika Cristina Chagas Lesbon, Elisangela Chicaroni Mattos, Heidge Fukumasu. USP-Botucatu: Rejane Maria Tommasini Grotto, Jayme A. Souza-Neto, Guilherme Targino Valente, Patricia Akemi Assato, Felipe Allan da Silva da Costa, Bianca Cechetto Carlos. Mendelics: Bibiana Santos, João Paulo Kitajima, Erika Freitas, David Schlesinger. Hemocentro Ribeirão Preto: Simone Kashima, Evandra Strazza Rodrigues, Svetoslav Nanev Slavov, Elaine Vieira dos Santos, Rafael dos Santos Bezerra, Luiz Carlos Junior de Alcantara, Marta Giovanetti, Vagner Fonseca, Flavia Aburjaile, Rodrigo Tocantins Calado. FAMERP-SJRP: Cecília Artico Banho, Lívia Sacchetto, Fábio Sossai Possebon, Leila Sabrina Ullmann, Cíntia Bittar, Guilherme Campos, Helena Lage Ferreira, Jorge A. Petroli Marchesi, Maisa C. Pereira Parra, Marília Moraes, Paula Rahal, Paulo Inacio da Costa, João Pessoa Araújo Jr., Maurício Lacerda Nogueira. Prefeitura de Sao Paulo: Melissa Palmieri. |
| EPI_ISL_1966476 | SECRETARIA DE SAUDE PUBLICA DE PRAIA GRANDE | Instituto Butantan / Mendelics | Instituto Butantan: Dimas Tadeu Covas, Sandra Coccuzzo Sampaio, Maria Carolina Elias, José Salvatore Leister Patané, Vincent Louis Viala, Antonio Jorge Martins, Ricardo Haddad, Claudia Renata dos Santos Barros, Elaine Cristina Marqueze, Raul Machado Neto, Debora Botequio Moretti, Jardelina de Souza Todao Bernardino, Loyze Paola Oliveira de Lima, Luiz Aurelio de Campos Crispin. Centro de Genômica Funcional da ESALQ: Luiz Lehmann Coutinho, Ricardo Augusto Brassaloti, Raquel de Lello Rocha Campos Cassano. NGS Soluções Genômicas: Pilar Drummond Sampaio Corrêa Mariani. FZEA-USP Pirassununga: Mirele Daiana Poleti, Jessika Cristina Chagas Lesbon, Elisangela Chicaroni Mattos, Heidge Fukumasu. USP-Botucatu: Rejane Maria Tommasini Grotto, Jayme A. Souza-Neto, Guilherme Targino Valente, Patricia Akemi Assato, Felipe Allan da Silva da Costa, Bianca Cechetto Carlos. Mendelics: Bibiana Santos, João Paulo Kitajima, Erika Freitas, David Schlesinger. Hemocentro Ribeirão Preto: Simone Kashima, Evandra Strazza Rodrigues, Svetoslav Nanev Slavov, Elaine Vieira dos Santos, Rafael dos Santos Bezerra, Luiz Carlos Junior de Alcantara, Marta Giovanetti, Vagner Fonseca, Flavia Aburjaile, Rodrigo Tocantins Calado. FAMERP-SJRP: Cecília Artico Banho, Lívia Sacchetto, Fábio Sossai Possebon, Leila Sabrina Ullmann, Cíntia Bittar, Guilherme Campos, Helena Lage Ferreira, Jorge A. Petroli Marchesi, Maisa C. Pereira Parra, Marília Moraes, Paula Rahal, Paulo Inacio da Costa, João Pessoa Araújo Jr., Maurício Lacerda Nogueira. Prefeitura de Sao Paulo: Melissa Palmieri. |
| EPI_ISL_1966477, EPI_ISL_1966478, EPI_ISL_1966479 | CENTRO DE SAUDE III RINCAO | Instituto Butantan / Mendelics | Instituto Butantan: Dimas Tadeu Covas, Sandra Coccuzzo Sampaio, Maria Carolina Elias, José Salvatore Leister Patané, Vincent Louis Viala, Antonio Jorge Martins, Ricardo Haddad, Claudia Renata dos Santos Barros, Elaine Cristina Marqueze, Raul Machado Neto, Debora Botequio Moretti, Jardelina de Souza Todao Bernardino, Loyze Paola Oliveira de Lima, Luiz Aurelio de Campos Crispin. Centro de Genômica Funcional da ESALQ: Luiz Lehmann Coutinho, Ricardo Augusto Brassaloti, Raquel de Lello Rocha Campos Cassano. NGS Soluções Genômicas: Pilar Drummond Sampaio Corrêa Mariani. FZEA-USP Pirassununga: Mirele Daiana Poleti, Jessika Cristina Chagas Lesbon, Elisangela Chicaroni Mattos, Heidge Fukumasu. USP-Botucatu: Rejane Maria Tommasini Grotto, Jayme A. Souza-Neto, Guilherme Targino Valente, Patricia Akemi Assato, Felipe Allan da Silva da Costa, Bianca Cechetto Carlos. Mendelics: Bibiana Santos, João Paulo Kitajima, Erika Freitas, David Schlesinger. Hemocentro Ribeirão Preto: Simone Kashima, Evandra Strazza Rodrigues, Svetoslav Nanev Slavov, Elaine Vieira dos Santos, Rafael dos Santos Bezerra, Luiz Carlos Junior de Alcantara, Marta Giovanetti, Vagner |

| | | | |
|---|---|---|---|
| | | | Fonseca, Flavia Aburjaile, Rodrigo Tocantins Calado. FAMERP-SJRP: Cecília Artico Banho, Lívia Sacchetto, Fábio Sossai Possebon, Leila Sabrina Ullmann, Cíntia Bittar, Guilherme Campos, Helena Lage Ferreira, Jorge A. Petroli Marchesi, Maisa C. Pereira Parra, Marília Moraes, Paula Rahal, Paulo Inacio da Costa, João Pessoa Araújo Jr., Maurício Lacerda Nogueira. Prefeitura de Sao Paulo: Melissa Palmieri. |
| EPI_ISL_1966480 | CS III SANTOPOLIS DO AGUAPEI | Instituto Butantan / Mendelics | Instituto Butantan: Dimas Tadeu Covas, Sandra Coccuzzo Sampaio, Maria Carolina Elias, José Salvatore Leister Patané, Vincent Louis Viala, Antonio Jorge Martins, Ricardo Haddad, Claudia Renata dos Santos Barros, Elaine Cristina Marqueze, Raul Machado Neto, Debora Botequio Moretti, Jardelina de Souza Todao Bernardino, Loyze Paola Oliveira de Lima, Luiz Aurelio de Campos Crispin. Centro de Genômica Funcional da ESALQ: Luiz Lehmann Coutinho, Ricardo Augusto Brassaloti, Raquel de Lello Rocha Campos Cassano. NGS Soluções Genômicas: Pilar Drummond Sampaio Corrêa Mariani. FZEA-USP Pirassununga: Mirele Daiana Poleti, Jessika Cristina Chagas Lesbon, Elisangela Chicaroni Mattos, Heidge Fukumasu. USP-Botucatu: Rejane Maria Tommasini Grotto, Jayme A. Souza-Neto, Guilherme Targino Valente, Patricia Akemi Assato, Felipe Allan da Silva da Costa, Bianca Cechetto Carlos. Mendelics: Bibiana Santos, João Paulo Kitajima, Erika Freitas, David Schlesinger. Hemocentro Ribeirão Preto: Simone Kashima, Evandra Strazza Rodrigues, Svetoslav Nanev Slavov, Elaine Vieira dos Santos, Rafael dos Santos Bezerra, Luiz Carlos Junior de Alcantara, Marta Giovanetti, Vagner Fonseca, Flavia Aburjaile, Rodrigo Tocantins Calado. FAMERP-SJRP: Cecília Artico Banho, Lívia Sacchetto, Fábio Sossai Possebon, Leila Sabrina Ullmann, Cíntia Bittar, Guilherme Campos, Helena Lage Ferreira, Jorge A. Petroli Marchesi, Maisa C. Pereira Parra, Marília Moraes, Paula Rahal, Paulo Inacio da Costa, João Pessoa Araújo Jr., Maurício Lacerda Nogueira. Prefeitura de Sao Paulo: Melissa Palmieri. |
| EPI_ISL_1966481, EPI_ISL_1966482, EPI_ISL_1966483, EPI_ISL_1966484, EPI_ISL_1966485, EPI_ISL_1966486, EPI_ISL_1966487, EPI_ISL_1966489, EPI_ISL_1966490, EPI_ISL_1966491, EPI_ISL_1966492, EPI_ISL_1966493, EPI_ISL_1966494, EPI_ISL_1966495, EPI_ISL_1966496 | | | |
| see above | SECAO CENTRO DE DIAGNOSTICO SECEDI | Instituto Butantan / Mendelics | Instituto Butantan: Dimas Tadeu Covas, Sandra Coccuzzo Sampaio, Maria Carolina Elias, José Salvatore Leister Patané, Vincent Louis Viala, Antonio Jorge Martins, Ricardo Haddad, Claudia Renata dos Santos Barros, Elaine Cristina Marqueze, Raul Machado Neto, Debora Botequio Moretti, Jardelina de Souza Todao Bernardino, Loyze Paola Oliveira de Lima, Luiz Aurelio de Campos Crispin. Centro de Genômica Funcional da ESALQ: Luiz Lehmann Coutinho, Ricardo Augusto Brassaloti, Raquel de Lello Rocha Campos Cassano. NGS Soluções Genômicas: Pilar Drummond Sampaio Corrêa Mariani. FZEA-USP Pirassununga: Mirele Daiana Poleti, Jessika Cristina Chagas Lesbon, Elisangela Chicaroni Mattos, Heidge Fukumasu. USP-Botucatu: Rejane Maria Tommasini Grotto, Jayme A. Souza-Neto, Guilherme Targino Valente, Patricia Akemi Assato, Felipe Allan da Silva da Costa, Bianca Cechetto Carlos. Mendelics: Bibiana Santos, João Paulo Kitajima, Erika Freitas, David Schlesinger. Hemocentro Ribeirão Preto: Simone Kashima, Evandra Strazza Rodrigues, Svetoslav Nanev Slavov, Elaine Vieira dos Santos, Rafael dos Santos Bezerra, Luiz Carlos Junior de Alcantara, Marta Giovanetti, Vagner Fonseca, Flavia Aburjaile, Rodrigo Tocantins Calado. FAMERP-SJRP: Cecília Artico Banho, Lívia Sacchetto, Fábio Sossai Possebon, Leila Sabrina Ullmann, Cíntia Bittar, Guilherme Campos, Helena Lage Ferreira, Jorge A. Petroli Marchesi, Maisa C. Pereira Parra, Marília Moraes, Paula Rahal, Paulo Inacio da Costa, João Pessoa Araújo Jr., Maurício Lacerda Nogueira. Prefeitura de Sao Paulo: Melissa Palmieri. |
| EPI_ISL_1966497, EPI_ISL_1966498, EPI_ISL_1966499, EPI_ISL_1966500, EPI_ISL_1966501, EPI_ISL_1966502, EPI_ISL_1966504, EPI_ISL_1966506 | Ambulatório IB | Instituto Butantan / Mendelics | Instituto Butantan: Dimas Tadeu Covas, Sandra Coccuzzo Sampaio, Maria Carolina Elias, José Salvatore Leister Patané, Vincent Louis Viala, Antonio Jorge Martins, Ricardo Haddad, Claudia Renata dos Santos Barros, Elaine Cristina Marqueze, Raul Machado Neto, Debora Botequio Moretti, Jardelina de Souza Todao Bernardino, Loyze Paola Oliveira de Lima, Luiz Aurelio de Campos Crispin. Centro de Genômica Funcional da ESALQ: Luiz Lehmann Coutinho, Ricardo Augusto Brassaloti, Raquel de Lello Rocha Campos Cassano. NGS Soluções Genômicas: Pilar Drummond Sampaio Corrêa Mariani. FZEA-USP Pirassununga: Mirele Daiana Poleti, Jessika Cristina Chagas Lesbon, Elisangela Chicaroni Mattos, Heidge Fukumasu. USP-Botucatu: Rejane Maria Tommasini Grotto, Jayme A. Souza-Neto, Guilherme Targino Valente, Patricia Akemi Assato, Felipe Allan da Silva da Costa, Bianca Cechetto Carlos. Mendelics: Bibiana Santos, João Paulo Kitajima, Erika Freitas, David Schlesinger. Hemocentro Ribeirão Preto: Simone Kashima, Evandra Strazza Rodrigues, Svetoslav Nanev Slavov, Elaine Vieira dos Santos, Rafael dos Santos Bezerra, Luiz Carlos Junior de Alcantara, Marta Giovanetti, Vagner Fonseca, Flavia Aburjaile, Rodrigo Tocantins Calado. FAMERP-SJRP: Cecília Artico Banho, Lívia Sacchetto, Fábio Sossai Possebon, Leila Sabrina Ullmann, Cíntia Bittar, Guilherme Campos, Helena Lage Ferreira, Jorge A. Petroli Marchesi, Maisa C. Pereira Parra, Marília Moraes, Paula Rahal, Paulo Inacio da Costa, João Pessoa Araújo Jr., Maurício Lacerda Nogueira. Prefeitura de Sao Paulo: Melissa Palmieri. |
| EPI_ISL_1966507 | CENTRO DE SAUDE SAO ROQUE DR JOSE CARVALHO BRITO | Instituto Butantan / Mendelics | Instituto Butantan: Dimas Tadeu Covas, Sandra Coccuzzo Sampaio, Maria Carolina Elias, José Salvatore Leister Patané, Vincent Louis Viala, Antonio Jorge Martins, Ricardo Haddad, Claudia Renata dos Santos Barros, Elaine Cristina Marqueze, Raul Machado Neto, Debora Botequio Moretti, Jardelina de Souza Todao Bernardino, Loyze Paola Oliveira de Lima, Luiz Aurelio de Campos Crispin. Centro de Genômica Funcional da ESALQ: Luiz Lehmann Coutinho, Ricardo Augusto Brassaloti, Raquel de Lello Rocha Campos Cassano. NGS Soluções Genômicas: Pilar Drummond Sampaio Corrêa Mariani. FZEA-USP Pirassununga: Mirele Daiana Poleti, Jessika Cristina Chagas Lesbon, Elisangela Chicaroni Mattos, Heidge Fukumasu. USP-Botucatu: Rejane Maria Tommasini Grotto, Jayme A. Souza-Neto, Guilherme Targino Valente, Patricia Akemi Assato, Felipe Allan da Silva da Costa, Bianca Cechetto Carlos. Mendelics: Bibiana Santos, João Paulo Kitajima, Erika Freitas, David Schlesinger. Hemocentro Ribeirão Preto: Simone Kashima, Evandra Strazza Rodrigues, Svetoslav Nanev Slavov, Elaine Vieira dos Santos, Rafael dos Santos Bezerra, Luiz Carlos Junior de Alcantara, Marta Giovanetti, Vagner Fonseca, Flavia Aburjaile, Rodrigo Tocantins Calado. FAMERP-SJRP: Cecília Artico Banho, Lívia Sacchetto, Fábio Sossai Possebon, Leila Sabrina Ullmann, Cíntia Bittar, Guilherme Campos, Helena Lage Ferreira, Jorge A. Petroli Marchesi, Maisa C. Pereira Parra, Marília Moraes, Paula Rahal, Paulo Inacio da Costa, João Pessoa Araújo Jr., Maurício Lacerda Nogueira. Prefeitura de Sao Paulo: Melissa Palmieri. |
| EPI_ISL_1966508, EPI_ISL_1966509 | PRONTO SOCORRO MUNICIPAL DE SEVERINIA | Instituto Butantan / Mendelics | Instituto Butantan: Dimas Tadeu Covas, Sandra Coccuzzo Sampaio, Maria Carolina Elias, José Salvatore Leister Patané, Vincent Louis Viala, Antonio Jorge Martins, Ricardo Haddad, Claudia Renata dos Santos Barros, Elaine Cristina Marqueze, Raul Machado Neto, Debora Botequio Moretti, Jardelina de Souza Todao Bernardino, Loyze Paola Oliveira de Lima, Luiz Aurelio de Campos Crispin. Centro de Genômica Funcional da ESALQ: Luiz Lehmann Coutinho, Ricardo Augusto Brassaloti, Raquel de Lello Rocha Campos Cassano. NGS Soluções Genômicas: Pilar Drummond Sampaio Corrêa Mariani. FZEA-USP Pirassununga: Mirele Daiana Poleti, Jessika Cristina Chagas Lesbon, Elisangela Chicaroni Mattos, Heidge Fukumasu. USP-Botucatu: Rejane Maria Tommasini Grotto, Jayme A. Souza-Neto, Guilherme Targino Valente, Patricia Akemi Assato, Felipe Allan da Silva da Costa, Bianca Cechetto Carlos. Mendelics: Bibiana Santos, João Paulo Kitajima, Erika Freitas, David Schlesinger. Hemocentro Ribeirão Preto: Simone Kashima, Evandra Strazza Rodrigues, Svetoslav Nanev Slavov, Elaine Vieira dos Santos, Rafael dos Santos Bezerra, Luiz Carlos Junior de Alcantara, Marta Giovanetti, Vagner Fonseca, Flavia Aburjaile, Rodrigo Tocantins Calado. FAMERP-SJRP: Cecília Artico Banho, Lívia Sacchetto, Fábio Sossai Possebon, Leila Sabrina Ullmann, Cíntia Bittar, Guilherme Campos, Helena Lage Ferreira, Jorge A. Petroli Marchesi, Maisa C. Pereira Parra, Marília Moraes, Paula Rahal, Paulo Inacio da Costa, João Pessoa Araújo Jr., Maurício Lacerda Nogueira. Prefeitura de Sao Paulo: Melissa Palmieri. |
| EPI_ISL_1966510 | UBS DE SUD MENNUCCI | Instituto Butantan / Mendelics | Instituto Butantan: Dimas Tadeu Covas, Sandra Coccuzzo Sampaio, Maria Carolina Elias, José Salvatore Leister Patané, Vincent Louis Viala, Antonio Jorge Martins, Ricardo Haddad, Claudia Renata dos Santos Barros, Elaine Cristina Marqueze, Raul Machado Neto, Debora Botequio Moretti, Jardelina de Souza Todao Bernardino, Loyze Paola Oliveira de Lima, Luiz Aurelio de Campos Crispin. Centro de Genômica Funcional da ESALQ: Luiz Lehmann Coutinho, Ricardo Augusto Brassaloti, Raquel de Lello Rocha Campos Cassano. NGS Soluções Genômicas: Pilar Drummond Sampaio Corrêa Mariani. FZEA-USP Pirassununga: Mirele Daiana Poleti, Jessika Cristina Chagas Lesbon, Elisangela Chicaroni Mattos, Heidge Fukumasu. USP-Botucatu: Rejane Maria Tommasini Grotto, Jayme A. Souza-Neto, Guilherme Targino Valente, Patricia Akemi Assato, Felipe Allan da Silva da Costa, Bianca Cechetto Carlos. Mendelics: Bibiana Santos, João Paulo Kitajima, Erika Freitas, David Schlesinger. Hemocentro Ribeirão Preto: Simone Kashima, Evandra Strazza Rodrigues, Svetoslav Nanev Slavov, Elaine Vieira dos Santos, Rafael dos Santos Bezerra, Luiz Carlos Junior de Alcantara, Marta Giovanetti, Vagner Fonseca, Flavia Aburjaile, Rodrigo Tocantins Calado. FAMERP-SJRP: Cecília Artico Banho, Lívia Sacchetto, Fábio Sossai Possebon, Leila Sabrina Ullmann, Cíntia Bittar, Guilherme Campos, Helena Lage Ferreira, Jorge A. Petroli Marchesi, Maisa C. Pereira Parra, Marília Moraes, Paula Rahal, Paulo Inacio da Costa, João Pessoa Araújo Jr., Maurício Lacerda Nogueira. Prefeitura de Sao Paulo: Melissa Palmieri. |
| EPI_ISL_1966511 | UBS 2 DE SUZANAPOLIS | Instituto Butantan / Mendelics | Instituto Butantan: Dimas Tadeu Covas, Sandra Coccuzzo Sampaio, Maria Carolina Elias, José Salvatore Leister Patané, Vincent Louis Viala, Antonio Jorge Martins, Ricardo Haddad, Claudia Renata dos Santos Barros, Elaine Cristina Marqueze, Raul Machado Neto, Debora Botequio Moretti, Jardelina de Souza Todao Bernardino, Loyze Paola Oliveira de Lima, Luiz Aurelio de Campos Crispin. Centro de Genômica Funcional da ESALQ: Luiz Lehmann Coutinho, Ricardo Augusto Brassaloti, Raquel de Lello Rocha Campos Cassano. NGS Soluções Genômicas: Pilar Drummond Sampaio Corrêa Mariani. FZEA-USP Pirassununga: Mirele Daiana Poleti, Jessika Cristina Chagas Lesbon, Elisangela Chicaroni Mattos, Heidge Fukumasu. USP-Botucatu: Rejane Maria Tommasini Grotto, Jayme A. Souza-Neto, Guilherme Targino Valente, Patricia Akemi Assato, Felipe Allan da Silva da Costa, Bianca Cechetto Carlos. Mendelics: Bibiana Santos, João Paulo Kitajima, Erika Freitas, David Schlesinger. Hemocentro Ribeirão Preto: Simone Kashima, Evandra Strazza Rodrigues, Svetoslav Nanev Slavov, Elaine Vieira dos Santos, Rafael dos Santos Bezerra, Luiz Carlos Junior de Alcantara, Marta Giovanetti, Vagner Fonseca, Flavia Aburjaile, Rodrigo Tocantins Calado. FAMERP-SJRP: Cecília Artico Banho, Lívia Sacchetto, Fábio Sossai Possebon, Leila Sabrina Ullmann, Cíntia Bittar, Guilherme Campos, Helena Lage Ferreira, Jorge A. Petroli Marchesi, Maisa C. Pereira Parra, Marília Moraes, Paula Rahal, Paulo Inacio da Costa, João Pessoa Araújo Jr., Maurício Lacerda Nogueira. Prefeitura de Sao Paulo: Melissa Palmieri. |
| EPI_ISL_1966512 | UNIDADE MISTA DE TAIACU | Instituto Butantan / Mendelics | Dimas Tadeu Covas, Antonio Jorge Martins, Claudia Renata dos Santos Barros, David Schlesinger, Debora Botequio Moretti, Elaine Cristina Marqueze, Elaine Vieira Santos, Evandra Strazza Rodrigues, Heidge Fukumasu, Jayme Augusto de Souza-Neto, José Salvatore Leister Patané, Luiz Alcantara, Luiz |

| | | | |
|---|---|---|---|
| | | | Lehmann Coutinho, Maria Carolina Elias, Maurício Lacerda Nogueira, Rafael dos Santos Bezerra, Raul Machado Neto, Rejane Maria Tommasini Grotto, Ricardo Haddad, Sandra Coccuzzo Sampaio Vessoni, Simone Kashima, Svetoslav Nanev Slavov, Vincent Louis Viala |
| EPI_ISL_1966513, EPI_ISL_1966514, EPI_ISL_1966515, EPI_ISL_1966516 | UNIDADE BASICA DE SAUDE DE TAQUARAL CAETANO PITELLI | Instituto Butantan / Mendelics | Instituto Butantan: Dimas Tadeu Covas, Sandra Coccuzzo Sampaio, Maria Carolina Elias, José Salvatore Leister Patané, Vincent Louis Viala, Antonio Jorge Martins, Ricardo Haddad, Claudia Renata dos Santos Barros, Elaine Cristina Marqueze, Raul Machado Neto, Debora Botequio Moretti, Jardelina de Souza Todao Bernardino, Loyze Paola Oliveira de Lima, Luiz Aurelio de Campos Crispin. Centro de Genômica Funcional da ESALQ: Luiz Lehmann Coutinho, Ricardo Augusto Brassaloti, Raquel de Lello Rocha Campos Cassano. NGS Soluções Genômicas: Pilar Drummond Sampaio Corrêa Mariani. FZEA-USP Pirassununga: Mirele Daiana Poleti, Jessika Cristina Chagas Lesbon, Elisangela Chicaroni Mattos, Heidge Fukumasu. USP-Botucatu: Rejane Maria Tommasini Grotto, Jayme A. Souza-Neto, Guilherme Targino Valente, Patricia Akemi Assato, Felipe Allan da Silva da Costa, Bianca Cechetto Carlos. Mendelics: Bibiana Santos, João Paulo Kitajima, Erika Freitas, David Schlesinger. Hemocentro Ribeirão Preto: Simone Kashima, Evandra Strazza Rodrigues, Svetoslav Nanev Slavov, Elaine Vieira dos Santos, Rafael dos Santos Bezerra, Luiz Carlos Junior de Alcantara, Marta Giovanetti, Vagner Fonseca, Flavia Aburjaile, Rodrigo Tocantins Calado. FAMERP-SJRP: Cecília Artico Banho, Lívia Sacchetto, Fábio Sossai Possebon, Leila Sabrina Ullmann, Cíntia Bittar, Guilherme Campos, Helena Lage Ferreira, Jorge A. Petroli Marchesi, Maisa C. Pereira Parra, Marília Moraes, Paula Rahal, Paulo Inacio da Costa, João Pessoa Araújo Jr., Maurício Lacerda Nogueira. Prefeitura de Sao Paulo: Melissa Palmieri. |
| EPI_ISL_1966518, EPI_ISL_1966519, EPI_ISL_1966520 | SAE SERVICO DE ATENDIMENTO ESPECIALIZADO | Instituto Butantan / Mendelics | Instituto Butantan: Dimas Tadeu Covas, Sandra Coccuzzo Sampaio, Maria Carolina Elias, José Salvatore Leister Patané, Vincent Louis Viala, Antonio Jorge Martins, Ricardo Haddad, Claudia Renata dos Santos Barros, Elaine Cristina Marqueze, Raul Machado Neto, Debora Botequio Moretti, Jardelina de Souza Todao Bernardino, Loyze Paola Oliveira de Lima, Luiz Aurelio de Campos Crispin. Centro de Genômica Funcional da ESALQ: Luiz Lehmann Coutinho, Ricardo Augusto Brassaloti, Raquel de Lello Rocha Campos Cassano. NGS Soluções Genômicas: Pilar Drummond Sampaio Corrêa Mariani. FZEA-USP Pirassununga: Mirele Daiana Poleti, Jessika Cristina Chagas Lesbon, Elisangela Chicaroni Mattos, Heidge Fukumasu. USP-Botucatu: Rejane Maria Tommasini Grotto, Jayme A. Souza-Neto, Guilherme Targino Valente, Patricia Akemi Assato, Felipe Allan da Silva da Costa, Bianca Cechetto Carlos. Mendelics: Bibiana Santos, João Paulo Kitajima, Erika Freitas, David Schlesinger. Hemocentro Ribeirão Preto: Simone Kashima, Evandra Strazza Rodrigues, Svetoslav Nanev Slavov, Elaine Vieira dos Santos, Rafael dos Santos Bezerra, Luiz Carlos Junior de Alcantara, Marta Giovanetti, Vagner Fonseca, Flavia Aburjaile, Rodrigo Tocantins Calado. FAMERP-SJRP: Cecília Artico Banho, Lívia Sacchetto, Fábio Sossai Possebon, Leila Sabrina Ullmann, Cíntia Bittar, Guilherme Campos, Helena Lage Ferreira, Jorge A. Petroli Marchesi, Maisa C. Pereira Parra, Marília Moraes, Paula Rahal, Paulo Inacio da Costa, João Pessoa Araújo Jr., Maurício Lacerda Nogueira. Prefeitura de Sao Paulo: Melissa Palmieri. |
| EPI_ISL_1966521 | UNIDADE MISTA DE VISTA ALEGRE DO ALTO VISTA ALEGRE DO ALTO | Instituto Butantan / Mendelics | Instituto Butantan: Dimas Tadeu Covas, Sandra Coccuzzo Sampaio, Maria Carolina Elias, José Salvatore Leister Patané, Vincent Louis Viala, Antonio Jorge Martins, Ricardo Haddad, Claudia Renata dos Santos Barros, Elaine Cristina Marqueze, Raul Machado Neto, Debora Botequio Moretti, Jardelina de Souza Todao Bernardino, Loyze Paola Oliveira de Lima, Luiz Aurelio de Campos Crispin. Centro de Genômica Funcional da ESALQ: Luiz Lehmann Coutinho, Ricardo Augusto Brassaloti, Raquel de Lello Rocha Campos Cassano. NGS Soluções Genômicas: Pilar Drummond Sampaio Corrêa Mariani. FZEA-USP Pirassununga: Mirele Daiana Poleti, Jessika Cristina Chagas Lesbon, Elisangela Chicaroni Mattos, Heidge Fukumasu. USP-Botucatu: Rejane Maria Tommasini Grotto, Jayme A. Souza-Neto, Guilherme Targino Valente, Patricia Akemi Assato, Felipe Allan da Silva da Costa, Bianca Cechetto Carlos. Mendelics: Bibiana Santos, João Paulo Kitajima, Erika Freitas, David Schlesinger. Hemocentro Ribeirão Preto: Simone Kashima, Evandra Strazza Rodrigues, Svetoslav Nanev Slavov, Elaine Vieira dos Santos, Rafael dos Santos Bezerra, Luiz Carlos Junior de Alcantara, Marta Giovanetti, Vagner Fonseca, Flavia Aburjaile, Rodrigo Tocantins Calado. FAMERP-SJRP: Cecília Artico Banho, Lívia Sacchetto, Fábio Sossai Possebon, Leila Sabrina Ullmann, Cíntia Bittar, Guilherme Campos, Helena Lage Ferreira, Jorge A. Petroli Marchesi, Maisa C. Pereira Parra, Marília Moraes, Paula Rahal, Paulo Inacio da Costa, João Pessoa Araújo Jr., Maurício Lacerda Nogueira. Prefeitura de Sao Paulo: Melissa Palmieri. |
| EPI_ISL_1966522 | CENTRO DE SAUDE DE ADAMANTINA | Instituto Butantan / FZEA-USP (Pirassununga) | Instituto Butantan: Dimas Tadeu Covas, Sandra Coccuzzo Sampaio, Maria Carolina Elias, José Salvatore Leister Patané, Vincent Louis Viala, Antonio Jorge Martins, Ricardo Haddad, Claudia Renata dos Santos Barros, Elaine Cristina Marqueze, Raul Machado Neto, Debora Botequio Moretti, Jardelina de Souza Todao Bernardino, Loyze Paola Oliveira de Lima, Luiz Aurelio de Campos Crispin. Centro de Genômica Funcional da ESALQ: Luiz Lehmann Coutinho, Ricardo Augusto Brassaloti, Raquel de Lello Rocha Campos Cassano. NGS Soluções Genômicas: Pilar Drummond Sampaio Corrêa Mariani. FZEA-USP Pirassununga: Mirele Daiana Poleti, Jessika Cristina Chagas Lesbon, Elisangela Chicaroni Mattos, Heidge Fukumasu. USP-Botucatu: Rejane Maria Tommasini Grotto, Jayme A. Souza-Neto, Guilherme Targino Valente, Patricia Akemi Assato, Felipe Allan da Silva da Costa, Bianca Cechetto Carlos. Mendelics: Bibiana Santos, João Paulo Kitajima, Erika Freitas, David Schlesinger. Hemocentro Ribeirão Preto: Simone Kashima, Evandra Strazza Rodrigues, Svetoslav Nanev Slavov, Elaine Vieira dos Santos, Rafael dos Santos Bezerra, Luiz Carlos Junior de Alcantara, Marta Giovanetti, Vagner Fonseca, Flavia Aburjaile, Rodrigo Tocantins Calado. FAMERP-SJRP: Cecília Artico Banho, Lívia Sacchetto, Fábio Sossai Possebon, Leila Sabrina Ullmann, Cíntia Bittar, Guilherme Campos, Helena Lage Ferreira, Jorge A. Petroli Marchesi, Maisa C. Pereira Parra, Marília Moraes, Paula Rahal, Paulo Inacio da Costa, João Pessoa Araújo Jr., Maurício Lacerda Nogueira. Prefeitura de Sao Paulo: Melissa Palmieri. |
| EPI_ISL_1966523, EPI_ISL_1966524 | CS DE BALSAMO | Instituto Butantan / FZEA-USP (Pirassununga) | Instituto Butantan: Dimas Tadeu Covas, Sandra Coccuzzo Sampaio, Maria Carolina Elias, José Salvatore Leister Patané, Vincent Louis Viala, Antonio Jorge Martins, Ricardo Haddad, Claudia Renata dos Santos Barros, Elaine Cristina Marqueze, Raul Machado Neto, Debora Botequio Moretti, Jardelina de Souza Todao Bernardino, Loyze Paola Oliveira de Lima, Luiz Aurelio de Campos Crispin. Centro de Genômica Funcional da ESALQ: Luiz Lehmann Coutinho, Ricardo Augusto Brassaloti, Raquel de Lello Rocha Campos Cassano. NGS Soluções Genômicas: Pilar Drummond Sampaio Corrêa Mariani. FZEA-USP Pirassununga: Mirele Daiana Poleti, Jessika Cristina Chagas Lesbon, Elisangela Chicaroni Mattos, Heidge Fukumasu. USP-Botucatu: Rejane Maria Tommasini Grotto, Jayme A. Souza-Neto, Guilherme Targino Valente, Patricia Akemi Assato, Felipe Allan da Silva da Costa, Bianca Cechetto Carlos. Mendelics: Bibiana Santos, João Paulo Kitajima, Erika Freitas, David Schlesinger. Hemocentro Ribeirão Preto: Simone Kashima, Evandra Strazza Rodrigues, Svetoslav Nanev Slavov, Elaine Vieira dos Santos, Rafael dos Santos Bezerra, Luiz Carlos Junior de Alcantara, Marta Giovanetti, Vagner Fonseca, Flavia Aburjaile, Rodrigo Tocantins Calado. FAMERP-SJRP: Cecília Artico Banho, Lívia Sacchetto, Fábio Sossai Possebon, Leila Sabrina Ullmann, Cíntia Bittar, Guilherme Campos, Helena Lage Ferreira, Jorge A. Petroli Marchesi, Maisa C. Pereira Parra, Marília Moraes, Paula Rahal, Paulo Inacio da Costa, João Pessoa Araújo Jr., Maurício Lacerda Nogueira. Prefeitura de Sao Paulo: Melissa Palmieri. |
| EPI_ISL_1966525 | VIGILANCIA EM SAUDE | Instituto Butantan / FZEA-USP (Pirassununga) | Instituto Butantan: Dimas Tadeu Covas, Sandra Coccuzzo Sampaio, Maria Carolina Elias, José Salvatore Leister Patané, Vincent Louis Viala, Antonio Jorge Martins, Ricardo Haddad, Claudia Renata dos Santos Barros, Elaine Cristina Marqueze, Raul Machado Neto, Debora Botequio Moretti, Jardelina de Souza Todao Bernardino, Loyze Paola Oliveira de Lima, Luiz Aurelio de Campos Crispin. Centro de Genômica Funcional da ESALQ: Luiz Lehmann Coutinho, Ricardo Augusto Brassaloti, Raquel de Lello Rocha Campos Cassano. NGS Soluções Genômicas: Pilar Drummond Sampaio Corrêa Mariani. FZEA-USP Pirassununga: Mirele Daiana Poleti, Jessika Cristina Chagas Lesbon, Elisangela Chicaroni Mattos, Heidge Fukumasu. USP-Botucatu: Rejane Maria Tommasini Grotto, Jayme A. Souza-Neto, Guilherme Targino Valente, Patricia Akemi Assato, Felipe Allan da Silva da Costa, Bianca Cechetto Carlos. Mendelics: Bibiana Santos, João Paulo Kitajima, Erika Freitas, David Schlesinger. Hemocentro Ribeirão Preto: Simone Kashima, Evandra Strazza Rodrigues, Svetoslav Nanev Slavov, Elaine Vieira dos Santos, Rafael dos Santos Bezerra, Luiz Carlos Junior de Alcantara, Marta Giovanetti, Vagner Fonseca, Flavia Aburjaile, Rodrigo Tocantins Calado. FAMERP-SJRP: Cecília Artico Banho, Lívia Sacchetto, Fábio Sossai Possebon, Leila Sabrina Ullmann, Cíntia Bittar, Guilherme Campos, Helena Lage Ferreira, Jorge A. Petroli Marchesi, Maisa C. Pereira Parra, Marília Moraes, Paula Rahal, Paulo Inacio da Costa, João Pessoa Araújo Jr., Maurício Lacerda Nogueira. Prefeitura de Sao Paulo: Melissa Palmieri. |
| EPI_ISL_1966526, EPI_ISL_1966527, EPI_ISL_1966528, EPI_ISL_1966529 | USF PAULISTA FERNANDOPOLIS ANTONIO PIVATO | Instituto Butantan / FZEA-USP (Pirassununga) | Instituto Butantan: Dimas Tadeu Covas, Sandra Coccuzzo Sampaio, Maria Carolina Elias, José Salvatore Leister Patané, Vincent Louis Viala, Antonio Jorge Martins, Ricardo Haddad, Claudia Renata dos Santos Barros, Elaine Cristina Marqueze, Raul Machado Neto, Debora Botequio Moretti, Jardelina de Souza Todao Bernardino, Loyze Paola Oliveira de Lima, Luiz Aurelio de Campos Crispin. Centro de Genômica Funcional da ESALQ: Luiz Lehmann Coutinho, Ricardo Augusto Brassaloti, Raquel de Lello Rocha Campos Cassano. NGS Soluções Genômicas: Pilar Drummond Sampaio Corrêa Mariani. FZEA-USP Pirassununga: Mirele Daiana Poleti, Jessika Cristina Chagas Lesbon, Elisangela Chicaroni Mattos, Heidge Fukumasu. USP-Botucatu: Rejane Maria Tommasini Grotto, Jayme A. Souza-Neto, Guilherme Targino Valente, Patricia Akemi Assato, Felipe Allan da Silva da Costa, Bianca Cechetto Carlos. Mendelics: Bibiana Santos, João Paulo Kitajima, Erika Freitas, David Schlesinger. Hemocentro Ribeirão Preto: Simone Kashima, Evandra Strazza Rodrigues, Svetoslav Nanev Slavov, Elaine Vieira dos Santos, Rafael dos Santos Bezerra, Luiz Carlos Junior de Alcantara, Marta Giovanetti, Vagner Fonseca, Flavia Aburjaile, Rodrigo Tocantins Calado. FAMERP-SJRP: Cecília Artico Banho, Lívia Sacchetto, Fábio Sossai Possebon, Leila Sabrina Ullmann, Cíntia Bittar, Guilherme Campos, Helena Lage Ferreira, Jorge A. Petroli Marchesi, Maisa C. Pereira Parra, Marília Moraes, Paula Rahal, Paulo Inacio da Costa, João Pessoa Araújo Jr., Maurício Lacerda Nogueira. Prefeitura de Sao Paulo: Melissa Palmieri. |
| EPI_ISL_1966530 | HOSPITAL MUNICIPAL DR TABAJARA RAMOS | Instituto Butantan / FZEA-USP (Pirassununga) | Instituto Butantan: Dimas Tadeu Covas, Sandra Coccuzzo Sampaio, Maria Carolina Elias, José Salvatore Leister Patané, Vincent Louis Viala, Antonio Jorge Martins, Ricardo Haddad, Claudia Renata dos Santos Barros, Elaine Cristina Marqueze, Raul Machado Neto, Debora Botequio Moretti, Jardelina de Souza Todao Bernardino, Loyze Paola Oliveira de Lima, Luiz Aurelio de Campos Crispin. Centro de Genômica Funcional da ESALQ: Luiz Lehmann Coutinho, Ricardo Augusto Brassaloti, Raquel de Lello Rocha Campos Cassano. NGS Soluções Genômicas: Pilar Drummond Sampaio Corrêa Mariani. |

| | | | |
|---|---|---|---|
| | | | FZEA-USP Pirassununga: Mirele Daiana Poleti, Jessika Cristina Chagas Lesbon, Elisangela Chicaroni Mattos, Heidge Fukumasu. USP-Botucatu: Rejane Maria Tommasini Grotto, Jayme A. Souza-Neto, Guilherme Targino Valente, Patricia Akemi Assato, Felipe Allan da Silva da Costa, Bianca Cechetto Carlos. Mendelics: Bibiana Santos, João Paulo Kitajima, Erika Freitas, David Schlesinger. Hemocentro Ribeirão Preto: Simone Kashima, Evandra Strazza Rodrigues, Svetoslav Nanev Slavov, Elaine Vieira dos Santos, Rafael dos Santos Bezerra, Luiz Carlos Junior de Alcantara, Marta Giovanetti, Vagner Fonseca, Flavia Aburjaile, Rodrigo Tocantins Calado. FAMERP-SJRP: Cecília Artico Banho, Lívia Sacchetto, Fábio Sossai Possebon, Leila Sabrina Ullmann, Cíntia Bittar, Guilherme Campos, Helena Lage Ferreira, Jorge A. Petroli Marchesi, Maisa C. Pereira Parra, Marília Moraes, Paula Rahal, Paulo Inacio da Costa, João Pessoa Araújo Jr., Maurício Lacerda Nogueira. Prefeitura de Sao Paulo: Melissa Palmieri. |
| EPI_ISL_1966532, EPI_ISL_1966533 | EMERGENCIA RESPIRATORIA DE NOVA GRANADA | Instituto Butantan / FZEA-USP (Pirassununga) | Instituto Butantan: Dimas Tadeu Covas, Sandra Coccuzzo Sampaio, Maria Carolina Elias, José Salvatore Leister Patané, Vincent Louis Viala, Antonio Jorge Martins, Ricardo Haddad, Claudia Renata dos Santos Barros, Elaine Cristina Marqueze, Raul Machado Neto, Debora Botequio Moretti, Jardelina de Souza Todao Bernardino, Loyze Paola Oliveira de Lima, Luiz Aurelio de Campos Crispin. Centro de Genômica Funcional da ESALQ: Luiz Lehmann Coutinho, Ricardo Augusto Brassaloti, Raquel de Lello Rocha Campos Cassano. NGS Soluções Genômicas: Pilar Drummond Sampaio Corrêa Mariani. FZEA-USP Pirassununga: Mirele Daiana Poleti, Jessika Cristina Chagas Lesbon, Elisangela Chicaroni Mattos, Heidge Fukumasu. USP-Botucatu: Rejane Maria Tommasini Grotto, Jayme A. Souza-Neto, Guilherme Targino Valente, Patricia Akemi Assato, Felipe Allan da Silva da Costa, Bianca Cechetto Carlos. Mendelics: Bibiana Santos, João Paulo Kitajima, Erika Freitas, David Schlesinger. Hemocentro Ribeirão Preto: Simone Kashima, Evandra Strazza Rodrigues, Svetoslav Nanev Slavov, Elaine Vieira dos Santos, Rafael dos Santos Bezerra, Luiz Carlos Junior de Alcantara, Marta Giovanetti, Vagner Fonseca, Flavia Aburjaile, Rodrigo Tocantins Calado. FAMERP-SJRP: Cecília Artico Banho, Lívia Sacchetto, Fábio Sossai Possebon, Leila Sabrina Ullmann, Cíntia Bittar, Guilherme Campos, Helena Lage Ferreira, Jorge A. Petroli Marchesi, Maisa C. Pereira Parra, Marília Moraes, Paula Rahal, Paulo Inacio da Costa, João Pessoa Araújo Jr., Maurício Lacerda Nogueira. Prefeitura de Sao Paulo: Melissa Palmieri. |
| EPI_ISL_1966534 | UBS DR HELIO MIGLIARI | Instituto Butantan / FZEA-USP (Pirassununga) | Instituto Butantan: Dimas Tadeu Covas, Sandra Coccuzzo Sampaio, Maria Carolina Elias, José Salvatore Leister Patané, Vincent Louis Viala, Antonio Jorge Martins, Ricardo Haddad, Claudia Renata dos Santos Barros, Elaine Cristina Marqueze, Raul Machado Neto, Debora Botequio Moretti, Jardelina de Souza Todao Bernardino, Loyze Paola Oliveira de Lima, Luiz Aurelio de Campos Crispin. Centro de Genômica Funcional da ESALQ: Luiz Lehmann Coutinho, Ricardo Augusto Brassaloti, Raquel de Lello Rocha Campos Cassano. NGS Soluções Genômicas: Pilar Drummond Sampaio Corrêa Mariani. FZEA-USP Pirassununga: Mirele Daiana Poleti, Jessika Cristina Chagas Lesbon, Elisangela Chicaroni Mattos, Heidge Fukumasu. USP-Botucatu: Rejane Maria Tommasini Grotto, Jayme A. Souza-Neto, Guilherme Targino Valente, Patricia Akemi Assato, Felipe Allan da Silva da Costa, Bianca Cechetto Carlos. Mendelics: Bibiana Santos, João Paulo Kitajima, Erika Freitas, David Schlesinger. Hemocentro Ribeirão Preto: Simone Kashima, Evandra Strazza Rodrigues, Svetoslav Nanev Slavov, Elaine Vieira dos Santos, Rafael dos Santos Bezerra, Luiz Carlos Junior de Alcantara, Marta Giovanetti, Vagner Fonseca, Flavia Aburjaile, Rodrigo Tocantins Calado. FAMERP-SJRP: Cecília Artico Banho, Lívia Sacchetto, Fábio Sossai Possebon, Leila Sabrina Ullmann, Cíntia Bittar, Guilherme Campos, Helena Lage Ferreira, Jorge A. Petroli Marchesi, Maisa C. Pereira Parra, Marília Moraes, Paula Rahal, Paulo Inacio da Costa, João Pessoa Araújo Jr., Maurício Lacerda Nogueira. Prefeitura de Sao Paulo: Melissa Palmieri. |
| EPI_ISL_1966535 | UBS REGIAO OESTE DE OURINHOS | Instituto Butantan / FZEA-USP (Pirassununga) | Instituto Butantan: Dimas Tadeu Covas, Sandra Coccuzzo Sampaio, Maria Carolina Elias, José Salvatore Leister Patané, Vincent Louis Viala, Antonio Jorge Martins, Ricardo Haddad, Claudia Renata dos Santos Barros, Elaine Cristina Marqueze, Raul Machado Neto, Debora Botequio Moretti, Jardelina de Souza Todao Bernardino, Loyze Paola Oliveira de Lima, Luiz Aurelio de Campos Crispin. Centro de Genômica Funcional da ESALQ: Luiz Lehmann Coutinho, Ricardo Augusto Brassaloti, Raquel de Lello Rocha Campos Cassano. NGS Soluções Genômicas: Pilar Drummond Sampaio Corrêa Mariani. FZEA-USP Pirassununga: Mirele Daiana Poleti, Jessika Cristina Chagas Lesbon, Elisangela Chicaroni Mattos, Heidge Fukumasu. USP-Botucatu: Rejane Maria Tommasini Grotto, Jayme A. Souza-Neto, Guilherme Targino Valente, Patricia Akemi Assato, Felipe Allan da Silva da Costa, Bianca Cechetto Carlos. Mendelics: Bibiana Santos, João Paulo Kitajima, Erika Freitas, David Schlesinger. Hemocentro Ribeirão Preto: Simone Kashima, Evandra Strazza Rodrigues, Svetoslav Nanev Slavov, Elaine Vieira dos Santos, Rafael dos Santos Bezerra, Luiz Carlos Junior de Alcantara, Marta Giovanetti, Vagner Fonseca, Flavia Aburjaile, Rodrigo Tocantins Calado. FAMERP-SJRP: Cecília Artico Banho, Lívia Sacchetto, Fábio Sossai Possebon, Leila Sabrina Ullmann, Cíntia Bittar, Guilherme Campos, Helena Lage Ferreira, Jorge A. Petroli Marchesi, Maisa C. Pereira Parra, Marília Moraes, Paula Rahal, Paulo Inacio da Costa, João Pessoa Araújo Jr., Maurício Lacerda Nogueira. Prefeitura de Sao Paulo: Melissa Palmieri. |
| EPI_ISL_1966536, EPI_ISL_1966537, EPI_ISL_1966538, EPI_ISL_1966539, EPI_ISL_1966540 | CENTRO DE SAUDE I TACITO LEITE DE CARVALHO E SILVA | Instituto Butantan / FZEA-USP (Pirassununga) | Instituto Butantan: Dimas Tadeu Covas, Sandra Coccuzzo Sampaio, Maria Carolina Elias, José Salvatore Leister Patané, Vincent Louis Viala, Antonio Jorge Martins, Ricardo Haddad, Claudia Renata dos Santos Barros, Elaine Cristina Marqueze, Raul Machado Neto, Debora Botequio Moretti, Jardelina de Souza Todao Bernardino, Loyze Paola Oliveira de Lima, Luiz Aurelio de Campos Crispin. Centro de Genômica Funcional da ESALQ: Luiz Lehmann Coutinho, Ricardo Augusto Brassaloti, Raquel de Lello Rocha Campos Cassano. NGS Soluções Genômicas: Pilar Drummond Sampaio Corrêa Mariani. FZEA-USP Pirassununga: Mirele Daiana Poleti, Jessika Cristina Chagas Lesbon, Elisangela Chicaroni Mattos, Heidge Fukumasu. USP-Botucatu: Rejane Maria Tommasini Grotto, Jayme A. Souza-Neto, Guilherme Targino Valente, Patricia Akemi Assato, Felipe Allan da Silva da Costa, Bianca Cechetto Carlos. Mendelics: Bibiana Santos, João Paulo Kitajima, Erika Freitas, David Schlesinger. Hemocentro Ribeirão Preto: Simone Kashima, Evandra Strazza Rodrigues, Svetoslav Nanev Slavov, Elaine Vieira dos Santos, Rafael dos Santos Bezerra, Luiz Carlos Junior de Alcantara, Marta Giovanetti, Vagner Fonseca, Flavia Aburjaile, Rodrigo Tocantins Calado. FAMERP-SJRP: Cecília Artico Banho, Lívia Sacchetto, Fábio Sossai Possebon, Leila Sabrina Ullmann, Cíntia Bittar, Guilherme Campos, Helena Lage Ferreira, Jorge A. Petroli Marchesi, Maisa C. Pereira Parra, Marília Moraes, Paula Rahal, Paulo Inacio da Costa, João Pessoa Araújo Jr., Maurício Lacerda Nogueira. Prefeitura de Sao Paulo: Melissa Palmieri. |
| EPI_ISL_1966542 | USF DR APARECIDO RODRIGUES MOUCO SCRPARDO | Instituto Butantan / FZEA-USP (Pirassununga) | Instituto Butantan: Dimas Tadeu Covas, Sandra Coccuzzo Sampaio, Maria Carolina Elias, José Salvatore Leister Patané, Vincent Louis Viala, Antonio Jorge Martins, Ricardo Haddad, Claudia Renata dos Santos Barros, Elaine Cristina Marqueze, Raul Machado Neto, Debora Botequio Moretti, Jardelina de Souza Todao Bernardino, Loyze Paola Oliveira de Lima, Luiz Aurelio de Campos Crispin. Centro de Genômica Funcional da ESALQ: Luiz Lehmann Coutinho, Ricardo Augusto Brassaloti, Raquel de Lello Rocha Campos Cassano. NGS Soluções Genômicas: Pilar Drummond Sampaio Corrêa Mariani. FZEA-USP Pirassununga: Mirele Daiana Poleti, Jessika Cristina Chagas Lesbon, Elisangela Chicaroni Mattos, Heidge Fukumasu. USP-Botucatu: Rejane Maria Tommasini Grotto, Jayme A. Souza-Neto, Guilherme Targino Valente, Patricia Akemi Assato, Felipe Allan da Silva da Costa, Bianca Cechetto Carlos. Mendelics: Bibiana Santos, João Paulo Kitajima, Erika Freitas, David Schlesinger. Hemocentro Ribeirão Preto: Simone Kashima, Evandra Strazza Rodrigues, Svetoslav Nanev Slavov, Elaine Vieira dos Santos, Rafael dos Santos Bezerra, Luiz Carlos Junior de Alcantara, Marta Giovanetti, Vagner Fonseca, Flavia Aburjaile, Rodrigo Tocantins Calado. FAMERP-SJRP: Cecília Artico Banho, Lívia Sacchetto, Fábio Sossai Possebon, Leila Sabrina Ullmann, Cíntia Bittar, Guilherme Campos, Helena Lage Ferreira, Jorge A. Petroli Marchesi, Maisa C. Pereira Parra, Marília Moraes, Paula Rahal, Paulo Inacio da Costa, João Pessoa Araújo Jr., Maurício Lacerda Nogueira. Prefeitura de Sao Paulo: Melissa Palmieri. |
| EPI_ISL_1966543, EPI_ISL_1966544 | UBS DR ABELARDO PINHEIRO GUIMARAES SCRPARDO | Instituto Butantan / FZEA-USP (Pirassununga) | Instituto Butantan: Dimas Tadeu Covas, Sandra Coccuzzo Sampaio, Maria Carolina Elias, José Salvatore Leister Patané, Vincent Louis Viala, Antonio Jorge Martins, Ricardo Haddad, Claudia Renata dos Santos Barros, Elaine Cristina Marqueze, Raul Machado Neto, Debora Botequio Moretti, Jardelina de Souza Todao Bernardino, Loyze Paola Oliveira de Lima, Luiz Aurelio de Campos Crispin. Centro de Genômica Funcional da ESALQ: Luiz Lehmann Coutinho, Ricardo Augusto Brassaloti, Raquel de Lello Rocha Campos Cassano. NGS Soluções Genômicas: Pilar Drummond Sampaio Corrêa Mariani. FZEA-USP Pirassununga: Mirele Daiana Poleti, Jessika Cristina Chagas Lesbon, Elisangela Chicaroni Mattos, Heidge Fukumasu. USP-Botucatu: Rejane Maria Tommasini Grotto, Jayme A. Souza-Neto, Guilherme Targino Valente, Patricia Akemi Assato, Felipe Allan da Silva da Costa, Bianca Cechetto Carlos. Mendelics: Bibiana Santos, João Paulo Kitajima, Erika Freitas, David Schlesinger. Hemocentro Ribeirão Preto: Simone Kashima, Evandra Strazza Rodrigues, Svetoslav Nanev Slavov, Elaine Vieira dos Santos, Rafael dos Santos Bezerra, Luiz Carlos Junior de Alcantara, Marta Giovanetti, Vagner Fonseca, Flavia Aburjaile, Rodrigo Tocantins Calado. FAMERP-SJRP: Cecília Artico Banho, Lívia Sacchetto, Fábio Sossai Possebon, Leila Sabrina Ullmann, Cíntia Bittar, Guilherme Campos, Helena Lage Ferreira, Jorge A. Petroli Marchesi, Maisa C. Pereira Parra, Marília Moraes, Paula Rahal, Paulo Inacio da Costa, João Pessoa Araújo Jr., Maurício Lacerda Nogueira. Prefeitura de Sao Paulo: Melissa Palmieri. |
| EPI_ISL_1966545, EPI_ISL_1966546 | UBS DR WALDOMIRO FERREIRA NEVES SCRPARDO | Instituto Butantan / FZEA-USP (Pirassununga) | Instituto Butantan: Dimas Tadeu Covas, Sandra Coccuzzo Sampaio, Maria Carolina Elias, José Salvatore Leister Patané, Vincent Louis Viala, Antonio Jorge Martins, Ricardo Haddad, Claudia Renata dos Santos Barros, Elaine Cristina Marqueze, Raul Machado Neto, Debora Botequio Moretti, Jardelina de Souza Todao Bernardino, Loyze Paola Oliveira de Lima, Luiz Aurelio de Campos Crispin. Centro de Genômica Funcional da ESALQ: Luiz Lehmann Coutinho, Ricardo Augusto Brassaloti, Raquel de Lello Rocha Campos Cassano. NGS Soluções Genômicas: Pilar Drummond Sampaio Corrêa Mariani. FZEA-USP Pirassununga: Mirele Daiana Poleti, Jessika Cristina Chagas Lesbon, Elisangela Chicaroni Mattos, Heidge Fukumasu. USP-Botucatu: Rejane Maria Tommasini Grotto, Jayme A. Souza-Neto, Guilherme Targino Valente, Patricia Akemi Assato, Felipe Allan da Silva da Costa, Bianca Cechetto Carlos. Mendelics: Bibiana Santos, João Paulo Kitajima, Erika Freitas, David Schlesinger. Hemocentro Ribeirão Preto: Simone Kashima, Evandra Strazza Rodrigues, Svetoslav Nanev Slavov, Elaine Vieira dos Santos, Rafael dos Santos Bezerra, Luiz Carlos Junior de Alcantara, Marta Giovanetti, Vagner Fonseca, Flavia Aburjaile, Rodrigo Tocantins Calado. FAMERP-SJRP: Cecília Artico Banho, Lívia Sacchetto, Fábio Sossai Possebon, Leila Sabrina Ullmann, Cíntia Bittar, Guilherme Campos, Helena Lage Ferreira, Jorge A. Petroli Marchesi, Maisa C. Pereira Parra, Marília Moraes, Paula Rahal, Paulo Inacio da Costa, João Pessoa Araújo Jr., Maurício Lacerda Nogueira. Prefeitura de Sao Paulo: Melissa Palmieri. |

| | | | |
|---|---|---|---|
| EPI_ISL_1966547 | CS DE SANTA RITA DOESTE | Instituto Butantan / FZEA-USP (Pirassununga) | Instituto Butantan: Dimas Tadeu Covas, Sandra Coccuzzo Sampaio, Maria Carolina Elias, José Salvatore Leister Patané, Vincent Louis Viala, Antonio Jorge Martins, Ricardo Haddad, Claudia Renata dos Santos Barros, Elaine Cristina Marqueze, Raul Machado Neto, Debora Botequio Moretti, Jardelina de Souza Todao Bernardino, Loyze Paola Oliveira de Lima, Luiz Aurelio de Campos Crispin. Centro de Genômica Funcional da ESALQ: Luiz Lehmann Coutinho, Ricardo Augusto Brassaloti, Raquel de Lello Rocha Campos Cassano. NGS Soluções Genômicas: Pilar Drummond Sampaio Corrêa Mariani. FZEA-USP Pirassununga: Mirele Daiana Poleti, Jessika Cristina Chagas Lesbon, Elisangela Chicaroni Mattos, Heidge Fukumasu. USP-Botucatu: Rejane Maria Tommasini Grotto, Jayme A. Souza-Neto, Guilherme Targino Valente, Patricia Akemi Assato, Felipe Allan da Silva da Costa, Bianca Cechetto Carlos. Mendelics: Bibiana Santos, João Paulo Kitajima, Erika Freitas, David Schlesinger. Hemocentro Ribeirão Preto: Simone Kashima, Evandra Strazza Rodrigues, Svetoslav Nanev Slavov, Elaine Vieira dos Santos, Rafael dos Santos Bezerra, Luiz Carlos Junior de Alcantara, Marta Giovanetti, Vagner Fonseca, Flavia Aburjaile, Rodrigo Tocantins Calado. FAMERP-SJRP: Cecília Artico Banho, Lívia Sacchetto, Fábio Sossai Possebon, Leila Sabrina Ullmann, Cíntia Bittar, Guilherme Campos, Helena Lage Ferreira, Jorge A. Petroli Marchesi, Maisa C. Pereira Parra, Marília Moraes, Paula Rahal, Paulo Inacio da Costa, João Pessoa Araújo Jr., Maurício Lacerda Nogueira. Prefeitura de Sao Paulo: Melissa Palmieri. |
| EPI_ISL_1966548 | UBS II DOUTOR ORLANDO BERTOLLI | Instituto Butantan / FZEA-USP (Pirassununga) | Instituto Butantan: Dimas Tadeu Covas, Sandra Coccuzzo Sampaio, Maria Carolina Elias, José Salvatore Leister Patané, Vincent Louis Viala, Antonio Jorge Martins, Ricardo Haddad, Claudia Renata dos Santos Barros, Elaine Cristina Marqueze, Raul Machado Neto, Debora Botequio Moretti, Jardelina de Souza Todao Bernardino, Loyze Paola Oliveira de Lima, Luiz Aurelio de Campos Crispin. Centro de Genômica Funcional da ESALQ: Luiz Lehmann Coutinho, Ricardo Augusto Brassaloti, Raquel de Lello Rocha Campos Cassano. NGS Soluções Genômicas: Pilar Drummond Sampaio Corrêa Mariani. FZEA-USP Pirassununga: Mirele Daiana Poleti, Jessika Cristina Chagas Lesbon, Elisangela Chicaroni Mattos, Heidge Fukumasu. USP-Botucatu: Rejane Maria Tommasini Grotto, Jayme A. Souza-Neto, Guilherme Targino Valente, Patricia Akemi Assato, Felipe Allan da Silva da Costa, Bianca Cechetto Carlos. Mendelics: Bibiana Santos, João Paulo Kitajima, Erika Freitas, David Schlesinger. Hemocentro Ribeirão Preto: Simone Kashima, Evandra Strazza Rodrigues, Svetoslav Nanev Slavov, Elaine Vieira dos Santos, Rafael dos Santos Bezerra, Luiz Carlos Junior de Alcantara, Marta Giovanetti, Vagner Fonseca, Flavia Aburjaile, Rodrigo Tocantins Calado. FAMERP-SJRP: Cecília Artico Banho, Lívia Sacchetto, Fábio Sossai Possebon, Leila Sabrina Ullmann, Cíntia Bittar, Guilherme Campos, Helena Lage Ferreira, Jorge A. Petroli Marchesi, Maisa C. Pereira Parra, Marília Moraes, Paula Rahal, Paulo Inacio da Costa, João Pessoa Araújo Jr., Maurício Lacerda Nogueira. Prefeitura de Sao Paulo: Melissa Palmieri. |
| EPI_ISL_1966549, EPI_ISL_1966550 | UNIDADE DE VIGILANCIA EM SAUDE | Instituto Butantan / FZEA-USP (Pirassununga) | Instituto Butantan: Dimas Tadeu Covas, Sandra Coccuzzo Sampaio, Maria Carolina Elias, José Salvatore Leister Patané, Vincent Louis Viala, Antonio Jorge Martins, Ricardo Haddad, Claudia Renata dos Santos Barros, Elaine Cristina Marqueze, Raul Machado Neto, Debora Botequio Moretti, Jardelina de Souza Todao Bernardino, Loyze Paola Oliveira de Lima, Luiz Aurelio de Campos Crispin. Centro de Genômica Funcional da ESALQ: Luiz Lehmann Coutinho, Ricardo Augusto Brassaloti, Raquel de Lello Rocha Campos Cassano. NGS Soluções Genômicas: Pilar Drummond Sampaio Corrêa Mariani. FZEA-USP Pirassununga: Mirele Daiana Poleti, Jessika Cristina Chagas Lesbon, Elisangela Chicaroni Mattos, Heidge Fukumasu. USP-Botucatu: Rejane Maria Tommasini Grotto, Jayme A. Souza-Neto, Guilherme Targino Valente, Patricia Akemi Assato, Felipe Allan da Silva da Costa, Bianca Cechetto Carlos. Mendelics: Bibiana Santos, João Paulo Kitajima, Erika Freitas, David Schlesinger. Hemocentro Ribeirão Preto: Simone Kashima, Evandra Strazza Rodrigues, Svetoslav Nanev Slavov, Elaine Vieira dos Santos, Rafael dos Santos Bezerra, Luiz Carlos Junior de Alcantara, Marta Giovanetti, Vagner Fonseca, Flavia Aburjaile, Rodrigo Tocantins Calado. FAMERP-SJRP: Cecília Artico Banho, Lívia Sacchetto, Fábio Sossai Possebon, Leila Sabrina Ullmann, Cíntia Bittar, Guilherme Campos, Helena Lage Ferreira, Jorge A. Petroli Marchesi, Maisa C. Pereira Parra, Marília Moraes, Paula Rahal, Paulo Inacio da Costa, João Pessoa Araújo Jr., Maurício Lacerda Nogueira. Prefeitura de Sao Paulo: Melissa Palmieri. |
| EPI_ISL_1966551 | SECRETARIA MUNICIPAL DE SAUDE | Instituto Butantan / FZEA-USP (Pirassununga) | Instituto Butantan: Dimas Tadeu Covas, Sandra Coccuzzo Sampaio, Maria Carolina Elias, José Salvatore Leister Patané, Vincent Louis Viala, Antonio Jorge Martins, Ricardo Haddad, Claudia Renata dos Santos Barros, Elaine Cristina Marqueze, Raul Machado Neto, Debora Botequio Moretti, Jardelina de Souza Todao Bernardino, Loyze Paola Oliveira de Lima, Luiz Aurelio de Campos Crispin. Centro de Genômica Funcional da ESALQ: Luiz Lehmann Coutinho, Ricardo Augusto Brassaloti, Raquel de Lello Rocha Campos Cassano. NGS Soluções Genômicas: Pilar Drummond Sampaio Corrêa Mariani. FZEA-USP Pirassununga: Mirele Daiana Poleti, Jessika Cristina Chagas Lesbon, Elisangela Chicaroni Mattos, Heidge Fukumasu. USP-Botucatu: Rejane Maria Tommasini Grotto, Jayme A. Souza-Neto, Guilherme Targino Valente, Patricia Akemi Assato, Felipe Allan da Silva da Costa, Bianca Cechetto Carlos. Mendelics: Bibiana Santos, João Paulo Kitajima, Erika Freitas, David Schlesinger. Hemocentro Ribeirão Preto: Simone Kashima, Evandra Strazza Rodrigues, Svetoslav Nanev Slavov, Elaine Vieira dos Santos, Rafael dos Santos Bezerra, Luiz Carlos Junior de Alcantara, Marta Giovanetti, Vagner Fonseca, Flavia Aburjaile, Rodrigo Tocantins Calado. FAMERP-SJRP: Cecília Artico Banho, Lívia Sacchetto, Fábio Sossai Possebon, Leila Sabrina Ullmann, Cíntia Bittar, Guilherme Campos, Helena Lage Ferreira, Jorge A. Petroli Marchesi, Maisa C. Pereira Parra, Marília Moraes, Paula Rahal, Paulo Inacio da Costa, João Pessoa Araújo Jr., Maurício Lacerda Nogueira. Prefeitura de Sao Paulo: Melissa Palmieri. |
| EPI_ISL_1966555 | UNIDADE BASICA DE SAUDE II DE TARABAI | Instituto Butantan / FZEA-USP (Pirassununga) | Instituto Butantan: Dimas Tadeu Covas, Sandra Coccuzzo Sampaio, Maria Carolina Elias, José Salvatore Leister Patané, Vincent Louis Viala, Antonio Jorge Martins, Ricardo Haddad, Claudia Renata dos Santos Barros, Elaine Cristina Marqueze, Raul Machado Neto, Debora Botequio Moretti, Jardelina de Souza Todao Bernardino, Loyze Paola Oliveira de Lima, Luiz Aurelio de Campos Crispin. Centro de Genômica Funcional da ESALQ: Luiz Lehmann Coutinho, Ricardo Augusto Brassaloti, Raquel de Lello Rocha Campos Cassano. NGS Soluções Genômicas: Pilar Drummond Sampaio Corrêa Mariani. FZEA-USP Pirassununga: Mirele Daiana Poleti, Jessika Cristina Chagas Lesbon, Elisangela Chicaroni Mattos, Heidge Fukumasu. USP-Botucatu: Rejane Maria Tommasini Grotto, Jayme A. Souza-Neto, Guilherme Targino Valente, Patricia Akemi Assato, Felipe Allan da Silva da Costa, Bianca Cechetto Carlos. Mendelics: Bibiana Santos, João Paulo Kitajima, Erika Freitas, David Schlesinger. Hemocentro Ribeirão Preto: Simone Kashima, Evandra Strazza Rodrigues, Svetoslav Nanev Slavov, Elaine Vieira dos Santos, Rafael dos Santos Bezerra, Luiz Carlos Junior de Alcantara, Marta Giovanetti, Vagner Fonseca, Flavia Aburjaile, Rodrigo Tocantins Calado. FAMERP-SJRP: Cecília Artico Banho, Lívia Sacchetto, Fábio Sossai Possebon, Leila Sabrina Ullmann, Cíntia Bittar, Guilherme Campos, Helena Lage Ferreira, Jorge A. Petroli Marchesi, Maisa C. Pereira Parra, Marília Moraes, Paula Rahal, Paulo Inacio da Costa, João Pessoa Araújo Jr., Maurício Lacerda Nogueira. Prefeitura de Sao Paulo: Melissa Palmieri. |
| EPI_ISL_1966556, EPI_ISL_1966557, EPI_ISL_1966558, EPI_ISL_1966559 | VIGILANCIA EM SAUDE SAE VIGILANCIA EPIDEMIOLOGICA | Instituto Butantan / Mendelics | Instituto Butantan: Dimas Tadeu Covas, Sandra Coccuzzo Sampaio, Maria Carolina Elias, José Salvatore Leister Patané, Vincent Louis Viala, Antonio Jorge Martins, Ricardo Haddad, Claudia Renata dos Santos Barros, Elaine Cristina Marqueze, Raul Machado Neto, Debora Botequio Moretti, Jardelina de Souza Todao Bernardino, Loyze Paola Oliveira de Lima, Luiz Aurelio de Campos Crispin. Centro de Genômica Funcional da ESALQ: Luiz Lehmann Coutinho, Ricardo Augusto Brassaloti, Raquel de Lello Rocha Campos Cassano. NGS Soluções Genômicas: Pilar Drummond Sampaio Corrêa Mariani. FZEA-USP Pirassununga: Mirele Daiana Poleti, Jessika Cristina Chagas Lesbon, Elisangela Chicaroni Mattos, Heidge Fukumasu. USP-Botucatu: Rejane Maria Tommasini Grotto, Jayme A. Souza-Neto, Guilherme Targino Valente, Patricia Akemi Assato, Felipe Allan da Silva da Costa, Bianca Cechetto Carlos. Mendelics: Bibiana Santos, João Paulo Kitajima, Erika Freitas, David Schlesinger. Hemocentro Ribeirão Preto: Simone Kashima, Evandra Strazza Rodrigues, Svetoslav Nanev Slavov, Elaine Vieira dos Santos, Rafael dos Santos Bezerra, Luiz Carlos Junior de Alcantara, Marta Giovanetti, Vagner Fonseca, Flavia Aburjaile, Rodrigo Tocantins Calado. FAMERP-SJRP: Cecília Artico Banho, Lívia Sacchetto, Fábio Sossai Possebon, Leila Sabrina Ullmann, Cíntia Bittar, Guilherme Campos, Helena Lage Ferreira, Jorge A. Petroli Marchesi, Maisa C. Pereira Parra, Marília Moraes, Paula Rahal, Paulo Inacio da Costa, João Pessoa Araújo Jr., Maurício Lacerda Nogueira. Prefeitura de Sao Paulo: Melissa Palmieri. |
| EPI_ISL_1966562 | NUCLEO DE SAUDE VILA FALCAO DE BAURU | Instituto Butantan / Mendelics | Instituto Butantan: Dimas Tadeu Covas, Sandra Coccuzzo Sampaio, Maria Carolina Elias, José Salvatore Leister Patané, Vincent Louis Viala, Antonio Jorge Martins, Ricardo Haddad, Claudia Renata dos Santos Barros, Elaine Cristina Marqueze, Raul Machado Neto, Debora Botequio Moretti, Jardelina de Souza Todao Bernardino, Loyze Paola Oliveira de Lima, Luiz Aurelio de Campos Crispin. Centro de Genômica Funcional da ESALQ: Luiz Lehmann Coutinho, Ricardo Augusto Brassaloti, Raquel de Lello Rocha Campos Cassano. NGS Soluções Genômicas: Pilar Drummond Sampaio Corrêa Mariani. FZEA-USP Pirassununga: Mirele Daiana Poleti, Jessika Cristina Chagas Lesbon, Elisangela Chicaroni Mattos, Heidge Fukumasu. USP-Botucatu: Rejane Maria Tommasini Grotto, Jayme A. Souza-Neto, Guilherme Targino Valente, Patricia Akemi Assato, Felipe Allan da Silva da Costa, Bianca Cechetto Carlos. Mendelics: Bibiana Santos, João Paulo Kitajima, Erika Freitas, David Schlesinger. Hemocentro Ribeirão Preto: Simone Kashima, Evandra Strazza Rodrigues, Svetoslav Nanev Slavov, Elaine Vieira dos Santos, Rafael dos Santos Bezerra, Luiz Carlos Junior de Alcantara, Marta Giovanetti, Vagner Fonseca, Flavia Aburjaile, Rodrigo Tocantins Calado. FAMERP-SJRP: Cecília Artico Banho, Lívia Sacchetto, Fábio Sossai Possebon, Leila Sabrina Ullmann, Cíntia Bittar, Guilherme Campos, Helena Lage Ferreira, Jorge A. Petroli Marchesi, Maisa C. Pereira Parra, Marília Moraes, Paula Rahal, Paulo Inacio da Costa, João Pessoa Araújo Jr., Maurício Lacerda Nogueira. Prefeitura de Sao Paulo: Melissa Palmieri. |
| EPI_ISL_1966565 | UPA BELA VISTA | Instituto Butantan / Mendelics | Instituto Butantan: Dimas Tadeu Covas, Sandra Coccuzzo Sampaio, Maria Carolina Elias, José Salvatore Leister Patané, Vincent Louis Viala, Antonio Jorge Martins, Ricardo Haddad, Claudia Renata dos Santos Barros, Elaine Cristina Marqueze, Raul Machado Neto, Debora Botequio Moretti, Jardelina de Souza Todao Bernardino, Loyze Paola Oliveira de Lima, Luiz Aurelio de Campos Crispin. Centro de Genômica Funcional da ESALQ: Luiz Lehmann Coutinho, Ricardo Augusto Brassaloti, Raquel de Lello Rocha Campos Cassano. NGS Soluções Genômicas: Pilar Drummond Sampaio Corrêa Mariani. FZEA-USP Pirassununga: Mirele Daiana Poleti, Jessika Cristina Chagas Lesbon, Elisangela Chicaroni Mattos, Heidge Fukumasu. USP-Botucatu: Rejane Maria Tommasini Grotto, Jayme A. Souza-Neto, Guilherme Targino Valente, Patricia Akemi Assato, Felipe Allan da Silva da Costa, Bianca Cechetto Carlos. Mendelics: Bibiana Santos, João Paulo Kitajima, Erika Freitas, David Schlesinger. Hemocentro Ribeirão Preto: Simone Kashima, Evandra Strazza |

| | | | |
|---|---|---|---|
| | | | Rodrigues, Svetoslav Nanev Slavov, Elaine Vieira dos Santos, Rafael dos Santos Bezerra, Luiz Carlos Junior de Alcantara, Marta Giovanetti, Vagner Fonseca, Flavia Aburjaile, Rodrigo Tocantins Calado. FAMERP-SJRP: Cecília Artico Banho, Lívia Sacchetto, Fábio Sossai Possebon, Leila Sabrina Ullmann, Cíntia Bittar, Guilherme Campos, Helena Lage Ferreira, Jorge A. Petroli Marchesi, Maisa C. Pereira Parra, Marília Moraes, Paula Rahal, Paulo Inacio da Costa, João Pessoa Araújo Jr., Maurício Lacerda Nogueira. Prefeitura de Sao Paulo: Melissa Palmieri. |
| EPI_ISL_1966567 | NUCLEO DE SAUDE VILA FALCAO DE BAURU | Instituto Butantan / Mendelics | Instituto Butantan: Dimas Tadeu Covas, Sandra Coccuzzo Sampaio, Maria Carolina Elias, José Salvatore Leister Patané, Vincent Louis Viala, Antonio Jorge Martins, Ricardo Haddad, Claudia Renata dos Santos Barros, Elaine Cristina Marqueze, Raul Machado Neto, Debora Botequio Moretti, Jardelina de Souza Todao Bernardino, Loyze Paola Oliveira de Lima, Luiz Aurelio de Campos Crispin. Centro de Genômica Funcional da ESALQ: Luiz Lehmann Coutinho, Ricardo Augusto Brassaloti, Raquel de Lello Rocha Campos Cassano. NGS Soluções Genômicas: Pilar Drummond Sampaio Corrêa Mariani. FZEA-USP Pirassununga: Mirele Daiana Poleti, Jessika Cristina Chagas Lesbon, Elisangela Chicaroni Mattos, Heidge Fukumasu. USP-Botucatu: Rejane Maria Tommasini Grotto, Jayme A. Souza-Neto, Guilherme Targino Valente, Patricia Akemi Assato, Felipe Allan da Silva da Costa, Bianca Cechetto Carlos. Mendelics: Bibiana Santos, João Paulo Kitajima, Erika Freitas, David Schlesinger. Hemocentro Ribeirão Preto: Simone Kashima, Evandra Strazza Rodrigues, Svetoslav Nanev Slavov, Elaine Vieira dos Santos, Rafael dos Santos Bezerra, Luiz Carlos Junior de Alcantara, Marta Giovanetti, Vagner Fonseca, Flavia Aburjaile, Rodrigo Tocantins Calado. FAMERP-SJRP: Cecília Artico Banho, Lívia Sacchetto, Fábio Sossai Possebon, Leila Sabrina Ullmann, Cíntia Bittar, Guilherme Campos, Helena Lage Ferreira, Jorge A. Petroli Marchesi, Maisa C. Pereira Parra, Marília Moraes, Paula Rahal, Paulo Inacio da Costa, João Pessoa Araújo Jr., Maurício Lacerda Nogueira. Prefeitura de Sao Paulo: Melissa Palmieri. |
| EPI_ISL_1966569 | UPA BELA VISTA | Instituto Butantan / Mendelics | Instituto Butantan: Dimas Tadeu Covas, Sandra Coccuzzo Sampaio, Maria Carolina Elias, José Salvatore Leister Patané, Vincent Louis Viala, Antonio Jorge Martins, Ricardo Haddad, Claudia Renata dos Santos Barros, Elaine Cristina Marqueze, Raul Machado Neto, Debora Botequio Moretti, Jardelina de Souza Todao Bernardino, Loyze Paola Oliveira de Lima, Luiz Aurelio de Campos Crispin. Centro de Genômica Funcional da ESALQ: Luiz Lehmann Coutinho, Ricardo Augusto Brassaloti, Raquel de Lello Rocha Campos Cassano. NGS Soluções Genômicas: Pilar Drummond Sampaio Corrêa Mariani. FZEA-USP Pirassununga: Mirele Daiana Poleti, Jessika Cristina Chagas Lesbon, Elisangela Chicaroni Mattos, Heidge Fukumasu. USP-Botucatu: Rejane Maria Tommasini Grotto, Jayme A. Souza-Neto, Guilherme Targino Valente, Patricia Akemi Assato, Felipe Allan da Silva da Costa, Bianca Cechetto Carlos. Mendelics: Bibiana Santos, João Paulo Kitajima, Erika Freitas, David Schlesinger. Hemocentro Ribeirão Preto: Simone Kashima, Evandra Strazza Rodrigues, Svetoslav Nanev Slavov, Elaine Vieira dos Santos, Rafael dos Santos Bezerra, Luiz Carlos Junior de Alcantara, Marta Giovanetti, Vagner Fonseca, Flavia Aburjaile, Rodrigo Tocantins Calado. FAMERP-SJRP: Cecília Artico Banho, Lívia Sacchetto, Fábio Sossai Possebon, Leila Sabrina Ullmann, Cíntia Bittar, Guilherme Campos, Helena Lage Ferreira, Jorge A. Petroli Marchesi, Maisa C. Pereira Parra, Marília Moraes, Paula Rahal, Paulo Inacio da Costa, João Pessoa Araújo Jr., Maurício Lacerda Nogueira. Prefeitura de Sao Paulo: Melissa Palmieri. |
| EPI_ISL_1966571 | NUCLEO DE SAUDE VILA FALCAO DE BAURU | Instituto Butantan / Mendelics | Instituto Butantan: Dimas Tadeu Covas, Sandra Coccuzzo Sampaio, Maria Carolina Elias, José Salvatore Leister Patané, Vincent Louis Viala, Antonio Jorge Martins, Ricardo Haddad, Claudia Renata dos Santos Barros, Elaine Cristina Marqueze, Raul Machado Neto, Debora Botequio Moretti, Jardelina de Souza Todao Bernardino, Loyze Paola Oliveira de Lima, Luiz Aurelio de Campos Crispin. Centro de Genômica Funcional da ESALQ: Luiz Lehmann Coutinho, Ricardo Augusto Brassaloti, Raquel de Lello Rocha Campos Cassano. NGS Soluções Genômicas: Pilar Drummond Sampaio Corrêa Mariani. FZEA-USP Pirassununga: Mirele Daiana Poleti, Jessika Cristina Chagas Lesbon, Elisangela Chicaroni Mattos, Heidge Fukumasu. USP-Botucatu: Rejane Maria Tommasini Grotto, Jayme A. Souza-Neto, Guilherme Targino Valente, Patricia Akemi Assato, Felipe Allan da Silva da Costa, Bianca Cechetto Carlos. Mendelics: Bibiana Santos, João Paulo Kitajima, Erika Freitas, David Schlesinger. Hemocentro Ribeirão Preto: Simone Kashima, Evandra Strazza Rodrigues, Svetoslav Nanev Slavov, Elaine Vieira dos Santos, Rafael dos Santos Bezerra, Luiz Carlos Junior de Alcantara, Marta Giovanetti, Vagner Fonseca, Flavia Aburjaile, Rodrigo Tocantins Calado. FAMERP-SJRP: Cecília Artico Banho, Lívia Sacchetto, Fábio Sossai Possebon, Leila Sabrina Ullmann, Cíntia Bittar, Guilherme Campos, Helena Lage Ferreira, Jorge A. Petroli Marchesi, Maisa C. Pereira Parra, Marília Moraes, Paula Rahal, Paulo Inacio da Costa, João Pessoa Araújo Jr., Maurício Lacerda Nogueira. Prefeitura de Sao Paulo: Melissa Palmieri. |
| EPI_ISL_1966573 | UPA BELA VISTA | Instituto Butantan / Mendelics | Instituto Butantan: Dimas Tadeu Covas, Sandra Coccuzzo Sampaio, Maria Carolina Elias, José Salvatore Leister Patané, Vincent Louis Viala, Antonio Jorge Martins, Ricardo Haddad, Claudia Renata dos Santos Barros, Elaine Cristina Marqueze, Raul Machado Neto, Debora Botequio Moretti, Jardelina de Souza Todao Bernardino, Loyze Paola Oliveira de Lima, Luiz Aurelio de Campos Crispin. Centro de Genômica Funcional da ESALQ: Luiz Lehmann Coutinho, Ricardo Augusto Brassaloti, Raquel de Lello Rocha Campos Cassano. NGS Soluções Genômicas: Pilar Drummond Sampaio Corrêa Mariani. FZEA-USP Pirassununga: Mirele Daiana Poleti, Jessika Cristina Chagas Lesbon, Elisangela Chicaroni Mattos, Heidge Fukumasu. USP-Botucatu: Rejane Maria Tommasini Grotto, Jayme A. Souza-Neto, Guilherme Targino Valente, Patricia Akemi Assato, Felipe Allan da Silva da Costa, Bianca Cechetto Carlos. Mendelics: Bibiana Santos, João Paulo Kitajima, Erika Freitas, David Schlesinger. Hemocentro Ribeirão Preto: Simone Kashima, Evandra Strazza Rodrigues, Svetoslav Nanev Slavov, Elaine Vieira dos Santos, Rafael dos Santos Bezerra, Luiz Carlos Junior de Alcantara, Marta Giovanetti, Vagner Fonseca, Flavia Aburjaile, Rodrigo Tocantins Calado. FAMERP-SJRP: Cecília Artico Banho, Lívia Sacchetto, Fábio Sossai Possebon, Leila Sabrina Ullmann, Cíntia Bittar, Guilherme Campos, Helena Lage Ferreira, Jorge A. Petroli Marchesi, Maisa C. Pereira Parra, Marília Moraes, Paula Rahal, Paulo Inacio da Costa, João Pessoa Araújo Jr., Maurício Lacerda Nogueira. Prefeitura de Sao Paulo: Melissa Palmieri. |
| EPI_ISL_1966576, EPI_ISL_1966578, EPI_ISL_1966579, EPI_ISL_1966580, EPI_ISL_1966581, EPI_ISL_1966582, EPI_ISL_1966583, EPI_ISL_1966584, EPI_ISL_1966585, EPI_ISL_1966586, EPI_ISL_1966587, EPI_ISL_1966589, EPI_ISL_1966590, EPI_ISL_1966592 | | | |
| see above | NUCLEO DE SAUDE VILA FALCAO DE BAURU | Instituto Butantan / Mendelics | Instituto Butantan: Dimas Tadeu Covas, Sandra Coccuzzo Sampaio, Maria Carolina Elias, José Salvatore Leister Patané, Vincent Louis Viala, Antonio Jorge Martins, Ricardo Haddad, Claudia Renata dos Santos Barros, Elaine Cristina Marqueze, Raul Machado Neto, Debora Botequio Moretti, Jardelina de Souza Todao Bernardino, Loyze Paola Oliveira de Lima, Luiz Aurelio de Campos Crispin. Centro de Genômica Funcional da ESALQ: Luiz Lehmann Coutinho, Ricardo Augusto Brassaloti, Raquel de Lello Rocha Campos Cassano. NGS Soluções Genômicas: Pilar Drummond Sampaio Corrêa Mariani. FZEA-USP Pirassununga: Mirele Daiana Poleti, Jessika Cristina Chagas Lesbon, Elisangela Chicaroni Mattos, Heidge Fukumasu. USP-Botucatu: Rejane Maria Tommasini Grotto, Jayme A. Souza-Neto, Guilherme Targino Valente, Patricia Akemi Assato, Felipe Allan da Silva da Costa, Bianca Cechetto Carlos. Mendelics: Bibiana Santos, João Paulo Kitajima, Erika Freitas, David Schlesinger. Hemocentro Ribeirão Preto: Simone Kashima, Evandra Strazza Rodrigues, Svetoslav Nanev Slavov, Elaine Vieira dos Santos, Rafael dos Santos Bezerra, Luiz Carlos Junior de Alcantara, Marta Giovanetti, Vagner Fonseca, Flavia Aburjaile, Rodrigo Tocantins Calado. FAMERP-SJRP: Cecília Artico Banho, Lívia Sacchetto, Fábio Sossai Possebon, Leila Sabrina Ullmann, Cíntia Bittar, Guilherme Campos, Helena Lage Ferreira, Jorge A. Petroli Marchesi, Maisa C. Pereira Parra, Marília Moraes, Paula Rahal, Paulo Inacio da Costa, João Pessoa Araújo Jr., Maurício Lacerda Nogueira. Prefeitura de Sao Paulo: Melissa Palmieri. |
| EPI_ISL_1966593, EPI_ISL_1966594, EPI_ISL_1966595, EPI_ISL_1966596, EPI_ISL_1966597, EPI_ISL_1966598, EPI_ISL_1966599, EPI_ISL_1966600, EPI_ISL_1966601 | UPA DE BEBEDOURO | Instituto Butantan / Mendelics | Instituto Butantan: Dimas Tadeu Covas, Sandra Coccuzzo Sampaio, Maria Carolina Elias, José Salvatore Leister Patané, Vincent Louis Viala, Antonio Jorge Martins, Ricardo Haddad, Claudia Renata dos Santos Barros, Elaine Cristina Marqueze, Raul Machado Neto, Debora Botequio Moretti, Jardelina de Souza Todao Bernardino, Loyze Paola Oliveira de Lima, Luiz Aurelio de Campos Crispin. Centro de Genômica Funcional da ESALQ: Luiz Lehmann Coutinho, Ricardo Augusto Brassaloti, Raquel de Lello Rocha Campos Cassano. NGS Soluções Genômicas: Pilar Drummond Sampaio Corrêa Mariani. FZEA-USP Pirassununga: Mirele Daiana Poleti, Jessika Cristina Chagas Lesbon, Elisangela Chicaroni Mattos, Heidge Fukumasu. USP-Botucatu: Rejane Maria Tommasini Grotto, Jayme A. Souza-Neto, Guilherme Targino Valente, Patricia Akemi Assato, Felipe Allan da Silva da Costa, Bianca Cechetto Carlos. Mendelics: Bibiana Santos, João Paulo Kitajima, Erika Freitas, David Schlesinger. Hemocentro Ribeirão Preto: Simone Kashima, Evandra Strazza Rodrigues, Svetoslav Nanev Slavov, Elaine Vieira dos Santos, Rafael dos Santos Bezerra, Luiz Carlos Junior de Alcantara, Marta Giovanetti, Vagner Fonseca, Flavia Aburjaile, Rodrigo Tocantins Calado. FAMERP-SJRP: Cecília Artico Banho, Lívia Sacchetto, Fábio Sossai Possebon, Leila Sabrina Ullmann, Cíntia Bittar, Guilherme Campos, Helena Lage Ferreira, Jorge A. Petroli Marchesi, Maisa C. Pereira Parra, Marília Moraes, Paula Rahal, Paulo Inacio da Costa, João Pessoa Araújo Jr., Maurício Lacerda Nogueira. Prefeitura de Sao Paulo: Melissa Palmieri. |
| EPI_ISL_1966603, EPI_ISL_1966605, EPI_ISL_1966606 | UNIDADE RESPIRATORIA NOVA HORTOLANDIA | Instituto Butantan / Mendelics | Instituto Butantan: Dimas Tadeu Covas, Sandra Coccuzzo Sampaio, Maria Carolina Elias, José Salvatore Leister Patané, Vincent Louis Viala, Antonio Jorge Martins, Ricardo Haddad, Claudia Renata dos Santos Barros, Elaine Cristina Marqueze, Raul Machado Neto, Debora Botequio Moretti, Jardelina de Souza Todao Bernardino, Loyze Paola Oliveira de Lima, Luiz Aurelio de Campos Crispin. Centro de Genômica Funcional da ESALQ: Luiz Lehmann Coutinho, Ricardo Augusto Brassaloti, Raquel de Lello Rocha Campos Cassano. NGS Soluções Genômicas: Pilar Drummond Sampaio Corrêa Mariani. FZEA-USP Pirassununga: Mirele Daiana Poleti, Jessika Cristina Chagas Lesbon, Elisangela Chicaroni Mattos, Heidge Fukumasu. USP-Botucatu: Rejane Maria Tommasini Grotto, Jayme A. Souza-Neto, Guilherme Targino Valente, Patricia Akemi Assato, Felipe Allan da Silva da Costa, Bianca Cechetto Carlos. Mendelics: Bibiana Santos, João Paulo Kitajima, Erika Freitas, David Schlesinger. Hemocentro Ribeirão Preto: Simone Kashima, Evandra Strazza Rodrigues, Svetoslav Nanev Slavov, Elaine Vieira dos Santos, Rafael dos Santos Bezerra, Luiz Carlos Junior de Alcantara, Marta Giovanetti, Vagner Fonseca, Flavia Aburjaile, Rodrigo Tocantins Calado. FAMERP-SJRP: Cecília Artico Banho, Lívia Sacchetto, Fábio Sossai Possebon, Leila Sabrina Ullmann, Cíntia Bittar, Guilherme Campos, Helena Lage Ferreira, Jorge A. Petroli Marchesi, Maisa C. Pereira Parra, Marília Moraes, Paula Rahal, Paulo Inacio da Costa, João Pessoa Araújo Jr., Maurício Lacerda Nogueira. Prefeitura de Sao Paulo: Melissa Palmieri. |
| EPI_ISL_1966607 | VIGILANCIA EPIDEMIOLOGICA DE IBATE | Instituto Butantan / Mendelics | Instituto Butantan: Dimas Tadeu Covas, Sandra Coccuzzo Sampaio, Maria Carolina Elias, José Salvatore Leister Patané, Vincent Louis Viala, Antonio |

| | | | |
|---|---|---|---|
| | | | Jorge Martins, Ricardo Haddad, Claudia Renata dos Santos Barros, Elaine Cristina Marqueze, Raul Machado Neto, Debora Botequio Moretti, Jardelina de Souza Todao Bernardino, Loyze Paola Oliveira de Lima, Luiz Aurelio de Campos Crispin. Centro de Genômica Funcional da ESALQ: Luiz Lehmann Coutinho, Ricardo Augusto Brassaloti, Raquel de Lello Rocha Campos Cassano. NGS Soluções Genômicas: Pilar Drummond Sampaio Corrêa Mariani. FZEA-USP Pirassununga: Mirele Daiana Poleti, Jessika Cristina Chagas Lesbon, Elisangela Chicaroni Mattos, Heidge Fukumasu. USP-Botucatu: Rejane Maria Tommasini Grotto, Jayme A. Souza-Neto, Guilherme Targino Valente, Patricia Akemi Assato, Felipe Allan da Silva da Costa, Bianca Cechetto Carlos. Mendelics: Bibiana Santos, João Paulo Kitajima, Erika Freitas, David Schlesinger. Hemocentro Ribeirão Preto: Simone Kashima, Evandra Strazza Rodrigues, Svetoslav Nanev Slavov, Elaine Vieira dos Santos, Rafael dos Santos Bezerra, Luiz Carlos Junior de Alcantara, Marta Giovanetti, Vagner Fonseca, Flavia Aburjaile, Rodrigo Tocantins Calado. FAMERP-SJRP: Cecília Artico Banho, Lívia Sacchetto, Fábio Sossai Possebon, Leila Sabrina Ullmann, Cíntia Bittar, Guilherme Campos, Helena Lage Ferreira, Jorge A. Petroli Marchesi, Maisa C. Pereira Parra, Marília Moraes, Paula Rahal, Paulo Inacio da Costa, João Pessoa Araújo Jr., Maurício Lacerda Nogueira. Prefeitura de Sao Paulo: Melissa Palmieri. |
| EPI_ISL_1966608, EPI_ISL_1966609, EPI_ISL_1966610, EPI_ISL_1966611 | LABORATORIO MUNICIPAL DE ANALISES CLINICAS DE ITANHAEM | Instituto Butantan / Mendelics | Instituto Butantan: Dimas Tadeu Covas, Sandra Coccuzzo Sampaio, Maria Carolina Elias, José Salvatore Leister Patané, Vincent Louis Viala, Antonio Jorge Martins, Ricardo Haddad, Claudia Renata dos Santos Barros, Elaine Cristina Marqueze, Raul Machado Neto, Debora Botequio Moretti, Jardelina de Souza Todao Bernardino, Loyze Paola Oliveira de Lima, Luiz Aurelio de Campos Crispin. Centro de Genômica Funcional da ESALQ: Luiz Lehmann Coutinho, Ricardo Augusto Brassaloti, Raquel de Lello Rocha Campos Cassano. NGS Soluções Genômicas: Pilar Drummond Sampaio Corrêa Mariani. FZEA-USP Pirassununga: Mirele Daiana Poleti, Jessika Cristina Chagas Lesbon, Elisangela Chicaroni Mattos, Heidge Fukumasu. USP-Botucatu: Rejane Maria Tommasini Grotto, Jayme A. Souza-Neto, Guilherme Targino Valente, Patricia Akemi Assato, Felipe Allan da Silva da Costa, Bianca Cechetto Carlos. Mendelics: Bibiana Santos, João Paulo Kitajima, Erika Freitas, David Schlesinger. Hemocentro Ribeirão Preto: Simone Kashima, Evandra Strazza Rodrigues, Svetoslav Nanev Slavov, Elaine Vieira dos Santos, Rafael dos Santos Bezerra, Luiz Carlos Junior de Alcantara, Marta Giovanetti, Vagner Fonseca, Flavia Aburjaile, Rodrigo Tocantins Calado. FAMERP-SJRP: Cecília Artico Banho, Lívia Sacchetto, Fábio Sossai Possebon, Leila Sabrina Ullmann, Cíntia Bittar, Guilherme Campos, Helena Lage Ferreira, Jorge A. Petroli Marchesi, Maisa C. Pereira Parra, Marília Moraes, Paula Rahal, Paulo Inacio da Costa, João Pessoa Araújo Jr., Maurício Lacerda Nogueira. Prefeitura de Sao Paulo: Melissa Palmieri. |
| EPI_ISL_1966612, EPI_ISL_1966613, EPI_ISL_1966614, EPI_ISL_1966615 | UNIDADE DE PRONTO ATENDIMENTO UPA DRA ANA OLIVIA BENTIVOGLIO | Instituto Butantan / Mendelics | Instituto Butantan: Dimas Tadeu Covas, Sandra Coccuzzo Sampaio, Maria Carolina Elias, José Salvatore Leister Patané, Vincent Louis Viala, Antonio Jorge Martins, Ricardo Haddad, Claudia Renata dos Santos Barros, Elaine Cristina Marqueze, Raul Machado Neto, Debora Botequio Moretti, Jardelina de Souza Todao Bernardino, Loyze Paola Oliveira de Lima, Luiz Aurelio de Campos Crispin. Centro de Genômica Funcional da ESALQ: Luiz Lehmann Coutinho, Ricardo Augusto Brassaloti, Raquel de Lello Rocha Campos Cassano. NGS Soluções Genômicas: Pilar Drummond Sampaio Corrêa Mariani. FZEA-USP Pirassununga: Mirele Daiana Poleti, Jessika Cristina Chagas Lesbon, Elisangela Chicaroni Mattos, Heidge Fukumasu. USP-Botucatu: Rejane Maria Tommasini Grotto, Jayme A. Souza-Neto, Guilherme Targino Valente, Patricia Akemi Assato, Felipe Allan da Silva da Costa, Bianca Cechetto Carlos. Mendelics: Bibiana Santos, João Paulo Kitajima, Erika Freitas, David Schlesinger. Hemocentro Ribeirão Preto: Simone Kashima, Evandra Strazza Rodrigues, Svetoslav Nanev Slavov, Elaine Vieira dos Santos, Rafael dos Santos Bezerra, Luiz Carlos Junior de Alcantara, Marta Giovanetti, Vagner Fonseca, Flavia Aburjaile, Rodrigo Tocantins Calado. FAMERP-SJRP: Cecília Artico Banho, Lívia Sacchetto, Fábio Sossai Possebon, Leila Sabrina Ullmann, Cíntia Bittar, Guilherme Campos, Helena Lage Ferreira, Jorge A. Petroli Marchesi, Maisa C. Pereira Parra, Marília Moraes, Paula Rahal, Paulo Inacio da Costa, João Pessoa Araújo Jr., Maurício Lacerda Nogueira. Prefeitura de Sao Paulo: Melissa Palmieri. |
| EPI_ISL_1966616, EPI_ISL_1966617, EPI_ISL_1966618, EPI_ISL_1966619, EPI_ISL_1966620, EPI_ISL_1966621, EPI_ISL_1966622, EPI_ISL_1966623, EPI_ISL_1966624, EPI_ISL_1966625 | POLICLINICA HORTOLANDIA | Instituto Butantan / Mendelics | Instituto Butantan: Dimas Tadeu Covas, Sandra Coccuzzo Sampaio, Maria Carolina Elias, José Salvatore Leister Patané, Vincent Louis Viala, Antonio Jorge Martins, Ricardo Haddad, Claudia Renata dos Santos Barros, Elaine Cristina Marqueze, Raul Machado Neto, Debora Botequio Moretti, Jardelina de Souza Todao Bernardino, Loyze Paola Oliveira de Lima, Luiz Aurelio de Campos Crispin. Centro de Genômica Funcional da ESALQ: Luiz Lehmann Coutinho, Ricardo Augusto Brassaloti, Raquel de Lello Rocha Campos Cassano. NGS Soluções Genômicas: Pilar Drummond Sampaio Corrêa Mariani. FZEA-USP Pirassununga: Mirele Daiana Poleti, Jessika Cristina Chagas Lesbon, Elisangela Chicaroni Mattos, Heidge Fukumasu. USP-Botucatu: Rejane Maria Tommasini Grotto, Jayme A. Souza-Neto, Guilherme Targino Valente, Patricia Akemi Assato, Felipe Allan da Silva da Costa, Bianca Cechetto Carlos. Mendelics: Bibiana Santos, João Paulo Kitajima, Erika Freitas, David Schlesinger. Hemocentro Ribeirão Preto: Simone Kashima, Evandra Strazza Rodrigues, Svetoslav Nanev Slavov, Elaine Vieira dos Santos, Rafael dos Santos Bezerra, Luiz Carlos Junior de Alcantara, Marta Giovanetti, Vagner Fonseca, Flavia Aburjaile, Rodrigo Tocantins Calado. FAMERP-SJRP: Cecília Artico Banho, Lívia Sacchetto, Fábio Sossai Possebon, Leila Sabrina Ullmann, Cíntia Bittar, Guilherme Campos, Helena Lage Ferreira, Jorge A. Petroli Marchesi, Maisa C. Pereira Parra, Marília Moraes, Paula Rahal, Paulo Inacio da Costa, João Pessoa Araújo Jr., Maurício Lacerda Nogueira. Prefeitura de Sao Paulo: Melissa Palmieri. |
| EPI_ISL_1966627, EPI_ISL_1966628, EPI_ISL_1966630, EPI_ISL_1966631, EPI_ISL_1966632, EPI_ISL_1966633 | SECAO CENTRO DE DIAGNOSTICO SECEDI | Instituto Butantan / Mendelics | Instituto Butantan: Dimas Tadeu Covas, Sandra Coccuzzo Sampaio, Maria Carolina Elias, José Salvatore Leister Patané, Vincent Louis Viala, Antonio Jorge Martins, Ricardo Haddad, Claudia Renata dos Santos Barros, Elaine Cristina Marqueze, Raul Machado Neto, Debora Botequio Moretti, Jardelina de Souza Todao Bernardino, Loyze Paola Oliveira de Lima, Luiz Aurelio de Campos Crispin. Centro de Genômica Funcional da ESALQ: Luiz Lehmann Coutinho, Ricardo Augusto Brassaloti, Raquel de Lello Rocha Campos Cassano. NGS Soluções Genômicas: Pilar Drummond Sampaio Corrêa Mariani. FZEA-USP Pirassununga: Mirele Daiana Poleti, Jessika Cristina Chagas Lesbon, Elisangela Chicaroni Mattos, Heidge Fukumasu. USP-Botucatu: Rejane Maria Tommasini Grotto, Jayme A. Souza-Neto, Guilherme Targino Valente, Patricia Akemi Assato, Felipe Allan da Silva da Costa, Bianca Cechetto Carlos. Mendelics: Bibiana Santos, João Paulo Kitajima, Erika Freitas, David Schlesinger. Hemocentro Ribeirão Preto: Simone Kashima, Evandra Strazza Rodrigues, Svetoslav Nanev Slavov, Elaine Vieira dos Santos, Rafael dos Santos Bezerra, Luiz Carlos Junior de Alcantara, Marta Giovanetti, Vagner Fonseca, Flavia Aburjaile, Rodrigo Tocantins Calado. FAMERP-SJRP: Cecília Artico Banho, Lívia Sacchetto, Fábio Sossai Possebon, Leila Sabrina Ullmann, Cíntia Bittar, Guilherme Campos, Helena Lage Ferreira, Jorge A. Petroli Marchesi, Maisa C. Pereira Parra, Marília Moraes, Paula Rahal, Paulo Inacio da Costa, João Pessoa Araújo Jr., Maurício Lacerda Nogueira. Prefeitura de Sao Paulo: Melissa Palmieri. |
| EPI_ISL_1966634, EPI_ISL_1966635, EPI_ISL_1966636, EPI_ISL_1966637, EPI_ISL_1966638, EPI_ISL_1966639 | CENTRO DE SAUDE III TABATINGA | Instituto Butantan / Mendelics | Instituto Butantan: Dimas Tadeu Covas, Sandra Coccuzzo Sampaio, Maria Carolina Elias, José Salvatore Leister Patané, Vincent Louis Viala, Antonio Jorge Martins, Ricardo Haddad, Claudia Renata dos Santos Barros, Elaine Cristina Marqueze, Raul Machado Neto, Debora Botequio Moretti, Jardelina de Souza Todao Bernardino, Loyze Paola Oliveira de Lima, Luiz Aurelio de Campos Crispin. Centro de Genômica Funcional da ESALQ: Luiz Lehmann Coutinho, Ricardo Augusto Brassaloti, Raquel de Lello Rocha Campos Cassano. NGS Soluções Genômicas: Pilar Drummond Sampaio Corrêa Mariani. FZEA-USP Pirassununga: Mirele Daiana Poleti, Jessika Cristina Chagas Lesbon, Elisangela Chicaroni Mattos, Heidge Fukumasu. USP-Botucatu: Rejane Maria Tommasini Grotto, Jayme A. Souza-Neto, Guilherme Targino Valente, Patricia Akemi Assato, Felipe Allan da Silva da Costa, Bianca Cechetto Carlos. Mendelics: Bibiana Santos, João Paulo Kitajima, Erika Freitas, David Schlesinger. Hemocentro Ribeirão Preto: Simone Kashima, Evandra Strazza Rodrigues, Svetoslav Nanev Slavov, Elaine Vieira dos Santos, Rafael dos Santos Bezerra, Luiz Carlos Junior de Alcantara, Marta Giovanetti, Vagner Fonseca, Flavia Aburjaile, Rodrigo Tocantins Calado. FAMERP-SJRP: Cecília Artico Banho, Lívia Sacchetto, Fábio Sossai Possebon, Leila Sabrina Ullmann, Cíntia Bittar, Guilherme Campos, Helena Lage Ferreira, Jorge A. Petroli Marchesi, Maisa C. Pereira Parra, Marília Moraes, Paula Rahal, Paulo Inacio da Costa, João Pessoa Araújo Jr., Maurício Lacerda Nogueira. Prefeitura de Sao Paulo: Melissa Palmieri. |
| EPI_ISL_1966640, EPI_ISL_1966641, EPI_ISL_1966642 | HOSPITAL MUNICIPAL SANTO ANTONIO | Instituto Butantan / Mendelics | Instituto Butantan: Dimas Tadeu Covas, Sandra Coccuzzo Sampaio, Maria Carolina Elias, José Salvatore Leister Patané, Vincent Louis Viala, Antonio Jorge Martins, Ricardo Haddad, Claudia Renata dos Santos Barros, Elaine Cristina Marqueze, Raul Machado Neto, Debora Botequio Moretti, Jardelina de Souza Todao Bernardino, Loyze Paola Oliveira de Lima, Luiz Aurelio de Campos Crispin. Centro de Genômica Funcional da ESALQ: Luiz Lehmann Coutinho, Ricardo Augusto Brassaloti, Raquel de Lello Rocha Campos Cassano. NGS Soluções Genômicas: Pilar Drummond Sampaio Corrêa Mariani. FZEA-USP Pirassununga: Mirele Daiana Poleti, Jessika Cristina Chagas Lesbon, Elisangela Chicaroni Mattos, Heidge Fukumasu. USP-Botucatu: Rejane Maria Tommasini Grotto, Jayme A. Souza-Neto, Guilherme Targino Valente, Patricia Akemi Assato, Felipe Allan da Silva da Costa, Bianca Cechetto Carlos. Mendelics: Bibiana Santos, João Paulo Kitajima, Erika Freitas, David Schlesinger. Hemocentro Ribeirão Preto: Simone Kashima, Evandra Strazza Rodrigues, Svetoslav Nanev Slavov, Elaine Vieira dos Santos, Rafael dos Santos Bezerra, Luiz Carlos Junior de Alcantara, Marta Giovanetti, Vagner Fonseca, Flavia Aburjaile, Rodrigo Tocantins Calado. FAMERP-SJRP: Cecília Artico Banho, Lívia Sacchetto, Fábio Sossai Possebon, Leila Sabrina Ullmann, Cíntia Bittar, Guilherme Campos, Helena Lage Ferreira, Jorge A. Petroli Marchesi, Maisa C. Pereira Parra, Marília Moraes, Paula Rahal, Paulo Inacio da Costa, João Pessoa Araújo Jr., Maurício Lacerda Nogueira. Prefeitura de Sao Paulo: Melissa Palmieri. |
| EPI_ISL_1966644, EPI_ISL_1966645 | HOSPITAL MUNICIPAL DR WALDEMAR TEBALDI | Instituto Butantan / Mendelics | Instituto Butantan: Dimas Tadeu Covas, Sandra Coccuzzo Sampaio, Maria Carolina Elias, José Salvatore Leister Patané, Vincent Louis Viala, Antonio Jorge Martins, Ricardo Haddad, Claudia Renata dos Santos Barros, Elaine Cristina Marqueze, Raul Machado Neto, Debora Botequio Moretti, Jardelina de Souza Todao Bernardino, Loyze Paola Oliveira de Lima, Luiz Aurelio de Campos Crispin. Centro de Genômica Funcional da ESALQ: Luiz Lehmann Coutinho, Ricardo Augusto Brassaloti, Raquel de Lello Rocha Campos Cassano. NGS Soluções Genômicas: Pilar Drummond Sampaio Corrêa Mariani. FZEA-USP Pirassununga: Mirele Daiana Poleti, Jessika Cristina Chagas Lesbon, Elisangela Chicaroni Mattos, Heidge Fukumasu. USP-Botucatu: Rejane Maria Tommasini Grotto, Jayme A. Souza-Neto, Guilherme Targino Valente, Patricia Akemi Assato, Felipe Allan da Silva da Costa, Bianca Cechetto Carlos. Mendelics: Bibiana Santos, João Paulo Kitajima, Erika Freitas, David Schlesinger. Hemocentro Ribeirão Preto: Simone Kashima, Evandra Strazza Rodrigues, Svetoslav Nanev Slavov, Elaine Vieira dos Santos, Rafael dos Santos Bezerra, Luiz Carlos Junior de Alcantara, Marta Giovanetti, Vagner |

| | | | |
|---|---|---|---|
| | | | Fonseca, Flavia Aburjaile, Rodrigo Tocantins Calado. FAMERP-SJRP: Cecília Artico Banho, Lívia Sacchetto, Fábio Sossai Possebon, Leila Sabrina Ullmann, Cíntia Bittar, Guilherme Campos, Helena Lage Ferreira, Jorge A. Petroli Marchesi, Maisa C. Pereira Parra, Marília Moraes, Paula Rahal, Paulo Inacio da Costa, João Pessoa Araújo Jr., Maurício Lacerda Nogueira. Prefeitura de Sao Paulo: Melissa Palmieri. |
| EPI_ISL_1966647, EPI_ISL_1966648, EPI_ISL_1966649 | UNIDADE MISTA DE BOM JESUS DOS PERDOES | Instituto Butantan / Mendelics | Instituto Butantan: Dimas Tadeu Covas, Sandra Coccuzzo Sampaio, Maria Carolina Elias, José Salvatore Leister Patané, Vincent Louis Viala, Antonio Jorge Martins, Ricardo Haddad, Claudia Renata dos Santos Barros, Elaine Cristina Marqueze, Raul Machado Neto, Debora Botequio Moretti, Jardelina de Souza Todao Bernardino, Loyze Paola Oliveira de Lima, Luiz Aurelio de Campos Crispin. Centro de Genômica Funcional da ESALQ: Luiz Lehmann Coutinho, Ricardo Augusto Brassaloti, Raquel de Lello Rocha Campos Cassano. NGS Soluções Genômicas: Pilar Drummond Sampaio Corrêa Mariani. FZEA-USP Pirassununga: Mirele Daiana Poleti, Jessika Cristina Chagas Lesbon, Elisangela Chicaroni Mattos, Heidge Fukumasu. USP-Botucatu: Rejane Maria Tommasini Grotto, Jayme A. Souza-Neto, Guilherme Targino Valente, Patricia Akemi Assato, Felipe Allan da Silva da Costa, Bianca Cechetto Carlos. Mendelics: Bibiana Santos, João Paulo Kitajima, Erika Freitas, David Schlesinger. Hemocentro Ribeirão Preto: Simone Kashima, Evandra Strazza Rodrigues, Svetoslav Nanev Slavov, Elaine Vieira dos Santos, Rafael dos Santos Bezerra, Luiz Carlos Junior de Alcantara, Marta Giovanetti, Vagner Fonseca, Flavia Aburjaile, Rodrigo Tocantins Calado. FAMERP-SJRP: Cecília Artico Banho, Lívia Sacchetto, Fábio Sossai Possebon, Leila Sabrina Ullmann, Cíntia Bittar, Guilherme Campos, Helena Lage Ferreira, Jorge A. Petroli Marchesi, Maisa C. Pereira Parra, Marília Moraes, Paula Rahal, Paulo Inacio da Costa, João Pessoa Araújo Jr., Maurício Lacerda Nogueira. Prefeitura de Sao Paulo: Melissa Palmieri. |
| EPI_ISL_1966650, EPI_ISL_1966651, EPI_ISL_1966652, EPI_ISL_1966654, EPI_ISL_1966655, EPI_ISL_1966656, EPI_ISL_1966657, EPI_ISL_1966658, EPI_ISL_1966660, EPI_ISL_1966661, EPI_ISL_1966662, EPI_ISL_1966663, EPI_ISL_1966664, EPI_ISL_1966665, EPI_ISL_1966666, EPI_ISL_1966668 | | | |
| see above | UPA UNIDADE DE PRONTO ATENDIMENTO 24 HORAS BOM JESUS | Instituto Butantan / Mendelics | Instituto Butantan: Dimas Tadeu Covas, Sandra Coccuzzo Sampaio, Maria Carolina Elias, José Salvatore Leister Patané, Vincent Louis Viala, Antonio Jorge Martins, Ricardo Haddad, Claudia Renata dos Santos Barros, Elaine Cristina Marqueze, Raul Machado Neto, Debora Botequio Moretti, Jardelina de Souza Todao Bernardino, Loyze Paola Oliveira de Lima, Luiz Aurelio de Campos Crispin. Centro de Genômica Funcional da ESALQ: Luiz Lehmann Coutinho, Ricardo Augusto Brassaloti, Raquel de Lello Rocha Campos Cassano. NGS Soluções Genômicas: Pilar Drummond Sampaio Corrêa Mariani. FZEA-USP Pirassununga: Mirele Daiana Poleti, Jessika Cristina Chagas Lesbon, Elisangela Chicaroni Mattos, Heidge Fukumasu. USP-Botucatu: Rejane Maria Tommasini Grotto, Jayme A. Souza-Neto, Guilherme Targino Valente, Patricia Akemi Assato, Felipe Allan da Silva da Costa, Bianca Cechetto Carlos. Mendelics: Bibiana Santos, João Paulo Kitajima, Erika Freitas, David Schlesinger. Hemocentro Ribeirão Preto: Simone Kashima, Evandra Strazza Rodrigues, Svetoslav Nanev Slavov, Elaine Vieira dos Santos, Rafael dos Santos Bezerra, Luiz Carlos Junior de Alcantara, Marta Giovanetti, Vagner Fonseca, Flavia Aburjaile, Rodrigo Tocantins Calado. FAMERP-SJRP: Cecília Artico Banho, Lívia Sacchetto, Fábio Sossai Possebon, Leila Sabrina Ullmann, Cíntia Bittar, Guilherme Campos, Helena Lage Ferreira, Jorge A. Petroli Marchesi, Maisa C. Pereira Parra, Marília Moraes, Paula Rahal, Paulo Inacio da Costa, João Pessoa Araújo Jr., Maurício Lacerda Nogueira. Prefeitura de Sao Paulo: Melissa Palmieri. |
| EPI_ISL_1966670, EPI_ISL_1966671, EPI_ISL_1966672, EPI_ISL_1966673 | HOSPITAL MUNICIPAL REYNALDO GUERRA CAJATI | Instituto Butantan / Mendelics | Instituto Butantan: Dimas Tadeu Covas, Sandra Coccuzzo Sampaio, Maria Carolina Elias, José Salvatore Leister Patané, Vincent Louis Viala, Antonio Jorge Martins, Ricardo Haddad, Claudia Renata dos Santos Barros, Elaine Cristina Marqueze, Raul Machado Neto, Debora Botequio Moretti, Jardelina de Souza Todao Bernardino, Loyze Paola Oliveira de Lima, Luiz Aurelio de Campos Crispin. Centro de Genômica Funcional da ESALQ: Luiz Lehmann Coutinho, Ricardo Augusto Brassaloti, Raquel de Lello Rocha Campos Cassano. NGS Soluções Genômicas: Pilar Drummond Sampaio Corrêa Mariani. FZEA-USP Pirassununga: Mirele Daiana Poleti, Jessika Cristina Chagas Lesbon, Elisangela Chicaroni Mattos, Heidge Fukumasu. USP-Botucatu: Rejane Maria Tommasini Grotto, Jayme A. Souza-Neto, Guilherme Targino Valente, Patricia Akemi Assato, Felipe Allan da Silva da Costa, Bianca Cechetto Carlos. Mendelics: Bibiana Santos, João Paulo Kitajima, Erika Freitas, David Schlesinger. Hemocentro Ribeirão Preto: Simone Kashima, Evandra Strazza Rodrigues, Svetoslav Nanev Slavov, Elaine Vieira dos Santos, Rafael dos Santos Bezerra, Luiz Carlos Junior de Alcantara, Marta Giovanetti, Vagner Fonseca, Flavia Aburjaile, Rodrigo Tocantins Calado. FAMERP-SJRP: Cecília Artico Banho, Lívia Sacchetto, Fábio Sossai Possebon, Leila Sabrina Ullmann, Cíntia Bittar, Guilherme Campos, Helena Lage Ferreira, Jorge A. Petroli Marchesi, Maisa C. Pereira Parra, Marília Moraes, Paula Rahal, Paulo Inacio da Costa, João Pessoa Araújo Jr., Maurício Lacerda Nogueira. Prefeitura de Sao Paulo: Melissa Palmieri. |
| EPI_ISL_1966674 | ESF NOVA ALEXANDRIA | Instituto Butantan / Mendelics | Instituto Butantan: Dimas Tadeu Covas, Sandra Coccuzzo Sampaio, Maria Carolina Elias, José Salvatore Leister Patané, Vincent Louis Viala, Antonio Jorge Martins, Ricardo Haddad, Claudia Renata dos Santos Barros, Elaine Cristina Marqueze, Raul Machado Neto, Debora Botequio Moretti, Jardelina de Souza Todao Bernardino, Loyze Paola Oliveira de Lima, Luiz Aurelio de Campos Crispin. Centro de Genômica Funcional da ESALQ: Luiz Lehmann Coutinho, Ricardo Augusto Brassaloti, Raquel de Lello Rocha Campos Cassano. NGS Soluções Genômicas: Pilar Drummond Sampaio Corrêa Mariani. FZEA-USP Pirassununga: Mirele Daiana Poleti, Jessika Cristina Chagas Lesbon, Elisangela Chicaroni Mattos, Heidge Fukumasu. USP-Botucatu: Rejane Maria Tommasini Grotto, Jayme A. Souza-Neto, Guilherme Targino Valente, Patricia Akemi Assato, Felipe Allan da Silva da Costa, Bianca Cechetto Carlos. Mendelics: Bibiana Santos, João Paulo Kitajima, Erika Freitas, David Schlesinger. Hemocentro Ribeirão Preto: Simone Kashima, Evandra Strazza Rodrigues, Svetoslav Nanev Slavov, Elaine Vieira dos Santos, Rafael dos Santos Bezerra, Luiz Carlos Junior de Alcantara, Marta Giovanetti, Vagner Fonseca, Flavia Aburjaile, Rodrigo Tocantins Calado. FAMERP-SJRP: Cecília Artico Banho, Lívia Sacchetto, Fábio Sossai Possebon, Leila Sabrina Ullmann, Cíntia Bittar, Guilherme Campos, Helena Lage Ferreira, Jorge A. Petroli Marchesi, Maisa C. Pereira Parra, Marília Moraes, Paula Rahal, Paulo Inacio da Costa, João Pessoa Araújo Jr., Maurício Lacerda Nogueira. Prefeitura de Sao Paulo: Melissa Palmieri. |
| EPI_ISL_1966675, EPI_ISL_1966676, EPI_ISL_1966677, EPI_ISL_1966679, EPI_ISL_1966680 | LABORATORIO DE FRANCA | Instituto Butantan / Mendelics | Instituto Butantan: Dimas Tadeu Covas, Sandra Coccuzzo Sampaio, Maria Carolina Elias, José Salvatore Leister Patané, Vincent Louis Viala, Antonio Jorge Martins, Ricardo Haddad, Claudia Renata dos Santos Barros, Elaine Cristina Marqueze, Raul Machado Neto, Debora Botequio Moretti, Jardelina de Souza Todao Bernardino, Loyze Paola Oliveira de Lima, Luiz Aurelio de Campos Crispin. Centro de Genômica Funcional da ESALQ: Luiz Lehmann Coutinho, Ricardo Augusto Brassaloti, Raquel de Lello Rocha Campos Cassano. NGS Soluções Genômicas: Pilar Drummond Sampaio Corrêa Mariani. FZEA-USP Pirassununga: Mirele Daiana Poleti, Jessika Cristina Chagas Lesbon, Elisangela Chicaroni Mattos, Heidge Fukumasu. USP-Botucatu: Rejane Maria Tommasini Grotto, Jayme A. Souza-Neto, Guilherme Targino Valente, Patricia Akemi Assato, Felipe Allan da Silva da Costa, Bianca Cechetto Carlos. Mendelics: Bibiana Santos, João Paulo Kitajima, Erika Freitas, David Schlesinger. Hemocentro Ribeirão Preto: Simone Kashima, Evandra Strazza Rodrigues, Svetoslav Nanev Slavov, Elaine Vieira dos Santos, Rafael dos Santos Bezerra, Luiz Carlos Junior de Alcantara, Marta Giovanetti, Vagner Fonseca, Flavia Aburjaile, Rodrigo Tocantins Calado. FAMERP-SJRP: Cecília Artico Banho, Lívia Sacchetto, Fábio Sossai Possebon, Leila Sabrina Ullmann, Cíntia Bittar, Guilherme Campos, Helena Lage Ferreira, Jorge A. Petroli Marchesi, Maisa C. Pereira Parra, Marília Moraes, Paula Rahal, Paulo Inacio da Costa, João Pessoa Araújo Jr., Maurício Lacerda Nogueira. Prefeitura de Sao Paulo: Melissa Palmieri. |
| EPI_ISL_1966681, EPI_ISL_1966682 | UNIDADE MISTA DE IGUAPE | Instituto Butantan / Mendelics | Instituto Butantan: Dimas Tadeu Covas, Sandra Coccuzzo Sampaio, Maria Carolina Elias, José Salvatore Leister Patané, Vincent Louis Viala, Antonio Jorge Martins, Ricardo Haddad, Claudia Renata dos Santos Barros, Elaine Cristina Marqueze, Raul Machado Neto, Debora Botequio Moretti, Jardelina de Souza Todao Bernardino, Loyze Paola Oliveira de Lima, Luiz Aurelio de Campos Crispin. Centro de Genômica Funcional da ESALQ: Luiz Lehmann Coutinho, Ricardo Augusto Brassaloti, Raquel de Lello Rocha Campos Cassano. NGS Soluções Genômicas: Pilar Drummond Sampaio Corrêa Mariani. FZEA-USP Pirassununga: Mirele Daiana Poleti, Jessika Cristina Chagas Lesbon, Elisangela Chicaroni Mattos, Heidge Fukumasu. USP-Botucatu: Rejane Maria Tommasini Grotto, Jayme A. Souza-Neto, Guilherme Targino Valente, Patricia Akemi Assato, Felipe Allan da Silva da Costa, Bianca Cechetto Carlos. Mendelics: Bibiana Santos, João Paulo Kitajima, Erika Freitas, David Schlesinger. Hemocentro Ribeirão Preto: Simone Kashima, Evandra Strazza Rodrigues, Svetoslav Nanev Slavov, Elaine Vieira dos Santos, Rafael dos Santos Bezerra, Luiz Carlos Junior de Alcantara, Marta Giovanetti, Vagner Fonseca, Flavia Aburjaile, Rodrigo Tocantins Calado. FAMERP-SJRP: Cecília Artico Banho, Lívia Sacchetto, Fábio Sossai Possebon, Leila Sabrina Ullmann, Cíntia Bittar, Guilherme Campos, Helena Lage Ferreira, Jorge A. Petroli Marchesi, Maisa C. Pereira Parra, Marília Moraes, Paula Rahal, Paulo Inacio da Costa, João Pessoa Araújo Jr., Maurício Lacerda Nogueira. Prefeitura de Sao Paulo: Melissa Palmieri. |
| EPI_ISL_1966683, EPI_ISL_1966684, EPI_ISL_1966685, EPI_ISL_1966686, EPI_ISL_1966687, EPI_ISL_1966688, EPI_ISL_1966690 | LABORATORIO MUNICIPAL DE ANALISES CLINICAS DE ITANHAEM | Instituto Butantan / Mendelics | Instituto Butantan: Dimas Tadeu Covas, Sandra Coccuzzo Sampaio, Maria Carolina Elias, José Salvatore Leister Patané, Vincent Louis Viala, Antonio Jorge Martins, Ricardo Haddad, Claudia Renata dos Santos Barros, Elaine Cristina Marqueze, Raul Machado Neto, Debora Botequio Moretti, Jardelina de Souza Todao Bernardino, Loyze Paola Oliveira de Lima, Luiz Aurelio de Campos Crispin. Centro de Genômica Funcional da ESALQ: Luiz Lehmann Coutinho, Ricardo Augusto Brassaloti, Raquel de Lello Rocha Campos Cassano. NGS Soluções Genômicas: Pilar Drummond Sampaio Corrêa Mariani. FZEA-USP Pirassununga: Mirele Daiana Poleti, Jessika Cristina Chagas Lesbon, Elisangela Chicaroni Mattos, Heidge Fukumasu. USP-Botucatu: Rejane Maria Tommasini Grotto, Jayme A. Souza-Neto, Guilherme Targino Valente, Patricia Akemi Assato, Felipe Allan da Silva da Costa, Bianca Cechetto Carlos. Mendelics: Bibiana Santos, João Paulo Kitajima, Erika Freitas, David Schlesinger. Hemocentro Ribeirão Preto: Simone Kashima, Evandra Strazza Rodrigues, Svetoslav Nanev Slavov, Elaine Vieira dos Santos, Rafael dos Santos Bezerra, Luiz Carlos Junior de Alcantara, Marta Giovanetti, Vagner Fonseca, Flavia Aburjaile, Rodrigo Tocantins Calado. FAMERP-SJRP: Cecília Artico Banho, Lívia Sacchetto, Fábio Sossai Possebon, Leila Sabrina Ullmann, Cíntia Bittar, Guilherme Campos, Helena Lage Ferreira, Jorge A. Petroli Marchesi, Maisa C. Pereira Parra, Marília Moraes, Paula Rahal, Paulo Inacio da Costa, João Pessoa Araújo Jr., Maurício Lacerda Nogueira. Prefeitura de Sao Paulo: Melissa Palmieri. |
| EPI_ISL_1966692 | PRONTO ATENDIMENTO MUNICIPAL DE JACUPIRANGA | Instituto Butantan / Mendelics | Instituto Butantan: Dimas Tadeu Covas, Sandra Coccuzzo Sampaio, Maria Carolina Elias, José Salvatore Leister Patané, Vincent Louis Viala, Antonio Jorge Martins, Ricardo Haddad, Claudia Renata dos Santos Barros, Elaine Cristina Marqueze, Raul Machado Neto, Debora Botequio Moretti, Jardelina de |

| | | | |
|---|---|---|---|
| | | | Souza Todao Bernardino, Loyze Paola Oliveira de Lima, Luiz Aurelio de Campos Crispin. Centro de Genômica Funcional da ESALQ: Luiz Lehmann Coutinho, Ricardo Augusto Brassaloti, Raquel de Lello Rocha Campos Cassano. NGS Soluções Genômicas: Pilar Drummond Sampaio Corrêa Mariani. FZEA-USP Pirassununga: Mirele Daiana Poleti, Jessika Cristina Chagas Lesbon, Elisangela Chicaroni Mattos, Heidge Fukumasu. USP-Botucatu: Rejane Maria Tommasini Grotto, Jayme A. Souza-Neto, Guilherme Targino Valente, Patricia Akemi Assato, Felipe Allan da Silva da Costa, Bianca Cechetto Carlos. Mendelics: Bibiana Santos, João Paulo Kitajima, Erika Freitas, David Schlesinger. Hemocentro Ribeirão Preto: Simone Kashima, Evandra Strazza Rodrigues, Svetoslav Nanev Slavov, Elaine Vieira dos Santos, Rafael dos Santos Bezerra, Luiz Carlos Junior de Alcantara, Marta Giovanetti, Vagner Fonseca, Flavia Aburjaile, Rodrigo Tocantins Calado. FAMERP-SJRP: Cecília Artico Banho, Lívia Sacchetto, Fábio Sossai Possebon, Leila Sabrina Ullmann, Cíntia Bittar, Guilherme Campos, Helena Lage Ferreira, Jorge A. Petroli Marchesi, Maisa C. Pereira Parra, Marília Moraes, Paula Rahal, Paulo Inacio da Costa, João Pessoa Araújo Jr., Maurício Lacerda Nogueira. Prefeitura de Sao Paulo: Melissa Palmieri. |
| EPI_ISL_1966693 | SECRETARIA MUNICIPAL DE SAUDE DE JARINU | Instituto Butantan / Mendelics | Instituto Butantan: Dimas Tadeu Covas, Sandra Coccuzzo Sampaio, Maria Carolina Elias, José Salvatore Leister Patané, Vincent Louis Viala, Antonio Jorge Martins, Ricardo Haddad, Claudia Renata dos Santos Barros, Elaine Cristina Marqueze, Raul Machado Neto, Debora Botequio Moretti, Jardelina de Souza Todao Bernardino, Loyze Paola Oliveira de Lima, Luiz Aurelio de Campos Crispin. Centro de Genômica Funcional da ESALQ: Luiz Lehmann Coutinho, Ricardo Augusto Brassaloti, Raquel de Lello Rocha Campos Cassano. NGS Soluções Genômicas: Pilar Drummond Sampaio Corrêa Mariani. FZEA-USP Pirassununga: Mirele Daiana Poleti, Jessika Cristina Chagas Lesbon, Elisangela Chicaroni Mattos, Heidge Fukumasu. USP-Botucatu: Rejane Maria Tommasini Grotto, Jayme A. Souza-Neto, Guilherme Targino Valente, Patricia Akemi Assato, Felipe Allan da Silva da Costa, Bianca Cechetto Carlos. Mendelics: Bibiana Santos, João Paulo Kitajima, Erika Freitas, David Schlesinger. Hemocentro Ribeirão Preto: Simone Kashima, Evandra Strazza Rodrigues, Svetoslav Nanev Slavov, Elaine Vieira dos Santos, Rafael dos Santos Bezerra, Luiz Carlos Junior de Alcantara, Marta Giovanetti, Vagner Fonseca, Flavia Aburjaile, Rodrigo Tocantins Calado. FAMERP-SJRP: Cecília Artico Banho, Lívia Sacchetto, Fábio Sossai Possebon, Leila Sabrina Ullmann, Cíntia Bittar, Guilherme Campos, Helena Lage Ferreira, Jorge A. Petroli Marchesi, Maisa C. Pereira Parra, Marília Moraes, Paula Rahal, Paulo Inacio da Costa, João Pessoa Araújo Jr., Maurício Lacerda Nogueira. Prefeitura de Sao Paulo: Melissa Palmieri. |
| EPI_ISL_1966694 | HOSPITAL SANTO ANTONIO DE JUQUIA JUQUIA | Instituto Butantan / Mendelics | Instituto Butantan: Dimas Tadeu Covas, Sandra Coccuzzo Sampaio, Maria Carolina Elias, José Salvatore Leister Patané, Vincent Louis Viala, Antonio Jorge Martins, Ricardo Haddad, Claudia Renata dos Santos Barros, Elaine Cristina Marqueze, Raul Machado Neto, Debora Botequio Moretti, Jardelina de Souza Todao Bernardino, Loyze Paola Oliveira de Lima, Luiz Aurelio de Campos Crispin. Centro de Genômica Funcional da ESALQ: Luiz Lehmann Coutinho, Ricardo Augusto Brassaloti, Raquel de Lello Rocha Campos Cassano. NGS Soluções Genômicas: Pilar Drummond Sampaio Corrêa Mariani. FZEA-USP Pirassununga: Mirele Daiana Poleti, Jessika Cristina Chagas Lesbon, Elisangela Chicaroni Mattos, Heidge Fukumasu. USP-Botucatu: Rejane Maria Tommasini Grotto, Jayme A. Souza-Neto, Guilherme Targino Valente, Patricia Akemi Assato, Felipe Allan da Silva da Costa, Bianca Cechetto Carlos. Mendelics: Bibiana Santos, João Paulo Kitajima, Erika Freitas, David Schlesinger. Hemocentro Ribeirão Preto: Simone Kashima, Evandra Strazza Rodrigues, Svetoslav Nanev Slavov, Elaine Vieira dos Santos, Rafael dos Santos Bezerra, Luiz Carlos Junior de Alcantara, Marta Giovanetti, Vagner Fonseca, Flavia Aburjaile, Rodrigo Tocantins Calado. FAMERP-SJRP: Cecília Artico Banho, Lívia Sacchetto, Fábio Sossai Possebon, Leila Sabrina Ullmann, Cíntia Bittar, Guilherme Campos, Helena Lage Ferreira, Jorge A. Petroli Marchesi, Maisa C. Pereira Parra, Marília Moraes, Paula Rahal, Paulo Inacio da Costa, João Pessoa Araújo Jr., Maurício Lacerda Nogueira. Prefeitura de Sao Paulo: Melissa Palmieri. |
| EPI_ISL_1966696 | UBS III DE PARIQUERA ACU PARIQUERA ACU | Instituto Butantan / Mendelics | Instituto Butantan: Dimas Tadeu Covas, Sandra Coccuzzo Sampaio, Maria Carolina Elias, José Salvatore Leister Patané, Vincent Louis Viala, Antonio Jorge Martins, Ricardo Haddad, Claudia Renata dos Santos Barros, Elaine Cristina Marqueze, Raul Machado Neto, Debora Botequio Moretti, Jardelina de Souza Todao Bernardino, Loyze Paola Oliveira de Lima, Luiz Aurelio de Campos Crispin. Centro de Genômica Funcional da ESALQ: Luiz Lehmann Coutinho, Ricardo Augusto Brassaloti, Raquel de Lello Rocha Campos Cassano. NGS Soluções Genômicas: Pilar Drummond Sampaio Corrêa Mariani. FZEA-USP Pirassununga: Mirele Daiana Poleti, Jessika Cristina Chagas Lesbon, Elisangela Chicaroni Mattos, Heidge Fukumasu. USP-Botucatu: Rejane Maria Tommasini Grotto, Jayme A. Souza-Neto, Guilherme Targino Valente, Patricia Akemi Assato, Felipe Allan da Silva da Costa, Bianca Cechetto Carlos. Mendelics: Bibiana Santos, João Paulo Kitajima, Erika Freitas, David Schlesinger. Hemocentro Ribeirão Preto: Simone Kashima, Evandra Strazza Rodrigues, Svetoslav Nanev Slavov, Elaine Vieira dos Santos, Rafael dos Santos Bezerra, Luiz Carlos Junior de Alcantara, Marta Giovanetti, Vagner Fonseca, Flavia Aburjaile, Rodrigo Tocantins Calado. FAMERP-SJRP: Cecília Artico Banho, Lívia Sacchetto, Fábio Sossai Possebon, Leila Sabrina Ullmann, Cíntia Bittar, Guilherme Campos, Helena Lage Ferreira, Jorge A. Petroli Marchesi, Maisa C. Pereira Parra, Marília Moraes, Paula Rahal, Paulo Inacio da Costa, João Pessoa Araújo Jr., Maurício Lacerda Nogueira. Prefeitura de Sao Paulo: Melissa Palmieri. |
| EPI_ISL_1966697, EPI_ISL_1966698, EPI_ISL_1966700, EPI_ISL_1966701, EPI_ISL_1966702, EPI_ISL_1966703 | AMBULATORIO MEDICO DE ESPECIALIDADES DE PERUIBE | Instituto Butantan / Mendelics | Instituto Butantan: Dimas Tadeu Covas, Sandra Coccuzzo Sampaio, Maria Carolina Elias, José Salvatore Leister Patané, Vincent Louis Viala, Antonio Jorge Martins, Ricardo Haddad, Claudia Renata dos Santos Barros, Elaine Cristina Marqueze, Raul Machado Neto, Debora Botequio Moretti, Jardelina de Souza Todao Bernardino, Loyze Paola Oliveira de Lima, Luiz Aurelio de Campos Crispin. Centro de Genômica Funcional da ESALQ: Luiz Lehmann Coutinho, Ricardo Augusto Brassaloti, Raquel de Lello Rocha Campos Cassano. NGS Soluções Genômicas: Pilar Drummond Sampaio Corrêa Mariani. FZEA-USP Pirassununga: Mirele Daiana Poleti, Jessika Cristina Chagas Lesbon, Elisangela Chicaroni Mattos, Heidge Fukumasu. USP-Botucatu: Rejane Maria Tommasini Grotto, Jayme A. Souza-Neto, Guilherme Targino Valente, Patricia Akemi Assato, Felipe Allan da Silva da Costa, Bianca Cechetto Carlos. Mendelics: Bibiana Santos, João Paulo Kitajima, Erika Freitas, David Schlesinger. Hemocentro Ribeirão Preto: Simone Kashima, Evandra Strazza Rodrigues, Svetoslav Nanev Slavov, Elaine Vieira dos Santos, Rafael dos Santos Bezerra, Luiz Carlos Junior de Alcantara, Marta Giovanetti, Vagner Fonseca, Flavia Aburjaile, Rodrigo Tocantins Calado. FAMERP-SJRP: Cecília Artico Banho, Lívia Sacchetto, Fábio Sossai Possebon, Leila Sabrina Ullmann, Cíntia Bittar, Guilherme Campos, Helena Lage Ferreira, Jorge A. Petroli Marchesi, Maisa C. Pereira Parra, Marília Moraes, Paula Rahal, Paulo Inacio da Costa, João Pessoa Araújo Jr., Maurício Lacerda Nogueira. Prefeitura de Sao Paulo: Melissa Palmieri. |
| EPI_ISL_1966704 | USAFA REAL | Instituto Butantan / Mendelics | Instituto Butantan: Dimas Tadeu Covas, Sandra Coccuzzo Sampaio, Maria Carolina Elias, José Salvatore Leister Patané, Vincent Louis Viala, Antonio Jorge Martins, Ricardo Haddad, Claudia Renata dos Santos Barros, Elaine Cristina Marqueze, Raul Machado Neto, Debora Botequio Moretti, Jardelina de Souza Todao Bernardino, Loyze Paola Oliveira de Lima, Luiz Aurelio de Campos Crispin. Centro de Genômica Funcional da ESALQ: Luiz Lehmann Coutinho, Ricardo Augusto Brassaloti, Raquel de Lello Rocha Campos Cassano. NGS Soluções Genômicas: Pilar Drummond Sampaio Corrêa Mariani. FZEA-USP Pirassununga: Mirele Daiana Poleti, Jessika Cristina Chagas Lesbon, Elisangela Chicaroni Mattos, Heidge Fukumasu. USP-Botucatu: Rejane Maria Tommasini Grotto, Jayme A. Souza-Neto, Guilherme Targino Valente, Patricia Akemi Assato, Felipe Allan da Silva da Costa, Bianca Cechetto Carlos. Mendelics: Bibiana Santos, João Paulo Kitajima, Erika Freitas, David Schlesinger. Hemocentro Ribeirão Preto: Simone Kashima, Evandra Strazza Rodrigues, Svetoslav Nanev Slavov, Elaine Vieira dos Santos, Rafael dos Santos Bezerra, Luiz Carlos Junior de Alcantara, Marta Giovanetti, Vagner Fonseca, Flavia Aburjaile, Rodrigo Tocantins Calado. FAMERP-SJRP: Cecília Artico Banho, Lívia Sacchetto, Fábio Sossai Possebon, Leila Sabrina Ullmann, Cíntia Bittar, Guilherme Campos, Helena Lage Ferreira, Jorge A. Petroli Marchesi, Maisa C. Pereira Parra, Marília Moraes, Paula Rahal, Paulo Inacio da Costa, João Pessoa Araújo Jr., Maurício Lacerda Nogueira. Prefeitura de Sao Paulo: Melissa Palmieri. |
| EPI_ISL_1966705 | USAFA ALOHA | Instituto Butantan / Mendelics | Instituto Butantan: Dimas Tadeu Covas, Sandra Coccuzzo Sampaio, Maria Carolina Elias, José Salvatore Leister Patané, Vincent Louis Viala, Antonio Jorge Martins, Ricardo Haddad, Claudia Renata dos Santos Barros, Elaine Cristina Marqueze, Raul Machado Neto, Debora Botequio Moretti, Jardelina de Souza Todao Bernardino, Loyze Paola Oliveira de Lima, Luiz Aurelio de Campos Crispin. Centro de Genômica Funcional da ESALQ: Luiz Lehmann Coutinho, Ricardo Augusto Brassaloti, Raquel de Lello Rocha Campos Cassano. NGS Soluções Genômicas: Pilar Drummond Sampaio Corrêa Mariani. FZEA-USP Pirassununga: Mirele Daiana Poleti, Jessika Cristina Chagas Lesbon, Elisangela Chicaroni Mattos, Heidge Fukumasu. USP-Botucatu: Rejane Maria Tommasini Grotto, Jayme A. Souza-Neto, Guilherme Targino Valente, Patricia Akemi Assato, Felipe Allan da Silva da Costa, Bianca Cechetto Carlos. Mendelics: Bibiana Santos, João Paulo Kitajima, Erika Freitas, David Schlesinger. Hemocentro Ribeirão Preto: Simone Kashima, Evandra Strazza Rodrigues, Svetoslav Nanev Slavov, Elaine Vieira dos Santos, Rafael dos Santos Bezerra, Luiz Carlos Junior de Alcantara, Marta Giovanetti, Vagner Fonseca, Flavia Aburjaile, Rodrigo Tocantins Calado. FAMERP-SJRP: Cecília Artico Banho, Lívia Sacchetto, Fábio Sossai Possebon, Leila Sabrina Ullmann, Cíntia Bittar, Guilherme Campos, Helena Lage Ferreira, Jorge A. Petroli Marchesi, Maisa C. Pereira Parra, Marília Moraes, Paula Rahal, Paulo Inacio da Costa, João Pessoa Araújo Jr., Maurício Lacerda Nogueira. Prefeitura de Sao Paulo: Melissa Palmieri. |
| EPI_ISL_1966706 | AMBULATORIO TUPI | Instituto Butantan / Mendelics | Instituto Butantan: Dimas Tadeu Covas, Sandra Coccuzzo Sampaio, Maria Carolina Elias, José Salvatore Leister Patané, Vincent Louis Viala, Antonio Jorge Martins, Ricardo Haddad, Claudia Renata dos Santos Barros, Elaine Cristina Marqueze, Raul Machado Neto, Debora Botequio Moretti, Jardelina de Souza Todao Bernardino, Loyze Paola Oliveira de Lima, Luiz Aurelio de Campos Crispin. Centro de Genômica Funcional da ESALQ: Luiz Lehmann Coutinho, Ricardo Augusto Brassaloti, Raquel de Lello Rocha Campos Cassano. NGS Soluções Genômicas: Pilar Drummond Sampaio Corrêa Mariani. FZEA-USP Pirassununga: Mirele Daiana Poleti, Jessika Cristina Chagas Lesbon, Elisangela Chicaroni Mattos, Heidge Fukumasu. USP-Botucatu: Rejane Maria Tommasini Grotto, Jayme A. Souza-Neto, Guilherme Targino Valente, Patricia Akemi Assato, Felipe Allan da Silva da Costa, Bianca Cechetto Carlos. Mendelics: Bibiana Santos, João Paulo Kitajima, Erika Freitas, David Schlesinger. Hemocentro Ribeirão Preto: Simone Kashima, Evandra Strazza Rodrigues, Svetoslav Nanev Slavov, Elaine Vieira dos Santos, Rafael dos Santos Bezerra, Luiz Carlos Junior de Alcantara, Marta Giovanetti, Vagner Fonseca, Flavia Aburjaile, Rodrigo Tocantins Calado. FAMERP-SJRP: Cecília Artico Banho, Lívia Sacchetto, Fábio Sossai Possebon, Leila Sabrina |

| | | | |
|---|---|---|---|
| | | | Ullmann, Cíntia Bittar, Guilherme Campos, Helena Lage Ferreira, Jorge A. Petroli Marchesi, Maisa C. Pereira Parra, Marília Moraes, Paula Rahal, Paulo Inacio da Costa, João Pessoa Araújo Jr., Maurício Lacerda Nogueira. Prefeitura de Sao Paulo: Melissa Palmieri. |
| EPI_ISL_1966707 | USAFA GUARAMAR | Instituto Butantan / Mendelics | Instituto Butantan: Dimas Tadeu Covas, Sandra Coccuzzo Sampaio, Maria Carolina Elias, José Salvatore Leister Patané, Vincent Louis Viala, Antonio Jorge Martins, Ricardo Haddad, Claudia Renata dos Santos Barros, Elaine Cristina Marqueze, Raul Machado Neto, Debora Botequio Moretti, Jardelina de Souza Todao Bernardino, Loyze Paola Oliveira de Lima, Luiz Aurelio de Campos Crispin. Centro de Genômica Funcional da ESALQ: Luiz Lehmann Coutinho, Ricardo Augusto Brassaloti, Raquel de Lello Rocha Campos Cassano. NGS Soluções Genômicas: Pilar Drummond Sampaio Corrêa Mariani. FZEA-USP Pirassununga: Mirele Daiana Poleti, Jessika Cristina Chagas Lesbon, Elisangela Chicaroni Mattos, Heidge Fukumasu. USP-Botucatu: Rejane Maria Tommasini Grotto, Jayme A. Souza-Neto, Guilherme Targino Valente, Patricia Akemi Assato, Felipe Allan da Silva da Costa, Bianca Cechetto Carlos. Mendelics: Bibiana Santos, João Paulo Kitajima, Erika Freitas, David Schlesinger. Hemocentro Ribeirão Preto: Simone Kashima, Evandra Strazza Rodrigues, Svetoslav Nanev Slavov, Elaine Vieira dos Santos, Rafael dos Santos Bezerra, Luiz Carlos Junior de Alcantara, Marta Giovanetti, Vagner Fonseca, Flavia Aburjaile, Rodrigo Tocantins Calado. FAMERP-SJRP: Cecília Artico Banho, Lívia Sacchetto, Fábio Sossai Possebon, Leila Sabrina Ullmann, Cíntia Bittar, Guilherme Campos, Helena Lage Ferreira, Jorge A. Petroli Marchesi, Maisa C. Pereira Parra, Marília Moraes, Paula Rahal, Paulo Inacio da Costa, João Pessoa Araújo Jr., Maurício Lacerda Nogueira. Prefeitura de Sao Paulo: Melissa Palmieri. |
| EPI_ISL_1966708 | UNIDADE DE PRONTO ATENDIMENTO UPA | Instituto Butantan / Mendelics | Instituto Butantan: Dimas Tadeu Covas, Sandra Coccuzzo Sampaio, Maria Carolina Elias, José Salvatore Leister Patané, Vincent Louis Viala, Antonio Jorge Martins, Ricardo Haddad, Claudia Renata dos Santos Barros, Elaine Cristina Marqueze, Raul Machado Neto, Debora Botequio Moretti, Jardelina de Souza Todao Bernardino, Loyze Paola Oliveira de Lima, Luiz Aurelio de Campos Crispin. Centro de Genômica Funcional da ESALQ: Luiz Lehmann Coutinho, Ricardo Augusto Brassaloti, Raquel de Lello Rocha Campos Cassano. NGS Soluções Genômicas: Pilar Drummond Sampaio Corrêa Mariani. FZEA-USP Pirassununga: Mirele Daiana Poleti, Jessika Cristina Chagas Lesbon, Elisangela Chicaroni Mattos, Heidge Fukumasu. USP-Botucatu: Rejane Maria Tommasini Grotto, Jayme A. Souza-Neto, Guilherme Targino Valente, Patricia Akemi Assato, Felipe Allan da Silva da Costa, Bianca Cechetto Carlos. Mendelics: Bibiana Santos, João Paulo Kitajima, Erika Freitas, David Schlesinger. Hemocentro Ribeirão Preto: Simone Kashima, Evandra Strazza Rodrigues, Svetoslav Nanev Slavov, Elaine Vieira dos Santos, Rafael dos Santos Bezerra, Luiz Carlos Junior de Alcantara, Marta Giovanetti, Vagner Fonseca, Flavia Aburjaile, Rodrigo Tocantins Calado. FAMERP-SJRP: Cecília Artico Banho, Lívia Sacchetto, Fábio Sossai Possebon, Leila Sabrina Ullmann, Cíntia Bittar, Guilherme Campos, Helena Lage Ferreira, Jorge A. Petroli Marchesi, Maisa C. Pereira Parra, Marília Moraes, Paula Rahal, Paulo Inacio da Costa, João Pessoa Araújo Jr., Maurício Lacerda Nogueira. Prefeitura de Sao Paulo: Melissa Palmieri. |
| EPI_ISL_1966710, EPI_ISL_1966711, EPI_ISL_1966712, EPI_ISL_1966713 | SECRETARIA MUNICIPAL DE SAUDE DE SANTA BARBARA D OESTE | Instituto Butantan / Mendelics | Instituto Butantan: Dimas Tadeu Covas, Sandra Coccuzzo Sampaio, Maria Carolina Elias, José Salvatore Leister Patané, Vincent Louis Viala, Antonio Jorge Martins, Ricardo Haddad, Claudia Renata dos Santos Barros, Elaine Cristina Marqueze, Raul Machado Neto, Debora Botequio Moretti, Jardelina de Souza Todao Bernardino, Loyze Paola Oliveira de Lima, Luiz Aurelio de Campos Crispin. Centro de Genômica Funcional da ESALQ: Luiz Lehmann Coutinho, Ricardo Augusto Brassaloti, Raquel de Lello Rocha Campos Cassano. NGS Soluções Genômicas: Pilar Drummond Sampaio Corrêa Mariani. FZEA-USP Pirassununga: Mirele Daiana Poleti, Jessika Cristina Chagas Lesbon, Elisangela Chicaroni Mattos, Heidge Fukumasu. USP-Botucatu: Rejane Maria Tommasini Grotto, Jayme A. Souza-Neto, Guilherme Targino Valente, Patricia Akemi Assato, Felipe Allan da Silva da Costa, Bianca Cechetto Carlos. Mendelics: Bibiana Santos, João Paulo Kitajima, Erika Freitas, David Schlesinger. Hemocentro Ribeirão Preto: Simone Kashima, Evandra Strazza Rodrigues, Svetoslav Nanev Slavov, Elaine Vieira dos Santos, Rafael dos Santos Bezerra, Luiz Carlos Junior de Alcantara, Marta Giovanetti, Vagner Fonseca, Flavia Aburjaile, Rodrigo Tocantins Calado. FAMERP-SJRP: Cecília Artico Banho, Lívia Sacchetto, Fábio Sossai Possebon, Leila Sabrina Ullmann, Cíntia Bittar, Guilherme Campos, Helena Lage Ferreira, Jorge A. Petroli Marchesi, Maisa C. Pereira Parra, Marília Moraes, Paula Rahal, Paulo Inacio da Costa, João Pessoa Araújo Jr., Maurício Lacerda Nogueira. Prefeitura de Sao Paulo: Melissa Palmieri. |
| EPI_ISL_1966716, EPI_ISL_1966717, EPI_ISL_1966718, EPI_ISL_1966719 | SECAO CENTRO DE DIAGNOSTICO SECEDI | Instituto Butantan / Mendelics | Instituto Butantan: Dimas Tadeu Covas, Sandra Coccuzzo Sampaio, Maria Carolina Elias, José Salvatore Leister Patané, Vincent Louis Viala, Antonio Jorge Martins, Ricardo Haddad, Claudia Renata dos Santos Barros, Elaine Cristina Marqueze, Raul Machado Neto, Debora Botequio Moretti, Jardelina de Souza Todao Bernardino, Loyze Paola Oliveira de Lima, Luiz Aurelio de Campos Crispin. Centro de Genômica Funcional da ESALQ: Luiz Lehmann Coutinho, Ricardo Augusto Brassaloti, Raquel de Lello Rocha Campos Cassano. NGS Soluções Genômicas: Pilar Drummond Sampaio Corrêa Mariani. FZEA-USP Pirassununga: Mirele Daiana Poleti, Jessika Cristina Chagas Lesbon, Elisangela Chicaroni Mattos, Heidge Fukumasu. USP-Botucatu: Rejane Maria Tommasini Grotto, Jayme A. Souza-Neto, Guilherme Targino Valente, Patricia Akemi Assato, Felipe Allan da Silva da Costa, Bianca Cechetto Carlos. Mendelics: Bibiana Santos, João Paulo Kitajima, Erika Freitas, David Schlesinger. Hemocentro Ribeirão Preto: Simone Kashima, Evandra Strazza Rodrigues, Svetoslav Nanev Slavov, Elaine Vieira dos Santos, Rafael dos Santos Bezerra, Luiz Carlos Junior de Alcantara, Marta Giovanetti, Vagner Fonseca, Flavia Aburjaile, Rodrigo Tocantins Calado. FAMERP-SJRP: Cecília Artico Banho, Lívia Sacchetto, Fábio Sossai Possebon, Leila Sabrina Ullmann, Cíntia Bittar, Guilherme Campos, Helena Lage Ferreira, Jorge A. Petroli Marchesi, Maisa C. Pereira Parra, Marília Moraes, Paula Rahal, Paulo Inacio da Costa, João Pessoa Araújo Jr., Maurício Lacerda Nogueira. Prefeitura de Sao Paulo: Melissa Palmieri. |
| EPI_ISL_1966720 | UNIDADE DE VIGILANCIA EM SAUDE | Instituto Butantan / Mendelics | Instituto Butantan: Dimas Tadeu Covas, Sandra Coccuzzo Sampaio, Maria Carolina Elias, José Salvatore Leister Patané, Vincent Louis Viala, Antonio Jorge Martins, Ricardo Haddad, Claudia Renata dos Santos Barros, Elaine Cristina Marqueze, Raul Machado Neto, Debora Botequio Moretti, Jardelina de Souza Todao Bernardino, Loyze Paola Oliveira de Lima, Luiz Aurelio de Campos Crispin. Centro de Genômica Funcional da ESALQ: Luiz Lehmann Coutinho, Ricardo Augusto Brassaloti, Raquel de Lello Rocha Campos Cassano. NGS Soluções Genômicas: Pilar Drummond Sampaio Corrêa Mariani. FZEA-USP Pirassununga: Mirele Daiana Poleti, Jessika Cristina Chagas Lesbon, Elisangela Chicaroni Mattos, Heidge Fukumasu. USP-Botucatu: Rejane Maria Tommasini Grotto, Jayme A. Souza-Neto, Guilherme Targino Valente, Patricia Akemi Assato, Felipe Allan da Silva da Costa, Bianca Cechetto Carlos. Mendelics: Bibiana Santos, João Paulo Kitajima, Erika Freitas, David Schlesinger. Hemocentro Ribeirão Preto: Simone Kashima, Evandra Strazza Rodrigues, Svetoslav Nanev Slavov, Elaine Vieira dos Santos, Rafael dos Santos Bezerra, Luiz Carlos Junior de Alcantara, Marta Giovanetti, Vagner Fonseca, Flavia Aburjaile, Rodrigo Tocantins Calado. FAMERP-SJRP: Cecília Artico Banho, Lívia Sacchetto, Fábio Sossai Possebon, Leila Sabrina Ullmann, Cíntia Bittar, Guilherme Campos, Helena Lage Ferreira, Jorge A. Petroli Marchesi, Maisa C. Pereira Parra, Marília Moraes, Paula Rahal, Paulo Inacio da Costa, João Pessoa Araújo Jr., Maurício Lacerda Nogueira. Prefeitura de Sao Paulo: Melissa Palmieri. |
| EPI_ISL_1966721, EPI_ISL_1966722, EPI_ISL_1966723 | UNIDADE DE PRONTO ATENDIMENTO DE VARZEA PAULISTA UPA II | Instituto Butantan / Mendelics | Instituto Butantan: Dimas Tadeu Covas, Sandra Coccuzzo Sampaio, Maria Carolina Elias, José Salvatore Leister Patané, Vincent Louis Viala, Antonio Jorge Martins, Ricardo Haddad, Claudia Renata dos Santos Barros, Elaine Cristina Marqueze, Raul Machado Neto, Debora Botequio Moretti, Jardelina de Souza Todao Bernardino, Loyze Paola Oliveira de Lima, Luiz Aurelio de Campos Crispin. Centro de Genômica Funcional da ESALQ: Luiz Lehmann Coutinho, Ricardo Augusto Brassaloti, Raquel de Lello Rocha Campos Cassano. NGS Soluções Genômicas: Pilar Drummond Sampaio Corrêa Mariani. FZEA-USP Pirassununga: Mirele Daiana Poleti, Jessika Cristina Chagas Lesbon, Elisangela Chicaroni Mattos, Heidge Fukumasu. USP-Botucatu: Rejane Maria Tommasini Grotto, Jayme A. Souza-Neto, Guilherme Targino Valente, Patricia Akemi Assato, Felipe Allan da Silva da Costa, Bianca Cechetto Carlos. Mendelics: Bibiana Santos, João Paulo Kitajima, Erika Freitas, David Schlesinger. Hemocentro Ribeirão Preto: Simone Kashima, Evandra Strazza Rodrigues, Svetoslav Nanev Slavov, Elaine Vieira dos Santos, Rafael dos Santos Bezerra, Luiz Carlos Junior de Alcantara, Marta Giovanetti, Vagner Fonseca, Flavia Aburjaile, Rodrigo Tocantins Calado. FAMERP-SJRP: Cecília Artico Banho, Lívia Sacchetto, Fábio Sossai Possebon, Leila Sabrina Ullmann, Cíntia Bittar, Guilherme Campos, Helena Lage Ferreira, Jorge A. Petroli Marchesi, Maisa C. Pereira Parra, Marília Moraes, Paula Rahal, Paulo Inacio da Costa, João Pessoa Araújo Jr., Maurício Lacerda Nogueira. Prefeitura de Sao Paulo: Melissa Palmieri. |
| EPI_ISL_1966724, EPI_ISL_1966725, EPI_ISL_1966726 | GRUPO TECNICO DE VIGILANCIA SANITARIA E EPIDEMIOLOGICA | Instituto Butantan | Instituto Butantan: Dimas Tadeu Covas, Sandra Coccuzzo Sampaio, Maria Carolina Elias, José Salvatore Leister Patané, Vincent Louis Viala, Antonio Jorge Martins, Ricardo Haddad, Claudia Renata dos Santos Barros, Elaine Cristina Marqueze, Raul Machado Neto, Debora Botequio Moretti, Jardelina de Souza Todao Bernardino, Loyze Paola Oliveira de Lima, Luiz Aurelio de Campos Crispin. Centro de Genômica Funcional da ESALQ: Luiz Lehmann Coutinho, Ricardo Augusto Brassaloti, Raquel de Lello Rocha Campos Cassano. NGS Soluções Genômicas: Pilar Drummond Sampaio Corrêa Mariani. FZEA-USP Pirassununga: Mirele Daiana Poleti, Jessika Cristina Chagas Lesbon, Elisangela Chicaroni Mattos, Heidge Fukumasu. USP-Botucatu: Rejane Maria Tommasini Grotto, Jayme A. Souza-Neto, Guilherme Targino Valente, Patricia Akemi Assato, Felipe Allan da Silva da Costa, Bianca Cechetto Carlos. Mendelics: Bibiana Santos, João Paulo Kitajima, Erika Freitas, David Schlesinger. Hemocentro Ribeirão Preto: Simone Kashima, Evandra Strazza Rodrigues, Svetoslav Nanev Slavov, Elaine Vieira dos Santos, Rafael dos Santos Bezerra, Luiz Carlos Junior de Alcantara, Marta Giovanetti, Vagner Fonseca, Flavia Aburjaile, Rodrigo Tocantins Calado. FAMERP-SJRP: Cecília Artico Banho, Lívia Sacchetto, Fábio Sossai Possebon, Leila Sabrina Ullmann, Cíntia Bittar, Guilherme Campos, Helena Lage Ferreira, Jorge A. Petroli Marchesi, Maisa C. Pereira Parra, Marília Moraes, Paula Rahal, Paulo Inacio da Costa, João Pessoa Araújo Jr., Maurício Lacerda Nogueira. Prefeitura de Sao Paulo: Melissa Palmieri. |
| EPI_ISL_1966727 | PPA MARCIO VALDEVINO BATISTA | Instituto Butantan | Instituto Butantan: Dimas Tadeu Covas, Sandra Coccuzzo Sampaio, Maria Carolina Elias, José Salvatore Leister Patané, Vincent Louis Viala, Antonio Jorge Martins, Ricardo Haddad, Claudia Renata dos Santos Barros, Elaine Cristina Marqueze, Raul Machado Neto, Debora Botequio Moretti, Jardelina de Souza Todao Bernardino, Loyze Paola Oliveira de Lima, Luiz Aurelio de Campos Crispin. Centro de Genômica Funcional da ESALQ: Luiz Lehmann Coutinho, Ricardo Augusto Brassaloti, Raquel de Lello Rocha Campos Cassano. NGS Soluções Genômicas: Pilar Drummond Sampaio Corrêa Mariani. |

| | | | |
|---|---|---|---|
| | | | FZEA-USP Pirassununga: Mirele Daiana Poleti, Jessika Cristina Chagas Lesbon, Elisangela Chicaroni Mattos, Heidge Fukumasu. USP-Botucatu: Rejane Maria Tommasini Grotto, Jayme A. Souza-Neto, Guilherme Targino Valente, Patricia Akemi Assato, Felipe Allan da Silva da Costa, Bianca Cechetto Carlos. Mendelics: Bibiana Santos, João Paulo Kitajima, Erika Freitas, David Schlesinger. Hemocentro Ribeirão Preto: Simone Kashima, Evandra Strazza Rodrigues, Svetoslav Nanev Slavov, Elaine Vieira dos Santos, Rafael dos Santos Bezerra, Luiz Carlos Junior de Alcantara, Marta Giovanetti, Vagner Fonseca, Flavia Aburjaile, Rodrigo Tocantins Calado. FAMERP-SJRP: Cecília Artico Banho, Lívia Sacchetto, Fábio Sossai Possebon, Leila Sabrina Ullmann, Cíntia Bittar, Guilherme Campos, Helena Lage Ferreira, Jorge A. Petroli Marchesi, Maisa C. Pereira Parra, Marília Moraes, Paula Rahal, Paulo Inacio da Costa, João Pessoa Araújo Jr., Maurício Lacerda Nogueira. Prefeitura de Sao Paulo: Melissa Palmieri. |
| EPI_ISL_1966728 | UBS JOSE SABINO FERREIRA | Instituto Butantan | Instituto Butantan: Dimas Tadeu Covas, Sandra Coccuzzo Sampaio, Maria Carolina Elias, José Salvatore Leister Patané, Vincent Louis Viala, Antonio Jorge Martins, Ricardo Haddad, Claudia Renata dos Santos Barros, Elaine Cristina Marqueze, Raul Machado Neto, Debora Botequio Moretti, Jardelina de Souza Todao Bernardino, Loyze Paola Oliveira de Lima, Luiz Aurelio de Campos Crispin. Centro de Genômica Funcional da ESALQ: Luiz Lehmann Coutinho, Ricardo Augusto Brassaloti, Raquel de Lello Rocha Campos Cassano. NGS Soluções Genômicas: Pilar Drummond Sampaio Corrêa Mariani. FZEA-USP Pirassununga: Mirele Daiana Poleti, Jessika Cristina Chagas Lesbon, Elisangela Chicaroni Mattos, Heidge Fukumasu. USP-Botucatu: Rejane Maria Tommasini Grotto, Jayme A. Souza-Neto, Guilherme Targino Valente, Patricia Akemi Assato, Felipe Allan da Silva da Costa, Bianca Cechetto Carlos. Mendelics: Bibiana Santos, João Paulo Kitajima, Erika Freitas, David Schlesinger. Hemocentro Ribeirão Preto: Simone Kashima, Evandra Strazza Rodrigues, Svetoslav Nanev Slavov, Elaine Vieira dos Santos, Rafael dos Santos Bezerra, Luiz Carlos Junior de Alcantara, Marta Giovanetti, Vagner Fonseca, Flavia Aburjaile, Rodrigo Tocantins Calado. FAMERP-SJRP: Cecília Artico Banho, Lívia Sacchetto, Fábio Sossai Possebon, Leila Sabrina Ullmann, Cíntia Bittar, Guilherme Campos, Helena Lage Ferreira, Jorge A. Petroli Marchesi, Maisa C. Pereira Parra, Marília Moraes, Paula Rahal, Paulo Inacio da Costa, João Pessoa Araújo Jr., Maurício Lacerda Nogueira. Prefeitura de Sao Paulo: Melissa Palmieri. |
| EPI_ISL_1966729 | UPA JOSE CAMPOS BARRETO ZEQUINHA BARRETO | Instituto Butantan | Instituto Butantan: Dimas Tadeu Covas, Sandra Coccuzzo Sampaio, Maria Carolina Elias, José Salvatore Leister Patané, Vincent Louis Viala, Antonio Jorge Martins, Ricardo Haddad, Claudia Renata dos Santos Barros, Elaine Cristina Marqueze, Raul Machado Neto, Debora Botequio Moretti, Jardelina de Souza Todao Bernardino, Loyze Paola Oliveira de Lima, Luiz Aurelio de Campos Crispin. Centro de Genômica Funcional da ESALQ: Luiz Lehmann Coutinho, Ricardo Augusto Brassaloti, Raquel de Lello Rocha Campos Cassano. NGS Soluções Genômicas: Pilar Drummond Sampaio Corrêa Mariani. FZEA-USP Pirassununga: Mirele Daiana Poleti, Jessika Cristina Chagas Lesbon, Elisangela Chicaroni Mattos, Heidge Fukumasu. USP-Botucatu: Rejane Maria Tommasini Grotto, Jayme A. Souza-Neto, Guilherme Targino Valente, Patricia Akemi Assato, Felipe Allan da Silva da Costa, Bianca Cechetto Carlos. Mendelics: Bibiana Santos, João Paulo Kitajima, Erika Freitas, David Schlesinger. Hemocentro Ribeirão Preto: Simone Kashima, Evandra Strazza Rodrigues, Svetoslav Nanev Slavov, Elaine Vieira dos Santos, Rafael dos Santos Bezerra, Luiz Carlos Junior de Alcantara, Marta Giovanetti, Vagner Fonseca, Flavia Aburjaile, Rodrigo Tocantins Calado. FAMERP-SJRP: Cecília Artico Banho, Lívia Sacchetto, Fábio Sossai Possebon, Leila Sabrina Ullmann, Cíntia Bittar, Guilherme Campos, Helena Lage Ferreira, Jorge A. Petroli Marchesi, Maisa C. Pereira Parra, Marília Moraes, Paula Rahal, Paulo Inacio da Costa, João Pessoa Araújo Jr., Maurício Lacerda Nogueira. Prefeitura de Sao Paulo: Melissa Palmieri. |
| EPI_ISL_1966730 | UBS JOSE SABINO FERREIRA | Instituto Butantan | Instituto Butantan: Dimas Tadeu Covas, Sandra Coccuzzo Sampaio, Maria Carolina Elias, José Salvatore Leister Patané, Vincent Louis Viala, Antonio Jorge Martins, Ricardo Haddad, Claudia Renata dos Santos Barros, Elaine Cristina Marqueze, Raul Machado Neto, Debora Botequio Moretti, Jardelina de Souza Todao Bernardino, Loyze Paola Oliveira de Lima, Luiz Aurelio de Campos Crispin. Centro de Genômica Funcional da ESALQ: Luiz Lehmann Coutinho, Ricardo Augusto Brassaloti, Raquel de Lello Rocha Campos Cassano. NGS Soluções Genômicas: Pilar Drummond Sampaio Corrêa Mariani. FZEA-USP Pirassununga: Mirele Daiana Poleti, Jessika Cristina Chagas Lesbon, Elisangela Chicaroni Mattos, Heidge Fukumasu. USP-Botucatu: Rejane Maria Tommasini Grotto, Jayme A. Souza-Neto, Guilherme Targino Valente, Patricia Akemi Assato, Felipe Allan da Silva da Costa, Bianca Cechetto Carlos. Mendelics: Bibiana Santos, João Paulo Kitajima, Erika Freitas, David Schlesinger. Hemocentro Ribeirão Preto: Simone Kashima, Evandra Strazza Rodrigues, Svetoslav Nanev Slavov, Elaine Vieira dos Santos, Rafael dos Santos Bezerra, Luiz Carlos Junior de Alcantara, Marta Giovanetti, Vagner Fonseca, Flavia Aburjaile, Rodrigo Tocantins Calado. FAMERP-SJRP: Cecília Artico Banho, Lívia Sacchetto, Fábio Sossai Possebon, Leila Sabrina Ullmann, Cíntia Bittar, Guilherme Campos, Helena Lage Ferreira, Jorge A. Petroli Marchesi, Maisa C. Pereira Parra, Marília Moraes, Paula Rahal, Paulo Inacio da Costa, João Pessoa Araújo Jr., Maurício Lacerda Nogueira. Prefeitura de Sao Paulo: Melissa Palmieri. |
| EPI_ISL_1966731, EPI_ISL_1966732, EPI_ISL_1966733, EPI_ISL_1966734 | UBS RAIMUNDA CAVALCANTE DE SOUZA | Instituto Butantan | Instituto Butantan: Dimas Tadeu Covas, Sandra Coccuzzo Sampaio, Maria Carolina Elias, José Salvatore Leister Patané, Vincent Louis Viala, Antonio Jorge Martins, Ricardo Haddad, Claudia Renata dos Santos Barros, Elaine Cristina Marqueze, Raul Machado Neto, Debora Botequio Moretti, Jardelina de Souza Todao Bernardino, Loyze Paola Oliveira de Lima, Luiz Aurelio de Campos Crispin. Centro de Genômica Funcional da ESALQ: Luiz Lehmann Coutinho, Ricardo Augusto Brassaloti, Raquel de Lello Rocha Campos Cassano. NGS Soluções Genômicas: Pilar Drummond Sampaio Corrêa Mariani. FZEA-USP Pirassununga: Mirele Daiana Poleti, Jessika Cristina Chagas Lesbon, Elisangela Chicaroni Mattos, Heidge Fukumasu. USP-Botucatu: Rejane Maria Tommasini Grotto, Jayme A. Souza-Neto, Guilherme Targino Valente, Patricia Akemi Assato, Felipe Allan da Silva da Costa, Bianca Cechetto Carlos. Mendelics: Bibiana Santos, João Paulo Kitajima, Erika Freitas, David Schlesinger. Hemocentro Ribeirão Preto: Simone Kashima, Evandra Strazza Rodrigues, Svetoslav Nanev Slavov, Elaine Vieira dos Santos, Rafael dos Santos Bezerra, Luiz Carlos Junior de Alcantara, Marta Giovanetti, Vagner Fonseca, Flavia Aburjaile, Rodrigo Tocantins Calado. FAMERP-SJRP: Cecília Artico Banho, Lívia Sacchetto, Fábio Sossai Possebon, Leila Sabrina Ullmann, Cíntia Bittar, Guilherme Campos, Helena Lage Ferreira, Jorge A. Petroli Marchesi, Maisa C. Pereira Parra, Marília Moraes, Paula Rahal, Paulo Inacio da Costa, João Pessoa Araújo Jr., Maurício Lacerda Nogueira. Prefeitura de Sao Paulo: Melissa Palmieri. |
| EPI_ISL_1966735, EPI_ISL_1966736 | PRONTO SOCORRO PESTANA P S ANDRE SACCO | Instituto Butantan | Instituto Butantan: Dimas Tadeu Covas, Sandra Coccuzzo Sampaio, Maria Carolina Elias, José Salvatore Leister Patané, Vincent Louis Viala, Antonio Jorge Martins, Ricardo Haddad, Claudia Renata dos Santos Barros, Elaine Cristina Marqueze, Raul Machado Neto, Debora Botequio Moretti, Jardelina de Souza Todao Bernardino, Loyze Paola Oliveira de Lima, Luiz Aurelio de Campos Crispin. Centro de Genômica Funcional da ESALQ: Luiz Lehmann Coutinho, Ricardo Augusto Brassaloti, Raquel de Lello Rocha Campos Cassano. NGS Soluções Genômicas: Pilar Drummond Sampaio Corrêa Mariani. FZEA-USP Pirassununga: Mirele Daiana Poleti, Jessika Cristina Chagas Lesbon, Elisangela Chicaroni Mattos, Heidge Fukumasu. USP-Botucatu: Rejane Maria Tommasini Grotto, Jayme A. Souza-Neto, Guilherme Targino Valente, Patricia Akemi Assato, Felipe Allan da Silva da Costa, Bianca Cechetto Carlos. Mendelics: Bibiana Santos, João Paulo Kitajima, Erika Freitas, David Schlesinger. Hemocentro Ribeirão Preto: Simone Kashima, Evandra Strazza Rodrigues, Svetoslav Nanev Slavov, Elaine Vieira dos Santos, Rafael dos Santos Bezerra, Luiz Carlos Junior de Alcantara, Marta Giovanetti, Vagner Fonseca, Flavia Aburjaile, Rodrigo Tocantins Calado. FAMERP-SJRP: Cecília Artico Banho, Lívia Sacchetto, Fábio Sossai Possebon, Leila Sabrina Ullmann, Cíntia Bittar, Guilherme Campos, Helena Lage Ferreira, Jorge A. Petroli Marchesi, Maisa C. Pereira Parra, Marília Moraes, Paula Rahal, Paulo Inacio da Costa, João Pessoa Araújo Jr., Maurício Lacerda Nogueira. Prefeitura de Sao Paulo: Melissa Palmieri. |
| EPI_ISL_1966737 | HOSP MUN JOSANIAS CASTANHA BRAGA | Instituto Butantan | Instituto Butantan: Dimas Tadeu Covas, Sandra Coccuzzo Sampaio, Maria Carolina Elias, José Salvatore Leister Patané, Vincent Louis Viala, Antonio Jorge Martins, Ricardo Haddad, Claudia Renata dos Santos Barros, Elaine Cristina Marqueze, Raul Machado Neto, Debora Botequio Moretti, Jardelina de Souza Todao Bernardino, Loyze Paola Oliveira de Lima, Luiz Aurelio de Campos Crispin. Centro de Genômica Funcional da ESALQ: Luiz Lehmann Coutinho, Ricardo Augusto Brassaloti, Raquel de Lello Rocha Campos Cassano. NGS Soluções Genômicas: Pilar Drummond Sampaio Corrêa Mariani. FZEA-USP Pirassununga: Mirele Daiana Poleti, Jessika Cristina Chagas Lesbon, Elisangela Chicaroni Mattos, Heidge Fukumasu. USP-Botucatu: Rejane Maria Tommasini Grotto, Jayme A. Souza-Neto, Guilherme Targino Valente, Patricia Akemi Assato, Felipe Allan da Silva da Costa, Bianca Cechetto Carlos. Mendelics: Bibiana Santos, João Paulo Kitajima, Erika Freitas, David Schlesinger. Hemocentro Ribeirão Preto: Simone Kashima, Evandra Strazza Rodrigues, Svetoslav Nanev Slavov, Elaine Vieira dos Santos, Rafael dos Santos Bezerra, Luiz Carlos Junior de Alcantara, Marta Giovanetti, Vagner Fonseca, Flavia Aburjaile, Rodrigo Tocantins Calado. FAMERP-SJRP: Cecília Artico Banho, Lívia Sacchetto, Fábio Sossai Possebon, Leila Sabrina Ullmann, Cíntia Bittar, Guilherme Campos, Helena Lage Ferreira, Jorge A. Petroli Marchesi, Maisa C. Pereira Parra, Marília Moraes, Paula Rahal, Paulo Inacio da Costa, João Pessoa Araújo Jr., Maurício Lacerda Nogueira. Prefeitura de Sao Paulo: Melissa Palmieri. |
| EPI_ISL_1966738 | HOSPITAL GERAL DE VILA NOVA CACHOEIRINHA SAO PAULO | Instituto Butantan | Instituto Butantan: Dimas Tadeu Covas, Sandra Coccuzzo Sampaio, Maria Carolina Elias, José Salvatore Leister Patané, Vincent Louis Viala, Antonio Jorge Martins, Ricardo Haddad, Claudia Renata dos Santos Barros, Elaine Cristina Marqueze, Raul Machado Neto, Debora Botequio Moretti, Jardelina de Souza Todao Bernardino, Loyze Paola Oliveira de Lima, Luiz Aurelio de Campos Crispin. Centro de Genômica Funcional da ESALQ: Luiz Lehmann Coutinho, Ricardo Augusto Brassaloti, Raquel de Lello Rocha Campos Cassano. NGS Soluções Genômicas: Pilar Drummond Sampaio Corrêa Mariani. FZEA-USP Pirassununga: Mirele Daiana Poleti, Jessika Cristina Chagas Lesbon, Elisangela Chicaroni Mattos, Heidge Fukumasu. USP-Botucatu: Rejane Maria Tommasini Grotto, Jayme A. Souza-Neto, Guilherme Targino Valente, Patricia Akemi Assato, Felipe Allan da Silva da Costa, Bianca Cechetto Carlos. Mendelics: Bibiana Santos, João Paulo Kitajima, Erika Freitas, David Schlesinger. Hemocentro Ribeirão Preto: Simone Kashima, Evandra Strazza Rodrigues, Svetoslav Nanev Slavov, Elaine Vieira dos Santos, Rafael dos Santos Bezerra, Luiz Carlos Junior de Alcantara, Marta Giovanetti, Vagner Fonseca, Flavia Aburjaile, Rodrigo Tocantins Calado. FAMERP-SJRP: Cecília Artico Banho, Lívia Sacchetto, Fábio Sossai Possebon, Leila Sabrina Ullmann, Cíntia Bittar, Guilherme Campos, Helena Lage Ferreira, Jorge A. Petroli Marchesi, Maisa C. Pereira Parra, Marília Moraes, Paula Rahal, Paulo Inacio da Costa, João Pessoa Araújo Jr., Maurício Lacerda Nogueira. Prefeitura de Sao Paulo: Melissa Palmieri. |

| | | | |
|---|---|---|---|
| EPI_ISL_1966739, EPI_ISL_1966740, EPI_ISL_1966741 | CS DE AMERICO DE CAMPOS | Instituto Butantan / Mendelics | Instituto Butantan: Dimas Tadeu Covas, Sandra Coccuzzo Sampaio, Maria Carolina Elias, José Salvatore Leister Patané, Vincent Louis Viala, Antonio Jorge Martins, Ricardo Haddad, Claudia Renata dos Santos Barros, Elaine Cristina Marqueze, Raul Machado Neto, Debora Botequio Moretti, Jardelina de Souza Todao Bernardino, Loyze Paola Oliveira de Lima, Luiz Aurelio de Campos Crispin. Centro de Genômica Funcional da ESALQ: Luiz Lehmann Coutinho, Ricardo Augusto Brassaloti, Raquel de Lello Rocha Campos Cassano. NGS Soluções Genômicas: Pilar Drummond Sampaio Corrêa Mariani. FZEA-USP Pirassununga: Mirele Daiana Poleti, Jessika Cristina Chagas Lesbon, Elisangela Chicaroni Mattos, Heidge Fukumasu. USP-Botucatu: Rejane Maria Tommasini Grotto, Jayme A. Souza-Neto, Guilherme Targino Valente, Patricia Akemi Assato, Felipe Allan da Silva da Costa, Bianca Cechetto Carlos. Mendelics: Bibiana Santos, João Paulo Kitajima, Erika Freitas, David Schlesinger. Hemocentro Ribeirão Preto: Simone Kashima, Evandra Strazza Rodrigues, Svetoslav Nanev Slavov, Elaine Vieira dos Santos, Rafael dos Santos Bezerra, Luiz Carlos Junior de Alcantara, Marta Giovanetti, Vagner Fonseca, Flavia Aburjaile, Rodrigo Tocantins Calado. FAMERP-SJRP: Cecília Artico Banho, Lívia Sacchetto, Fábio Sossai Possebon, Leila Sabrina Ullmann, Cíntia Bittar, Guilherme Campos, Helena Lage Ferreira, Jorge A. Petroli Marchesi, Maisa C. Pereira Parra, Marília Moraes, Paula Rahal, Paulo Inacio da Costa, João Pessoa Araújo Jr., Maurício Lacerda Nogueira. Prefeitura de Sao Paulo: Melissa Palmieri. |
| EPI_ISL_1966742, EPI_ISL_1966743, EPI_ISL_1966744 | CENTRO DE SAUDE DR RENATO DE CARVALHO RIBEIRO ANGATUBA | Instituto Butantan / Mendelics | Instituto Butantan: Dimas Tadeu Covas, Sandra Coccuzzo Sampaio, Maria Carolina Elias, José Salvatore Leister Patané, Vincent Louis Viala, Antonio Jorge Martins, Ricardo Haddad, Claudia Renata dos Santos Barros, Elaine Cristina Marqueze, Raul Machado Neto, Debora Botequio Moretti, Jardelina de Souza Todao Bernardino, Loyze Paola Oliveira de Lima, Luiz Aurelio de Campos Crispin. Centro de Genômica Funcional da ESALQ: Luiz Lehmann Coutinho, Ricardo Augusto Brassaloti, Raquel de Lello Rocha Campos Cassano. NGS Soluções Genômicas: Pilar Drummond Sampaio Corrêa Mariani. FZEA-USP Pirassununga: Mirele Daiana Poleti, Jessika Cristina Chagas Lesbon, Elisangela Chicaroni Mattos, Heidge Fukumasu. USP-Botucatu: Rejane Maria Tommasini Grotto, Jayme A. Souza-Neto, Guilherme Targino Valente, Patricia Akemi Assato, Felipe Allan da Silva da Costa, Bianca Cechetto Carlos. Mendelics: Bibiana Santos, João Paulo Kitajima, Erika Freitas, David Schlesinger. Hemocentro Ribeirão Preto: Simone Kashima, Evandra Strazza Rodrigues, Svetoslav Nanev Slavov, Elaine Vieira dos Santos, Rafael dos Santos Bezerra, Luiz Carlos Junior de Alcantara, Marta Giovanetti, Vagner Fonseca, Flavia Aburjaile, Rodrigo Tocantins Calado. FAMERP-SJRP: Cecília Artico Banho, Lívia Sacchetto, Fábio Sossai Possebon, Leila Sabrina Ullmann, Cíntia Bittar, Guilherme Campos, Helena Lage Ferreira, Jorge A. Petroli Marchesi, Maisa C. Pereira Parra, Marília Moraes, Paula Rahal, Paulo Inacio da Costa, João Pessoa Araújo Jr., Maurício Lacerda Nogueira. Prefeitura de Sao Paulo: Melissa Palmieri. |
| EPI_ISL_1966745 | VIGILANCIA EPIDIMIOLOGICA DE ARACARIGUAMA | Instituto Butantan / Mendelics | Instituto Butantan: Dimas Tadeu Covas, Sandra Coccuzzo Sampaio, Maria Carolina Elias, José Salvatore Leister Patané, Vincent Louis Viala, Antonio Jorge Martins, Ricardo Haddad, Claudia Renata dos Santos Barros, Elaine Cristina Marqueze, Raul Machado Neto, Debora Botequio Moretti, Jardelina de Souza Todao Bernardino, Loyze Paola Oliveira de Lima, Luiz Aurelio de Campos Crispin. Centro de Genômica Funcional da ESALQ: Luiz Lehmann Coutinho, Ricardo Augusto Brassaloti, Raquel de Lello Rocha Campos Cassano. NGS Soluções Genômicas: Pilar Drummond Sampaio Corrêa Mariani. FZEA-USP Pirassununga: Mirele Daiana Poleti, Jessika Cristina Chagas Lesbon, Elisangela Chicaroni Mattos, Heidge Fukumasu. USP-Botucatu: Rejane Maria Tommasini Grotto, Jayme A. Souza-Neto, Guilherme Targino Valente, Patricia Akemi Assato, Felipe Allan da Silva da Costa, Bianca Cechetto Carlos. Mendelics: Bibiana Santos, João Paulo Kitajima, Erika Freitas, David Schlesinger. Hemocentro Ribeirão Preto: Simone Kashima, Evandra Strazza Rodrigues, Svetoslav Nanev Slavov, Elaine Vieira dos Santos, Rafael dos Santos Bezerra, Luiz Carlos Junior de Alcantara, Marta Giovanetti, Vagner Fonseca, Flavia Aburjaile, Rodrigo Tocantins Calado. FAMERP-SJRP: Cecília Artico Banho, Lívia Sacchetto, Fábio Sossai Possebon, Leila Sabrina Ullmann, Cíntia Bittar, Guilherme Campos, Helena Lage Ferreira, Jorge A. Petroli Marchesi, Maisa C. Pereira Parra, Marília Moraes, Paula Rahal, Paulo Inacio da Costa, João Pessoa Araújo Jr., Maurício Lacerda Nogueira. Prefeitura de Sao Paulo: Melissa Palmieri. |
| EPI_ISL_1966746, EPI_ISL_1966747, EPI_ISL_1966748 | SMS SECRETARIA MUNICIPAL DE SAUDE DE BOITUVA | Instituto Butantan / Mendelics | Instituto Butantan: Dimas Tadeu Covas, Sandra Coccuzzo Sampaio, Maria Carolina Elias, José Salvatore Leister Patané, Vincent Louis Viala, Antonio Jorge Martins, Ricardo Haddad, Claudia Renata dos Santos Barros, Elaine Cristina Marqueze, Raul Machado Neto, Debora Botequio Moretti, Jardelina de Souza Todao Bernardino, Loyze Paola Oliveira de Lima, Luiz Aurelio de Campos Crispin. Centro de Genômica Funcional da ESALQ: Luiz Lehmann Coutinho, Ricardo Augusto Brassaloti, Raquel de Lello Rocha Campos Cassano. NGS Soluções Genômicas: Pilar Drummond Sampaio Corrêa Mariani. FZEA-USP Pirassununga: Mirele Daiana Poleti, Jessika Cristina Chagas Lesbon, Elisangela Chicaroni Mattos, Heidge Fukumasu. USP-Botucatu: Rejane Maria Tommasini Grotto, Jayme A. Souza-Neto, Guilherme Targino Valente, Patricia Akemi Assato, Felipe Allan da Silva da Costa, Bianca Cechetto Carlos. Mendelics: Bibiana Santos, João Paulo Kitajima, Erika Freitas, David Schlesinger. Hemocentro Ribeirão Preto: Simone Kashima, Evandra Strazza Rodrigues, Svetoslav Nanev Slavov, Elaine Vieira dos Santos, Rafael dos Santos Bezerra, Luiz Carlos Junior de Alcantara, Marta Giovanetti, Vagner Fonseca, Flavia Aburjaile, Rodrigo Tocantins Calado. FAMERP-SJRP: Cecília Artico Banho, Lívia Sacchetto, Fábio Sossai Possebon, Leila Sabrina Ullmann, Cíntia Bittar, Guilherme Campos, Helena Lage Ferreira, Jorge A. Petroli Marchesi, Maisa C. Pereira Parra, Marília Moraes, Paula Rahal, Paulo Inacio da Costa, João Pessoa Araújo Jr., Maurício Lacerda Nogueira. Prefeitura de Sao Paulo: Melissa Palmieri. |
| EPI_ISL_1966749 | CENTRO DE SAUDE III BORBOREMA | Instituto Butantan / Mendelics | Instituto Butantan: Dimas Tadeu Covas, Sandra Coccuzzo Sampaio, Maria Carolina Elias, José Salvatore Leister Patané, Vincent Louis Viala, Antonio Jorge Martins, Ricardo Haddad, Claudia Renata dos Santos Barros, Elaine Cristina Marqueze, Raul Machado Neto, Debora Botequio Moretti, Jardelina de Souza Todao Bernardino, Loyze Paola Oliveira de Lima, Luiz Aurelio de Campos Crispin. Centro de Genômica Funcional da ESALQ: Luiz Lehmann Coutinho, Ricardo Augusto Brassaloti, Raquel de Lello Rocha Campos Cassano. NGS Soluções Genômicas: Pilar Drummond Sampaio Corrêa Mariani. FZEA-USP Pirassununga: Mirele Daiana Poleti, Jessika Cristina Chagas Lesbon, Elisangela Chicaroni Mattos, Heidge Fukumasu. USP-Botucatu: Rejane Maria Tommasini Grotto, Jayme A. Souza-Neto, Guilherme Targino Valente, Patricia Akemi Assato, Felipe Allan da Silva da Costa, Bianca Cechetto Carlos. Mendelics: Bibiana Santos, João Paulo Kitajima, Erika Freitas, David Schlesinger. Hemocentro Ribeirão Preto: Simone Kashima, Evandra Strazza Rodrigues, Svetoslav Nanev Slavov, Elaine Vieira dos Santos, Rafael dos Santos Bezerra, Luiz Carlos Junior de Alcantara, Marta Giovanetti, Vagner Fonseca, Flavia Aburjaile, Rodrigo Tocantins Calado. FAMERP-SJRP: Cecília Artico Banho, Lívia Sacchetto, Fábio Sossai Possebon, Leila Sabrina Ullmann, Cíntia Bittar, Guilherme Campos, Helena Lage Ferreira, Jorge A. Petroli Marchesi, Maisa C. Pereira Parra, Marília Moraes, Paula Rahal, Paulo Inacio da Costa, João Pessoa Araújo Jr., Maurício Lacerda Nogueira. Prefeitura de Sao Paulo: Melissa Palmieri. |
| EPI_ISL_1966751 | PRONTO ATENDIMENTO MUNICIPAL ITALO SANTUCCI | Instituto Butantan / Mendelics | Instituto Butantan: Dimas Tadeu Covas, Sandra Coccuzzo Sampaio, Maria Carolina Elias, José Salvatore Leister Patané, Vincent Louis Viala, Antonio Jorge Martins, Ricardo Haddad, Claudia Renata dos Santos Barros, Elaine Cristina Marqueze, Raul Machado Neto, Debora Botequio Moretti, Jardelina de Souza Todao Bernardino, Loyze Paola Oliveira de Lima, Luiz Aurelio de Campos Crispin. Centro de Genômica Funcional da ESALQ: Luiz Lehmann Coutinho, Ricardo Augusto Brassaloti, Raquel de Lello Rocha Campos Cassano. NGS Soluções Genômicas: Pilar Drummond Sampaio Corrêa Mariani. FZEA-USP Pirassununga: Mirele Daiana Poleti, Jessika Cristina Chagas Lesbon, Elisangela Chicaroni Mattos, Heidge Fukumasu. USP-Botucatu: Rejane Maria Tommasini Grotto, Jayme A. Souza-Neto, Guilherme Targino Valente, Patricia Akemi Assato, Felipe Allan da Silva da Costa, Bianca Cechetto Carlos. Mendelics: Bibiana Santos, João Paulo Kitajima, Erika Freitas, David Schlesinger. Hemocentro Ribeirão Preto: Simone Kashima, Evandra Strazza Rodrigues, Svetoslav Nanev Slavov, Elaine Vieira dos Santos, Rafael dos Santos Bezerra, Luiz Carlos Junior de Alcantara, Marta Giovanetti, Vagner Fonseca, Flavia Aburjaile, Rodrigo Tocantins Calado. FAMERP-SJRP: Cecília Artico Banho, Lívia Sacchetto, Fábio Sossai Possebon, Leila Sabrina Ullmann, Cíntia Bittar, Guilherme Campos, Helena Lage Ferreira, Jorge A. Petroli Marchesi, Maisa C. Pereira Parra, Marília Moraes, Paula Rahal, Paulo Inacio da Costa, João Pessoa Araújo Jr., Maurício Lacerda Nogueira. Prefeitura de Sao Paulo: Melissa Palmieri. |
| EPI_ISL_1966753 | UNIDADE SENTINELA COVID19 | Instituto Butantan / Mendelics | Instituto Butantan: Dimas Tadeu Covas, Sandra Coccuzzo Sampaio, Maria Carolina Elias, José Salvatore Leister Patané, Vincent Louis Viala, Antonio Jorge Martins, Ricardo Haddad, Claudia Renata dos Santos Barros, Elaine Cristina Marqueze, Raul Machado Neto, Debora Botequio Moretti, Jardelina de Souza Todao Bernardino, Loyze Paola Oliveira de Lima, Luiz Aurelio de Campos Crispin. Centro de Genômica Funcional da ESALQ: Luiz Lehmann Coutinho, Ricardo Augusto Brassaloti, Raquel de Lello Rocha Campos Cassano. NGS Soluções Genômicas: Pilar Drummond Sampaio Corrêa Mariani. FZEA-USP Pirassununga: Mirele Daiana Poleti, Jessika Cristina Chagas Lesbon, Elisangela Chicaroni Mattos, Heidge Fukumasu. USP-Botucatu: Rejane Maria Tommasini Grotto, Jayme A. Souza-Neto, Guilherme Targino Valente, Patricia Akemi Assato, Felipe Allan da Silva da Costa, Bianca Cechetto Carlos. Mendelics: Bibiana Santos, João Paulo Kitajima, Erika Freitas, David Schlesinger. Hemocentro Ribeirão Preto: Simone Kashima, Evandra Strazza Rodrigues, Svetoslav Nanev Slavov, Elaine Vieira dos Santos, Rafael dos Santos Bezerra, Luiz Carlos Junior de Alcantara, Marta Giovanetti, Vagner Fonseca, Flavia Aburjaile, Rodrigo Tocantins Calado. FAMERP-SJRP: Cecília Artico Banho, Lívia Sacchetto, Fábio Sossai Possebon, Leila Sabrina Ullmann, Cíntia Bittar, Guilherme Campos, Helena Lage Ferreira, Jorge A. Petroli Marchesi, Maisa C. Pereira Parra, Marília Moraes, Paula Rahal, Paulo Inacio da Costa, João Pessoa Araújo Jr., Maurício Lacerda Nogueira. Prefeitura de Sao Paulo: Melissa Palmieri. |
| EPI_ISL_1966755 | UNIDADE DE SAUDE DA FAMILIA I COLOMBIA | Instituto Butantan / Mendelics | Instituto Butantan: Dimas Tadeu Covas, Sandra Coccuzzo Sampaio, Maria Carolina Elias, José Salvatore Leister Patané, Vincent Louis Viala, Antonio Jorge Martins, Ricardo Haddad, Claudia Renata dos Santos Barros, Elaine Cristina Marqueze, Raul Machado Neto, Debora Botequio Moretti, Jardelina de Souza Todao Bernardino, Loyze Paola Oliveira de Lima, Luiz Aurelio de Campos Crispin. Centro de Genômica Funcional da ESALQ: Luiz Lehmann Coutinho, Ricardo Augusto Brassaloti, Raquel de Lello Rocha Campos Cassano. NGS Soluções Genômicas: Pilar Drummond Sampaio Corrêa Mariani. FZEA-USP Pirassununga: Mirele Daiana Poleti, Jessika Cristina Chagas Lesbon, Elisangela Chicaroni Mattos, Heidge Fukumasu. USP-Botucatu: Rejane Maria Tommasini Grotto, Jayme A. Souza-Neto, Guilherme Targino Valente, Patricia Akemi Assato, Felipe Allan da Silva da Costa, Bianca Cechetto Carlos. Mendelics: Bibiana Santos, João Paulo Kitajima, Erika Freitas, David Schlesinger. Hemocentro Ribeirão Preto: Simone Kashima, Evandra Strazza |

| | | | |
|---|---|---|---|
| | | | Rodrigues, Svetoslav Nanev Slavov, Elaine Vieira dos Santos, Rafael dos Santos Bezerra, Luiz Carlos Junior de Alcantara, Marta Giovanetti, Vagner Fonseca, Flavia Aburjaile, Rodrigo Tocantins Calado. FAMERP-SJRP: Cecília Artico Banho, Lívia Sacchetto, Fábio Sossai Possebon, Leila Sabrina Ullmann, Cíntia Bittar, Guilherme Campos, Helena Lage Ferreira, Jorge A. Petroli Marchesi, Maisa C. Pereira Parra, Marília Moraes, Paula Rahal, Paulo Inacio da Costa, João Pessoa Araújo Jr., Maurício Lacerda Nogueira. Prefeitura de Sao Paulo: Melissa Palmieri. |
| EPI_ISL_1966756, EPI_ISL_1966757, EPI_ISL_1966758, EPI_ISL_1966759, EPI_ISL_1966760 | LABORATORIO DE FRANCA | Instituto Butantan / Mendelics | Instituto Butantan: Dimas Tadeu Covas, Sandra Coccuzzo Sampaio, Maria Carolina Elias, José Salvatore Leister Patané, Vincent Louis Viala, Antonio Jorge Martins, Ricardo Haddad, Claudia Renata dos Santos Barros, Elaine Cristina Marqueze, Raul Machado Neto, Debora Botequio Moretti, Jardelina de Souza Todao Bernardino, Loyze Paola Oliveira de Lima, Luiz Aurelio de Campos Crispin. Centro de Genômica Funcional da ESALQ: Luiz Lehmann Coutinho, Ricardo Augusto Brassaloti, Raquel de Lello Rocha Campos Cassano. NGS Soluções Genômicas: Pilar Drummond Sampaio Corrêa Mariani. FZEA-USP Pirassununga: Mirele Daiana Poleti, Jessika Cristina Chagas Lesbon, Elisangela Chicaroni Mattos, Heidge Fukumasu. USP-Botucatu: Rejane Maria Tommasini Grotto, Jayme A. Souza-Neto, Guilherme Targino Valente, Patricia Akemi Assato, Felipe Allan da Silva da Costa, Bianca Cechetto Carlos. Mendelics: Bibiana Santos, João Paulo Kitajima, Erika Freitas, David Schlesinger. Hemocentro Ribeirão Preto: Simone Kashima, Evandra Strazza Rodrigues, Svetoslav Nanev Slavov, Elaine Vieira dos Santos, Rafael dos Santos Bezerra, Luiz Carlos Junior de Alcantara, Marta Giovanetti, Vagner Fonseca, Flavia Aburjaile, Rodrigo Tocantins Calado. FAMERP-SJRP: Cecília Artico Banho, Lívia Sacchetto, Fábio Sossai Possebon, Leila Sabrina Ullmann, Cíntia Bittar, Guilherme Campos, Helena Lage Ferreira, Jorge A. Petroli Marchesi, Maisa C. Pereira Parra, Marília Moraes, Paula Rahal, Paulo Inacio da Costa, João Pessoa Araújo Jr., Maurício Lacerda Nogueira. Prefeitura de Sao Paulo: Melissa Palmieri. |
| EPI_ISL_1966761, EPI_ISL_1966762 | UBS GLENDA RODELLA DUMAS | Instituto Butantan / Mendelics | Instituto Butantan: Dimas Tadeu Covas, Sandra Coccuzzo Sampaio, Maria Carolina Elias, José Salvatore Leister Patané, Vincent Louis Viala, Antonio Jorge Martins, Ricardo Haddad, Claudia Renata dos Santos Barros, Elaine Cristina Marqueze, Raul Machado Neto, Debora Botequio Moretti, Jardelina de Souza Todao Bernardino, Loyze Paola Oliveira de Lima, Luiz Aurelio de Campos Crispin. Centro de Genômica Funcional da ESALQ: Luiz Lehmann Coutinho, Ricardo Augusto Brassaloti, Raquel de Lello Rocha Campos Cassano. NGS Soluções Genômicas: Pilar Drummond Sampaio Corrêa Mariani. FZEA-USP Pirassununga: Mirele Daiana Poleti, Jessika Cristina Chagas Lesbon, Elisangela Chicaroni Mattos, Heidge Fukumasu. USP-Botucatu: Rejane Maria Tommasini Grotto, Jayme A. Souza-Neto, Guilherme Targino Valente, Patricia Akemi Assato, Felipe Allan da Silva da Costa, Bianca Cechetto Carlos. Mendelics: Bibiana Santos, João Paulo Kitajima, Erika Freitas, David Schlesinger. Hemocentro Ribeirão Preto: Simone Kashima, Evandra Strazza Rodrigues, Svetoslav Nanev Slavov, Elaine Vieira dos Santos, Rafael dos Santos Bezerra, Luiz Carlos Junior de Alcantara, Marta Giovanetti, Vagner Fonseca, Flavia Aburjaile, Rodrigo Tocantins Calado. FAMERP-SJRP: Cecília Artico Banho, Lívia Sacchetto, Fábio Sossai Possebon, Leila Sabrina Ullmann, Cíntia Bittar, Guilherme Campos, Helena Lage Ferreira, Jorge A. Petroli Marchesi, Maisa C. Pereira Parra, Marília Moraes, Paula Rahal, Paulo Inacio da Costa, João Pessoa Araújo Jr., Maurício Lacerda Nogueira. Prefeitura de Sao Paulo: Melissa Palmieri. |
| EPI_ISL_1966763 | HOSPITAL MUNICIPAL DE IBIUNA IBIUNA SP | Instituto Butantan / Mendelics | Instituto Butantan: Dimas Tadeu Covas, Sandra Coccuzzo Sampaio, Maria Carolina Elias, José Salvatore Leister Patané, Vincent Louis Viala, Antonio Jorge Martins, Ricardo Haddad, Claudia Renata dos Santos Barros, Elaine Cristina Marqueze, Raul Machado Neto, Debora Botequio Moretti, Jardelina de Souza Todao Bernardino, Loyze Paola Oliveira de Lima, Luiz Aurelio de Campos Crispin. Centro de Genômica Funcional da ESALQ: Luiz Lehmann Coutinho, Ricardo Augusto Brassaloti, Raquel de Lello Rocha Campos Cassano. NGS Soluções Genômicas: Pilar Drummond Sampaio Corrêa Mariani. FZEA-USP Pirassununga: Mirele Daiana Poleti, Jessika Cristina Chagas Lesbon, Elisangela Chicaroni Mattos, Heidge Fukumasu. USP-Botucatu: Rejane Maria Tommasini Grotto, Jayme A. Souza-Neto, Guilherme Targino Valente, Patricia Akemi Assato, Felipe Allan da Silva da Costa, Bianca Cechetto Carlos. Mendelics: Bibiana Santos, João Paulo Kitajima, Erika Freitas, David Schlesinger. Hemocentro Ribeirão Preto: Simone Kashima, Evandra Strazza Rodrigues, Svetoslav Nanev Slavov, Elaine Vieira dos Santos, Rafael dos Santos Bezerra, Luiz Carlos Junior de Alcantara, Marta Giovanetti, Vagner Fonseca, Flavia Aburjaile, Rodrigo Tocantins Calado. FAMERP-SJRP: Cecília Artico Banho, Lívia Sacchetto, Fábio Sossai Possebon, Leila Sabrina Ullmann, Cíntia Bittar, Guilherme Campos, Helena Lage Ferreira, Jorge A. Petroli Marchesi, Maisa C. Pereira Parra, Marília Moraes, Paula Rahal, Paulo Inacio da Costa, João Pessoa Araújo Jr., Maurício Lacerda Nogueira. Prefeitura de Sao Paulo: Melissa Palmieri. |
| EPI_ISL_1966764, EPI_ISL_1966765 | CS DE ICEM | Instituto Butantan / Mendelics | Instituto Butantan: Dimas Tadeu Covas, Sandra Coccuzzo Sampaio, Maria Carolina Elias, José Salvatore Leister Patané, Vincent Louis Viala, Antonio Jorge Martins, Ricardo Haddad, Claudia Renata dos Santos Barros, Elaine Cristina Marqueze, Raul Machado Neto, Debora Botequio Moretti, Jardelina de Souza Todao Bernardino, Loyze Paola Oliveira de Lima, Luiz Aurelio de Campos Crispin. Centro de Genômica Funcional da ESALQ: Luiz Lehmann Coutinho, Ricardo Augusto Brassaloti, Raquel de Lello Rocha Campos Cassano. NGS Soluções Genômicas: Pilar Drummond Sampaio Corrêa Mariani. FZEA-USP Pirassununga: Mirele Daiana Poleti, Jessika Cristina Chagas Lesbon, Elisangela Chicaroni Mattos, Heidge Fukumasu. USP-Botucatu: Rejane Maria Tommasini Grotto, Jayme A. Souza-Neto, Guilherme Targino Valente, Patricia Akemi Assato, Felipe Allan da Silva da Costa, Bianca Cechetto Carlos. Mendelics: Bibiana Santos, João Paulo Kitajima, Erika Freitas, David Schlesinger. Hemocentro Ribeirão Preto: Simone Kashima, Evandra Strazza Rodrigues, Svetoslav Nanev Slavov, Elaine Vieira dos Santos, Rafael dos Santos Bezerra, Luiz Carlos Junior de Alcantara, Marta Giovanetti, Vagner Fonseca, Flavia Aburjaile, Rodrigo Tocantins Calado. FAMERP-SJRP: Cecília Artico Banho, Lívia Sacchetto, Fábio Sossai Possebon, Leila Sabrina Ullmann, Cíntia Bittar, Guilherme Campos, Helena Lage Ferreira, Jorge A. Petroli Marchesi, Maisa C. Pereira Parra, Marília Moraes, Paula Rahal, Paulo Inacio da Costa, João Pessoa Araújo Jr., Maurício Lacerda Nogueira. Prefeitura de Sao Paulo: Melissa Palmieri. |
| EPI_ISL_1966766, EPI_ISL_1966767, EPI_ISL_1966768 | POLICLINICA COVID 19 ITAPETININGA | Instituto Butantan / Mendelics | Instituto Butantan: Dimas Tadeu Covas, Sandra Coccuzzo Sampaio, Maria Carolina Elias, José Salvatore Leister Patané, Vincent Louis Viala, Antonio Jorge Martins, Ricardo Haddad, Claudia Renata dos Santos Barros, Elaine Cristina Marqueze, Raul Machado Neto, Debora Botequio Moretti, Jardelina de Souza Todao Bernardino, Loyze Paola Oliveira de Lima, Luiz Aurelio de Campos Crispin. Centro de Genômica Funcional da ESALQ: Luiz Lehmann Coutinho, Ricardo Augusto Brassaloti, Raquel de Lello Rocha Campos Cassano. NGS Soluções Genômicas: Pilar Drummond Sampaio Corrêa Mariani. FZEA-USP Pirassununga: Mirele Daiana Poleti, Jessika Cristina Chagas Lesbon, Elisangela Chicaroni Mattos, Heidge Fukumasu. USP-Botucatu: Rejane Maria Tommasini Grotto, Jayme A. Souza-Neto, Guilherme Targino Valente, Patricia Akemi Assato, Felipe Allan da Silva da Costa, Bianca Cechetto Carlos. Mendelics: Bibiana Santos, João Paulo Kitajima, Erika Freitas, David Schlesinger. Hemocentro Ribeirão Preto: Simone Kashima, Evandra Strazza Rodrigues, Svetoslav Nanev Slavov, Elaine Vieira dos Santos, Rafael dos Santos Bezerra, Luiz Carlos Junior de Alcantara, Marta Giovanetti, Vagner Fonseca, Flavia Aburjaile, Rodrigo Tocantins Calado. FAMERP-SJRP: Cecília Artico Banho, Lívia Sacchetto, Fábio Sossai Possebon, Leila Sabrina Ullmann, Cíntia Bittar, Guilherme Campos, Helena Lage Ferreira, Jorge A. Petroli Marchesi, Maisa C. Pereira Parra, Marília Moraes, Paula Rahal, Paulo Inacio da Costa, João Pessoa Araújo Jr., Maurício Lacerda Nogueira. Prefeitura de Sao Paulo: Melissa Palmieri. |
| EPI_ISL_1966769 | HOSPITAL REGIONAL DE ITAPETININGA | Instituto Butantan / Mendelics | Instituto Butantan: Dimas Tadeu Covas, Sandra Coccuzzo Sampaio, Maria Carolina Elias, José Salvatore Leister Patané, Vincent Louis Viala, Antonio Jorge Martins, Ricardo Haddad, Claudia Renata dos Santos Barros, Elaine Cristina Marqueze, Raul Machado Neto, Debora Botequio Moretti, Jardelina de Souza Todao Bernardino, Loyze Paola Oliveira de Lima, Luiz Aurelio de Campos Crispin. Centro de Genômica Funcional da ESALQ: Luiz Lehmann Coutinho, Ricardo Augusto Brassaloti, Raquel de Lello Rocha Campos Cassano. NGS Soluções Genômicas: Pilar Drummond Sampaio Corrêa Mariani. FZEA-USP Pirassununga: Mirele Daiana Poleti, Jessika Cristina Chagas Lesbon, Elisangela Chicaroni Mattos, Heidge Fukumasu. USP-Botucatu: Rejane Maria Tommasini Grotto, Jayme A. Souza-Neto, Guilherme Targino Valente, Patricia Akemi Assato, Felipe Allan da Silva da Costa, Bianca Cechetto Carlos. Mendelics: Bibiana Santos, João Paulo Kitajima, Erika Freitas, David Schlesinger. Hemocentro Ribeirão Preto: Simone Kashima, Evandra Strazza Rodrigues, Svetoslav Nanev Slavov, Elaine Vieira dos Santos, Rafael dos Santos Bezerra, Luiz Carlos Junior de Alcantara, Marta Giovanetti, Vagner Fonseca, Flavia Aburjaile, Rodrigo Tocantins Calado. FAMERP-SJRP: Cecília Artico Banho, Lívia Sacchetto, Fábio Sossai Possebon, Leila Sabrina Ullmann, Cíntia Bittar, Guilherme Campos, Helena Lage Ferreira, Jorge A. Petroli Marchesi, Maisa C. Pereira Parra, Marília Moraes, Paula Rahal, Paulo Inacio da Costa, João Pessoa Araújo Jr., Maurício Lacerda Nogueira. Prefeitura de Sao Paulo: Melissa Palmieri. |
| EPI_ISL_1966771 | SECRETARIA MUNICIPAL DA SAUDE DE ITAPEVA SP | Instituto Butantan / Mendelics | Instituto Butantan: Dimas Tadeu Covas, Sandra Coccuzzo Sampaio, Maria Carolina Elias, José Salvatore Leister Patané, Vincent Louis Viala, Antonio Jorge Martins, Ricardo Haddad, Claudia Renata dos Santos Barros, Elaine Cristina Marqueze, Raul Machado Neto, Debora Botequio Moretti, Jardelina de Souza Todao Bernardino, Loyze Paola Oliveira de Lima, Luiz Aurelio de Campos Crispin. Centro de Genômica Funcional da ESALQ: Luiz Lehmann Coutinho, Ricardo Augusto Brassaloti, Raquel de Lello Rocha Campos Cassano. NGS Soluções Genômicas: Pilar Drummond Sampaio Corrêa Mariani. FZEA-USP Pirassununga: Mirele Daiana Poleti, Jessika Cristina Chagas Lesbon, Elisangela Chicaroni Mattos, Heidge Fukumasu. USP-Botucatu: Rejane Maria Tommasini Grotto, Jayme A. Souza-Neto, Guilherme Targino Valente, Patricia Akemi Assato, Felipe Allan da Silva da Costa, Bianca Cechetto Carlos. Mendelics: Bibiana Santos, João Paulo Kitajima, Erika Freitas, David Schlesinger. Hemocentro Ribeirão Preto: Simone Kashima, Evandra Strazza Rodrigues, Svetoslav Nanev Slavov, Elaine Vieira dos Santos, Rafael dos Santos Bezerra, Luiz Carlos Junior de Alcantara, Marta Giovanetti, Vagner Fonseca, Flavia Aburjaile, Rodrigo Tocantins Calado. FAMERP-SJRP: Cecília Artico Banho, Lívia Sacchetto, Fábio Sossai Possebon, Leila Sabrina Ullmann, Cíntia Bittar, Guilherme Campos, Helena Lage Ferreira, Jorge A. Petroli Marchesi, Maisa C. Pereira Parra, Marília Moraes, Paula Rahal, Paulo Inacio da Costa, João Pessoa Araújo Jr., Maurício Lacerda Nogueira. Prefeitura de Sao Paulo: Melissa Palmieri. |
| EPI_ISL_1966772 | SECRETARIA MUNICIPAL DA SAUDE DE ITARARE | Instituto Butantan / Mendelics | Instituto Butantan: Dimas Tadeu Covas, Sandra Coccuzzo Sampaio, Maria Carolina Elias, José Salvatore Leister Patané, Vincent Louis Viala, Antonio Jorge Martins, Ricardo Haddad, Claudia Renata dos Santos Barros, Elaine Cristina Marqueze, Raul Machado Neto, Debora Botequio Moretti, Jardelina de |

| | | | |
|---|---|---|---|
| | | | Souza Todao Bernardino, Loyze Paola Oliveira de Lima, Luiz Aurelio de Campos Crispin. Centro de Genômica Funcional da ESALQ: Luiz Lehmann Coutinho, Ricardo Augusto Brassaloti, Raquel de Lello Rocha Campos Cassano. NGS Soluções Genômicas: Pilar Drummond Sampaio Corrêa Mariani. FZEA-USP Pirassununga: Mirele Daiana Poleti, Jessika Cristina Chagas Lesbon, Elisangela Chicaroni Mattos, Heidge Fukumasu. USP-Botucatu: Rejane Maria Tommasini Grotto, Jayme A. Souza-Neto, Guilherme Targino Valente, Patricia Akemi Assato, Felipe Allan da Silva da Costa, Bianca Cechetto Carlos. Mendelics: Bibiana Santos, João Paulo Kitajima, Erika Freitas, David Schlesinger. Hemocentro Ribeirão Preto: Simone Kashima, Evandra Strazza Rodrigues, Svetoslav Nanev Slavov, Elaine Vieira dos Santos, Rafael dos Santos Bezerra, Luiz Carlos Junior de Alcantara, Marta Giovanetti, Vagner Fonseca, Flavia Aburjaile, Rodrigo Tocantins Calado. FAMERP-SJRP: Cecília Artico Banho, Lívia Sacchetto, Fábio Sossai Possebon, Leila Sabrina Ullmann, Cíntia Bittar, Guilherme Campos, Helena Lage Ferreira, Jorge A. Petroli Marchesi, Maisa C. Pereira Parra, Marília Moraes, Paula Rahal, Paulo Inacio da Costa, João Pessoa Araújo Jr., Maurício Lacerda Nogueira. Prefeitura de Sao Paulo: Melissa Palmieri. |
| EPI_ISL_1966773, EPI_ISL_1966774, EPI_ISL_1966775 | VIGILANCIA EPIDEMIOLOGICA JARDINOPOLIS SP | Instituto Butantan / Mendelics | Instituto Butantan: Dimas Tadeu Covas, Sandra Coccuzzo Sampaio, Maria Carolina Elias, José Salvatore Leister Patané, Vincent Louis Viala, Antonio Jorge Martins, Ricardo Haddad, Claudia Renata dos Santos Barros, Elaine Cristina Marqueze, Raul Machado Neto, Debora Botequio Moretti, Jardelina de Souza Todao Bernardino, Loyze Paola Oliveira de Lima, Luiz Aurelio de Campos Crispin. Centro de Genômica Funcional da ESALQ: Luiz Lehmann Coutinho, Ricardo Augusto Brassaloti, Raquel de Lello Rocha Campos Cassano. NGS Soluções Genômicas: Pilar Drummond Sampaio Corrêa Mariani. FZEA-USP Pirassununga: Mirele Daiana Poleti, Jessika Cristina Chagas Lesbon, Elisangela Chicaroni Mattos, Heidge Fukumasu. USP-Botucatu: Rejane Maria Tommasini Grotto, Jayme A. Souza-Neto, Guilherme Targino Valente, Patricia Akemi Assato, Felipe Allan da Silva da Costa, Bianca Cechetto Carlos. Mendelics: Bibiana Santos, João Paulo Kitajima, Erika Freitas, David Schlesinger. Hemocentro Ribeirão Preto: Simone Kashima, Evandra Strazza Rodrigues, Svetoslav Nanev Slavov, Elaine Vieira dos Santos, Rafael dos Santos Bezerra, Luiz Carlos Junior de Alcantara, Marta Giovanetti, Vagner Fonseca, Flavia Aburjaile, Rodrigo Tocantins Calado. FAMERP-SJRP: Cecília Artico Banho, Lívia Sacchetto, Fábio Sossai Possebon, Leila Sabrina Ullmann, Cíntia Bittar, Guilherme Campos, Helena Lage Ferreira, Jorge A. Petroli Marchesi, Maisa C. Pereira Parra, Marília Moraes, Paula Rahal, Paulo Inacio da Costa, João Pessoa Araújo Jr., Maurício Lacerda Nogueira. Prefeitura de Sao Paulo: Melissa Palmieri. |
| EPI_ISL_1966776 | AMB MEDICO DR FRANCISCO FELIX DE MENDONCA SAUDADE | Instituto Butantan / Mendelics | Instituto Butantan: Dimas Tadeu Covas, Sandra Coccuzzo Sampaio, Maria Carolina Elias, José Salvatore Leister Patané, Vincent Louis Viala, Antonio Jorge Martins, Ricardo Haddad, Claudia Renata dos Santos Barros, Elaine Cristina Marqueze, Raul Machado Neto, Debora Botequio Moretti, Jardelina de Souza Todao Bernardino, Loyze Paola Oliveira de Lima, Luiz Aurelio de Campos Crispin. Centro de Genômica Funcional da ESALQ: Luiz Lehmann Coutinho, Ricardo Augusto Brassaloti, Raquel de Lello Rocha Campos Cassano. NGS Soluções Genômicas: Pilar Drummond Sampaio Corrêa Mariani. FZEA-USP Pirassununga: Mirele Daiana Poleti, Jessika Cristina Chagas Lesbon, Elisangela Chicaroni Mattos, Heidge Fukumasu. USP-Botucatu: Rejane Maria Tommasini Grotto, Jayme A. Souza-Neto, Guilherme Targino Valente, Patricia Akemi Assato, Felipe Allan da Silva da Costa, Bianca Cechetto Carlos. Mendelics: Bibiana Santos, João Paulo Kitajima, Erika Freitas, David Schlesinger. Hemocentro Ribeirão Preto: Simone Kashima, Evandra Strazza Rodrigues, Svetoslav Nanev Slavov, Elaine Vieira dos Santos, Rafael dos Santos Bezerra, Luiz Carlos Junior de Alcantara, Marta Giovanetti, Vagner Fonseca, Flavia Aburjaile, Rodrigo Tocantins Calado. FAMERP-SJRP: Cecília Artico Banho, Lívia Sacchetto, Fábio Sossai Possebon, Leila Sabrina Ullmann, Cíntia Bittar, Guilherme Campos, Helena Lage Ferreira, Jorge A. Petroli Marchesi, Maisa C. Pereira Parra, Marília Moraes, Paula Rahal, Paulo Inacio da Costa, João Pessoa Araújo Jr., Maurício Lacerda Nogueira. Prefeitura de Sao Paulo: Melissa Palmieri. |
| EPI_ISL_1966777, EPI_ISL_1966778, EPI_ISL_1966779, EPI_ISL_1966780, EPI_ISL_1966781 | AMBULATORIO DR JOAO LANIA CENTRO JOSE BONIFACIO | Instituto Butantan / Mendelics | Instituto Butantan: Dimas Tadeu Covas, Sandra Coccuzzo Sampaio, Maria Carolina Elias, José Salvatore Leister Patané, Vincent Louis Viala, Antonio Jorge Martins, Ricardo Haddad, Claudia Renata dos Santos Barros, Elaine Cristina Marqueze, Raul Machado Neto, Debora Botequio Moretti, Jardelina de Souza Todao Bernardino, Loyze Paola Oliveira de Lima, Luiz Aurelio de Campos Crispin. Centro de Genômica Funcional da ESALQ: Luiz Lehmann Coutinho, Ricardo Augusto Brassaloti, Raquel de Lello Rocha Campos Cassano. NGS Soluções Genômicas: Pilar Drummond Sampaio Corrêa Mariani. FZEA-USP Pirassununga: Mirele Daiana Poleti, Jessika Cristina Chagas Lesbon, Elisangela Chicaroni Mattos, Heidge Fukumasu. USP-Botucatu: Rejane Maria Tommasini Grotto, Jayme A. Souza-Neto, Guilherme Targino Valente, Patricia Akemi Assato, Felipe Allan da Silva da Costa, Bianca Cechetto Carlos. Mendelics: Bibiana Santos, João Paulo Kitajima, Erika Freitas, David Schlesinger. Hemocentro Ribeirão Preto: Simone Kashima, Evandra Strazza Rodrigues, Svetoslav Nanev Slavov, Elaine Vieira dos Santos, Rafael dos Santos Bezerra, Luiz Carlos Junior de Alcantara, Marta Giovanetti, Vagner Fonseca, Flavia Aburjaile, Rodrigo Tocantins Calado. FAMERP-SJRP: Cecília Artico Banho, Lívia Sacchetto, Fábio Sossai Possebon, Leila Sabrina Ullmann, Cíntia Bittar, Guilherme Campos, Helena Lage Ferreira, Jorge A. Petroli Marchesi, Maisa C. Pereira Parra, Marília Moraes, Paula Rahal, Paulo Inacio da Costa, João Pessoa Araújo Jr., Maurício Lacerda Nogueira. Prefeitura de Sao Paulo: Melissa Palmieri. |
| EPI_ISL_1966782, EPI_ISL_1966783 | UBS DE MENDONCA | Instituto Butantan / Mendelics | Instituto Butantan: Dimas Tadeu Covas, Sandra Coccuzzo Sampaio, Maria Carolina Elias, José Salvatore Leister Patané, Vincent Louis Viala, Antonio Jorge Martins, Ricardo Haddad, Claudia Renata dos Santos Barros, Elaine Cristina Marqueze, Raul Machado Neto, Debora Botequio Moretti, Jardelina de Souza Todao Bernardino, Loyze Paola Oliveira de Lima, Luiz Aurelio de Campos Crispin. Centro de Genômica Funcional da ESALQ: Luiz Lehmann Coutinho, Ricardo Augusto Brassaloti, Raquel de Lello Rocha Campos Cassano. NGS Soluções Genômicas: Pilar Drummond Sampaio Corrêa Mariani. FZEA-USP Pirassununga: Mirele Daiana Poleti, Jessika Cristina Chagas Lesbon, Elisangela Chicaroni Mattos, Heidge Fukumasu. USP-Botucatu: Rejane Maria Tommasini Grotto, Jayme A. Souza-Neto, Guilherme Targino Valente, Patricia Akemi Assato, Felipe Allan da Silva da Costa, Bianca Cechetto Carlos. Mendelics: Bibiana Santos, João Paulo Kitajima, Erika Freitas, David Schlesinger. Hemocentro Ribeirão Preto: Simone Kashima, Evandra Strazza Rodrigues, Svetoslav Nanev Slavov, Elaine Vieira dos Santos, Rafael dos Santos Bezerra, Luiz Carlos Junior de Alcantara, Marta Giovanetti, Vagner Fonseca, Flavia Aburjaile, Rodrigo Tocantins Calado. FAMERP-SJRP: Cecília Artico Banho, Lívia Sacchetto, Fábio Sossai Possebon, Leila Sabrina Ullmann, Cíntia Bittar, Guilherme Campos, Helena Lage Ferreira, Jorge A. Petroli Marchesi, Maisa C. Pereira Parra, Marília Moraes, Paula Rahal, Paulo Inacio da Costa, João Pessoa Araújo Jr., Maurício Lacerda Nogueira. Prefeitura de Sao Paulo: Melissa Palmieri. |
| EPI_ISL_1966784, EPI_ISL_1966785 | CS DE MIRASSOLANDIA | Instituto Butantan / Mendelics | Instituto Butantan: Dimas Tadeu Covas, Sandra Coccuzzo Sampaio, Maria Carolina Elias, José Salvatore Leister Patané, Vincent Louis Viala, Antonio Jorge Martins, Ricardo Haddad, Claudia Renata dos Santos Barros, Elaine Cristina Marqueze, Raul Machado Neto, Debora Botequio Moretti, Jardelina de Souza Todao Bernardino, Loyze Paola Oliveira de Lima, Luiz Aurelio de Campos Crispin. Centro de Genômica Funcional da ESALQ: Luiz Lehmann Coutinho, Ricardo Augusto Brassaloti, Raquel de Lello Rocha Campos Cassano. NGS Soluções Genômicas: Pilar Drummond Sampaio Corrêa Mariani. FZEA-USP Pirassununga: Mirele Daiana Poleti, Jessika Cristina Chagas Lesbon, Elisangela Chicaroni Mattos, Heidge Fukumasu. USP-Botucatu: Rejane Maria Tommasini Grotto, Jayme A. Souza-Neto, Guilherme Targino Valente, Patricia Akemi Assato, Felipe Allan da Silva da Costa, Bianca Cechetto Carlos. Mendelics: Bibiana Santos, João Paulo Kitajima, Erika Freitas, David Schlesinger. Hemocentro Ribeirão Preto: Simone Kashima, Evandra Strazza Rodrigues, Svetoslav Nanev Slavov, Elaine Vieira dos Santos, Rafael dos Santos Bezerra, Luiz Carlos Junior de Alcantara, Marta Giovanetti, Vagner Fonseca, Flavia Aburjaile, Rodrigo Tocantins Calado. FAMERP-SJRP: Cecília Artico Banho, Lívia Sacchetto, Fábio Sossai Possebon, Leila Sabrina Ullmann, Cíntia Bittar, Guilherme Campos, Helena Lage Ferreira, Jorge A. Petroli Marchesi, Maisa C. Pereira Parra, Marília Moraes, Paula Rahal, Paulo Inacio da Costa, João Pessoa Araújo Jr., Maurício Lacerda Nogueira. Prefeitura de Sao Paulo: Melissa Palmieri. |
| EPI_ISL_1966786 | PSF SETOR LESTE NHANDEARA | Instituto Butantan / Mendelics | Instituto Butantan: Dimas Tadeu Covas, Sandra Coccuzzo Sampaio, Maria Carolina Elias, José Salvatore Leister Patané, Vincent Louis Viala, Antonio Jorge Martins, Ricardo Haddad, Claudia Renata dos Santos Barros, Elaine Cristina Marqueze, Raul Machado Neto, Debora Botequio Moretti, Jardelina de Souza Todao Bernardino, Loyze Paola Oliveira de Lima, Luiz Aurelio de Campos Crispin. Centro de Genômica Funcional da ESALQ: Luiz Lehmann Coutinho, Ricardo Augusto Brassaloti, Raquel de Lello Rocha Campos Cassano. NGS Soluções Genômicas: Pilar Drummond Sampaio Corrêa Mariani. FZEA-USP Pirassununga: Mirele Daiana Poleti, Jessika Cristina Chagas Lesbon, Elisangela Chicaroni Mattos, Heidge Fukumasu. USP-Botucatu: Rejane Maria Tommasini Grotto, Jayme A. Souza-Neto, Guilherme Targino Valente, Patricia Akemi Assato, Felipe Allan da Silva da Costa, Bianca Cechetto Carlos. Mendelics: Bibiana Santos, João Paulo Kitajima, Erika Freitas, David Schlesinger. Hemocentro Ribeirão Preto: Simone Kashima, Evandra Strazza Rodrigues, Svetoslav Nanev Slavov, Elaine Vieira dos Santos, Rafael dos Santos Bezerra, Luiz Carlos Junior de Alcantara, Marta Giovanetti, Vagner Fonseca, Flavia Aburjaile, Rodrigo Tocantins Calado. FAMERP-SJRP: Cecília Artico Banho, Lívia Sacchetto, Fábio Sossai Possebon, Leila Sabrina Ullmann, Cíntia Bittar, Guilherme Campos, Helena Lage Ferreira, Jorge A. Petroli Marchesi, Maisa C. Pereira Parra, Marília Moraes, Paula Rahal, Paulo Inacio da Costa, João Pessoa Araújo Jr., Maurício Lacerda Nogueira. Prefeitura de Sao Paulo: Melissa Palmieri. |
| EPI_ISL_1966787, EPI_ISL_1966788, EPI_ISL_1966789, EPI_ISL_1966790, EPI_ISL_1966791 | CS DE NIPOA | Instituto Butantan / Mendelics | Instituto Butantan: Dimas Tadeu Covas, Sandra Coccuzzo Sampaio, Maria Carolina Elias, José Salvatore Leister Patané, Vincent Louis Viala, Antonio Jorge Martins, Ricardo Haddad, Claudia Renata dos Santos Barros, Elaine Cristina Marqueze, Raul Machado Neto, Debora Botequio Moretti, Jardelina de Souza Todao Bernardino, Loyze Paola Oliveira de Lima, Luiz Aurelio de Campos Crispin. Centro de Genômica Funcional da ESALQ: Luiz Lehmann Coutinho, Ricardo Augusto Brassaloti, Raquel de Lello Rocha Campos Cassano. NGS Soluções Genômicas: Pilar Drummond Sampaio Corrêa Mariani. FZEA-USP Pirassununga: Mirele Daiana Poleti, Jessika Cristina Chagas Lesbon, Elisangela Chicaroni Mattos, Heidge Fukumasu. USP-Botucatu: Rejane Maria Tommasini Grotto, Jayme A. Souza-Neto, Guilherme Targino Valente, Patricia Akemi Assato, Felipe Allan da Silva da Costa, Bianca Cechetto Carlos. Mendelics: Bibiana Santos, João Paulo Kitajima, Erika Freitas, David Schlesinger. Hemocentro Ribeirão Preto: Simone Kashima, Evandra Strazza Rodrigues, Svetoslav Nanev Slavov, Elaine Vieira dos Santos, Rafael dos Santos Bezerra, Luiz Carlos Junior de Alcantara, Marta Giovanetti, Vagner Fonseca, Flavia Aburjaile, Rodrigo Tocantins Calado. FAMERP-SJRP: Cecília Artico Banho, Lívia Sacchetto, Fábio Sossai Possebon, Leila Sabrina |

| | | | |
|---|---|---|---|
| | | | Ullmann, Cíntia Bittar, Guilherme Campos, Helena Lage Ferreira, Jorge A. Petroli Marchesi, Maisa C. Pereira Parra, Marília Moraes, Paula Rahal, Paulo Inacio da Costa, João Pessoa Araújo Jr., Maurício Lacerda Nogueira. Prefeitura de Sao Paulo: Melissa Palmieri. |
| EPI_ISL_1966792, EPI_ISL_1966793, EPI_ISL_1966794 | CENTRO DE SAUDE DE OSVALDO CRUZ | Instituto Butantan / Mendelics | Instituto Butantan: Dimas Tadeu Covas, Sandra Coccuzzo Sampaio, Maria Carolina Elias, José Salvatore Leister Patané, Vincent Louis Viala, Antonio Jorge Martins, Ricardo Haddad, Claudia Renata dos Santos Barros, Elaine Cristina Marqueze, Raul Machado Neto, Debora Botequio Moretti, Jardelina de Souza Todao Bernardino, Loyze Paola Oliveira de Lima, Luiz Aurelio de Campos Crispin. Centro de Genômica Funcional da ESALQ: Luiz Lehmann Coutinho, Ricardo Augusto Brassaloti, Raquel de Lello Rocha Campos Cassano. NGS Soluções Genômicas: Pilar Drummond Sampaio Corrêa Mariani. FZEA-USP Pirassununga: Mirele Daiana Poleti, Jessika Cristina Chagas Lesbon, Elisangela Chicaroni Mattos, Heidge Fukumasu. USP-Botucatu: Rejane Maria Tommasini Grotto, Jayme A. Souza-Neto, Guilherme Targino Valente, Patricia Akemi Assato, Felipe Allan da Silva da Costa, Bianca Cechetto Carlos. Mendelics: Bibiana Santos, João Paulo Kitajima, Erika Freitas, David Schlesinger. Hemocentro Ribeirão Preto: Simone Kashima, Evandra Strazza Rodrigues, Svetoslav Nanev Slavov, Elaine Vieira dos Santos, Rafael dos Santos Bezerra, Luiz Carlos Junior de Alcantara, Marta Giovanetti, Vagner Fonseca, Flavia Aburjaile, Rodrigo Tocantins Calado. FAMERP-SJRP: Cecília Artico Banho, Lívia Sacchetto, Fábio Sossai Possebon, Leila Sabrina Ullmann, Cíntia Bittar, Guilherme Campos, Helena Lage Ferreira, Jorge A. Petroli Marchesi, Maisa C. Pereira Parra, Marília Moraes, Paula Rahal, Paulo Inacio da Costa, João Pessoa Araújo Jr., Maurício Lacerda Nogueira. Prefeitura de Sao Paulo: Melissa Palmieri. |
| EPI_ISL_1966795 | CS DE PALESTINA | Instituto Butantan / Mendelics | Instituto Butantan: Dimas Tadeu Covas, Sandra Coccuzzo Sampaio, Maria Carolina Elias, José Salvatore Leister Patané, Vincent Louis Viala, Antonio Jorge Martins, Ricardo Haddad, Claudia Renata dos Santos Barros, Elaine Cristina Marqueze, Raul Machado Neto, Debora Botequio Moretti, Jardelina de Souza Todao Bernardino, Loyze Paola Oliveira de Lima, Luiz Aurelio de Campos Crispin. Centro de Genômica Funcional da ESALQ: Luiz Lehmann Coutinho, Ricardo Augusto Brassaloti, Raquel de Lello Rocha Campos Cassano. NGS Soluções Genômicas: Pilar Drummond Sampaio Corrêa Mariani. FZEA-USP Pirassununga: Mirele Daiana Poleti, Jessika Cristina Chagas Lesbon, Elisangela Chicaroni Mattos, Heidge Fukumasu. USP-Botucatu: Rejane Maria Tommasini Grotto, Jayme A. Souza-Neto, Guilherme Targino Valente, Patricia Akemi Assato, Felipe Allan da Silva da Costa, Bianca Cechetto Carlos. Mendelics: Bibiana Santos, João Paulo Kitajima, Erika Freitas, David Schlesinger. Hemocentro Ribeirão Preto: Simone Kashima, Evandra Strazza Rodrigues, Svetoslav Nanev Slavov, Elaine Vieira dos Santos, Rafael dos Santos Bezerra, Luiz Carlos Junior de Alcantara, Marta Giovanetti, Vagner Fonseca, Flavia Aburjaile, Rodrigo Tocantins Calado. FAMERP-SJRP: Cecília Artico Banho, Lívia Sacchetto, Fábio Sossai Possebon, Leila Sabrina Ullmann, Cíntia Bittar, Guilherme Campos, Helena Lage Ferreira, Jorge A. Petroli Marchesi, Maisa C. Pereira Parra, Marília Moraes, Paula Rahal, Paulo Inacio da Costa, João Pessoa Araújo Jr., Maurício Lacerda Nogueira. Prefeitura de Sao Paulo: Melissa Palmieri. |
| EPI_ISL_1966796 | CENTRO DE SAUDE TEREZINHA DE MORAES ARSILLA | Instituto Butantan / Mendelics | Instituto Butantan: Dimas Tadeu Covas, Sandra Coccuzzo Sampaio, Maria Carolina Elias, José Salvatore Leister Patané, Vincent Louis Viala, Antonio Jorge Martins, Ricardo Haddad, Claudia Renata dos Santos Barros, Elaine Cristina Marqueze, Raul Machado Neto, Debora Botequio Moretti, Jardelina de Souza Todao Bernardino, Loyze Paola Oliveira de Lima, Luiz Aurelio de Campos Crispin. Centro de Genômica Funcional da ESALQ: Luiz Lehmann Coutinho, Ricardo Augusto Brassaloti, Raquel de Lello Rocha Campos Cassano. NGS Soluções Genômicas: Pilar Drummond Sampaio Corrêa Mariani. FZEA-USP Pirassununga: Mirele Daiana Poleti, Jessika Cristina Chagas Lesbon, Elisangela Chicaroni Mattos, Heidge Fukumasu. USP-Botucatu: Rejane Maria Tommasini Grotto, Jayme A. Souza-Neto, Guilherme Targino Valente, Patricia Akemi Assato, Felipe Allan da Silva da Costa, Bianca Cechetto Carlos. Mendelics: Bibiana Santos, João Paulo Kitajima, Erika Freitas, David Schlesinger. Hemocentro Ribeirão Preto: Simone Kashima, Evandra Strazza Rodrigues, Svetoslav Nanev Slavov, Elaine Vieira dos Santos, Rafael dos Santos Bezerra, Luiz Carlos Junior de Alcantara, Marta Giovanetti, Vagner Fonseca, Flavia Aburjaile, Rodrigo Tocantins Calado. FAMERP-SJRP: Cecília Artico Banho, Lívia Sacchetto, Fábio Sossai Possebon, Leila Sabrina Ullmann, Cíntia Bittar, Guilherme Campos, Helena Lage Ferreira, Jorge A. Petroli Marchesi, Maisa C. Pereira Parra, Marília Moraes, Paula Rahal, Paulo Inacio da Costa, João Pessoa Araújo Jr., Maurício Lacerda Nogueira. Prefeitura de Sao Paulo: Melissa Palmieri. |
| EPI_ISL_1966797, EPI_ISL_1966798, EPI_ISL_1966799 | PSF DR ANTONIO PIRES DE ALMEIDA PORTO FELIZ | Instituto Butantan / Mendelics | Instituto Butantan: Dimas Tadeu Covas, Sandra Coccuzzo Sampaio, Maria Carolina Elias, José Salvatore Leister Patané, Vincent Louis Viala, Antonio Jorge Martins, Ricardo Haddad, Claudia Renata dos Santos Barros, Elaine Cristina Marqueze, Raul Machado Neto, Debora Botequio Moretti, Jardelina de Souza Todao Bernardino, Loyze Paola Oliveira de Lima, Luiz Aurelio de Campos Crispin. Centro de Genômica Funcional da ESALQ: Luiz Lehmann Coutinho, Ricardo Augusto Brassaloti, Raquel de Lello Rocha Campos Cassano. NGS Soluções Genômicas: Pilar Drummond Sampaio Corrêa Mariani. FZEA-USP Pirassununga: Mirele Daiana Poleti, Jessika Cristina Chagas Lesbon, Elisangela Chicaroni Mattos, Heidge Fukumasu. USP-Botucatu: Rejane Maria Tommasini Grotto, Jayme A. Souza-Neto, Guilherme Targino Valente, Patricia Akemi Assato, Felipe Allan da Silva da Costa, Bianca Cechetto Carlos. Mendelics: Bibiana Santos, João Paulo Kitajima, Erika Freitas, David Schlesinger. Hemocentro Ribeirão Preto: Simone Kashima, Evandra Strazza Rodrigues, Svetoslav Nanev Slavov, Elaine Vieira dos Santos, Rafael dos Santos Bezerra, Luiz Carlos Junior de Alcantara, Marta Giovanetti, Vagner Fonseca, Flavia Aburjaile, Rodrigo Tocantins Calado. FAMERP-SJRP: Cecília Artico Banho, Lívia Sacchetto, Fábio Sossai Possebon, Leila Sabrina Ullmann, Cíntia Bittar, Guilherme Campos, Helena Lage Ferreira, Jorge A. Petroli Marchesi, Maisa C. Pereira Parra, Marília Moraes, Paula Rahal, Paulo Inacio da Costa, João Pessoa Araújo Jr., Maurício Lacerda Nogueira. Prefeitura de Sao Paulo: Melissa Palmieri. |
| EPI_ISL_1966800 | UNIDADE DE PRONTO ATENDIMENTO UPA | Instituto Butantan / Mendelics | Instituto Butantan: Dimas Tadeu Covas, Sandra Coccuzzo Sampaio, Maria Carolina Elias, José Salvatore Leister Patané, Vincent Louis Viala, Antonio Jorge Martins, Ricardo Haddad, Claudia Renata dos Santos Barros, Elaine Cristina Marqueze, Raul Machado Neto, Debora Botequio Moretti, Jardelina de Souza Todao Bernardino, Loyze Paola Oliveira de Lima, Luiz Aurelio de Campos Crispin. Centro de Genômica Funcional da ESALQ: Luiz Lehmann Coutinho, Ricardo Augusto Brassaloti, Raquel de Lello Rocha Campos Cassano. NGS Soluções Genômicas: Pilar Drummond Sampaio Corrêa Mariani. FZEA-USP Pirassununga: Mirele Daiana Poleti, Jessika Cristina Chagas Lesbon, Elisangela Chicaroni Mattos, Heidge Fukumasu. USP-Botucatu: Rejane Maria Tommasini Grotto, Jayme A. Souza-Neto, Guilherme Targino Valente, Patricia Akemi Assato, Felipe Allan da Silva da Costa, Bianca Cechetto Carlos. Mendelics: Bibiana Santos, João Paulo Kitajima, Erika Freitas, David Schlesinger. Hemocentro Ribeirão Preto: Simone Kashima, Evandra Strazza Rodrigues, Svetoslav Nanev Slavov, Elaine Vieira dos Santos, Rafael dos Santos Bezerra, Luiz Carlos Junior de Alcantara, Marta Giovanetti, Vagner Fonseca, Flavia Aburjaile, Rodrigo Tocantins Calado. FAMERP-SJRP: Cecília Artico Banho, Lívia Sacchetto, Fábio Sossai Possebon, Leila Sabrina Ullmann, Cíntia Bittar, Guilherme Campos, Helena Lage Ferreira, Jorge A. Petroli Marchesi, Maisa C. Pereira Parra, Marília Moraes, Paula Rahal, Paulo Inacio da Costa, João Pessoa Araújo Jr., Maurício Lacerda Nogueira. Prefeitura de Sao Paulo: Melissa Palmieri. |
| EPI_ISL_1966801, EPI_ISL_1966802, EPI_ISL_1966803, EPI_ISL_1966804, EPI_ISL_1966805, EPI_ISL_1966806 | CENTRO DE SAUDE DE RIBEIRAO DO SUL | Instituto Butantan / Mendelics | Instituto Butantan: Dimas Tadeu Covas, Sandra Coccuzzo Sampaio, Maria Carolina Elias, José Salvatore Leister Patané, Vincent Louis Viala, Antonio Jorge Martins, Ricardo Haddad, Claudia Renata dos Santos Barros, Elaine Cristina Marqueze, Raul Machado Neto, Debora Botequio Moretti, Jardelina de Souza Todao Bernardino, Loyze Paola Oliveira de Lima, Luiz Aurelio de Campos Crispin. Centro de Genômica Funcional da ESALQ: Luiz Lehmann Coutinho, Ricardo Augusto Brassaloti, Raquel de Lello Rocha Campos Cassano. NGS Soluções Genômicas: Pilar Drummond Sampaio Corrêa Mariani. FZEA-USP Pirassununga: Mirele Daiana Poleti, Jessika Cristina Chagas Lesbon, Elisangela Chicaroni Mattos, Heidge Fukumasu. USP-Botucatu: Rejane Maria Tommasini Grotto, Jayme A. Souza-Neto, Guilherme Targino Valente, Patricia Akemi Assato, Felipe Allan da Silva da Costa, Bianca Cechetto Carlos. Mendelics: Bibiana Santos, João Paulo Kitajima, Erika Freitas, David Schlesinger. Hemocentro Ribeirão Preto: Simone Kashima, Evandra Strazza Rodrigues, Svetoslav Nanev Slavov, Elaine Vieira dos Santos, Rafael dos Santos Bezerra, Luiz Carlos Junior de Alcantara, Marta Giovanetti, Vagner Fonseca, Flavia Aburjaile, Rodrigo Tocantins Calado. FAMERP-SJRP: Cecília Artico Banho, Lívia Sacchetto, Fábio Sossai Possebon, Leila Sabrina Ullmann, Cíntia Bittar, Guilherme Campos, Helena Lage Ferreira, Jorge A. Petroli Marchesi, Maisa C. Pereira Parra, Marília Moraes, Paula Rahal, Paulo Inacio da Costa, João Pessoa Araújo Jr., Maurício Lacerda Nogueira. Prefeitura de Sao Paulo: Melissa Palmieri. |
| EPI_ISL_1966807 | UBS DR WALDOMIRO FERREIRA NEVES SCRPARDO | Instituto Butantan / Mendelics | Instituto Butantan: Dimas Tadeu Covas, Sandra Coccuzzo Sampaio, Maria Carolina Elias, José Salvatore Leister Patané, Vincent Louis Viala, Antonio Jorge Martins, Ricardo Haddad, Claudia Renata dos Santos Barros, Elaine Cristina Marqueze, Raul Machado Neto, Debora Botequio Moretti, Jardelina de Souza Todao Bernardino, Loyze Paola Oliveira de Lima, Luiz Aurelio de Campos Crispin. Centro de Genômica Funcional da ESALQ: Luiz Lehmann Coutinho, Ricardo Augusto Brassaloti, Raquel de Lello Rocha Campos Cassano. NGS Soluções Genômicas: Pilar Drummond Sampaio Corrêa Mariani. FZEA-USP Pirassununga: Mirele Daiana Poleti, Jessika Cristina Chagas Lesbon, Elisangela Chicaroni Mattos, Heidge Fukumasu. USP-Botucatu: Rejane Maria Tommasini Grotto, Jayme A. Souza-Neto, Guilherme Targino Valente, Patricia Akemi Assato, Felipe Allan da Silva da Costa, Bianca Cechetto Carlos. Mendelics: Bibiana Santos, João Paulo Kitajima, Erika Freitas, David Schlesinger. Hemocentro Ribeirão Preto: Simone Kashima, Evandra Strazza Rodrigues, Svetoslav Nanev Slavov, Elaine Vieira dos Santos, Rafael dos Santos Bezerra, Luiz Carlos Junior de Alcantara, Marta Giovanetti, Vagner Fonseca, Flavia Aburjaile, Rodrigo Tocantins Calado. FAMERP-SJRP: Cecília Artico Banho, Lívia Sacchetto, Fábio Sossai Possebon, Leila Sabrina Ullmann, Cíntia Bittar, Guilherme Campos, Helena Lage Ferreira, Jorge A. Petroli Marchesi, Maisa C. Pereira Parra, Marília Moraes, Paula Rahal, Paulo Inacio da Costa, João Pessoa Araújo Jr., Maurício Lacerda Nogueira. Prefeitura de Sao Paulo: Melissa Palmieri. |
| EPI_ISL_1966808 | CS II DR JOSE CARQUEIJO SCRPARDO | Instituto Butantan / Mendelics | Instituto Butantan: Dimas Tadeu Covas, Sandra Coccuzzo Sampaio, Maria Carolina Elias, José Salvatore Leister Patané, Vincent Louis Viala, Antonio Jorge Martins, Ricardo Haddad, Claudia Renata dos Santos Barros, Elaine Cristina Marqueze, Raul Machado Neto, Debora Botequio Moretti, Jardelina de Souza Todao Bernardino, Loyze Paola Oliveira de Lima, Luiz Aurelio de Campos Crispin. Centro de Genômica Funcional da ESALQ: Luiz Lehmann Coutinho, Ricardo Augusto Brassaloti, Raquel de Lello Rocha Campos Cassano. NGS Soluções Genômicas: Pilar Drummond Sampaio Corrêa Mariani. |

| | | | |
|---|---|---|---|
| | | | FZEA-USP Pirassununga: Mirele Daiana Poleti, Jessika Cristina Chagas Lesbon, Elisangela Chicaroni Mattos, Heidge Fukumasu. USP-Botucatu: Rejane Maria Tommasini Grotto, Jayme A. Souza-Neto, Guilherme Targino Valente, Patricia Akemi Assato, Felipe Allan da Silva da Costa, Bianca Cechetto Carlos. Mendelics: Bibiana Santos, João Paulo Kitajima, Erika Freitas, David Schlesinger. Hemocentro Ribeirão Preto: Simone Kashima, Evandra Strazza Rodrigues, Svetoslav Nanev Slavov, Elaine Vieira dos Santos, Rafael dos Santos Bezerra, Luiz Carlos Junior de Alcantara, Marta Giovanetti, Vagner Fonseca, Flavia Aburjaile, Rodrigo Tocantins Calado. FAMERP-SJRP: Cecília Artico Banho, Lívia Sacchetto, Fábio Sossai Possebon, Leila Sabrina Ullmann, Cíntia Bittar, Guilherme Campos, Helena Lage Ferreira, Jorge A. Petroli Marchesi, Maisa C. Pereira Parra, Marília Moraes, Paula Rahal, Paulo Inacio da Costa, João Pessoa Araújo Jr., Maurício Lacerda Nogueira. Prefeitura de Sao Paulo: Melissa Palmieri. |
| EPI_ISL_1966809 | UBS DR ABELARDO PINHEIRO GUIMARAES SCRPARDO | Instituto Butantan / Mendelics | Instituto Butantan: Dimas Tadeu Covas, Sandra Coccuzzo Sampaio, Maria Carolina Elias, José Salvatore Leister Patané, Vincent Louis Viala, Antonio Jorge Martins, Ricardo Haddad, Claudia Renata dos Santos Barros, Elaine Cristina Marqueze, Raul Machado Neto, Debora Botequio Moretti, Jardelina de Souza Todao Bernardino, Loyze Paola Oliveira de Lima, Luiz Aurelio de Campos Crispin. Centro de Genômica Funcional da ESALQ: Luiz Lehmann Coutinho, Ricardo Augusto Brassaloti, Raquel de Lello Rocha Campos Cassano. NGS Soluções Genômicas: Pilar Drummond Sampaio Corrêa Mariani. FZEA-USP Pirassununga: Mirele Daiana Poleti, Jessika Cristina Chagas Lesbon, Elisangela Chicaroni Mattos, Heidge Fukumasu. USP-Botucatu: Rejane Maria Tommasini Grotto, Jayme A. Souza-Neto, Guilherme Targino Valente, Patricia Akemi Assato, Felipe Allan da Silva da Costa, Bianca Cechetto Carlos. Mendelics: Bibiana Santos, João Paulo Kitajima, Erika Freitas, David Schlesinger. Hemocentro Ribeirão Preto: Simone Kashima, Evandra Strazza Rodrigues, Svetoslav Nanev Slavov, Elaine Vieira dos Santos, Rafael dos Santos Bezerra, Luiz Carlos Junior de Alcantara, Marta Giovanetti, Vagner Fonseca, Flavia Aburjaile, Rodrigo Tocantins Calado. FAMERP-SJRP: Cecília Artico Banho, Lívia Sacchetto, Fábio Sossai Possebon, Leila Sabrina Ullmann, Cíntia Bittar, Guilherme Campos, Helena Lage Ferreira, Jorge A. Petroli Marchesi, Maisa C. Pereira Parra, Marília Moraes, Paula Rahal, Paulo Inacio da Costa, João Pessoa Araújo Jr., Maurício Lacerda Nogueira. Prefeitura de Sao Paulo: Melissa Palmieri. |
| EPI_ISL_1966810 | USF PARQUE DAS NACOES SCRPARDO | Instituto Butantan / Mendelics | Instituto Butantan: Dimas Tadeu Covas, Sandra Coccuzzo Sampaio, Maria Carolina Elias, José Salvatore Leister Patané, Vincent Louis Viala, Antonio Jorge Martins, Ricardo Haddad, Claudia Renata dos Santos Barros, Elaine Cristina Marqueze, Raul Machado Neto, Debora Botequio Moretti, Jardelina de Souza Todao Bernardino, Loyze Paola Oliveira de Lima, Luiz Aurelio de Campos Crispin. Centro de Genômica Funcional da ESALQ: Luiz Lehmann Coutinho, Ricardo Augusto Brassaloti, Raquel de Lello Rocha Campos Cassano. NGS Soluções Genômicas: Pilar Drummond Sampaio Corrêa Mariani. FZEA-USP Pirassununga: Mirele Daiana Poleti, Jessika Cristina Chagas Lesbon, Elisangela Chicaroni Mattos, Heidge Fukumasu. USP-Botucatu: Rejane Maria Tommasini Grotto, Jayme A. Souza-Neto, Guilherme Targino Valente, Patricia Akemi Assato, Felipe Allan da Silva da Costa, Bianca Cechetto Carlos. Mendelics: Bibiana Santos, João Paulo Kitajima, Erika Freitas, David Schlesinger. Hemocentro Ribeirão Preto: Simone Kashima, Evandra Strazza Rodrigues, Svetoslav Nanev Slavov, Elaine Vieira dos Santos, Rafael dos Santos Bezerra, Luiz Carlos Junior de Alcantara, Marta Giovanetti, Vagner Fonseca, Flavia Aburjaile, Rodrigo Tocantins Calado. FAMERP-SJRP: Cecília Artico Banho, Lívia Sacchetto, Fábio Sossai Possebon, Leila Sabrina Ullmann, Cíntia Bittar, Guilherme Campos, Helena Lage Ferreira, Jorge A. Petroli Marchesi, Maisa C. Pereira Parra, Marília Moraes, Paula Rahal, Paulo Inacio da Costa, João Pessoa Araújo Jr., Maurício Lacerda Nogueira. Prefeitura de Sao Paulo: Melissa Palmieri. |
| EPI_ISL_1966811 | UBS DR ABELARDO PINHEIRO GUIMARAES SCRPARDO | Instituto Butantan / Mendelics | Instituto Butantan: Dimas Tadeu Covas, Sandra Coccuzzo Sampaio, Maria Carolina Elias, José Salvatore Leister Patané, Vincent Louis Viala, Antonio Jorge Martins, Ricardo Haddad, Claudia Renata dos Santos Barros, Elaine Cristina Marqueze, Raul Machado Neto, Debora Botequio Moretti, Jardelina de Souza Todao Bernardino, Loyze Paola Oliveira de Lima, Luiz Aurelio de Campos Crispin. Centro de Genômica Funcional da ESALQ: Luiz Lehmann Coutinho, Ricardo Augusto Brassaloti, Raquel de Lello Rocha Campos Cassano. NGS Soluções Genômicas: Pilar Drummond Sampaio Corrêa Mariani. FZEA-USP Pirassununga: Mirele Daiana Poleti, Jessika Cristina Chagas Lesbon, Elisangela Chicaroni Mattos, Heidge Fukumasu. USP-Botucatu: Rejane Maria Tommasini Grotto, Jayme A. Souza-Neto, Guilherme Targino Valente, Patricia Akemi Assato, Felipe Allan da Silva da Costa, Bianca Cechetto Carlos. Mendelics: Bibiana Santos, João Paulo Kitajima, Erika Freitas, David Schlesinger. Hemocentro Ribeirão Preto: Simone Kashima, Evandra Strazza Rodrigues, Svetoslav Nanev Slavov, Elaine Vieira dos Santos, Rafael dos Santos Bezerra, Luiz Carlos Junior de Alcantara, Marta Giovanetti, Vagner Fonseca, Flavia Aburjaile, Rodrigo Tocantins Calado. FAMERP-SJRP: Cecília Artico Banho, Lívia Sacchetto, Fábio Sossai Possebon, Leila Sabrina Ullmann, Cíntia Bittar, Guilherme Campos, Helena Lage Ferreira, Jorge A. Petroli Marchesi, Maisa C. Pereira Parra, Marília Moraes, Paula Rahal, Paulo Inacio da Costa, João Pessoa Araújo Jr., Maurício Lacerda Nogueira. Prefeitura de Sao Paulo: Melissa Palmieri. |
| EPI_ISL_1966813 | UBS DR WALDOMIRO FERREIRA NEVES SCRPARDO | Instituto Butantan / Mendelics | Instituto Butantan: Dimas Tadeu Covas, Sandra Coccuzzo Sampaio, Maria Carolina Elias, José Salvatore Leister Patané, Vincent Louis Viala, Antonio Jorge Martins, Ricardo Haddad, Claudia Renata dos Santos Barros, Elaine Cristina Marqueze, Raul Machado Neto, Debora Botequio Moretti, Jardelina de Souza Todao Bernardino, Loyze Paola Oliveira de Lima, Luiz Aurelio de Campos Crispin. Centro de Genômica Funcional da ESALQ: Luiz Lehmann Coutinho, Ricardo Augusto Brassaloti, Raquel de Lello Rocha Campos Cassano. NGS Soluções Genômicas: Pilar Drummond Sampaio Corrêa Mariani. FZEA-USP Pirassununga: Mirele Daiana Poleti, Jessika Cristina Chagas Lesbon, Elisangela Chicaroni Mattos, Heidge Fukumasu. USP-Botucatu: Rejane Maria Tommasini Grotto, Jayme A. Souza-Neto, Guilherme Targino Valente, Patricia Akemi Assato, Felipe Allan da Silva da Costa, Bianca Cechetto Carlos. Mendelics: Bibiana Santos, João Paulo Kitajima, Erika Freitas, David Schlesinger. Hemocentro Ribeirão Preto: Simone Kashima, Evandra Strazza Rodrigues, Svetoslav Nanev Slavov, Elaine Vieira dos Santos, Rafael dos Santos Bezerra, Luiz Carlos Junior de Alcantara, Marta Giovanetti, Vagner Fonseca, Flavia Aburjaile, Rodrigo Tocantins Calado. FAMERP-SJRP: Cecília Artico Banho, Lívia Sacchetto, Fábio Sossai Possebon, Leila Sabrina Ullmann, Cíntia Bittar, Guilherme Campos, Helena Lage Ferreira, Jorge A. Petroli Marchesi, Maisa C. Pereira Parra, Marília Moraes, Paula Rahal, Paulo Inacio da Costa, João Pessoa Araújo Jr., Maurício Lacerda Nogueira. Prefeitura de Sao Paulo: Melissa Palmieri. |
| EPI_ISL_1966814, EPI_ISL_1966815 | CENTRO DE SAUDE II SAO MIGUEL ARCANJO | Instituto Butantan / Mendelics | Instituto Butantan: Dimas Tadeu Covas, Sandra Coccuzzo Sampaio, Maria Carolina Elias, José Salvatore Leister Patané, Vincent Louis Viala, Antonio Jorge Martins, Ricardo Haddad, Claudia Renata dos Santos Barros, Elaine Cristina Marqueze, Raul Machado Neto, Debora Botequio Moretti, Jardelina de Souza Todao Bernardino, Loyze Paola Oliveira de Lima, Luiz Aurelio de Campos Crispin. Centro de Genômica Funcional da ESALQ: Luiz Lehmann Coutinho, Ricardo Augusto Brassaloti, Raquel de Lello Rocha Campos Cassano. NGS Soluções Genômicas: Pilar Drummond Sampaio Corrêa Mariani. FZEA-USP Pirassununga: Mirele Daiana Poleti, Jessika Cristina Chagas Lesbon, Elisangela Chicaroni Mattos, Heidge Fukumasu. USP-Botucatu: Rejane Maria Tommasini Grotto, Jayme A. Souza-Neto, Guilherme Targino Valente, Patricia Akemi Assato, Felipe Allan da Silva da Costa, Bianca Cechetto Carlos. Mendelics: Bibiana Santos, João Paulo Kitajima, Erika Freitas, David Schlesinger. Hemocentro Ribeirão Preto: Simone Kashima, Evandra Strazza Rodrigues, Svetoslav Nanev Slavov, Elaine Vieira dos Santos, Rafael dos Santos Bezerra, Luiz Carlos Junior de Alcantara, Marta Giovanetti, Vagner Fonseca, Flavia Aburjaile, Rodrigo Tocantins Calado. FAMERP-SJRP: Cecília Artico Banho, Lívia Sacchetto, Fábio Sossai Possebon, Leila Sabrina Ullmann, Cíntia Bittar, Guilherme Campos, Helena Lage Ferreira, Jorge A. Petroli Marchesi, Maisa C. Pereira Parra, Marília Moraes, Paula Rahal, Paulo Inacio da Costa, João Pessoa Araújo Jr., Maurício Lacerda Nogueira. Prefeitura de Sao Paulo: Melissa Palmieri. |
| EPI_ISL_1966816 | AMBULATORIO COVID 19 | Instituto Butantan / Mendelics | Instituto Butantan: Dimas Tadeu Covas, Sandra Coccuzzo Sampaio, Maria Carolina Elias, José Salvatore Leister Patané, Vincent Louis Viala, Antonio Jorge Martins, Ricardo Haddad, Claudia Renata dos Santos Barros, Elaine Cristina Marqueze, Raul Machado Neto, Debora Botequio Moretti, Jardelina de Souza Todao Bernardino, Loyze Paola Oliveira de Lima, Luiz Aurelio de Campos Crispin. Centro de Genômica Funcional da ESALQ: Luiz Lehmann Coutinho, Ricardo Augusto Brassaloti, Raquel de Lello Rocha Campos Cassano. NGS Soluções Genômicas: Pilar Drummond Sampaio Corrêa Mariani. FZEA-USP Pirassununga: Mirele Daiana Poleti, Jessika Cristina Chagas Lesbon, Elisangela Chicaroni Mattos, Heidge Fukumasu. USP-Botucatu: Rejane Maria Tommasini Grotto, Jayme A. Souza-Neto, Guilherme Targino Valente, Patricia Akemi Assato, Felipe Allan da Silva da Costa, Bianca Cechetto Carlos. Mendelics: Bibiana Santos, João Paulo Kitajima, Erika Freitas, David Schlesinger. Hemocentro Ribeirão Preto: Simone Kashima, Evandra Strazza Rodrigues, Svetoslav Nanev Slavov, Elaine Vieira dos Santos, Rafael dos Santos Bezerra, Luiz Carlos Junior de Alcantara, Marta Giovanetti, Vagner Fonseca, Flavia Aburjaile, Rodrigo Tocantins Calado. FAMERP-SJRP: Cecília Artico Banho, Lívia Sacchetto, Fábio Sossai Possebon, Leila Sabrina Ullmann, Cíntia Bittar, Guilherme Campos, Helena Lage Ferreira, Jorge A. Petroli Marchesi, Maisa C. Pereira Parra, Marília Moraes, Paula Rahal, Paulo Inacio da Costa, João Pessoa Araújo Jr., Maurício Lacerda Nogueira. Prefeitura de Sao Paulo: Melissa Palmieri. |
| EPI_ISL_1966817 | UNIDADE BASICA DE SAUDE DE TARUMA | Instituto Butantan / Mendelics | Instituto Butantan: Dimas Tadeu Covas, Sandra Coccuzzo Sampaio, Maria Carolina Elias, José Salvatore Leister Patané, Vincent Louis Viala, Antonio Jorge Martins, Ricardo Haddad, Claudia Renata dos Santos Barros, Elaine Cristina Marqueze, Raul Machado Neto, Debora Botequio Moretti, Jardelina de Souza Todao Bernardino, Loyze Paola Oliveira de Lima, Luiz Aurelio de Campos Crispin. Centro de Genômica Funcional da ESALQ: Luiz Lehmann Coutinho, Ricardo Augusto Brassaloti, Raquel de Lello Rocha Campos Cassano. NGS Soluções Genômicas: Pilar Drummond Sampaio Corrêa Mariani. FZEA-USP Pirassununga: Mirele Daiana Poleti, Jessika Cristina Chagas Lesbon, Elisangela Chicaroni Mattos, Heidge Fukumasu. USP-Botucatu: Rejane Maria Tommasini Grotto, Jayme A. Souza-Neto, Guilherme Targino Valente, Patricia Akemi Assato, Felipe Allan da Silva da Costa, Bianca Cechetto Carlos. Mendelics: Bibiana Santos, João Paulo Kitajima, Erika Freitas, David Schlesinger. Hemocentro Ribeirão Preto: Simone Kashima, Evandra Strazza Rodrigues, Svetoslav Nanev Slavov, Elaine Vieira dos Santos, Rafael dos Santos Bezerra, Luiz Carlos Junior de Alcantara, Marta Giovanetti, Vagner Fonseca, Flavia Aburjaile, Rodrigo Tocantins Calado. FAMERP-SJRP: Cecília Artico Banho, Lívia Sacchetto, Fábio Sossai Possebon, Leila Sabrina Ullmann, Cíntia Bittar, Guilherme Campos, Helena Lage Ferreira, Jorge A. Petroli Marchesi, Maisa C. Pereira Parra, Marília Moraes, Paula Rahal, Paulo Inacio da Costa, João Pessoa Araújo Jr., Maurício Lacerda Nogueira. Prefeitura de Sao Paulo: Melissa Palmieri. |

| | | | |
|---|---|---|---|
| EPI_ISL_1966820 | CENTRO DE SAUDE II TIETE DR RUI SILVEIRA MELO | Instituto Butantan / Mendelics | Instituto Butantan: Dimas Tadeu Covas, Sandra Coccuzzo Sampaio, Maria Carolina Elias, José Salvatore Leister Patané, Vincent Louis Viala, Antonio Jorge Martins, Ricardo Haddad, Claudia Renata dos Santos Barros, Elaine Cristina Marqueze, Raul Machado Neto, Debora Botequio Moretti, Jardelina de Souza Todao Bernardino, Loyze Paola Oliveira de Lima, Luiz Aurelio de Campos Crispin. Centro de Genômica Funcional da ESALQ: Luiz Lehmann Coutinho, Ricardo Augusto Brassaloti, Raquel de Lello Rocha Campos Cassano. NGS Soluções Genômicas: Pilar Drummond Sampaio Corrêa Mariani. FZEA-USP Pirassununga: Mirele Daiana Poleti, Jessika Cristina Chagas Lesbon, Elisangela Chicaroni Mattos, Heidge Fukumasu. USP-Botucatu: Rejane Maria Tommasini Grotto, Jayme A. Souza-Neto, Guilherme Targino Valente, Patricia Akemi Assato, Felipe Allan da Silva da Costa, Bianca Cechetto Carlos. Mendelics: Bibiana Santos, João Paulo Kitajima, Erika Freitas, David Schlesinger. Hemocentro Ribeirão Preto: Simone Kashima, Evandra Strazza Rodrigues, Svetoslav Nanev Slavov, Elaine Vieira dos Santos, Rafael dos Santos Bezerra, Luiz Carlos Junior de Alcantara, Marta Giovanetti, Vagner Fonseca, Flavia Aburjaile, Rodrigo Tocantins Calado. FAMERP-SJRP: Cecília Artico Banho, Lívia Sacchetto, Fábio Sossai Possebon, Leila Sabrina Ullmann, Cíntia Bittar, Guilherme Campos, Helena Lage Ferreira, Jorge A. Petroli Marchesi, Maisa C. Pereira Parra, Marília Moraes, Paula Rahal, Paulo Inacio da Costa, João Pessoa Araújo Jr., Maurício Lacerda Nogueira. Prefeitura de Sao Paulo: Melissa Palmieri. |
| EPI_ISL_1966822 | UNIDADE DE SAUDE DA FAMILIA DE UBIRAJARA | Instituto Butantan / Mendelics | Instituto Butantan: Dimas Tadeu Covas, Sandra Coccuzzo Sampaio, Maria Carolina Elias, José Salvatore Leister Patané, Vincent Louis Viala, Antonio Jorge Martins, Ricardo Haddad, Claudia Renata dos Santos Barros, Elaine Cristina Marqueze, Raul Machado Neto, Debora Botequio Moretti, Jardelina de Souza Todao Bernardino, Loyze Paola Oliveira de Lima, Luiz Aurelio de Campos Crispin. Centro de Genômica Funcional da ESALQ: Luiz Lehmann Coutinho, Ricardo Augusto Brassaloti, Raquel de Lello Rocha Campos Cassano. NGS Soluções Genômicas: Pilar Drummond Sampaio Corrêa Mariani. FZEA-USP Pirassununga: Mirele Daiana Poleti, Jessika Cristina Chagas Lesbon, Elisangela Chicaroni Mattos, Heidge Fukumasu. USP-Botucatu: Rejane Maria Tommasini Grotto, Jayme A. Souza-Neto, Guilherme Targino Valente, Patricia Akemi Assato, Felipe Allan da Silva da Costa, Bianca Cechetto Carlos. Mendelics: Bibiana Santos, João Paulo Kitajima, Erika Freitas, David Schlesinger. Hemocentro Ribeirão Preto: Simone Kashima, Evandra Strazza Rodrigues, Svetoslav Nanev Slavov, Elaine Vieira dos Santos, Rafael dos Santos Bezerra, Luiz Carlos Junior de Alcantara, Marta Giovanetti, Vagner Fonseca, Flavia Aburjaile, Rodrigo Tocantins Calado. FAMERP-SJRP: Cecília Artico Banho, Lívia Sacchetto, Fábio Sossai Possebon, Leila Sabrina Ullmann, Cíntia Bittar, Guilherme Campos, Helena Lage Ferreira, Jorge A. Petroli Marchesi, Maisa C. Pereira Parra, Marília Moraes, Paula Rahal, Paulo Inacio da Costa, João Pessoa Araújo Jr., Maurício Lacerda Nogueira. Prefeitura de Sao Paulo: Melissa Palmieri. |
| EPI_ISL_1966823 | PRONTO ATENDIMENTO MUNICIPAL ALUMINIO | Instituto Butantan / ESALQ-USP (Piracicaba) | Instituto Butantan: Dimas Tadeu Covas, Sandra Coccuzzo Sampaio, Maria Carolina Elias, José Salvatore Leister Patané, Vincent Louis Viala, Antonio Jorge Martins, Ricardo Haddad, Claudia Renata dos Santos Barros, Elaine Cristina Marqueze, Raul Machado Neto, Debora Botequio Moretti, Jardelina de Souza Todao Bernardino, Loyze Paola Oliveira de Lima, Luiz Aurelio de Campos Crispin. Centro de Genômica Funcional da ESALQ: Luiz Lehmann Coutinho, Ricardo Augusto Brassaloti, Raquel de Lello Rocha Campos Cassano. NGS Soluções Genômicas: Pilar Drummond Sampaio Corrêa Mariani. FZEA-USP Pirassununga: Mirele Daiana Poleti, Jessika Cristina Chagas Lesbon, Elisangela Chicaroni Mattos, Heidge Fukumasu. USP-Botucatu: Rejane Maria Tommasini Grotto, Jayme A. Souza-Neto, Guilherme Targino Valente, Patricia Akemi Assato, Felipe Allan da Silva da Costa, Bianca Cechetto Carlos. Mendelics: Bibiana Santos, João Paulo Kitajima, Erika Freitas, David Schlesinger. Hemocentro Ribeirão Preto: Simone Kashima, Evandra Strazza Rodrigues, Svetoslav Nanev Slavov, Elaine Vieira dos Santos, Rafael dos Santos Bezerra, Luiz Carlos Junior de Alcantara, Marta Giovanetti, Vagner Fonseca, Flavia Aburjaile, Rodrigo Tocantins Calado. FAMERP-SJRP: Cecília Artico Banho, Lívia Sacchetto, Fábio Sossai Possebon, Leila Sabrina Ullmann, Cíntia Bittar, Guilherme Campos, Helena Lage Ferreira, Jorge A. Petroli Marchesi, Maisa C. Pereira Parra, Marília Moraes, Paula Rahal, Paulo Inacio da Costa, João Pessoa Araújo Jr., Maurício Lacerda Nogueira. Prefeitura de Sao Paulo: Melissa Palmieri. |
| EPI_ISL_1966824, EPI_ISL_1966825, EPI_ISL_1966826, EPI_ISL_1966827, EPI_ISL_1966828, EPI_ISL_1966829 | CENTRO DE SAUDE DR RENATO DE CARVALHO RIBEIRO ANGATUBA | Instituto Butantan / ESALQ-USP (Piracicaba) | Instituto Butantan: Dimas Tadeu Covas, Sandra Coccuzzo Sampaio, Maria Carolina Elias, José Salvatore Leister Patané, Vincent Louis Viala, Antonio Jorge Martins, Ricardo Haddad, Claudia Renata dos Santos Barros, Elaine Cristina Marqueze, Raul Machado Neto, Debora Botequio Moretti, Jardelina de Souza Todao Bernardino, Loyze Paola Oliveira de Lima, Luiz Aurelio de Campos Crispin. Centro de Genômica Funcional da ESALQ: Luiz Lehmann Coutinho, Ricardo Augusto Brassaloti, Raquel de Lello Rocha Campos Cassano. NGS Soluções Genômicas: Pilar Drummond Sampaio Corrêa Mariani. FZEA-USP Pirassununga: Mirele Daiana Poleti, Jessika Cristina Chagas Lesbon, Elisangela Chicaroni Mattos, Heidge Fukumasu. USP-Botucatu: Rejane Maria Tommasini Grotto, Jayme A. Souza-Neto, Guilherme Targino Valente, Patricia Akemi Assato, Felipe Allan da Silva da Costa, Bianca Cechetto Carlos. Mendelics: Bibiana Santos, João Paulo Kitajima, Erika Freitas, David Schlesinger. Hemocentro Ribeirão Preto: Simone Kashima, Evandra Strazza Rodrigues, Svetoslav Nanev Slavov, Elaine Vieira dos Santos, Rafael dos Santos Bezerra, Luiz Carlos Junior de Alcantara, Marta Giovanetti, Vagner Fonseca, Flavia Aburjaile, Rodrigo Tocantins Calado. FAMERP-SJRP: Cecília Artico Banho, Lívia Sacchetto, Fábio Sossai Possebon, Leila Sabrina Ullmann, Cíntia Bittar, Guilherme Campos, Helena Lage Ferreira, Jorge A. Petroli Marchesi, Maisa C. Pereira Parra, Marília Moraes, Paula Rahal, Paulo Inacio da Costa, João Pessoa Araújo Jr., Maurício Lacerda Nogueira. Prefeitura de Sao Paulo: Melissa Palmieri. |
| EPI_ISL_1966830, EPI_ISL_1966831 | VIGILANCIA EPIDIMIOLOGICA DE ARACARIGUAMA | Instituto Butantan / ESALQ-USP (Piracicaba) | Instituto Butantan: Dimas Tadeu Covas, Sandra Coccuzzo Sampaio, Maria Carolina Elias, José Salvatore Leister Patané, Vincent Louis Viala, Antonio Jorge Martins, Ricardo Haddad, Claudia Renata dos Santos Barros, Elaine Cristina Marqueze, Raul Machado Neto, Debora Botequio Moretti, Jardelina de Souza Todao Bernardino, Loyze Paola Oliveira de Lima, Luiz Aurelio de Campos Crispin. Centro de Genômica Funcional da ESALQ: Luiz Lehmann Coutinho, Ricardo Augusto Brassaloti, Raquel de Lello Rocha Campos Cassano. NGS Soluções Genômicas: Pilar Drummond Sampaio Corrêa Mariani. FZEA-USP Pirassununga: Mirele Daiana Poleti, Jessika Cristina Chagas Lesbon, Elisangela Chicaroni Mattos, Heidge Fukumasu. USP-Botucatu: Rejane Maria Tommasini Grotto, Jayme A. Souza-Neto, Guilherme Targino Valente, Patricia Akemi Assato, Felipe Allan da Silva da Costa, Bianca Cechetto Carlos. Mendelics: Bibiana Santos, João Paulo Kitajima, Erika Freitas, David Schlesinger. Hemocentro Ribeirão Preto: Simone Kashima, Evandra Strazza Rodrigues, Svetoslav Nanev Slavov, Elaine Vieira dos Santos, Rafael dos Santos Bezerra, Luiz Carlos Junior de Alcantara, Marta Giovanetti, Vagner Fonseca, Flavia Aburjaile, Rodrigo Tocantins Calado. FAMERP-SJRP: Cecília Artico Banho, Lívia Sacchetto, Fábio Sossai Possebon, Leila Sabrina Ullmann, Cíntia Bittar, Guilherme Campos, Helena Lage Ferreira, Jorge A. Petroli Marchesi, Maisa C. Pereira Parra, Marília Moraes, Paula Rahal, Paulo Inacio da Costa, João Pessoa Araújo Jr., Maurício Lacerda Nogueira. Prefeitura de Sao Paulo: Melissa Palmieri. |
| EPI_ISL_1966833 | CENTRO DE SAUDE I AVARE | Instituto Butantan / ESALQ-USP (Piracicaba) | Instituto Butantan: Dimas Tadeu Covas, Sandra Coccuzzo Sampaio, Maria Carolina Elias, José Salvatore Leister Patané, Vincent Louis Viala, Antonio Jorge Martins, Ricardo Haddad, Claudia Renata dos Santos Barros, Elaine Cristina Marqueze, Raul Machado Neto, Debora Botequio Moretti, Jardelina de Souza Todao Bernardino, Loyze Paola Oliveira de Lima, Luiz Aurelio de Campos Crispin. Centro de Genômica Funcional da ESALQ: Luiz Lehmann Coutinho, Ricardo Augusto Brassaloti, Raquel de Lello Rocha Campos Cassano. NGS Soluções Genômicas: Pilar Drummond Sampaio Corrêa Mariani. FZEA-USP Pirassununga: Mirele Daiana Poleti, Jessika Cristina Chagas Lesbon, Elisangela Chicaroni Mattos, Heidge Fukumasu. USP-Botucatu: Rejane Maria Tommasini Grotto, Jayme A. Souza-Neto, Guilherme Targino Valente, Patricia Akemi Assato, Felipe Allan da Silva da Costa, Bianca Cechetto Carlos. Mendelics: Bibiana Santos, João Paulo Kitajima, Erika Freitas, David Schlesinger. Hemocentro Ribeirão Preto: Simone Kashima, Evandra Strazza Rodrigues, Svetoslav Nanev Slavov, Elaine Vieira dos Santos, Rafael dos Santos Bezerra, Luiz Carlos Junior de Alcantara, Marta Giovanetti, Vagner Fonseca, Flavia Aburjaile, Rodrigo Tocantins Calado. FAMERP-SJRP: Cecília Artico Banho, Lívia Sacchetto, Fábio Sossai Possebon, Leila Sabrina Ullmann, Cíntia Bittar, Guilherme Campos, Helena Lage Ferreira, Jorge A. Petroli Marchesi, Maisa C. Pereira Parra, Marília Moraes, Paula Rahal, Paulo Inacio da Costa, João Pessoa Araújo Jr., Maurício Lacerda Nogueira. Prefeitura de Sao Paulo: Melissa Palmieri. |
| EPI_ISL_1966834 | CS DE BALSAMO | Instituto Butantan / ESALQ-USP (Piracicaba) | Instituto Butantan: Dimas Tadeu Covas, Sandra Coccuzzo Sampaio, Maria Carolina Elias, José Salvatore Leister Patané, Vincent Louis Viala, Antonio Jorge Martins, Ricardo Haddad, Claudia Renata dos Santos Barros, Elaine Cristina Marqueze, Raul Machado Neto, Debora Botequio Moretti, Jardelina de Souza Todao Bernardino, Loyze Paola Oliveira de Lima, Luiz Aurelio de Campos Crispin. Centro de Genômica Funcional da ESALQ: Luiz Lehmann Coutinho, Ricardo Augusto Brassaloti, Raquel de Lello Rocha Campos Cassano. NGS Soluções Genômicas: Pilar Drummond Sampaio Corrêa Mariani. FZEA-USP Pirassununga: Mirele Daiana Poleti, Jessika Cristina Chagas Lesbon, Elisangela Chicaroni Mattos, Heidge Fukumasu. USP-Botucatu: Rejane Maria Tommasini Grotto, Jayme A. Souza-Neto, Guilherme Targino Valente, Patricia Akemi Assato, Felipe Allan da Silva da Costa, Bianca Cechetto Carlos. Mendelics: Bibiana Santos, João Paulo Kitajima, Erika Freitas, David Schlesinger. Hemocentro Ribeirão Preto: Simone Kashima, Evandra Strazza Rodrigues, Svetoslav Nanev Slavov, Elaine Vieira dos Santos, Rafael dos Santos Bezerra, Luiz Carlos Junior de Alcantara, Marta Giovanetti, Vagner Fonseca, Flavia Aburjaile, Rodrigo Tocantins Calado. FAMERP-SJRP: Cecília Artico Banho, Lívia Sacchetto, Fábio Sossai Possebon, Leila Sabrina Ullmann, Cíntia Bittar, Guilherme Campos, Helena Lage Ferreira, Jorge A. Petroli Marchesi, Maisa C. Pereira Parra, Marília Moraes, Paula Rahal, Paulo Inacio da Costa, João Pessoa Araújo Jr., Maurício Lacerda Nogueira. Prefeitura de Sao Paulo: Melissa Palmieri. |
| EPI_ISL_1966835, EPI_ISL_1966836, EPI_ISL_1966837, EPI_ISL_1966838, EPI_ISL_1966839 | GRUPO TECNICO DE VIGILANCIA SANITARIA E EPIDEMIOLOGICA | Instituto Butantan / ESALQ-USP (Piracicaba) | Instituto Butantan: Dimas Tadeu Covas, Sandra Coccuzzo Sampaio, Maria Carolina Elias, José Salvatore Leister Patané, Vincent Louis Viala, Antonio Jorge Martins, Ricardo Haddad, Claudia Renata dos Santos Barros, Elaine Cristina Marqueze, Raul Machado Neto, Debora Botequio Moretti, Jardelina de Souza Todao Bernardino, Loyze Paola Oliveira de Lima, Luiz Aurelio de Campos Crispin. Centro de Genômica Funcional da ESALQ: Luiz Lehmann Coutinho, Ricardo Augusto Brassaloti, Raquel de Lello Rocha Campos Cassano. NGS Soluções Genômicas: Pilar Drummond Sampaio Corrêa Mariani. FZEA-USP Pirassununga: Mirele Daiana Poleti, Jessika Cristina Chagas Lesbon, Elisangela Chicaroni Mattos, Heidge Fukumasu. USP-Botucatu: Rejane Maria Tommasini Grotto, Jayme A. Souza-Neto, Guilherme Targino Valente, Patricia Akemi Assato, Felipe Allan da Silva da Costa, Bianca Cechetto Carlos. Mendelics: Bibiana Santos, João Paulo Kitajima, Erika Freitas, David Schlesinger. Hemocentro Ribeirão Preto: Simone Kashima, Evandra Strazza |

| | | | |
|---|---|---|---|
| | | | Rodrigues, Svetoslav Nanev Slavov, Elaine Vieira dos Santos, Rafael dos Santos Bezerra, Luiz Carlos Junior de Alcantara, Marta Giovanetti, Vagner Fonseca, Flavia Aburjaile, Rodrigo Tocantins Calado. FAMERP-SJRP: Cecília Artico Banho, Lívia Sacchetto, Fábio Sossai Possebon, Leila Sabrina Ullmann, Cíntia Bittar, Guilherme Campos, Helena Lage Ferreira, Jorge A. Petroli Marchesi, Maisa C. Pereira Parra, Marília Moraes, Paula Rahal, Paulo Inacio da Costa, João Pessoa Araújo Jr., Maurício Lacerda Nogueira. Prefeitura de Sao Paulo: Melissa Palmieri. |
| EPI_ISL_1966840, EPI_ISL_1966841, EPI_ISL_1966842, EPI_ISL_1966843, EPI_ISL_1966844 | SMS SECRETARIA MUNICIPAL DE SAUDE DE BOITUVA | Instituto Butantan / ESALQ-USP (Piracicaba) | Instituto Butantan: Dimas Tadeu Covas, Sandra Coccuzzo Sampaio, Maria Carolina Elias, José Salvatore Leister Patané, Vincent Louis Viala, Antonio Jorge Martins, Ricardo Haddad, Claudia Renata dos Santos Barros, Elaine Cristina Marqueze, Raul Machado Neto, Debora Botequio Moretti, Jardelina de Souza Todao Bernardino, Loyze Paola Oliveira de Lima, Luiz Aurelio de Campos Crispin. Centro de Genômica Funcional da ESALQ: Luiz Lehmann Coutinho, Ricardo Augusto Brassaloti, Raquel de Lello Rocha Campos Cassano. NGS Soluções Genômicas: Pilar Drummond Sampaio Corrêa Mariani. FZEA-USP Pirassununga: Mirele Daiana Poleti, Jessika Cristina Chagas Lesbon, Elisangela Chicaroni Mattos, Heidge Fukumasu. USP-Botucatu: Rejane Maria Tommasini Grotto, Jayme A. Souza-Neto, Guilherme Targino Valente, Patricia Akemi Assato, Felipe Allan da Silva da Costa, Bianca Cechetto Carlos. Mendelics: Bibiana Santos, João Paulo Kitajima, Erika Freitas, David Schlesinger. Hemocentro Ribeirão Preto: Simone Kashima, Evandra Strazza Rodrigues, Svetoslav Nanev Slavov, Elaine Vieira dos Santos, Rafael dos Santos Bezerra, Luiz Carlos Junior de Alcantara, Marta Giovanetti, Vagner Fonseca, Flavia Aburjaile, Rodrigo Tocantins Calado. FAMERP-SJRP: Cecília Artico Banho, Lívia Sacchetto, Fábio Sossai Possebon, Leila Sabrina Ullmann, Cíntia Bittar, Guilherme Campos, Helena Lage Ferreira, Jorge A. Petroli Marchesi, Maisa C. Pereira Parra, Marília Moraes, Paula Rahal, Paulo Inacio da Costa, João Pessoa Araújo Jr., Maurício Lacerda Nogueira. Prefeitura de Sao Paulo: Melissa Palmieri. |
| EPI_ISL_1966845 | PRONTO ATENDIMENTO MUNICIPAL ITALO SANTUCCI | Instituto Butantan / ESALQ-USP (Piracicaba) | Instituto Butantan: Dimas Tadeu Covas, Sandra Coccuzzo Sampaio, Maria Carolina Elias, José Salvatore Leister Patané, Vincent Louis Viala, Antonio Jorge Martins, Ricardo Haddad, Claudia Renata dos Santos Barros, Elaine Cristina Marqueze, Raul Machado Neto, Debora Botequio Moretti, Jardelina de Souza Todao Bernardino, Loyze Paola Oliveira de Lima, Luiz Aurelio de Campos Crispin. Centro de Genômica Funcional da ESALQ: Luiz Lehmann Coutinho, Ricardo Augusto Brassaloti, Raquel de Lello Rocha Campos Cassano. NGS Soluções Genômicas: Pilar Drummond Sampaio Corrêa Mariani. FZEA-USP Pirassununga: Mirele Daiana Poleti, Jessika Cristina Chagas Lesbon, Elisangela Chicaroni Mattos, Heidge Fukumasu. USP-Botucatu: Rejane Maria Tommasini Grotto, Jayme A. Souza-Neto, Guilherme Targino Valente, Patricia Akemi Assato, Felipe Allan da Silva da Costa, Bianca Cechetto Carlos. Mendelics: Bibiana Santos, João Paulo Kitajima, Erika Freitas, David Schlesinger. Hemocentro Ribeirão Preto: Simone Kashima, Evandra Strazza Rodrigues, Svetoslav Nanev Slavov, Elaine Vieira dos Santos, Rafael dos Santos Bezerra, Luiz Carlos Junior de Alcantara, Marta Giovanetti, Vagner Fonseca, Flavia Aburjaile, Rodrigo Tocantins Calado. FAMERP-SJRP: Cecília Artico Banho, Lívia Sacchetto, Fábio Sossai Possebon, Leila Sabrina Ullmann, Cíntia Bittar, Guilherme Campos, Helena Lage Ferreira, Jorge A. Petroli Marchesi, Maisa C. Pereira Parra, Marília Moraes, Paula Rahal, Paulo Inacio da Costa, João Pessoa Araújo Jr., Maurício Lacerda Nogueira. Prefeitura de Sao Paulo: Melissa Palmieri. |
| EPI_ISL_1966846 | HOSPITAL MUNICIPAL REYNALDO GUERRA CAJATI | Instituto Butantan / ESALQ-USP (Piracicaba) | Instituto Butantan: Dimas Tadeu Covas, Sandra Coccuzzo Sampaio, Maria Carolina Elias, José Salvatore Leister Patané, Vincent Louis Viala, Antonio Jorge Martins, Ricardo Haddad, Claudia Renata dos Santos Barros, Elaine Cristina Marqueze, Raul Machado Neto, Debora Botequio Moretti, Jardelina de Souza Todao Bernardino, Loyze Paola Oliveira de Lima, Luiz Aurelio de Campos Crispin. Centro de Genômica Funcional da ESALQ: Luiz Lehmann Coutinho, Ricardo Augusto Brassaloti, Raquel de Lello Rocha Campos Cassano. NGS Soluções Genômicas: Pilar Drummond Sampaio Corrêa Mariani. FZEA-USP Pirassununga: Mirele Daiana Poleti, Jessika Cristina Chagas Lesbon, Elisangela Chicaroni Mattos, Heidge Fukumasu. USP-Botucatu: Rejane Maria Tommasini Grotto, Jayme A. Souza-Neto, Guilherme Targino Valente, Patricia Akemi Assato, Felipe Allan da Silva da Costa, Bianca Cechetto Carlos. Mendelics: Bibiana Santos, João Paulo Kitajima, Erika Freitas, David Schlesinger. Hemocentro Ribeirão Preto: Simone Kashima, Evandra Strazza Rodrigues, Svetoslav Nanev Slavov, Elaine Vieira dos Santos, Rafael dos Santos Bezerra, Luiz Carlos Junior de Alcantara, Marta Giovanetti, Vagner Fonseca, Flavia Aburjaile, Rodrigo Tocantins Calado. FAMERP-SJRP: Cecília Artico Banho, Lívia Sacchetto, Fábio Sossai Possebon, Leila Sabrina Ullmann, Cíntia Bittar, Guilherme Campos, Helena Lage Ferreira, Jorge A. Petroli Marchesi, Maisa C. Pereira Parra, Marília Moraes, Paula Rahal, Paulo Inacio da Costa, João Pessoa Araújo Jr., Maurício Lacerda Nogueira. Prefeitura de Sao Paulo: Melissa Palmieri. |
| EPI_ISL_1966847, EPI_ISL_1966848, EPI_ISL_1966849, EPI_ISL_1966850, EPI_ISL_1966851, EPI_ISL_1966852 | UBS SAO JUDAS | Instituto Butantan / ESALQ-USP (Piracicaba) | Instituto Butantan: Dimas Tadeu Covas, Sandra Coccuzzo Sampaio, Maria Carolina Elias, José Salvatore Leister Patané, Vincent Louis Viala, Antonio Jorge Martins, Ricardo Haddad, Claudia Renata dos Santos Barros, Elaine Cristina Marqueze, Raul Machado Neto, Debora Botequio Moretti, Jardelina de Souza Todao Bernardino, Loyze Paola Oliveira de Lima, Luiz Aurelio de Campos Crispin. Centro de Genômica Funcional da ESALQ: Luiz Lehmann Coutinho, Ricardo Augusto Brassaloti, Raquel de Lello Rocha Campos Cassano. NGS Soluções Genômicas: Pilar Drummond Sampaio Corrêa Mariani. FZEA-USP Pirassununga: Mirele Daiana Poleti, Jessika Cristina Chagas Lesbon, Elisangela Chicaroni Mattos, Heidge Fukumasu. USP-Botucatu: Rejane Maria Tommasini Grotto, Jayme A. Souza-Neto, Guilherme Targino Valente, Patricia Akemi Assato, Felipe Allan da Silva da Costa, Bianca Cechetto Carlos. Mendelics: Bibiana Santos, João Paulo Kitajima, Erika Freitas, David Schlesinger. Hemocentro Ribeirão Preto: Simone Kashima, Evandra Strazza Rodrigues, Svetoslav Nanev Slavov, Elaine Vieira dos Santos, Rafael dos Santos Bezerra, Luiz Carlos Junior de Alcantara, Marta Giovanetti, Vagner Fonseca, Flavia Aburjaile, Rodrigo Tocantins Calado. FAMERP-SJRP: Cecília Artico Banho, Lívia Sacchetto, Fábio Sossai Possebon, Leila Sabrina Ullmann, Cíntia Bittar, Guilherme Campos, Helena Lage Ferreira, Jorge A. Petroli Marchesi, Maisa C. Pereira Parra, Marília Moraes, Paula Rahal, Paulo Inacio da Costa, João Pessoa Araújo Jr., Maurício Lacerda Nogueira. Prefeitura de Sao Paulo: Melissa Palmieri. |
| EPI_ISL_1966853 | ESF ROSA ROSSI MAIA | Instituto Butantan / ESALQ-USP (Piracicaba) | Instituto Butantan: Dimas Tadeu Covas, Sandra Coccuzzo Sampaio, Maria Carolina Elias, José Salvatore Leister Patané, Vincent Louis Viala, Antonio Jorge Martins, Ricardo Haddad, Claudia Renata dos Santos Barros, Elaine Cristina Marqueze, Raul Machado Neto, Debora Botequio Moretti, Jardelina de Souza Todao Bernardino, Loyze Paola Oliveira de Lima, Luiz Aurelio de Campos Crispin. Centro de Genômica Funcional da ESALQ: Luiz Lehmann Coutinho, Ricardo Augusto Brassaloti, Raquel de Lello Rocha Campos Cassano. NGS Soluções Genômicas: Pilar Drummond Sampaio Corrêa Mariani. FZEA-USP Pirassununga: Mirele Daiana Poleti, Jessika Cristina Chagas Lesbon, Elisangela Chicaroni Mattos, Heidge Fukumasu. USP-Botucatu: Rejane Maria Tommasini Grotto, Jayme A. Souza-Neto, Guilherme Targino Valente, Patricia Akemi Assato, Felipe Allan da Silva da Costa, Bianca Cechetto Carlos. Mendelics: Bibiana Santos, João Paulo Kitajima, Erika Freitas, David Schlesinger. Hemocentro Ribeirão Preto: Simone Kashima, Evandra Strazza Rodrigues, Svetoslav Nanev Slavov, Elaine Vieira dos Santos, Rafael dos Santos Bezerra, Luiz Carlos Junior de Alcantara, Marta Giovanetti, Vagner Fonseca, Flavia Aburjaile, Rodrigo Tocantins Calado. FAMERP-SJRP: Cecília Artico Banho, Lívia Sacchetto, Fábio Sossai Possebon, Leila Sabrina Ullmann, Cíntia Bittar, Guilherme Campos, Helena Lage Ferreira, Jorge A. Petroli Marchesi, Maisa C. Pereira Parra, Marília Moraes, Paula Rahal, Paulo Inacio da Costa, João Pessoa Araújo Jr., Maurício Lacerda Nogueira. Prefeitura de Sao Paulo: Melissa Palmieri. |
| EPI_ISL_1966854, EPI_ISL_1966855, EPI_ISL_1966856, EPI_ISL_1966857 | UNIDADE SENTINELA COVID19 | Instituto Butantan / ESALQ-USP (Piracicaba) | Instituto Butantan: Dimas Tadeu Covas, Sandra Coccuzzo Sampaio, Maria Carolina Elias, José Salvatore Leister Patané, Vincent Louis Viala, Antonio Jorge Martins, Ricardo Haddad, Claudia Renata dos Santos Barros, Elaine Cristina Marqueze, Raul Machado Neto, Debora Botequio Moretti, Jardelina de Souza Todao Bernardino, Loyze Paola Oliveira de Lima, Luiz Aurelio de Campos Crispin. Centro de Genômica Funcional da ESALQ: Luiz Lehmann Coutinho, Ricardo Augusto Brassaloti, Raquel de Lello Rocha Campos Cassano. NGS Soluções Genômicas: Pilar Drummond Sampaio Corrêa Mariani. FZEA-USP Pirassununga: Mirele Daiana Poleti, Jessika Cristina Chagas Lesbon, Elisangela Chicaroni Mattos, Heidge Fukumasu. USP-Botucatu: Rejane Maria Tommasini Grotto, Jayme A. Souza-Neto, Guilherme Targino Valente, Patricia Akemi Assato, Felipe Allan da Silva da Costa, Bianca Cechetto Carlos. Mendelics: Bibiana Santos, João Paulo Kitajima, Erika Freitas, David Schlesinger. Hemocentro Ribeirão Preto: Simone Kashima, Evandra Strazza Rodrigues, Svetoslav Nanev Slavov, Elaine Vieira dos Santos, Rafael dos Santos Bezerra, Luiz Carlos Junior de Alcantara, Marta Giovanetti, Vagner Fonseca, Flavia Aburjaile, Rodrigo Tocantins Calado. FAMERP-SJRP: Cecília Artico Banho, Lívia Sacchetto, Fábio Sossai Possebon, Leila Sabrina Ullmann, Cíntia Bittar, Guilherme Campos, Helena Lage Ferreira, Jorge A. Petroli Marchesi, Maisa C. Pereira Parra, Marília Moraes, Paula Rahal, Paulo Inacio da Costa, João Pessoa Araújo Jr., Maurício Lacerda Nogueira. Prefeitura de Sao Paulo: Melissa Palmieri. |
| EPI_ISL_1966859, EPI_ISL_1966860 | UNIDADE SENTINELA VILA APARECIDA | Instituto Butantan / ESALQ-USP (Piracicaba) | Instituto Butantan: Dimas Tadeu Covas, Sandra Coccuzzo Sampaio, Maria Carolina Elias, José Salvatore Leister Patané, Vincent Louis Viala, Antonio Jorge Martins, Ricardo Haddad, Claudia Renata dos Santos Barros, Elaine Cristina Marqueze, Raul Machado Neto, Debora Botequio Moretti, Jardelina de Souza Todao Bernardino, Loyze Paola Oliveira de Lima, Luiz Aurelio de Campos Crispin. Centro de Genômica Funcional da ESALQ: Luiz Lehmann Coutinho, Ricardo Augusto Brassaloti, Raquel de Lello Rocha Campos Cassano. NGS Soluções Genômicas: Pilar Drummond Sampaio Corrêa Mariani. FZEA-USP Pirassununga: Mirele Daiana Poleti, Jessika Cristina Chagas Lesbon, Elisangela Chicaroni Mattos, Heidge Fukumasu. USP-Botucatu: Rejane Maria Tommasini Grotto, Jayme A. Souza-Neto, Guilherme Targino Valente, Patricia Akemi Assato, Felipe Allan da Silva da Costa, Bianca Cechetto Carlos. Mendelics: Bibiana Santos, João Paulo Kitajima, Erika Freitas, David Schlesinger. Hemocentro Ribeirão Preto: Simone Kashima, Evandra Strazza Rodrigues, Svetoslav Nanev Slavov, Elaine Vieira dos Santos, Rafael dos Santos Bezerra, Luiz Carlos Junior de Alcantara, Marta Giovanetti, Vagner Fonseca, Flavia Aburjaile, Rodrigo Tocantins Calado. FAMERP-SJRP: Cecília Artico Banho, Lívia Sacchetto, Fábio Sossai Possebon, Leila Sabrina Ullmann, Cíntia Bittar, Guilherme Campos, Helena Lage Ferreira, Jorge A. Petroli Marchesi, Maisa C. Pereira Parra, Marília Moraes, Paula Rahal, Paulo Inacio da Costa, João Pessoa Araújo Jr., Maurício Lacerda Nogueira. Prefeitura de Sao Paulo: Melissa Palmieri. |
| EPI_ISL_1966861 | UNIDADE SENTINELA COVID19 | Instituto Butantan / ESALQ-USP (Piracicaba) | Instituto Butantan: Dimas Tadeu Covas, Sandra Coccuzzo Sampaio, Maria Carolina Elias, José Salvatore Leister Patané, Vincent Louis Viala, Antonio Jorge Martins, Ricardo Haddad, Claudia Renata dos Santos Barros, Elaine Cristina Marqueze, Raul Machado Neto, Debora Botequio Moretti, Jardelina de |

| | | | |
|---|---|---|---|
| | | | Souza Todao Bernardino, Loyze Paola Oliveira de Lima, Luiz Aurelio de Campos Crispin. Centro de Genômica Funcional da ESALQ: Luiz Lehmann Coutinho, Ricardo Augusto Brassaloti, Raquel de Lello Rocha Campos Cassano. NGS Soluções Genômicas: Pilar Drummond Sampaio Corrêa Mariani. FZEA-USP Pirassununga: Mirele Daiana Poleti, Jessika Cristina Chagas Lesbon, Elisangela Chicaroni Mattos, Heidge Fukumasu. USP-Botucatu: Rejane Maria Tommasini Grotto, Jayme A. Souza-Neto, Guilherme Targino Valente, Patricia Akemi Assato, Felipe Allan da Silva da Costa, Bianca Cechetto Carlos. Mendelics: Bibiana Santos, João Paulo Kitajima, Erika Freitas, David Schlesinger. Hemocentro Ribeirão Preto: Simone Kashima, Evandra Strazza Rodrigues, Svetoslav Nanev Slavov, Elaine Vieira dos Santos, Rafael dos Santos Bezerra, Luiz Carlos Junior de Alcantara, Marta Giovanetti, Vagner Fonseca, Flavia Aburjaile, Rodrigo Tocantins Calado. FAMERP-SJRP: Cecília Artico Banho, Lívia Sacchetto, Fábio Sossai Possebon, Leila Sabrina Ullmann, Cíntia Bittar, Guilherme Campos, Helena Lage Ferreira, Jorge A. Petroli Marchesi, Maisa C. Pereira Parra, Marília Moraes, Paula Rahal, Paulo Inacio da Costa, João Pessoa Araújo Jr., Maurício Lacerda Nogueira. Prefeitura de Sao Paulo: Melissa Palmieri. |
| EPI_ISL_1966863, EPI_ISL_1966864, EPI_ISL_1966865, EPI_ISL_1966866, EPI_ISL_1966867 | UNIDADE MISTA DE SAUDE CAPELA DO ALTO | Instituto Butantan / ESALQ-USP (Piracicaba) | Instituto Butantan: Dimas Tadeu Covas, Sandra Coccuzzo Sampaio, Maria Carolina Elias, José Salvatore Leister Patané, Vincent Louis Viala, Antonio Jorge Martins, Ricardo Haddad, Claudia Renata dos Santos Barros, Elaine Cristina Marqueze, Raul Machado Neto, Debora Botequio Moretti, Jardelina de Souza Todao Bernardino, Loyze Paola Oliveira de Lima, Luiz Aurelio de Campos Crispin. Centro de Genômica Funcional da ESALQ: Luiz Lehmann Coutinho, Ricardo Augusto Brassaloti, Raquel de Lello Rocha Campos Cassano. NGS Soluções Genômicas: Pilar Drummond Sampaio Corrêa Mariani. FZEA-USP Pirassununga: Mirele Daiana Poleti, Jessika Cristina Chagas Lesbon, Elisangela Chicaroni Mattos, Heidge Fukumasu. USP-Botucatu: Rejane Maria Tommasini Grotto, Jayme A. Souza-Neto, Guilherme Targino Valente, Patricia Akemi Assato, Felipe Allan da Silva da Costa, Bianca Cechetto Carlos. Mendelics: Bibiana Santos, João Paulo Kitajima, Erika Freitas, David Schlesinger. Hemocentro Ribeirão Preto: Simone Kashima, Evandra Strazza Rodrigues, Svetoslav Nanev Slavov, Elaine Vieira dos Santos, Rafael dos Santos Bezerra, Luiz Carlos Junior de Alcantara, Marta Giovanetti, Vagner Fonseca, Flavia Aburjaile, Rodrigo Tocantins Calado. FAMERP-SJRP: Cecília Artico Banho, Lívia Sacchetto, Fábio Sossai Possebon, Leila Sabrina Ullmann, Cíntia Bittar, Guilherme Campos, Helena Lage Ferreira, Jorge A. Petroli Marchesi, Maisa C. Pereira Parra, Marília Moraes, Paula Rahal, Paulo Inacio da Costa, João Pessoa Araújo Jr., Maurício Lacerda Nogueira. Prefeitura de Sao Paulo: Melissa Palmieri. |
| EPI_ISL_1966868, EPI_ISL_1966869 | CS DE CATIGUA | Instituto Butantan / ESALQ-USP (Piracicaba) | Instituto Butantan: Dimas Tadeu Covas, Sandra Coccuzzo Sampaio, Maria Carolina Elias, José Salvatore Leister Patané, Vincent Louis Viala, Antonio Jorge Martins, Ricardo Haddad, Claudia Renata dos Santos Barros, Elaine Cristina Marqueze, Raul Machado Neto, Debora Botequio Moretti, Jardelina de Souza Todao Bernardino, Loyze Paola Oliveira de Lima, Luiz Aurelio de Campos Crispin. Centro de Genômica Funcional da ESALQ: Luiz Lehmann Coutinho, Ricardo Augusto Brassaloti, Raquel de Lello Rocha Campos Cassano. NGS Soluções Genômicas: Pilar Drummond Sampaio Corrêa Mariani. FZEA-USP Pirassununga: Mirele Daiana Poleti, Jessika Cristina Chagas Lesbon, Elisangela Chicaroni Mattos, Heidge Fukumasu. USP-Botucatu: Rejane Maria Tommasini Grotto, Jayme A. Souza-Neto, Guilherme Targino Valente, Patricia Akemi Assato, Felipe Allan da Silva da Costa, Bianca Cechetto Carlos. Mendelics: Bibiana Santos, João Paulo Kitajima, Erika Freitas, David Schlesinger. Hemocentro Ribeirão Preto: Simone Kashima, Evandra Strazza Rodrigues, Svetoslav Nanev Slavov, Elaine Vieira dos Santos, Rafael dos Santos Bezerra, Luiz Carlos Junior de Alcantara, Marta Giovanetti, Vagner Fonseca, Flavia Aburjaile, Rodrigo Tocantins Calado. FAMERP-SJRP: Cecília Artico Banho, Lívia Sacchetto, Fábio Sossai Possebon, Leila Sabrina Ullmann, Cíntia Bittar, Guilherme Campos, Helena Lage Ferreira, Jorge A. Petroli Marchesi, Maisa C. Pereira Parra, Marília Moraes, Paula Rahal, Paulo Inacio da Costa, João Pessoa Araújo Jr., Maurício Lacerda Nogueira. Prefeitura de Sao Paulo: Melissa Palmieri. |
| EPI_ISL_1966870, EPI_ISL_1966871, EPI_ISL_1966872, EPI_ISL_1966873, EPI_ISL_1966874, EPI_ISL_1966875 | UBS ATALAIA DR MARIO AGUIAR FILHO | Instituto Butantan / ESALQ-USP (Piracicaba) | Instituto Butantan: Dimas Tadeu Covas, Sandra Coccuzzo Sampaio, Maria Carolina Elias, José Salvatore Leister Patané, Vincent Louis Viala, Antonio Jorge Martins, Ricardo Haddad, Claudia Renata dos Santos Barros, Elaine Cristina Marqueze, Raul Machado Neto, Debora Botequio Moretti, Jardelina de Souza Todao Bernardino, Loyze Paola Oliveira de Lima, Luiz Aurelio de Campos Crispin. Centro de Genômica Funcional da ESALQ: Luiz Lehmann Coutinho, Ricardo Augusto Brassaloti, Raquel de Lello Rocha Campos Cassano. NGS Soluções Genômicas: Pilar Drummond Sampaio Corrêa Mariani. FZEA-USP Pirassununga: Mirele Daiana Poleti, Jessika Cristina Chagas Lesbon, Elisangela Chicaroni Mattos, Heidge Fukumasu. USP-Botucatu: Rejane Maria Tommasini Grotto, Jayme A. Souza-Neto, Guilherme Targino Valente, Patricia Akemi Assato, Felipe Allan da Silva da Costa, Bianca Cechetto Carlos. Mendelics: Bibiana Santos, João Paulo Kitajima, Erika Freitas, David Schlesinger. Hemocentro Ribeirão Preto: Simone Kashima, Evandra Strazza Rodrigues, Svetoslav Nanev Slavov, Elaine Vieira dos Santos, Rafael dos Santos Bezerra, Luiz Carlos Junior de Alcantara, Marta Giovanetti, Vagner Fonseca, Flavia Aburjaile, Rodrigo Tocantins Calado. FAMERP-SJRP: Cecília Artico Banho, Lívia Sacchetto, Fábio Sossai Possebon, Leila Sabrina Ullmann, Cíntia Bittar, Guilherme Campos, Helena Lage Ferreira, Jorge A. Petroli Marchesi, Maisa C. Pereira Parra, Marília Moraes, Paula Rahal, Paulo Inacio da Costa, João Pessoa Araújo Jr., Maurício Lacerda Nogueira. Prefeitura de Sao Paulo: Melissa Palmieri. |
| EPI_ISL_1966876 | PRONTO SOCORRO CAUCAIA DO ALTO | Instituto Butantan / ESALQ-USP (Piracicaba) | Instituto Butantan: Dimas Tadeu Covas, Sandra Coccuzzo Sampaio, Maria Carolina Elias, José Salvatore Leister Patané, Vincent Louis Viala, Antonio Jorge Martins, Ricardo Haddad, Claudia Renata dos Santos Barros, Elaine Cristina Marqueze, Raul Machado Neto, Debora Botequio Moretti, Jardelina de Souza Todao Bernardino, Loyze Paola Oliveira de Lima, Luiz Aurelio de Campos Crispin. Centro de Genômica Funcional da ESALQ: Luiz Lehmann Coutinho, Ricardo Augusto Brassaloti, Raquel de Lello Rocha Campos Cassano. NGS Soluções Genômicas: Pilar Drummond Sampaio Corrêa Mariani. FZEA-USP Pirassununga: Mirele Daiana Poleti, Jessika Cristina Chagas Lesbon, Elisangela Chicaroni Mattos, Heidge Fukumasu. USP-Botucatu: Rejane Maria Tommasini Grotto, Jayme A. Souza-Neto, Guilherme Targino Valente, Patricia Akemi Assato, Felipe Allan da Silva da Costa, Bianca Cechetto Carlos. Mendelics: Bibiana Santos, João Paulo Kitajima, Erika Freitas, David Schlesinger. Hemocentro Ribeirão Preto: Simone Kashima, Evandra Strazza Rodrigues, Svetoslav Nanev Slavov, Elaine Vieira dos Santos, Rafael dos Santos Bezerra, Luiz Carlos Junior de Alcantara, Marta Giovanetti, Vagner Fonseca, Flavia Aburjaile, Rodrigo Tocantins Calado. FAMERP-SJRP: Cecília Artico Banho, Lívia Sacchetto, Fábio Sossai Possebon, Leila Sabrina Ullmann, Cíntia Bittar, Guilherme Campos, Helena Lage Ferreira, Jorge A. Petroli Marchesi, Maisa C. Pereira Parra, Marília Moraes, Paula Rahal, Paulo Inacio da Costa, João Pessoa Araújo Jr., Maurício Lacerda Nogueira. Prefeitura de Sao Paulo: Melissa Palmieri. |
| EPI_ISL_1966877, EPI_ISL_1966878, EPI_ISL_1966879, EPI_ISL_1966880, EPI_ISL_1966881, EPI_ISL_1966882, EPI_ISL_1966883, EPI_ISL_1966884, EPI_ISL_1966885, EPI_ISL_1966886, EPI_ISL_1966887, EPI_ISL_1966888 | | | |
| see above | UBS ATALAIA DR MARIO AGUIAR FILHO | Instituto Butantan / ESALQ-USP (Piracicaba) | Instituto Butantan: Dimas Tadeu Covas, Sandra Coccuzzo Sampaio, Maria Carolina Elias, José Salvatore Leister Patané, Vincent Louis Viala, Antonio Jorge Martins, Ricardo Haddad, Claudia Renata dos Santos Barros, Elaine Cristina Marqueze, Raul Machado Neto, Debora Botequio Moretti, Jardelina de Souza Todao Bernardino, Loyze Paola Oliveira de Lima, Luiz Aurelio de Campos Crispin. Centro de Genômica Funcional da ESALQ: Luiz Lehmann Coutinho, Ricardo Augusto Brassaloti, Raquel de Lello Rocha Campos Cassano. NGS Soluções Genômicas: Pilar Drummond Sampaio Corrêa Mariani. FZEA-USP Pirassununga: Mirele Daiana Poleti, Jessika Cristina Chagas Lesbon, Elisangela Chicaroni Mattos, Heidge Fukumasu. USP-Botucatu: Rejane Maria Tommasini Grotto, Jayme A. Souza-Neto, Guilherme Targino Valente, Patricia Akemi Assato, Felipe Allan da Silva da Costa, Bianca Cechetto Carlos. Mendelics: Bibiana Santos, João Paulo Kitajima, Erika Freitas, David Schlesinger. Hemocentro Ribeirão Preto: Simone Kashima, Evandra Strazza Rodrigues, Svetoslav Nanev Slavov, Elaine Vieira dos Santos, Rafael dos Santos Bezerra, Luiz Carlos Junior de Alcantara, Marta Giovanetti, Vagner Fonseca, Flavia Aburjaile, Rodrigo Tocantins Calado. FAMERP-SJRP: Cecília Artico Banho, Lívia Sacchetto, Fábio Sossai Possebon, Leila Sabrina Ullmann, Cíntia Bittar, Guilherme Campos, Helena Lage Ferreira, Jorge A. Petroli Marchesi, Maisa C. Pereira Parra, Marília Moraes, Paula Rahal, Paulo Inacio da Costa, João Pessoa Araújo Jr., Maurício Lacerda Nogueira. Prefeitura de Sao Paulo: Melissa Palmieri. |
| EPI_ISL_1966889 | UPA ATALAIA DRA RITA DE CASSIA SORIO | Instituto Butantan / ESALQ-USP (Piracicaba) | Instituto Butantan: Dimas Tadeu Covas, Sandra Coccuzzo Sampaio, Maria Carolina Elias, José Salvatore Leister Patané, Vincent Louis Viala, Antonio Jorge Martins, Ricardo Haddad, Claudia Renata dos Santos Barros, Elaine Cristina Marqueze, Raul Machado Neto, Debora Botequio Moretti, Jardelina de Souza Todao Bernardino, Loyze Paola Oliveira de Lima, Luiz Aurelio de Campos Crispin. Centro de Genômica Funcional da ESALQ: Luiz Lehmann Coutinho, Ricardo Augusto Brassaloti, Raquel de Lello Rocha Campos Cassano. NGS Soluções Genômicas: Pilar Drummond Sampaio Corrêa Mariani. FZEA-USP Pirassununga: Mirele Daiana Poleti, Jessika Cristina Chagas Lesbon, Elisangela Chicaroni Mattos, Heidge Fukumasu. USP-Botucatu: Rejane Maria Tommasini Grotto, Jayme A. Souza-Neto, Guilherme Targino Valente, Patricia Akemi Assato, Felipe Allan da Silva da Costa, Bianca Cechetto Carlos. Mendelics: Bibiana Santos, João Paulo Kitajima, Erika Freitas, David Schlesinger. Hemocentro Ribeirão Preto: Simone Kashima, Evandra Strazza Rodrigues, Svetoslav Nanev Slavov, Elaine Vieira dos Santos, Rafael dos Santos Bezerra, Luiz Carlos Junior de Alcantara, Marta Giovanetti, Vagner Fonseca, Flavia Aburjaile, Rodrigo Tocantins Calado. FAMERP-SJRP: Cecília Artico Banho, Lívia Sacchetto, Fábio Sossai Possebon, Leila Sabrina Ullmann, Cíntia Bittar, Guilherme Campos, Helena Lage Ferreira, Jorge A. Petroli Marchesi, Maisa C. Pereira Parra, Marília Moraes, Paula Rahal, Paulo Inacio da Costa, João Pessoa Araújo Jr., Maurício Lacerda Nogueira. Prefeitura de Sao Paulo: Melissa Palmieri. |
| EPI_ISL_1966890 | UBS ATALAIA DR MARIO AGUIAR FILHO | Instituto Butantan / ESALQ-USP (Piracicaba) | Instituto Butantan: Dimas Tadeu Covas, Sandra Coccuzzo Sampaio, Maria Carolina Elias, José Salvatore Leister Patané, Vincent Louis Viala, Antonio Jorge Martins, Ricardo Haddad, Claudia Renata dos Santos Barros, Elaine Cristina Marqueze, Raul Machado Neto, Debora Botequio Moretti, Jardelina de Souza Todao Bernardino, Loyze Paola Oliveira de Lima, Luiz Aurelio de Campos Crispin. Centro de Genômica Funcional da ESALQ: Luiz Lehmann Coutinho, Ricardo Augusto Brassaloti, Raquel de Lello Rocha Campos Cassano. NGS Soluções Genômicas: Pilar Drummond Sampaio Corrêa Mariani. FZEA-USP Pirassununga: Mirele Daiana Poleti, Jessika Cristina Chagas Lesbon, Elisangela Chicaroni Mattos, Heidge Fukumasu. USP-Botucatu: Rejane Maria Tommasini Grotto, Jayme A. Souza-Neto, Guilherme Targino Valente, Patricia Akemi Assato, Felipe Allan da Silva da Costa, Bianca Cechetto Carlos. Mendelics: Bibiana Santos, João Paulo Kitajima, Erika Freitas, David Schlesinger. Hemocentro Ribeirão Preto: Simone Kashima, Evandra Strazza |

| | | | |
|---|---|---|---|
| | | | Rodrigues, Svetoslav Nanev Slavov, Elaine Vieira dos Santos, Rafael dos Santos Bezerra, Luiz Carlos Junior de Alcantara, Marta Giovanetti, Vagner Fonseca, Flavia Aburjaile, Rodrigo Tocantins Calado. FAMERP-SJRP: Cecília Artico Banho, Lívia Sacchetto, Fábio Sossai Possebon, Leila Sabrina Ullmann, Cíntia Bittar, Guilherme Campos, Helena Lage Ferreira, Jorge A. Petroli Marchesi, Maisa C. Pereira Parra, Marília Moraes, Paula Rahal, Paulo Inacio da Costa, João Pessoa Araújo Jr., Maurício Lacerda Nogueira. Prefeitura de Sao Paulo: Melissa Palmieri. |
| EPI_ISL_1966891, EPI_ISL_1966892, EPI_ISL_1966893, EPI_ISL_1966894 | UPA ATALAIA DRA RITA DE CASSIA SORIO | Instituto Butantan / ESALQ-USP (Piracicaba) | Instituto Butantan: Dimas Tadeu Covas, Sandra Coccuzzo Sampaio, Maria Carolina Elias, José Salvatore Leister Patané, Vincent Louis Viala, Antonio Jorge Martins, Ricardo Haddad, Claudia Renata dos Santos Barros, Elaine Cristina Marqueze, Raul Machado Neto, Debora Botequio Moretti, Jardelina de Souza Todao Bernardino, Loyze Paola Oliveira de Lima, Luiz Aurelio de Campos Crispin. Centro de Genômica Funcional da ESALQ: Luiz Lehmann Coutinho, Ricardo Augusto Brassaloti, Raquel de Lello Rocha Campos Cassano. NGS Soluções Genômicas: Pilar Drummond Sampaio Corrêa Mariani. FZEA-USP Pirassununga: Mirele Daiana Poleti, Jessika Cristina Chagas Lesbon, Elisangela Chicaroni Mattos, Heidge Fukumasu. USP-Botucatu: Rejane Maria Tommasini Grotto, Jayme A. Souza-Neto, Guilherme Targino Valente, Patricia Akemi Assato, Felipe Allan da Silva da Costa, Bianca Cechetto Carlos. Mendelics: Bibiana Santos, João Paulo Kitajima, Erika Freitas, David Schlesinger. Hemocentro Ribeirão Preto: Simone Kashima, Evandra Strazza Rodrigues, Svetoslav Nanev Slavov, Elaine Vieira dos Santos, Rafael dos Santos Bezerra, Luiz Carlos Junior de Alcantara, Marta Giovanetti, Vagner Fonseca, Flavia Aburjaile, Rodrigo Tocantins Calado. FAMERP-SJRP: Cecília Artico Banho, Lívia Sacchetto, Fábio Sossai Possebon, Leila Sabrina Ullmann, Cíntia Bittar, Guilherme Campos, Helena Lage Ferreira, Jorge A. Petroli Marchesi, Maisa C. Pereira Parra, Marília Moraes, Paula Rahal, Paulo Inacio da Costa, João Pessoa Araújo Jr., Maurício Lacerda Nogueira. Prefeitura de Sao Paulo: Melissa Palmieri. |
| EPI_ISL_1966895, EPI_ISL_1966896, EPI_ISL_1966897, EPI_ISL_1966898, EPI_ISL_1966899, EPI_ISL_1966900, EPI_ISL_1966902, EPI_ISL_1966903, EPI_ISL_1966904 | CENTRO DE APOIO EPIDEMIOLOGICO | Instituto Butantan / ESALQ-USP (Piracicaba) | Instituto Butantan: Dimas Tadeu Covas, Sandra Coccuzzo Sampaio, Maria Carolina Elias, José Salvatore Leister Patané, Vincent Louis Viala, Antonio Jorge Martins, Ricardo Haddad, Claudia Renata dos Santos Barros, Elaine Cristina Marqueze, Raul Machado Neto, Debora Botequio Moretti, Jardelina de Souza Todao Bernardino, Loyze Paola Oliveira de Lima, Luiz Aurelio de Campos Crispin. Centro de Genômica Funcional da ESALQ: Luiz Lehmann Coutinho, Ricardo Augusto Brassaloti, Raquel de Lello Rocha Campos Cassano. NGS Soluções Genômicas: Pilar Drummond Sampaio Corrêa Mariani. FZEA-USP Pirassununga: Mirele Daiana Poleti, Jessika Cristina Chagas Lesbon, Elisangela Chicaroni Mattos, Heidge Fukumasu. USP-Botucatu: Rejane Maria Tommasini Grotto, Jayme A. Souza-Neto, Guilherme Targino Valente, Patricia Akemi Assato, Felipe Allan da Silva da Costa, Bianca Cechetto Carlos. Mendelics: Bibiana Santos, João Paulo Kitajima, Erika Freitas, David Schlesinger. Hemocentro Ribeirão Preto: Simone Kashima, Evandra Strazza Rodrigues, Svetoslav Nanev Slavov, Elaine Vieira dos Santos, Rafael dos Santos Bezerra, Luiz Carlos Junior de Alcantara, Marta Giovanetti, Vagner Fonseca, Flavia Aburjaile, Rodrigo Tocantins Calado. FAMERP-SJRP: Cecília Artico Banho, Lívia Sacchetto, Fábio Sossai Possebon, Leila Sabrina Ullmann, Cíntia Bittar, Guilherme Campos, Helena Lage Ferreira, Jorge A. Petroli Marchesi, Maisa C. Pereira Parra, Marília Moraes, Paula Rahal, Paulo Inacio da Costa, João Pessoa Araújo Jr., Maurício Lacerda Nogueira. Prefeitura de Sao Paulo: Melissa Palmieri. |
| EPI_ISL_1966905 | LABORATORIO DE FRANCA | Instituto Butantan / ESALQ-USP (Piracicaba) | Instituto Butantan: Dimas Tadeu Covas, Sandra Coccuzzo Sampaio, Maria Carolina Elias, José Salvatore Leister Patané, Vincent Louis Viala, Antonio Jorge Martins, Ricardo Haddad, Claudia Renata dos Santos Barros, Elaine Cristina Marqueze, Raul Machado Neto, Debora Botequio Moretti, Jardelina de Souza Todao Bernardino, Loyze Paola Oliveira de Lima, Luiz Aurelio de Campos Crispin. Centro de Genômica Funcional da ESALQ: Luiz Lehmann Coutinho, Ricardo Augusto Brassaloti, Raquel de Lello Rocha Campos Cassano. NGS Soluções Genômicas: Pilar Drummond Sampaio Corrêa Mariani. FZEA-USP Pirassununga: Mirele Daiana Poleti, Jessika Cristina Chagas Lesbon, Elisangela Chicaroni Mattos, Heidge Fukumasu. USP-Botucatu: Rejane Maria Tommasini Grotto, Jayme A. Souza-Neto, Guilherme Targino Valente, Patricia Akemi Assato, Felipe Allan da Silva da Costa, Bianca Cechetto Carlos. Mendelics: Bibiana Santos, João Paulo Kitajima, Erika Freitas, David Schlesinger. Hemocentro Ribeirão Preto: Simone Kashima, Evandra Strazza Rodrigues, Svetoslav Nanev Slavov, Elaine Vieira dos Santos, Rafael dos Santos Bezerra, Luiz Carlos Junior de Alcantara, Marta Giovanetti, Vagner Fonseca, Flavia Aburjaile, Rodrigo Tocantins Calado. FAMERP-SJRP: Cecília Artico Banho, Lívia Sacchetto, Fábio Sossai Possebon, Leila Sabrina Ullmann, Cíntia Bittar, Guilherme Campos, Helena Lage Ferreira, Jorge A. Petroli Marchesi, Maisa C. Pereira Parra, Marília Moraes, Paula Rahal, Paulo Inacio da Costa, João Pessoa Araújo Jr., Maurício Lacerda Nogueira. Prefeitura de Sao Paulo: Melissa Palmieri. |
| EPI_ISL_1966907, EPI_ISL_1966908 | USF I DE GUAIMBE | Instituto Butantan / ESALQ-USP (Piracicaba) | Instituto Butantan: Dimas Tadeu Covas, Sandra Coccuzzo Sampaio, Maria Carolina Elias, José Salvatore Leister Patané, Vincent Louis Viala, Antonio Jorge Martins, Ricardo Haddad, Claudia Renata dos Santos Barros, Elaine Cristina Marqueze, Raul Machado Neto, Debora Botequio Moretti, Jardelina de Souza Todao Bernardino, Loyze Paola Oliveira de Lima, Luiz Aurelio de Campos Crispin. Centro de Genômica Funcional da ESALQ: Luiz Lehmann Coutinho, Ricardo Augusto Brassaloti, Raquel de Lello Rocha Campos Cassano. NGS Soluções Genômicas: Pilar Drummond Sampaio Corrêa Mariani. FZEA-USP Pirassununga: Mirele Daiana Poleti, Jessika Cristina Chagas Lesbon, Elisangela Chicaroni Mattos, Heidge Fukumasu. USP-Botucatu: Rejane Maria Tommasini Grotto, Jayme A. Souza-Neto, Guilherme Targino Valente, Patricia Akemi Assato, Felipe Allan da Silva da Costa, Bianca Cechetto Carlos. Mendelics: Bibiana Santos, João Paulo Kitajima, Erika Freitas, David Schlesinger. Hemocentro Ribeirão Preto: Simone Kashima, Evandra Strazza Rodrigues, Svetoslav Nanev Slavov, Elaine Vieira dos Santos, Rafael dos Santos Bezerra, Luiz Carlos Junior de Alcantara, Marta Giovanetti, Vagner Fonseca, Flavia Aburjaile, Rodrigo Tocantins Calado. FAMERP-SJRP: Cecília Artico Banho, Lívia Sacchetto, Fábio Sossai Possebon, Leila Sabrina Ullmann, Cíntia Bittar, Guilherme Campos, Helena Lage Ferreira, Jorge A. Petroli Marchesi, Maisa C. Pereira Parra, Marília Moraes, Paula Rahal, Paulo Inacio da Costa, João Pessoa Araújo Jr., Maurício Lacerda Nogueira. Prefeitura de Sao Paulo: Melissa Palmieri. |
| EPI_ISL_1966909 | CS II DR JAHYR DE PAULA RIBEIRO GUARA | Instituto Butantan / ESALQ-USP (Piracicaba) | Instituto Butantan: Dimas Tadeu Covas, Sandra Coccuzzo Sampaio, Maria Carolina Elias, José Salvatore Leister Patané, Vincent Louis Viala, Antonio Jorge Martins, Ricardo Haddad, Claudia Renata dos Santos Barros, Elaine Cristina Marqueze, Raul Machado Neto, Debora Botequio Moretti, Jardelina de Souza Todao Bernardino, Loyze Paola Oliveira de Lima, Luiz Aurelio de Campos Crispin. Centro de Genômica Funcional da ESALQ: Luiz Lehmann Coutinho, Ricardo Augusto Brassaloti, Raquel de Lello Rocha Campos Cassano. NGS Soluções Genômicas: Pilar Drummond Sampaio Corrêa Mariani. FZEA-USP Pirassununga: Mirele Daiana Poleti, Jessika Cristina Chagas Lesbon, Elisangela Chicaroni Mattos, Heidge Fukumasu. USP-Botucatu: Rejane Maria Tommasini Grotto, Jayme A. Souza-Neto, Guilherme Targino Valente, Patricia Akemi Assato, Felipe Allan da Silva da Costa, Bianca Cechetto Carlos. Mendelics: Bibiana Santos, João Paulo Kitajima, Erika Freitas, David Schlesinger. Hemocentro Ribeirão Preto: Simone Kashima, Evandra Strazza Rodrigues, Svetoslav Nanev Slavov, Elaine Vieira dos Santos, Rafael dos Santos Bezerra, Luiz Carlos Junior de Alcantara, Marta Giovanetti, Vagner Fonseca, Flavia Aburjaile, Rodrigo Tocantins Calado. FAMERP-SJRP: Cecília Artico Banho, Lívia Sacchetto, Fábio Sossai Possebon, Leila Sabrina Ullmann, Cíntia Bittar, Guilherme Campos, Helena Lage Ferreira, Jorge A. Petroli Marchesi, Maisa C. Pereira Parra, Marília Moraes, Paula Rahal, Paulo Inacio da Costa, João Pessoa Araújo Jr., Maurício Lacerda Nogueira. Prefeitura de Sao Paulo: Melissa Palmieri. |
| EPI_ISL_1966910, EPI_ISL_1966911 | PR S DA FAMILIA UNIDADE DE SAUDE ADALBERTO ROCHA GUAREI | Instituto Butantan / ESALQ-USP (Piracicaba) | Instituto Butantan: Dimas Tadeu Covas, Sandra Coccuzzo Sampaio, Maria Carolina Elias, José Salvatore Leister Patané, Vincent Louis Viala, Antonio Jorge Martins, Ricardo Haddad, Claudia Renata dos Santos Barros, Elaine Cristina Marqueze, Raul Machado Neto, Debora Botequio Moretti, Jardelina de Souza Todao Bernardino, Loyze Paola Oliveira de Lima, Luiz Aurelio de Campos Crispin. Centro de Genômica Funcional da ESALQ: Luiz Lehmann Coutinho, Ricardo Augusto Brassaloti, Raquel de Lello Rocha Campos Cassano. NGS Soluções Genômicas: Pilar Drummond Sampaio Corrêa Mariani. FZEA-USP Pirassununga: Mirele Daiana Poleti, Jessika Cristina Chagas Lesbon, Elisangela Chicaroni Mattos, Heidge Fukumasu. USP-Botucatu: Rejane Maria Tommasini Grotto, Jayme A. Souza-Neto, Guilherme Targino Valente, Patricia Akemi Assato, Felipe Allan da Silva da Costa, Bianca Cechetto Carlos. Mendelics: Bibiana Santos, João Paulo Kitajima, Erika Freitas, David Schlesinger. Hemocentro Ribeirão Preto: Simone Kashima, Evandra Strazza Rodrigues, Svetoslav Nanev Slavov, Elaine Vieira dos Santos, Rafael dos Santos Bezerra, Luiz Carlos Junior de Alcantara, Marta Giovanetti, Vagner Fonseca, Flavia Aburjaile, Rodrigo Tocantins Calado. FAMERP-SJRP: Cecília Artico Banho, Lívia Sacchetto, Fábio Sossai Possebon, Leila Sabrina Ullmann, Cíntia Bittar, Guilherme Campos, Helena Lage Ferreira, Jorge A. Petroli Marchesi, Maisa C. Pereira Parra, Marília Moraes, Paula Rahal, Paulo Inacio da Costa, João Pessoa Araújo Jr., Maurício Lacerda Nogueira. Prefeitura de Sao Paulo: Melissa Palmieri. |
| EPI_ISL_1966912 | CENTRO DE SAUDE II DR ARCY BANDEIRA IBIUNA SP | Instituto Butantan / ESALQ-USP (Piracicaba) | Instituto Butantan: Dimas Tadeu Covas, Sandra Coccuzzo Sampaio, Maria Carolina Elias, José Salvatore Leister Patané, Vincent Louis Viala, Antonio Jorge Martins, Ricardo Haddad, Claudia Renata dos Santos Barros, Elaine Cristina Marqueze, Raul Machado Neto, Debora Botequio Moretti, Jardelina de Souza Todao Bernardino, Loyze Paola Oliveira de Lima, Luiz Aurelio de Campos Crispin. Centro de Genômica Funcional da ESALQ: Luiz Lehmann Coutinho, Ricardo Augusto Brassaloti, Raquel de Lello Rocha Campos Cassano. NGS Soluções Genômicas: Pilar Drummond Sampaio Corrêa Mariani. FZEA-USP Pirassununga: Mirele Daiana Poleti, Jessika Cristina Chagas Lesbon, Elisangela Chicaroni Mattos, Heidge Fukumasu. USP-Botucatu: Rejane Maria Tommasini Grotto, Jayme A. Souza-Neto, Guilherme Targino Valente, Patricia Akemi Assato, Felipe Allan da Silva da Costa, Bianca Cechetto Carlos. Mendelics: Bibiana Santos, João Paulo Kitajima, Erika Freitas, David Schlesinger. Hemocentro Ribeirão Preto: Simone Kashima, Evandra Strazza Rodrigues, Svetoslav Nanev Slavov, Elaine Vieira dos Santos, Rafael dos Santos Bezerra, Luiz Carlos Junior de Alcantara, Marta Giovanetti, Vagner Fonseca, Flavia Aburjaile, Rodrigo Tocantins Calado. FAMERP-SJRP: Cecília Artico Banho, Lívia Sacchetto, Fábio Sossai Possebon, Leila Sabrina Ullmann, Cíntia Bittar, Guilherme Campos, Helena Lage Ferreira, Jorge A. Petroli Marchesi, Maisa C. Pereira Parra, Marília Moraes, Paula Rahal, Paulo Inacio da Costa, João Pessoa Araújo Jr., Maurício Lacerda Nogueira. Prefeitura de Sao Paulo: Melissa Palmieri. |
| EPI_ISL_1966913, EPI_ISL_1966914 | HOSPITAL MUNICIPAL DE IBIUNA IBIUNA SP | Instituto Butantan / ESALQ-USP (Piracicaba) | Instituto Butantan: Dimas Tadeu Covas, Sandra Coccuzzo Sampaio, Maria Carolina Elias, José Salvatore Leister Patané, Vincent Louis Viala, Antonio Jorge Martins, Ricardo Haddad, Claudia Renata dos Santos Barros, Elaine Cristina Marqueze, Raul Machado Neto, Debora Botequio Moretti, Jardelina de |

| | | | |
|---|---|---|---|
| | | | Souza Todao Bernardino, Loyze Paola Oliveira de Lima, Luiz Aurelio de Campos Crispin. Centro de Genômica Funcional da ESALQ: Luiz Lehmann Coutinho, Ricardo Augusto Brassaloti, Raquel de Lello Rocha Campos Cassano. NGS Soluções Genômicas: Pilar Drummond Sampaio Corrêa Mariani. FZEA-USP Pirassununga: Mirele Daiana Poleti, Jessika Cristina Chagas Lesbon, Elisangela Chicaroni Mattos, Heidge Fukumasu. USP-Botucatu: Rejane Maria Tommasini Grotto, Jayme A. Souza-Neto, Guilherme Targino Valente, Patricia Akemi Assato, Felipe Allan da Silva da Costa, Bianca Cechetto Carlos. Mendelics: Bibiana Santos, João Paulo Kitajima, Erika Freitas, David Schlesinger. Hemocentro Ribeirão Preto: Simone Kashima, Evandra Strazza Rodrigues, Svetoslav Nanev Slavov, Elaine Vieira dos Santos, Rafael dos Santos Bezerra, Luiz Carlos Junior de Alcantara, Marta Giovanetti, Vagner Fonseca, Flavia Aburjaile, Rodrigo Tocantins Calado. FAMERP-SJRP: Cecília Artico Banho, Lívia Sacchetto, Fábio Sossai Possebon, Leila Sabrina Ullmann, Cíntia Bittar, Guilherme Campos, Helena Lage Ferreira, Jorge A. Petroli Marchesi, Maisa C. Pereira Parra, Marília Moraes, Paula Rahal, Paulo Inacio da Costa, João Pessoa Araújo Jr., Maurício Lacerda Nogueira. Prefeitura de Sao Paulo: Melissa Palmieri. |
| EPI_ISL_1966915 | UNIDADE MISTA DE IGUAPE | Instituto Butantan / ESALQ-USP (Piracicaba) | Instituto Butantan: Dimas Tadeu Covas, Sandra Coccuzzo Sampaio, Maria Carolina Elias, José Salvatore Leister Patané, Vincent Louis Viala, Antonio Jorge Martins, Ricardo Haddad, Claudia Renata dos Santos Barros, Elaine Cristina Marqueze, Raul Machado Neto, Debora Botequio Moretti, Jardelina de Souza Todao Bernardino, Loyze Paola Oliveira de Lima, Luiz Aurelio de Campos Crispin. Centro de Genômica Funcional da ESALQ: Luiz Lehmann Coutinho, Ricardo Augusto Brassaloti, Raquel de Lello Rocha Campos Cassano. NGS Soluções Genômicas: Pilar Drummond Sampaio Corrêa Mariani. FZEA-USP Pirassununga: Mirele Daiana Poleti, Jessika Cristina Chagas Lesbon, Elisangela Chicaroni Mattos, Heidge Fukumasu. USP-Botucatu: Rejane Maria Tommasini Grotto, Jayme A. Souza-Neto, Guilherme Targino Valente, Patricia Akemi Assato, Felipe Allan da Silva da Costa, Bianca Cechetto Carlos. Mendelics: Bibiana Santos, João Paulo Kitajima, Erika Freitas, David Schlesinger. Hemocentro Ribeirão Preto: Simone Kashima, Evandra Strazza Rodrigues, Svetoslav Nanev Slavov, Elaine Vieira dos Santos, Rafael dos Santos Bezerra, Luiz Carlos Junior de Alcantara, Marta Giovanetti, Vagner Fonseca, Flavia Aburjaile, Rodrigo Tocantins Calado. FAMERP-SJRP: Cecília Artico Banho, Lívia Sacchetto, Fábio Sossai Possebon, Leila Sabrina Ullmann, Cíntia Bittar, Guilherme Campos, Helena Lage Ferreira, Jorge A. Petroli Marchesi, Maisa C. Pereira Parra, Marília Moraes, Paula Rahal, Paulo Inacio da Costa, João Pessoa Araújo Jr., Maurício Lacerda Nogueira. Prefeitura de Sao Paulo: Melissa Palmieri. |
| EPI_ISL_1966916 | PENITENCIARIA ODON RAMOS MARANHAO IPERO | Instituto Butantan / ESALQ-USP (Piracicaba) | Instituto Butantan: Dimas Tadeu Covas, Sandra Coccuzzo Sampaio, Maria Carolina Elias, José Salvatore Leister Patané, Vincent Louis Viala, Antonio Jorge Martins, Ricardo Haddad, Claudia Renata dos Santos Barros, Elaine Cristina Marqueze, Raul Machado Neto, Debora Botequio Moretti, Jardelina de Souza Todao Bernardino, Loyze Paola Oliveira de Lima, Luiz Aurelio de Campos Crispin. Centro de Genômica Funcional da ESALQ: Luiz Lehmann Coutinho, Ricardo Augusto Brassaloti, Raquel de Lello Rocha Campos Cassano. NGS Soluções Genômicas: Pilar Drummond Sampaio Corrêa Mariani. FZEA-USP Pirassununga: Mirele Daiana Poleti, Jessika Cristina Chagas Lesbon, Elisangela Chicaroni Mattos, Heidge Fukumasu. USP-Botucatu: Rejane Maria Tommasini Grotto, Jayme A. Souza-Neto, Guilherme Targino Valente, Patricia Akemi Assato, Felipe Allan da Silva da Costa, Bianca Cechetto Carlos. Mendelics: Bibiana Santos, João Paulo Kitajima, Erika Freitas, David Schlesinger. Hemocentro Ribeirão Preto: Simone Kashima, Evandra Strazza Rodrigues, Svetoslav Nanev Slavov, Elaine Vieira dos Santos, Rafael dos Santos Bezerra, Luiz Carlos Junior de Alcantara, Marta Giovanetti, Vagner Fonseca, Flavia Aburjaile, Rodrigo Tocantins Calado. FAMERP-SJRP: Cecília Artico Banho, Lívia Sacchetto, Fábio Sossai Possebon, Leila Sabrina Ullmann, Cíntia Bittar, Guilherme Campos, Helena Lage Ferreira, Jorge A. Petroli Marchesi, Maisa C. Pereira Parra, Marília Moraes, Paula Rahal, Paulo Inacio da Costa, João Pessoa Araújo Jr., Maurício Lacerda Nogueira. Prefeitura de Sao Paulo: Melissa Palmieri. |
| EPI_ISL_1966918 | UBS JARDIM VITORINO IPERO | Instituto Butantan / ESALQ-USP (Piracicaba) | Instituto Butantan: Dimas Tadeu Covas, Sandra Coccuzzo Sampaio, Maria Carolina Elias, José Salvatore Leister Patané, Vincent Louis Viala, Antonio Jorge Martins, Ricardo Haddad, Claudia Renata dos Santos Barros, Elaine Cristina Marqueze, Raul Machado Neto, Debora Botequio Moretti, Jardelina de Souza Todao Bernardino, Loyze Paola Oliveira de Lima, Luiz Aurelio de Campos Crispin. Centro de Genômica Funcional da ESALQ: Luiz Lehmann Coutinho, Ricardo Augusto Brassaloti, Raquel de Lello Rocha Campos Cassano. NGS Soluções Genômicas: Pilar Drummond Sampaio Corrêa Mariani. FZEA-USP Pirassununga: Mirele Daiana Poleti, Jessika Cristina Chagas Lesbon, Elisangela Chicaroni Mattos, Heidge Fukumasu. USP-Botucatu: Rejane Maria Tommasini Grotto, Jayme A. Souza-Neto, Guilherme Targino Valente, Patricia Akemi Assato, Felipe Allan da Silva da Costa, Bianca Cechetto Carlos. Mendelics: Bibiana Santos, João Paulo Kitajima, Erika Freitas, David Schlesinger. Hemocentro Ribeirão Preto: Simone Kashima, Evandra Strazza Rodrigues, Svetoslav Nanev Slavov, Elaine Vieira dos Santos, Rafael dos Santos Bezerra, Luiz Carlos Junior de Alcantara, Marta Giovanetti, Vagner Fonseca, Flavia Aburjaile, Rodrigo Tocantins Calado. FAMERP-SJRP: Cecília Artico Banho, Lívia Sacchetto, Fábio Sossai Possebon, Leila Sabrina Ullmann, Cíntia Bittar, Guilherme Campos, Helena Lage Ferreira, Jorge A. Petroli Marchesi, Maisa C. Pereira Parra, Marília Moraes, Paula Rahal, Paulo Inacio da Costa, João Pessoa Araújo Jr., Maurício Lacerda Nogueira. Prefeitura de Sao Paulo: Melissa Palmieri. |
| EPI_ISL_1966919, EPI_ISL_1966920, EPI_ISL_1966921 | HOSPITAL MUNICIPAL DE ITABERA | Instituto Butantan / ESALQ-USP (Piracicaba) | Instituto Butantan: Dimas Tadeu Covas, Sandra Coccuzzo Sampaio, Maria Carolina Elias, José Salvatore Leister Patané, Vincent Louis Viala, Antonio Jorge Martins, Ricardo Haddad, Claudia Renata dos Santos Barros, Elaine Cristina Marqueze, Raul Machado Neto, Debora Botequio Moretti, Jardelina de Souza Todao Bernardino, Loyze Paola Oliveira de Lima, Luiz Aurelio de Campos Crispin. Centro de Genômica Funcional da ESALQ: Luiz Lehmann Coutinho, Ricardo Augusto Brassaloti, Raquel de Lello Rocha Campos Cassano. NGS Soluções Genômicas: Pilar Drummond Sampaio Corrêa Mariani. FZEA-USP Pirassununga: Mirele Daiana Poleti, Jessika Cristina Chagas Lesbon, Elisangela Chicaroni Mattos, Heidge Fukumasu. USP-Botucatu: Rejane Maria Tommasini Grotto, Jayme A. Souza-Neto, Guilherme Targino Valente, Patricia Akemi Assato, Felipe Allan da Silva da Costa, Bianca Cechetto Carlos. Mendelics: Bibiana Santos, João Paulo Kitajima, Erika Freitas, David Schlesinger. Hemocentro Ribeirão Preto: Simone Kashima, Evandra Strazza Rodrigues, Svetoslav Nanev Slavov, Elaine Vieira dos Santos, Rafael dos Santos Bezerra, Luiz Carlos Junior de Alcantara, Marta Giovanetti, Vagner Fonseca, Flavia Aburjaile, Rodrigo Tocantins Calado. FAMERP-SJRP: Cecília Artico Banho, Lívia Sacchetto, Fábio Sossai Possebon, Leila Sabrina Ullmann, Cíntia Bittar, Guilherme Campos, Helena Lage Ferreira, Jorge A. Petroli Marchesi, Maisa C. Pereira Parra, Marília Moraes, Paula Rahal, Paulo Inacio da Costa, João Pessoa Araújo Jr., Maurício Lacerda Nogueira. Prefeitura de Sao Paulo: Melissa Palmieri. |
| EPI_ISL_1966923, EPI_ISL_1966924, EPI_ISL_1966925, EPI_ISL_1966926 | HOSPITAL REGIONAL DE ITAPETININGA | Instituto Butantan / ESALQ-USP (Piracicaba) | Instituto Butantan: Dimas Tadeu Covas, Sandra Coccuzzo Sampaio, Maria Carolina Elias, José Salvatore Leister Patané, Vincent Louis Viala, Antonio Jorge Martins, Ricardo Haddad, Claudia Renata dos Santos Barros, Elaine Cristina Marqueze, Raul Machado Neto, Debora Botequio Moretti, Jardelina de Souza Todao Bernardino, Loyze Paola Oliveira de Lima, Luiz Aurelio de Campos Crispin. Centro de Genômica Funcional da ESALQ: Luiz Lehmann Coutinho, Ricardo Augusto Brassaloti, Raquel de Lello Rocha Campos Cassano. NGS Soluções Genômicas: Pilar Drummond Sampaio Corrêa Mariani. FZEA-USP Pirassununga: Mirele Daiana Poleti, Jessika Cristina Chagas Lesbon, Elisangela Chicaroni Mattos, Heidge Fukumasu. USP-Botucatu: Rejane Maria Tommasini Grotto, Jayme A. Souza-Neto, Guilherme Targino Valente, Patricia Akemi Assato, Felipe Allan da Silva da Costa, Bianca Cechetto Carlos. Mendelics: Bibiana Santos, João Paulo Kitajima, Erika Freitas, David Schlesinger. Hemocentro Ribeirão Preto: Simone Kashima, Evandra Strazza Rodrigues, Svetoslav Nanev Slavov, Elaine Vieira dos Santos, Rafael dos Santos Bezerra, Luiz Carlos Junior de Alcantara, Marta Giovanetti, Vagner Fonseca, Flavia Aburjaile, Rodrigo Tocantins Calado. FAMERP-SJRP: Cecília Artico Banho, Lívia Sacchetto, Fábio Sossai Possebon, Leila Sabrina Ullmann, Cíntia Bittar, Guilherme Campos, Helena Lage Ferreira, Jorge A. Petroli Marchesi, Maisa C. Pereira Parra, Marília Moraes, Paula Rahal, Paulo Inacio da Costa, João Pessoa Araújo Jr., Maurício Lacerda Nogueira. Prefeitura de Sao Paulo: Melissa Palmieri. |
| EPI_ISL_1966927, EPI_ISL_1966928, EPI_ISL_1966929, EPI_ISL_1966930, EPI_ISL_1966931, EPI_ISL_1966932, EPI_ISL_1966933, EPI_ISL_1966934 | POLICLINICA COVID 19 ITAPETININGA | Instituto Butantan / ESALQ-USP (Piracicaba) | Instituto Butantan: Dimas Tadeu Covas, Sandra Coccuzzo Sampaio, Maria Carolina Elias, José Salvatore Leister Patané, Vincent Louis Viala, Antonio Jorge Martins, Ricardo Haddad, Claudia Renata dos Santos Barros, Elaine Cristina Marqueze, Raul Machado Neto, Debora Botequio Moretti, Jardelina de Souza Todao Bernardino, Loyze Paola Oliveira de Lima, Luiz Aurelio de Campos Crispin. Centro de Genômica Funcional da ESALQ: Luiz Lehmann Coutinho, Ricardo Augusto Brassaloti, Raquel de Lello Rocha Campos Cassano. NGS Soluções Genômicas: Pilar Drummond Sampaio Corrêa Mariani. FZEA-USP Pirassununga: Mirele Daiana Poleti, Jessika Cristina Chagas Lesbon, Elisangela Chicaroni Mattos, Heidge Fukumasu. USP-Botucatu: Rejane Maria Tommasini Grotto, Jayme A. Souza-Neto, Guilherme Targino Valente, Patricia Akemi Assato, Felipe Allan da Silva da Costa, Bianca Cechetto Carlos. Mendelics: Bibiana Santos, João Paulo Kitajima, Erika Freitas, David Schlesinger. Hemocentro Ribeirão Preto: Simone Kashima, Evandra Strazza Rodrigues, Svetoslav Nanev Slavov, Elaine Vieira dos Santos, Rafael dos Santos Bezerra, Luiz Carlos Junior de Alcantara, Marta Giovanetti, Vagner Fonseca, Flavia Aburjaile, Rodrigo Tocantins Calado. FAMERP-SJRP: Cecília Artico Banho, Lívia Sacchetto, Fábio Sossai Possebon, Leila Sabrina Ullmann, Cíntia Bittar, Guilherme Campos, Helena Lage Ferreira, Jorge A. Petroli Marchesi, Maisa C. Pereira Parra, Marília Moraes, Paula Rahal, Paulo Inacio da Costa, João Pessoa Araújo Jr., Maurício Lacerda Nogueira. Prefeitura de Sao Paulo: Melissa Palmieri. |
| EPI_ISL_1966935, EPI_ISL_1966936, EPI_ISL_1966937, EPI_ISL_1966938, EPI_ISL_1966939, EPI_ISL_1966940, EPI_ISL_1966941 | VIGILANCIA SANITARIA E VIG EPIDEMIOLOGICA DE ITAPEVI | Instituto Butantan / ESALQ-USP (Piracicaba) | Instituto Butantan: Dimas Tadeu Covas, Sandra Coccuzzo Sampaio, Maria Carolina Elias, José Salvatore Leister Patané, Vincent Louis Viala, Antonio Jorge Martins, Ricardo Haddad, Claudia Renata dos Santos Barros, Elaine Cristina Marqueze, Raul Machado Neto, Debora Botequio Moretti, Jardelina de Souza Todao Bernardino, Loyze Paola Oliveira de Lima, Luiz Aurelio de Campos Crispin. Centro de Genômica Funcional da ESALQ: Luiz Lehmann Coutinho, Ricardo Augusto Brassaloti, Raquel de Lello Rocha Campos Cassano. NGS Soluções Genômicas: Pilar Drummond Sampaio Corrêa Mariani. FZEA-USP Pirassununga: Mirele Daiana Poleti, Jessika Cristina Chagas Lesbon, Elisangela Chicaroni Mattos, Heidge Fukumasu. USP-Botucatu: Rejane Maria Tommasini Grotto, Jayme A. Souza-Neto, Guilherme Targino Valente, Patricia Akemi Assato, Felipe Allan da Silva da Costa, Bianca Cechetto Carlos. Mendelics: Bibiana Santos, João Paulo Kitajima, Erika Freitas, David Schlesinger. Hemocentro Ribeirão Preto: Simone Kashima, Evandra Strazza Rodrigues, Svetoslav Nanev Slavov, Elaine Vieira dos Santos, Rafael dos Santos Bezerra, Luiz Carlos Junior de Alcantara, Marta Giovanetti, Vagner Fonseca, Flavia Aburjaile, Rodrigo Tocantins Calado. FAMERP-SJRP: Cecília Artico Banho, Lívia Sacchetto, Fábio Sossai Possebon, Leila Sabrina |

| | | | |
|---|---|---|---|
| | | | Ullmann, Cíntia Bittar, Guilherme Campos, Helena Lage Ferreira, Jorge A. Petroli Marchesi, Maisa C. Pereira Parra, Marília Moraes, Paula Rahal, Paulo Inacio da Costa, João Pessoa Araújo Jr., Maurício Lacerda Nogueira. Prefeitura de Sao Paulo: Melissa Palmieri. |
| EPI_ISL_1966942, EPI_ISL_1966945, EPI_ISL_1966946, EPI_ISL_1966947, EPI_ISL_1966949 | HOSPITAL MUNICIPAL DE ITAPIRA | Instituto Butantan / ESALQ-USP (Piracicaba) | Instituto Butantan: Dimas Tadeu Covas, Sandra Coccuzzo Sampaio, Maria Carolina Elias, José Salvatore Leister Patané, Vincent Louis Viala, Antonio Jorge Martins, Ricardo Haddad, Claudia Renata dos Santos Barros, Elaine Cristina Marqueze, Raul Machado Neto, Debora Botequio Moretti, Jardelina de Souza Todao Bernardino, Loyze Paola Oliveira de Lima, Luiz Aurelio de Campos Crispin. Centro de Genômica Funcional da ESALQ: Luiz Lehmann Coutinho, Ricardo Augusto Brassaloti, Raquel de Lello Rocha Campos Cassano. NGS Soluções Genômicas: Pilar Drummond Sampaio Corrêa Mariani. FZEA-USP Pirassununga: Mirele Daiana Poleti, Jessika Cristina Chagas Lesbon, Elisangela Chicaroni Mattos, Heidge Fukumasu. USP-Botucatu: Rejane Maria Tommasini Grotto, Jayme A. Souza-Neto, Guilherme Targino Valente, Patricia Akemi Assato, Felipe Allan da Silva da Costa, Bianca Cechetto Carlos. Mendelics: Bibiana Santos, João Paulo Kitajima, Erika Freitas, David Schlesinger. Hemocentro Ribeirão Preto: Simone Kashima, Evandra Strazza Rodrigues, Svetoslav Nanev Slavov, Elaine Vieira dos Santos, Rafael dos Santos Bezerra, Luiz Carlos Junior de Alcantara, Marta Giovanetti, Vagner Fonseca, Flavia Aburjaile, Rodrigo Tocantins Calado. FAMERP-SJRP: Cecília Artico Banho, Lívia Sacchetto, Fábio Sossai Possebon, Leila Sabrina Ullmann, Cíntia Bittar, Guilherme Campos, Helena Lage Ferreira, Jorge A. Petroli Marchesi, Maisa C. Pereira Parra, Marília Moraes, Paula Rahal, Paulo Inacio da Costa, João Pessoa Araújo Jr., Maurício Lacerda Nogueira. Prefeitura de Sao Paulo: Melissa Palmieri. |
| EPI_ISL_1966950, EPI_ISL_1966951 | SECRETARIA MUNICIPAL DA SAUDE DE ITARARE | Instituto Butantan / ESALQ-USP (Piracicaba) | Instituto Butantan: Dimas Tadeu Covas, Sandra Coccuzzo Sampaio, Maria Carolina Elias, José Salvatore Leister Patané, Vincent Louis Viala, Antonio Jorge Martins, Ricardo Haddad, Claudia Renata dos Santos Barros, Elaine Cristina Marqueze, Raul Machado Neto, Debora Botequio Moretti, Jardelina de Souza Todao Bernardino, Loyze Paola Oliveira de Lima, Luiz Aurelio de Campos Crispin. Centro de Genômica Funcional da ESALQ: Luiz Lehmann Coutinho, Ricardo Augusto Brassaloti, Raquel de Lello Rocha Campos Cassano. NGS Soluções Genômicas: Pilar Drummond Sampaio Corrêa Mariani. FZEA-USP Pirassununga: Mirele Daiana Poleti, Jessika Cristina Chagas Lesbon, Elisangela Chicaroni Mattos, Heidge Fukumasu. USP-Botucatu: Rejane Maria Tommasini Grotto, Jayme A. Souza-Neto, Guilherme Targino Valente, Patricia Akemi Assato, Felipe Allan da Silva da Costa, Bianca Cechetto Carlos. Mendelics: Bibiana Santos, João Paulo Kitajima, Erika Freitas, David Schlesinger. Hemocentro Ribeirão Preto: Simone Kashima, Evandra Strazza Rodrigues, Svetoslav Nanev Slavov, Elaine Vieira dos Santos, Rafael dos Santos Bezerra, Luiz Carlos Junior de Alcantara, Marta Giovanetti, Vagner Fonseca, Flavia Aburjaile, Rodrigo Tocantins Calado. FAMERP-SJRP: Cecília Artico Banho, Lívia Sacchetto, Fábio Sossai Possebon, Leila Sabrina Ullmann, Cíntia Bittar, Guilherme Campos, Helena Lage Ferreira, Jorge A. Petroli Marchesi, Maisa C. Pereira Parra, Marília Moraes, Paula Rahal, Paulo Inacio da Costa, João Pessoa Araújo Jr., Maurício Lacerda Nogueira. Prefeitura de Sao Paulo: Melissa Palmieri. |
| EPI_ISL_1966952, EPI_ISL_1966953 | PRONTO ATENDIMENTO MUNICIPAL DE JACUPIRANGA | Instituto Butantan / ESALQ-USP (Piracicaba) | Instituto Butantan: Dimas Tadeu Covas, Sandra Coccuzzo Sampaio, Maria Carolina Elias, José Salvatore Leister Patané, Vincent Louis Viala, Antonio Jorge Martins, Ricardo Haddad, Claudia Renata dos Santos Barros, Elaine Cristina Marqueze, Raul Machado Neto, Debora Botequio Moretti, Jardelina de Souza Todao Bernardino, Loyze Paola Oliveira de Lima, Luiz Aurelio de Campos Crispin. Centro de Genômica Funcional da ESALQ: Luiz Lehmann Coutinho, Ricardo Augusto Brassaloti, Raquel de Lello Rocha Campos Cassano. NGS Soluções Genômicas: Pilar Drummond Sampaio Corrêa Mariani. FZEA-USP Pirassununga: Mirele Daiana Poleti, Jessika Cristina Chagas Lesbon, Elisangela Chicaroni Mattos, Heidge Fukumasu. USP-Botucatu: Rejane Maria Tommasini Grotto, Jayme A. Souza-Neto, Guilherme Targino Valente, Patricia Akemi Assato, Felipe Allan da Silva da Costa, Bianca Cechetto Carlos. Mendelics: Bibiana Santos, João Paulo Kitajima, Erika Freitas, David Schlesinger. Hemocentro Ribeirão Preto: Simone Kashima, Evandra Strazza Rodrigues, Svetoslav Nanev Slavov, Elaine Vieira dos Santos, Rafael dos Santos Bezerra, Luiz Carlos Junior de Alcantara, Marta Giovanetti, Vagner Fonseca, Flavia Aburjaile, Rodrigo Tocantins Calado. FAMERP-SJRP: Cecília Artico Banho, Lívia Sacchetto, Fábio Sossai Possebon, Leila Sabrina Ullmann, Cíntia Bittar, Guilherme Campos, Helena Lage Ferreira, Jorge A. Petroli Marchesi, Maisa C. Pereira Parra, Marília Moraes, Paula Rahal, Paulo Inacio da Costa, João Pessoa Araújo Jr., Maurício Lacerda Nogueira. Prefeitura de Sao Paulo: Melissa Palmieri. |
| EPI_ISL_1966954 | HOSPITAL SANTO ANTONIO DE JUQUIA JUQUIA | Instituto Butantan / ESALQ-USP (Piracicaba) | Instituto Butantan: Dimas Tadeu Covas, Sandra Coccuzzo Sampaio, Maria Carolina Elias, José Salvatore Leister Patané, Vincent Louis Viala, Antonio Jorge Martins, Ricardo Haddad, Claudia Renata dos Santos Barros, Elaine Cristina Marqueze, Raul Machado Neto, Debora Botequio Moretti, Jardelina de Souza Todao Bernardino, Loyze Paola Oliveira de Lima, Luiz Aurelio de Campos Crispin. Centro de Genômica Funcional da ESALQ: Luiz Lehmann Coutinho, Ricardo Augusto Brassaloti, Raquel de Lello Rocha Campos Cassano. NGS Soluções Genômicas: Pilar Drummond Sampaio Corrêa Mariani. FZEA-USP Pirassununga: Mirele Daiana Poleti, Jessika Cristina Chagas Lesbon, Elisangela Chicaroni Mattos, Heidge Fukumasu. USP-Botucatu: Rejane Maria Tommasini Grotto, Jayme A. Souza-Neto, Guilherme Targino Valente, Patricia Akemi Assato, Felipe Allan da Silva da Costa, Bianca Cechetto Carlos. Mendelics: Bibiana Santos, João Paulo Kitajima, Erika Freitas, David Schlesinger. Hemocentro Ribeirão Preto: Simone Kashima, Evandra Strazza Rodrigues, Svetoslav Nanev Slavov, Elaine Vieira dos Santos, Rafael dos Santos Bezerra, Luiz Carlos Junior de Alcantara, Marta Giovanetti, Vagner Fonseca, Flavia Aburjaile, Rodrigo Tocantins Calado. FAMERP-SJRP: Cecília Artico Banho, Lívia Sacchetto, Fábio Sossai Possebon, Leila Sabrina Ullmann, Cíntia Bittar, Guilherme Campos, Helena Lage Ferreira, Jorge A. Petroli Marchesi, Maisa C. Pereira Parra, Marília Moraes, Paula Rahal, Paulo Inacio da Costa, João Pessoa Araújo Jr., Maurício Lacerda Nogueira. Prefeitura de Sao Paulo: Melissa Palmieri. |
| EPI_ISL_1966955, EPI_ISL_1966956 | CENTRO DE SAUDE II MAIRINQUE MAIRINQUE | Instituto Butantan / ESALQ-USP (Piracicaba) | Instituto Butantan: Dimas Tadeu Covas, Sandra Coccuzzo Sampaio, Maria Carolina Elias, José Salvatore Leister Patané, Vincent Louis Viala, Antonio Jorge Martins, Ricardo Haddad, Claudia Renata dos Santos Barros, Elaine Cristina Marqueze, Raul Machado Neto, Debora Botequio Moretti, Jardelina de Souza Todao Bernardino, Loyze Paola Oliveira de Lima, Luiz Aurelio de Campos Crispin. Centro de Genômica Funcional da ESALQ: Luiz Lehmann Coutinho, Ricardo Augusto Brassaloti, Raquel de Lello Rocha Campos Cassano. NGS Soluções Genômicas: Pilar Drummond Sampaio Corrêa Mariani. FZEA-USP Pirassununga: Mirele Daiana Poleti, Jessika Cristina Chagas Lesbon, Elisangela Chicaroni Mattos, Heidge Fukumasu. USP-Botucatu: Rejane Maria Tommasini Grotto, Jayme A. Souza-Neto, Guilherme Targino Valente, Patricia Akemi Assato, Felipe Allan da Silva da Costa, Bianca Cechetto Carlos. Mendelics: Bibiana Santos, João Paulo Kitajima, Erika Freitas, David Schlesinger. Hemocentro Ribeirão Preto: Simone Kashima, Evandra Strazza Rodrigues, Svetoslav Nanev Slavov, Elaine Vieira dos Santos, Rafael dos Santos Bezerra, Luiz Carlos Junior de Alcantara, Marta Giovanetti, Vagner Fonseca, Flavia Aburjaile, Rodrigo Tocantins Calado. FAMERP-SJRP: Cecília Artico Banho, Lívia Sacchetto, Fábio Sossai Possebon, Leila Sabrina Ullmann, Cíntia Bittar, Guilherme Campos, Helena Lage Ferreira, Jorge A. Petroli Marchesi, Maisa C. Pereira Parra, Marília Moraes, Paula Rahal, Paulo Inacio da Costa, João Pessoa Araújo Jr., Maurício Lacerda Nogueira. Prefeitura de Sao Paulo: Melissa Palmieri. |
| EPI_ISL_1966957, EPI_ISL_1966959, EPI_ISL_1966960 | CENTRO DE SAUDE DE MARACAI | Instituto Butantan / ESALQ-USP (Piracicaba) | Instituto Butantan: Dimas Tadeu Covas, Sandra Coccuzzo Sampaio, Maria Carolina Elias, José Salvatore Leister Patané, Vincent Louis Viala, Antonio Jorge Martins, Ricardo Haddad, Claudia Renata dos Santos Barros, Elaine Cristina Marqueze, Raul Machado Neto, Debora Botequio Moretti, Jardelina de Souza Todao Bernardino, Loyze Paola Oliveira de Lima, Luiz Aurelio de Campos Crispin. Centro de Genômica Funcional da ESALQ: Luiz Lehmann Coutinho, Ricardo Augusto Brassaloti, Raquel de Lello Rocha Campos Cassano. NGS Soluções Genômicas: Pilar Drummond Sampaio Corrêa Mariani. FZEA-USP Pirassununga: Mirele Daiana Poleti, Jessika Cristina Chagas Lesbon, Elisangela Chicaroni Mattos, Heidge Fukumasu. USP-Botucatu: Rejane Maria Tommasini Grotto, Jayme A. Souza-Neto, Guilherme Targino Valente, Patricia Akemi Assato, Felipe Allan da Silva da Costa, Bianca Cechetto Carlos. Mendelics: Bibiana Santos, João Paulo Kitajima, Erika Freitas, David Schlesinger. Hemocentro Ribeirão Preto: Simone Kashima, Evandra Strazza Rodrigues, Svetoslav Nanev Slavov, Elaine Vieira dos Santos, Rafael dos Santos Bezerra, Luiz Carlos Junior de Alcantara, Marta Giovanetti, Vagner Fonseca, Flavia Aburjaile, Rodrigo Tocantins Calado. FAMERP-SJRP: Cecília Artico Banho, Lívia Sacchetto, Fábio Sossai Possebon, Leila Sabrina Ullmann, Cíntia Bittar, Guilherme Campos, Helena Lage Ferreira, Jorge A. Petroli Marchesi, Maisa C. Pereira Parra, Marília Moraes, Paula Rahal, Paulo Inacio da Costa, João Pessoa Araújo Jr., Maurício Lacerda Nogueira. Prefeitura de Sao Paulo: Melissa Palmieri. |
| EPI_ISL_1966961, EPI_ISL_1966962, EPI_ISL_1966963, EPI_ISL_1966964 | CS DE MONTE APRAZIVEL | Instituto Butantan / ESALQ-USP (Piracicaba) | Instituto Butantan: Dimas Tadeu Covas, Sandra Coccuzzo Sampaio, Maria Carolina Elias, José Salvatore Leister Patané, Vincent Louis Viala, Antonio Jorge Martins, Ricardo Haddad, Claudia Renata dos Santos Barros, Elaine Cristina Marqueze, Raul Machado Neto, Debora Botequio Moretti, Jardelina de Souza Todao Bernardino, Loyze Paola Oliveira de Lima, Luiz Aurelio de Campos Crispin. Centro de Genômica Funcional da ESALQ: Luiz Lehmann Coutinho, Ricardo Augusto Brassaloti, Raquel de Lello Rocha Campos Cassano. NGS Soluções Genômicas: Pilar Drummond Sampaio Corrêa Mariani. FZEA-USP Pirassununga: Mirele Daiana Poleti, Jessika Cristina Chagas Lesbon, Elisangela Chicaroni Mattos, Heidge Fukumasu. USP-Botucatu: Rejane Maria Tommasini Grotto, Jayme A. Souza-Neto, Guilherme Targino Valente, Patricia Akemi Assato, Felipe Allan da Silva da Costa, Bianca Cechetto Carlos. Mendelics: Bibiana Santos, João Paulo Kitajima, Erika Freitas, David Schlesinger. Hemocentro Ribeirão Preto: Simone Kashima, Evandra Strazza Rodrigues, Svetoslav Nanev Slavov, Elaine Vieira dos Santos, Rafael dos Santos Bezerra, Luiz Carlos Junior de Alcantara, Marta Giovanetti, Vagner Fonseca, Flavia Aburjaile, Rodrigo Tocantins Calado. FAMERP-SJRP: Cecília Artico Banho, Lívia Sacchetto, Fábio Sossai Possebon, Leila Sabrina Ullmann, Cíntia Bittar, Guilherme Campos, Helena Lage Ferreira, Jorge A. Petroli Marchesi, Maisa C. Pereira Parra, Marília Moraes, Paula Rahal, Paulo Inacio da Costa, João Pessoa Araújo Jr., Maurício Lacerda Nogueira. Prefeitura de Sao Paulo: Melissa Palmieri. |
| EPI_ISL_1966965, EPI_ISL_1966966, EPI_ISL_1966967, EPI_ISL_1966968, EPI_ISL_1966970, EPI_ISL_1966971, EPI_ISL_1966972, EPI_ISL_1966973, EPI_ISL_1966974, EPI_ISL_1966975, EPI_ISL_1966976, EPI_ISL_1966977, EPI_ISL_1966978, EPI_ISL_1966979, EPI_ISL_1966980, EPI_ISL_1966981, EPI_ISL_1966982, EPI_ISL_1966983 | | | |
| see above | CS II EGIDIO BRUNHARA MORRO AGUDO | Instituto Butantan / ESALQ-USP (Piracicaba) | Instituto Butantan: Dimas Tadeu Covas, Sandra Coccuzzo Sampaio, Maria Carolina Elias, José Salvatore Leister Patané, Vincent Louis Viala, Antonio Jorge Martins, Ricardo Haddad, Claudia Renata dos Santos Barros, Elaine Cristina Marqueze, Raul Machado Neto, Debora Botequio Moretti, Jardelina de |

| | | | |
|---|---|---|---|
| | | | Souza Todao Bernardino, Loyze Paola Oliveira de Lima, Luiz Aurelio de Campos Crispin. Centro de Genômica Funcional da ESALQ: Luiz Lehmann Coutinho, Ricardo Augusto Brassaloti, Raquel de Lello Rocha Campos Cassano. NGS Soluções Genômicas: Pilar Drummond Sampaio Corrêa Mariani. FZEA-USP Pirassununga: Mirele Daiana Poleti, Jessika Cristina Chagas Lesbon, Elisangela Chicaroni Mattos, Heidge Fukumasu. USP-Botucatu: Rejane Maria Tommasini Grotto, Jayme A. Souza-Neto, Guilherme Targino Valente, Patricia Akemi Assato, Felipe Allan da Silva da Costa, Bianca Cechetto Carlos. Mendelics: Bibiana Santos, João Paulo Kitajima, Erika Freitas, David Schlesinger. Hemocentro Ribeirão Preto: Simone Kashima, Evandra Strazza Rodrigues, Svetoslav Nanev Slavov, Elaine Vieira dos Santos, Rafael dos Santos Bezerra, Luiz Carlos Junior de Alcantara, Marta Giovanetti, Vagner Fonseca, Flavia Aburjaile, Rodrigo Tocantins Calado. FAMERP-SJRP: Cecília Artico Banho, Lívia Sacchetto, Fábio Sossai Possebon, Leila Sabrina Ullmann, Cíntia Bittar, Guilherme Campos, Helena Lage Ferreira, Jorge A. Petroli Marchesi, Maisa C. Pereira Parra, Marília Moraes, Paula Rahal, Paulo Inacio da Costa, João Pessoa Araújo Jr., Maurício Lacerda Nogueira. Prefeitura de Sao Paulo: Melissa Palmieri. |
| EPI_ISL_1966984, EPI_ISL_1966985 | CS DE NEVES PAULISTA | Instituto Butantan / ESALQ-USP (Piracicaba) | Instituto Butantan: Dimas Tadeu Covas, Sandra Coccuzzo Sampaio, Maria Carolina Elias, José Salvatore Leister Patané, Vincent Louis Viala, Antonio Jorge Martins, Ricardo Haddad, Claudia Renata dos Santos Barros, Elaine Cristina Marqueze, Raul Machado Neto, Debora Botequio Moretti, Jardelina de Souza Todao Bernardino, Loyze Paola Oliveira de Lima, Luiz Aurelio de Campos Crispin. Centro de Genômica Funcional da ESALQ: Luiz Lehmann Coutinho, Ricardo Augusto Brassaloti, Raquel de Lello Rocha Campos Cassano. NGS Soluções Genômicas: Pilar Drummond Sampaio Corrêa Mariani. FZEA-USP Pirassununga: Mirele Daiana Poleti, Jessika Cristina Chagas Lesbon, Elisangela Chicaroni Mattos, Heidge Fukumasu. USP-Botucatu: Rejane Maria Tommasini Grotto, Jayme A. Souza-Neto, Guilherme Targino Valente, Patricia Akemi Assato, Felipe Allan da Silva da Costa, Bianca Cechetto Carlos. Mendelics: Bibiana Santos, João Paulo Kitajima, Erika Freitas, David Schlesinger. Hemocentro Ribeirão Preto: Simone Kashima, Evandra Strazza Rodrigues, Svetoslav Nanev Slavov, Elaine Vieira dos Santos, Rafael dos Santos Bezerra, Luiz Carlos Junior de Alcantara, Marta Giovanetti, Vagner Fonseca, Flavia Aburjaile, Rodrigo Tocantins Calado. FAMERP-SJRP: Cecília Artico Banho, Lívia Sacchetto, Fábio Sossai Possebon, Leila Sabrina Ullmann, Cíntia Bittar, Guilherme Campos, Helena Lage Ferreira, Jorge A. Petroli Marchesi, Maisa C. Pereira Parra, Marília Moraes, Paula Rahal, Paulo Inacio da Costa, João Pessoa Araújo Jr., Maurício Lacerda Nogueira. Prefeitura de Sao Paulo: Melissa Palmieri. |
| EPI_ISL_1966986, EPI_ISL_1966987 | UNIDADE MISTA DE SAUDE DE ORIENTE | Instituto Butantan / ESALQ-USP (Piracicaba) | Instituto Butantan: Dimas Tadeu Covas, Sandra Coccuzzo Sampaio, Maria Carolina Elias, José Salvatore Leister Patané, Vincent Louis Viala, Antonio Jorge Martins, Ricardo Haddad, Claudia Renata dos Santos Barros, Elaine Cristina Marqueze, Raul Machado Neto, Debora Botequio Moretti, Jardelina de Souza Todao Bernardino, Loyze Paola Oliveira de Lima, Luiz Aurelio de Campos Crispin. Centro de Genômica Funcional da ESALQ: Luiz Lehmann Coutinho, Ricardo Augusto Brassaloti, Raquel de Lello Rocha Campos Cassano. NGS Soluções Genômicas: Pilar Drummond Sampaio Corrêa Mariani. FZEA-USP Pirassununga: Mirele Daiana Poleti, Jessika Cristina Chagas Lesbon, Elisangela Chicaroni Mattos, Heidge Fukumasu. USP-Botucatu: Rejane Maria Tommasini Grotto, Jayme A. Souza-Neto, Guilherme Targino Valente, Patricia Akemi Assato, Felipe Allan da Silva da Costa, Bianca Cechetto Carlos. Mendelics: Bibiana Santos, João Paulo Kitajima, Erika Freitas, David Schlesinger. Hemocentro Ribeirão Preto: Simone Kashima, Evandra Strazza Rodrigues, Svetoslav Nanev Slavov, Elaine Vieira dos Santos, Rafael dos Santos Bezerra, Luiz Carlos Junior de Alcantara, Marta Giovanetti, Vagner Fonseca, Flavia Aburjaile, Rodrigo Tocantins Calado. FAMERP-SJRP: Cecília Artico Banho, Lívia Sacchetto, Fábio Sossai Possebon, Leila Sabrina Ullmann, Cíntia Bittar, Guilherme Campos, Helena Lage Ferreira, Jorge A. Petroli Marchesi, Maisa C. Pereira Parra, Marília Moraes, Paula Rahal, Paulo Inacio da Costa, João Pessoa Araújo Jr., Maurício Lacerda Nogueira. Prefeitura de Sao Paulo: Melissa Palmieri. |
| EPI_ISL_1966988 | CS DE ORINDIUVA | Instituto Butantan / ESALQ-USP (Piracicaba) | Instituto Butantan: Dimas Tadeu Covas, Sandra Coccuzzo Sampaio, Maria Carolina Elias, José Salvatore Leister Patané, Vincent Louis Viala, Antonio Jorge Martins, Ricardo Haddad, Claudia Renata dos Santos Barros, Elaine Cristina Marqueze, Raul Machado Neto, Debora Botequio Moretti, Jardelina de Souza Todao Bernardino, Loyze Paola Oliveira de Lima, Luiz Aurelio de Campos Crispin. Centro de Genômica Funcional da ESALQ: Luiz Lehmann Coutinho, Ricardo Augusto Brassaloti, Raquel de Lello Rocha Campos Cassano. NGS Soluções Genômicas: Pilar Drummond Sampaio Corrêa Mariani. FZEA-USP Pirassununga: Mirele Daiana Poleti, Jessika Cristina Chagas Lesbon, Elisangela Chicaroni Mattos, Heidge Fukumasu. USP-Botucatu: Rejane Maria Tommasini Grotto, Jayme A. Souza-Neto, Guilherme Targino Valente, Patricia Akemi Assato, Felipe Allan da Silva da Costa, Bianca Cechetto Carlos. Mendelics: Bibiana Santos, João Paulo Kitajima, Erika Freitas, David Schlesinger. Hemocentro Ribeirão Preto: Simone Kashima, Evandra Strazza Rodrigues, Svetoslav Nanev Slavov, Elaine Vieira dos Santos, Rafael dos Santos Bezerra, Luiz Carlos Junior de Alcantara, Marta Giovanetti, Vagner Fonseca, Flavia Aburjaile, Rodrigo Tocantins Calado. FAMERP-SJRP: Cecília Artico Banho, Lívia Sacchetto, Fábio Sossai Possebon, Leila Sabrina Ullmann, Cíntia Bittar, Guilherme Campos, Helena Lage Ferreira, Jorge A. Petroli Marchesi, Maisa C. Pereira Parra, Marília Moraes, Paula Rahal, Paulo Inacio da Costa, João Pessoa Araújo Jr., Maurício Lacerda Nogueira. Prefeitura de Sao Paulo: Melissa Palmieri. |
| EPI_ISL_1966989 | CS II DR MIGUEL VITALIANO ORLANDIA | Instituto Butantan / ESALQ-USP (Piracicaba) | Instituto Butantan: Dimas Tadeu Covas, Sandra Coccuzzo Sampaio, Maria Carolina Elias, José Salvatore Leister Patané, Vincent Louis Viala, Antonio Jorge Martins, Ricardo Haddad, Claudia Renata dos Santos Barros, Elaine Cristina Marqueze, Raul Machado Neto, Debora Botequio Moretti, Jardelina de Souza Todao Bernardino, Loyze Paola Oliveira de Lima, Luiz Aurelio de Campos Crispin. Centro de Genômica Funcional da ESALQ: Luiz Lehmann Coutinho, Ricardo Augusto Brassaloti, Raquel de Lello Rocha Campos Cassano. NGS Soluções Genômicas: Pilar Drummond Sampaio Corrêa Mariani. FZEA-USP Pirassununga: Mirele Daiana Poleti, Jessika Cristina Chagas Lesbon, Elisangela Chicaroni Mattos, Heidge Fukumasu. USP-Botucatu: Rejane Maria Tommasini Grotto, Jayme A. Souza-Neto, Guilherme Targino Valente, Patricia Akemi Assato, Felipe Allan da Silva da Costa, Bianca Cechetto Carlos. Mendelics: Bibiana Santos, João Paulo Kitajima, Erika Freitas, David Schlesinger. Hemocentro Ribeirão Preto: Simone Kashima, Evandra Strazza Rodrigues, Svetoslav Nanev Slavov, Elaine Vieira dos Santos, Rafael dos Santos Bezerra, Luiz Carlos Junior de Alcantara, Marta Giovanetti, Vagner Fonseca, Flavia Aburjaile, Rodrigo Tocantins Calado. FAMERP-SJRP: Cecília Artico Banho, Lívia Sacchetto, Fábio Sossai Possebon, Leila Sabrina Ullmann, Cíntia Bittar, Guilherme Campos, Helena Lage Ferreira, Jorge A. Petroli Marchesi, Maisa C. Pereira Parra, Marília Moraes, Paula Rahal, Paulo Inacio da Costa, João Pessoa Araújo Jr., Maurício Lacerda Nogueira. Prefeitura de Sao Paulo: Melissa Palmieri. |
| EPI_ISL_1966990 | UBS DR HELIO MIGLIARI | Instituto Butantan / ESALQ-USP (Piracicaba) | Instituto Butantan: Dimas Tadeu Covas, Sandra Coccuzzo Sampaio, Maria Carolina Elias, José Salvatore Leister Patané, Vincent Louis Viala, Antonio Jorge Martins, Ricardo Haddad, Claudia Renata dos Santos Barros, Elaine Cristina Marqueze, Raul Machado Neto, Debora Botequio Moretti, Jardelina de Souza Todao Bernardino, Loyze Paola Oliveira de Lima, Luiz Aurelio de Campos Crispin. Centro de Genômica Funcional da ESALQ: Luiz Lehmann Coutinho, Ricardo Augusto Brassaloti, Raquel de Lello Rocha Campos Cassano. NGS Soluções Genômicas: Pilar Drummond Sampaio Corrêa Mariani. FZEA-USP Pirassununga: Mirele Daiana Poleti, Jessika Cristina Chagas Lesbon, Elisangela Chicaroni Mattos, Heidge Fukumasu. USP-Botucatu: Rejane Maria Tommasini Grotto, Jayme A. Souza-Neto, Guilherme Targino Valente, Patricia Akemi Assato, Felipe Allan da Silva da Costa, Bianca Cechetto Carlos. Mendelics: Bibiana Santos, João Paulo Kitajima, Erika Freitas, David Schlesinger. Hemocentro Ribeirão Preto: Simone Kashima, Evandra Strazza Rodrigues, Svetoslav Nanev Slavov, Elaine Vieira dos Santos, Rafael dos Santos Bezerra, Luiz Carlos Junior de Alcantara, Marta Giovanetti, Vagner Fonseca, Flavia Aburjaile, Rodrigo Tocantins Calado. FAMERP-SJRP: Cecília Artico Banho, Lívia Sacchetto, Fábio Sossai Possebon, Leila Sabrina Ullmann, Cíntia Bittar, Guilherme Campos, Helena Lage Ferreira, Jorge A. Petroli Marchesi, Maisa C. Pereira Parra, Marília Moraes, Paula Rahal, Paulo Inacio da Costa, João Pessoa Araújo Jr., Maurício Lacerda Nogueira. Prefeitura de Sao Paulo: Melissa Palmieri. |
| EPI_ISL_1966992, EPI_ISL_1966993 | CS I DE OURINHOS | Instituto Butantan / ESALQ-USP (Piracicaba) | Instituto Butantan: Dimas Tadeu Covas, Sandra Coccuzzo Sampaio, Maria Carolina Elias, José Salvatore Leister Patané, Vincent Louis Viala, Antonio Jorge Martins, Ricardo Haddad, Claudia Renata dos Santos Barros, Elaine Cristina Marqueze, Raul Machado Neto, Debora Botequio Moretti, Jardelina de Souza Todao Bernardino, Loyze Paola Oliveira de Lima, Luiz Aurelio de Campos Crispin. Centro de Genômica Funcional da ESALQ: Luiz Lehmann Coutinho, Ricardo Augusto Brassaloti, Raquel de Lello Rocha Campos Cassano. NGS Soluções Genômicas: Pilar Drummond Sampaio Corrêa Mariani. FZEA-USP Pirassununga: Mirele Daiana Poleti, Jessika Cristina Chagas Lesbon, Elisangela Chicaroni Mattos, Heidge Fukumasu. USP-Botucatu: Rejane Maria Tommasini Grotto, Jayme A. Souza-Neto, Guilherme Targino Valente, Patricia Akemi Assato, Felipe Allan da Silva da Costa, Bianca Cechetto Carlos. Mendelics: Bibiana Santos, João Paulo Kitajima, Erika Freitas, David Schlesinger. Hemocentro Ribeirão Preto: Simone Kashima, Evandra Strazza Rodrigues, Svetoslav Nanev Slavov, Elaine Vieira dos Santos, Rafael dos Santos Bezerra, Luiz Carlos Junior de Alcantara, Marta Giovanetti, Vagner Fonseca, Flavia Aburjaile, Rodrigo Tocantins Calado. FAMERP-SJRP: Cecília Artico Banho, Lívia Sacchetto, Fábio Sossai Possebon, Leila Sabrina Ullmann, Cíntia Bittar, Guilherme Campos, Helena Lage Ferreira, Jorge A. Petroli Marchesi, Maisa C. Pereira Parra, Marília Moraes, Paula Rahal, Paulo Inacio da Costa, João Pessoa Araújo Jr., Maurício Lacerda Nogueira. Prefeitura de Sao Paulo: Melissa Palmieri. |
| EPI_ISL_1966994 | UBS DR HELIO MIGLIARI | Instituto Butantan / ESALQ-USP (Piracicaba) | Instituto Butantan: Dimas Tadeu Covas, Sandra Coccuzzo Sampaio, Maria Carolina Elias, José Salvatore Leister Patané, Vincent Louis Viala, Antonio Jorge Martins, Ricardo Haddad, Claudia Renata dos Santos Barros, Elaine Cristina Marqueze, Raul Machado Neto, Debora Botequio Moretti, Jardelina de Souza Todao Bernardino, Loyze Paola Oliveira de Lima, Luiz Aurelio de Campos Crispin. Centro de Genômica Funcional da ESALQ: Luiz Lehmann Coutinho, Ricardo Augusto Brassaloti, Raquel de Lello Rocha Campos Cassano. NGS Soluções Genômicas: Pilar Drummond Sampaio Corrêa Mariani. FZEA-USP Pirassununga: Mirele Daiana Poleti, Jessika Cristina Chagas Lesbon, Elisangela Chicaroni Mattos, Heidge Fukumasu. USP-Botucatu: Rejane Maria Tommasini Grotto, Jayme A. Souza-Neto, Guilherme Targino Valente, Patricia Akemi Assato, Felipe Allan da Silva da Costa, Bianca Cechetto Carlos. Mendelics: Bibiana Santos, João Paulo Kitajima, Erika Freitas, David Schlesinger. Hemocentro Ribeirão Preto: Simone Kashima, Evandra Strazza Rodrigues, Svetoslav Nanev Slavov, Elaine Vieira dos Santos, Rafael dos Santos Bezerra, Luiz Carlos Junior de Alcantara, Marta Giovanetti, Vagner Fonseca, Flavia Aburjaile, Rodrigo Tocantins Calado. FAMERP-SJRP: Cecília Artico Banho, Lívia Sacchetto, Fábio Sossai Possebon, Leila Sabrina |

| | | | |
|---|---|---|---|
| | | | Ullmann, Cíntia Bittar, Guilherme Campos, Helena Lage Ferreira, Jorge A. Petroli Marchesi, Maisa C. Pereira Parra, Marília Moraes, Paula Rahal, Paulo Inacio da Costa, João Pessoa Araújo Jr., Maurício Lacerda Nogueira. Prefeitura de Sao Paulo: Melissa Palmieri. |
| EPI_ISL_1966995 | UBS III DE PARIQUERA ACU PARIQUERA ACU | Instituto Butantan / ESALQ-USP (Piracicaba) | Instituto Butantan: Dimas Tadeu Covas, Sandra Coccuzzo Sampaio, Maria Carolina Elias, José Salvatore Leister Patané, Vincent Louis Viala, Antonio Jorge Martins, Ricardo Haddad, Claudia Renata dos Santos Barros, Elaine Cristina Marqueze, Raul Machado Neto, Debora Botequio Moretti, Jardelina de Souza Todao Bernardino, Loyze Paola Oliveira de Lima, Luiz Aurelio de Campos Crispin. Centro de Genômica Funcional da ESALQ: Luiz Lehmann Coutinho, Ricardo Augusto Brassaloti, Raquel de Lello Rocha Campos Cassano. NGS Soluções Genômicas: Pilar Drummond Sampaio Corrêa Mariani. FZEA-USP Pirassununga: Mirele Daiana Poleti, Jessika Cristina Chagas Lesbon, Elisangela Chicaroni Mattos, Heidge Fukumasu. USP-Botucatu: Rejane Maria Tommasini Grotto, Jayme A. Souza-Neto, Guilherme Targino Valente, Patricia Akemi Assato, Felipe Allan da Silva da Costa, Bianca Cechetto Carlos. Mendelics: Bibiana Santos, João Paulo Kitajima, Erika Freitas, David Schlesinger. Hemocentro Ribeirão Preto: Simone Kashima, Evandra Strazza Rodrigues, Svetoslav Nanev Slavov, Elaine Vieira dos Santos, Rafael dos Santos Bezerra, Luiz Carlos Junior de Alcantara, Marta Giovanetti, Vagner Fonseca, Flavia Aburjaile, Rodrigo Tocantins Calado. FAMERP-SJRP: Cecília Artico Banho, Lívia Sacchetto, Fábio Sossai Possebon, Leila Sabrina Ullmann, Cíntia Bittar, Guilherme Campos, Helena Lage Ferreira, Jorge A. Petroli Marchesi, Maisa C. Pereira Parra, Marília Moraes, Paula Rahal, Paulo Inacio da Costa, João Pessoa Araújo Jr., Maurício Lacerda Nogueira. Prefeitura de Sao Paulo: Melissa Palmieri. |
| EPI_ISL_1966996, EPI_ISL_1966997, EPI_ISL_1966998, EPI_ISL_1966999, EPI_ISL_1967000, EPI_ISL_1967001, EPI_ISL_1967002, EPI_ISL_1967003 | LABORATORIO MUNICIPAL DE PIRACICABA | Instituto Butantan / ESALQ-USP (Piracicaba) | Instituto Butantan: Dimas Tadeu Covas, Sandra Coccuzzo Sampaio, Maria Carolina Elias, José Salvatore Leister Patané, Vincent Louis Viala, Antonio Jorge Martins, Ricardo Haddad, Claudia Renata dos Santos Barros, Elaine Cristina Marqueze, Raul Machado Neto, Debora Botequio Moretti, Jardelina de Souza Todao Bernardino, Loyze Paola Oliveira de Lima, Luiz Aurelio de Campos Crispin. Centro de Genômica Funcional da ESALQ: Luiz Lehmann Coutinho, Ricardo Augusto Brassaloti, Raquel de Lello Rocha Campos Cassano. NGS Soluções Genômicas: Pilar Drummond Sampaio Corrêa Mariani. FZEA-USP Pirassununga: Mirele Daiana Poleti, Jessika Cristina Chagas Lesbon, Elisangela Chicaroni Mattos, Heidge Fukumasu. USP-Botucatu: Rejane Maria Tommasini Grotto, Jayme A. Souza-Neto, Guilherme Targino Valente, Patricia Akemi Assato, Felipe Allan da Silva da Costa, Bianca Cechetto Carlos. Mendelics: Bibiana Santos, João Paulo Kitajima, Erika Freitas, David Schlesinger. Hemocentro Ribeirão Preto: Simone Kashima, Evandra Strazza Rodrigues, Svetoslav Nanev Slavov, Elaine Vieira dos Santos, Rafael dos Santos Bezerra, Luiz Carlos Junior de Alcantara, Marta Giovanetti, Vagner Fonseca, Flavia Aburjaile, Rodrigo Tocantins Calado. FAMERP-SJRP: Cecília Artico Banho, Lívia Sacchetto, Fábio Sossai Possebon, Leila Sabrina Ullmann, Cíntia Bittar, Guilherme Campos, Helena Lage Ferreira, Jorge A. Petroli Marchesi, Maisa C. Pereira Parra, Marília Moraes, Paula Rahal, Paulo Inacio da Costa, João Pessoa Araújo Jr., Maurício Lacerda Nogueira. Prefeitura de Sao Paulo: Melissa Palmieri. |
| EPI_ISL_1967004 | CSII DR WASHINGTON LUIS M RODRIGUES DA SILVA PITANGUEIRAS | Instituto Butantan / ESALQ-USP (Piracicaba) | Instituto Butantan: Dimas Tadeu Covas, Sandra Coccuzzo Sampaio, Maria Carolina Elias, José Salvatore Leister Patané, Vincent Louis Viala, Antonio Jorge Martins, Ricardo Haddad, Claudia Renata dos Santos Barros, Elaine Cristina Marqueze, Raul Machado Neto, Debora Botequio Moretti, Jardelina de Souza Todao Bernardino, Loyze Paola Oliveira de Lima, Luiz Aurelio de Campos Crispin. Centro de Genômica Funcional da ESALQ: Luiz Lehmann Coutinho, Ricardo Augusto Brassaloti, Raquel de Lello Rocha Campos Cassano. NGS Soluções Genômicas: Pilar Drummond Sampaio Corrêa Mariani. FZEA-USP Pirassununga: Mirele Daiana Poleti, Jessika Cristina Chagas Lesbon, Elisangela Chicaroni Mattos, Heidge Fukumasu. USP-Botucatu: Rejane Maria Tommasini Grotto, Jayme A. Souza-Neto, Guilherme Targino Valente, Patricia Akemi Assato, Felipe Allan da Silva da Costa, Bianca Cechetto Carlos. Mendelics: Bibiana Santos, João Paulo Kitajima, Erika Freitas, David Schlesinger. Hemocentro Ribeirão Preto: Simone Kashima, Evandra Strazza Rodrigues, Svetoslav Nanev Slavov, Elaine Vieira dos Santos, Rafael dos Santos Bezerra, Luiz Carlos Junior de Alcantara, Marta Giovanetti, Vagner Fonseca, Flavia Aburjaile, Rodrigo Tocantins Calado. FAMERP-SJRP: Cecília Artico Banho, Lívia Sacchetto, Fábio Sossai Possebon, Leila Sabrina Ullmann, Cíntia Bittar, Guilherme Campos, Helena Lage Ferreira, Jorge A. Petroli Marchesi, Maisa C. Pereira Parra, Marília Moraes, Paula Rahal, Paulo Inacio da Costa, João Pessoa Araújo Jr., Maurício Lacerda Nogueira. Prefeitura de Sao Paulo: Melissa Palmieri. |
| EPI_ISL_1967005 | PA DE IBITIUVA DR OTAVIO BENETTI PITANGUEIRAS | Instituto Butantan / ESALQ-USP (Piracicaba) | Instituto Butantan: Dimas Tadeu Covas, Sandra Coccuzzo Sampaio, Maria Carolina Elias, José Salvatore Leister Patané, Vincent Louis Viala, Antonio Jorge Martins, Ricardo Haddad, Claudia Renata dos Santos Barros, Elaine Cristina Marqueze, Raul Machado Neto, Debora Botequio Moretti, Jardelina de Souza Todao Bernardino, Loyze Paola Oliveira de Lima, Luiz Aurelio de Campos Crispin. Centro de Genômica Funcional da ESALQ: Luiz Lehmann Coutinho, Ricardo Augusto Brassaloti, Raquel de Lello Rocha Campos Cassano. NGS Soluções Genômicas: Pilar Drummond Sampaio Corrêa Mariani. FZEA-USP Pirassununga: Mirele Daiana Poleti, Jessika Cristina Chagas Lesbon, Elisangela Chicaroni Mattos, Heidge Fukumasu. USP-Botucatu: Rejane Maria Tommasini Grotto, Jayme A. Souza-Neto, Guilherme Targino Valente, Patricia Akemi Assato, Felipe Allan da Silva da Costa, Bianca Cechetto Carlos. Mendelics: Bibiana Santos, João Paulo Kitajima, Erika Freitas, David Schlesinger. Hemocentro Ribeirão Preto: Simone Kashima, Evandra Strazza Rodrigues, Svetoslav Nanev Slavov, Elaine Vieira dos Santos, Rafael dos Santos Bezerra, Luiz Carlos Junior de Alcantara, Marta Giovanetti, Vagner Fonseca, Flavia Aburjaile, Rodrigo Tocantins Calado. FAMERP-SJRP: Cecília Artico Banho, Lívia Sacchetto, Fábio Sossai Possebon, Leila Sabrina Ullmann, Cíntia Bittar, Guilherme Campos, Helena Lage Ferreira, Jorge A. Petroli Marchesi, Maisa C. Pereira Parra, Marília Moraes, Paula Rahal, Paulo Inacio da Costa, João Pessoa Araújo Jr., Maurício Lacerda Nogueira. Prefeitura de Sao Paulo: Melissa Palmieri. |
| EPI_ISL_1967006, EPI_ISL_1967007, EPI_ISL_1967008, EPI_ISL_1967010, EPI_ISL_1967011, EPI_ISL_1967012, EPI_ISL_1967013 | CSII DR WASHINGTON LUIS M RODRIGUES DA SILVA PITANGUEIRAS | Instituto Butantan / ESALQ-USP (Piracicaba) | Instituto Butantan: Dimas Tadeu Covas, Sandra Coccuzzo Sampaio, Maria Carolina Elias, José Salvatore Leister Patané, Vincent Louis Viala, Antonio Jorge Martins, Ricardo Haddad, Claudia Renata dos Santos Barros, Elaine Cristina Marqueze, Raul Machado Neto, Debora Botequio Moretti, Jardelina de Souza Todao Bernardino, Loyze Paola Oliveira de Lima, Luiz Aurelio de Campos Crispin. Centro de Genômica Funcional da ESALQ: Luiz Lehmann Coutinho, Ricardo Augusto Brassaloti, Raquel de Lello Rocha Campos Cassano. NGS Soluções Genômicas: Pilar Drummond Sampaio Corrêa Mariani. FZEA-USP Pirassununga: Mirele Daiana Poleti, Jessika Cristina Chagas Lesbon, Elisangela Chicaroni Mattos, Heidge Fukumasu. USP-Botucatu: Rejane Maria Tommasini Grotto, Jayme A. Souza-Neto, Guilherme Targino Valente, Patricia Akemi Assato, Felipe Allan da Silva da Costa, Bianca Cechetto Carlos. Mendelics: Bibiana Santos, João Paulo Kitajima, Erika Freitas, David Schlesinger. Hemocentro Ribeirão Preto: Simone Kashima, Evandra Strazza Rodrigues, Svetoslav Nanev Slavov, Elaine Vieira dos Santos, Rafael dos Santos Bezerra, Luiz Carlos Junior de Alcantara, Marta Giovanetti, Vagner Fonseca, Flavia Aburjaile, Rodrigo Tocantins Calado. FAMERP-SJRP: Cecília Artico Banho, Lívia Sacchetto, Fábio Sossai Possebon, Leila Sabrina Ullmann, Cíntia Bittar, Guilherme Campos, Helena Lage Ferreira, Jorge A. Petroli Marchesi, Maisa C. Pereira Parra, Marília Moraes, Paula Rahal, Paulo Inacio da Costa, João Pessoa Araújo Jr., Maurício Lacerda Nogueira. Prefeitura de Sao Paulo: Melissa Palmieri. |
| EPI_ISL_1967014, EPI_ISL_1967015, EPI_ISL_1967016, EPI_ISL_1967017, EPI_ISL_1967018, EPI_ISL_1967019 | SANTA CASA DE PORTO FELIZ | Instituto Butantan / ESALQ-USP (Piracicaba) | Instituto Butantan: Dimas Tadeu Covas, Sandra Coccuzzo Sampaio, Maria Carolina Elias, José Salvatore Leister Patané, Vincent Louis Viala, Antonio Jorge Martins, Ricardo Haddad, Claudia Renata dos Santos Barros, Elaine Cristina Marqueze, Raul Machado Neto, Debora Botequio Moretti, Jardelina de Souza Todao Bernardino, Loyze Paola Oliveira de Lima, Luiz Aurelio de Campos Crispin. Centro de Genômica Funcional da ESALQ: Luiz Lehmann Coutinho, Ricardo Augusto Brassaloti, Raquel de Lello Rocha Campos Cassano. NGS Soluções Genômicas: Pilar Drummond Sampaio Corrêa Mariani. FZEA-USP Pirassununga: Mirele Daiana Poleti, Jessika Cristina Chagas Lesbon, Elisangela Chicaroni Mattos, Heidge Fukumasu. USP-Botucatu: Rejane Maria Tommasini Grotto, Jayme A. Souza-Neto, Guilherme Targino Valente, Patricia Akemi Assato, Felipe Allan da Silva da Costa, Bianca Cechetto Carlos. Mendelics: Bibiana Santos, João Paulo Kitajima, Erika Freitas, David Schlesinger. Hemocentro Ribeirão Preto: Simone Kashima, Evandra Strazza Rodrigues, Svetoslav Nanev Slavov, Elaine Vieira dos Santos, Rafael dos Santos Bezerra, Luiz Carlos Junior de Alcantara, Marta Giovanetti, Vagner Fonseca, Flavia Aburjaile, Rodrigo Tocantins Calado. FAMERP-SJRP: Cecília Artico Banho, Lívia Sacchetto, Fábio Sossai Possebon, Leila Sabrina Ullmann, Cíntia Bittar, Guilherme Campos, Helena Lage Ferreira, Jorge A. Petroli Marchesi, Maisa C. Pereira Parra, Marília Moraes, Paula Rahal, Paulo Inacio da Costa, João Pessoa Araújo Jr., Maurício Lacerda Nogueira. Prefeitura de Sao Paulo: Melissa Palmieri. |
| EPI_ISL_1967020, EPI_ISL_1967021, EPI_ISL_1967022, EPI_ISL_1967023, EPI_ISL_1967024 | PSF DR ANTONIO PIRES DE ALMEIDA PORTO FELIZ | Instituto Butantan / ESALQ-USP (Piracicaba) | Instituto Butantan: Dimas Tadeu Covas, Sandra Coccuzzo Sampaio, Maria Carolina Elias, José Salvatore Leister Patané, Vincent Louis Viala, Antonio Jorge Martins, Ricardo Haddad, Claudia Renata dos Santos Barros, Elaine Cristina Marqueze, Raul Machado Neto, Debora Botequio Moretti, Jardelina de Souza Todao Bernardino, Loyze Paola Oliveira de Lima, Luiz Aurelio de Campos Crispin. Centro de Genômica Funcional da ESALQ: Luiz Lehmann Coutinho, Ricardo Augusto Brassaloti, Raquel de Lello Rocha Campos Cassano. NGS Soluções Genômicas: Pilar Drummond Sampaio Corrêa Mariani. FZEA-USP Pirassununga: Mirele Daiana Poleti, Jessika Cristina Chagas Lesbon, Elisangela Chicaroni Mattos, Heidge Fukumasu. USP-Botucatu: Rejane Maria Tommasini Grotto, Jayme A. Souza-Neto, Guilherme Targino Valente, Patricia Akemi Assato, Felipe Allan da Silva da Costa, Bianca Cechetto Carlos. Mendelics: Bibiana Santos, João Paulo Kitajima, Erika Freitas, David Schlesinger. Hemocentro Ribeirão Preto: Simone Kashima, Evandra Strazza Rodrigues, Svetoslav Nanev Slavov, Elaine Vieira dos Santos, Rafael dos Santos Bezerra, Luiz Carlos Junior de Alcantara, Marta Giovanetti, Vagner Fonseca, Flavia Aburjaile, Rodrigo Tocantins Calado. FAMERP-SJRP: Cecília Artico Banho, Lívia Sacchetto, Fábio Sossai Possebon, Leila Sabrina Ullmann, Cíntia Bittar, Guilherme Campos, Helena Lage Ferreira, Jorge A. Petroli Marchesi, Maisa C. Pereira Parra, Marília Moraes, Paula Rahal, Paulo Inacio da Costa, João Pessoa Araújo Jr., Maurício Lacerda Nogueira. Prefeitura de Sao Paulo: Melissa Palmieri. |
| EPI_ISL_1967025, EPI_ISL_1967026 | HOSPITAL MUNICIPAL MARIA ROSA CARDOSO | Instituto Butantan / ESALQ-USP (Piracicaba) | Instituto Butantan: Dimas Tadeu Covas, Sandra Coccuzzo Sampaio, Maria Carolina Elias, José Salvatore Leister Patané, Vincent Louis Viala, Antonio Jorge Martins, Ricardo Haddad, Claudia Renata dos Santos Barros, Elaine Cristina Marqueze, Raul Machado Neto, Debora Botequio Moretti, Jardelina de Souza Todao Bernardino, Loyze Paola Oliveira de Lima, Luiz Aurelio de Campos Crispin. Centro de Genômica Funcional da ESALQ: Luiz Lehmann Coutinho, Ricardo Augusto Brassaloti, Raquel de Lello Rocha Campos Cassano. NGS Soluções Genômicas: Pilar Drummond Sampaio Corrêa Mariani. |

| | | | |
|---|---|---|---|
| | | | FZEA-USP Pirassununga: Mirele Daiana Poleti, Jessika Cristina Chagas Lesbon, Elisangela Chicaroni Mattos, Heidge Fukumasu. USP-Botucatu: Rejane Maria Tommasini Grotto, Jayme A. Souza-Neto, Guilherme Targino Valente, Patricia Akemi Assato, Felipe Allan da Silva da Costa, Bianca Cechetto Carlos. Mendelics: Bibiana Santos, João Paulo Kitajima, Erika Freitas, David Schlesinger. Hemocentro Ribeirão Preto: Simone Kashima, Evandra Strazza Rodrigues, Svetoslav Nanev Slavov, Elaine Vieira dos Santos, Rafael dos Santos Bezerra, Luiz Carlos Junior de Alcantara, Marta Giovanetti, Vagner Fonseca, Flavia Aburjaile, Rodrigo Tocantins Calado. FAMERP-SJRP: Cecília Artico Banho, Lívia Sacchetto, Fábio Sossai Possebon, Leila Sabrina Ullmann, Cíntia Bittar, Guilherme Campos, Helena Lage Ferreira, Jorge A. Petroli Marchesi, Maisa C. Pereira Parra, Marília Moraes, Paula Rahal, Paulo Inacio da Costa, João Pessoa Araújo Jr., Maurício Lacerda Nogueira. Prefeitura de Sao Paulo: Melissa Palmieri. |
| EPI_ISL_1967028 | AMBULATORIO DE MOLESTIAS INFECCIOSAS DE SALTO | Instituto Butantan / ESALQ-USP (Piracicaba) | Instituto Butantan: Dimas Tadeu Covas, Sandra Coccuzzo Sampaio, Maria Carolina Elias, José Salvatore Leister Patané, Vincent Louis Viala, Antonio Jorge Martins, Ricardo Haddad, Claudia Renata dos Santos Barros, Elaine Cristina Marqueze, Raul Machado Neto, Debora Botequio Moretti, Jardelina de Souza Todao Bernardino, Loyze Paola Oliveira de Lima, Luiz Aurelio de Campos Crispin. Centro de Genômica Funcional da ESALQ: Luiz Lehmann Coutinho, Ricardo Augusto Brassaloti, Raquel de Lello Rocha Campos Cassano. NGS Soluções Genômicas: Pilar Drummond Sampaio Corrêa Mariani. FZEA-USP Pirassununga: Mirele Daiana Poleti, Jessika Cristina Chagas Lesbon, Elisangela Chicaroni Mattos, Heidge Fukumasu. USP-Botucatu: Rejane Maria Tommasini Grotto, Jayme A. Souza-Neto, Guilherme Targino Valente, Patricia Akemi Assato, Felipe Allan da Silva da Costa, Bianca Cechetto Carlos. Mendelics: Bibiana Santos, João Paulo Kitajima, Erika Freitas, David Schlesinger. Hemocentro Ribeirão Preto: Simone Kashima, Evandra Strazza Rodrigues, Svetoslav Nanev Slavov, Elaine Vieira dos Santos, Rafael dos Santos Bezerra, Luiz Carlos Junior de Alcantara, Marta Giovanetti, Vagner Fonseca, Flavia Aburjaile, Rodrigo Tocantins Calado. FAMERP-SJRP: Cecília Artico Banho, Lívia Sacchetto, Fábio Sossai Possebon, Leila Sabrina Ullmann, Cíntia Bittar, Guilherme Campos, Helena Lage Ferreira, Jorge A. Petroli Marchesi, Maisa C. Pereira Parra, Marília Moraes, Paula Rahal, Paulo Inacio da Costa, João Pessoa Araújo Jr., Maurício Lacerda Nogueira. Prefeitura de Sao Paulo: Melissa Palmieri. |
| EPI_ISL_1967029, EPI_ISL_1967030, EPI_ISL_1967031, EPI_ISL_1967032 | HOSP E MATERNIDADE MUNICIPAL N SRA MONTE SERRAT | Instituto Butantan / ESALQ-USP (Piracicaba) | Instituto Butantan: Dimas Tadeu Covas, Sandra Coccuzzo Sampaio, Maria Carolina Elias, José Salvatore Leister Patané, Vincent Louis Viala, Antonio Jorge Martins, Ricardo Haddad, Claudia Renata dos Santos Barros, Elaine Cristina Marqueze, Raul Machado Neto, Debora Botequio Moretti, Jardelina de Souza Todao Bernardino, Loyze Paola Oliveira de Lima, Luiz Aurelio de Campos Crispin. Centro de Genômica Funcional da ESALQ: Luiz Lehmann Coutinho, Ricardo Augusto Brassaloti, Raquel de Lello Rocha Campos Cassano. NGS Soluções Genômicas: Pilar Drummond Sampaio Corrêa Mariani. FZEA-USP Pirassununga: Mirele Daiana Poleti, Jessika Cristina Chagas Lesbon, Elisangela Chicaroni Mattos, Heidge Fukumasu. USP-Botucatu: Rejane Maria Tommasini Grotto, Jayme A. Souza-Neto, Guilherme Targino Valente, Patricia Akemi Assato, Felipe Allan da Silva da Costa, Bianca Cechetto Carlos. Mendelics: Bibiana Santos, João Paulo Kitajima, Erika Freitas, David Schlesinger. Hemocentro Ribeirão Preto: Simone Kashima, Evandra Strazza Rodrigues, Svetoslav Nanev Slavov, Elaine Vieira dos Santos, Rafael dos Santos Bezerra, Luiz Carlos Junior de Alcantara, Marta Giovanetti, Vagner Fonseca, Flavia Aburjaile, Rodrigo Tocantins Calado. FAMERP-SJRP: Cecília Artico Banho, Lívia Sacchetto, Fábio Sossai Possebon, Leila Sabrina Ullmann, Cíntia Bittar, Guilherme Campos, Helena Lage Ferreira, Jorge A. Petroli Marchesi, Maisa C. Pereira Parra, Marília Moraes, Paula Rahal, Paulo Inacio da Costa, João Pessoa Araújo Jr., Maurício Lacerda Nogueira. Prefeitura de Sao Paulo: Melissa Palmieri. |
| EPI_ISL_1967033, EPI_ISL_1967034, EPI_ISL_1967035, EPI_ISL_1967036, EPI_ISL_1967037, EPI_ISL_1967038, EPI_ISL_1967039, EPI_ISL_1967040, EPI_ISL_1967041 | PSF MARIA JOSE SALTO DE PIRAPORA | Instituto Butantan / ESALQ-USP (Piracicaba) | Instituto Butantan: Dimas Tadeu Covas, Sandra Coccuzzo Sampaio, Maria Carolina Elias, José Salvatore Leister Patané, Vincent Louis Viala, Antonio Jorge Martins, Ricardo Haddad, Claudia Renata dos Santos Barros, Elaine Cristina Marqueze, Raul Machado Neto, Debora Botequio Moretti, Jardelina de Souza Todao Bernardino, Loyze Paola Oliveira de Lima, Luiz Aurelio de Campos Crispin. Centro de Genômica Funcional da ESALQ: Luiz Lehmann Coutinho, Ricardo Augusto Brassaloti, Raquel de Lello Rocha Campos Cassano. NGS Soluções Genômicas: Pilar Drummond Sampaio Corrêa Mariani. FZEA-USP Pirassununga: Mirele Daiana Poleti, Jessika Cristina Chagas Lesbon, Elisangela Chicaroni Mattos, Heidge Fukumasu. USP-Botucatu: Rejane Maria Tommasini Grotto, Jayme A. Souza-Neto, Guilherme Targino Valente, Patricia Akemi Assato, Felipe Allan da Silva da Costa, Bianca Cechetto Carlos. Mendelics: Bibiana Santos, João Paulo Kitajima, Erika Freitas, David Schlesinger. Hemocentro Ribeirão Preto: Simone Kashima, Evandra Strazza Rodrigues, Svetoslav Nanev Slavov, Elaine Vieira dos Santos, Rafael dos Santos Bezerra, Luiz Carlos Junior de Alcantara, Marta Giovanetti, Vagner Fonseca, Flavia Aburjaile, Rodrigo Tocantins Calado. FAMERP-SJRP: Cecília Artico Banho, Lívia Sacchetto, Fábio Sossai Possebon, Leila Sabrina Ullmann, Cíntia Bittar, Guilherme Campos, Helena Lage Ferreira, Jorge A. Petroli Marchesi, Maisa C. Pereira Parra, Marília Moraes, Paula Rahal, Paulo Inacio da Costa, João Pessoa Araújo Jr., Maurício Lacerda Nogueira. Prefeitura de Sao Paulo: Melissa Palmieri. |
| EPI_ISL_1967042, EPI_ISL_1967043, EPI_ISL_1967044, EPI_ISL_1967045, EPI_ISL_1967046, EPI_ISL_1967047, EPI_ISL_1967048, EPI_ISL_1967049 | UNIDADE DE PRONTO ATENDIMENTO UPA I SANTA ISABEL | Instituto Butantan / ESALQ-USP (Piracicaba) | Instituto Butantan: Dimas Tadeu Covas, Sandra Coccuzzo Sampaio, Maria Carolina Elias, José Salvatore Leister Patané, Vincent Louis Viala, Antonio Jorge Martins, Ricardo Haddad, Claudia Renata dos Santos Barros, Elaine Cristina Marqueze, Raul Machado Neto, Debora Botequio Moretti, Jardelina de Souza Todao Bernardino, Loyze Paola Oliveira de Lima, Luiz Aurelio de Campos Crispin. Centro de Genômica Funcional da ESALQ: Luiz Lehmann Coutinho, Ricardo Augusto Brassaloti, Raquel de Lello Rocha Campos Cassano. NGS Soluções Genômicas: Pilar Drummond Sampaio Corrêa Mariani. FZEA-USP Pirassununga: Mirele Daiana Poleti, Jessika Cristina Chagas Lesbon, Elisangela Chicaroni Mattos, Heidge Fukumasu. USP-Botucatu: Rejane Maria Tommasini Grotto, Jayme A. Souza-Neto, Guilherme Targino Valente, Patricia Akemi Assato, Felipe Allan da Silva da Costa, Bianca Cechetto Carlos. Mendelics: Bibiana Santos, João Paulo Kitajima, Erika Freitas, David Schlesinger. Hemocentro Ribeirão Preto: Simone Kashima, Evandra Strazza Rodrigues, Svetoslav Nanev Slavov, Elaine Vieira dos Santos, Rafael dos Santos Bezerra, Luiz Carlos Junior de Alcantara, Marta Giovanetti, Vagner Fonseca, Flavia Aburjaile, Rodrigo Tocantins Calado. FAMERP-SJRP: Cecília Artico Banho, Lívia Sacchetto, Fábio Sossai Possebon, Leila Sabrina Ullmann, Cíntia Bittar, Guilherme Campos, Helena Lage Ferreira, Jorge A. Petroli Marchesi, Maisa C. Pereira Parra, Marília Moraes, Paula Rahal, Paulo Inacio da Costa, João Pessoa Araújo Jr., Maurício Lacerda Nogueira. Prefeitura de Sao Paulo: Melissa Palmieri. |
| EPI_ISL_1967050 | CS DE SANTA RITA DOESTE | Instituto Butantan / ESALQ-USP (Piracicaba) | Instituto Butantan: Dimas Tadeu Covas, Sandra Coccuzzo Sampaio, Maria Carolina Elias, José Salvatore Leister Patané, Vincent Louis Viala, Antonio Jorge Martins, Ricardo Haddad, Claudia Renata dos Santos Barros, Elaine Cristina Marqueze, Raul Machado Neto, Debora Botequio Moretti, Jardelina de Souza Todao Bernardino, Loyze Paola Oliveira de Lima, Luiz Aurelio de Campos Crispin. Centro de Genômica Funcional da ESALQ: Luiz Lehmann Coutinho, Ricardo Augusto Brassaloti, Raquel de Lello Rocha Campos Cassano. NGS Soluções Genômicas: Pilar Drummond Sampaio Corrêa Mariani. FZEA-USP Pirassununga: Mirele Daiana Poleti, Jessika Cristina Chagas Lesbon, Elisangela Chicaroni Mattos, Heidge Fukumasu. USP-Botucatu: Rejane Maria Tommasini Grotto, Jayme A. Souza-Neto, Guilherme Targino Valente, Patricia Akemi Assato, Felipe Allan da Silva da Costa, Bianca Cechetto Carlos. Mendelics: Bibiana Santos, João Paulo Kitajima, Erika Freitas, David Schlesinger. Hemocentro Ribeirão Preto: Simone Kashima, Evandra Strazza Rodrigues, Svetoslav Nanev Slavov, Elaine Vieira dos Santos, Rafael dos Santos Bezerra, Luiz Carlos Junior de Alcantara, Marta Giovanetti, Vagner Fonseca, Flavia Aburjaile, Rodrigo Tocantins Calado. FAMERP-SJRP: Cecília Artico Banho, Lívia Sacchetto, Fábio Sossai Possebon, Leila Sabrina Ullmann, Cíntia Bittar, Guilherme Campos, Helena Lage Ferreira, Jorge A. Petroli Marchesi, Maisa C. Pereira Parra, Marília Moraes, Paula Rahal, Paulo Inacio da Costa, João Pessoa Araújo Jr., Maurício Lacerda Nogueira. Prefeitura de Sao Paulo: Melissa Palmieri. |
| EPI_ISL_1967052, EPI_ISL_1967053, EPI_ISL_1967054, EPI_ISL_1967056, EPI_ISL_1967057 | UNIDADE DE VIGILANCIA EM SAUDE | Instituto Butantan / ESALQ-USP (Piracicaba) | Instituto Butantan: Dimas Tadeu Covas, Sandra Coccuzzo Sampaio, Maria Carolina Elias, José Salvatore Leister Patané, Vincent Louis Viala, Antonio Jorge Martins, Ricardo Haddad, Claudia Renata dos Santos Barros, Elaine Cristina Marqueze, Raul Machado Neto, Debora Botequio Moretti, Jardelina de Souza Todao Bernardino, Loyze Paola Oliveira de Lima, Luiz Aurelio de Campos Crispin. Centro de Genômica Funcional da ESALQ: Luiz Lehmann Coutinho, Ricardo Augusto Brassaloti, Raquel de Lello Rocha Campos Cassano. NGS Soluções Genômicas: Pilar Drummond Sampaio Corrêa Mariani. FZEA-USP Pirassununga: Mirele Daiana Poleti, Jessika Cristina Chagas Lesbon, Elisangela Chicaroni Mattos, Heidge Fukumasu. USP-Botucatu: Rejane Maria Tommasini Grotto, Jayme A. Souza-Neto, Guilherme Targino Valente, Patricia Akemi Assato, Felipe Allan da Silva da Costa, Bianca Cechetto Carlos. Mendelics: Bibiana Santos, João Paulo Kitajima, Erika Freitas, David Schlesinger. Hemocentro Ribeirão Preto: Simone Kashima, Evandra Strazza Rodrigues, Svetoslav Nanev Slavov, Elaine Vieira dos Santos, Rafael dos Santos Bezerra, Luiz Carlos Junior de Alcantara, Marta Giovanetti, Vagner Fonseca, Flavia Aburjaile, Rodrigo Tocantins Calado. FAMERP-SJRP: Cecília Artico Banho, Lívia Sacchetto, Fábio Sossai Possebon, Leila Sabrina Ullmann, Cíntia Bittar, Guilherme Campos, Helena Lage Ferreira, Jorge A. Petroli Marchesi, Maisa C. Pereira Parra, Marília Moraes, Paula Rahal, Paulo Inacio da Costa, João Pessoa Araújo Jr., Maurício Lacerda Nogueira. Prefeitura de Sao Paulo: Melissa Palmieri. |
| EPI_ISL_1967058, EPI_ISL_1967059 | CENTRO DE SAUDE ESF IV ZONA RURAL DOMINGOS DE S SJRIOPARDO | Instituto Butantan / ESALQ-USP (Piracicaba) | Instituto Butantan: Dimas Tadeu Covas, Sandra Coccuzzo Sampaio, Maria Carolina Elias, José Salvatore Leister Patané, Vincent Louis Viala, Antonio Jorge Martins, Ricardo Haddad, Claudia Renata dos Santos Barros, Elaine Cristina Marqueze, Raul Machado Neto, Debora Botequio Moretti, Jardelina de Souza Todao Bernardino, Loyze Paola Oliveira de Lima, Luiz Aurelio de Campos Crispin. Centro de Genômica Funcional da ESALQ: Luiz Lehmann Coutinho, Ricardo Augusto Brassaloti, Raquel de Lello Rocha Campos Cassano. NGS Soluções Genômicas: Pilar Drummond Sampaio Corrêa Mariani. FZEA-USP Pirassununga: Mirele Daiana Poleti, Jessika Cristina Chagas Lesbon, Elisangela Chicaroni Mattos, Heidge Fukumasu. USP-Botucatu: Rejane Maria Tommasini Grotto, Jayme A. Souza-Neto, Guilherme Targino Valente, Patricia Akemi Assato, Felipe Allan da Silva da Costa, Bianca Cechetto Carlos. Mendelics: Bibiana Santos, João Paulo Kitajima, Erika Freitas, David Schlesinger. Hemocentro Ribeirão Preto: Simone Kashima, Evandra Strazza Rodrigues, Svetoslav Nanev Slavov, Elaine Vieira dos Santos, Rafael dos Santos Bezerra, Luiz Carlos Junior de Alcantara, Marta Giovanetti, Vagner Fonseca, Flavia Aburjaile, Rodrigo Tocantins Calado. FAMERP-SJRP: Cecília Artico Banho, Lívia Sacchetto, Fábio Sossai Possebon, Leila Sabrina Ullmann, Cíntia Bittar, Guilherme Campos, Helena Lage Ferreira, Jorge A. Petroli Marchesi, Maisa C. Pereira Parra, Marília Moraes, Paula Rahal, Paulo Inacio da Costa, João Pessoa Araújo Jr., Maurício Lacerda Nogueira. Prefeitura de Sao Paulo: Melissa Palmieri. |

| | | | |
|---|---|---|---|
| EPI_ISL_1967062, EPI_ISL_1967063, EPI_ISL_1967064 | CENTRO DE SAUDE II SAO MIGUEL ARCANJO | Instituto Butantan / ESALQ-USP (Piracicaba) | Instituto Butantan: Dimas Tadeu Covas, Sandra Coccuzzo Sampaio, Maria Carolina Elias, José Salvatore Leister Patané, Vincent Louis Viala, Antonio Jorge Martins, Ricardo Haddad, Claudia Renata dos Santos Barros, Elaine Cristina Marqueze, Raul Machado Neto, Debora Botequio Moretti, Jardelina de Souza Todao Bernardino, Loyze Paola Oliveira de Lima, Luiz Aurelio de Campos Crispin. Centro de Genômica Funcional da ESALQ: Luiz Lehmann Coutinho, Ricardo Augusto Brassaloti, Raquel de Lello Rocha Campos Cassano. NGS Soluções Genômicas: Pilar Drummond Sampaio Corrêa Mariani. FZEA-USP Pirassununga: Mirele Daiana Poleti, Jessika Cristina Chagas Lesbon, Elisangela Chicaroni Mattos, Heidge Fukumasu. USP-Botucatu: Rejane Maria Tommasini Grotto, Jayme A. Souza-Neto, Guilherme Targino Valente, Patricia Akemi Assato, Felipe Allan da Silva da Costa, Bianca Cechetto Carlos. Mendelics: Bibiana Santos, João Paulo Kitajima, Erika Freitas, David Schlesinger. Hemocentro Ribeirão Preto: Simone Kashima, Evandra Strazza Rodrigues, Svetoslav Nanev Slavov, Elaine Vieira dos Santos, Rafael dos Santos Bezerra, Luiz Carlos Junior de Alcantara, Marta Giovanetti, Vagner Fonseca, Flavia Aburjaile, Rodrigo Tocantins Calado. FAMERP-SJRP: Cecília Artico Banho, Lívia Sacchetto, Fábio Sossai Possebon, Leila Sabrina Ullmann, Cíntia Bittar, Guilherme Campos, Helena Lage Ferreira, Jorge A. Petroli Marchesi, Maisa C. Pereira Parra, Marília Moraes, Paula Rahal, Paulo Inacio da Costa, João Pessoa Araújo Jr., Maurício Lacerda Nogueira. Prefeitura de Sao Paulo: Melissa Palmieri. |
| EPI_ISL_1967065, EPI_ISL_1967066 | HOSPITAL SAO MIGUEL ARCANJO | Instituto Butantan / ESALQ-USP (Piracicaba) | Instituto Butantan: Dimas Tadeu Covas, Sandra Coccuzzo Sampaio, Maria Carolina Elias, José Salvatore Leister Patané, Vincent Louis Viala, Antonio Jorge Martins, Ricardo Haddad, Claudia Renata dos Santos Barros, Elaine Cristina Marqueze, Raul Machado Neto, Debora Botequio Moretti, Jardelina de Souza Todao Bernardino, Loyze Paola Oliveira de Lima, Luiz Aurelio de Campos Crispin. Centro de Genômica Funcional da ESALQ: Luiz Lehmann Coutinho, Ricardo Augusto Brassaloti, Raquel de Lello Rocha Campos Cassano. NGS Soluções Genômicas: Pilar Drummond Sampaio Corrêa Mariani. FZEA-USP Pirassununga: Mirele Daiana Poleti, Jessika Cristina Chagas Lesbon, Elisangela Chicaroni Mattos, Heidge Fukumasu. USP-Botucatu: Rejane Maria Tommasini Grotto, Jayme A. Souza-Neto, Guilherme Targino Valente, Patricia Akemi Assato, Felipe Allan da Silva da Costa, Bianca Cechetto Carlos. Mendelics: Bibiana Santos, João Paulo Kitajima, Erika Freitas, David Schlesinger. Hemocentro Ribeirão Preto: Simone Kashima, Evandra Strazza Rodrigues, Svetoslav Nanev Slavov, Elaine Vieira dos Santos, Rafael dos Santos Bezerra, Luiz Carlos Junior de Alcantara, Marta Giovanetti, Vagner Fonseca, Flavia Aburjaile, Rodrigo Tocantins Calado. FAMERP-SJRP: Cecília Artico Banho, Lívia Sacchetto, Fábio Sossai Possebon, Leila Sabrina Ullmann, Cíntia Bittar, Guilherme Campos, Helena Lage Ferreira, Jorge A. Petroli Marchesi, Maisa C. Pereira Parra, Marília Moraes, Paula Rahal, Paulo Inacio da Costa, João Pessoa Araújo Jr., Maurício Lacerda Nogueira. Prefeitura de Sao Paulo: Melissa Palmieri. |
| EPI_ISL_1967067 | HOSP PRONTO SOCORRO ITAQUERA | Instituto Butantan / ESALQ-USP (Piracicaba) | Instituto Butantan: Dimas Tadeu Covas, Sandra Coccuzzo Sampaio, Maria Carolina Elias, José Salvatore Leister Patané, Vincent Louis Viala, Antonio Jorge Martins, Ricardo Haddad, Claudia Renata dos Santos Barros, Elaine Cristina Marqueze, Raul Machado Neto, Debora Botequio Moretti, Jardelina de Souza Todao Bernardino, Loyze Paola Oliveira de Lima, Luiz Aurelio de Campos Crispin. Centro de Genômica Funcional da ESALQ: Luiz Lehmann Coutinho, Ricardo Augusto Brassaloti, Raquel de Lello Rocha Campos Cassano. NGS Soluções Genômicas: Pilar Drummond Sampaio Corrêa Mariani. FZEA-USP Pirassununga: Mirele Daiana Poleti, Jessika Cristina Chagas Lesbon, Elisangela Chicaroni Mattos, Heidge Fukumasu. USP-Botucatu: Rejane Maria Tommasini Grotto, Jayme A. Souza-Neto, Guilherme Targino Valente, Patricia Akemi Assato, Felipe Allan da Silva da Costa, Bianca Cechetto Carlos. Mendelics: Bibiana Santos, João Paulo Kitajima, Erika Freitas, David Schlesinger. Hemocentro Ribeirão Preto: Simone Kashima, Evandra Strazza Rodrigues, Svetoslav Nanev Slavov, Elaine Vieira dos Santos, Rafael dos Santos Bezerra, Luiz Carlos Junior de Alcantara, Marta Giovanetti, Vagner Fonseca, Flavia Aburjaile, Rodrigo Tocantins Calado. FAMERP-SJRP: Cecília Artico Banho, Lívia Sacchetto, Fábio Sossai Possebon, Leila Sabrina Ullmann, Cíntia Bittar, Guilherme Campos, Helena Lage Ferreira, Jorge A. Petroli Marchesi, Maisa C. Pereira Parra, Marília Moraes, Paula Rahal, Paulo Inacio da Costa, João Pessoa Araújo Jr., Maurício Lacerda Nogueira. Prefeitura de Sao Paulo: Melissa Palmieri. |
| EPI_ISL_1967069, EPI_ISL_1967070, EPI_ISL_1967071, EPI_ISL_1967072 | HOSP MUN CIDADE TIRADENTES CARMEN PRUDENTE | Instituto Butantan / ESALQ-USP (Piracicaba) | Instituto Butantan: Dimas Tadeu Covas, Sandra Coccuzzo Sampaio, Maria Carolina Elias, José Salvatore Leister Patané, Vincent Louis Viala, Antonio Jorge Martins, Ricardo Haddad, Claudia Renata dos Santos Barros, Elaine Cristina Marqueze, Raul Machado Neto, Debora Botequio Moretti, Jardelina de Souza Todao Bernardino, Loyze Paola Oliveira de Lima, Luiz Aurelio de Campos Crispin. Centro de Genômica Funcional da ESALQ: Luiz Lehmann Coutinho, Ricardo Augusto Brassaloti, Raquel de Lello Rocha Campos Cassano. NGS Soluções Genômicas: Pilar Drummond Sampaio Corrêa Mariani. FZEA-USP Pirassununga: Mirele Daiana Poleti, Jessika Cristina Chagas Lesbon, Elisangela Chicaroni Mattos, Heidge Fukumasu. USP-Botucatu: Rejane Maria Tommasini Grotto, Jayme A. Souza-Neto, Guilherme Targino Valente, Patricia Akemi Assato, Felipe Allan da Silva da Costa, Bianca Cechetto Carlos. Mendelics: Bibiana Santos, João Paulo Kitajima, Erika Freitas, David Schlesinger. Hemocentro Ribeirão Preto: Simone Kashima, Evandra Strazza Rodrigues, Svetoslav Nanev Slavov, Elaine Vieira dos Santos, Rafael dos Santos Bezerra, Luiz Carlos Junior de Alcantara, Marta Giovanetti, Vagner Fonseca, Flavia Aburjaile, Rodrigo Tocantins Calado. FAMERP-SJRP: Cecília Artico Banho, Lívia Sacchetto, Fábio Sossai Possebon, Leila Sabrina Ullmann, Cíntia Bittar, Guilherme Campos, Helena Lage Ferreira, Jorge A. Petroli Marchesi, Maisa C. Pereira Parra, Marília Moraes, Paula Rahal, Paulo Inacio da Costa, João Pessoa Araújo Jr., Maurício Lacerda Nogueira. Prefeitura de Sao Paulo: Melissa Palmieri. |
| EPI_ISL_1967075 | HOSP MUN JOSANIAS CASTANHA BRAGA | Instituto Butantan / ESALQ-USP (Piracicaba) | Instituto Butantan: Dimas Tadeu Covas, Sandra Coccuzzo Sampaio, Maria Carolina Elias, José Salvatore Leister Patané, Vincent Louis Viala, Antonio Jorge Martins, Ricardo Haddad, Claudia Renata dos Santos Barros, Elaine Cristina Marqueze, Raul Machado Neto, Debora Botequio Moretti, Jardelina de Souza Todao Bernardino, Loyze Paola Oliveira de Lima, Luiz Aurelio de Campos Crispin. Centro de Genômica Funcional da ESALQ: Luiz Lehmann Coutinho, Ricardo Augusto Brassaloti, Raquel de Lello Rocha Campos Cassano. NGS Soluções Genômicas: Pilar Drummond Sampaio Corrêa Mariani. FZEA-USP Pirassununga: Mirele Daiana Poleti, Jessika Cristina Chagas Lesbon, Elisangela Chicaroni Mattos, Heidge Fukumasu. USP-Botucatu: Rejane Maria Tommasini Grotto, Jayme A. Souza-Neto, Guilherme Targino Valente, Patricia Akemi Assato, Felipe Allan da Silva da Costa, Bianca Cechetto Carlos. Mendelics: Bibiana Santos, João Paulo Kitajima, Erika Freitas, David Schlesinger. Hemocentro Ribeirão Preto: Simone Kashima, Evandra Strazza Rodrigues, Svetoslav Nanev Slavov, Elaine Vieira dos Santos, Rafael dos Santos Bezerra, Luiz Carlos Junior de Alcantara, Marta Giovanetti, Vagner Fonseca, Flavia Aburjaile, Rodrigo Tocantins Calado. FAMERP-SJRP: Cecília Artico Banho, Lívia Sacchetto, Fábio Sossai Possebon, Leila Sabrina Ullmann, Cíntia Bittar, Guilherme Campos, Helena Lage Ferreira, Jorge A. Petroli Marchesi, Maisa C. Pereira Parra, Marília Moraes, Paula Rahal, Paulo Inacio da Costa, João Pessoa Araújo Jr., Maurício Lacerda Nogueira. Prefeitura de Sao Paulo: Melissa Palmieri. |
| EPI_ISL_1967076 | UPA VILA SANTA CATARINA | Instituto Butantan / ESALQ-USP (Piracicaba) | Instituto Butantan: Dimas Tadeu Covas, Sandra Coccuzzo Sampaio, Maria Carolina Elias, José Salvatore Leister Patané, Vincent Louis Viala, Antonio Jorge Martins, Ricardo Haddad, Claudia Renata dos Santos Barros, Elaine Cristina Marqueze, Raul Machado Neto, Debora Botequio Moretti, Jardelina de Souza Todao Bernardino, Loyze Paola Oliveira de Lima, Luiz Aurelio de Campos Crispin. Centro de Genômica Funcional da ESALQ: Luiz Lehmann Coutinho, Ricardo Augusto Brassaloti, Raquel de Lello Rocha Campos Cassano. NGS Soluções Genômicas: Pilar Drummond Sampaio Corrêa Mariani. FZEA-USP Pirassununga: Mirele Daiana Poleti, Jessika Cristina Chagas Lesbon, Elisangela Chicaroni Mattos, Heidge Fukumasu. USP-Botucatu: Rejane Maria Tommasini Grotto, Jayme A. Souza-Neto, Guilherme Targino Valente, Patricia Akemi Assato, Felipe Allan da Silva da Costa, Bianca Cechetto Carlos. Mendelics: Bibiana Santos, João Paulo Kitajima, Erika Freitas, David Schlesinger. Hemocentro Ribeirão Preto: Simone Kashima, Evandra Strazza Rodrigues, Svetoslav Nanev Slavov, Elaine Vieira dos Santos, Rafael dos Santos Bezerra, Luiz Carlos Junior de Alcantara, Marta Giovanetti, Vagner Fonseca, Flavia Aburjaile, Rodrigo Tocantins Calado. FAMERP-SJRP: Cecília Artico Banho, Lívia Sacchetto, Fábio Sossai Possebon, Leila Sabrina Ullmann, Cíntia Bittar, Guilherme Campos, Helena Lage Ferreira, Jorge A. Petroli Marchesi, Maisa C. Pereira Parra, Marília Moraes, Paula Rahal, Paulo Inacio da Costa, João Pessoa Araújo Jr., Maurício Lacerda Nogueira. Prefeitura de Sao Paulo: Melissa Palmieri. |
| EPI_ISL_1967077, EPI_ISL_1967078, EPI_ISL_1967079, EPI_ISL_1967080, EPI_ISL_1967081, EPI_ISL_1967082, EPI_ISL_1967083, EPI_ISL_1967084, EPI_ISL_1967085, EPI_ISL_1967086, EPI_ISL_1967087, EPI_ISL_1967088, EPI_ISL_1967089, EPI_ISL_1967090 | | | |
| see above | HOSPITAL GERAL JESUS TEIXEIRA DA COSTA GUAIANASES SAO PAULO | Instituto Butantan / ESALQ-USP (Piracicaba) | Instituto Butantan: Dimas Tadeu Covas, Sandra Coccuzzo Sampaio, Maria Carolina Elias, José Salvatore Leister Patané, Vincent Louis Viala, Antonio Jorge Martins, Ricardo Haddad, Claudia Renata dos Santos Barros, Elaine Cristina Marqueze, Raul Machado Neto, Debora Botequio Moretti, Jardelina de Souza Todao Bernardino, Loyze Paola Oliveira de Lima, Luiz Aurelio de Campos Crispin. Centro de Genômica Funcional da ESALQ: Luiz Lehmann Coutinho, Ricardo Augusto Brassaloti, Raquel de Lello Rocha Campos Cassano. NGS Soluções Genômicas: Pilar Drummond Sampaio Corrêa Mariani. FZEA-USP Pirassununga: Mirele Daiana Poleti, Jessika Cristina Chagas Lesbon, Elisangela Chicaroni Mattos, Heidge Fukumasu. USP-Botucatu: Rejane Maria Tommasini Grotto, Jayme A. Souza-Neto, Guilherme Targino Valente, Patricia Akemi Assato, Felipe Allan da Silva da Costa, Bianca Cechetto Carlos. Mendelics: Bibiana Santos, João Paulo Kitajima, Erika Freitas, David Schlesinger. Hemocentro Ribeirão Preto: Simone Kashima, Evandra Strazza Rodrigues, Svetoslav Nanev Slavov, Elaine Vieira dos Santos, Rafael dos Santos Bezerra, Luiz Carlos Junior de Alcantara, Marta Giovanetti, Vagner Fonseca, Flavia Aburjaile, Rodrigo Tocantins Calado. FAMERP-SJRP: Cecília Artico Banho, Lívia Sacchetto, Fábio Sossai Possebon, Leila Sabrina Ullmann, Cíntia Bittar, Guilherme Campos, Helena Lage Ferreira, Jorge A. Petroli Marchesi, Maisa C. Pereira Parra, Marília Moraes, Paula Rahal, Paulo Inacio da Costa, João Pessoa Araújo Jr., Maurício Lacerda Nogueira. Prefeitura de Sao Paulo: Melissa Palmieri. |
| EPI_ISL_1967091 | HOSP MUN CIDADE TIRADENTES CARMEN PRUDENTE | Instituto Butantan / ESALQ-USP (Piracicaba) | Instituto Butantan: Dimas Tadeu Covas, Sandra Coccuzzo Sampaio, Maria Carolina Elias, José Salvatore Leister Patané, Vincent Louis Viala, Antonio Jorge Martins, Ricardo Haddad, Claudia Renata dos Santos Barros, Elaine Cristina Marqueze, Raul Machado Neto, Debora Botequio Moretti, Jardelina de Souza Todao Bernardino, Loyze Paola Oliveira de Lima, Luiz Aurelio de Campos Crispin. Centro de Genômica Funcional da ESALQ: Luiz Lehmann Coutinho, Ricardo Augusto Brassaloti, Raquel de Lello Rocha Campos Cassano. NGS Soluções Genômicas: Pilar Drummond Sampaio Corrêa Mariani. FZEA-USP Pirassununga: Mirele Daiana Poleti, Jessika Cristina Chagas Lesbon, Elisangela Chicaroni Mattos, Heidge Fukumasu. USP-Botucatu: Rejane |

| | | | |
|---|---|---|---|
| | | | Maria Tommasini Grotto, Jayme A. Souza-Neto, Guilherme Targino Valente, Patricia Akemi Assato, Felipe Allan da Silva da Costa, Bianca Cechetto Carlos. Mendelics: Bibiana Santos, João Paulo Kitajima, Erika Freitas, David Schlesinger. Hemocentro Ribeirão Preto: Simone Kashima, Evandra Strazza Rodrigues, Svetoslav Nanev Slavov, Elaine Vieira dos Santos, Rafael dos Santos Bezerra, Luiz Carlos Junior de Alcantara, Marta Giovanetti, Vagner Fonseca, Flavia Aburjaile, Rodrigo Tocantins Calado. FAMERP-SJRP: Cecília Artico Banho, Lívia Sacchetto, Fábio Sossai Possebon, Leila Sabrina Ullmann, Cíntia Bittar, Guilherme Campos, Helena Lage Ferreira, Jorge A. Petroli Marchesi, Maisa C. Pereira Parra, Marília Moraes, Paula Rahal, Paulo Inacio da Costa, João Pessoa Araújo Jr., Maurício Lacerda Nogueira. Prefeitura de Sao Paulo: Melissa Palmieri. |
| EPI_ISL_1967092, EPI_ISL_1967093, EPI_ISL_1967094, EPI_ISL_1967095 | HOSP DO SERV PUB ESTFCO MORATO DE OLIVEIRA SAO PAULO | Instituto Butantan / ESALQ-USP (Piracicaba) | Instituto Butantan: Dimas Tadeu Covas, Sandra Coccuzzo Sampaio, Maria Carolina Elias, José Salvatore Leister Patané, Vincent Louis Viala, Antonio Jorge Martins, Ricardo Haddad, Claudia Renata dos Santos Barros, Elaine Cristina Marqueze, Raul Machado Neto, Debora Botequio Moretti, Jardelina de Souza Todao Bernardino, Loyze Paola Oliveira de Lima, Luiz Aurelio de Campos Crispin. Centro de Genômica Funcional da ESALQ: Luiz Lehmann Coutinho, Ricardo Augusto Brassaloti, Raquel de Lello Rocha Campos Cassano. NGS Soluções Genômicas: Pilar Drummond Sampaio Corrêa Mariani. FZEA-USP Pirassununga: Mirele Daiana Poleti, Jessika Cristina Chagas Lesbon, Elisangela Chicaroni Mattos, Heidge Fukumasu. USP-Botucatu: Rejane Maria Tommasini Grotto, Jayme A. Souza-Neto, Guilherme Targino Valente, Patricia Akemi Assato, Felipe Allan da Silva da Costa, Bianca Cechetto Carlos. Mendelics: Bibiana Santos, João Paulo Kitajima, Erika Freitas, David Schlesinger. Hemocentro Ribeirão Preto: Simone Kashima, Evandra Strazza Rodrigues, Svetoslav Nanev Slavov, Elaine Vieira dos Santos, Rafael dos Santos Bezerra, Luiz Carlos Junior de Alcantara, Marta Giovanetti, Vagner Fonseca, Flavia Aburjaile, Rodrigo Tocantins Calado. FAMERP-SJRP: Cecília Artico Banho, Lívia Sacchetto, Fábio Sossai Possebon, Leila Sabrina Ullmann, Cíntia Bittar, Guilherme Campos, Helena Lage Ferreira, Jorge A. Petroli Marchesi, Maisa C. Pereira Parra, Marília Moraes, Paula Rahal, Paulo Inacio da Costa, João Pessoa Araújo Jr., Maurício Lacerda Nogueira. Prefeitura de Sao Paulo: Melissa Palmieri. |
| EPI_ISL_1967096, EPI_ISL_1967097 | CENTRO DE SAUDE SAO ROQUE DR JOSE CARVALHO BRITO | Instituto Butantan / ESALQ-USP (Piracicaba) | Instituto Butantan: Dimas Tadeu Covas, Sandra Coccuzzo Sampaio, Maria Carolina Elias, José Salvatore Leister Patané, Vincent Louis Viala, Antonio Jorge Martins, Ricardo Haddad, Claudia Renata dos Santos Barros, Elaine Cristina Marqueze, Raul Machado Neto, Debora Botequio Moretti, Jardelina de Souza Todao Bernardino, Loyze Paola Oliveira de Lima, Luiz Aurelio de Campos Crispin. Centro de Genômica Funcional da ESALQ: Luiz Lehmann Coutinho, Ricardo Augusto Brassaloti, Raquel de Lello Rocha Campos Cassano. NGS Soluções Genômicas: Pilar Drummond Sampaio Corrêa Mariani. FZEA-USP Pirassununga: Mirele Daiana Poleti, Jessika Cristina Chagas Lesbon, Elisangela Chicaroni Mattos, Heidge Fukumasu. USP-Botucatu: Rejane Maria Tommasini Grotto, Jayme A. Souza-Neto, Guilherme Targino Valente, Patricia Akemi Assato, Felipe Allan da Silva da Costa, Bianca Cechetto Carlos. Mendelics: Bibiana Santos, João Paulo Kitajima, Erika Freitas, David Schlesinger. Hemocentro Ribeirão Preto: Simone Kashima, Evandra Strazza Rodrigues, Svetoslav Nanev Slavov, Elaine Vieira dos Santos, Rafael dos Santos Bezerra, Luiz Carlos Junior de Alcantara, Marta Giovanetti, Vagner Fonseca, Flavia Aburjaile, Rodrigo Tocantins Calado. FAMERP-SJRP: Cecília Artico Banho, Lívia Sacchetto, Fábio Sossai Possebon, Leila Sabrina Ullmann, Cíntia Bittar, Guilherme Campos, Helena Lage Ferreira, Jorge A. Petroli Marchesi, Maisa C. Pereira Parra, Marília Moraes, Paula Rahal, Paulo Inacio da Costa, João Pessoa Araújo Jr., Maurício Lacerda Nogueira. Prefeitura de Sao Paulo: Melissa Palmieri. |
| EPI_ISL_1967098, EPI_ISL_1967099, EPI_ISL_1967100, EPI_ISL_1967101, EPI_ISL_1967102, EPI_ISL_1967103, EPI_ISL_1967104, EPI_ISL_1967105, EPI_ISL_1967106, EPI_ISL_1967107, EPI_ISL_1967108, EPI_ISL_1967109, EPI_ISL_1967110, EPI_ISL_1967111, EPI_ISL_1967112, EPI_ISL_1967113, EPI_ISL_1967114, EPI_ISL_1967115, EPI_ISL_1967116, EPI_ISL_1967117, EPI_ISL_1967118, EPI_ISL_1967119, EPI_ISL_1967120, EPI_ISL_1967121, EPI_ISL_1967122, EPI_ISL_1967123, EPI_ISL_1967124, EPI_ISL_1967125, EPI_ISL_1967126, EPI_ISL_1967127, EPI_ISL_1967128, EPI_ISL_1967129, EPI_ISL_1967130, EPI_ISL_1967131, EPI_ISL_1967132, EPI_ISL_1967133, EPI_ISL_1967134, EPI_ISL_1967135, EPI_ISL_1967136, EPI_ISL_1967137, EPI_ISL_1967138, EPI_ISL_1967139, EPI_ISL_1967140, EPI_ISL_1967141, EPI_ISL_1967142, EPI_ISL_1967143, EPI_ISL_1967144, EPI_ISL_1967145, EPI_ISL_1967146, EPI_ISL_1967147 | | | |
| see above | SECRETARIA MUNICIPAL DE SAUDE SOROCABA | Instituto Butantan / ESALQ-USP (Piracicaba) | Instituto Butantan: Dimas Tadeu Covas, Sandra Coccuzzo Sampaio, Maria Carolina Elias, José Salvatore Leister Patané, Vincent Louis Viala, Antonio Jorge Martins, Ricardo Haddad, Claudia Renata dos Santos Barros, Elaine Cristina Marqueze, Raul Machado Neto, Debora Botequio Moretti, Jardelina de Souza Todao Bernardino, Loyze Paola Oliveira de Lima, Luiz Aurelio de Campos Crispin. Centro de Genômica Funcional da ESALQ: Luiz Lehmann Coutinho, Ricardo Augusto Brassaloti, Raquel de Lello Rocha Campos Cassano. NGS Soluções Genômicas: Pilar Drummond Sampaio Corrêa Mariani. FZEA-USP Pirassununga: Mirele Daiana Poleti, Jessika Cristina Chagas Lesbon, Elisangela Chicaroni Mattos, Heidge Fukumasu. USP-Botucatu: Rejane Maria Tommasini Grotto, Jayme A. Souza-Neto, Guilherme Targino Valente, Patricia Akemi Assato, Felipe Allan da Silva da Costa, Bianca Cechetto Carlos. Mendelics: Bibiana Santos, João Paulo Kitajima, Erika Freitas, David Schlesinger. Hemocentro Ribeirão Preto: Simone Kashima, Evandra Strazza Rodrigues, Svetoslav Nanev Slavov, Elaine Vieira dos Santos, Rafael dos Santos Bezerra, Luiz Carlos Junior de Alcantara, Marta Giovanetti, Vagner Fonseca, Flavia Aburjaile, Rodrigo Tocantins Calado. FAMERP-SJRP: Cecília Artico Banho, Lívia Sacchetto, Fábio Sossai Possebon, Leila Sabrina Ullmann, Cíntia Bittar, Guilherme Campos, Helena Lage Ferreira, Jorge A. Petroli Marchesi, Maisa C. Pereira Parra, Marília Moraes, Paula Rahal, Paulo Inacio da Costa, João Pessoa Araújo Jr., Maurício Lacerda Nogueira. Prefeitura de Sao Paulo: Melissa Palmieri. |
| EPI_ISL_1967148, EPI_ISL_1967149, EPI_ISL_1967150, EPI_ISL_1967151, EPI_ISL_1967152, EPI_ISL_1967153, EPI_ISL_1967154, EPI_ISL_1967155, EPI_ISL_1967156, EPI_ISL_1967158 | LABORATORIO MUNICIPAL DE SUZANO | Instituto Butantan / ESALQ-USP (Piracicaba) | Instituto Butantan: Dimas Tadeu Covas, Sandra Coccuzzo Sampaio, Maria Carolina Elias, José Salvatore Leister Patané, Vincent Louis Viala, Antonio Jorge Martins, Ricardo Haddad, Claudia Renata dos Santos Barros, Elaine Cristina Marqueze, Raul Machado Neto, Debora Botequio Moretti, Jardelina de Souza Todao Bernardino, Loyze Paola Oliveira de Lima, Luiz Aurelio de Campos Crispin. Centro de Genômica Funcional da ESALQ: Luiz Lehmann Coutinho, Ricardo Augusto Brassaloti, Raquel de Lello Rocha Campos Cassano. NGS Soluções Genômicas: Pilar Drummond Sampaio Corrêa Mariani. FZEA-USP Pirassununga: Mirele Daiana Poleti, Jessika Cristina Chagas Lesbon, Elisangela Chicaroni Mattos, Heidge Fukumasu. USP-Botucatu: Rejane Maria Tommasini Grotto, Jayme A. Souza-Neto, Guilherme Targino Valente, Patricia Akemi Assato, Felipe Allan da Silva da Costa, Bianca Cechetto Carlos. Mendelics: Bibiana Santos, João Paulo Kitajima, Erika Freitas, David Schlesinger. Hemocentro Ribeirão Preto: Simone Kashima, Evandra Strazza Rodrigues, Svetoslav Nanev Slavov, Elaine Vieira dos Santos, Rafael dos Santos Bezerra, Luiz Carlos Junior de Alcantara, Marta Giovanetti, Vagner Fonseca, Flavia Aburjaile, Rodrigo Tocantins Calado. FAMERP-SJRP: Cecília Artico Banho, Lívia Sacchetto, Fábio Sossai Possebon, Leila Sabrina Ullmann, Cíntia Bittar, Guilherme Campos, Helena Lage Ferreira, Jorge A. Petroli Marchesi, Maisa C. Pereira Parra, Marília Moraes, Paula Rahal, Paulo Inacio da Costa, João Pessoa Araújo Jr., Maurício Lacerda Nogueira. Prefeitura de Sao Paulo: Melissa Palmieri. |
| EPI_ISL_1967159, EPI_ISL_1967160, EPI_ISL_1967161, EPI_ISL_1967162 | UBS DE TABAPUA | Instituto Butantan / ESALQ-USP (Piracicaba) | Instituto Butantan: Dimas Tadeu Covas, Sandra Coccuzzo Sampaio, Maria Carolina Elias, José Salvatore Leister Patané, Vincent Louis Viala, Antonio Jorge Martins, Ricardo Haddad, Claudia Renata dos Santos Barros, Elaine Cristina Marqueze, Raul Machado Neto, Debora Botequio Moretti, Jardelina de Souza Todao Bernardino, Loyze Paola Oliveira de Lima, Luiz Aurelio de Campos Crispin. Centro de Genômica Funcional da ESALQ: Luiz Lehmann Coutinho, Ricardo Augusto Brassaloti, Raquel de Lello Rocha Campos Cassano. NGS Soluções Genômicas: Pilar Drummond Sampaio Corrêa Mariani. FZEA-USP Pirassununga: Mirele Daiana Poleti, Jessika Cristina Chagas Lesbon, Elisangela Chicaroni Mattos, Heidge Fukumasu. USP-Botucatu: Rejane Maria Tommasini Grotto, Jayme A. Souza-Neto, Guilherme Targino Valente, Patricia Akemi Assato, Felipe Allan da Silva da Costa, Bianca Cechetto Carlos. Mendelics: Bibiana Santos, João Paulo Kitajima, Erika Freitas, David Schlesinger. Hemocentro Ribeirão Preto: Simone Kashima, Evandra Strazza Rodrigues, Svetoslav Nanev Slavov, Elaine Vieira dos Santos, Rafael dos Santos Bezerra, Luiz Carlos Junior de Alcantara, Marta Giovanetti, Vagner Fonseca, Flavia Aburjaile, Rodrigo Tocantins Calado. FAMERP-SJRP: Cecília Artico Banho, Lívia Sacchetto, Fábio Sossai Possebon, Leila Sabrina Ullmann, Cíntia Bittar, Guilherme Campos, Helena Lage Ferreira, Jorge A. Petroli Marchesi, Maisa C. Pereira Parra, Marília Moraes, Paula Rahal, Paulo Inacio da Costa, João Pessoa Araújo Jr., Maurício Lacerda Nogueira. Prefeitura de Sao Paulo: Melissa Palmieri. |
| EPI_ISL_1967163 | UBS ENRIQUE GUILHERME ERNESTO MARZEUSKI | Instituto Butantan / ESALQ-USP (Piracicaba) | Instituto Butantan: Dimas Tadeu Covas, Sandra Coccuzzo Sampaio, Maria Carolina Elias, José Salvatore Leister Patané, Vincent Louis Viala, Antonio Jorge Martins, Ricardo Haddad, Claudia Renata dos Santos Barros, Elaine Cristina Marqueze, Raul Machado Neto, Debora Botequio Moretti, Jardelina de Souza Todao Bernardino, Loyze Paola Oliveira de Lima, Luiz Aurelio de Campos Crispin. Centro de Genômica Funcional da ESALQ: Luiz Lehmann Coutinho, Ricardo Augusto Brassaloti, Raquel de Lello Rocha Campos Cassano. NGS Soluções Genômicas: Pilar Drummond Sampaio Corrêa Mariani. FZEA-USP Pirassununga: Mirele Daiana Poleti, Jessika Cristina Chagas Lesbon, Elisangela Chicaroni Mattos, Heidge Fukumasu. USP-Botucatu: Rejane Maria Tommasini Grotto, Jayme A. Souza-Neto, Guilherme Targino Valente, Patricia Akemi Assato, Felipe Allan da Silva da Costa, Bianca Cechetto Carlos. Mendelics: Bibiana Santos, João Paulo Kitajima, Erika Freitas, David Schlesinger. Hemocentro Ribeirão Preto: Simone Kashima, Evandra Strazza Rodrigues, Svetoslav Nanev Slavov, Elaine Vieira dos Santos, Rafael dos Santos Bezerra, Luiz Carlos Junior de Alcantara, Marta Giovanetti, Vagner Fonseca, Flavia Aburjaile, Rodrigo Tocantins Calado. FAMERP-SJRP: Cecília Artico Banho, Lívia Sacchetto, Fábio Sossai Possebon, Leila Sabrina Ullmann, Cíntia Bittar, Guilherme Campos, Helena Lage Ferreira, Jorge A. Petroli Marchesi, Maisa C. Pereira Parra, Marília Moraes, Paula Rahal, Paulo Inacio da Costa, João Pessoa Araújo Jr., Maurício Lacerda Nogueira. Prefeitura de Sao Paulo: Melissa Palmieri. |
| EPI_ISL_1967165, EPI_ISL_1967166, EPI_ISL_1967167, EPI_ISL_1967168, EPI_ISL_1967169, EPI_ISL_1967170, EPI_ISL_1967171, EPI_ISL_1967172, EPI_ISL_1967173, EPI_ISL_1967174, EPI_ISL_1967175, EPI_ISL_1967176, EPI_ISL_1967177, EPI_ISL_1967178, EPI_ISL_1967179, EPI_ISL_1967180, EPI_ISL_1967181, EPI_ISL_1967182, EPI_ISL_1967183, EPI_ISL_1967184, EPI_ISL_1967185, EPI_ISL_1967186, EPI_ISL_1967187, EPI_ISL_1967188, EPI_ISL_1967189, EPI_ISL_1967190, EPI_ISL_1967191 | | | |
| see above | HOSPITAL DE CAMPANHA COVID 19 MUNICIPIO DE TAUBATE | Instituto Butantan / ESALQ-USP (Piracicaba) | Instituto Butantan: Dimas Tadeu Covas, Sandra Coccuzzo Sampaio, Maria Carolina Elias, José Salvatore Leister Patané, Vincent Louis Viala, Antonio Jorge Martins, Ricardo Haddad, Claudia Renata dos Santos Barros, Elaine Cristina Marqueze, Raul Machado Neto, Debora Botequio Moretti, Jardelina de Souza Todao Bernardino, Loyze Paola Oliveira de Lima, Luiz Aurelio de Campos Crispin. Centro de Genômica Funcional da ESALQ: Luiz Lehmann Coutinho, Ricardo Augusto Brassaloti, Raquel de Lello Rocha Campos Cassano. NGS Soluções Genômicas: Pilar Drummond Sampaio Corrêa Mariani. FZEA-USP Pirassununga: Mirele Daiana Poleti, Jessika Cristina Chagas Lesbon, Elisangela Chicaroni Mattos, Heidge Fukumasu. USP-Botucatu: Rejane Maria Tommasini Grotto, Jayme A. Souza-Neto, Guilherme Targino Valente, Patricia Akemi Assato, Felipe Allan da Silva da Costa, Bianca Cechetto |

| | | | |
|---|---|---|---|
| | | | Carlos. Mendelics: Bibiana Santos, João Paulo Kitajima, Erika Freitas, David Schlesinger. Hemocentro Ribeirão Preto: Simone Kashima, Evandra Strazza Rodrigues, Svetoslav Nanev Slavov, Elaine Vieira dos Santos, Rafael dos Santos Bezerra, Luiz Carlos Junior de Alcantara, Marta Giovanetti, Vagner Fonseca, Flavia Aburjaile, Rodrigo Tocantins Calado. FAMERP-SJRP: Cecília Artico Banho, Lívia Sacchetto, Fábio Sossai Possebon, Leila Sabrina Ullmann, Cíntia Bittar, Guilherme Campos, Helena Lage Ferreira, Jorge A. Petroli Marchesi, Maisa C. Pereira Parra, Marília Moraes, Paula Rahal, Paulo Inacio da Costa, João Pessoa Araújo Jr., Maurício Lacerda Nogueira. Prefeitura de Sao Paulo: Melissa Palmieri. |
| EPI_ISL_1967192, EPI_ISL_1967193, EPI_ISL_1967194, EPI_ISL_1967195 | SANTA CASA DE MISERICORDIA DE TIETE | Instituto Butantan / ESALQ-USP (Piracicaba) | Instituto Butantan: Dimas Tadeu Covas, Sandra Coccuzzo Sampaio, Maria Carolina Elias, José Salvatore Leister Patané, Vincent Louis Viala, Antonio Jorge Martins, Ricardo Haddad, Claudia Renata dos Santos Barros, Elaine Cristina Marqueze, Raul Machado Neto, Debora Botequio Moretti, Jardelina de Souza Todao Bernardino, Loyze Paola Oliveira de Lima, Luiz Aurelio de Campos Crispin. Centro de Genômica Funcional da ESALQ: Luiz Lehmann Coutinho, Ricardo Augusto Brassaloti, Raquel de Lello Rocha Campos Cassano. NGS Soluções Genômicas: Pilar Drummond Sampaio Corrêa Mariani. FZEA-USP Pirassununga: Mirele Daiana Poleti, Jessika Cristina Chagas Lesbon, Elisangela Chicaroni Mattos, Heidge Fukumasu. USP-Botucatu: Rejane Maria Tommasini Grotto, Jayme A. Souza-Neto, Guilherme Targino Valente, Patricia Akemi Assato, Felipe Allan da Silva da Costa, Bianca Cechetto Carlos. Mendelics: Bibiana Santos, João Paulo Kitajima, Erika Freitas, David Schlesinger. Hemocentro Ribeirão Preto: Simone Kashima, Evandra Strazza Rodrigues, Svetoslav Nanev Slavov, Elaine Vieira dos Santos, Rafael dos Santos Bezerra, Luiz Carlos Junior de Alcantara, Marta Giovanetti, Vagner Fonseca, Flavia Aburjaile, Rodrigo Tocantins Calado. FAMERP-SJRP: Cecília Artico Banho, Lívia Sacchetto, Fábio Sossai Possebon, Leila Sabrina Ullmann, Cíntia Bittar, Guilherme Campos, Helena Lage Ferreira, Jorge A. Petroli Marchesi, Maisa C. Pereira Parra, Marília Moraes, Paula Rahal, Paulo Inacio da Costa, João Pessoa Araújo Jr., Maurício Lacerda Nogueira. Prefeitura de Sao Paulo: Melissa Palmieri. |
| EPI_ISL_1967197 | DEPARTAMENTO DE SAUDE COLETIVA | Instituto Butantan / ESALQ-USP (Piracicaba) | Instituto Butantan: Dimas Tadeu Covas, Sandra Coccuzzo Sampaio, Maria Carolina Elias, José Salvatore Leister Patané, Vincent Louis Viala, Antonio Jorge Martins, Ricardo Haddad, Claudia Renata dos Santos Barros, Elaine Cristina Marqueze, Raul Machado Neto, Debora Botequio Moretti, Jardelina de Souza Todao Bernardino, Loyze Paola Oliveira de Lima, Luiz Aurelio de Campos Crispin. Centro de Genômica Funcional da ESALQ: Luiz Lehmann Coutinho, Ricardo Augusto Brassaloti, Raquel de Lello Rocha Campos Cassano. NGS Soluções Genômicas: Pilar Drummond Sampaio Corrêa Mariani. FZEA-USP Pirassununga: Mirele Daiana Poleti, Jessika Cristina Chagas Lesbon, Elisangela Chicaroni Mattos, Heidge Fukumasu. USP-Botucatu: Rejane Maria Tommasini Grotto, Jayme A. Souza-Neto, Guilherme Targino Valente, Patricia Akemi Assato, Felipe Allan da Silva da Costa, Bianca Cechetto Carlos. Mendelics: Bibiana Santos, João Paulo Kitajima, Erika Freitas, David Schlesinger. Hemocentro Ribeirão Preto: Simone Kashima, Evandra Strazza Rodrigues, Svetoslav Nanev Slavov, Elaine Vieira dos Santos, Rafael dos Santos Bezerra, Luiz Carlos Junior de Alcantara, Marta Giovanetti, Vagner Fonseca, Flavia Aburjaile, Rodrigo Tocantins Calado. FAMERP-SJRP: Cecília Artico Banho, Lívia Sacchetto, Fábio Sossai Possebon, Leila Sabrina Ullmann, Cíntia Bittar, Guilherme Campos, Helena Lage Ferreira, Jorge A. Petroli Marchesi, Maisa C. Pereira Parra, Marília Moraes, Paula Rahal, Paulo Inacio da Costa, João Pessoa Araújo Jr., Maurício Lacerda Nogueira. Prefeitura de Sao Paulo: Melissa Palmieri. |
| EPI_ISL_1967198, EPI_ISL_1967199, EPI_ISL_1967200, EPI_ISL_1967201, EPI_ISL_1967202, EPI_ISL_1967203 | VIGILANCIA EM SAUDE SAE VIGILANCIA EPIDEMIOLOGICA | Instituto Butantan / Mendelics | Instituto Butantan: Dimas Tadeu Covas, Sandra Coccuzzo Sampaio, Maria Carolina Elias, José Salvatore Leister Patané, Vincent Louis Viala, Antonio Jorge Martins, Ricardo Haddad, Claudia Renata dos Santos Barros, Elaine Cristina Marqueze, Raul Machado Neto, Debora Botequio Moretti, Jardelina de Souza Todao Bernardino, Loyze Paola Oliveira de Lima, Luiz Aurelio de Campos Crispin. Centro de Genômica Funcional da ESALQ: Luiz Lehmann Coutinho, Ricardo Augusto Brassaloti, Raquel de Lello Rocha Campos Cassano. NGS Soluções Genômicas: Pilar Drummond Sampaio Corrêa Mariani. FZEA-USP Pirassununga: Mirele Daiana Poleti, Jessika Cristina Chagas Lesbon, Elisangela Chicaroni Mattos, Heidge Fukumasu. USP-Botucatu: Rejane Maria Tommasini Grotto, Jayme A. Souza-Neto, Guilherme Targino Valente, Patricia Akemi Assato, Felipe Allan da Silva da Costa, Bianca Cechetto Carlos. Mendelics: Bibiana Santos, João Paulo Kitajima, Erika Freitas, David Schlesinger. Hemocentro Ribeirão Preto: Simone Kashima, Evandra Strazza Rodrigues, Svetoslav Nanev Slavov, Elaine Vieira dos Santos, Rafael dos Santos Bezerra, Luiz Carlos Junior de Alcantara, Marta Giovanetti, Vagner Fonseca, Flavia Aburjaile, Rodrigo Tocantins Calado. FAMERP-SJRP: Cecília Artico Banho, Lívia Sacchetto, Fábio Sossai Possebon, Leila Sabrina Ullmann, Cíntia Bittar, Guilherme Campos, Helena Lage Ferreira, Jorge A. Petroli Marchesi, Maisa C. Pereira Parra, Marília Moraes, Paula Rahal, Paulo Inacio da Costa, João Pessoa Araújo Jr., Maurício Lacerda Nogueira. Prefeitura de Sao Paulo: Melissa Palmieri. |
| EPI_ISL_1967204, EPI_ISL_1967205, EPI_ISL_1967206, EPI_ISL_1967207, EPI_ISL_1967208, EPI_ISL_1967209, EPI_ISL_1967210, EPI_ISL_1967211 | UBS DR ALFREDO DANTAS DE SOUZA UMUARAMA | Instituto Butantan / Mendelics | Instituto Butantan: Dimas Tadeu Covas, Sandra Coccuzzo Sampaio, Maria Carolina Elias, José Salvatore Leister Patané, Vincent Louis Viala, Antonio Jorge Martins, Ricardo Haddad, Claudia Renata dos Santos Barros, Elaine Cristina Marqueze, Raul Machado Neto, Debora Botequio Moretti, Jardelina de Souza Todao Bernardino, Loyze Paola Oliveira de Lima, Luiz Aurelio de Campos Crispin. Centro de Genômica Funcional da ESALQ: Luiz Lehmann Coutinho, Ricardo Augusto Brassaloti, Raquel de Lello Rocha Campos Cassano. NGS Soluções Genômicas: Pilar Drummond Sampaio Corrêa Mariani. FZEA-USP Pirassununga: Mirele Daiana Poleti, Jessika Cristina Chagas Lesbon, Elisangela Chicaroni Mattos, Heidge Fukumasu. USP-Botucatu: Rejane Maria Tommasini Grotto, Jayme A. Souza-Neto, Guilherme Targino Valente, Patricia Akemi Assato, Felipe Allan da Silva da Costa, Bianca Cechetto Carlos. Mendelics: Bibiana Santos, João Paulo Kitajima, Erika Freitas, David Schlesinger. Hemocentro Ribeirão Preto: Simone Kashima, Evandra Strazza Rodrigues, Svetoslav Nanev Slavov, Elaine Vieira dos Santos, Rafael dos Santos Bezerra, Luiz Carlos Junior de Alcantara, Marta Giovanetti, Vagner Fonseca, Flavia Aburjaile, Rodrigo Tocantins Calado. FAMERP-SJRP: Cecília Artico Banho, Lívia Sacchetto, Fábio Sossai Possebon, Leila Sabrina Ullmann, Cíntia Bittar, Guilherme Campos, Helena Lage Ferreira, Jorge A. Petroli Marchesi, Maisa C. Pereira Parra, Marília Moraes, Paula Rahal, Paulo Inacio da Costa, João Pessoa Araújo Jr., Maurício Lacerda Nogueira. Prefeitura de Sao Paulo: Melissa Palmieri. |
| EPI_ISL_1967212 | CENTRO DE SAUDE DE BARBOSA | Instituto Butantan / Mendelics | Instituto Butantan: Dimas Tadeu Covas, Sandra Coccuzzo Sampaio, Maria Carolina Elias, José Salvatore Leister Patané, Vincent Louis Viala, Antonio Jorge Martins, Ricardo Haddad, Claudia Renata dos Santos Barros, Elaine Cristina Marqueze, Raul Machado Neto, Debora Botequio Moretti, Jardelina de Souza Todao Bernardino, Loyze Paola Oliveira de Lima, Luiz Aurelio de Campos Crispin. Centro de Genômica Funcional da ESALQ: Luiz Lehmann Coutinho, Ricardo Augusto Brassaloti, Raquel de Lello Rocha Campos Cassano. NGS Soluções Genômicas: Pilar Drummond Sampaio Corrêa Mariani. FZEA-USP Pirassununga: Mirele Daiana Poleti, Jessika Cristina Chagas Lesbon, Elisangela Chicaroni Mattos, Heidge Fukumasu. USP-Botucatu: Rejane Maria Tommasini Grotto, Jayme A. Souza-Neto, Guilherme Targino Valente, Patricia Akemi Assato, Felipe Allan da Silva da Costa, Bianca Cechetto Carlos. Mendelics: Bibiana Santos, João Paulo Kitajima, Erika Freitas, David Schlesinger. Hemocentro Ribeirão Preto: Simone Kashima, Evandra Strazza Rodrigues, Svetoslav Nanev Slavov, Elaine Vieira dos Santos, Rafael dos Santos Bezerra, Luiz Carlos Junior de Alcantara, Marta Giovanetti, Vagner Fonseca, Flavia Aburjaile, Rodrigo Tocantins Calado. FAMERP-SJRP: Cecília Artico Banho, Lívia Sacchetto, Fábio Sossai Possebon, Leila Sabrina Ullmann, Cíntia Bittar, Guilherme Campos, Helena Lage Ferreira, Jorge A. Petroli Marchesi, Maisa C. Pereira Parra, Marília Moraes, Paula Rahal, Paulo Inacio da Costa, João Pessoa Araújo Jr., Maurício Lacerda Nogueira. Prefeitura de Sao Paulo: Melissa Palmieri. |
| EPI_ISL_1967213 | NUCLEO DE SAUDE MARY DOTA DE BAURU | Instituto Butantan / Mendelics | Instituto Butantan: Dimas Tadeu Covas, Sandra Coccuzzo Sampaio, Maria Carolina Elias, José Salvatore Leister Patané, Vincent Louis Viala, Antonio Jorge Martins, Ricardo Haddad, Claudia Renata dos Santos Barros, Elaine Cristina Marqueze, Raul Machado Neto, Debora Botequio Moretti, Jardelina de Souza Todao Bernardino, Loyze Paola Oliveira de Lima, Luiz Aurelio de Campos Crispin. Centro de Genômica Funcional da ESALQ: Luiz Lehmann Coutinho, Ricardo Augusto Brassaloti, Raquel de Lello Rocha Campos Cassano. NGS Soluções Genômicas: Pilar Drummond Sampaio Corrêa Mariani. FZEA-USP Pirassununga: Mirele Daiana Poleti, Jessika Cristina Chagas Lesbon, Elisangela Chicaroni Mattos, Heidge Fukumasu. USP-Botucatu: Rejane Maria Tommasini Grotto, Jayme A. Souza-Neto, Guilherme Targino Valente, Patricia Akemi Assato, Felipe Allan da Silva da Costa, Bianca Cechetto Carlos. Mendelics: Bibiana Santos, João Paulo Kitajima, Erika Freitas, David Schlesinger. Hemocentro Ribeirão Preto: Simone Kashima, Evandra Strazza Rodrigues, Svetoslav Nanev Slavov, Elaine Vieira dos Santos, Rafael dos Santos Bezerra, Luiz Carlos Junior de Alcantara, Marta Giovanetti, Vagner Fonseca, Flavia Aburjaile, Rodrigo Tocantins Calado. FAMERP-SJRP: Cecília Artico Banho, Lívia Sacchetto, Fábio Sossai Possebon, Leila Sabrina Ullmann, Cíntia Bittar, Guilherme Campos, Helena Lage Ferreira, Jorge A. Petroli Marchesi, Maisa C. Pereira Parra, Marília Moraes, Paula Rahal, Paulo Inacio da Costa, João Pessoa Araújo Jr., Maurício Lacerda Nogueira. Prefeitura de Sao Paulo: Melissa Palmieri. |
| EPI_ISL_1967216 | UPA MARY DOTA | Instituto Butantan / Mendelics | Instituto Butantan: Dimas Tadeu Covas, Sandra Coccuzzo Sampaio, Maria Carolina Elias, José Salvatore Leister Patané, Vincent Louis Viala, Antonio Jorge Martins, Ricardo Haddad, Claudia Renata dos Santos Barros, Elaine Cristina Marqueze, Raul Machado Neto, Debora Botequio Moretti, Jardelina de Souza Todao Bernardino, Loyze Paola Oliveira de Lima, Luiz Aurelio de Campos Crispin. Centro de Genômica Funcional da ESALQ: Luiz Lehmann Coutinho, Ricardo Augusto Brassaloti, Raquel de Lello Rocha Campos Cassano. NGS Soluções Genômicas: Pilar Drummond Sampaio Corrêa Mariani. FZEA-USP Pirassununga: Mirele Daiana Poleti, Jessika Cristina Chagas Lesbon, Elisangela Chicaroni Mattos, Heidge Fukumasu. USP-Botucatu: Rejane Maria Tommasini Grotto, Jayme A. Souza-Neto, Guilherme Targino Valente, Patricia Akemi Assato, Felipe Allan da Silva da Costa, Bianca Cechetto Carlos. Mendelics: Bibiana Santos, João Paulo Kitajima, Erika Freitas, David Schlesinger. Hemocentro Ribeirão Preto: Simone Kashima, Evandra Strazza Rodrigues, Svetoslav Nanev Slavov, Elaine Vieira dos Santos, Rafael dos Santos Bezerra, Luiz Carlos Junior de Alcantara, Marta Giovanetti, Vagner Fonseca, Flavia Aburjaile, Rodrigo Tocantins Calado. FAMERP-SJRP: Cecília Artico Banho, Lívia Sacchetto, Fábio Sossai Possebon, Leila Sabrina Ullmann, Cíntia Bittar, Guilherme Campos, Helena Lage Ferreira, Jorge A. Petroli Marchesi, Maisa C. Pereira Parra, Marília Moraes, Paula Rahal, Paulo Inacio da Costa, João Pessoa Araújo Jr., Maurício Lacerda Nogueira. Prefeitura de Sao Paulo: Melissa Palmieri. |
| EPI_ISL_1967217 | NUCLEO DE SAUDE MARY DOTA DE BAURU | Instituto Butantan / Mendelics | Instituto Butantan: Dimas Tadeu Covas, Sandra Coccuzzo Sampaio, Maria Carolina Elias, José Salvatore Leister Patané, Vincent Louis Viala, Antonio |

| | | | |
|---|---|---|---|
| | | | Jorge Martins, Ricardo Haddad, Claudia Renata dos Santos Barros, Elaine Cristina Marqueze, Raul Machado Neto, Debora Botequio Moretti, Jardelina de Souza Todao Bernardino, Loyze Paola Oliveira de Lima, Luiz Aurelio de Campos Crispin. Centro de Genômica Funcional da ESALQ: Luiz Lehmann Coutinho, Ricardo Augusto Brassaloti, Raquel de Lello Rocha Campos Cassano. NGS Soluções Genômicas: Pilar Drummond Sampaio Corrêa Mariani. FZEA-USP Pirassununga: Mirele Daiana Poleti, Jessika Cristina Chagas Lesbon, Elisangela Chicaroni Mattos, Heidge Fukumasu. USP-Botucatu: Rejane Maria Tommasini Grotto, Jayme A. Souza-Neto, Guilherme Targino Valente, Patricia Akemi Assato, Felipe Allan da Silva da Costa, Bianca Cechetto Carlos. Mendelics: Bibiana Santos, João Paulo Kitajima, Erika Freitas, David Schlesinger. Hemocentro Ribeirão Preto: Simone Kashima, Evandra Strazza Rodrigues, Svetoslav Nanev Slavov, Elaine Vieira dos Santos, Rafael dos Santos Bezerra, Luiz Carlos Junior de Alcantara, Marta Giovanetti, Vagner Fonseca, Flavia Aburjaile, Rodrigo Tocantins Calado. FAMERP-SJRP: Cecília Artico Banho, Lívia Sacchetto, Fábio Sossai Possebon, Leila Sabrina Ullmann, Cíntia Bittar, Guilherme Campos, Helena Lage Ferreira, Jorge A. Petroli Marchesi, Maisa C. Pereira Parra, Marília Moraes, Paula Rahal, Paulo Inacio da Costa, João Pessoa Araújo Jr., Maurício Lacerda Nogueira. Prefeitura de Sao Paulo: Melissa Palmieri. |
| EPI_ISL_1967218, EPI_ISL_1967219, EPI_ISL_1967220, EPI_ISL_1967221, EPI_ISL_1967222, EPI_ISL_1967223, EPI_ISL_1967224, EPI_ISL_1967225, EPI_ISL_1967226, EPI_ISL_1967227 | UPA DE BEBEDOURO | Instituto Butantan / Mendelics | Instituto Butantan: Dimas Tadeu Covas, Sandra Coccuzzo Sampaio, Maria Carolina Elias, José Salvatore Leister Patané, Vincent Louis Viala, Antonio Jorge Martins, Ricardo Haddad, Claudia Renata dos Santos Barros, Elaine Cristina Marqueze, Raul Machado Neto, Debora Botequio Moretti, Jardelina de Souza Todao Bernardino, Loyze Paola Oliveira de Lima, Luiz Aurelio de Campos Crispin. Centro de Genômica Funcional da ESALQ: Luiz Lehmann Coutinho, Ricardo Augusto Brassaloti, Raquel de Lello Rocha Campos Cassano. NGS Soluções Genômicas: Pilar Drummond Sampaio Corrêa Mariani. FZEA-USP Pirassununga: Mirele Daiana Poleti, Jessika Cristina Chagas Lesbon, Elisangela Chicaroni Mattos, Heidge Fukumasu. USP-Botucatu: Rejane Maria Tommasini Grotto, Jayme A. Souza-Neto, Guilherme Targino Valente, Patricia Akemi Assato, Felipe Allan da Silva da Costa, Bianca Cechetto Carlos. Mendelics: Bibiana Santos, João Paulo Kitajima, Erika Freitas, David Schlesinger. Hemocentro Ribeirão Preto: Simone Kashima, Evandra Strazza Rodrigues, Svetoslav Nanev Slavov, Elaine Vieira dos Santos, Rafael dos Santos Bezerra, Luiz Carlos Junior de Alcantara, Marta Giovanetti, Vagner Fonseca, Flavia Aburjaile, Rodrigo Tocantins Calado. FAMERP-SJRP: Cecília Artico Banho, Lívia Sacchetto, Fábio Sossai Possebon, Leila Sabrina Ullmann, Cíntia Bittar, Guilherme Campos, Helena Lage Ferreira, Jorge A. Petroli Marchesi, Maisa C. Pereira Parra, Marília Moraes, Paula Rahal, Paulo Inacio da Costa, João Pessoa Araújo Jr., Maurício Lacerda Nogueira. Prefeitura de Sao Paulo: Melissa Palmieri. |
| EPI_ISL_1967228 | UBS 09 JOAO CREVELARO BIRIGUI | Instituto Butantan / Mendelics | Instituto Butantan: Dimas Tadeu Covas, Sandra Coccuzzo Sampaio, Maria Carolina Elias, José Salvatore Leister Patané, Vincent Louis Viala, Antonio Jorge Martins, Ricardo Haddad, Claudia Renata dos Santos Barros, Elaine Cristina Marqueze, Raul Machado Neto, Debora Botequio Moretti, Jardelina de Souza Todao Bernardino, Loyze Paola Oliveira de Lima, Luiz Aurelio de Campos Crispin. Centro de Genômica Funcional da ESALQ: Luiz Lehmann Coutinho, Ricardo Augusto Brassaloti, Raquel de Lello Rocha Campos Cassano. NGS Soluções Genômicas: Pilar Drummond Sampaio Corrêa Mariani. FZEA-USP Pirassununga: Mirele Daiana Poleti, Jessika Cristina Chagas Lesbon, Elisangela Chicaroni Mattos, Heidge Fukumasu. USP-Botucatu: Rejane Maria Tommasini Grotto, Jayme A. Souza-Neto, Guilherme Targino Valente, Patricia Akemi Assato, Felipe Allan da Silva da Costa, Bianca Cechetto Carlos. Mendelics: Bibiana Santos, João Paulo Kitajima, Erika Freitas, David Schlesinger. Hemocentro Ribeirão Preto: Simone Kashima, Evandra Strazza Rodrigues, Svetoslav Nanev Slavov, Elaine Vieira dos Santos, Rafael dos Santos Bezerra, Luiz Carlos Junior de Alcantara, Marta Giovanetti, Vagner Fonseca, Flavia Aburjaile, Rodrigo Tocantins Calado. FAMERP-SJRP: Cecília Artico Banho, Lívia Sacchetto, Fábio Sossai Possebon, Leila Sabrina Ullmann, Cíntia Bittar, Guilherme Campos, Helena Lage Ferreira, Jorge A. Petroli Marchesi, Maisa C. Pereira Parra, Marília Moraes, Paula Rahal, Paulo Inacio da Costa, João Pessoa Araújo Jr., Maurício Lacerda Nogueira. Prefeitura de Sao Paulo: Melissa Palmieri. |
| EPI_ISL_1967229 | UBS 01 VILA BANDEIRANTES BIRIGUI | Instituto Butantan / Mendelics | Instituto Butantan: Dimas Tadeu Covas, Sandra Coccuzzo Sampaio, Maria Carolina Elias, José Salvatore Leister Patané, Vincent Louis Viala, Antonio Jorge Martins, Ricardo Haddad, Claudia Renata dos Santos Barros, Elaine Cristina Marqueze, Raul Machado Neto, Debora Botequio Moretti, Jardelina de Souza Todao Bernardino, Loyze Paola Oliveira de Lima, Luiz Aurelio de Campos Crispin. Centro de Genômica Funcional da ESALQ: Luiz Lehmann Coutinho, Ricardo Augusto Brassaloti, Raquel de Lello Rocha Campos Cassano. NGS Soluções Genômicas: Pilar Drummond Sampaio Corrêa Mariani. FZEA-USP Pirassununga: Mirele Daiana Poleti, Jessika Cristina Chagas Lesbon, Elisangela Chicaroni Mattos, Heidge Fukumasu. USP-Botucatu: Rejane Maria Tommasini Grotto, Jayme A. Souza-Neto, Guilherme Targino Valente, Patricia Akemi Assato, Felipe Allan da Silva da Costa, Bianca Cechetto Carlos. Mendelics: Bibiana Santos, João Paulo Kitajima, Erika Freitas, David Schlesinger. Hemocentro Ribeirão Preto: Simone Kashima, Evandra Strazza Rodrigues, Svetoslav Nanev Slavov, Elaine Vieira dos Santos, Rafael dos Santos Bezerra, Luiz Carlos Junior de Alcantara, Marta Giovanetti, Vagner Fonseca, Flavia Aburjaile, Rodrigo Tocantins Calado. FAMERP-SJRP: Cecília Artico Banho, Lívia Sacchetto, Fábio Sossai Possebon, Leila Sabrina Ullmann, Cíntia Bittar, Guilherme Campos, Helena Lage Ferreira, Jorge A. Petroli Marchesi, Maisa C. Pereira Parra, Marília Moraes, Paula Rahal, Paulo Inacio da Costa, João Pessoa Araújo Jr., Maurício Lacerda Nogueira. Prefeitura de Sao Paulo: Melissa Palmieri. |
| EPI_ISL_1967230 | UBS 10 MANOEL SANCHES GRANEIRO COLINAS BIRIGUI | Instituto Butantan / Mendelics | Instituto Butantan: Dimas Tadeu Covas, Sandra Coccuzzo Sampaio, Maria Carolina Elias, José Salvatore Leister Patané, Vincent Louis Viala, Antonio Jorge Martins, Ricardo Haddad, Claudia Renata dos Santos Barros, Elaine Cristina Marqueze, Raul Machado Neto, Debora Botequio Moretti, Jardelina de Souza Todao Bernardino, Loyze Paola Oliveira de Lima, Luiz Aurelio de Campos Crispin. Centro de Genômica Funcional da ESALQ: Luiz Lehmann Coutinho, Ricardo Augusto Brassaloti, Raquel de Lello Rocha Campos Cassano. NGS Soluções Genômicas: Pilar Drummond Sampaio Corrêa Mariani. FZEA-USP Pirassununga: Mirele Daiana Poleti, Jessika Cristina Chagas Lesbon, Elisangela Chicaroni Mattos, Heidge Fukumasu. USP-Botucatu: Rejane Maria Tommasini Grotto, Jayme A. Souza-Neto, Guilherme Targino Valente, Patricia Akemi Assato, Felipe Allan da Silva da Costa, Bianca Cechetto Carlos. Mendelics: Bibiana Santos, João Paulo Kitajima, Erika Freitas, David Schlesinger. Hemocentro Ribeirão Preto: Simone Kashima, Evandra Strazza Rodrigues, Svetoslav Nanev Slavov, Elaine Vieira dos Santos, Rafael dos Santos Bezerra, Luiz Carlos Junior de Alcantara, Marta Giovanetti, Vagner Fonseca, Flavia Aburjaile, Rodrigo Tocantins Calado. FAMERP-SJRP: Cecília Artico Banho, Lívia Sacchetto, Fábio Sossai Possebon, Leila Sabrina Ullmann, Cíntia Bittar, Guilherme Campos, Helena Lage Ferreira, Jorge A. Petroli Marchesi, Maisa C. Pereira Parra, Marília Moraes, Paula Rahal, Paulo Inacio da Costa, João Pessoa Araújo Jr., Maurício Lacerda Nogueira. Prefeitura de Sao Paulo: Melissa Palmieri. |
| EPI_ISL_1967231 | UBS 04 JARDIM COSTA RICA BIRIGUI | Instituto Butantan / Mendelics | Instituto Butantan: Dimas Tadeu Covas, Sandra Coccuzzo Sampaio, Maria Carolina Elias, José Salvatore Leister Patané, Vincent Louis Viala, Antonio Jorge Martins, Ricardo Haddad, Claudia Renata dos Santos Barros, Elaine Cristina Marqueze, Raul Machado Neto, Debora Botequio Moretti, Jardelina de Souza Todao Bernardino, Loyze Paola Oliveira de Lima, Luiz Aurelio de Campos Crispin. Centro de Genômica Funcional da ESALQ: Luiz Lehmann Coutinho, Ricardo Augusto Brassaloti, Raquel de Lello Rocha Campos Cassano. NGS Soluções Genômicas: Pilar Drummond Sampaio Corrêa Mariani. FZEA-USP Pirassununga: Mirele Daiana Poleti, Jessika Cristina Chagas Lesbon, Elisangela Chicaroni Mattos, Heidge Fukumasu. USP-Botucatu: Rejane Maria Tommasini Grotto, Jayme A. Souza-Neto, Guilherme Targino Valente, Patricia Akemi Assato, Felipe Allan da Silva da Costa, Bianca Cechetto Carlos. Mendelics: Bibiana Santos, João Paulo Kitajima, Erika Freitas, David Schlesinger. Hemocentro Ribeirão Preto: Simone Kashima, Evandra Strazza Rodrigues, Svetoslav Nanev Slavov, Elaine Vieira dos Santos, Rafael dos Santos Bezerra, Luiz Carlos Junior de Alcantara, Marta Giovanetti, Vagner Fonseca, Flavia Aburjaile, Rodrigo Tocantins Calado. FAMERP-SJRP: Cecília Artico Banho, Lívia Sacchetto, Fábio Sossai Possebon, Leila Sabrina Ullmann, Cíntia Bittar, Guilherme Campos, Helena Lage Ferreira, Jorge A. Petroli Marchesi, Maisa C. Pereira Parra, Marília Moraes, Paula Rahal, Paulo Inacio da Costa, João Pessoa Araújo Jr., Maurício Lacerda Nogueira. Prefeitura de Sao Paulo: Melissa Palmieri. |
| EPI_ISL_1967232 | UBS 09 JOAO CREVELARO BIRIGUI | Instituto Butantan / Mendelics | Instituto Butantan: Dimas Tadeu Covas, Sandra Coccuzzo Sampaio, Maria Carolina Elias, José Salvatore Leister Patané, Vincent Louis Viala, Antonio Jorge Martins, Ricardo Haddad, Claudia Renata dos Santos Barros, Elaine Cristina Marqueze, Raul Machado Neto, Debora Botequio Moretti, Jardelina de Souza Todao Bernardino, Loyze Paola Oliveira de Lima, Luiz Aurelio de Campos Crispin. Centro de Genômica Funcional da ESALQ: Luiz Lehmann Coutinho, Ricardo Augusto Brassaloti, Raquel de Lello Rocha Campos Cassano. NGS Soluções Genômicas: Pilar Drummond Sampaio Corrêa Mariani. FZEA-USP Pirassununga: Mirele Daiana Poleti, Jessika Cristina Chagas Lesbon, Elisangela Chicaroni Mattos, Heidge Fukumasu. USP-Botucatu: Rejane Maria Tommasini Grotto, Jayme A. Souza-Neto, Guilherme Targino Valente, Patricia Akemi Assato, Felipe Allan da Silva da Costa, Bianca Cechetto Carlos. Mendelics: Bibiana Santos, João Paulo Kitajima, Erika Freitas, David Schlesinger. Hemocentro Ribeirão Preto: Simone Kashima, Evandra Strazza Rodrigues, Svetoslav Nanev Slavov, Elaine Vieira dos Santos, Rafael dos Santos Bezerra, Luiz Carlos Junior de Alcantara, Marta Giovanetti, Vagner Fonseca, Flavia Aburjaile, Rodrigo Tocantins Calado. FAMERP-SJRP: Cecília Artico Banho, Lívia Sacchetto, Fábio Sossai Possebon, Leila Sabrina Ullmann, Cíntia Bittar, Guilherme Campos, Helena Lage Ferreira, Jorge A. Petroli Marchesi, Maisa C. Pereira Parra, Marília Moraes, Paula Rahal, Paulo Inacio da Costa, João Pessoa Araújo Jr., Maurício Lacerda Nogueira. Prefeitura de Sao Paulo: Melissa Palmieri. |
| EPI_ISL_1967233 | UBS 01 VILA BANDEIRANTES BIRIGUI | Instituto Butantan / Mendelics | Instituto Butantan: Dimas Tadeu Covas, Sandra Coccuzzo Sampaio, Maria Carolina Elias, José Salvatore Leister Patané, Vincent Louis Viala, Antonio Jorge Martins, Ricardo Haddad, Claudia Renata dos Santos Barros, Elaine Cristina Marqueze, Raul Machado Neto, Debora Botequio Moretti, Jardelina de Souza Todao Bernardino, Loyze Paola Oliveira de Lima, Luiz Aurelio de Campos Crispin. Centro de Genômica Funcional da ESALQ: Luiz Lehmann Coutinho, Ricardo Augusto Brassaloti, Raquel de Lello Rocha Campos Cassano. NGS Soluções Genômicas: Pilar Drummond Sampaio Corrêa Mariani. FZEA-USP Pirassununga: Mirele Daiana Poleti, Jessika Cristina Chagas Lesbon, Elisangela Chicaroni Mattos, Heidge Fukumasu. USP-Botucatu: Rejane Maria Tommasini Grotto, Jayme A. Souza-Neto, Guilherme Targino Valente, Patricia Akemi Assato, Felipe Allan da Silva da Costa, Bianca Cechetto Carlos. Mendelics: Bibiana Santos, João Paulo Kitajima, Erika Freitas, David Schlesinger. Hemocentro Ribeirão Preto: Simone Kashima, Evandra Strazza Rodrigues, Svetoslav Nanev Slavov, Elaine Vieira dos Santos, Rafael dos Santos Bezerra, Luiz Carlos Junior de Alcantara, Marta Giovanetti, Vagner |

| | | | |
|---|---|---|---|
| | | | Fonseca, Flavia Aburjaile, Rodrigo Tocantins Calado. FAMERP-SJRP: Cecília Artico Banho, Lívia Sacchetto, Fábio Sossai Possebon, Leila Sabrina Ullmann, Cíntia Bittar, Guilherme Campos, Helena Lage Ferreira, Jorge A. Petroli Marchesi, Maisa C. Pereira Parra, Marília Moraes, Paula Rahal, Paulo Inacio da Costa, João Pessoa Araújo Jr., Maurício Lacerda Nogueira. Prefeitura de Sao Paulo: Melissa Palmieri. |
| EPI_ISL_1967234 | UBS 02 JARDIM TOSELAR BIRIGUI | Instituto Butantan / Mendelics | Instituto Butantan: Dimas Tadeu Covas, Sandra Coccuzzo Sampaio, Maria Carolina Elias, José Salvatore Leister Patané, Vincent Louis Viala, Antonio Jorge Martins, Ricardo Haddad, Claudia Renata dos Santos Barros, Elaine Cristina Marqueze, Raul Machado Neto, Debora Botequio Moretti, Jardelina de Souza Todao Bernardino, Loyze Paola Oliveira de Lima, Luiz Aurelio de Campos Crispin. Centro de Genômica Funcional da ESALQ: Luiz Lehmann Coutinho, Ricardo Augusto Brassaloti, Raquel de Lello Rocha Campos Cassano. NGS Soluções Genômicas: Pilar Drummond Sampaio Corrêa Mariani. FZEA-USP Pirassununga: Mirele Daiana Poleti, Jessika Cristina Chagas Lesbon, Elisangela Chicaroni Mattos, Heidge Fukumasu. USP-Botucatu: Rejane Maria Tommasini Grotto, Jayme A. Souza-Neto, Guilherme Targino Valente, Patricia Akemi Assato, Felipe Allan da Silva da Costa, Bianca Cechetto Carlos. Mendelics: Bibiana Santos, João Paulo Kitajima, Erika Freitas, David Schlesinger. Hemocentro Ribeirão Preto: Simone Kashima, Evandra Strazza Rodrigues, Svetoslav Nanev Slavov, Elaine Vieira dos Santos, Rafael dos Santos Bezerra, Luiz Carlos Junior de Alcantara, Marta Giovanetti, Vagner Fonseca, Flavia Aburjaile, Rodrigo Tocantins Calado. FAMERP-SJRP: Cecília Artico Banho, Lívia Sacchetto, Fábio Sossai Possebon, Leila Sabrina Ullmann, Cíntia Bittar, Guilherme Campos, Helena Lage Ferreira, Jorge A. Petroli Marchesi, Maisa C. Pereira Parra, Marília Moraes, Paula Rahal, Paulo Inacio da Costa, João Pessoa Araújo Jr., Maurício Lacerda Nogueira. Prefeitura de Sao Paulo: Melissa Palmieri. |
| EPI_ISL_1967235 | UBS 11 DR SERGIO AUGUSTO CLARK XAVIER SOARES PORTAL II | Instituto Butantan / Mendelics | Instituto Butantan: Dimas Tadeu Covas, Sandra Coccuzzo Sampaio, Maria Carolina Elias, José Salvatore Leister Patané, Vincent Louis Viala, Antonio Jorge Martins, Ricardo Haddad, Claudia Renata dos Santos Barros, Elaine Cristina Marqueze, Raul Machado Neto, Debora Botequio Moretti, Jardelina de Souza Todao Bernardino, Loyze Paola Oliveira de Lima, Luiz Aurelio de Campos Crispin. Centro de Genômica Funcional da ESALQ: Luiz Lehmann Coutinho, Ricardo Augusto Brassaloti, Raquel de Lello Rocha Campos Cassano. NGS Soluções Genômicas: Pilar Drummond Sampaio Corrêa Mariani. FZEA-USP Pirassununga: Mirele Daiana Poleti, Jessika Cristina Chagas Lesbon, Elisangela Chicaroni Mattos, Heidge Fukumasu. USP-Botucatu: Rejane Maria Tommasini Grotto, Jayme A. Souza-Neto, Guilherme Targino Valente, Patricia Akemi Assato, Felipe Allan da Silva da Costa, Bianca Cechetto Carlos. Mendelics: Bibiana Santos, João Paulo Kitajima, Erika Freitas, David Schlesinger. Hemocentro Ribeirão Preto: Simone Kashima, Evandra Strazza Rodrigues, Svetoslav Nanev Slavov, Elaine Vieira dos Santos, Rafael dos Santos Bezerra, Luiz Carlos Junior de Alcantara, Marta Giovanetti, Vagner Fonseca, Flavia Aburjaile, Rodrigo Tocantins Calado. FAMERP-SJRP: Cecília Artico Banho, Lívia Sacchetto, Fábio Sossai Possebon, Leila Sabrina Ullmann, Cíntia Bittar, Guilherme Campos, Helena Lage Ferreira, Jorge A. Petroli Marchesi, Maisa C. Pereira Parra, Marília Moraes, Paula Rahal, Paulo Inacio da Costa, João Pessoa Araújo Jr., Maurício Lacerda Nogueira. Prefeitura de Sao Paulo: Melissa Palmieri. |
| EPI_ISL_1967236, EPI_ISL_1967237, EPI_ISL_1967238, EPI_ISL_1967239, EPI_ISL_1967240, EPI_ISL_1967241 | PRONTO SOCORRO CENTRAL GUIOMAR FERREIRA ROEBBELEN | Instituto Butantan / Mendelics | Instituto Butantan: Dimas Tadeu Covas, Sandra Coccuzzo Sampaio, Maria Carolina Elias, José Salvatore Leister Patané, Vincent Louis Viala, Antonio Jorge Martins, Ricardo Haddad, Claudia Renata dos Santos Barros, Elaine Cristina Marqueze, Raul Machado Neto, Debora Botequio Moretti, Jardelina de Souza Todao Bernardino, Loyze Paola Oliveira de Lima, Luiz Aurelio de Campos Crispin. Centro de Genômica Funcional da ESALQ: Luiz Lehmann Coutinho, Ricardo Augusto Brassaloti, Raquel de Lello Rocha Campos Cassano. NGS Soluções Genômicas: Pilar Drummond Sampaio Corrêa Mariani. FZEA-USP Pirassununga: Mirele Daiana Poleti, Jessika Cristina Chagas Lesbon, Elisangela Chicaroni Mattos, Heidge Fukumasu. USP-Botucatu: Rejane Maria Tommasini Grotto, Jayme A. Souza-Neto, Guilherme Targino Valente, Patricia Akemi Assato, Felipe Allan da Silva da Costa, Bianca Cechetto Carlos. Mendelics: Bibiana Santos, João Paulo Kitajima, Erika Freitas, David Schlesinger. Hemocentro Ribeirão Preto: Simone Kashima, Evandra Strazza Rodrigues, Svetoslav Nanev Slavov, Elaine Vieira dos Santos, Rafael dos Santos Bezerra, Luiz Carlos Junior de Alcantara, Marta Giovanetti, Vagner Fonseca, Flavia Aburjaile, Rodrigo Tocantins Calado. FAMERP-SJRP: Cecília Artico Banho, Lívia Sacchetto, Fábio Sossai Possebon, Leila Sabrina Ullmann, Cíntia Bittar, Guilherme Campos, Helena Lage Ferreira, Jorge A. Petroli Marchesi, Maisa C. Pereira Parra, Marília Moraes, Paula Rahal, Paulo Inacio da Costa, João Pessoa Araújo Jr., Maurício Lacerda Nogueira. Prefeitura de Sao Paulo: Melissa Palmieri. |
| EPI_ISL_1967242, EPI_ISL_1967243 | VIGILANCIA EPIDEMIOLOGICA DE IBATE | Instituto Butantan / Mendelics | Instituto Butantan: Dimas Tadeu Covas, Sandra Coccuzzo Sampaio, Maria Carolina Elias, José Salvatore Leister Patané, Vincent Louis Viala, Antonio Jorge Martins, Ricardo Haddad, Claudia Renata dos Santos Barros, Elaine Cristina Marqueze, Raul Machado Neto, Debora Botequio Moretti, Jardelina de Souza Todao Bernardino, Loyze Paola Oliveira de Lima, Luiz Aurelio de Campos Crispin. Centro de Genômica Funcional da ESALQ: Luiz Lehmann Coutinho, Ricardo Augusto Brassaloti, Raquel de Lello Rocha Campos Cassano. NGS Soluções Genômicas: Pilar Drummond Sampaio Corrêa Mariani. FZEA-USP Pirassununga: Mirele Daiana Poleti, Jessika Cristina Chagas Lesbon, Elisangela Chicaroni Mattos, Heidge Fukumasu. USP-Botucatu: Rejane Maria Tommasini Grotto, Jayme A. Souza-Neto, Guilherme Targino Valente, Patricia Akemi Assato, Felipe Allan da Silva da Costa, Bianca Cechetto Carlos. Mendelics: Bibiana Santos, João Paulo Kitajima, Erika Freitas, David Schlesinger. Hemocentro Ribeirão Preto: Simone Kashima, Evandra Strazza Rodrigues, Svetoslav Nanev Slavov, Elaine Vieira dos Santos, Rafael dos Santos Bezerra, Luiz Carlos Junior de Alcantara, Marta Giovanetti, Vagner Fonseca, Flavia Aburjaile, Rodrigo Tocantins Calado. FAMERP-SJRP: Cecília Artico Banho, Lívia Sacchetto, Fábio Sossai Possebon, Leila Sabrina Ullmann, Cíntia Bittar, Guilherme Campos, Helena Lage Ferreira, Jorge A. Petroli Marchesi, Maisa C. Pereira Parra, Marília Moraes, Paula Rahal, Paulo Inacio da Costa, João Pessoa Araújo Jr., Maurício Lacerda Nogueira. Prefeitura de Sao Paulo: Melissa Palmieri. |
| EPI_ISL_1967244, EPI_ISL_1967245, EPI_ISL_1967246, EPI_ISL_1967247, EPI_ISL_1967248, EPI_ISL_1967249, EPI_ISL_1967250 | LABORATORIO MUNICIPAL DE ANALISES CLINICAS DE ITANHAEM | Instituto Butantan / Mendelics | Instituto Butantan: Dimas Tadeu Covas, Sandra Coccuzzo Sampaio, Maria Carolina Elias, José Salvatore Leister Patané, Vincent Louis Viala, Antonio Jorge Martins, Ricardo Haddad, Claudia Renata dos Santos Barros, Elaine Cristina Marqueze, Raul Machado Neto, Debora Botequio Moretti, Jardelina de Souza Todao Bernardino, Loyze Paola Oliveira de Lima, Luiz Aurelio de Campos Crispin. Centro de Genômica Funcional da ESALQ: Luiz Lehmann Coutinho, Ricardo Augusto Brassaloti, Raquel de Lello Rocha Campos Cassano. NGS Soluções Genômicas: Pilar Drummond Sampaio Corrêa Mariani. FZEA-USP Pirassununga: Mirele Daiana Poleti, Jessika Cristina Chagas Lesbon, Elisangela Chicaroni Mattos, Heidge Fukumasu. USP-Botucatu: Rejane Maria Tommasini Grotto, Jayme A. Souza-Neto, Guilherme Targino Valente, Patricia Akemi Assato, Felipe Allan da Silva da Costa, Bianca Cechetto Carlos. Mendelics: Bibiana Santos, João Paulo Kitajima, Erika Freitas, David Schlesinger. Hemocentro Ribeirão Preto: Simone Kashima, Evandra Strazza Rodrigues, Svetoslav Nanev Slavov, Elaine Vieira dos Santos, Rafael dos Santos Bezerra, Luiz Carlos Junior de Alcantara, Marta Giovanetti, Vagner Fonseca, Flavia Aburjaile, Rodrigo Tocantins Calado. FAMERP-SJRP: Cecília Artico Banho, Lívia Sacchetto, Fábio Sossai Possebon, Leila Sabrina Ullmann, Cíntia Bittar, Guilherme Campos, Helena Lage Ferreira, Jorge A. Petroli Marchesi, Maisa C. Pereira Parra, Marília Moraes, Paula Rahal, Paulo Inacio da Costa, João Pessoa Araújo Jr., Maurício Lacerda Nogueira. Prefeitura de Sao Paulo: Melissa Palmieri. |
| EPI_ISL_1967251 | CENTRO DE DETENCAO PROVISORIA DE NOVA INDEPENDENCIA | Instituto Butantan / Mendelics | Instituto Butantan: Dimas Tadeu Covas, Sandra Coccuzzo Sampaio, Maria Carolina Elias, José Salvatore Leister Patané, Vincent Louis Viala, Antonio Jorge Martins, Ricardo Haddad, Claudia Renata dos Santos Barros, Elaine Cristina Marqueze, Raul Machado Neto, Debora Botequio Moretti, Jardelina de Souza Todao Bernardino, Loyze Paola Oliveira de Lima, Luiz Aurelio de Campos Crispin. Centro de Genômica Funcional da ESALQ: Luiz Lehmann Coutinho, Ricardo Augusto Brassaloti, Raquel de Lello Rocha Campos Cassano. NGS Soluções Genômicas: Pilar Drummond Sampaio Corrêa Mariani. FZEA-USP Pirassununga: Mirele Daiana Poleti, Jessika Cristina Chagas Lesbon, Elisangela Chicaroni Mattos, Heidge Fukumasu. USP-Botucatu: Rejane Maria Tommasini Grotto, Jayme A. Souza-Neto, Guilherme Targino Valente, Patricia Akemi Assato, Felipe Allan da Silva da Costa, Bianca Cechetto Carlos. Mendelics: Bibiana Santos, João Paulo Kitajima, Erika Freitas, David Schlesinger. Hemocentro Ribeirão Preto: Simone Kashima, Evandra Strazza Rodrigues, Svetoslav Nanev Slavov, Elaine Vieira dos Santos, Rafael dos Santos Bezerra, Luiz Carlos Junior de Alcantara, Marta Giovanetti, Vagner Fonseca, Flavia Aburjaile, Rodrigo Tocantins Calado. FAMERP-SJRP: Cecília Artico Banho, Lívia Sacchetto, Fábio Sossai Possebon, Leila Sabrina Ullmann, Cíntia Bittar, Guilherme Campos, Helena Lage Ferreira, Jorge A. Petroli Marchesi, Maisa C. Pereira Parra, Marília Moraes, Paula Rahal, Paulo Inacio da Costa, João Pessoa Araújo Jr., Maurício Lacerda Nogueira. Prefeitura de Sao Paulo: Melissa Palmieri. |
| EPI_ISL_1967252, EPI_ISL_1967253, EPI_ISL_1967255, EPI_ISL_1967256, EPI_ISL_1967257 | SECRETARIA MUNICIPAL DE SAUDE PORTO FERREIRA | Instituto Butantan / Mendelics | Instituto Butantan: Dimas Tadeu Covas, Sandra Coccuzzo Sampaio, Maria Carolina Elias, José Salvatore Leister Patané, Vincent Louis Viala, Antonio Jorge Martins, Ricardo Haddad, Claudia Renata dos Santos Barros, Elaine Cristina Marqueze, Raul Machado Neto, Debora Botequio Moretti, Jardelina de Souza Todao Bernardino, Loyze Paola Oliveira de Lima, Luiz Aurelio de Campos Crispin. Centro de Genômica Funcional da ESALQ: Luiz Lehmann Coutinho, Ricardo Augusto Brassaloti, Raquel de Lello Rocha Campos Cassano. NGS Soluções Genômicas: Pilar Drummond Sampaio Corrêa Mariani. FZEA-USP Pirassununga: Mirele Daiana Poleti, Jessika Cristina Chagas Lesbon, Elisangela Chicaroni Mattos, Heidge Fukumasu. USP-Botucatu: Rejane Maria Tommasini Grotto, Jayme A. Souza-Neto, Guilherme Targino Valente, Patricia Akemi Assato, Felipe Allan da Silva da Costa, Bianca Cechetto Carlos. Mendelics: Bibiana Santos, João Paulo Kitajima, Erika Freitas, David Schlesinger. Hemocentro Ribeirão Preto: Simone Kashima, Evandra Strazza Rodrigues, Svetoslav Nanev Slavov, Elaine Vieira dos Santos, Rafael dos Santos Bezerra, Luiz Carlos Junior de Alcantara, Marta Giovanetti, Vagner Fonseca, Flavia Aburjaile, Rodrigo Tocantins Calado. FAMERP-SJRP: Cecília Artico Banho, Lívia Sacchetto, Fábio Sossai Possebon, Leila Sabrina Ullmann, Cíntia Bittar, Guilherme Campos, Helena Lage Ferreira, Jorge A. Petroli Marchesi, Maisa C. Pereira Parra, Marília Moraes, Paula Rahal, Paulo Inacio da Costa, João Pessoa Araújo Jr., Maurício Lacerda Nogueira. Prefeitura de Sao Paulo: Melissa Palmieri. |
| EPI_ISL_1967258, EPI_ISL_1967259, EPI_ISL_1967260, EPI_ISL_1967261, EPI_ISL_1967262, EPI_ISL_1967263, EPI_ISL_1967264, EPI_ISL_1967265, EPI_ISL_1967266, EPI_ISL_1967267, EPI_ISL_1967268, EPI_ISL_1967269, EPI_ISL_1967270, EPI_ISL_1967271, EPI_ISL_1967273, EPI_ISL_1967274 | | | |
| see above | SECAO CENTRO DE DIAGNOSTICO SECEDI | Instituto Butantan / Mendelics | Instituto Butantan: Dimas Tadeu Covas, Sandra Coccuzzo Sampaio, Maria Carolina Elias, José Salvatore Leister Patané, Vincent Louis Viala, Antonio Jorge Martins, Ricardo Haddad, Claudia Renata dos Santos Barros, Elaine Cristina Marqueze, Raul Machado Neto, Debora Botequio Moretti, Jardelina de |

| | | | |
|---|---|---|---|
| | | | Souza Todao Bernardino, Loyze Paola Oliveira de Lima, Luiz Aurelio de Campos Crispin. Centro de Genômica Funcional da ESALQ: Luiz Lehmann Coutinho, Ricardo Augusto Brassaloti, Raquel de Lello Rocha Campos Cassano. NGS Soluções Genômicas: Pilar Drummond Sampaio Corrêa Mariani. FZEA-USP Pirassununga: Mirele Daiana Poleti, Jessika Cristina Chagas Lesbon, Elisangela Chicaroni Mattos, Heidge Fukumasu. USP-Botucatu: Rejane Maria Tommasini Grotto, Jayme A. Souza-Neto, Guilherme Targino Valente, Patricia Akemi Assato, Felipe Allan da Silva da Costa, Bianca Cechetto Carlos. Mendelics: Bibiana Santos, João Paulo Kitajima, Erika Freitas, David Schlesinger. Hemocentro Ribeirão Preto: Simone Kashima, Evandra Strazza Rodrigues, Svetoslav Nanev Slavov, Elaine Vieira dos Santos, Rafael dos Santos Bezerra, Luiz Carlos Junior de Alcantara, Marta Giovanetti, Vagner Fonseca, Flavia Aburjaile, Rodrigo Tocantins Calado. FAMERP-SJRP: Cecília Artico Banho, Lívia Sacchetto, Fábio Sossai Possebon, Leila Sabrina Ullmann, Cíntia Bittar, Guilherme Campos, Helena Lage Ferreira, Jorge A. Petroli Marchesi, Maisa C. Pereira Parra, Marília Moraes, Paula Rahal, Paulo Inacio da Costa, João Pessoa Araújo Jr., Maurício Lacerda Nogueira. Prefeitura de Sao Paulo: Melissa Palmieri. |
| EPI_ISL_1967278, EPI_ISL_1967279 | CENTRO DE SAUDE III TABATINGA | Instituto Butantan / Mendelics | Instituto Butantan: Dimas Tadeu Covas, Sandra Coccuzzo Sampaio, Maria Carolina Elias, José Salvatore Leister Patané, Vincent Louis Viala, Antonio Jorge Martins, Ricardo Haddad, Claudia Renata dos Santos Barros, Elaine Cristina Marqueze, Raul Machado Neto, Debora Botequio Moretti, Jardelina de Souza Todao Bernardino, Loyze Paola Oliveira de Lima, Luiz Aurelio de Campos Crispin. Centro de Genômica Funcional da ESALQ: Luiz Lehmann Coutinho, Ricardo Augusto Brassaloti, Raquel de Lello Rocha Campos Cassano. NGS Soluções Genômicas: Pilar Drummond Sampaio Corrêa Mariani. FZEA-USP Pirassununga: Mirele Daiana Poleti, Jessika Cristina Chagas Lesbon, Elisangela Chicaroni Mattos, Heidge Fukumasu. USP-Botucatu: Rejane Maria Tommasini Grotto, Jayme A. Souza-Neto, Guilherme Targino Valente, Patricia Akemi Assato, Felipe Allan da Silva da Costa, Bianca Cechetto Carlos. Mendelics: Bibiana Santos, João Paulo Kitajima, Erika Freitas, David Schlesinger. Hemocentro Ribeirão Preto: Simone Kashima, Evandra Strazza Rodrigues, Svetoslav Nanev Slavov, Elaine Vieira dos Santos, Rafael dos Santos Bezerra, Luiz Carlos Junior de Alcantara, Marta Giovanetti, Vagner Fonseca, Flavia Aburjaile, Rodrigo Tocantins Calado. FAMERP-SJRP: Cecília Artico Banho, Lívia Sacchetto, Fábio Sossai Possebon, Leila Sabrina Ullmann, Cíntia Bittar, Guilherme Campos, Helena Lage Ferreira, Jorge A. Petroli Marchesi, Maisa C. Pereira Parra, Marília Moraes, Paula Rahal, Paulo Inacio da Costa, João Pessoa Araújo Jr., Maurício Lacerda Nogueira. Prefeitura de Sao Paulo: Melissa Palmieri. |
| EPI_ISL_1967280 | UNIDADE BASICA DE SAUDE DE TAQUARAL CAETANO PITELLI | Instituto Butantan / Mendelics | Instituto Butantan: Dimas Tadeu Covas, Sandra Coccuzzo Sampaio, Maria Carolina Elias, José Salvatore Leister Patané, Vincent Louis Viala, Antonio Jorge Martins, Ricardo Haddad, Claudia Renata dos Santos Barros, Elaine Cristina Marqueze, Raul Machado Neto, Debora Botequio Moretti, Jardelina de Souza Todao Bernardino, Loyze Paola Oliveira de Lima, Luiz Aurelio de Campos Crispin. Centro de Genômica Funcional da ESALQ: Luiz Lehmann Coutinho, Ricardo Augusto Brassaloti, Raquel de Lello Rocha Campos Cassano. NGS Soluções Genômicas: Pilar Drummond Sampaio Corrêa Mariani. FZEA-USP Pirassununga: Mirele Daiana Poleti, Jessika Cristina Chagas Lesbon, Elisangela Chicaroni Mattos, Heidge Fukumasu. USP-Botucatu: Rejane Maria Tommasini Grotto, Jayme A. Souza-Neto, Guilherme Targino Valente, Patricia Akemi Assato, Felipe Allan da Silva da Costa, Bianca Cechetto Carlos. Mendelics: Bibiana Santos, João Paulo Kitajima, Erika Freitas, David Schlesinger. Hemocentro Ribeirão Preto: Simone Kashima, Evandra Strazza Rodrigues, Svetoslav Nanev Slavov, Elaine Vieira dos Santos, Rafael dos Santos Bezerra, Luiz Carlos Junior de Alcantara, Marta Giovanetti, Vagner Fonseca, Flavia Aburjaile, Rodrigo Tocantins Calado. FAMERP-SJRP: Cecília Artico Banho, Lívia Sacchetto, Fábio Sossai Possebon, Leila Sabrina Ullmann, Cíntia Bittar, Guilherme Campos, Helena Lage Ferreira, Jorge A. Petroli Marchesi, Maisa C. Pereira Parra, Marília Moraes, Paula Rahal, Paulo Inacio da Costa, João Pessoa Araújo Jr., Maurício Lacerda Nogueira. Prefeitura de Sao Paulo: Melissa Palmieri. |
| EPI_ISL_1967281 | PRONTO ATENDIMENTO MUNICIPAL | Instituto Butantan / Mendelics | Instituto Butantan: Dimas Tadeu Covas, Sandra Coccuzzo Sampaio, Maria Carolina Elias, José Salvatore Leister Patané, Vincent Louis Viala, Antonio Jorge Martins, Ricardo Haddad, Claudia Renata dos Santos Barros, Elaine Cristina Marqueze, Raul Machado Neto, Debora Botequio Moretti, Jardelina de Souza Todao Bernardino, Loyze Paola Oliveira de Lima, Luiz Aurelio de Campos Crispin. Centro de Genômica Funcional da ESALQ: Luiz Lehmann Coutinho, Ricardo Augusto Brassaloti, Raquel de Lello Rocha Campos Cassano. NGS Soluções Genômicas: Pilar Drummond Sampaio Corrêa Mariani. FZEA-USP Pirassununga: Mirele Daiana Poleti, Jessika Cristina Chagas Lesbon, Elisangela Chicaroni Mattos, Heidge Fukumasu. USP-Botucatu: Rejane Maria Tommasini Grotto, Jayme A. Souza-Neto, Guilherme Targino Valente, Patricia Akemi Assato, Felipe Allan da Silva da Costa, Bianca Cechetto Carlos. Mendelics: Bibiana Santos, João Paulo Kitajima, Erika Freitas, David Schlesinger. Hemocentro Ribeirão Preto: Simone Kashima, Evandra Strazza Rodrigues, Svetoslav Nanev Slavov, Elaine Vieira dos Santos, Rafael dos Santos Bezerra, Luiz Carlos Junior de Alcantara, Marta Giovanetti, Vagner Fonseca, Flavia Aburjaile, Rodrigo Tocantins Calado. FAMERP-SJRP: Cecília Artico Banho, Lívia Sacchetto, Fábio Sossai Possebon, Leila Sabrina Ullmann, Cíntia Bittar, Guilherme Campos, Helena Lage Ferreira, Jorge A. Petroli Marchesi, Maisa C. Pereira Parra, Marília Moraes, Paula Rahal, Paulo Inacio da Costa, João Pessoa Araújo Jr., Maurício Lacerda Nogueira. Prefeitura de Sao Paulo: Melissa Palmieri. |
| EPI_ISL_1967282 | SANTA CASA DE ATIBAIA PRO SAUDE | Instituto Butantan / Mendelics | Instituto Butantan: Dimas Tadeu Covas, Sandra Coccuzzo Sampaio, Maria Carolina Elias, José Salvatore Leister Patané, Vincent Louis Viala, Antonio Jorge Martins, Ricardo Haddad, Claudia Renata dos Santos Barros, Elaine Cristina Marqueze, Raul Machado Neto, Debora Botequio Moretti, Jardelina de Souza Todao Bernardino, Loyze Paola Oliveira de Lima, Luiz Aurelio de Campos Crispin. Centro de Genômica Funcional da ESALQ: Luiz Lehmann Coutinho, Ricardo Augusto Brassaloti, Raquel de Lello Rocha Campos Cassano. NGS Soluções Genômicas: Pilar Drummond Sampaio Corrêa Mariani. FZEA-USP Pirassununga: Mirele Daiana Poleti, Jessika Cristina Chagas Lesbon, Elisangela Chicaroni Mattos, Heidge Fukumasu. USP-Botucatu: Rejane Maria Tommasini Grotto, Jayme A. Souza-Neto, Guilherme Targino Valente, Patricia Akemi Assato, Felipe Allan da Silva da Costa, Bianca Cechetto Carlos. Mendelics: Bibiana Santos, João Paulo Kitajima, Erika Freitas, David Schlesinger. Hemocentro Ribeirão Preto: Simone Kashima, Evandra Strazza Rodrigues, Svetoslav Nanev Slavov, Elaine Vieira dos Santos, Rafael dos Santos Bezerra, Luiz Carlos Junior de Alcantara, Marta Giovanetti, Vagner Fonseca, Flavia Aburjaile, Rodrigo Tocantins Calado. FAMERP-SJRP: Cecília Artico Banho, Lívia Sacchetto, Fábio Sossai Possebon, Leila Sabrina Ullmann, Cíntia Bittar, Guilherme Campos, Helena Lage Ferreira, Jorge A. Petroli Marchesi, Maisa C. Pereira Parra, Marília Moraes, Paula Rahal, Paulo Inacio da Costa, João Pessoa Araújo Jr., Maurício Lacerda Nogueira. Prefeitura de Sao Paulo: Melissa Palmieri. |
| EPI_ISL_1967283 | VIGILANCIA EM SAUDE DE FCO MORATO | Instituto Butantan / Mendelics | Instituto Butantan: Dimas Tadeu Covas, Sandra Coccuzzo Sampaio, Maria Carolina Elias, José Salvatore Leister Patané, Vincent Louis Viala, Antonio Jorge Martins, Ricardo Haddad, Claudia Renata dos Santos Barros, Elaine Cristina Marqueze, Raul Machado Neto, Debora Botequio Moretti, Jardelina de Souza Todao Bernardino, Loyze Paola Oliveira de Lima, Luiz Aurelio de Campos Crispin. Centro de Genômica Funcional da ESALQ: Luiz Lehmann Coutinho, Ricardo Augusto Brassaloti, Raquel de Lello Rocha Campos Cassano. NGS Soluções Genômicas: Pilar Drummond Sampaio Corrêa Mariani. FZEA-USP Pirassununga: Mirele Daiana Poleti, Jessika Cristina Chagas Lesbon, Elisangela Chicaroni Mattos, Heidge Fukumasu. USP-Botucatu: Rejane Maria Tommasini Grotto, Jayme A. Souza-Neto, Guilherme Targino Valente, Patricia Akemi Assato, Felipe Allan da Silva da Costa, Bianca Cechetto Carlos. Mendelics: Bibiana Santos, João Paulo Kitajima, Erika Freitas, David Schlesinger. Hemocentro Ribeirão Preto: Simone Kashima, Evandra Strazza Rodrigues, Svetoslav Nanev Slavov, Elaine Vieira dos Santos, Rafael dos Santos Bezerra, Luiz Carlos Junior de Alcantara, Marta Giovanetti, Vagner Fonseca, Flavia Aburjaile, Rodrigo Tocantins Calado. FAMERP-SJRP: Cecília Artico Banho, Lívia Sacchetto, Fábio Sossai Possebon, Leila Sabrina Ullmann, Cíntia Bittar, Guilherme Campos, Helena Lage Ferreira, Jorge A. Petroli Marchesi, Maisa C. Pereira Parra, Marília Moraes, Paula Rahal, Paulo Inacio da Costa, João Pessoa Araújo Jr., Maurício Lacerda Nogueira. Prefeitura de Sao Paulo: Melissa Palmieri. |
| EPI_ISL_1967284 | UBS RECREIO SAO JORGE | Instituto Butantan / Mendelics | Instituto Butantan: Dimas Tadeu Covas, Sandra Coccuzzo Sampaio, Maria Carolina Elias, José Salvatore Leister Patané, Vincent Louis Viala, Antonio Jorge Martins, Ricardo Haddad, Claudia Renata dos Santos Barros, Elaine Cristina Marqueze, Raul Machado Neto, Debora Botequio Moretti, Jardelina de Souza Todao Bernardino, Loyze Paola Oliveira de Lima, Luiz Aurelio de Campos Crispin. Centro de Genômica Funcional da ESALQ: Luiz Lehmann Coutinho, Ricardo Augusto Brassaloti, Raquel de Lello Rocha Campos Cassano. NGS Soluções Genômicas: Pilar Drummond Sampaio Corrêa Mariani. FZEA-USP Pirassununga: Mirele Daiana Poleti, Jessika Cristina Chagas Lesbon, Elisangela Chicaroni Mattos, Heidge Fukumasu. USP-Botucatu: Rejane Maria Tommasini Grotto, Jayme A. Souza-Neto, Guilherme Targino Valente, Patricia Akemi Assato, Felipe Allan da Silva da Costa, Bianca Cechetto Carlos. Mendelics: Bibiana Santos, João Paulo Kitajima, Erika Freitas, David Schlesinger. Hemocentro Ribeirão Preto: Simone Kashima, Evandra Strazza Rodrigues, Svetoslav Nanev Slavov, Elaine Vieira dos Santos, Rafael dos Santos Bezerra, Luiz Carlos Junior de Alcantara, Marta Giovanetti, Vagner Fonseca, Flavia Aburjaile, Rodrigo Tocantins Calado. FAMERP-SJRP: Cecília Artico Banho, Lívia Sacchetto, Fábio Sossai Possebon, Leila Sabrina Ullmann, Cíntia Bittar, Guilherme Campos, Helena Lage Ferreira, Jorge A. Petroli Marchesi, Maisa C. Pereira Parra, Marília Moraes, Paula Rahal, Paulo Inacio da Costa, João Pessoa Araújo Jr., Maurício Lacerda Nogueira. Prefeitura de Sao Paulo: Melissa Palmieri. |
| EPI_ISL_1967285, EPI_ISL_1967286 | UBS JARDIM CABUCU | Instituto Butantan / Mendelics | Instituto Butantan: Dimas Tadeu Covas, Sandra Coccuzzo Sampaio, Maria Carolina Elias, José Salvatore Leister Patané, Vincent Louis Viala, Antonio Jorge Martins, Ricardo Haddad, Claudia Renata dos Santos Barros, Elaine Cristina Marqueze, Raul Machado Neto, Debora Botequio Moretti, Jardelina de Souza Todao Bernardino, Loyze Paola Oliveira de Lima, Luiz Aurelio de Campos Crispin. Centro de Genômica Funcional da ESALQ: Luiz Lehmann Coutinho, Ricardo Augusto Brassaloti, Raquel de Lello Rocha Campos Cassano. NGS Soluções Genômicas: Pilar Drummond Sampaio Corrêa Mariani. FZEA-USP Pirassununga: Mirele Daiana Poleti, Jessika Cristina Chagas Lesbon, Elisangela Chicaroni Mattos, Heidge Fukumasu. USP-Botucatu: Rejane Maria Tommasini Grotto, Jayme A. Souza-Neto, Guilherme Targino Valente, Patricia Akemi Assato, Felipe Allan da Silva da Costa, Bianca Cechetto Carlos. Mendelics: Bibiana Santos, João Paulo Kitajima, Erika Freitas, David Schlesinger. Hemocentro Ribeirão Preto: Simone Kashima, Evandra Strazza Rodrigues, Svetoslav Nanev Slavov, Elaine Vieira dos Santos, Rafael dos Santos Bezerra, Luiz Carlos Junior de Alcantara, Marta Giovanetti, Vagner Fonseca, Flavia Aburjaile, Rodrigo Tocantins Calado. FAMERP-SJRP: Cecília Artico Banho, Lívia Sacchetto, Fábio Sossai Possebon, Leila Sabrina |

| | | | |
|---|---|---|---|
| | | | Ullmann, Cíntia Bittar, Guilherme Campos, Helena Lage Ferreira, Jorge A. Petroli Marchesi, Maisa C. Pereira Parra, Marília Moraes, Paula Rahal, Paulo Inacio da Costa, João Pessoa Araújo Jr., Maurício Lacerda Nogueira. Prefeitura de Sao Paulo: Melissa Palmieri. |
| EPI_ISL_1967287 | UBS VILA GALVAO | Instituto Butantan / Mendelics | Instituto Butantan: Dimas Tadeu Covas, Sandra Coccuzzo Sampaio, Maria Carolina Elias, José Salvatore Leister Patané, Vincent Louis Viala, Antonio Jorge Martins, Ricardo Haddad, Claudia Renata dos Santos Barros, Elaine Cristina Marqueze, Raul Machado Neto, Debora Botequio Moretti, Jardelina de Souza Todao Bernardino, Loyze Paola Oliveira de Lima, Luiz Aurelio de Campos Crispin. Centro de Genômica Funcional da ESALQ: Luiz Lehmann Coutinho, Ricardo Augusto Brassaloti, Raquel de Lello Rocha Campos Cassano. NGS Soluções Genômicas: Pilar Drummond Sampaio Corrêa Mariani. FZEA-USP Pirassununga: Mirele Daiana Poleti, Jessika Cristina Chagas Lesbon, Elisangela Chicaroni Mattos, Heidge Fukumasu. USP-Botucatu: Rejane Maria Tommasini Grotto, Jayme A. Souza-Neto, Guilherme Targino Valente, Patricia Akemi Assato, Felipe Allan da Silva da Costa, Bianca Cechetto Carlos. Mendelics: Bibiana Santos, João Paulo Kitajima, Erika Freitas, David Schlesinger. Hemocentro Ribeirão Preto: Simone Kashima, Evandra Strazza Rodrigues, Svetoslav Nanev Slavov, Elaine Vieira dos Santos, Rafael dos Santos Bezerra, Luiz Carlos Junior de Alcantara, Marta Giovanetti, Vagner Fonseca, Flavia Aburjaile, Rodrigo Tocantins Calado. FAMERP-SJRP: Cecília Artico Banho, Lívia Sacchetto, Fábio Sossai Possebon, Leila Sabrina Ullmann, Cíntia Bittar, Guilherme Campos, Helena Lage Ferreira, Jorge A. Petroli Marchesi, Maisa C. Pereira Parra, Marília Moraes, Paula Rahal, Paulo Inacio da Costa, João Pessoa Araújo Jr., Maurício Lacerda Nogueira. Prefeitura de Sao Paulo: Melissa Palmieri. |
| EPI_ISL_1967288 | UBS CAVADAS | Instituto Butantan / Mendelics | Instituto Butantan: Dimas Tadeu Covas, Sandra Coccuzzo Sampaio, Maria Carolina Elias, José Salvatore Leister Patané, Vincent Louis Viala, Antonio Jorge Martins, Ricardo Haddad, Claudia Renata dos Santos Barros, Elaine Cristina Marqueze, Raul Machado Neto, Debora Botequio Moretti, Jardelina de Souza Todao Bernardino, Loyze Paola Oliveira de Lima, Luiz Aurelio de Campos Crispin. Centro de Genômica Funcional da ESALQ: Luiz Lehmann Coutinho, Ricardo Augusto Brassaloti, Raquel de Lello Rocha Campos Cassano. NGS Soluções Genômicas: Pilar Drummond Sampaio Corrêa Mariani. FZEA-USP Pirassununga: Mirele Daiana Poleti, Jessika Cristina Chagas Lesbon, Elisangela Chicaroni Mattos, Heidge Fukumasu. USP-Botucatu: Rejane Maria Tommasini Grotto, Jayme A. Souza-Neto, Guilherme Targino Valente, Patricia Akemi Assato, Felipe Allan da Silva da Costa, Bianca Cechetto Carlos. Mendelics: Bibiana Santos, João Paulo Kitajima, Erika Freitas, David Schlesinger. Hemocentro Ribeirão Preto: Simone Kashima, Evandra Strazza Rodrigues, Svetoslav Nanev Slavov, Elaine Vieira dos Santos, Rafael dos Santos Bezerra, Luiz Carlos Junior de Alcantara, Marta Giovanetti, Vagner Fonseca, Flavia Aburjaile, Rodrigo Tocantins Calado. FAMERP-SJRP: Cecília Artico Banho, Lívia Sacchetto, Fábio Sossai Possebon, Leila Sabrina Ullmann, Cíntia Bittar, Guilherme Campos, Helena Lage Ferreira, Jorge A. Petroli Marchesi, Maisa C. Pereira Parra, Marília Moraes, Paula Rahal, Paulo Inacio da Costa, João Pessoa Araújo Jr., Maurício Lacerda Nogueira. Prefeitura de Sao Paulo: Melissa Palmieri. |
| EPI_ISL_1967289, EPI_ISL_1967290, EPI_ISL_1967291, EPI_ISL_1967292, EPI_ISL_1967293 | UNIDADE RESPIRATORIA NOVA HORTOLANDIA | Instituto Butantan / Mendelics | Instituto Butantan: Dimas Tadeu Covas, Sandra Coccuzzo Sampaio, Maria Carolina Elias, José Salvatore Leister Patané, Vincent Louis Viala, Antonio Jorge Martins, Ricardo Haddad, Claudia Renata dos Santos Barros, Elaine Cristina Marqueze, Raul Machado Neto, Debora Botequio Moretti, Jardelina de Souza Todao Bernardino, Loyze Paola Oliveira de Lima, Luiz Aurelio de Campos Crispin. Centro de Genômica Funcional da ESALQ: Luiz Lehmann Coutinho, Ricardo Augusto Brassaloti, Raquel de Lello Rocha Campos Cassano. NGS Soluções Genômicas: Pilar Drummond Sampaio Corrêa Mariani. FZEA-USP Pirassununga: Mirele Daiana Poleti, Jessika Cristina Chagas Lesbon, Elisangela Chicaroni Mattos, Heidge Fukumasu. USP-Botucatu: Rejane Maria Tommasini Grotto, Jayme A. Souza-Neto, Guilherme Targino Valente, Patricia Akemi Assato, Felipe Allan da Silva da Costa, Bianca Cechetto Carlos. Mendelics: Bibiana Santos, João Paulo Kitajima, Erika Freitas, David Schlesinger. Hemocentro Ribeirão Preto: Simone Kashima, Evandra Strazza Rodrigues, Svetoslav Nanev Slavov, Elaine Vieira dos Santos, Rafael dos Santos Bezerra, Luiz Carlos Junior de Alcantara, Marta Giovanetti, Vagner Fonseca, Flavia Aburjaile, Rodrigo Tocantins Calado. FAMERP-SJRP: Cecília Artico Banho, Lívia Sacchetto, Fábio Sossai Possebon, Leila Sabrina Ullmann, Cíntia Bittar, Guilherme Campos, Helena Lage Ferreira, Jorge A. Petroli Marchesi, Maisa C. Pereira Parra, Marília Moraes, Paula Rahal, Paulo Inacio da Costa, João Pessoa Araújo Jr., Maurício Lacerda Nogueira. Prefeitura de Sao Paulo: Melissa Palmieri. |
| EPI_ISL_1967294, EPI_ISL_1967295 | UPA II JUNDIAI | Instituto Butantan / Mendelics | Instituto Butantan: Dimas Tadeu Covas, Sandra Coccuzzo Sampaio, Maria Carolina Elias, José Salvatore Leister Patané, Vincent Louis Viala, Antonio Jorge Martins, Ricardo Haddad, Claudia Renata dos Santos Barros, Elaine Cristina Marqueze, Raul Machado Neto, Debora Botequio Moretti, Jardelina de Souza Todao Bernardino, Loyze Paola Oliveira de Lima, Luiz Aurelio de Campos Crispin. Centro de Genômica Funcional da ESALQ: Luiz Lehmann Coutinho, Ricardo Augusto Brassaloti, Raquel de Lello Rocha Campos Cassano. NGS Soluções Genômicas: Pilar Drummond Sampaio Corrêa Mariani. FZEA-USP Pirassununga: Mirele Daiana Poleti, Jessika Cristina Chagas Lesbon, Elisangela Chicaroni Mattos, Heidge Fukumasu. USP-Botucatu: Rejane Maria Tommasini Grotto, Jayme A. Souza-Neto, Guilherme Targino Valente, Patricia Akemi Assato, Felipe Allan da Silva da Costa, Bianca Cechetto Carlos. Mendelics: Bibiana Santos, João Paulo Kitajima, Erika Freitas, David Schlesinger. Hemocentro Ribeirão Preto: Simone Kashima, Evandra Strazza Rodrigues, Svetoslav Nanev Slavov, Elaine Vieira dos Santos, Rafael dos Santos Bezerra, Luiz Carlos Junior de Alcantara, Marta Giovanetti, Vagner Fonseca, Flavia Aburjaile, Rodrigo Tocantins Calado. FAMERP-SJRP: Cecília Artico Banho, Lívia Sacchetto, Fábio Sossai Possebon, Leila Sabrina Ullmann, Cíntia Bittar, Guilherme Campos, Helena Lage Ferreira, Jorge A. Petroli Marchesi, Maisa C. Pereira Parra, Marília Moraes, Paula Rahal, Paulo Inacio da Costa, João Pessoa Araújo Jr., Maurício Lacerda Nogueira. Prefeitura de Sao Paulo: Melissa Palmieri. |
| EPI_ISL_1967296, EPI_ISL_1967297 | AMBULATORIO DE ESPECIALIDADE V E MOGI MIRIM | Instituto Butantan / Mendelics | Instituto Butantan: Dimas Tadeu Covas, Sandra Coccuzzo Sampaio, Maria Carolina Elias, José Salvatore Leister Patané, Vincent Louis Viala, Antonio Jorge Martins, Ricardo Haddad, Claudia Renata dos Santos Barros, Elaine Cristina Marqueze, Raul Machado Neto, Debora Botequio Moretti, Jardelina de Souza Todao Bernardino, Loyze Paola Oliveira de Lima, Luiz Aurelio de Campos Crispin. Centro de Genômica Funcional da ESALQ: Luiz Lehmann Coutinho, Ricardo Augusto Brassaloti, Raquel de Lello Rocha Campos Cassano. NGS Soluções Genômicas: Pilar Drummond Sampaio Corrêa Mariani. FZEA-USP Pirassununga: Mirele Daiana Poleti, Jessika Cristina Chagas Lesbon, Elisangela Chicaroni Mattos, Heidge Fukumasu. USP-Botucatu: Rejane Maria Tommasini Grotto, Jayme A. Souza-Neto, Guilherme Targino Valente, Patricia Akemi Assato, Felipe Allan da Silva da Costa, Bianca Cechetto Carlos. Mendelics: Bibiana Santos, João Paulo Kitajima, Erika Freitas, David Schlesinger. Hemocentro Ribeirão Preto: Simone Kashima, Evandra Strazza Rodrigues, Svetoslav Nanev Slavov, Elaine Vieira dos Santos, Rafael dos Santos Bezerra, Luiz Carlos Junior de Alcantara, Marta Giovanetti, Vagner Fonseca, Flavia Aburjaile, Rodrigo Tocantins Calado. FAMERP-SJRP: Cecília Artico Banho, Lívia Sacchetto, Fábio Sossai Possebon, Leila Sabrina Ullmann, Cíntia Bittar, Guilherme Campos, Helena Lage Ferreira, Jorge A. Petroli Marchesi, Maisa C. Pereira Parra, Marília Moraes, Paula Rahal, Paulo Inacio da Costa, João Pessoa Araújo Jr., Maurício Lacerda Nogueira. Prefeitura de Sao Paulo: Melissa Palmieri. |
| EPI_ISL_1967298 | UPA VILA SANTA CATARINA | Instituto Butantan / Mendelics | Instituto Butantan: Dimas Tadeu Covas, Sandra Coccuzzo Sampaio, Maria Carolina Elias, José Salvatore Leister Patané, Vincent Louis Viala, Antonio Jorge Martins, Ricardo Haddad, Claudia Renata dos Santos Barros, Elaine Cristina Marqueze, Raul Machado Neto, Debora Botequio Moretti, Jardelina de Souza Todao Bernardino, Loyze Paola Oliveira de Lima, Luiz Aurelio de Campos Crispin. Centro de Genômica Funcional da ESALQ: Luiz Lehmann Coutinho, Ricardo Augusto Brassaloti, Raquel de Lello Rocha Campos Cassano. NGS Soluções Genômicas: Pilar Drummond Sampaio Corrêa Mariani. FZEA-USP Pirassununga: Mirele Daiana Poleti, Jessika Cristina Chagas Lesbon, Elisangela Chicaroni Mattos, Heidge Fukumasu. USP-Botucatu: Rejane Maria Tommasini Grotto, Jayme A. Souza-Neto, Guilherme Targino Valente, Patricia Akemi Assato, Felipe Allan da Silva da Costa, Bianca Cechetto Carlos. Mendelics: Bibiana Santos, João Paulo Kitajima, Erika Freitas, David Schlesinger. Hemocentro Ribeirão Preto: Simone Kashima, Evandra Strazza Rodrigues, Svetoslav Nanev Slavov, Elaine Vieira dos Santos, Rafael dos Santos Bezerra, Luiz Carlos Junior de Alcantara, Marta Giovanetti, Vagner Fonseca, Flavia Aburjaile, Rodrigo Tocantins Calado. FAMERP-SJRP: Cecília Artico Banho, Lívia Sacchetto, Fábio Sossai Possebon, Leila Sabrina Ullmann, Cíntia Bittar, Guilherme Campos, Helena Lage Ferreira, Jorge A. Petroli Marchesi, Maisa C. Pereira Parra, Marília Moraes, Paula Rahal, Paulo Inacio da Costa, João Pessoa Araújo Jr., Maurício Lacerda Nogueira. Prefeitura de Sao Paulo: Melissa Palmieri. |
| EPI_ISL_1967299 | HOSPITAL E PRONTO SOCORRO PORTINARI | Instituto Butantan / Mendelics | Instituto Butantan: Dimas Tadeu Covas, Sandra Coccuzzo Sampaio, Maria Carolina Elias, José Salvatore Leister Patané, Vincent Louis Viala, Antonio Jorge Martins, Ricardo Haddad, Claudia Renata dos Santos Barros, Elaine Cristina Marqueze, Raul Machado Neto, Debora Botequio Moretti, Jardelina de Souza Todao Bernardino, Loyze Paola Oliveira de Lima, Luiz Aurelio de Campos Crispin. Centro de Genômica Funcional da ESALQ: Luiz Lehmann Coutinho, Ricardo Augusto Brassaloti, Raquel de Lello Rocha Campos Cassano. NGS Soluções Genômicas: Pilar Drummond Sampaio Corrêa Mariani. FZEA-USP Pirassununga: Mirele Daiana Poleti, Jessika Cristina Chagas Lesbon, Elisangela Chicaroni Mattos, Heidge Fukumasu. USP-Botucatu: Rejane Maria Tommasini Grotto, Jayme A. Souza-Neto, Guilherme Targino Valente, Patricia Akemi Assato, Felipe Allan da Silva da Costa, Bianca Cechetto Carlos. Mendelics: Bibiana Santos, João Paulo Kitajima, Erika Freitas, David Schlesinger. Hemocentro Ribeirão Preto: Simone Kashima, Evandra Strazza Rodrigues, Svetoslav Nanev Slavov, Elaine Vieira dos Santos, Rafael dos Santos Bezerra, Luiz Carlos Junior de Alcantara, Marta Giovanetti, Vagner Fonseca, Flavia Aburjaile, Rodrigo Tocantins Calado. FAMERP-SJRP: Cecília Artico Banho, Lívia Sacchetto, Fábio Sossai Possebon, Leila Sabrina Ullmann, Cíntia Bittar, Guilherme Campos, Helena Lage Ferreira, Jorge A. Petroli Marchesi, Maisa C. Pereira Parra, Marília Moraes, Paula Rahal, Paulo Inacio da Costa, João Pessoa Araújo Jr., Maurício Lacerda Nogueira. Prefeitura de Sao Paulo: Melissa Palmieri. |
| EPI_ISL_1967300, EPI_ISL_1967301, EPI_ISL_1967302, EPI_ISL_1967303 | SECRETARIA MUNICIPAL DE SAUDE | Instituto Butantan / Mendelics | Instituto Butantan: Dimas Tadeu Covas, Sandra Coccuzzo Sampaio, Maria Carolina Elias, José Salvatore Leister Patané, Vincent Louis Viala, Antonio Jorge Martins, Ricardo Haddad, Claudia Renata dos Santos Barros, Elaine Cristina Marqueze, Raul Machado Neto, Debora Botequio Moretti, Jardelina de Souza Todao Bernardino, Loyze Paola Oliveira de Lima, Luiz Aurelio de Campos Crispin. Centro de Genômica Funcional da ESALQ: Luiz Lehmann Coutinho, Ricardo Augusto Brassaloti, Raquel de Lello Rocha Campos Cassano. NGS Soluções Genômicas: Pilar Drummond Sampaio Corrêa Mariani. |

| | | | |
|---|---|---|---|
| | | | FZEA-USP Pirassununga: Mirele Daiana Poleti, Jessika Cristina Chagas Lesbon, Elisangela Chicaroni Mattos, Heidge Fukumasu. USP-Botucatu: Rejane Maria Tommasini Grotto, Jayme A. Souza-Neto, Guilherme Targino Valente, Patricia Akemi Assato, Felipe Allan da Silva da Costa, Bianca Cechetto Carlos. Mendelics: Bibiana Santos, João Paulo Kitajima, Erika Freitas, David Schlesinger. Hemocentro Ribeirão Preto: Simone Kashima, Evandra Strazza Rodrigues, Svetoslav Nanev Slavov, Elaine Vieira dos Santos, Rafael dos Santos Bezerra, Luiz Carlos Junior de Alcantara, Marta Giovanetti, Vagner Fonseca, Flavia Aburjaile, Rodrigo Tocantins Calado. FAMERP-SJRP: Cecília Artico Banho, Lívia Sacchetto, Fábio Sossai Possebon, Leila Sabrina Ullmann, Cíntia Bittar, Guilherme Campos, Helena Lage Ferreira, Jorge A. Petroli Marchesi, Maisa C. Pereira Parra, Marília Moraes, Paula Rahal, Paulo Inacio da Costa, João Pessoa Araújo Jr., Maurício Lacerda Nogueira. Prefeitura de Sao Paulo: Melissa Palmieri. |
| EPI_ISL_1967305 | PSF PARQUE EMILIA | Instituto Butantan / Mendelics | Instituto Butantan: Dimas Tadeu Covas, Sandra Coccuzzo Sampaio, Maria Carolina Elias, José Salvatore Leister Patané, Vincent Louis Viala, Antonio Jorge Martins, Ricardo Haddad, Claudia Renata dos Santos Barros, Elaine Cristina Marqueze, Raul Machado Neto, Debora Botequio Moretti, Jardelina de Souza Todao Bernardino, Loyze Paola Oliveira de Lima, Luiz Aurelio de Campos Crispin. Centro de Genômica Funcional da ESALQ: Luiz Lehmann Coutinho, Ricardo Augusto Brassaloti, Raquel de Lello Rocha Campos Cassano. NGS Soluções Genômicas: Pilar Drummond Sampaio Corrêa Mariani. FZEA-USP Pirassununga: Mirele Daiana Poleti, Jessika Cristina Chagas Lesbon, Elisangela Chicaroni Mattos, Heidge Fukumasu. USP-Botucatu: Rejane Maria Tommasini Grotto, Jayme A. Souza-Neto, Guilherme Targino Valente, Patricia Akemi Assato, Felipe Allan da Silva da Costa, Bianca Cechetto Carlos. Mendelics: Bibiana Santos, João Paulo Kitajima, Erika Freitas, David Schlesinger. Hemocentro Ribeirão Preto: Simone Kashima, Evandra Strazza Rodrigues, Svetoslav Nanev Slavov, Elaine Vieira dos Santos, Rafael dos Santos Bezerra, Luiz Carlos Junior de Alcantara, Marta Giovanetti, Vagner Fonseca, Flavia Aburjaile, Rodrigo Tocantins Calado. FAMERP-SJRP: Cecília Artico Banho, Lívia Sacchetto, Fábio Sossai Possebon, Leila Sabrina Ullmann, Cíntia Bittar, Guilherme Campos, Helena Lage Ferreira, Jorge A. Petroli Marchesi, Maisa C. Pereira Parra, Marília Moraes, Paula Rahal, Paulo Inacio da Costa, João Pessoa Araújo Jr., Maurício Lacerda Nogueira. Prefeitura de Sao Paulo: Melissa Palmieri. |
| EPI_ISL_1967306, EPI_ISL_1967307, EPI_ISL_1967308, EPI_ISL_1967309 | UPA DR AKIRA TADA | Instituto Butantan / Mendelics | Instituto Butantan: Dimas Tadeu Covas, Sandra Coccuzzo Sampaio, Maria Carolina Elias, José Salvatore Leister Patané, Vincent Louis Viala, Antonio Jorge Martins, Ricardo Haddad, Claudia Renata dos Santos Barros, Elaine Cristina Marqueze, Raul Machado Neto, Debora Botequio Moretti, Jardelina de Souza Todao Bernardino, Loyze Paola Oliveira de Lima, Luiz Aurelio de Campos Crispin. Centro de Genômica Funcional da ESALQ: Luiz Lehmann Coutinho, Ricardo Augusto Brassaloti, Raquel de Lello Rocha Campos Cassano. NGS Soluções Genômicas: Pilar Drummond Sampaio Corrêa Mariani. FZEA-USP Pirassununga: Mirele Daiana Poleti, Jessika Cristina Chagas Lesbon, Elisangela Chicaroni Mattos, Heidge Fukumasu. USP-Botucatu: Rejane Maria Tommasini Grotto, Jayme A. Souza-Neto, Guilherme Targino Valente, Patricia Akemi Assato, Felipe Allan da Silva da Costa, Bianca Cechetto Carlos. Mendelics: Bibiana Santos, João Paulo Kitajima, Erika Freitas, David Schlesinger. Hemocentro Ribeirão Preto: Simone Kashima, Evandra Strazza Rodrigues, Svetoslav Nanev Slavov, Elaine Vieira dos Santos, Rafael dos Santos Bezerra, Luiz Carlos Junior de Alcantara, Marta Giovanetti, Vagner Fonseca, Flavia Aburjaile, Rodrigo Tocantins Calado. FAMERP-SJRP: Cecília Artico Banho, Lívia Sacchetto, Fábio Sossai Possebon, Leila Sabrina Ullmann, Cíntia Bittar, Guilherme Campos, Helena Lage Ferreira, Jorge A. Petroli Marchesi, Maisa C. Pereira Parra, Marília Moraes, Paula Rahal, Paulo Inacio da Costa, João Pessoa Araújo Jr., Maurício Lacerda Nogueira. Prefeitura de Sao Paulo: Melissa Palmieri. |
| EPI_ISL_1967310, EPI_ISL_1967311 | PRONTO SOCORRO MUNICIPAL TAMBAU | Instituto Butantan / Mendelics | Instituto Butantan: Dimas Tadeu Covas, Sandra Coccuzzo Sampaio, Maria Carolina Elias, José Salvatore Leister Patané, Vincent Louis Viala, Antonio Jorge Martins, Ricardo Haddad, Claudia Renata dos Santos Barros, Elaine Cristina Marqueze, Raul Machado Neto, Debora Botequio Moretti, Jardelina de Souza Todao Bernardino, Loyze Paola Oliveira de Lima, Luiz Aurelio de Campos Crispin. Centro de Genômica Funcional da ESALQ: Luiz Lehmann Coutinho, Ricardo Augusto Brassaloti, Raquel de Lello Rocha Campos Cassano. NGS Soluções Genômicas: Pilar Drummond Sampaio Corrêa Mariani. FZEA-USP Pirassununga: Mirele Daiana Poleti, Jessika Cristina Chagas Lesbon, Elisangela Chicaroni Mattos, Heidge Fukumasu. USP-Botucatu: Rejane Maria Tommasini Grotto, Jayme A. Souza-Neto, Guilherme Targino Valente, Patricia Akemi Assato, Felipe Allan da Silva da Costa, Bianca Cechetto Carlos. Mendelics: Bibiana Santos, João Paulo Kitajima, Erika Freitas, David Schlesinger. Hemocentro Ribeirão Preto: Simone Kashima, Evandra Strazza Rodrigues, Svetoslav Nanev Slavov, Elaine Vieira dos Santos, Rafael dos Santos Bezerra, Luiz Carlos Junior de Alcantara, Marta Giovanetti, Vagner Fonseca, Flavia Aburjaile, Rodrigo Tocantins Calado. FAMERP-SJRP: Cecília Artico Banho, Lívia Sacchetto, Fábio Sossai Possebon, Leila Sabrina Ullmann, Cíntia Bittar, Guilherme Campos, Helena Lage Ferreira, Jorge A. Petroli Marchesi, Maisa C. Pereira Parra, Marília Moraes, Paula Rahal, Paulo Inacio da Costa, João Pessoa Araújo Jr., Maurício Lacerda Nogueira. Prefeitura de Sao Paulo: Melissa Palmieri. |
| EPI_ISL_1967312, EPI_ISL_1967313, EPI_ISL_1967314 | UNIDADE DE PRONTO ATENDIMENTO DE VARZEA PAULISTA UPA II | Instituto Butantan / Mendelics | Instituto Butantan: Dimas Tadeu Covas, Sandra Coccuzzo Sampaio, Maria Carolina Elias, José Salvatore Leister Patané, Vincent Louis Viala, Antonio Jorge Martins, Ricardo Haddad, Claudia Renata dos Santos Barros, Elaine Cristina Marqueze, Raul Machado Neto, Debora Botequio Moretti, Jardelina de Souza Todao Bernardino, Loyze Paola Oliveira de Lima, Luiz Aurelio de Campos Crispin. Centro de Genômica Funcional da ESALQ: Luiz Lehmann Coutinho, Ricardo Augusto Brassaloti, Raquel de Lello Rocha Campos Cassano. NGS Soluções Genômicas: Pilar Drummond Sampaio Corrêa Mariani. FZEA-USP Pirassununga: Mirele Daiana Poleti, Jessika Cristina Chagas Lesbon, Elisangela Chicaroni Mattos, Heidge Fukumasu. USP-Botucatu: Rejane Maria Tommasini Grotto, Jayme A. Souza-Neto, Guilherme Targino Valente, Patricia Akemi Assato, Felipe Allan da Silva da Costa, Bianca Cechetto Carlos. Mendelics: Bibiana Santos, João Paulo Kitajima, Erika Freitas, David Schlesinger. Hemocentro Ribeirão Preto: Simone Kashima, Evandra Strazza Rodrigues, Svetoslav Nanev Slavov, Elaine Vieira dos Santos, Rafael dos Santos Bezerra, Luiz Carlos Junior de Alcantara, Marta Giovanetti, Vagner Fonseca, Flavia Aburjaile, Rodrigo Tocantins Calado. FAMERP-SJRP: Cecília Artico Banho, Lívia Sacchetto, Fábio Sossai Possebon, Leila Sabrina Ullmann, Cíntia Bittar, Guilherme Campos, Helena Lage Ferreira, Jorge A. Petroli Marchesi, Maisa C. Pereira Parra, Marília Moraes, Paula Rahal, Paulo Inacio da Costa, João Pessoa Araújo Jr., Maurício Lacerda Nogueira. Prefeitura de Sao Paulo: Melissa Palmieri. |
| EPI_ISL_2003111 | Pronto Socorro Central de Bauru | Instituto Adolfo Lutz, Interdiciplinary Procedures Center, Strategic Laboratory | Claudio Tavares Sacchi, Claudia Regina Gonçalves, Erica Valessa Ramos Gomes, Karoline Rodrigues Campos, Caio Vinicius Dias Lopes, Leonardo Jose Tadeu de Araujo |
| EPI_ISL_2003112 | CS de Paulo de Faria | Instituto Adolfo Lutz, Interdiciplinary Procedures Center, Strategic Laboratory | Claudio Tavares Sacchi, Claudia Regina Gonçalves, Erica Valessa Ramos Gomes, Karoline Rodrigues Campos, Caio Vinicius Dias Lopes, Leonardo Jose Tadeu de Araujo |
| EPI_ISL_2003113 | Hospital Municipal Dr Mario Gatti Campinas | Instituto Adolfo Lutz, Interdiciplinary Procedures Center, Strategic Laboratory | Claudio Tavares Sacchi, Claudia Regina Gonçalves, Erica Valessa Ramos Gomes, Karoline Rodrigues Campos, Caio Vinicius Dias Lopes, Leonardo Jose Tadeu de Araujo |
| EPI_ISL_2003114, EPI_ISL_2003115, EPI_ISL_2003116, EPI_ISL_2003117, EPI_ISL_2003118, EPI_ISL_2003119, EPI_ISL_2003120, EPI_ISL_2003121, EPI_ISL_2003122, EPI_ISL_2003123, EPI_ISL_2003124, EPI_ISL_2003125, EPI_ISL_2003126, EPI_ISL_2003127, EPI_ISL_2003128, EPI_ISL_2003129, EPI_ISL_2003130, EPI_ISL_2003131, EPI_ISL_2003132, EPI_ISL_2003133 | | | |
| see above | Instituto Adolfo Lutz Central | Instituto Adolfo Lutz, Interdiciplinary Procedures Center, Strategic Laboratory | Claudio Tavares Sacchi, Claudia Regina Gonçalves, Erica Valessa Ramos Gomes, Karoline Rodrigues Campos, Caio Vinicius Dias Lopes, Leonardo Jose Tadeu de Araujo |
| EPI_ISL_2003134, EPI_ISL_2003135, EPI_ISL_2003136, EPI_ISL_2003137, EPI_ISL_2003138, EPI_ISL_2003139, EPI_ISL_2003140, EPI_ISL_2003141, EPI_ISL_2003142, EPI_ISL_2003143, EPI_ISL_2003144, EPI_ISL_2003145, EPI_ISL_2003146, EPI_ISL_2003147, EPI_ISL_2003148, EPI_ISL_2003149 | | | |
| see above | Instituto Adolfo Lutz - Regional de Santos | Instituto Adolfo Lutz, Interdiciplinary Procedures Center, Strategic Laboratory | Claudio Tavares Sacchi, Claudia Regina Gonçalves, Erica Valessa Ramos Gomes, Karoline Rodrigues Campos, Caio Vinicius Dias Lopes, Leonardo Jose Tadeu de Araujo |
| EPI_ISL_2003151 | Instituto Adolfo Lutz - Regional de Bauru | Instituto Adolfo Lutz, Interdiciplinary Procedures Center, Strategic Laboratory | Claudio Tavares Sacchi, Claudia Regina Gonçalves, Erica Valessa Ramos Gomes, Karoline Rodrigues Campos, Caio Vinicius Dias Lopes, Leonardo Jose Tadeu de Araujo |
| EPI_ISL_2003152 | Instituto Adolfo Lutz - Regional de Marilia | Instituto Adolfo Lutz, Interdiciplinary Procedures Center, Strategic Laboratory | Claudio Tavares Sacchi, Claudia Regina Gonçalves, Erica Valessa Ramos Gomes, Karoline Rodrigues Campos, Caio Vinicius Dias Lopes, Leonardo Jose Tadeu de Araujo |
| EPI_ISL_2003162, EPI_ISL_2003163, EPI_ISL_2003164, EPI_ISL_2003166, EPI_ISL_2003167, EPI_ISL_2003168, EPI_ISL_2003169 | Instituto Adolfo Lutz - Regional de Campinas | Instituto Adolfo Lutz, Interdiciplinary Procedures Center, Strategic Laboratory | Claudio Tavares Sacchi, Claudia Regina Gonçalves, Erica Valessa Ramos Gomes, Karoline Rodrigues Campos, Caio Vinicius Dias Lopes, Leonardo Jose Tadeu de Araujo |
| EPI_ISL_2008906, EPI_ISL_2008907, EPI_ISL_2008908, EPI_ISL_2008909, EPI_ISL_2008910, EPI_ISL_2008911, EPI_ISL_2008912, EPI_ISL_2008913, EPI_ISL_2008914, EPI_ISL_2008915, EPI_ISL_2008916, EPI_ISL_2008917, EPI_ISL_2008918, EPI_ISL_2008919, EPI_ISL_2008920, EPI_ISL_2008921, EPI_ISL_2008922, EPI_ISL_2008923, EPI_ISL_2008924, EPI_ISL_2008925, EPI_ISL_2008926, EPI_ISL_2008927, EPI_ISL_2008928, EPI_ISL_2008929, EPI_ISL_2008930, EPI_ISL_2008931, EPI_ISL_2008932, EPI_ISL_2008933, EPI_ISL_2008934, EPI_ISL_2008935, EPI_ISL_2008938, EPI_ISL_2008940, EPI_ISL_2008941, EPI_ISL_2008944, EPI_ISL_2008945, EPI_ISL_2008946, EPI_ISL_2008947, EPI_ISL_2008948, EPI_ISL_2008949, EPI_ISL_2008950, EPI_ISL_2008951, EPI_ISL_2008952, EPI_ISL_2008953, EPI_ISL_2008954, EPI_ISL_2008955, EPI_ISL_2008956, EPI_ISL_2008957, EPI_ISL_2008958, EPI_ISL_2008959, EPI_ISL_2008960, EPI_ISL_2008961, EPI_ISL_2008962, EPI_ISL_2008963, EPI_ISL_2008964, EPI_ISL_2008965, EPI_ISL_2008966, EPI_ISL_2008967 | | | |
| see above | Laboratorio de Pesquisa em Virologia, FAMERP, SJRP | Laboratorio de Pesquisa em Virologia, FAMERP, SJRP | Cecília Artico Banho; Lívia Sacchetto; Fábio Sossai Possebon; Leila Sabrina Ullmann; Cíntia Bittar; Guilherme Campos; Helena Lage Ferreira; Jorge A. Petroli Marchesi; Maisa C. Pereira Parra; Marília Moraes; Paula Rahal; Paulo Inacio da Costa; João Pessoa Araújo Jr.; Maurício L. Nogueira. |
| EPI_ISL_2017282, EPI_ISL_2017283, EPI_ISL_2017284, EPI_ISL_2017285, EPI_ISL_2017286, EPI_ISL_2017287, EPI_ISL_2017288, EPI_ISL_2017290, EPI_ISL_2017292, EPI_ISL_2017293, EPI_ISL_2017294, EPI_ISL_2017295, EPI_ISL_2017296, EPI_ISL_2017297, EPI_ISL_2017298, EPI_ISL_2017299, EPI_ISL_2017300, | | | |

| | | | |
|---|---|---|---|
| EPI_ISL_2017301, EPI_ISL_2017303, EPI_ISL_2017304, EPI_ISL_2017305, EPI_ISL_2017324, EPI_ISL_2017325, EPI_ISL_2017326, EPI_ISL_2017327, EPI_ISL_2017328, EPI_ISL_2017329, EPI_ISL_2017330, EPI_ISL_2017331, EPI_ISL_2017332, EPI_ISL_2017333, EPI_ISL_2017334, EPI_ISL_2017335, EPI_ISL_2017336, EPI_ISL_2017337, EPI_ISL_2017338, EPI_ISL_2017339, EPI_ISL_2017340, EPI_ISL_2017341, EPI_ISL_2017342, EPI_ISL_2017343, EPI_ISL_2017344, EPI_ISL_2017345, EPI_ISL_2017346, EPI_ISL_2017347, EPI_ISL_2017348, EPI_ISL_2017349, EPI_ISL_2017350, EPI_ISL_2017351, EPI_ISL_2017352, EPI_ISL_2017353, EPI_ISL_2017354, EPI_ISL_2017355, EPI_ISL_2017356, EPI_ISL_2017357, EPI_ISL_2017358, EPI_ISL_2017359, EPI_ISL_2017360 | | | |
| see above | HLAGYN - Laboratorio de Imunologia de Transplantes de Goias | HLAGYN - Laboratorio de Imunologia de Transplantes de Goias | Fernando Antonio Vinhal dos Santos, Erika Lopes Rocha Batista, Alessandro Leonardo Alvares Magalhaes, Raphael Bessa Parmigiane, Frederico Rodrigues Vinhal, Sabrina Sara Moreira Duarte, Danielle de Paiva Rezende, Lucas Carlos Gomes Pereira, Paola Cristina Resende Silva |
| EPI_ISL_2017361, EPI_ISL_2017362, EPI_ISL_2017363, EPI_ISL_2017364, EPI_ISL_2017365, EPI_ISL_2017366, EPI_ISL_2017367, EPI_ISL_2017368, EPI_ISL_2017369, EPI_ISL_2017370, EPI_ISL_2017371, EPI_ISL_2017372, EPI_ISL_2017373, EPI_ISL_2017374, EPI_ISL_2017375, EPI_ISL_2017376, EPI_ISL_2017377, EPI_ISL_2017378, EPI_ISL_2017379, EPI_ISL_2017380, EPI_ISL_2017381, EPI_ISL_2017382, EPI_ISL_2017383, EPI_ISL_2017384, EPI_ISL_2017385, EPI_ISL_2017386, EPI_ISL_2017387, EPI_ISL_2017388, EPI_ISL_2017389, EPI_ISL_2017390, EPI_ISL_2017391, EPI_ISL_2017392, EPI_ISL_2017393, EPI_ISL_2017394, EPI_ISL_2017395, EPI_ISL_2017396, EPI_ISL_2017397, EPI_ISL_2017398, EPI_ISL_2017399, EPI_ISL_2017400, EPI_ISL_2017401, EPI_ISL_2017402, EPI_ISL_2017403, EPI_ISL_2017404, EPI_ISL_2017405, EPI_ISL_2017406, EPI_ISL_2017407, EPI_ISL_2017408, EPI_ISL_2017409, EPI_ISL_2017410, EPI_ISL_2017411, EPI_ISL_2017412, EPI_ISL_2017413, EPI_ISL_2017414, EPI_ISL_2017415, EPI_ISL_2017416, EPI_ISL_2017417, EPI_ISL_2017418, EPI_ISL_2017419, EPI_ISL_2017420, EPI_ISL_2017421, EPI_ISL_2017423, EPI_ISL_2017424, EPI_ISL_2017425, EPI_ISL_2017426, EPI_ISL_2017427, EPI_ISL_2017428, EPI_ISL_2017429, EPI_ISL_2017431, EPI_ISL_2017432, EPI_ISL_2017433, EPI_ISL_2017434, EPI_ISL_2017435, EPI_ISL_2017436, EPI_ISL_2017437, EPI_ISL_2017438, EPI_ISL_2017439, EPI_ISL_2017440, EPI_ISL_2017446, EPI_ISL_2017448, EPI_ISL_2017451, EPI_ISL_2017452, EPI_ISL_2017453, EPI_ISL_2017454, EPI_ISL_2017456, EPI_ISL_2017459, EPI_ISL_2017462, EPI_ISL_2017463, EPI_ISL_2017464, EPI_ISL_2017467, EPI_ISL_2017468, EPI_ISL_2017469, EPI_ISL_2017470, EPI_ISL_2017471, EPI_ISL_2017472, EPI_ISL_2017473, EPI_ISL_2017474, EPI_ISL_2017475, EPI_ISL_2017476, EPI_ISL_2017477, EPI_ISL_2017479, EPI_ISL_2017480, EPI_ISL_2017481, EPI_ISL_2017482, EPI_ISL_2017483, EPI_ISL_2017484 | | | |
| see above | HLAGYN - Laboratorio de Imunologia de Transplantes de Goias | HLAGYN - Laboratorio de Imunologia de Transplantes de Goias | Fernando Antonio Vinhal dos Santos, Erika Lopes Rocha Batista, Alessandro Leonardo Alvares Magalhaes, Frederico Rodrigues Vinhal, Sabrina Sara Moreira Duarte, Danielle de Paiva Rezende, Lucas Carlos Gomes Pereira, Paola Cristina Resende Silva |
| EPI_ISL_2086585, EPI_ISL_2086587, EPI_ISL_2086588, EPI_ISL_2086589, EPI_ISL_2086590, EPI_ISL_2086591, EPI_ISL_2086592, EPI_ISL_2086593, EPI_ISL_2086594, EPI_ISL_2086595, EPI_ISL_2086596, EPI_ISL_2086597, EPI_ISL_2086598, EPI_ISL_2086599, EPI_ISL_2086600, EPI_ISL_2086603, EPI_ISL_2086604, EPI_ISL_2086605, EPI_ISL_2086607, EPI_ISL_2086608, EPI_ISL_2086609, EPI_ISL_2086610, EPI_ISL_2086611, EPI_ISL_2086612, EPI_ISL_2086613, EPI_ISL_2086614, EPI_ISL_2086615, EPI_ISL_2086616, EPI_ISL_2086617, EPI_ISL_2086618, EPI_ISL_2086619, EPI_ISL_2086620, EPI_ISL_2086621, EPI_ISL_2086622, EPI_ISL_2086763, EPI_ISL_2086764, EPI_ISL_2086765, EPI_ISL_2086766, EPI_ISL_2086767 | | | |
| see above | LABCOVID_HCPA | LABRESIS_HCPA | Wink PL, Martins AF, Volpato F, Monteiro F, Zavascki AP, Barth AL |
| EPI_ISL_2249447, EPI_ISL_2249448, EPI_ISL_2249449, EPI_ISL_2249450, EPI_ISL_2249451, EPI_ISL_2249452, EPI_ISL_2249454, EPI_ISL_2249455, EPI_ISL_2249456, EPI_ISL_2249457 | Laboratório Central de Saúde Pública do Paraá | Coordenação Geral de Laboratórios de Saúde Pública (CGLAB/DAEVS/SVS/MS) | Vagner Fonseca, et al. |
| EPI_ISL_2344233, EPI_ISL_2344235, EPI_ISL_2344236, EPI_ISL_2344242, EPI_ISL_2344244, EPI_ISL_2344258, EPI_ISL_2344259, EPI_ISL_2344260, EPI_ISL_2344262, EPI_ISL_2344265, EPI_ISL_2344271, EPI_ISL_2344272, EPI_ISL_2344273, EPI_ISL_2344275, EPI_ISL_2344420, EPI_ISL_2344421, EPI_ISL_2344425, EPI_ISL_2344426, EPI_ISL_2344428, EPI_ISL_2344431, EPI_ISL_2344440, EPI_ISL_2344450, EPI_ISL_2344451, EPI_ISL_2344455, EPI_ISL_2344456, EPI_ISL_2344457, EPI_ISL_2425433 | | | |
| see above | Instituto Butantan | Instituto de Medicina Tropical de Sao Paulo | Brazil-UK Centre for Arbovirus Discovery Diagnosis Genomics and Epidemiology (CADDE) Genomic Network - Instituto de Medicina Tropical |
| EPI_ISL_476282, EPI_ISL_476312, EPI_ISL_476313, EPI_ISL_476318, EPI_ISL_476370 | DB Diagnósticos do Brasil | Instituto de Medicina Tropical da Univesidade de São Paulo | Samples: Nelson Gaburo Jr; Sequencing: Ingra Morales Claro, Jaqueline Goes de Jesus, Erika Regina Manuli, Flavia Cristina da Silva Sales, Thais de Moura Coletti, Camila Alves Maia da Silva, Mariana Severo Ramundo, Giula Magalhaes Ferreira, Darlan da Silva Candido, Julien Theze, Nuno Faria, Ester Sabino |
| EPI_ISL_510535 | Molecular Virology, Instituto Carlos Chagas / Fiocruz Paraná | Universidade Federal do Parana (UFPR) | Suzukawa,A., Tscha,M., Zanluca,C., Raboni,S., Duarte dos Santos,C. |
| EPI_ISL_722135 | DB Diagnosticos do Brasil | Laboratório de Parasitologia Médica - Instituto de Medicina Tropical - Universidade de São Paulo | Brazil-UK Centre for Arbovirus Discovery Diagnosis Genomics and Epidemiology (CADDE) Genomic Network - Instituto de Medicina Tropical |
| EPI_ISL_804814, EPI_ISL_804815, EPI_ISL_804819, EPI_ISL_804820, EPI_ISL_804821, EPI_ISL_804823, EPI_ISL_804824 | DB Diagnosticos do Brasil | Laboratório de Parasitologia Médica - Instituto de Medicina Tropical - Universidade de São Paulo | Nuno Faria, Ingra Morales Claro, Darlan Candido, Lucas A. Moyses Franco, Pamela dos Santos Andrade, Thais de Moura Coletti, Camila A. Maia da Silva, Flavia Cristina Sales, Erika Regina Manuli, Renato A. Santana, Nelson Gaburo, Cecília da Cunha Camilo, Nelson Abrahim Fraiji, Myuki Alfaia Esashika Crispim, Maria do Perpétuo Socorro Sampaio Carvalho, Andrew Rambaut, Nick Loman, Oliver G. Pybus, Ester C. Sabino; DB; HEMOAM; CDL; CADDE Genomic Network. |
| EPI_ISL_833167, EPI_ISL_833169, EPI_ISL_833171, EPI_ISL_833172, EPI_ISL_833173, EPI_ISL_833174 | DB Diagnosticos do Brasil | Instituto Adolfo Lutz, Interdiciplinary Procedures Center, Strategic Laboratory | Claudio Tavares Sacchi, Claudia Regina Gonçalves, Erica Valessa Ramos Gomes, Karoline Rodrigues Campos |
| EPI_ISL_872191, EPI_ISL_872192 | Conjunto Hospitalar do Mandaqui de Sao Paulo | Instituto Adolfo Lutz, Interdiciplinary Procedures Center, Strategic Laboratory | Claudio Tavares Sacchi, Claudia Regina Gonçalves, Erica Valessa Ramos Gomes, Karoline Rodrigues Campos, Katia Correa de Oliveira Santos, Ana Lucia de Carvalho Avelino, Fabiana Cristina Pereira dos Santos |
| EPI_ISL_875688 | National Influenza Center - Instituto Adolfo Lutz | Instituto Adolfo Lutz, Interdiciplinary Procedures Center, Strategic Laboratory | Claudio Tavares Sacchi, Claudia Regina Gonçalves, Erica Valessa Ramos Gomes, Karoline Rodrigues Campos, Katia Correa de Oliveira Santos, Ana Lucia de Carvalho Avelino, Clovis Roberto Abe Constantino |
| EPI_ISL_875689 | Hospital do Servidor Publico | Instituto Adolfo Lutz, Interdiciplinary Procedures Center, Strategic Laboratory | Claudio Tavares Sacchi, Claudia Regina Gonçalves, Erica Valessa Ramos Gomes, Karoline Rodrigues Campos |
| EPI_ISL_904120, EPI_ISL_904121 | LACEN - Laboratório Central de Saúde Pública do Pará | Evandro Chagas Institute | Santos, M.C.; Silva, A.M.; Junior, W.D.C.; Barbagelata, L.S.; Ferreira, J.A.; Sousa, E.M.A.; da Silva, P.S.; Pinheiro, K.C.; L.C.; Sousa Junior, E.C. |
| EPI_ISL_906068, EPI_ISL_906069 | Instituto Adolfo Lutz - Regional de Campinas | Instituto Adolfo Lutz, Interdiciplinary Procedures Center, Strategic Laboratory | Claudio Tavares Sacchi, Claudia Regina Gonçalves, Erica Valessa Ramos Gomes, Karoline Rodrigues Campos |
| EPI_ISL_906071 | LACEN-PI DR. Costa Alvarenga | Instituto Adolfo Lutz, Interdiciplinary Procedures Center, Strategic Laboratory | Claudio Tavares Sacchi, Claudia Regina Gonçalves, Erica Valessa Ramos Gomes, Karoline Rodrigues Campos |
| EPI_ISL_906075 | Hospital Geral de Vila Penteado Dr Jose Pangella Sao Paulo | Instituto Adolfo Lutz, Interdiciplinary Procedures Center, Strategic Laboratory | Claudio Tavares Sacchi, Claudia Regina Gonçalves, Erica Valessa Ramos Gomes, Karoline Rodrigues Campos |
| EPI_ISL_906076, EPI_ISL_906077 | Hospital Sao Luiz Sao Caetano | Instituto Adolfo Lutz, Interdiciplinary Procedures Center, Strategic Laboratory | Claudio Tavares Sacchi, Claudia Regina Gonçalves, Erica Valessa Ramos Gomes, Karoline Rodrigues Campos |
| EPI_ISL_906080, EPI_ISL_906081 | Hospital Beneficiencia Portuguesa | Instituto Adolfo Lutz, Interdiciplinary Procedures Center, Strategic Laboratory | Claudio Tavares Sacchi, Claudia Regina Gonçalves, Erica Valessa Ramos Gomes, Karoline Rodrigues Campos |
| EPI_ISL_918499, EPI_ISL_918500, EPI_ISL_918502, EPI_ISL_918503, EPI_ISL_918504, EPI_ISL_918505, EPI_ISL_918506, EPI_ISL_918507, EPI_ISL_918508, EPI_ISL_918509, EPI_ISL_918510, EPI_ISL_918511 | | | |
| see above | LACEN - Laboratório Central de Saúde Pública do Amazonas | Evandro Chagas Institute | Santos, M.C.; Silva, A.M.; Junior, W.D.C.; Barbagelata, L.S.; Ferreira, J.A.; Sousa, E.M.A.; da Silva, P.S.; Pinheiro, K.C.; L.C.; Sousa Junior, E.C. |
| EPI_ISL_940614, EPI_ISL_940615, EPI_ISL_940616, EPI_ISL_940617, EPI_ISL_940618 | LACEN-PI DR. Costa Alvarenga | Instituto Adolfo Lutz, Interdiciplinary Procedures Center, Strategic Laboratory | Claudio Tavares Sacchi, Claudia Regina Gonçalves, Erica Valessa Ramos Gomes, Karoline Rodrigues Campos |
| EPI_ISL_940619, EPI_ISL_940620, EPI_ISL_940621, EPI_ISL_940622, EPI_ISL_940623, EPI_ISL_940624, EPI_ISL_940625 | Hospital Sao Joaquim - Beneficiencia Portuguesa | Instituto Adolfo Lutz, Interdiciplinary Procedures Center, Strategic Laboratory | Claudio Tavares Sacchi, Claudia Regina Gonçalves, Erica Valessa Ramos Gomes, Karoline Rodrigues Campos |
| EPI_ISL_940626, EPI_ISL_940627 | Hospital Central Sao Caetano do Sul | Instituto Adolfo Lutz, Interdiciplinary Procedures Center, Strategic Laboratory | Claudio Tavares Sacchi, Claudia Regina Gonçalves, Erica Valessa Ramos Gomes, Karoline Rodrigues Campos |
| EPI_ISL_940630, EPI_ISL_943967, EPI_ISL_943968, EPI_ISL_943969, EPI_ISL_943970, EPI_ISL_943971, EPI_ISL_943972 | Hospital Geral de Sao Paulo | Instituto Adolfo Lutz, Interdiciplinary Procedures Center, Strategic Laboratory | Claudio Tavares Sacchi, Claudia Regina Gonçalves, Erica Valessa Ramos Gomes, Karoline Rodrigues Campos |
| EPI_ISL_943987 | LACEN do Estado de Tocantins | Instituto Adolfo Lutz, Interdiciplinary Procedures Center, Strategic Laboratory | Claudio Tavares Sacchi, Claudia Regina Gonçalves, Erica Valessa Ramos Gomes, Karoline Rodrigues Campos |

| | | | |
|---|---|---|---|
| EPI_ISL_943990 | LACEN do Estado de Goias | Instituto Adolfo Lutz, Interdiciplinary Procedures Center, Strategic Laboratory | Claudio Tavares Sacchi, Claudia Regina Gonçalves, Erica Valessa Ramos Gomes, Karoline Rodrigues Campos |
| EPI_ISL_981383, EPI_ISL_981385, EPI_ISL_981387 | IAL Regional de Bauru | Instituto Adolfo Lutz, Interdiciplinary Procedures Center, Strategic Laboratory | Claudio Tavares Sacchi, Claudia Regina Gonçalves, Erica Valessa Ramos Gomes, Karoline Rodrigues Campos |
| EPI_ISL_983865, EPI_ISL_984619, EPI_ISL_984620, EPI_ISL_984621 | Central Laboratory of Public Health of Rio Grande do Sul (Lacen-RS) | State Center for Health Surveillance of the Health Department of the State of Rio Grande do Sul (CEVS/SES-RS) | Aline Campos, Cynthia Molina, Lara Crescente, Leticia Garay, Ludmila Fiorenzano Baethgen, Richard Salvato, Tatiana Gregianini |
| EPI_ISL_985303, EPI_ISL_985304, EPI_ISL_985305, EPI_ISL_985306, EPI_ISL_985307, EPI_ISL_985308, EPI_ISL_985309, EPI_ISL_985310, EPI_ISL_985311, EPI_ISL_985312, EPI_ISL_985313, EPI_ISL_985314, EPI_ISL_985315, EPI_ISL_985316, EPI_ISL_985317 | | | |
| see above | LACEN do Estado de Goias | Instituto Adolfo Lutz, Interdiciplinary Procedures Center, Strategic Laboratory | Claudio Tavares Sacchi, Claudia Regina Gonçalves, Erica Valessa Ramos Gomes, Karoline Rodrigues Campos |
| EPI_ISL_985318, EPI_ISL_985319 | LACEN de Santa Catarina | Instituto Adolfo Lutz, Interdiciplinary Procedures Center, Strategic Laboratory | Claudio Tavares Sacchi, Claudia Regina Gonçalves, Erica Valessa Ramos Gomes, Karoline Rodrigues Campos |
